# The effects on global health outcomes of switching from regular salt to potassium-enriched salt: a modelling study

**DOI:** 10.64898/2026.04.06.26350270

**Authors:** Liping Huang, Xiaoyue Xu, Kunihiro Matsushita, Tammy M. Brady, Lawrence J. Appel, Ewout J. Hoorn, Aletta E. Schutte, Jason HY Wu, Maoyi Tian, Leopold N Aminde, Kathy Trieu, Bruce Neal, Matti Marklund

## Abstract

**Objective:** To estimate the benefit and risk of replacing regular salt with potassium-enriched salt.

**Design:** Comparative risk assessment modelling.

**Setting:** Worldwide

**Participants:** Adult populations aged 25 and above.

**Intervention:** (1) worldwide replacement of all salt (discretionary salt used for seasoning or cooking in the home, and non-discretionary salt used in processed and restaurant foods); (2) worldwide replacement of just discretionary salt; (3) worldwide replacement of just non-discretionary salt; (4) replacement of discretionary salt just for people with diagnosed hypertension; and (5) replacement of discretionary salt just for people with treated hypertension.

**Main outcome measures:** For scenarios 1-3, we estimated benefits including deaths, new cases and disability-adjusted-life-years (DALYs) from cardiovascular disease and chronic kidney disease (CKD), from blood pressure-lowering as well as harms (CVD deaths) caused by hyperkalaemia among people with CKD stages G3-G5.

**Results:** Replacement of all salt worldwide could prevent 2.96 (95% uncertainty interval 2.81-3.12) million deaths, 10.17 (9.59-10.70) million new cases of disease and 69.43 (65.61-72.92) million disability-adjusted life years (DALYs) each year. These figures represent 14.6%, 13.1% and 16.5% of the annual global disease burden attributable to CVD and CKD. Replacement of all discretionary salt (1.85, 1.74-1.97 million deaths) would have a greater impact on mortality than replacement of all non-discretionary salt (1.56, 1.46-1.67 million deaths). In people with CKD Stage G3-G5, there would be a net benefit - replacement of all salt would prevent 0.75 (0.71-0.80) million deaths but might cause 0.10 (0.09-0.11) million deaths from hyperkalaemia. Discretionary salt replacement only among diagnosed or treated hypertensives would prevent 0.59 (0.55-0.63) million and 0.48 (0.45-0.52) million deaths, respectively.

**Conclusion:** Switching regular salt to potassium-enriched salt appears to offer large potential for health gains under diverse scenarios, including for people with CKD.

**Funding:** This work did not receive specific funding.

**What is already known on this topic:**

- Excess dietary sodium and low dietary potassium intake both cause high blood pressure, which causes a significant burden of cardiovascular disease (CVD) and chronic kidney disease (CKD).
- Efforts to cut dietary sodium intake as a strategy to control blood pressure have mostly been unsuccessful, with no country expected to meet the World Health Organisation (WHO) 2025 goal of reducing sodium intake by 30%.
- Switching regular salt to potassium-enriched salt has shown clear protection against CVD and causes minimal change in taste and has low cost to scale up, but concerns remain about the potential of causing deaths due to hyperkalaemia among people with advanced chronic kidney disease.

**What this study adds:**

- The study modelled five different possible approaches to the implementation of potassium-enriched salt that will suit a range of different circumstances, including countries with different levels of discretionary versus non-discretionary salt intake.
- The study indicates switching to potassium-enriched salt can prevent very large numbers of CVD and CKD events worldwide, while the potential for causing harm in people with CKD is small in comparison.
- There was also net benefit in analyses restricted to just people with CKD, where benefits of blood pressure lowering outweigh potential harms from hyperkalaemia.

## INTRODUCTION

Cardiovascular disease (CVD) remains the leading cause of death globally and chronic kidney disease (CKD) is projected to become the fifth leading cause of years of life lost by 2040.^1^ High blood pressure (BP) is a leading modifiable risk factor for CVD and CKD accounting for more than 10 million cardiovascular deaths and almost 0.5 million CKD deaths in 2021.^2^ Reducing sodium intake and increasing potassium intake are two dietary interventions that reduce BP. Current average global sodium intake (4.3 g/day) exceeds the World Health Organization (WHO) recommendation of 2.0 g/day^3^ and the Global Burden of Disease Study (GBD) identifies excess sodium intake as the leading dietary risk for CVD.^4^ In the meantime, average global potassium intake is 2.3 g/day,^5^ which is significantly below the WHO’s recommended minimum intake of 3.5 g/day.^6^ In 2013 the member states of the United Nations committed to reducing population sodium intake by 30% by 2025.^3^ Despite significant focus by the WHO and member states, no country is projected to meet this goal.^3^

The key challenge with current efforts focused on cutting sodium intake is that they require acceptance of a less salty taste and this has proved largely insurmountable for consumers and the food industry worldwide.^3^ By contrast, switching from regular salt to a potassium-enriched salt substitute can be done with no detectable effect on taste for most people and excellent overall acceptability.^7^ Beneficial effects of potassium-enriched salt on BP are long-established and recent large trials and overviews of the trials have shown parallel reductions in the risks of major cardiovascular events and premature death.^8,9^ In the largest and longest trial done to date, 92% of participants in the intervention group were still using a product with composition 25% potassium chloride and 75% sodium chloride after 5 years.^8^

Switching to potassium-enriched salt reduces BP by jointly lowering sodium intake and increasing potassium intake. A potential adverse consequence of increasing potassium intake is the risk of hyperkalaemia in people with CKD. To date, the majority of trials have asked participants to self-exclude if they are at elevated risk of hyperkalaemia and there is no evidence of clinical harm in any study or overview.^9^ One trial that included all participants from elderly care facilities, irrespective of kidney function, showed an increased risk of biochemical hyperkalaemia but there was no increase in adverse clinical outcomes.^10^

Prior modelling has reported large net health gains with potassium-enriched salt for China,^11^ India^12^ and Indonesia.^13^ The aim of this study was to estimate the benefits and potential harms of a switch from regular salt to potassium-enriched salt under a range of different possible intervention scenarios for 193 countries.

## METHODS

### Ethics

This modelling study used only published aggregated data and was not reviewed by an ethics committee.

### Research design

This study employed a comparative risk assessment framework adapted from our prior modelling studies.^11,12^ The model (**Figure 1**) first estimated the impact of strategies for replacing regular salt with potassium-enriched salt on dietary sodium and potassium intake for each country. The estimation incorporated baseline intake levels of sodium and the proportion of sodium intake from discretionary salt (i.e., used for seasoning or cooking in the home) versus other sources of sodium that are non-discretionary (i.e., in processed and restaurant foods including salty sauces and condiments) for each country. For every intervention scenario the composition of the potassium-enriched salt was fixed as 75% sodium chloride and 25% potassium chloride since this is the ratio for which there is best evidence of clinical benefits and acceptability.^8^ Based on the effects on dietary sodium and potassium intake, the model projected the impact on systolic BP (SBP) and the subsequent effect of SBP change on the risks of CVD and CKD. Additionally, the model estimated the increase in serum potassium among people with CKD stages G3-5, (eGFR<60) and the potential number of cardiovascular deaths caused by hyperkalaemia in this group.

**Figure 1.**
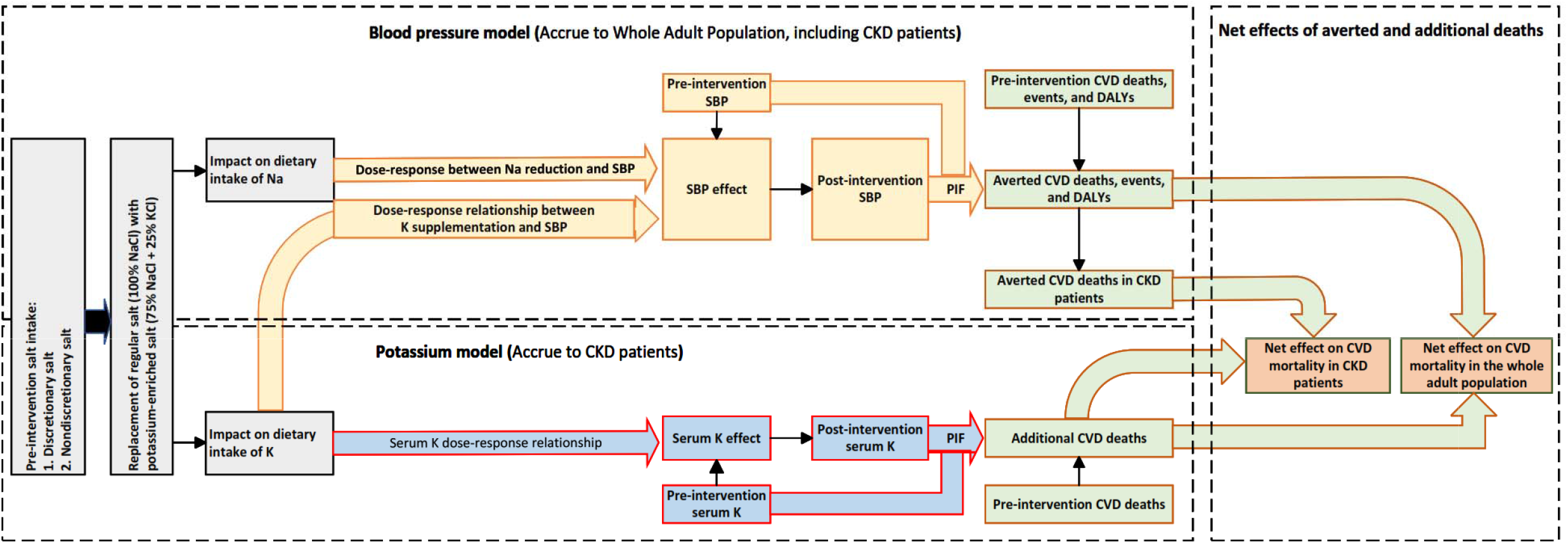
Conceptual framework for modelling the impact of using potassium-enriched salt on health outcomes. Model inputs and detailed information are presented in Supplementary Table 2. Discretionary salt: salt used for seasoning or cooking in the home Nondiscretionary salt: salt in processed and restaurant foods

### Participants

For each of the 193 countries and territories, the adult population (aged 25+ years) was divided into five-year age intervals from 25-29 to 90-94 years, with an additional group of 95+ years. Younger age groups were not included due to their much lower burden of CVD and CKD, limited data on sodium and potassium intake, and uncertainties about the effects of sodium and potassium on SBP and risks of CVD and CKD. The analyses were done separately for women and men and then combined.

### Intervention scenarios

We estimated the impact of a switch to potassium-enriched salt under five scenarios: (1) worldwide replacement of all salt (including both discretionary salt and non-discretionary salt; (2) worldwide replacement of just discretionary salt; (3) worldwide replacement of just non-discretionary salt; (4) worldwide replacement of discretionary salt just for people diagnosed with hypertension (whether treated or not); and (5) worldwide replacement of discretionary salt just for people treated for hypertension (**Supplementary Table 1**).

For every scenario we modelled the potential benefits mediated through SBP lowering. For the first three scenarios we also modelled the potential harms mediated through hyperkalaemia among people with CKD stages G3-5. For scenarios (4) and (5), we modelled only the potential benefits on the assumption that clinical decision-making would prevent use of potassium-enriched salt amongst people with CKD stages G3-5. For these model estimates, we assumed arbitrarily that 20% of people under each scenario would not be recommended to use potassium-enriched salt due to concerns about hyperkalaemia. The prevalence of CKD stages G3-5 among US adults with hypertension was 8.9% in 2015 to 2018^14^ so the assumed 20% untreated is deemed conservative.

### Data sources and key model assumptions

**Supplementary Table 2** lists key model inputs. For pre-intervention mean sodium intake, we used country-specific estimates from the 2021 GBD study. Country-specific estimates of dietary sources of sodium (discretionary versus non-discretionary) were derived from a linear regression based on Gross Domestic Product (GDP)^15^ updated by substituting the 2010 World Bank GDP figures used in the original report with 2021 data from the World Bank or the International Monetary Fund.

For BP inputs to the model, we used country-, age- and sex-specific estimates and standard deviations of SBP from the 2021 GBD study. Corresponding hypertension prevalence estimates and the proportions treated were extracted from the 2019 Non-Communicable Disease Risk Factor Collaboration.^16^ To estimate the effect of sodium reduction on SBP we used linear dose-response relationships from a meta-regression of trials^17^ that enabled us to calculate the change in SBP achieved for a given sodium reduction, in a given age group, separately for individuals with and without hypertension. Dose-response data from salt substitute trials were not used because there were few studies and different compositions of potassium-enriched salt studied. In line with the WHO recommendation for maximum daily intake of 2g/day sodium, no additional effect on SBP was modelled for reductions below this level. To estimate the effect of increased potassium intake on SBP, we used data from a meta-analysis of randomized controlled trials.^18^ Dose response effects for potassium intake on SBP varied by baseline sodium intake level with stronger relationships and greater maximum achievable SBP falls in those with higher baseline sodium consumption (**Supplementary text 1**).^18^ The effects of sodium reduction and increased potassium intake on SBP derived from these two estimation processes were assumed to be additive.

We used 2021 GBD Study country-, age- and sex-specific estimates for current CVD and CKD burden.^19^ The associations of SBP changes with the risks of each type of CVD and CKD were based on the 2019 GBD Study.^20^ The theoretical minimum risk exposure level for SBP was 110-115 mmHg for each outcome, and no additional protective effect on CVD or CKD risk was modelled for SBP falling below this level.

For modelling potential harms of hyperkalaemia-related mortality, we used data on the prevalence of each stage of CKD (G3-5) for country-, age- and sex-specific population groups obtained from the 2021 GBD Study group, supplemented with data from a further study that enabled separate estimates to be made for stage G3a (eGFR 45 to 59) and stage G3b (eGFR 30 to 44) CKD.^21^ The effects of increasing dietary potassium intake on serum potassium levels in people with stage G3a kidney disease were derived from a randomised trial^22^ and extrapolated to people with stages G3b, G4 and G5 using data from a cohort study.^23^ A meta-analysis of cohort studies was used to translate the effects on serum potassium levels into risks of cardiovascular death.^24^

### Statistical analysis

Following the methods used in our prior work^11^ (**Supplementary text 2**), we estimated the potential number of averted deaths, new cases and DALYs attributable to CVD and CKD resulting from a shift in SBP distribution due to changes in dietary sodium and potassium intake. For scenarios 1-3 we then also estimated the additional deaths from CVD caused by increased intake of potassium in individuals with CKD stages G3-5 and the net averted deaths were calculated for each simulation as the difference between the deaths averted by SBP lowering and the deaths caused by hyperkalaemia. We also estimated prevented deaths from CVD in individuals with CKD stages G3-5 alone. We summed up CKD stage-specific effects to derive the total additional deaths for all people with CKD stage G3-5 attributable to hyperkalaemia in each country, age and sex specific group. For new cases and DALYs, we excluded inflammatory heart diseases (rheumatic heart disease, endocarditis and other cardiomyopathy) from the estimations as the risks of these conditions are largely independent of BP.

### Uncertainty analysis

We quantified uncertainty in all analyses with probabilistic sensitivity analyses using n=1,000 Monte Carlo simulations, jointly incorporating uncertainties in input parameters (see **Supplementary Text 3**). We derived the corresponding central estimate for each endpoint (including benefits, harms, and net effects) from the 50^th^ percentile and the 95% uncertainty interval (UI) from the 2.5^th^ and 97.5^th^ centiles of 1000 modelling runs. We also conducted 11 deterministic sensitivity analyses by varying key input data (**Supplementary table 3**). All analyses were performed using R version 4.2.3 and RStudio version 2025.09.1 Build 401.

## RESULTS

### 1 Effects of replacing both discretionary and non-discretionary salt

Switching from regular salt to potassium-enriched salt worldwide was estimated to avert 2.96 (95% UI, 2.81 to 3.12) million deaths, 10.17 (9.59 to 10.70) million new cases, and 69.4 (65.6 to 72.9) million DALYs from CVD or CKD each year (**Table 1**). These figures represent 14.6%, 13.1% and 16.5% of the annual global disease burden attributable to CVD and CKD. For every metric, the greatest disease burden averted was for ischemic heart disease, hypertensive heart disease, stroke, and CKD (**Supplementary Figure 1**). The greatest absolute benefits were in China and India which have large populations and high levels of salt consumption (**Figure 2, Supplementary Figures 2-3**), but there were 133/193 (69%) countries, predominantly in Asia, North America, and Latin America where at least 10% of cardiovascular deaths would be prevented by switching to potassium-enriched salt (**Figure 3, Supplementary Table 4**). Overall, about 40% of the benefits are achieved in women, and this finding was broadly consistent across intervention strategies (**Supplementary Table 4**).

**Table 1.** Numbers of deaths, non-fatal events, and disability-adjusted life years from cardiovascular disease and kidney disease prevented each year, worldwide, under five different implementation scenarios for the use of potassium-enriched salt.

|  |  | Deaths averted in<br>millions<sup>#</sup><br>(95% UI) | Non-fatal events<br>averted in millions<br>(95% UI) | Disability Adjusted Life<br>Years prevented in<br>millions<br>(95% UI) |
| --- | --- | --- | --- | --- |
| *Adult population aged 25 years and above (n=4.63 billion; 95% UI: 4.60 to 4.67)* |  |  |  |  |
| **1. Replacement of discretionary salt and non-discretionary salt** | CVD and CKD | 2.96 (2.81-3.12) | 10.17 (9.59-10.70) | 69.43 (65.61-72.92) |
|  | • CVD* | 2.79 (2.65-2.95) | 8.00 (7.49-8.46) | 64.86 (61.20-68.22) |
|  | • CKD | 0.17 (0.17-0.18) | 2.18 (2.06-2.30) | 4.59 (4.38-4.78) |
| **2. Replacement of discretionary salt** | CVD and CKD | 1.85 (1.74-1.97) | 6.19 (5.78-6.59) | 44.28 (41.33-46.88) |
|  | • CVD* | 1.75 (1.64-1.87) | 4.96 (4.61-5.31) | 41.50 (38.67-43.97) |
|  | • CKD | 0.10 (0.10-0.11) | 1.23 (1.15-1.31) | 2.78 (2.64-2.94) |
| **3. Replacement of non-discretionary salt** | CVD and CKD | 1.56 (1.46-1.67) | 5.46 (5.11-5.83) | 36.12 (33.67-38.35) |
|  | • CVD* | 1.46 (1.37-1.57) | 4.23 (3.92-4.52) | 33.72 (31.35-35.84) |
|  | • CKD | 0.09 (0.09-0.10) | 1.23 (1.15-1.32) | 2.41 (2.28-2.53) |
| *^80% of adults with diagnosed hypertension aged 25 years and above (n=659 million; 95% UI: 645 to 673)* |  |  |  |  |
| **4. Replacement of discretionary salt among those with diagnosed hypertension** | CVD and CKD | 0.59 (0.55-0.63) | 1.72 (1.61-1.85) | 11.65 (10.87-12.48) |
|  | • CVD* | 0.55 (0.52-0.60) | 1.34 (1.24-1.45) | 10.91 (10.16-11.70) |
|  | • CKD | 0.03 (0.03-0.03) | 0.38 (0.36-0.41) | 0.73 (0.70-0.77) |
| *^80% of adults with treated hypertension aged 25 years and above (n=524 million; 95% UI: 512 to 536)* |  |  |  |  |
| **5. Replacement of discretionary salt among those with treated hypertension** | CVD and CKD | 0.48 (0.45-0.52) | 1.41 (1.31-1.52) | 9.34 (8.65-9.98) |
|  | • CVD* | 0.45 (0.42-0.49) | 1.09 (1.00-1.18) | 8.73 (8.07-9.35) |
|  | • CKD | 0.03 (0.03-0.03) | 0.32 (0.30-0.35) | 0.60 (0.57-0.64) |

\*For CVD deaths included are for rheumatic heart disease, ischemic heart disease, stroke (including ischemic stroke, intracerebral haemorrhage, and subarachnoid haemorrhage), hypertensive heart disease, non-rheumatic calcific aortic valve disease, other cardiomyopathy, atrial fibrillation and flutter, aortic aneurysm, peripheral artery disease, and endocarditis. For non-fatal CVD events, the outcomes included are ischemic heart disease, stroke (including ischemic stroke, intracerebral haemorrhage, and subarachnoid haemorrhage) non-rheumatic calcific aortic valve disease, atrial fibrillation and flutter and peripheral artery disease. For DALYs attributable to CVD outcomes included are ischemic heart disease, stroke (including ischemic stroke, intracerebral haemorrhage, and subarachnoid haemorrhage), hypertensive heart disease, non-rheumatic calcific aortic valve disease, atrial fibrillation and flutter, aortic aneurysm and peripheral artery disease

**Figure 2.**
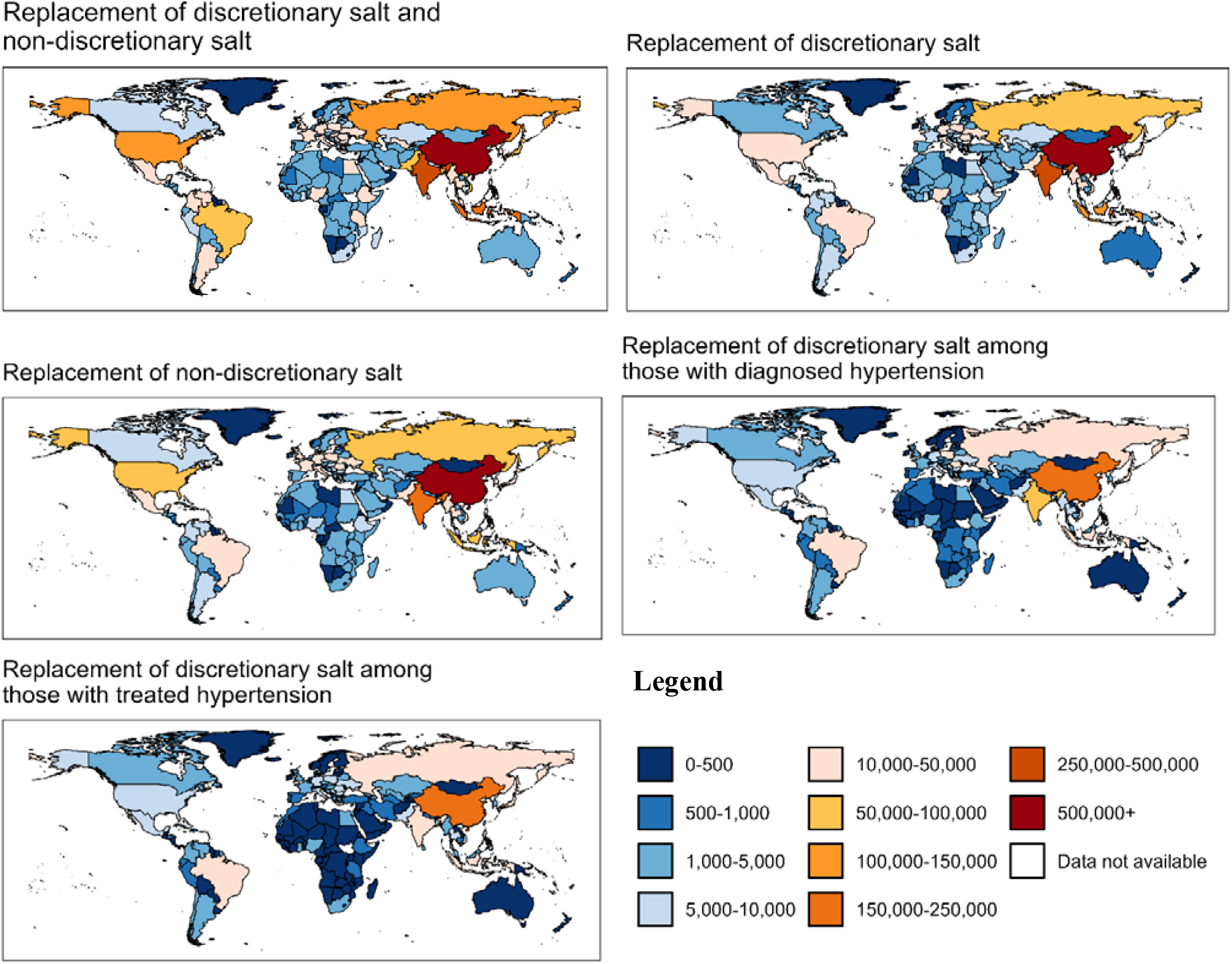
Numbers of deaths from cardiovascular disease and kidney disease prevented per year under different implementation scenarios for potassium-enriched salt for 193 countries or territories.

**Figure 3.**
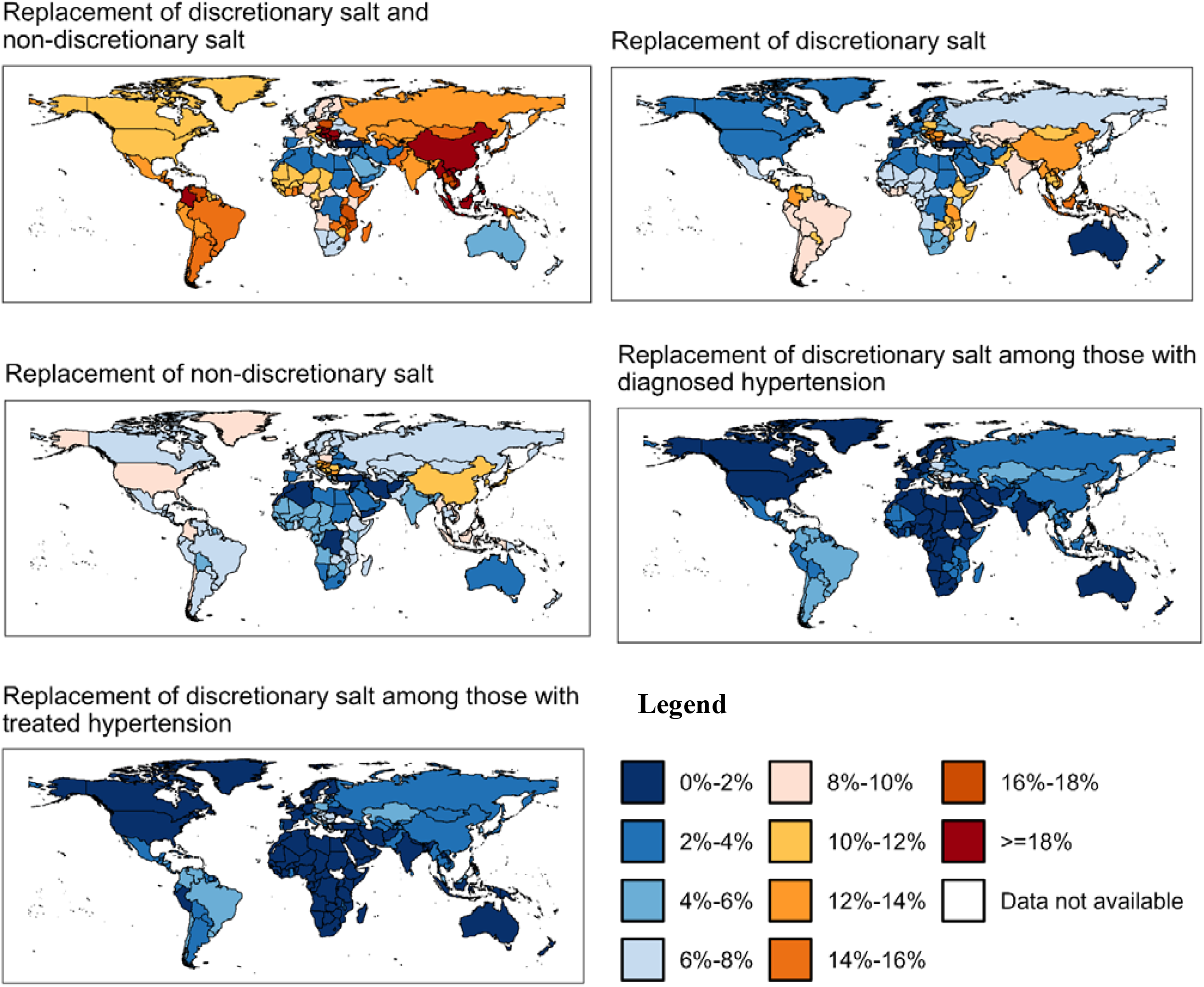
Proportion of national deaths from cardiovascular disease and kidney disease prevented per year under different implementation scenarios for potassium-enriched salt, for 193 countries or territories.

### 2 Effects of replacing discretionary salt alone

Switching just discretionary salt use to potassium-enriched salt was estimated to avert 1.85 (95% UI 1.74 to 1.97) million deaths, equating to 63% (1.85/2.96) (**Supplementary text 1**) of the benefit achieved with switching both discretionary and non-discretionary salt, and 9.2% of annual deaths attributable to CVD and CKD (**Table 1**). There were correspondingly lower proportions of new cases and DALYs averted with this approach, but the pattern of prevention across the main causes of CVD and CKD was similar to that observed for switching discretionary and non-discretionary salt together (**Supplementary Figure 1**). The geographic distributions of the absolute and proportional benefits were also similar (**Figures and 3, Supplementary Figures 2-5**). Switching discretionary salt use for China, India, Indonesia and Russia would achieve over 60% of the total global benefits (**Supplementary Figures 6-8**), but only 58/193 (30.1%) countries would prevent 10 % or more of CVD deaths each year with a strategy targeting discretionary salt alone (**Figure 3, Supplementary Table 4**).

### 3 Effects of replacing non-discretionary salt alone

Switching non-discretionary salt use alone would prevent 1.56 (95% UI 1.46 to 1.67) million deaths, which is 53% (1.56/2.96) (**Supplementary text 1**) of the benefit achieved with switching both discretionary and non-discretionary salt. Prevention of new cases and DALYs would be similarly lower (**Table 1**) and the pattern of prevention across CVD and CKD subtypes would be broadly similar to other strategies (**Supplementary Figure 1**). The projected health gain equates to 7.7% of annual deaths from CVD and CKD worldwide. The United States displaces Russia as one of the four countries projected to achieve the greatest benefit from this strategy which reflects the predominance of non-discretionary salt as the dietary source of sodium in the United States (**Supplementary Table 4, Supplementary Figures 6-8)**. There were only 20/193 (10%) countries achieving a 10% or greater reduction in cardiovascular deaths with an intervention strategy focused on non-discretionary salt alone (**Figure 3, Supplementary Table 4**).

### 4-5 Effects of replacing discretionary salt among people with diagnosed hypertension, treated hypertension or CKD

Replacing discretionary salt with potassium-enriched salt for just the 658 million people with diagnosed hypertension would prevent 0.59 (0.55 to 0.63) million deaths, and among the 523 million people with treated hypertension would prevent 0.48 (0.45 to 0.52) million deaths (**Table 1**). Among the 239 million adults worldwide with CKD stage G3 to G5, there would be 0.75 (95% UI 0.71 to 0.80) million CVD deaths prevented by switching discretionary and non-discretionary salt to potassium-enriched salt. This benefit would, however, be associated with the potential for causing 0.10 (0.09 to 0.11) million deaths from hyperkalaemia (**Table 2**).

**Table 2.** Numbers of cardiovascular deaths prevented by blood pressure lowering and numbers of cardiovascular deaths caused by hyperkalaemia each year, worldwide, in patients with chronic kidney disease stages 3-5, under three different population-wide implementation scenarios for the use of potassium-enriched salt

|  | Number of people with<br>stage 3-5 kidney disease<br>receiving potassium-enriched salt | Deaths prevented | Deaths caused by<br>hyperkalaemia |
| --- | --- | --- | --- |
|  | Millions (95% UI) | Millions (95% UI) | Millions (95% UI) |
| **1. Replacement of discretionary salt and non-discretionary salt** | 239 (236-242) | 0.75 (0.71-0.80) | 0.10 (0.09-0.11) |
| **2. Replacement of discretionary salt** | 239 (236-242) | 0.44 (0.41-0.47) | 0.05 (0.04-0.05) |
| **3. Replacement of non-discretionary salt** | 239 (236-242) | 0.40 (0.38-0.44) | 0.04 (0.04-0.05) |

UI=uncertainty interval

### Sensitivity analyses

In the deterministic sensitivity analyses, the magnitude of the estimated benefit varied across assumptions, but there were still large net benefits under each. Half replacement of all regular salt in all adults could avert 1.72 million deaths and one fifth replacement could avert 0.7 million deaths. We did five further sensitivity analyses on the effects of full replacement of all regular salt in all adults: (1) attenuated dose-response between sodium reduction and SBP; (2) additive effects of sodium reduction and potassium increase on blood pressure reduced from 100 per cent to 80 per cent; (3) effect of potassium supplementation on blood pressure reduced by half; (4) no effect of potassium supplementation in non-hypertensive individuals; and (5) for a salt substitute with a composition of 15% potassium chloride and 85% sodium chloride. The estimated number of averted deaths ranged from 2.02 to 2.52 million for these analyses (**Supplementary Table 6**). Using an alternative method to estimate the proportion of discretionary salt resulted in a greater benefit from replacing discretionary salt and a smaller benefit from replacing non-discretionary salt but had little impact on the overall estimated benefit. Finally, assuming a higher prevalence of CKD stages 3-5 in low and middle-income countries among individuals with diagnosed or treated hypertension led to slightly lower health gains, but the benefits remained substantial among these population groups (**Supplementary Table 6**).

## DISCUSSION

The global health impact of replacing regular salt with potassium-enriched salt projected by these analyses is large – more than 13 million deaths or new cases of CVD or CKD, and more than 69 million DALYs could be prevented each year if a global switch of regular salt to potassium-enriched salt could be implemented. The projected benefits are much greater than those anticipated from sodium reduction strategies that focus solely on cutting salt intake because the potassium supplementation included in this intervention provides additional health gains not achieved by sodium reduction alone.^19^ While replacement of salt from both discretionary and non-discretionary sources prevents the largest disease burden, population-wide interventions targeted at either alone would also deliver large health gains. Intervention strategies that switch just discretionary salt in people with diagnosed hypertension achieve only about one-fifth of the potential benefit but may be more immediately achievable because they target populations already under clinical care and should minimise hyperkalaemia risk.

### Explanation of findings

CVD is a frequent complication of CKD and hypertension is highly prevalent among this population group.^25^ Our analysis projected that more than seven CVD deaths could be prevented for every one death that might be caused in people with CKD. Importantly, more than one quarter (0.75/2.79 million) of CVD deaths that might be prevented by a global switch to potassium-enriched salt are projected to be averted in people with CKD. There is a strong rationale for expecting increased rates of hyperkalaemia in people with advanced CKD and the potential harm from hyperkalaemia from a public intervention raises an ethical concern, though the assumed harms have never been demonstrated in trials. The estimates we made for population-wide intervention assumed that there was no capacity to prevent people with advanced CKD from using potassium-enriched salt, and the magnitude of the projected harm is likely to have been substantially overestimated as a consequence. In practice, those with advanced CKD are more likely to be symptomatic and diagnosed and routinely advised to limit salt intake and avoid products high in potassium. Strategies to ameliorate risks of hyperkalaemia include better screening for CKD, standardised warning labels on potassium-enriched salt products, listing of potassium-enriched salt when used as an ingredient in foods, and reporting of potassium content in the nutrient information panels on packaged foods. If hyperkalaemia risks are considered particularly high for a population, then an implementation strategy that focuses on switching discretionary salt use in people with hypertension should minimise risk, because clinician screening should prevent at-risk people with CKD from being recommended potassium-enriched salt.

The largest anticipated absolute benefits are in countries like China, India, and Indonesia, which have large populations and a high burden of CVD and CKD. Targeting discretionary salt has greater overall benefits than targeting non-discretionary salt because a greater proportion of salt consumed worldwide derives from discretionary sources at present.^15^ The recently launched WHO guideline on using lower-sodium salt substitutes with added potassium focuses on switching potassium-enriched salt for regular table salt as part of efforts to reduce sodium intake.^26^ The importance of targeting non-discretionary salt will grow because economic development is strongly correlated with diets based on processed and restaurant foods.^15^ For regions like Europe and the United States over 70% of salt intake comes from these sources. The already greater impact of targeting non-discretionary sources of salt in higher income countries identified in these analyses presages a worldwide shift towards a greater overall impact of interventions targeting non-discretionary sources of salt over the coming years. There are some persisting uncertainties about the technical performance of potassium-enriched salt in processed and restaurant foods, but the UK Committee on Toxicity considered that replacing up to 25-30% with potassium chloride in most food categories is feasible and safe.^27^

Full implementation of a switch to potassium-enriched salt will likely take decades but can learn from the prior switch of the worlds salt supply from regular salt to iodised salt. The key practical challenges to achieving the health gains modelled here will be raising awareness amongst clinicians and the general community, increasing availability and supply in retail outlets, and reducing the price to that of regular salt. Ensuring the food industry switches to using potassium-enriched salt while actively engaging with governments, professional societies, and guideline bodies to drive change will also be important. The price of potassium-enriched salt varies considerably but on average is double the cost of regular salt.^28^ This increased cost relates to both potassium chloride being more expensive than sodium chloride and to potassium-enriched salt being marketed as a niche health product. This may have a higher impact on uptake in low and middle-income countries, though sodium chloride is a low-cost product and even at twice the price, potassium-enriched salt is a low-cost commodity. Economies of scale in the production of potassium chloride should bring down prices and increase availability. Reducing price will be key to achieving industry uptake and to supporting public health efforts such as mandated government interventions. Raising awareness can be achieved by updating clinical guidelines for the management of hypertension, as done recently in Europe^29^ and the United States^30^ alongside education campaigns targeting end users in the food industry and government, as well as consumers.

### Strengths and limitations

Our study is the first to project the effects of switching regular salt use for potassium-enriched salt in 193 separate jurisdictions and worldwide. A key strength of the analysis is the use of a consistent method to evaluate the potential health impacts of a range of different implementation scenarios. The modelling pathway was adapted from previous studies but upgraded to include additional detail about dietary intake levels. This enabled better understanding of how the benefits and potential risks might trade off against each other for each country and for specific population groups. The analyses also enable us to account for uncertainty across the model pathway and generate more robust estimates of uncertainty.

The currency of the analyses is limited by being based on data from a few years ago and may not fully reflect the present status of each country. There is also a paucity of data describing the proportion of salt intake from discretionary versus non-discretionary sources and the comparisons between the effects of strategies targeting each source of sodium are therefore somewhat uncertain. Additional country-specific data could provide more precise estimates for each nation as well as identify the most feasible strategies for implementation. The headline number of 2.96 million deaths averted were projected based on multiple assumptions about the relationships between sodium, potassium and BP, which probably fail to capture the full complexity of the effects in different populations. For example, the dose-response curves for the effects of sodium and potassium on blood pressure may change with background use of antihypertensive medications. The evidence for the dose-response relationship between potassium supplementation and blood pressure is also relatively sparse compared to that for sodium, particularly among individuals without hypertension and projected health benefits were lower when the modelled effects of potassium were reduced. The data available to estimate the effects of potassium supplementation on hyperkalaemia risk were limited, though we used assumptions that were more likely to over-estimate than under-estimate potential harms. The model makes an estimate for just one point in time and does not account for future changes in population demographics and exposures. Finally, all the modelled scenarios would take many years and much effort to implement and do not represent immediately achievable health gains.

## Conclusion

These analyses complement the recently released World Health Organization guideline recommendations on the use of low sodium salt substitutes with added potassium. The findings identify countries where switching to potassium-enriched salt might be prioritised and provide a quantitative indication of the likely balance of risks and benefits for each. A translational research agenda that shows how implementation can be achieved in diverse settings is now required.

## Data Availability

All data used in this paper are publicly available or can be requested from GBD. References to data sources are presented in Supplementary Table 1. Publicly available data can be shared upon request. For data requested from GBD, potential users can request from GBD or it can be shared with consent from GBD.

https://vizhub.healthdata.org/gbd-results/

## Contributors statement

LH, BN and MM conceptualised the study. LH conducted the formal analysis with supervision from MM. XX led the work while LH took maternity leave for 6 months. BN and MM provided oversight of the work and helped draft the early iterations of the report. All other authors reviewed the manuscript and provided critical feedback.

## Acknowledgements

We acknowledge the GBD team for providing the input data for this work. This work did not receive specific funding, but fellowship support received by individual authors enabled their contributions to the work. Specifically, Dr. Huang, Dr. Schutte, Dr. Aminde and Dr. Neal are supported by investigator grants from the National Health and Medical Research Council of Australia; Dr. Hoorn is supported by the Dutch Kidney Foundation; Dr Trieu is supported by the National Heart Foundation of Australia; Dr. Marklund, Dr. Matsushita, Dr. Brady and Dr. Appel are supported by Resolve to Save Lives.

## References

1. Foreman KJ, Marquez N, Dolgert A, et al. Forecasting life expectancy, years of life lost, and all-cause and cause-specific mortality for 250 causes of death: reference and alternative scenarios for 2016-40 for 195 countries and territories. Lancet 2018; 392(10159): 2052–90.

2. Vaduganathan M, Mensah GA, Turco JV, Fuster V, Roth GA. The Global Burden of Cardiovascular Diseases and Risk: A Compass for Future Health. J Am Coll Cardiol 2022; 80(25): 2361–71.

3. WHO Global Report on sodium intake reduction. Geneva World Health Organization, 2023.

4. Institute for Health Metrics and Evaluation (IHME) diet high in sodium. Global Burden of Disease; 2019.

5. Reddin C, Ferguson J, Murphy R, et al. Global mean potassium intake: a systematic review and Bayesian meta-analysis. European Journal of Nutrition 2023; 62(5): 2027–37.

6. Guideline: Potassium intake for adults and children. Geneva: World Health Organization; 2012.

7. Kissock KR, Ghammachi N, Hoek AC, et al. Knowledge, attitudes, and behaviours related to reduced-sodium salt: a systematic review. J Hum Hypertens 2026; 40(1): 1–9.

8. Neal B, Wu Y, Feng X, et al. Effect of Salt Substitution on Cardiovascular Events and Death. N Engl J Med 2021; 385(12): 1067–77.

9. Yin X, Rodgers A, Perkovic A, et al. Effects of salt substitutes on clinical outcomes: a systematic review and meta-analysis. Heart 2022; 108(20): 1608–15.

10. Yuan Y, Jin A, Neal B, et al. Salt substitution and salt-supply restriction for lowering blood pressure in elderly care facilities: a cluster-randomized trial. Nat Med 2023; 29(4): 973–81.

11. Marklund M, Singh G, Greer R, et al. Estimated population wide benefits and risks in China of lowering sodium through potassium enriched salt substitution: modelling study. BMJ 2020; 369: m824.

12. Marklund M, Tullu F, Raj Thout S, et al. Estimated Benefits and Risks of Using a Reduced-Sodium, Potassium-Enriched Salt Substitute in India: A Modeling Study. Hypertension 2022; 79(10): 2188–98.

13. Aminde LN, Nugraheni WP, Mubasyiroh R, et al. Cost-effectiveness analysis of low-sodium potassium-rich salt substitutes in Indonesia: an equity modelling study. Lancet Reg Health Southeast Asia 2024; 26: 100432.

14. Zeng X, Zeng Q, Zhou L, Zhu H, Luo J. Prevalence of Chronic Kidney Disease Among US Adults With Hypertension, 1999 to 2018. Hypertension 2023; 80(10): 2149–58.

15. Bhat S, Marklund M, Henry ME, et al. A Systematic Review of the Sources of Dietary Salt Around the World. Adv Nutr 2020; 11(3): 677–86.

16. Zhou B, Carrillo-Larco RM, Danaei G, et al. Worldwide trends in hypertension prevalence and progress in treatment and control from 1990 to 2019: a pooled analysis of 1201 population-representative studies with 104 million participants. The Lancet 2021; 398(10304): 957–80.

17. Mozaffarian D, Fahimi S, Singh GM, et al. Global sodium consumption and death from cardiovascular causes. New England Journal of Medicine 2014; 371(7): 624–34.

18. Filippini T, Naska A, Kasdagli MI, et al. Potassium intake and blood pressure: a dose-response meta-analysis of randomized controlled trials. Journal of the American Heart Association 2020; 9(12): e015719.

19. Institute for Health Metrics and Evaluation. GBD Results. 2021. https://vizhub.healthdata.org/gbd-results/ (accessed August 13 2024).

20. Global Burden of Disease Collaborative Network. Global Burden of Disease Study 2019 (GBD 2019) Relative Risks. Seattle, United States of America: Institute for Health Metrics and Evaluation (IHME) 2020.

21. Matsushita K, Mahmoodi BK, Woodward M, et al. Comparison of risk prediction using the CKD-EPI equation and the MDRD study equation for estimated glomerular filtration rate. Jama 2012; 307(18): 1941–51.

22. Turban S, Juraschek SP, Miller III ER, et al. Randomized trial on the effects of dietary potassium on blood pressure and serum potassium levels in adults with chronic kidney disease. Nutrients 2021; 13(8): 2678.

23. Ogata S, Akashi Y, Kato S, et al. Association Between Dietary Potassium Intake Estimated From Multiple 24-Hour Urine Collections and Serum Potassium in Patients With CKD. Kidney Int Rep 2023; 8(3): 584–95.

24. Kovesdy CP, Matsushita K, Sang Y, et al. Serum potassium and adverse outcomes across the range of kidney function: a CKD Prognosis Consortium meta-analysis. Eur Heart J 2018; 39(17): 1535–42.

25. Burnier M, Damianaki A. Hypertension as Cardiovascular Risk Factor in Chronic Kidney Disease. Circ Res 2023; 132(8): 1050–63.

26. Use of lower-sodium salt substitutes: WHO guideline. Geneva: World Health Organization; 2025.

27. Committe on Toxicity: Committee on Toxicity of Chemicals in Food CPatE. Statement on potassium-based replacements for sodium chloride and sodium-based additives.

28. Yin X, Liu H, Webster J, et al. Availability, Formulation, Labeling, and Price of Low-sodium Salt Worldwide: Environmental Scan. JMIR Public Health Surveill 2021; 7(7): e27423.

29. McEvoy JW, McCarthy CP, Bruno RM, et al. 2024 ESC Guidelines for the management of elevated blood pressure and hypertension. Eur Heart J 2024; 45(38): 3912–4018.

30. Gulati M, Moore MM, Cibotti-Sun M. 2025 High Blood Pressure Guideline-at-a-Glance. J Am Coll Cardiol 2025; 86(18): 1560–6.

