## Supplementary material for "The effects on global health outcomes of switching from regular salt to potassium-enriched salt: a modelling study"

### **Supplementary text 1. Additional information on evidence about the effect of potassium on blood pressure and cardiovascular disease, model assumptions, input data, and interpretation of results**

***The effect of potassium on blood pressure and cardiovascular risk***

The effect of excessive sodium on blood pressure (BP) and cardiovascular (CV) risk is well established and likewise the potential benefits of sodium reduction are widely accepted. By contrast, the role of potassium has received less attention. Nonetheless, the World Health Organisation recommends a minimum daily intake of 3.5g/day of potassium to support cardiovascular health,^1^ and this recommendation is based on data showing associations of potassium intake with blood pressure and cardiovascular risk, as well as intervention studies that show blood pressure-lowering effects of potassium supplementation.

**Physiology.** Diets throughout hominid evolution were potassium-rich and sodium-poor, with significant temporal fluctuation in consumption levels.^2^ Homeostatic mechanisms evolved to maintain plasma sodium and potassium levels within tightly controlled ranges. Potassium is the principal intracellular cation and sodium is the principal extracellular cation. Electrical potential across cell membranes is maintained by the actions of the sodium–potassium pump. Increased dietary potassium intake raises plasma potassium which causes the sodium–potassium pump to move potassium into cells and sodium out of cells to preserve the resting membrane potential and stabilise electrical activity.^3,4^ In parallel, when dietary potassium intake rises there is an increase in renal potassium excretion associated with sodium loss (natriuresis),^5^ water loss (diuresis), plasma volume contraction and lower blood pressure. Potassium also appears to contribute to blood pressure regulation by direct relaxing effects on vascular smooth muscle which reduces vascular resistance.^6^ Compared to evolutionary consumption levels, sodium intake is now very high and potassium intake very low. Potassium consumption levels are about one quarter that consumed on a palaeolithic diet and sodium consumption levels at least 4 times higher.^7^

**Potassium and blood pressure in cross-sectional observational studies.** Dietary potassium consumption levels and urinary indicators of dietary potassium consumption are consistently associated with blood pressure in adequately powered, well-conducted studies. Associations of potassium intake and blood pressure are observed among both hypertensive and non-hypertensive individuals. For example:

- In the large, multi-country INTERSALT study,^8^ potassium was negatively associated with blood pressure level after adjusting for confounders.
- In the Prospective Urban Rural Epidemiology (PURE) study, higher potassium excretion was associated with lower systolic blood pressure, with stronger associations at higher sodium intakes and in older or hypertensive individuals.^9^
- Multiple population-based studies found an inverse association of dietary potassium intake and systolic blood pressure in both people with and without hypertension.^10,11^
- A meta-analysis of 12 studies found that urinary potassium excretion is higher in people without hypertension than in those with hypertension, supporting a population-level association between higher habitual potassium intake and lower BP.^12^

**Potassium and cardiovascular risk in prospective cohort studies.** Meta‑analyses of prospective observational studies link higher potassium intake to lower stroke risk and suggest reductions in a broad range of cardiovascular endpoints^13-15^ - analysis of 11 prospective studies showed that higher potassium intake was associated with a significantly lower risk of stroke and with non-significant trends towards lower risks of coronary heart disease and total cardiovascular disease.^14^ In dose‑response analyses of 10 prospective cohorts,^15^ each 1,000 mg/day higher potassium intake was associated with an 11% lower stroke risk, with the lowest risk at about 3,500 mg/day (≈90 mmol/day). Another analysis of 14 cohort studies found a similar dose-response relationship between potassium intake and risk of stroke.^16^

**Potassium and blood pressure in randomised controlled trials**. Multiple meta-analyses^14,17-19^ of trials show a blood pressure-lowering effect of increasing potassium intake, particularly among people with hypertension. Associations may be non-linear and vary between population subgroups with greater reductions among people with hypertension^20^ and uncertainty about effects in people without hypertension who are under-represented in the trials. For example, in 32 RCTs included in the dose-response analysis in 2020, only 5 studies were conducted among people without hypertension.^20^ In general, point estimates of effect suggest blood pressure lowering amongst individuals without hypertension but confidence intervals about the effect estimates are wide and span unity.

The effects of potassium supplementation on blood pressure appear to be modified by the baseline level of sodium intake, with greater reductions when sodium intake is high. Besides having an independent effect on lowering blood pressure, potassium may mitigate the effect of excessive sodium intake on blood pressure and lower cardiovascular risk.^21-23^ Furthermore, the sodium-to-potassium ratio is significantly correlated with both blood pressure level and cardiovascular risks in multiple studies,^24-30^ and in many, the relationship of outcomes with the sodium-to-potassium ratio is stronger than for either sodium or potassium alone.^28-30^ This provides a strong rationale for simultaneously reducing sodium intake and increasing potassium intake to ultimately reduce cardiovascular risk.

***Maximum effect of sodium reduction and potassium increase on blood pressure***

In the primary analysis, we applied dose-response data of sodium reduction on systolic blood pressure (SBP) from Mozaffarian et al. (2014).^31^ For the general population, each 100 mmol reduction in sodium intake was associated with a 3.735 mmHg (SD: 0.73) decrease in SBP. For each year difference in age from 50, there is an additional more/less effect of 0.105 (SD:0.029) mmHg per 100 mmol sodium reduction, but the effect does not go any less for age below 40. Among individuals with hypertension, an additional SBP reduction of 1.874 mmHg (SD: 0.884) per 100 mmol sodium reduction was applied. If pre-intervention sodium intake was below the theoretical minimum risk level (2 g/day), no effect from sodium reduction or potassium increase was estimated. If post-intervention sodium intake fell below 2 g/day, it was adjusted to 2g/day, and changes in sodium and potassium intake were recalculated accordingly.

For the effect of potassium increase on SBP, we used dose-response data from a meta-analysis conducted by Filippini et al.^20^ Specifically, for baseline sodium intake below 3g/day, each mmol potassium increase would reduce SBP by 0.07 (SD:0.05) mmHg and if the effect of potassium increase on SBP reduction exceeds 2.2 mmHg calculated from dose-response data, this was reset to 2.2 mmHg. For baseline sodium intake between 3-4g/day each mmol potassium increase would reduce SBP by 0.11 (SD:0.04) mmHg and if the effect of potassium increase on SBP reduction exceeds 3.3 mmHg calculated from dose-response data, this was reset to 3.3 mmHg. For baseline sodium intake above 4g/day each mmol potassium increase would reduce SBP by 0.11 (SD:0.05) mmHg and if the effect of potassium increase on SBP exceeds 6.5 mmHg calculated from dose-response data, this was reset to 6.5 mmHg. Although in the meta-analysis by Filippini, only 32 trials were included, it is the best available data to quantify the dose-response relationship between potassium supplementation and blood pressure. However, the effect of potassium supplementation on blood pressure lowering is well studied in animal studies and other large epidemiological studies.

Meta-analysis of salt substitute trials^32^ have shown larger blood pressure reduction compared to sodium reduction^33^ and potassium supplementation^20^ alone. However, we did not use data from salt substitute trials in the model because there were few studies and different compositions of potassium-enriched salt studied, and no direct dose-response data are available. The effects of sodium reduction and potassium increase were summed to estimate the total SBP reduction. If pre-intervention SBP was below the theoretical minimum risk level (110–115 mmHg), no reduction was applied. If post-intervention SBP fell below this threshold, it was reset to the minimum risk level.

***Diseases included in the model***

For every intervention scenario, we modelled the impact on CVD and CKD. The subsets of CVD included were rheumatic heart disease, ischemic heart disease, stroke (including ischemic stroke, intracerebral haemorrhage, and subarachnoid haemorrhage), hypertensive heart disease, non-rheumatic calcific aortic valve disease, other cardiomyopathy, atrial fibrillation and flutter, aortic aneurysm, peripheral artery disease, and endocarditis. The subset of CKD included diabetes mellitus type 1, diabetes mellitus type 2, hypertension, glomerulonephritis, and other unspecified causes. These diseases were included based on the data availability of the relative risk between SBP and the disease from GBD 2019. The disease burden measures estimated were deaths, new cases (not available for rheumatic heart disease, endocarditis, hypertensive heart disease, aortic aneurysm, and other cardiomyopathies) and disability-adjusted life years (excluding rheumatic heart disease, endocarditis and other cardiomyopathy). Rheumatic heart disease and endocarditis are include in death outcome because high SBP has been associated with an increasing risk of fatal heart failure which is considered an intermediate cause of death.

***Effect of dietary potassium increase on serum potassium***

To estimate the effect of increased dietary intake of potassium on serum potassium, we used data from a randomised controlled feeding trial that provided meals that are lower or higher in potassium for people with CKD stage 3.^34^ Each g/day increase in dietary potassium intake was estimated to increase serum potassium by 0.23 (0.08 to 0.38) mmol/L. In another cohort study that investigated the effect of dietary potassium intake on serum potassium,^35^ the effect of dietary potassium on serum potassium among people with CKD stage 4 is 1.5 times the effect among people with CKD stage 3, and among people with CKD stage 5, the effect is double the effect among people with CKD stage 3. In our model, we used the data from the randomised controlled feeding trial for people with CKD stages 3a and 3b and data from the cohort study to extrapolate the effect among people with CKD stages 4 and 5.

***Understanding the proportion of effect from replacing discretionary/non-discretionary salt alone over replacing both discretionary and non-discretionary salt***

Switching from regular salt to potassium-enriched salt of both discretionary and non-discretionary salt was estimated to prevent 2.96 million deaths (95% UI: 2.81–3.12 million). Switching just discretionary salt use to potassium-enriched salt was estimated to avert 1.85 (95% UI 1.74 to 1.97) million deaths, equating to 62.5% (1.85/2.96) of the benefit achieved with switching both discretionary and non-discretionary salt, and switching non-discretionary salt use alone would prevent 1.56 (95% UI 1.46 to 1.67) million deaths, which is 52.7% (1.56/2.96) of the benefit achieved with switching both discretionary and non-discretionary salt.

It should be noted that deaths prevented from switching just discretionary salt use to potassium-enriched salt and switching just discretionary salt use to potassium-enriched salt don’t sum up to the deaths preventable by switching both discretionary and non-discretionary salt. Instead, they add up to more than the deaths preventable by switching both discretionary and non-discretionary salt. The reasons are due to the model assumptions explained above. The maximum effect of intervention on sodium/potassium and SBP are more likely to be reached with the joint intervention than with either of the separate interventions. Accordingly, the separate effects of the two implemented separately add to more than the joint effect of the two combined.

### **Supplementary text 2. Statistical analyses**

***Potential benefits mediated through blood pressure lowering***

We calculated the potential impact fraction (PIF) for each CVD and CKD outcome resulting from a shift in SBP distribution due to dietary sodium and potassium intake changes. The subsets of CVD included were rheumatic heart disease, ischemic heart disease, stroke (including ischemic stroke, intracerebral haemorrhage, and subarachnoid haemorrhage), hypertensive heart disease, non-rheumatic calcific aortic valve disease, other cardiomyopathy, atrial fibrillation and flutter, aortic aneurysm, peripheral artery disease, and endocarditis. The main causes of CKD varied between countries but included diabetes mellitus type 1, diabetes mellitus type 2, hypertension, glomerulonephritis, and other unspecified causes. The disease burden measures estimated were deaths, new cases (not available from GBD for hypertensive heart disease, aortic aneurysm, and other cardiomyopathies) and disability-adjusted life years. The PIF for each outcome (o) was defined as:

$${PIF}_{oas}=\frac{{ʃ^{m}}_{x=0}{RR}_{oa}\left( X \right)P_{as}\left( X \right)dx-{ʃ^{m}}_{x=0}{RR}_{oa}\left( X \right){P^{'}}_{as}\left( X \right)dx}{{ʃ^{m}}_{x=0}{RR}_{oa}\left( X \right)P_{as}\left( X \right)dx}$$

$P_{as}\left( X \right)$ and ${P^{'}}_{as}\left( X \right)$ are the pre-intervention and post-intervention SBP distributions in each age (a) and sex (s) group in each country. ${RR}_{oa}\left( X \right)$ is the relative risk as a function of SBP (X), outcome(O), and age (a): ${RR}_{oa}\left( X \right)=e^{(\frac{lnRRoa\left( x-TMREL \right)}{10})}$. LnRRoa is the increase in the natural logarithm of the relative risk of outcome with age for each 10 mmHg increase in SBP. TMREL is the theoretical minimum risk exposure level (110-115 mmHg) of SBP.

The CVD and CKD prevented was estimated by multiplying a country-, age-, sex-, and cause-specific PIF by the estimated current number of events for the same group. In each group, we also estimated prevented deaths from CVD in individuals with CKD alone. For new cases and DALYs, we excluded inflammatory heart diseases (rheumatic heart disease, endocarditis and other cardiomyopathy) from the estimations as the risks of these conditions are largely independent of BP. The theoretical minimum risk exposure level of blood pressure was propagated by sampling between the 110-115 mm Hg interval each time the potential impact fraction was calculated.

***Potential harms mediated through hyperkalaemia***

To estimate the additional deaths from CVD caused by increased intake of potassium in individuals with CKD stages G3-5, we computed the PIF for deaths from CVD for each stage of CKD(e) as:

$${PIF}_{e}=\frac{{\sum^{m}}_{x=0}RR\left( X \right)P_{e}\left( X \right)-{\sum^{m}}_{x=0}RR\left( X \right){P^{'}}_{e}\left( X \right)}{{\sum^{m}}_{x=0}RR\left( X \right)P_{e}\left( X \right)}$$

$P_{e}\left( X \right)$ and ${P^{'}}_{e}\left( X \right)$ are the pre-intervention and post-intervention distributions of serum potassium in patients with CKD stage. $RR\left( X \right)$ is the relative risk of death from CVD at serum potassium concentration x, compared to a reference serum potassium of 4.2 mmol/L. We defined hyperkalaemia deaths as cardiovascular deaths caused by serum potassium levels above 5.5 mmol/L to align with the clinical diagnosis of hyperkalaemia. Thus, for hyperkalaemia mortality, this was assumed as 1.00 if serum potassium <5.5 mmol/L. The additional hyperkalaemia deaths from the intervention were computed by multiplying the negative of the PIF in each group by the number of deaths from CVD in the same group before the intervention.

### **Supplementary text 3. Probabilistic sensitivity analyses**

We quantified uncertainty in all analyses with probabilistic sensitivity analyses using n=1,000 Monte Carlo simulations, jointly incorporating uncertainties in input parameters. For each simulation, a draw was made from the distributions of: pre-intervention mean sodium intake; intercept and slope of regression between GDP per capita and contribution of discretionary salt to total sodium intake; pre-intervention mean and SD of SBP; theoretical minimum risk exposure level of SBP; prevalence of diagnosed hypertension and treated hypertension; main effect and age- and hypertension-interaction of sodium reduction on SBP; effect of dietary potassium increase on SBP; effects of SBP on CVD and CKD outcome; current number of events (e.g., deaths) for each CVD and CKD outcome; prevalence of stage-specific CKD; and the hazard ratio for deaths from CVD in patients people with CKD compared with others in the same age group; population size; pre-intervention mean serum potassium concentration by different stage of CKD; association between dietary potassium increase and serum potassium concentration; and relative risk of serum potassium with deaths from CVD.

### **Supplementary table 1. Details of five intervention scenarios to reduce the global intake of sodium by replacing regular salt with potassium-enriched salt**

|  | **Population** | **Salt replaced** | **Proportional replacement *** | **Risks and benefits modelled** |
| --- | --- | --- | --- | --- |
| **1. Replacement of discretionary salt and non-discretionary salt** | Global population aged 25 years and above (n=4.63 billion; 95% UI: 4.59 to 4.67) | Discretionary salt and salt from processed and restaurant foods | 100% | Benefits from blood pressure lowering and potential harms from raising potassium consumption in people with kidney disease. |
| **2. Replacement of discretionary salt** | Global population aged 25 years and above (n=4.63 billion; 95% UI: 4.59 to 4.67) | Discretionary salt | 100% |  |
| **3. Replacement of non-discretionary salt** | Global population aged 25 years and above (n=4.63 billion; 95% UI: 4.59 to 4.67) | Salt from processed and restaurant foods | 100% |  |
| **4. Replacement of discretionary salt among those with diagnosed hypertension** | 80% of people with diagnosed hypertension aged 25 years and above (n=658 million; 95% UI: 644 to 673) | Discretionary salt | 100% | Benefits from blood pressure lowering with no potential harms from raising potassium consumption in people with kidney disease.** |
| **5. Replacement of discretionary salt among those with treated hypertension** | 80% of patients with treated hypertension aged 25 years and above (n=523 million; 95% UI: 512 to 537) | Discretionary salt | 100% |  |

* Replacement of 100% sodium chloride with potassium-enriched salt with a composition of 25% potassium chloride and 75% sodium chloride.

** Assumes 20% of population not treated on basis of clinical advice about possible hyperkalaemia risk.

### **Supplementary table 2. Input data for estimating the impact of a global switch from regular salt to potassium-enriched salt on health outcomes**

| **Input data** | **Group** | **Year** | **Value and distribution** | **Note** | **Source** |
| --- | --- | --- | --- | --- | --- |
| **Blood pressure model** | | | | | |
| Pre-intervention sodium intake | Country, sex, age | 2021 | Normal distribution |  | GBD Study 2021^36^ |
| Sources of dietary sodium intake (discretionary vs. other) | Country |  | Normal distribution | A random draw was done for both the intercept and coefficient of the regression model to estimate the proportion of discretionary salt | Bhat et al^37^ |
| GDP data | Country | 2021 | NA | Used for estimating percentage of sodium intake from discretionary vs. other. 2021 GDP was deflated to 2010 GDP using an inflation rate of 1.24 to be consistent with the original regression model.  For Eritrea and Cuba, 2021 GDP data is not available, 2019 data from IMF is used for Eritrea and 2020 data from World Bank is used for Cuba | World Bank^38^  International Monetary Fund^39^ |
| Pre-intervention SBP | Country, sex, age | 2021 | Gamma distribution | Requested from GBD working group | GBD Study 2021 |
| Prevalence of hypertension and prevalence of treated hypertension | Country, sex, age | 2019 | Normal distribution | Data was available for age 30 to 79 in five years group. For age 25 to 29, same prevalence was assumed as age 30 to 34, and for age above 80, same prevalence was assumed as age 75 to 79 | NCD risk factor collaboration 2019^40^ |
| Dose-response relationship between sodium reduction and SBP change, accounting for modifying effects of age and hypertension | NA | NA | Normal distribution | Data extracted from S1 Text | Mozaffarian et al^31^  Filippini et al^41^ |
| Dose-response relationship between potassium increase and SBP change | NA | NA | Normal distribution | A ceiling effect is applied based on baseline sodium intake | Filippini et al^20^ |
| SBP effect on CVD and CKD risk | Age | 2019 | Log-normal distribution |  | GBD Study 2019^42^ |
| Current CVD and CKD burden | Country, sex, age | 2021 | Normal distribution |  | GBD Study 2021^43^ |
| Prevalence of total CKD stages G3a–G5, % | Country, sex, age | 2021 | Normal distribution | Requested from GBD working group | GBD Study 2021 |
| Age-specific HR of CVD mortality in  CKD compared with non-CKD | Age | NA | Log-normal distribution |  | Hallan et al^44^ |
| Serum potassium model | | | | | |
| Stage-specific CKD prevalence (%) | CKD stage | 2021 | Normal distribution | Requested from GBD working group | GBD Study 2021 |
| Population estimate | Country, sex, age | 2021 | Normal distribution |  | GBD Study 2021^45^ |
| Stage-specific HR of CVD mortality  compared with non-CKD | CKD stage | NA | Log-normal distribution |  | Matsushita et al^46^ |
| Mean (SD) current serum potassium distribution (mmo/L) | CKD stage | NA | Normal distribution |  | Kovesdy et al^47^ |
| Dose-response relationship between dietary potassium effect on serum  potassium level (mmol/L per g/day) | CKD stage≥G3a | NA | Normal distribution | For stage 3a and 3b, data from Turban et al was used. For stages 4 & 5, the relative relationship with stage 3 from Ogata et al was used, i.e. the effect for stage 4 was 1.5 times the effect of stage 3 and for stage 5, double that of stage 3. | Turban et al^34^  Ogata et al^35^ |
| Serum potassium effect on CVD mortality | NA | NA | HR (95% CI) compared to serum K 4.2 mmol/L available for each 0.05 increment of serum K in the interval of 4.2 to 6.5 mmol/L |  | Kovesdy et al^47^ |

### **Supplementary table 3. Sensitivity analyses of intervention scenarios to reduce the global intake of sodium by replacing regular salt with potassium-enriched salt**

|  | **Population** | **Salt replaced** | **Proportional replacement** | **Risks and benefits modelled** | **Difference in input data from the main analyses** |
| --- | --- | --- | --- | --- | --- |
| **1. Replacement of discretionary salt and non-discretionary salt** | Global population aged 25 years and above (n=4.63 billion; 95% UI: 4.59 to 4.67) | Discretionary salt and salt from processed and restaurant foods | 50% | Benefits from blood pressure lowering and potential harms from raising potassium consumption in people with kidney disease. | Same as the primary analysis |
| **2. Replacement of discretionary salt and non-discretionary salt** | Global population aged 25 years and above (n=4.63 billion; 95% UI: 4.59 to 4.67) | Discretionary salt and salt from processed and restaurant foods | 20% |  | Same as the primary analysis |
| **3. Replacement of discretionary salt and non-discretionary salt** | Global population aged 25 years and above (n=4.63 billion; 95% UI: 4.59 to 4.67) | Discretionary salt and salt from processed and restaurant foods | 100% |  | In the main analysis, we used dose-response data of sodium reduction on SBP from meta-regression by Mozaffarian et al. (2014).^31^ In this sensitivity analysis, we used dose-response data of sodium reduction on SBP from a more recent meta-analysis by Filippini et al (2021).^41^ This new data allowed for different magnitudes of effects for hypertensive versus non-hypertensive individuals but not for different age groups. All other parameters stayed the same as scenario one in the main analysis. |
| **4. Replacement of discretionary salt and non-discretionary salt** | Global population aged 25 years and above (n=4.63 billion; 95% UI: 4.59 to 4.67) | Discretionary salt and salt from processed and restaurant foods | 100% |  | In the main analysis, we assumed that the effect of sodium reduction and potassium increase on SBP is additive. In this sensitivity analysis, we assumed the overall effect on SBP from sodium reduction and potassium increase is only 80% of the additive effect. All other parameters stayed the same as scenario one in the main analysis. |
| **5. Replacement of discretionary salt and non-discretionary salt** | Global population aged 25 years and above (n=4.63 billion; 95% UI: 4.59 to 4.67) | Discretionary salt and salt from processed and restaurant foods | 100% |  | In this analysis, we assumed that the effect of potassium increase on SBP is only 50% of the effect estimated in the main analysis. All other parameters stayed the same as scenario one in the main analysis. |
| **6. Replacement of discretionary salt and non-discretionary salt** | Global population aged 25 years and above (n=4.63 billion; 95% UI: 4.59 to 4.67) | Discretionary salt and salt from processed and restaurant foods | 100% |  | In the main analysis, we assumed potassium supplementation has the same effect among individuals with/without hypertension. In this analysis, we assumed potassium supplementation is only effective at reducing blood pressure among individuals with hypertension. |
| **7. Replacement of discretionary salt and non-discretionary salt** | Global population aged 25 years and above (n=4.63 billion; 95% UI: 4.59 to 4.67) | Discretionary salt and salt from processed and restaurant foods | 100% |  | In the main analyses, we fixed the mix of salt substitute as 75% NaCl +25% KCl. In this analysis, we modelled the effect of full replacement of both discretionary and non-discretionary salt using a mix of 85% NaCl+15% KCl potassium-enriched salt. All other parameters stayed the same as scenario one in the main analysis. |
| **8. Replacement of discretionary salt** | Global population aged 25 years and above (n=4.63 billion; 95% UI: 4.59 to 4.67) | Discretionary salt | 100% |  | In the main analysis, proportion of discretionary salt was estimated from regression-based model for all countries. The regression model was developed using data from 24 countries with reported data on proportion of discretionary salt. The main analysis used only regression-generated estimates for all countries to 1) provide consistency across countries and 2) potentially provide more recent estimates for the 24 countries with reported data as a lot of the reported data were quite old. In this analysis, we used the reported data for the 24 countries with data and for the rest of the countries, we used the regression-generated estimate. All other parameters stayed the same as scenarios two and three the main analyses. |
| **9. Replacement of non-discretionary salt** | Global population aged 25 years and above (n=4.63 billion; 95% UI: 4.59 to 4.67) | Salt from processed and restaurant foods | 100% |  |  |
| **10. Replacement of discretionary salt among those with diagnosed hypertension** | 80% of people with diagnosed hypertension aged 25 years and above in high-income countries and 70% of people with diagnosed hypertension aged 25 years and above in low and middle income countries (n=601 million; 95% UI: 588 to 614) | Discretionary salt | 100% | Benefits from blood pressure lowering with no potential harms from raising potassium consumption in people with CKD stages 3-5. | In the main analysis, we assumed 20% individuals with diagnosed hypertension across all countries have CKD stages 3-5 and would be advised by clinicians to not use potassium-enriched salt. In this analysis, we assumed that the prevalence of CKD stages 3-5 among hypertensive individuals in low and middle-income countries is higher than 20%, i.e. 30% individuals with hypertension in low and middle-income countries have CKD stages 3-5 and 20% individuals with hypertension in high income countries have CKD stages 3-5, and they would be advised by clinicians to not use potassium-enriched salt. All other parameters stayed the same as scenarios four and five in the main analyses. |
| **11. Replacement of discretionary salt among those with treated hypertension** | 80% of people with treated hypertension aged 25 years and above in high-income countries and 70% of people with treated hypertension aged 25 years and above in low and middle income countries (n=480 million; 95% UI: 470 to 492) | Discretionary salt | 100% |  |  |

### **Supplementary table 4. Effect of replacing regular salt with potassium-enriched salt to reduce CVD and CKD burden in the total population for each country**

#### a. **Prevented CVD deaths**

| **Country** | **Sex** | **Baseline disease burden Median (95% UI)** | **Replacement of discretionary salt and non-discretionary salt** | | **Replacement of discretionary salt** | | **Replacement of non-discretionary salt** | | **Replacement of discretionary salt among those with diagnosed hypertension** | | **Replacement of discretionary salt among those with treated hypertension** | |
| --- | --- | --- | --- | --- | --- | --- | --- | --- | --- | --- | --- | --- |
|  |  |  | **No. of events averted Median (95% UI)** | **Percentage of national disease burden averted % (95% UI)** | **No. of events averted Median (95% UI)** | **Percentage of national disease burden averted % (95% UI)** | **No. of events averted Median (95% UI)** | **Percentage of national disease burden averted % (95% UI)** | **No. of events averted Median (95% UI)** | **Percentage of national disease burden averted % (95% UI)** | **No. of events averted Median (95% UI)** | **Percentage of national disease burden averted % (95% UI)** |
| **Global, millions** | **Female** | 8.88 (8.76-9.01) | 1.13 (1.05-1.22) | 12.77 (11.83-13.70) | 0.71 (0.65-0.77) | 7.96 (7.30-8.66) | 0.59 (0.54-0.64) | 6.64 (6.06-7.25) | 0.26 (0.23-0.28) | 2.91 (2.65-3.19) | 0.21 (0.19-0.24) | 2.41 (2.18-2.66) |
| **Global, millions** | **Male** | 9.91 (9.77-10.06) | 1.66 (1.53-1.79) | 16.73 (15.53-18.02) | 1.05 (0.95-1.15) | 10.57 (9.67-11.58) | 0.87 (0.79-0.96) | 8.84 (8.00-9.67) | 0.30 (0.27-0.33) | 2.98 (2.71-3.31) | 0.24 (0.22-0.26) | 2.41 (2.18-2.68) |
| Afghanistan | Female | 21392 (19619-23080) | 128 (22-374) | 0.60 (0.10-1.74) | 116 (19-351) | 0.54 (0.09-1.62) | 92 (15-267) | 0.43 (0.07-1.21) | 28 (3-116) | 0.13 (0.01-0.53) | 23 (2-95) | 0.11 (0.01-0.45) |
| Afghanistan | Male | 23046 (21404-24703) | 1533 (919-2258) | 6.69 (4.09-9.72) | 1207 (730-1764) | 5.27 (3.21-7.62) | 760 (476-1090) | 3.30 (2.05-4.71) | 228 (132-358) | 0.98 (0.57-1.53) | 182 (105-285) | 0.79 (0.46-1.23) |
| Albania | Female | 6965 (6560-7347) | 1358 (1082-1637) | 19.57 (15.49-23.39) | 885 (682-1114) | 12.72 (9.88-15.84) | 621 (470-789) | 8.95 (6.75-11.21) | 463 (349-606) | 6.63 (5.08-8.75) | 413 (306-549) | 5.92 (4.36-7.86) |
| Albania | Male | 6715 (6351-7088) | 1585 (1282-1865) | 23.60 (19.41-27.46) | 1092 (857-1329) | 16.24 (12.75-19.63) | 772 (594-1001) | 11.51 (8.96-14.75) | 295 (210-394) | 4.37 (3.12-5.78) | 215 (147-307) | 3.20 (2.19-4.52) |
| Algeria | Female | 47079 (43337-50949) | 147 (14-505) | 0.31 (0.03-1.07) | 136 (11-483) | 0.29 (0.03-1.02) | 121 (10-395) | 0.26 (0.02-0.84) | 37 (0-196) | 0.08 (0.00-0.43) | 31 (0-180) | 0.07 (0.00-0.37) |
| Algeria | Male | 47246 (44302-50294) | 2326 (1352-3430) | 4.94 (2.90-7.19) | 1828 (1111-2760) | 3.89 (2.37-5.75) | 1286 (776-1915) | 2.71 (1.66-4.05) | 482 (245-829) | 1.02 (0.52-1.73) | 400 (188-690) | 0.85 (0.40-1.46) |
| American Samoa | Female | 58 (55-61) | 6 (4-7) | 9.61 (7.30-11.92) | 4 (3-4) | 6.05 (4.46-7.62) | 3 (2-4) | 5.57 (4.15-7.01) | 1 (1-2) | 2.22 (1.57-2.99) | 1 (1-1) | 1.72 (1.14-2.44) |
| American Samoa | Male | 78 (74-82) | 8 (6-10) | 10.04 (7.48-12.72) | 5 (4-6) | 6.36 (4.69-8.09) | 5 (3-6) | 5.87 (4.30-7.43) | 2 (1-2) | 2.04 (1.41-2.71) | 1 (1-2) | 1.51 (1.03-2.13) |
| Andorra | Female | 88 (81-95) | 3 (2-6) | 3.77 (1.73-6.62) | 2 (1-3) | 2.30 (1.09-3.81) | 3 (1-5) | 3.42 (1.55-5.74) | 1 (0-2) | 1.14 (0.50-2.03) | 1 (0-2) | 1.03 (0.44-1.86) |
| Andorra | Male | 64 (60-69) | 6 (5-8) | 9.81 (7.51-12.10) | 3 (2-3) | 3.96 (2.93-5.07) | 5 (3-6) | 7.12 (5.43-8.98) | 1 (1-2) | 1.85 (1.25-2.53) | 1 (1-2) | 1.71 (1.17-2.35) |
| Angola | Female | 15710 (14560-16794) | 1437 (961-1963) | 9.14 (6.15-12.45) | 1095 (727-1491) | 6.96 (4.69-9.34) | 701 (487-953) | 4.47 (3.09-6.01) | 353 (218-521) | 2.25 (1.40-3.29) | 162 (90-268) | 1.03 (0.57-1.72) |
| Angola | Male | 14731 (13762-15729) | 1329 (924-1750) | 9.00 (6.39-11.91) | 998 (692-1326) | 6.76 (4.79-8.91) | 634 (453-839) | 4.29 (3.11-5.64) | 214 (132-323) | 1.45 (0.90-2.17) | 131 (78-209) | 0.89 (0.53-1.39) |
| Antigua and Barbuda | Female | 95 (93-97) | 11 (8-14) | 11.31 (8.30-14.60) | 7 (5-9) | 6.95 (5.12-9.06) | 6 (5-8) | 6.49 (4.80-8.53) | 3 (2-5) | 3.55 (2.47-4.84) | 3 (2-4) | 3.00 (2.02-4.25) |
| Antigua and Barbuda | Male | 97 (93-100) | 14 (11-17) | 14.31 (10.88-17.31) | 8 (6-10) | 8.49 (6.30-10.76) | 8 (6-10) | 7.90 (5.98-9.94) | 3 (2-4) | 3.39 (2.39-4.67) | 3 (2-4) | 2.85 (1.95-4.02) |
| Argentina | Female | 39841 (38792-41002) | 5278 (4240-6349) | 13.25 (10.58-15.91) | 3340 (2615-4205) | 8.40 (6.56-10.54) | 2637 (2013-3327) | 6.61 (5.06-8.37) | 1735 (1312-2247) | 4.36 (3.32-5.61) | 1541 (1176-2028) | 3.88 (2.93-5.09) |
| Argentina | Male | 40802 (39960-41652) | 6460 (5361-7668) | 15.78 (13.13-18.68) | 4083 (3279-4955) | 9.98 (8.08-12.14) | 3198 (2569-3957) | 7.84 (6.30-9.69) | 1911 (1512-2389) | 4.68 (3.69-5.82) | 1657 (1306-2088) | 4.07 (3.20-5.09) |
| Armenia | Female | 6756 (6524-7001) | 815 (610-1065) | 12.08 (9.08-15.52) | 560 (409-726) | 8.26 (6.08-10.78) | 380 (276-507) | 5.61 (4.09-7.53) | 255 (175-360) | 3.77 (2.60-5.30) | 228 (152-328) | 3.39 (2.24-4.82) |
| Armenia | Male | 6544 (6355-6733) | 1045 (842-1235) | 15.98 (12.89-18.87) | 696 (545-839) | 10.62 (8.41-12.76) | 469 (366-587) | 7.16 (5.63-8.98) | 223 (167-295) | 3.41 (2.52-4.53) | 151 (110-204) | 2.31 (1.69-3.11) |
| Australia | Female | 22274 (21286-23345) | 542 (197-1049) | 2.44 (0.88-4.69) | 235 (84-449) | 1.06 (0.38-2.02) | 527 (191-986) | 2.35 (0.85-4.47) | 105 (32-210) | 0.47 (0.14-0.94) | 94 (28-188) | 0.42 (0.13-0.85) |
| Australia | Male | 23195 (22607-23868) | 1287 (810-1860) | 5.54 (3.50-8.00) | 397 (233-596) | 1.71 (0.99-2.55) | 1165 (745-1644) | 5.03 (3.27-7.09) | 155 (83-246) | 0.67 (0.36-1.06) | 143 (76-228) | 0.62 (0.32-0.98) |
| Austria | Female | 15527 (14830-16208) | 1906 (1413-2372) | 12.26 (9.16-15.27) | 589 (360-869) | 3.79 (2.30-5.60) | 1522 (1081-1973) | 9.81 (7.01-12.72) | 302 (175-475) | 1.95 (1.15-3.03) | 273 (159-428) | 1.76 (1.02-2.75) |
| Austria | Male | 12898 (12531-13284) | 1998 (1603-2424) | 15.51 (12.43-18.86) | 601 (421-827) | 4.67 (3.27-6.45) | 1562 (1223-1943) | 12.14 (9.53-15.17) | 300 (199-437) | 2.32 (1.54-3.42) | 293 (196-428) | 2.28 (1.52-3.34) |
| Azerbaijan | Female | 17537 (16744-18237) | 1992 (1488-2555) | 11.36 (8.47-14.50) | 1382 (1022-1774) | 7.89 (5.85-10.08) | 947 (703-1250) | 5.40 (4.00-7.06) | 680 (499-908) | 3.88 (2.84-5.16) | 577 (412-783) | 3.28 (2.35-4.46) |
| Azerbaijan | Male | 17710 (17075-18407) | 2900 (2355-3440) | 16.40 (13.27-19.30) | 1937 (1543-2347) | 10.90 (8.69-13.24) | 1321 (1051-1671) | 7.44 (5.98-9.39) | 651 (494-843) | 3.68 (2.79-4.75) | 470 (351-632) | 2.66 (1.97-3.55) |
| Bahrain | Female | 522 (495-550) | 2 (0-8) | 0.41 (0.06-1.61) | 2 (0-7) | 0.38 (0.04-1.36) | 2 (0-8) | 0.38 (0.05-1.52) | 1 (0-3) | 0.11 (0.00-0.55) | 0 (0-3) | 0.09 (0.00-0.50) |
| Bahrain | Male | 830 (788-870) | 59 (38-81) | 7.07 (4.64-9.79) | 35 (23-47) | 4.22 (2.81-5.67) | 41 (27-56) | 4.90 (3.36-6.78) | 9 (5-13) | 1.06 (0.61-1.56) | 7 (4-11) | 0.89 (0.50-1.35) |
| Bangladesh | Female | 161076 (149458-171768) | 21812 (17721-25866) | 13.57 (11.14-15.95) | 15243 (11974-18648) | 9.43 (7.57-11.41) | 9562 (7370-12055) | 5.97 (4.65-7.38) | 3956 (2998-5120) | 2.46 (1.87-3.10) | 3156 (2399-4117) | 1.96 (1.51-2.53) |
| Bangladesh | Male | 206865 (193758-220131) | 28109 (23014-33116) | 13.63 (11.21-15.98) | 19695 (15760-23862) | 9.52 (7.72-11.46) | 12308 (9710-15311) | 5.97 (4.72-7.33) | 3329 (2522-4195) | 1.61 (1.22-2.03) | 2597 (1980-3334) | 1.26 (0.96-1.60) |
| Barbados | Female | 472 (455-488) | 26 (15-39) | 5.55 (3.28-8.19) | 19 (12-27) | 4.05 (2.58-5.63) | 18 (12-25) | 3.82 (2.46-5.30) | 11 (7-16) | 2.36 (1.42-3.33) | 10 (6-15) | 2.19 (1.29-3.12) |
| Barbados | Male | 418 (398-438) | 43 (32-54) | 10.23 (7.59-12.87) | 25 (19-33) | 6.05 (4.56-7.82) | 24 (18-31) | 5.73 (4.30-7.44) | 12 (9-16) | 2.87 (2.08-3.86) | 11 (8-15) | 2.61 (1.89-3.54) |
| Belarus | Female | 41540 (39532-43376) | 1512 (742-2502) | 3.65 (1.79-6.10) | 1301 (675-2053) | 3.14 (1.64-4.95) | 1022 (569-1604) | 2.47 (1.37-3.86) | 830 (407-1342) | 2.00 (0.97-3.24) | 665 (336-1125) | 1.61 (0.82-2.68) |
| Belarus | Male | 33253 (31757-34694) | 3777 (2952-4636) | 11.34 (8.80-13.84) | 2509 (1915-3119) | 7.52 (5.78-9.34) | 1788 (1386-2287) | 5.39 (4.17-6.87) | 1175 (856-1517) | 3.53 (2.57-4.53) | 812 (577-1082) | 2.44 (1.75-3.23) |
| Belgium | Female | 13206 (12672-13745) | 1328 (1009-1620) | 10.07 (7.70-12.22) | 422 (281-606) | 3.20 (2.15-4.55) | 1039 (768-1333) | 7.89 (5.84-9.99) | 225 (149-325) | 1.71 (1.13-2.48) | 216 (143-318) | 1.64 (1.06-2.37) |
| Belgium | Male | 11937 (11594-12277) | 1548 (1266-1861) | 12.96 (10.52-15.55) | 496 (354-659) | 4.14 (2.97-5.49) | 1207 (984-1470) | 10.13 (8.19-12.22) | 234 (165-321) | 1.95 (1.37-2.68) | 232 (161-318) | 1.94 (1.34-2.65) |
| Belize | Female | 198 (192-203) | 20 (15-25) | 10.15 (7.53-12.59) | 14 (10-18) | 7.08 (5.27-9.01) | 10 (7-13) | 4.93 (3.69-6.34) | 6 (4-8) | 3.09 (2.16-4.08) | 5 (3-7) | 2.53 (1.72-3.47) |
| Belize | Male | 225 (218-232) | 32 (25-39) | 14.25 (11.38-17.04) | 22 (17-27) | 9.63 (7.60-11.77) | 15 (12-19) | 6.69 (5.21-8.32) | 7 (5-9) | 3.06 (2.27-4.02) | 6 (4-8) | 2.50 (1.79-3.37) |
| Benin | Female | 5721 (5326-6095) | 796 (636-941) | 13.93 (11.24-16.24) | 561 (437-679) | 9.78 (7.72-11.79) | 338 (264-427) | 5.93 (4.72-7.38) | 139 (100-181) | 2.44 (1.79-3.13) | 99 (68-135) | 1.73 (1.22-2.31) |
| Benin | Male | 5436 (5069-5776) | 604 (453-742) | 11.14 (8.51-13.57) | 432 (322-537) | 7.95 (5.97-9.91) | 265 (195-339) | 4.88 (3.62-6.16) | 57 (39-76) | 1.05 (0.71-1.39) | 41 (28-57) | 0.75 (0.51-1.04) |
| Bermuda | Female | 83 (79-88) | 8 (6-10) | 9.54 (6.98-12.32) | 1 (0-1) | 0.66 (0.45-1.25) | 8 (6-10) | 9.16 (6.74-11.84) | 0 (0-1) | 0.34 (0.23-0.68) | 0 (0-1) | 0.29 (0.19-0.61) |
| Bermuda | Male | 113 (109-118) | 14 (11-17) | 12.45 (9.93-14.85) | 1 (1-2) | 0.85 (0.64-1.48) | 14 (11-16) | 11.92 (9.51-14.26) | 0 (0-1) | 0.35 (0.24-0.66) | 0 (0-1) | 0.30 (0.20-0.57) |
| Bhutan | Female | 548 (504-588) | 72 (57-87) | 13.23 (10.40-15.64) | 50 (38-62) | 9.20 (7.04-11.26) | 33 (24-41) | 5.93 (4.47-7.52) | 16 (12-21) | 2.95 (2.19-3.82) | 13 (9-17) | 2.33 (1.72-3.09) |
| Bhutan | Male | 720 (665-771) | 98 (77-117) | 13.58 (10.90-16.06) | 68 (53-83) | 9.41 (7.40-11.49) | 44 (33-55) | 6.11 (4.71-7.53) | 17 (12-22) | 2.30 (1.75-2.95) | 11 (8-15) | 1.58 (1.16-2.12) |
| Bolivia | Female | 7062 (6450-7675) | 866 (643-1066) | 12.24 (9.32-14.88) | 598 (433-758) | 8.45 (6.30-10.52) | 385 (277-501) | 5.47 (4.00-6.95) | 237 (164-325) | 3.35 (2.33-4.57) | 211 (143-300) | 3.00 (2.04-4.14) |
| Bolivia | Male | 6571 (6062-7117) | 989 (733-1216) | 15.13 (11.44-18.39) | 678 (509-857) | 10.40 (7.77-12.88) | 441 (319-567) | 6.74 (4.95-8.43) | 205 (142-287) | 3.14 (2.19-4.32) | 171 (116-248) | 2.60 (1.76-3.70) |
| Bosnia and Herzegovina | Female | 9983 (9417-10513) | 1907 (1523-2309) | 19.12 (15.41-22.88) | 1224 (956-1548) | 12.34 (9.62-15.46) | 889 (684-1157) | 8.93 (6.87-11.51) | 760 (583-983) | 7.64 (5.88-9.75) | 699 (533-914) | 7.01 (5.40-9.05) |
| Bosnia and Herzegovina | Male | 8253 (7857-8703) | 1920 (1596-2208) | 23.25 (19.51-26.43) | 1303 (1046-1575) | 15.71 (12.87-18.91) | 946 (747-1160) | 11.45 (9.17-13.93) | 596 (461-758) | 7.21 (5.63-9.19) | 554 (424-711) | 6.66 (5.13-8.55) |
| Botswana | Female | 1444 (1350-1538) | 113 (73-156) | 7.86 (5.09-10.80) | 85 (58-116) | 5.88 (4.02-8.08) | 64 (44-85) | 4.39 (3.03-6.00) | 34 (22-50) | 2.38 (1.52-3.45) | 30 (18-44) | 2.07 (1.27-3.03) |
| Botswana | Male | 1360 (1275-1447) | 105 (70-144) | 7.73 (5.15-10.37) | 78 (52-105) | 5.71 (3.81-7.68) | 57 (39-79) | 4.24 (2.90-5.76) | 20 (12-30) | 1.49 (0.88-2.21) | 15 (8-22) | 1.08 (0.61-1.64) |
| Brazil | Female | 169326 (165896-172779) | 21693 (17649-25936) | 12.84 (10.39-15.23) | 14365 (11361-17707) | 8.49 (6.72-10.44) | 10476 (8096-13080) | 6.17 (4.78-7.71) | 8079 (6368-10161) | 4.77 (3.75-5.98) | 7852 (6142-9856) | 4.63 (3.66-5.80) |
| Brazil | Male | 190832 (188335-193615) | 30809 (25282-35984) | 16.13 (13.31-18.80) | 20237 (16343-23849) | 10.58 (8.62-12.48) | 14718 (11757-17739) | 7.70 (6.21-9.31) | 9244 (7418-11156) | 4.83 (3.90-5.84) | 8673 (6968-10503) | 4.54 (3.65-5.50) |
| Brunei | Female | 197 (186-206) | 33 (26-40) | 17.13 (13.59-20.25) | 16 (12-20) | 8.18 (6.32-10.09) | 22 (17-26) | 11.00 (8.58-13.44) | 8 (6-10) | 3.91 (2.94-4.96) | 7 (5-9) | 3.69 (2.70-4.71) |
| Brunei | Male | 283 (269-297) | 60 (49-70) | 21.38 (17.25-24.60) | 29 (23-36) | 10.20 (8.01-12.41) | 39 (31-47) | 13.74 (10.90-16.48) | 11 (8-14) | 3.81 (2.92-4.83) | 10 (8-13) | 3.49 (2.72-4.39) |
| Bulgaria | Female | 39169 (37977-40281) | 8649 (6888-10429) | 22.05 (17.42-26.82) | 5384 (4070-6898) | 13.77 (10.42-17.63) | 4444 (3376-5716) | 11.31 (8.59-14.70) | 3221 (2366-4291) | 8.27 (6.04-10.96) | 2900 (2051-3958) | 7.39 (5.23-10.09) |
| Bulgaria | Male | 34645 (33711-35592) | 9139 (7548-10534) | 26.38 (21.92-30.29) | 5929 (4734-7116) | 17.12 (13.68-20.55) | 4906 (3891-6056) | 14.23 (11.28-17.48) | 2837 (2106-3658) | 8.19 (6.06-10.55) | 2447 (1778-3285) | 7.06 (5.10-9.48) |
| Burkina Faso | Female | 9425 (8627-10215) | 1045 (741-1351) | 11.12 (7.87-14.17) | 752 (543-974) | 8.04 (5.79-10.37) | 457 (320-602) | 4.85 (3.45-6.41) | 174 (115-251) | 1.84 (1.21-2.66) | 123 (76-180) | 1.30 (0.81-1.92) |
| Burkina Faso | Male | 11191 (10376-11914) | 1236 (911-1564) | 11.06 (8.15-13.77) | 882 (641-1154) | 7.92 (5.76-10.21) | 531 (383-709) | 4.73 (3.47-6.28) | 138 (91-202) | 1.24 (0.83-1.80) | 74 (47-116) | 0.66 (0.43-1.02) |
| Burundi | Female | 4932 (4549-5294) | 921 (744-1117) | 18.69 (15.46-22.14) | 652 (516-803) | 13.25 (10.70-15.93) | 388 (301-488) | 7.92 (6.22-9.69) | 180 (131-250) | 3.68 (2.69-4.93) | 124 (82-183) | 2.53 (1.68-3.64) |
| Burundi | Male | 5758 (5359-6173) | 773 (586-953) | 13.47 (10.37-16.29) | 558 (422-696) | 9.66 (7.34-11.81) | 329 (244-426) | 5.73 (4.25-7.26) | 107 (70-151) | 1.85 (1.21-2.57) | 73 (43-113) | 1.28 (0.76-1.95) |
| Cambodia | Female | 17809 (16695-18894) | 2904 (2168-3651) | 16.32 (12.17-20.14) | 1995 (1490-2556) | 11.23 (8.32-14.15) | 1241 (897-1624) | 6.99 (5.03-9.05) | 571 (404-778) | 3.21 (2.26-4.35) | 452 (304-637) | 2.54 (1.75-3.57) |
| Cambodia | Male | 15041 (14009-15994) | 2883 (2049-3548) | 19.21 (13.69-23.57) | 2011 (1424-2544) | 13.36 (9.51-16.76) | 1256 (874-1646) | 8.37 (5.87-10.75) | 416 (283-575) | 2.78 (1.88-3.80) | 308 (200-439) | 2.05 (1.34-2.91) |
| Cameroon | Female | 14489 (13407-15602) | 1626 (1201-2037) | 11.25 (8.36-13.91) | 1157 (867-1484) | 8.03 (6.02-10.05) | 717 (528-922) | 4.94 (3.72-6.24) | 300 (213-408) | 2.06 (1.48-2.80) | 175 (116-252) | 1.21 (0.81-1.73) |
| Cameroon | Male | 15772 (14683-17019) | 1847 (1421-2334) | 11.73 (9.07-14.50) | 1317 (1004-1681) | 8.36 (6.46-10.47) | 811 (621-1043) | 5.14 (3.96-6.57) | 222 (154-310) | 1.42 (0.97-1.95) | 137 (91-206) | 0.87 (0.57-1.26) |
| Canada | Female | 35988 (34511-37464) | 2777 (1914-3643) | 7.71 (5.40-10.00) | 920 (617-1298) | 2.55 (1.70-3.63) | 2271 (1590-2963) | 6.28 (4.44-8.19) | 398 (255-588) | 1.10 (0.71-1.61) | 389 (248-579) | 1.08 (0.69-1.59) |
| Canada | Male | 37216 (36215-38229) | 4477 (3523-5370) | 12.02 (9.59-14.37) | 1418 (1012-1903) | 3.81 (2.72-5.09) | 3523 (2751-4346) | 9.48 (7.40-11.61) | 653 (460-904) | 1.75 (1.24-2.42) | 654 (458-909) | 1.75 (1.24-2.42) |
| Cape Verde | Female | 539 (505-574) | 55 (42-69) | 10.27 (7.72-12.83) | 39 (29-49) | 7.19 (5.46-9.20) | 25 (19-32) | 4.60 (3.47-6.02) | 18 (13-24) | 3.30 (2.38-4.43) | 15 (11-21) | 2.87 (1.99-3.90) |
| Cape Verde | Male | 559 (526-592) | 63 (49-77) | 11.22 (8.70-13.58) | 44 (34-54) | 7.79 (6.09-9.66) | 28 (22-36) | 5.02 (3.86-6.36) | 13 (10-17) | 2.33 (1.74-3.04) | 10 (7-14) | 1.80 (1.30-2.42) |
| Central African Republic | Female | 3534 (3197-3872) | 315 (202-433) | 8.86 (5.88-11.90) | 245 (158-329) | 6.90 (4.54-9.19) | 151 (102-207) | 4.27 (2.96-5.78) | 68 (40-101) | 1.93 (1.17-2.78) | 36 (20-58) | 1.03 (0.58-1.58) |
| Central African Republic | Male | 3739 (3384-4070) | 333 (209-460) | 8.87 (5.68-12.11) | 255 (161-341) | 6.82 (4.34-9.08) | 157 (100-216) | 4.20 (2.70-5.64) | 40 (24-60) | 1.08 (0.62-1.59) | 23 (13-35) | 0.62 (0.36-0.92) |
| Chad | Female | 6597 (6095-7153) | 740 (533-949) | 11.23 (7.98-14.14) | 530 (391-695) | 8.05 (5.85-10.31) | 323 (228-424) | 4.88 (3.47-6.33) | 136 (90-193) | 2.07 (1.37-2.90) | 89 (54-132) | 1.34 (0.81-1.97) |
| Chad | Male | 8501 (7878-9118) | 951 (696-1182) | 11.19 (8.17-13.71) | 681 (494-873) | 8.02 (5.86-10.04) | 409 (292-531) | 4.82 (3.40-6.18) | 130 (84-192) | 1.53 (0.99-2.22) | 87 (55-132) | 1.02 (0.64-1.56) |
| Chile | Female | 12774 (12404-13162) | 1814 (1439-2156) | 14.21 (11.34-16.78) | 1089 (839-1339) | 8.55 (6.57-10.51) | 986 (747-1229) | 7.71 (5.85-9.63) | 593 (444-747) | 4.64 (3.49-5.91) | 547 (408-699) | 4.27 (3.19-5.45) |
| Chile | Male | 13508 (13206-13809) | 2242 (1862-2631) | 16.61 (13.77-19.42) | 1325 (1077-1592) | 9.80 (7.99-11.74) | 1203 (962-1464) | 8.91 (7.11-10.86) | 625 (484-775) | 4.62 (3.58-5.71) | 545 (425-681) | 4.04 (3.13-4.99) |
| China | Female | 2207797 (2101879-2307504) | 409425 (333857-483501) | 18.59 (15.17-21.66) | 253787 (202028-310807) | 11.53 (9.06-13.88) | 209594 (166370-257125) | 9.52 (7.50-11.63) | 85277 (65230-106084) | 3.87 (2.96-4.81) | 72071 (54477-91157) | 3.26 (2.46-4.15) |
| China | Male | 2833759 (2708002-2961880) | 636447 (529915-742711) | 22.56 (18.63-26.11) | 411785 (335169-496244) | 14.56 (11.81-17.52) | 343911 (275567-424821) | 12.17 (9.71-14.87) | 113992 (89650-145941) | 4.03 (3.18-5.06) | 91886 (72257-116585) | 3.25 (2.55-4.11) |
| Colombia | Female | 33777 (32303-35250) | 4949 (4082-5992) | 14.66 (12.20-17.58) | 3308 (2679-4035) | 9.79 (8.03-11.83) | 2317 (1861-2917) | 6.88 (5.53-8.55) | 1710 (1349-2173) | 5.06 (4.01-6.33) | 1489 (1174-1904) | 4.41 (3.49-5.55) |
| Colombia | Male | 38208 (36743-39840) | 8215 (6814-9468) | 21.51 (18.05-24.68) | 5517 (4468-6601) | 14.43 (11.78-17.21) | 3899 (3128-4736) | 10.19 (8.18-12.41) | 2253 (1774-2839) | 5.90 (4.65-7.39) | 1865 (1429-2380) | 4.87 (3.76-6.21) |
| Comoros | Female | 543 (501-586) | 100 (81-123) | 18.50 (15.14-22.32) | 70 (55-88) | 12.91 (10.24-15.90) | 44 (33-56) | 8.01 (6.14-10.14) | 21 (15-28) | 3.79 (2.75-5.15) | 14 (9-20) | 2.49 (1.70-3.65) |
| Comoros | Male | 443 (408-475) | 61 (47-74) | 13.74 (10.83-16.48) | 43 (32-53) | 9.72 (7.57-11.78) | 26 (20-34) | 6.00 (4.62-7.55) | 8 (6-12) | 1.90 (1.33-2.61) | 6 (4-8) | 1.29 (0.87-1.87) |
| Congo | Female | 4405 (4067-4704) | 398 (268-531) | 9.08 (6.22-12.03) | 303 (213-406) | 6.89 (4.76-9.12) | 199 (139-265) | 4.55 (3.18-5.99) | 88 (54-132) | 2.00 (1.22-2.94) | 53 (29-86) | 1.20 (0.67-1.92) |
| Congo | Male | 3722 (3470-3999) | 346 (242-452) | 9.24 (6.47-12.06) | 257 (179-330) | 6.86 (4.86-8.92) | 165 (116-218) | 4.42 (3.16-5.77) | 51 (33-76) | 1.38 (0.91-2.04) | 38 (24-59) | 1.03 (0.64-1.58) |
| Costa Rica | Female | 2676 (2584-2773) | 356 (295-430) | 13.32 (11.02-15.95) | 224 (182-277) | 8.34 (6.75-10.37) | 185 (147-233) | 6.91 (5.45-8.65) | 135 (108-172) | 5.06 (4.01-6.45) | 132 (105-168) | 4.93 (3.89-6.31) |
| Costa Rica | Male | 3212 (3124-3304) | 563 (468-657) | 17.52 (14.69-20.44) | 346 (284-413) | 10.74 (8.83-12.75) | 286 (232-343) | 8.91 (7.23-10.65) | 168 (135-204) | 5.22 (4.24-6.38) | 160 (129-196) | 4.97 (4.02-6.12) |
| Cote d'Ivoire | Female | 11126 (10304-11906) | 1401 (1074-1719) | 12.61 (9.75-15.09) | 979 (731-1222) | 8.79 (6.67-10.82) | 617 (461-789) | 5.58 (4.22-6.98) | 254 (179-352) | 2.29 (1.64-3.13) | 169 (113-244) | 1.51 (1.03-2.21) |
| Cote d'Ivoire | Male | 16028 (15041-17180) | 2020 (1552-2472) | 12.60 (9.68-15.34) | 1414 (1069-1770) | 8.82 (6.75-10.88) | 893 (687-1161) | 5.57 (4.34-7.08) | 260 (178-362) | 1.61 (1.12-2.22) | 182 (119-271) | 1.13 (0.73-1.68) |
| Croatia | Female | 13042 (12597-13521) | 2507 (1973-3072) | 19.23 (15.10-23.39) | 1445 (1114-1836) | 11.02 (8.52-14.09) | 1378 (1010-1779) | 10.58 (7.75-13.65) | 981 (756-1271) | 7.51 (5.79-9.71) | 867 (659-1137) | 6.63 (5.01-8.65) |
| Croatia | Male | 9655 (9392-9933) | 2286 (1921-2596) | 23.67 (19.87-26.81) | 1371 (1097-1649) | 14.23 (11.36-17.16) | 1307 (1036-1612) | 13.54 (10.86-16.56) | 827 (648-1023) | 8.57 (6.76-10.56) | 600 (455-771) | 6.21 (4.73-7.94) |
| Cuba | Female | 19355 (18700-19967) | 1824 (1358-2326) | 9.45 (7.04-11.97) | 1193 (889-1573) | 6.16 (4.58-8.07) | 911 (681-1191) | 4.70 (3.52-6.15) | 626 (459-839) | 3.23 (2.35-4.30) | 579 (417-788) | 2.99 (2.15-4.05) |
| Cuba | Male | 20581 (19904-21222) | 2590 (2039-3092) | 12.57 (9.90-15.09) | 1672 (1296-2058) | 8.14 (6.24-9.97) | 1257 (985-1591) | 6.12 (4.76-7.70) | 695 (521-895) | 3.39 (2.51-4.32) | 639 (473-831) | 3.11 (2.29-4.02) |
| Cyprus | Female | 1397 (1311-1487) | 102 (64-142) | 7.33 (4.56-10.31) | 52 (35-75) | 3.73 (2.48-5.33) | 72 (47-102) | 5.17 (3.32-7.25) | 26 (16-41) | 1.88 (1.16-2.93) | 24 (15-37) | 1.74 (1.07-2.67) |
| Cyprus | Male | 1550 (1465-1631) | 184 (147-230) | 11.92 (9.55-14.68) | 87 (67-114) | 5.61 (4.29-7.27) | 122 (95-155) | 7.91 (6.19-9.97) | 40 (30-55) | 2.58 (1.92-3.55) | 37 (27-53) | 2.39 (1.74-3.36) |
| Czech Republic | Female | 23737 (22779-24565) | 4507 (3586-5432) | 18.96 (15.23-22.82) | 2292 (1750-2948) | 9.67 (7.43-12.43) | 2745 (2119-3448) | 11.54 (8.95-14.56) | 1363 (1024-1795) | 5.72 (4.33-7.52) | 1277 (967-1683) | 5.40 (4.09-7.14) |
| Czech Republic | Male | 21666 (21029-22323) | 5085 (4261-5860) | 23.42 (19.67-26.93) | 2732 (2136-3363) | 12.63 (9.96-15.49) | 3236 (2598-3933) | 14.92 (12.01-18.22) | 1591 (1228-2010) | 7.36 (5.71-9.23) | 1519 (1185-1928) | 7.03 (5.47-8.90) |
| Democratic Republic of the Congo | Female | 49969 (45577-53953) | 1095 (324-2300) | 2.21 (0.65-4.59) | 1058 (319-2110) | 2.14 (0.64-4.24) | 833 (279-1589) | 1.68 (0.55-3.18) | 270 (72-637) | 0.54 (0.14-1.30) | 154 (39-409) | 0.31 (0.08-0.81) |
| Democratic Republic of the Congo | Male | 43607 (40187-47131) | 1545 (671-2691) | 3.54 (1.58-6.03) | 1380 (612-2317) | 3.18 (1.41-5.18) | 973 (449-1603) | 2.24 (1.02-3.55) | 256 (96-500) | 0.59 (0.22-1.13) | 196 (70-381) | 0.45 (0.16-0.86) |
| Denmark | Female | 6118 (5892-6346) | 423 (275-565) | 6.88 (4.52-9.21) | 85 (45-148) | 1.39 (0.75-2.42) | 379 (252-513) | 6.19 (4.14-8.39) | 37 (19-67) | 0.61 (0.31-1.10) | 25 (13-48) | 0.41 (0.21-0.78) |
| Denmark | Male | 6481 (6312-6639) | 733 (582-880) | 11.31 (8.99-13.56) | 139 (84-211) | 2.15 (1.31-3.25) | 642 (501-776) | 9.91 (7.75-11.93) | 66 (39-105) | 1.03 (0.60-1.63) | 47 (27-79) | 0.73 (0.42-1.22) |
| Djibouti | Female | 492 (454-531) | 94 (75-115) | 19.17 (15.47-22.79) | 65 (50-80) | 13.19 (10.35-16.00) | 42 (33-53) | 8.48 (6.65-10.66) | 19 (13-25) | 3.76 (2.65-5.13) | 12 (8-19) | 2.51 (1.65-3.73) |
| Djibouti | Male | 776 (710-836) | 104 (79-129) | 13.46 (10.42-16.47) | 74 (55-91) | 9.50 (7.16-11.76) | 47 (35-62) | 6.12 (4.56-7.83) | 14 (9-20) | 1.81 (1.20-2.59) | 10 (6-14) | 1.25 (0.74-1.85) |
| Dominica | Female | 110 (104-116) | 12 (8-15) | 10.56 (7.71-13.59) | 8 (6-10) | 7.11 (5.28-9.23) | 6 (4-8) | 5.18 (3.72-6.99) | 4 (3-6) | 3.89 (2.74-5.28) | 4 (3-5) | 3.34 (2.33-4.63) |
| Dominica | Male | 95 (90-100) | 14 (11-16) | 14.21 (11.21-17.08) | 9 (7-11) | 9.48 (7.18-11.81) | 7 (5-8) | 6.88 (5.15-8.63) | 3 (2-5) | 3.56 (2.57-4.77) | 3 (2-4) | 3.03 (2.17-4.20) |
| Dominican Republic | Female | 10598 (9986-11180) | 1028 (756-1296) | 9.68 (7.19-12.09) | 694 (524-886) | 6.55 (4.93-8.32) | 513 (382-656) | 4.85 (3.61-6.24) | 399 (294-522) | 3.77 (2.75-4.90) | 343 (245-451) | 3.24 (2.32-4.24) |
| Dominican Republic | Male | 13324 (12470-14172) | 1823 (1433-2202) | 13.73 (10.91-16.37) | 1194 (943-1481) | 8.95 (7.09-10.99) | 884 (676-1108) | 6.64 (5.15-8.27) | 551 (413-709) | 4.15 (3.14-5.25) | 477 (357-622) | 3.57 (2.71-4.68) |
| Ecuador | Female | 9753 (9382-10156) | 1248 (990-1499) | 12.79 (10.15-15.32) | 840 (644-1053) | 8.62 (6.62-10.73) | 582 (444-730) | 5.96 (4.59-7.46) | 337 (251-441) | 3.45 (2.57-4.50) | 309 (228-405) | 3.16 (2.35-4.13) |
| Ecuador | Male | 10866 (10384-11367) | 1655 (1281-1984) | 15.23 (11.84-18.19) | 1101 (865-1341) | 10.13 (7.93-12.31) | 768 (577-956) | 7.07 (5.36-8.74) | 332 (246-436) | 3.07 (2.29-3.94) | 270 (198-359) | 2.50 (1.84-3.31) |
| Egypt | Female | 117414 (110804-123873) | 640 (132-2177) | 0.55 (0.12-1.85) | 598 (109-2026) | 0.51 (0.10-1.75) | 519 (82-1648) | 0.45 (0.07-1.42) | 180 (5-863) | 0.15 (0.00-0.74) | 149 (4-734) | 0.13 (0.00-0.63) |
| Egypt | Male | 145799 (137994-153565) | 9677 (6177-13732) | 6.64 (4.24-9.45) | 7482 (4962-10402) | 5.11 (3.35-7.11) | 5184 (3478-7252) | 3.55 (2.39-4.97) | 1752 (1000-2676) | 1.20 (0.68-1.80) | 1450 (832-2249) | 0.99 (0.56-1.53) |
| El Salvador | Female | 4863 (4582-5153) | 628 (509-752) | 12.95 (10.50-15.34) | 425 (334-524) | 8.74 (6.94-10.75) | 286 (222-353) | 5.86 (4.59-7.25) | 219 (167-280) | 4.51 (3.44-5.68) | 200 (154-256) | 4.13 (3.15-5.29) |
| El Salvador | Male | 4591 (4342-4841) | 793 (645-940) | 17.24 (14.16-20.34) | 526 (421-629) | 11.41 (9.21-13.66) | 351 (278-431) | 7.68 (6.12-9.41) | 180 (137-228) | 3.93 (3.04-4.95) | 162 (124-206) | 3.53 (2.74-4.44) |
| Equatorial Guinea | Female | 658 (594-716) | 60 (41-81) | 9.24 (6.34-12.16) | 43 (31-58) | 6.61 (4.62-8.73) | 32 (22-43) | 4.90 (3.49-6.44) | 13 (8-20) | 2.02 (1.23-3.00) | 9 (5-14) | 1.30 (0.74-2.11) |
| Equatorial Guinea | Male | 573 (527-625) | 55 (40-71) | 9.57 (6.93-12.29) | 39 (28-49) | 6.71 (4.83-8.56) | 28 (21-37) | 4.89 (3.60-6.27) | 8 (5-12) | 1.39 (0.92-2.06) | 6 (4-9) | 0.99 (0.61-1.56) |
| Eritrea | Female | 3704 (3406-3996) | 671 (513-832) | 18.15 (13.97-22.14) | 478 (359-597) | 12.90 (9.77-15.90) | 288 (213-373) | 7.78 (5.73-9.96) | 93 (63-130) | 2.49 (1.73-3.51) | 63 (41-95) | 1.72 (1.11-2.55) |
| Eritrea | Male | 3032 (2818-3249) | 366 (272-458) | 12.05 (9.16-15.17) | 266 (192-335) | 8.79 (6.58-10.93) | 160 (113-208) | 5.31 (3.80-6.81) | 32 (21-46) | 1.04 (0.70-1.50) | 23 (14-33) | 0.75 (0.47-1.08) |
| Estonia | Female | 4263 (4077-4423) | 7 (0-80) | 0.17 (0.00-1.88) | 7 (0-71) | 0.16 (0.00-1.68) | 7 (0-76) | 0.17 (0.00-1.78) | 2 (0-26) | 0.05 (0.00-0.60) | 2 (0-19) | 0.04 (0.00-0.46) |
| Estonia | Male | 2799 (2719-2873) | 266 (178-360) | 9.49 (6.36-12.87) | 145 (100-197) | 5.20 (3.59-6.99) | 179 (121-239) | 6.38 (4.33-8.49) | 41 (27-59) | 1.47 (0.96-2.11) | 31 (20-45) | 1.10 (0.71-1.61) |
| Ethiopia | Female | 31742 (29934-33524) | 5599 (4311-6815) | 17.70 (13.58-21.59) | 3949 (3008-4929) | 12.50 (9.40-15.55) | 2429 (1767-3082) | 7.66 (5.59-9.62) | 739 (518-1035) | 2.33 (1.63-3.24) | 373 (229-553) | 1.18 (0.72-1.76) |
| Ethiopia | Male | 34090 (32251-35730) | 4308 (3155-5331) | 12.65 (9.32-15.54) | 3073 (2236-3884) | 9.01 (6.61-11.40) | 1870 (1359-2411) | 5.49 (4.02-7.03) | 464 (316-658) | 1.37 (0.93-1.90) | 316 (209-461) | 0.93 (0.61-1.32) |
| Federated States of Micronesia | Female | 137 (128-146) | 19 (14-24) | 13.81 (10.25-17.05) | 13 (9-17) | 9.56 (6.94-11.97) | 9 (6-11) | 6.28 (4.62-7.96) | 4 (3-5) | 2.89 (2.00-3.82) | 2 (2-3) | 1.76 (1.14-2.40) |
| Federated States of Micronesia | Male | 188 (175-200) | 29 (21-36) | 15.50 (11.14-19.10) | 20 (14-26) | 10.73 (7.58-13.47) | 13 (9-17) | 7.02 (4.92-9.09) | 5 (3-6) | 2.41 (1.60-3.19) | 3 (2-4) | 1.41 (0.89-1.97) |
| Fiji | Female | 1122 (1053-1190) | 182 (148-221) | 16.20 (13.42-19.45) | 123 (99-152) | 10.95 (8.91-13.34) | 82 (65-104) | 7.34 (5.82-9.18) | 54 (41-69) | 4.75 (3.63-6.11) | 41 (28-54) | 3.62 (2.60-4.75) |
| Fiji | Male | 1637 (1518-1756) | 283 (224-339) | 17.33 (13.68-20.59) | 191 (147-238) | 11.65 (9.06-14.29) | 128 (98-163) | 7.83 (6.06-9.81) | 52 (37-69) | 3.17 (2.32-4.18) | 36 (25-51) | 2.21 (1.54-3.11) |
| Finland | Female | 11286 (10727-11820) | 715 (422-1051) | 6.38 (3.76-9.33) | 270 (155-425) | 2.38 (1.39-3.79) | 631 (386-916) | 5.59 (3.46-8.13) | 148 (80-241) | 1.31 (0.71-2.13) | 129 (71-209) | 1.14 (0.62-1.84) |
| Finland | Male | 10562 (10270-10877) | 1351 (1095-1615) | 12.79 (10.36-15.36) | 417 (292-568) | 3.95 (2.78-5.36) | 1077 (847-1314) | 10.18 (8.08-12.54) | 202 (139-282) | 1.92 (1.32-2.67) | 187 (127-264) | 1.77 (1.22-2.50) |
| France | Female | 78492 (75208-81572) | 5495 (3206-7749) | 7.04 (4.05-9.96) | 2309 (1409-3396) | 2.94 (1.82-4.28) | 4259 (2645-6083) | 5.42 (3.39-7.71) | 1114 (665-1721) | 1.42 (0.85-2.20) | 1020 (594-1564) | 1.30 (0.77-2.00) |
| France | Male | 64274 (62428-66079) | 6856 (5391-8345) | 10.67 (8.34-12.85) | 2626 (1911-3510) | 4.09 (3.03-5.47) | 5039 (3951-6234) | 7.86 (6.12-9.72) | 1368 (995-1884) | 2.13 (1.56-2.93) | 1267 (916-1761) | 1.97 (1.42-2.77) |
| Gabon | Female | 1330 (1217-1440) | 128 (86-170) | 9.62 (6.54-12.67) | 89 (61-120) | 6.74 (4.67-8.91) | 68 (47-91) | 5.09 (3.60-6.81) | 29 (19-45) | 2.23 (1.41-3.27) | 22 (13-34) | 1.65 (0.96-2.52) |
| Gabon | Male | 1373 (1274-1474) | 127 (88-167) | 9.32 (6.47-12.10) | 90 (62-118) | 6.55 (4.52-8.49) | 67 (47-90) | 4.89 (3.45-6.52) | 19 (12-29) | 1.42 (0.90-2.09) | 14 (9-22) | 1.05 (0.64-1.63) |
| Georgia | Female | 11585 (11246-11915) | 1506 (1178-1888) | 13.04 (10.17-16.35) | 1048 (788-1332) | 9.06 (6.86-11.54) | 709 (527-926) | 6.11 (4.57-7.97) | 619 (465-815) | 5.35 (4.02-7.03) | 563 (412-765) | 4.88 (3.59-6.61) |
| Georgia | Male | 10742 (10494-10975) | 1858 (1547-2189) | 17.29 (14.32-20.40) | 1251 (1017-1516) | 11.67 (9.51-14.19) | 847 (668-1045) | 7.87 (6.25-9.76) | 565 (441-719) | 5.25 (4.12-6.71) | 448 (347-591) | 4.17 (3.21-5.49) |
| Germany | Female | 159326 (152352-166715) | 11062 (6584-15962) | 6.94 (4.07-10.02) | 4204 (2529-6461) | 2.63 (1.57-3.99) | 9276 (5763-13300) | 5.82 (3.66-8.39) | 2183 (1239-3452) | 1.37 (0.76-2.16) | 2072 (1164-3356) | 1.31 (0.72-2.10) |
| Germany | Male | 146438 (142344-150377) | 18020 (14175-21763) | 12.32 (9.64-14.77) | 5847 (3995-7875) | 3.99 (2.73-5.37) | 14069 (10822-17709) | 9.59 (7.42-11.96) | 3073 (2025-4278) | 2.09 (1.38-2.90) | 2912 (1949-4057) | 1.99 (1.32-2.75) |
| Ghana | Female | 22485 (20870-23998) | 3307 (2645-3927) | 14.72 (11.81-17.25) | 2312 (1763-2831) | 10.30 (8.14-12.51) | 1440 (1099-1806) | 6.40 (4.98-8.06) | 807 (603-1041) | 3.60 (2.71-4.60) | 654 (474-861) | 2.91 (2.14-3.85) |
| Ghana | Male | 23185 (21846-24600) | 3402 (2697-4060) | 14.65 (11.79-17.27) | 2381 (1837-2923) | 10.26 (7.99-12.43) | 1497 (1128-1857) | 6.43 (4.94-8.05) | 444 (324-594) | 1.92 (1.41-2.57) | 342 (249-464) | 1.48 (1.08-2.01) |
| Greece | Female | 25128 (24118-26133) | 1852 (1143-2554) | 7.39 (4.52-10.20) | 1121 (760-1578) | 4.49 (2.99-6.20) | 1133 (732-1594) | 4.52 (2.93-6.29) | 668 (432-945) | 2.66 (1.70-3.74) | 659 (428-945) | 2.63 (1.68-3.72) |
| Greece | Male | 22104 (21454-22669) | 2563 (1980-3113) | 11.57 (8.94-14.02) | 1456 (1098-1834) | 6.60 (4.94-8.25) | 1469 (1116-1859) | 6.66 (5.03-8.41) | 771 (565-994) | 3.49 (2.56-4.50) | 760 (552-983) | 3.43 (2.48-4.43) |
| Greenland | Female | 36 (34-38) | 3 (2-4) | 9.06 (6.65-11.41) | 1 (1-1) | 2.61 (1.70-3.68) | 3 (2-3) | 7.50 (5.58-9.59) | 0 (0-0) | 0.94 (0.57-1.36) | 0 (0-0) | 0.76 (0.47-1.15) |
| Greenland | Male | 62 (59-65) | 8 (6-10) | 13.23 (9.17-16.11) | 2 (1-3) | 3.77 (2.35-5.36) | 7 (5-8) | 10.83 (7.54-13.46) | 1 (0-1) | 1.22 (0.75-1.81) | 1 (0-1) | 1.03 (0.63-1.54) |
| Grenada | Female | 122 (119-125) | 12 (9-16) | 9.92 (7.27-12.59) | 8 (6-11) | 6.63 (4.95-8.64) | 6 (5-8) | 5.06 (3.74-6.58) | 4 (3-6) | 3.52 (2.44-4.73) | 4 (3-5) | 3.10 (2.17-4.20) |
| Grenada | Male | 121 (117-125) | 17 (12-20) | 13.66 (10.13-16.45) | 11 (8-14) | 8.89 (6.51-11.23) | 8 (6-11) | 6.68 (4.89-8.53) | 4 (3-6) | 3.50 (2.38-4.68) | 3 (2-5) | 2.67 (1.78-3.81) |
| Guatemala | Female | 6880 (6636-7126) | 903 (733-1067) | 13.15 (10.63-15.50) | 616 (488-745) | 8.93 (7.00-10.82) | 418 (322-518) | 6.08 (4.67-7.55) | 245 (187-317) | 3.56 (2.75-4.57) | 175 (127-232) | 2.54 (1.84-3.38) |
| Guatemala | Male | 7052 (6771-7332) | 1200 (955-1409) | 16.99 (13.72-19.97) | 798 (633-957) | 11.35 (9.03-13.62) | 540 (427-655) | 7.66 (6.06-9.30) | 225 (169-290) | 3.20 (2.41-4.12) | 164 (119-219) | 2.31 (1.69-3.08) |
| Guinea | Female | 7720 (7186-8267) | 863 (657-1076) | 11.22 (8.49-13.94) | 623 (468-792) | 8.11 (6.11-10.29) | 383 (280-490) | 4.95 (3.63-6.32) | 222 (157-299) | 2.90 (2.05-3.83) | 108 (69-161) | 1.41 (0.89-2.08) |
| Guinea | Male | 7797 (7293-8317) | 862 (648-1066) | 11.08 (8.47-13.51) | 618 (460-775) | 7.94 (6.05-9.91) | 372 (276-478) | 4.82 (3.58-6.12) | 133 (91-186) | 1.71 (1.19-2.41) | 79 (48-119) | 1.02 (0.62-1.52) |
| Guinea-Bissau | Female | 1288 (1188-1384) | 152 (112-189) | 11.86 (8.79-14.40) | 110 (80-139) | 8.55 (6.29-10.58) | 67 (48-87) | 5.14 (3.82-6.64) | 31 (22-44) | 2.44 (1.68-3.39) | 21 (13-30) | 1.60 (1.04-2.37) |
| Guinea-Bissau | Male | 1268 (1190-1354) | 152 (110-190) | 11.98 (8.78-14.81) | 109 (79-140) | 8.64 (6.21-10.82) | 66 (49-87) | 5.20 (3.88-6.75) | 20 (13-29) | 1.60 (1.05-2.26) | 13 (8-21) | 1.05 (0.60-1.64) |
| Guyana | Female | 969 (932-1010) | 99 (73-125) | 10.21 (7.52-12.89) | 67 (49-86) | 6.90 (5.01-8.94) | 52 (38-67) | 5.36 (3.93-6.91) | 31 (23-42) | 3.23 (2.34-4.26) | 25 (17-34) | 2.56 (1.77-3.49) |
| Guyana | Male | 1060 (1008-1111) | 155 (118-187) | 14.61 (11.29-17.56) | 100 (75-126) | 9.42 (7.11-11.87) | 77 (57-98) | 7.23 (5.50-9.28) | 34 (25-44) | 3.17 (2.34-4.15) | 28 (20-37) | 2.61 (1.90-3.47) |
| Haiti | Female | 14294 (13220-15361) | 1375 (1006-1788) | 9.68 (7.19-12.29) | 1023 (760-1305) | 7.14 (5.40-9.08) | 642 (480-840) | 4.49 (3.39-5.74) | 419 (302-561) | 2.92 (2.15-3.89) | 228 (159-317) | 1.59 (1.15-2.20) |
| Haiti | Male | 11927 (11066-12762) | 1657 (1313-1949) | 13.90 (11.15-16.11) | 1165 (923-1425) | 9.74 (7.69-11.70) | 715 (559-888) | 6.03 (4.69-7.45) | 308 (230-396) | 2.59 (1.94-3.31) | 178 (125-242) | 1.50 (1.07-2.02) |
| Honduras | Female | 8568 (7997-9131) | 1183 (940-1438) | 13.83 (11.08-16.54) | 825 (643-1031) | 9.62 (7.45-11.75) | 523 (402-675) | 6.12 (4.71-7.74) | 401 (294-522) | 4.72 (3.48-6.10) | 378 (274-490) | 4.44 (3.28-5.72) |
| Honduras | Male | 8470 (7872-9029) | 1574 (1250-1887) | 18.58 (14.66-22.04) | 1067 (841-1305) | 12.62 (9.98-15.27) | 683 (532-866) | 8.07 (6.28-10.14) | 373 (280-503) | 4.43 (3.31-5.81) | 338 (248-455) | 3.98 (2.94-5.36) |
| Hungary | Female | 31194 (30163-32134) | 6636 (5397-7872) | 21.31 (17.37-25.31) | 3801 (2954-4759) | 12.20 (9.48-15.40) | 3710 (2861-4591) | 11.86 (9.13-14.82) | 2273 (1687-3022) | 7.26 (5.43-9.65) | 2041 (1532-2763) | 6.58 (4.87-8.81) |
| Hungary | Male | 24321 (23655-24978) | 6478 (5573-7328) | 26.65 (22.90-29.96) | 3984 (3227-4750) | 16.39 (13.29-19.55) | 3864 (3142-4679) | 15.92 (12.92-19.24) | 1979 (1502-2505) | 8.13 (6.18-10.30) | 1792 (1400-2296) | 7.38 (5.75-9.48) |
| Iceland | Female | 345 (328-362) | 23 (14-33) | 6.72 (4.03-9.37) | 5 (3-8) | 1.44 (0.74-2.46) | 21 (13-29) | 6.08 (3.82-8.48) | 3 (1-5) | 0.81 (0.40-1.45) | 3 (1-5) | 0.74 (0.36-1.31) |
| Iceland | Male | 343 (330-355) | 46 (37-56) | 13.37 (10.85-16.16) | 9 (6-14) | 2.61 (1.66-3.94) | 40 (32-49) | 11.70 (9.48-14.13) | 5 (3-8) | 1.46 (0.88-2.30) | 5 (3-7) | 1.38 (0.81-2.19) |
| India | Female | 1212629 (1174542-1252060) | 142499 (113702-165417) | 11.77 (9.38-13.54) | 99014 (77450-119810) | 8.18 (6.38-9.81) | 62203 (47566-75997) | 5.13 (3.91-6.23) | 22853 (17164-29362) | 1.89 (1.41-2.42) | 20226 (15128-25387) | 1.67 (1.24-2.11) |
| India | Male | 1598207 (1545065-1648495) | 239347 (189630-277639) | 15.00 (12.04-17.36) | 165174 (128677-196088) | 10.34 (8.10-12.27) | 103208 (79938-127792) | 6.47 (5.06-7.93) | 28743 (21878-36005) | 1.80 (1.37-2.25) | 24088 (18137-30611) | 1.51 (1.14-1.90) |
| Indonesia | Female | 355331 (339077-371891) | 69678 (57305-82579) | 19.64 (16.16-23.01) | 46582 (37867-56305) | 13.12 (10.53-15.82) | 30980 (24826-38473) | 8.73 (6.98-10.70) | 12838 (9883-16336) | 3.62 (2.78-4.58) | 6957 (5252-9055) | 1.96 (1.48-2.54) |
| Indonesia | Male | 400811 (381781-420580) | 90643 (74200-105884) | 22.63 (18.59-26.07) | 61144 (48525-74146) | 15.27 (12.11-18.13) | 40720 (32204-49894) | 10.19 (8.08-12.38) | 11303 (8621-14336) | 2.82 (2.16-3.55) | 6573 (4831-8584) | 1.64 (1.20-2.11) |
| Iran | Female | 76137 (73740-78520) | 210 (10-881) | 0.28 (0.01-1.16) | 199 (10-801) | 0.26 (0.01-1.04) | 183 (10-712) | 0.24 (0.01-0.94) | 60 (0-344) | 0.08 (0.00-0.46) | 51 (0-294) | 0.07 (0.00-0.40) |
| Iran | Male | 87042 (84964-89125) | 4397 (2652-6278) | 5.06 (3.06-7.21) | 3441 (2159-4854) | 3.95 (2.48-5.60) | 2409 (1530-3427) | 2.76 (1.76-3.94) | 833 (446-1285) | 0.95 (0.50-1.48) | 673 (356-1052) | 0.77 (0.41-1.19) |
| Iraq | Female | 36594 (34594-38506) | 152 (24-537) | 0.42 (0.06-1.45) | 141 (19-498) | 0.39 (0.05-1.36) | 128 (14-414) | 0.35 (0.04-1.13) | 49 (2-224) | 0.13 (0.01-0.62) | 36 (1-183) | 0.10 (0.00-0.50) |
| Iraq | Male | 48870 (45843-51972) | 3108 (1882-4350) | 6.35 (3.95-8.93) | 2395 (1480-3342) | 4.92 (3.06-6.83) | 1693 (1134-2303) | 3.49 (2.31-4.71) | 914 (518-1367) | 1.87 (1.06-2.79) | 691 (384-1066) | 1.41 (0.81-2.17) |
| Ireland | Female | 3802 (3620-3982) | 91 (33-170) | 2.38 (0.87-4.41) | 13 (6-29) | 0.34 (0.15-0.78) | 91 (33-168) | 2.38 (0.86-4.39) | 5 (2-13) | 0.14 (0.05-0.35) | 5 (2-12) | 0.13 (0.05-0.32) |
| Ireland | Male | 4229 (4090-4360) | 406 (310-496) | 9.57 (7.37-11.79) | 32 (21-57) | 0.75 (0.50-1.37) | 388 (300-477) | 9.15 (7.08-11.30) | 11 (7-22) | 0.26 (0.17-0.51) | 10 (6-20) | 0.24 (0.15-0.47) |
| Israel | Female | 5006 (4798-5219) | 341 (228-463) | 6.78 (4.65-9.16) | 118 (74-176) | 2.36 (1.49-3.49) | 282 (194-382) | 5.65 (3.88-7.53) | 61 (37-93) | 1.21 (0.74-1.84) | 55 (33-84) | 1.11 (0.67-1.68) |
| Israel | Male | 5117 (4969-5274) | 584 (468-701) | 11.40 (9.15-13.71) | 186 (134-250) | 3.64 (2.60-4.85) | 457 (357-563) | 8.96 (7.03-10.94) | 82 (57-114) | 1.60 (1.11-2.24) | 76 (52-106) | 1.48 (1.01-2.05) |
| Italy | Female | 117004 (111062-122674) | 11449 (7308-15870) | 9.75 (6.23-13.45) | 5231 (3302-7688) | 4.47 (2.84-6.46) | 8028 (5285-11459) | 6.86 (4.53-9.71) | 2992 (1855-4459) | 2.56 (1.56-3.76) | 2790 (1694-4245) | 2.38 (1.46-3.56) |
| Italy | Male | 94077 (92026-96367) | 12411 (9515-15135) | 13.19 (10.05-16.12) | 5506 (4002-7265) | 5.84 (4.26-7.72) | 8473 (6413-10803) | 9.02 (6.78-11.42) | 2861 (2072-3815) | 3.04 (2.20-4.07) | 2757 (1989-3668) | 2.94 (2.12-3.93) |
| Jamaica | Female | 3296 (3171-3424) | 344 (261-436) | 10.48 (7.98-13.19) | 241 (182-312) | 7.32 (5.46-9.55) | 165 (125-216) | 4.99 (3.78-6.50) | 140 (104-189) | 4.25 (3.12-5.69) | 122 (89-165) | 3.71 (2.69-5.05) |
| Jamaica | Male | 2912 (2783-3053) | 411 (328-496) | 14.09 (11.26-16.94) | 279 (218-342) | 9.59 (7.53-11.71) | 190 (148-238) | 6.52 (5.10-8.14) | 116 (87-150) | 3.96 (3.01-5.14) | 96 (71-127) | 3.28 (2.46-4.36) |
| Japan | Female | 189773 (179770-200043) | 24363 (18600-30748) | 12.82 (9.73-16.09) | 10150 (7080-13582) | 5.36 (3.72-7.19) | 17264 (12537-22427) | 9.13 (6.64-11.75) | 5020 (3426-7015) | 2.64 (1.82-3.66) | 4663 (3158-6419) | 2.46 (1.67-3.38) |
| Japan | Male | 171605 (168258-174792) | 27234 (22422-31871) | 15.89 (13.02-18.60) | 10946 (8469-13645) | 6.37 (4.93-8.00) | 18759 (15209-22662) | 10.94 (8.91-13.18) | 5623 (4207-7199) | 3.27 (2.46-4.20) | 4929 (3723-6300) | 2.87 (2.18-3.70) |
| Jordan | Female | 5144 (4861-5428) | 16 (2-68) | 0.32 (0.04-1.31) | 15 (2-66) | 0.30 (0.03-1.26) | 14 (1-55) | 0.27 (0.03-1.09) | 5 (0-32) | 0.10 (0.00-0.62) | 5 (0-30) | 0.10 (0.00-0.60) |
| Jordan | Male | 6184 (5882-6508) | 396 (253-566) | 6.45 (4.11-9.14) | 309 (199-426) | 4.99 (3.25-6.84) | 214 (141-296) | 3.47 (2.28-4.76) | 103 (57-153) | 1.67 (0.92-2.47) | 94 (52-139) | 1.54 (0.84-2.26) |
| Kazakhstan | Female | 33362 (32589-34201) | 3736 (2929-4757) | 11.19 (8.79-14.19) | 2427 (1864-3087) | 7.27 (5.62-9.26) | 1911 (1453-2525) | 5.74 (4.33-7.64) | 1524 (1157-1957) | 4.56 (3.49-5.86) | 1420 (1072-1845) | 4.25 (3.22-5.56) |
| Kazakhstan | Male | 30384 (29547-31182) | 4849 (4025-5701) | 15.98 (13.18-18.60) | 3061 (2453-3645) | 10.10 (8.06-12.02) | 2375 (1908-2901) | 7.82 (6.27-9.51) | 1456 (1149-1787) | 4.80 (3.78-5.89) | 1239 (974-1537) | 4.07 (3.21-5.10) |
| Kenya | Female | 19420 (18153-20649) | 3586 (2865-4363) | 18.52 (15.05-22.13) | 2495 (1978-3072) | 12.83 (10.27-15.56) | 1571 (1211-1983) | 8.12 (6.31-10.06) | 683 (506-933) | 3.52 (2.60-4.72) | 349 (235-500) | 1.78 (1.21-2.54) |
| Kenya | Male | 17160 (16335-18043) | 22 (0-176) | 0.13 (0.00-1.02) | 22 (0-176) | 0.13 (0.00-1.02) | 22 (0-140) | 0.13 (0.00-0.81) | 3 (0-32) | 0.02 (0.00-0.19) | 1 (0-15) | 0.01 (0.00-0.09) |
| Kiribati | Female | 100 (94-106) | 13 (9-17) | 12.99 (9.11-16.35) | 9 (6-12) | 9.12 (6.42-11.58) | 6 (4-8) | 5.72 (3.93-7.47) | 2 (1-3) | 2.00 (1.32-2.77) | 1 (1-2) | 1.04 (0.61-1.56) |
| Kiribati | Male | 164 (153-176) | 25 (18-31) | 15.28 (11.14-18.58) | 18 (13-22) | 10.82 (7.80-13.46) | 11 (8-14) | 6.76 (4.86-8.59) | 3 (2-4) | 1.92 (1.28-2.57) | 2 (1-3) | 1.10 (0.70-1.57) |
| Kuwait | Female | 849 (815-884) | 42 (25-58) | 4.90 (2.90-6.82) | 26 (16-36) | 3.01 (1.84-4.31) | 33 (20-46) | 3.87 (2.41-5.46) | 11 (6-17) | 1.30 (0.70-2.02) | 10 (6-16) | 1.23 (0.64-1.92) |
| Kuwait | Male | 3267 (3112-3426) | 414 (324-499) | 12.65 (10.02-15.13) | 202 (151-255) | 6.16 (4.72-7.77) | 275 (213-340) | 8.42 (6.53-10.32) | 71 (53-92) | 2.16 (1.61-2.82) | 64 (48-83) | 1.96 (1.49-2.54) |
| Kyrgyzstan | Female | 7381 (7091-7684) | 831 (635-1053) | 11.28 (8.59-14.33) | 598 (456-762) | 8.10 (6.14-10.36) | 368 (275-477) | 4.99 (3.78-6.49) | 310 (230-422) | 4.21 (3.15-5.68) | 291 (212-392) | 3.95 (2.85-5.33) |
| Kyrgyzstan | Male | 7674 (7350-7976) | 1252 (994-1504) | 16.36 (12.98-19.52) | 871 (673-1065) | 11.35 (8.86-13.88) | 534 (411-671) | 6.97 (5.40-8.70) | 276 (202-356) | 3.60 (2.68-4.64) | 202 (148-269) | 2.65 (1.93-3.48) |
| Laos | Female | 7051 (6650-7448) | 1260 (988-1513) | 17.86 (14.26-21.39) | 861 (671-1061) | 12.23 (9.66-14.84) | 549 (423-683) | 7.79 (6.05-9.55) | 205 (145-268) | 2.90 (2.07-3.76) | 149 (99-202) | 2.10 (1.41-2.84) |
| Laos | Male | 8281 (7797-8782) | 1770 (1437-2085) | 21.32 (17.46-24.94) | 1220 (971-1466) | 14.69 (11.67-17.52) | 777 (610-962) | 9.38 (7.36-11.46) | 224 (164-299) | 2.70 (1.98-3.60) | 159 (110-219) | 1.92 (1.34-2.62) |
| Latvia | Female | 8670 (8347-8970) | 330 (158-570) | 3.82 (1.83-6.58) | 261 (125-410) | 2.99 (1.43-4.74) | 259 (131-424) | 3.01 (1.51-4.91) | 153 (71-242) | 1.78 (0.82-2.79) | 127 (58-203) | 1.47 (0.67-2.30) |
| Latvia | Male | 5706 (5560-5850) | 839 (692-986) | 14.74 (12.03-17.42) | 471 (375-570) | 8.26 (6.58-9.92) | 480 (383-582) | 8.42 (6.74-10.17) | 208 (157-260) | 3.65 (2.76-4.54) | 168 (125-216) | 2.94 (2.20-3.77) |
| Lebanon | Female | 4712 (4422-5040) | 8 (2-34) | 0.17 (0.03-0.75) | 7 (1-33) | 0.16 (0.03-0.72) | 7 (1-30) | 0.14 (0.02-0.64) | 2 (0-14) | 0.04 (0.00-0.29) | 2 (0-14) | 0.04 (0.00-0.29) |
| Lebanon | Male | 5758 (5427-6065) | 242 (144-369) | 4.21 (2.58-6.37) | 192 (119-292) | 3.35 (2.04-5.03) | 138 (81-206) | 2.40 (1.43-3.57) | 63 (33-107) | 1.10 (0.57-1.84) | 60 (30-100) | 1.05 (0.53-1.73) |
| Lesotho | Female | 2122 (1950-2288) | 168 (104-239) | 7.94 (4.92-11.19) | 135 (83-189) | 6.39 (4.09-8.71) | 87 (55-120) | 4.08 (2.66-5.62) | 53 (30-79) | 2.50 (1.44-3.67) | 49 (29-75) | 2.34 (1.36-3.49) |
| Lesotho | Male | 1423 (1289-1561) | 117 (73-164) | 8.18 (5.29-11.48) | 91 (58-127) | 6.36 (4.13-8.99) | 58 (37-82) | 4.03 (2.63-5.65) | 16 (9-26) | 1.14 (0.61-1.78) | 11 (6-19) | 0.80 (0.43-1.31) |
| Liberia | Female | 2728 (2534-2916) | 314 (237-396) | 11.54 (8.86-14.20) | 228 (173-287) | 8.33 (6.42-10.42) | 137 (104-175) | 5.02 (3.86-6.41) | 65 (46-87) | 2.37 (1.71-3.17) | 41 (27-59) | 1.49 (1.01-2.12) |
| Liberia | Male | 2685 (2519-2860) | 322 (241-395) | 11.98 (9.15-14.50) | 231 (175-292) | 8.60 (6.44-10.74) | 139 (102-177) | 5.18 (3.87-6.47) | 49 (35-69) | 1.83 (1.28-2.57) | 34 (23-50) | 1.28 (0.86-1.81) |
| Libya | Female | 6127 (5717-6531) | 34 (10-92) | 0.56 (0.17-1.51) | 31 (8-85) | 0.50 (0.13-1.38) | 27 (6-71) | 0.43 (0.10-1.18) | 7 (0-33) | 0.11 (0.00-0.55) | 5 (0-27) | 0.09 (0.00-0.44) |
| Libya | Male | 7151 (6655-7610) | 464 (303-655) | 6.50 (4.25-9.03) | 355 (234-486) | 4.94 (3.36-6.71) | 258 (177-361) | 3.61 (2.51-4.97) | 91 (52-140) | 1.26 (0.73-1.95) | 71 (38-113) | 1.00 (0.54-1.58) |
| Lithuania | Female | 11660 (11153-12129) | 1190 (845-1537) | 10.19 (7.33-13.19) | 633 (451-872) | 5.44 (3.84-7.43) | 710 (496-949) | 6.08 (4.29-8.09) | 368 (261-516) | 3.16 (2.24-4.36) | 301 (196-435) | 2.61 (1.70-3.73) |
| Lithuania | Male | 8182 (7965-8426) | 958 (748-1172) | 11.72 (9.20-14.23) | 511 (394-648) | 6.25 (4.79-7.92) | 571 (436-721) | 6.97 (5.35-8.76) | 248 (185-324) | 3.03 (2.25-3.96) | 179 (129-242) | 2.19 (1.59-2.93) |
| Luxembourg | Female | 620 (598-644) | 44 (28-60) | 7.10 (4.50-9.72) | 3 (2-5) | 0.48 (0.33-0.72) | 43 (28-59) | 6.93 (4.46-9.48) | 1 (1-2) | 0.22 (0.15-0.34) | 1 (1-2) | 0.20 (0.13-0.31) |
| Luxembourg | Male | 583 (568-600) | 66 (51-79) | 11.33 (8.82-13.62) | 4 (3-6) | 0.70 (0.53-1.01) | 64 (49-77) | 10.92 (8.52-13.17) | 2 (1-3) | 0.32 (0.24-0.49) | 2 (1-3) | 0.31 (0.23-0.46) |
| Macedonia | Female | 7068 (6716-7407) | 1344 (1050-1637) | 19.04 (15.05-22.94) | 878 (672-1121) | 12.47 (9.63-15.74) | 628 (465-814) | 8.85 (6.68-11.47) | 527 (394-698) | 7.45 (5.57-9.75) | 472 (339-644) | 6.67 (4.77-9.13) |
| Macedonia | Male | 6031 (5717-6325) | 1461 (1215-1681) | 24.21 (20.28-27.74) | 996 (800-1205) | 16.57 (13.23-19.84) | 718 (569-888) | 11.88 (9.39-14.67) | 482 (356-632) | 7.98 (5.98-10.39) | 411 (303-549) | 6.83 (5.01-9.04) |
| Madagascar | Female | 17321 (16077-18541) | 3363 (2614-4088) | 19.48 (15.33-23.10) | 2387 (1830-2934) | 13.82 (10.67-16.58) | 1443 (1085-1832) | 8.33 (6.28-10.36) | 577 (408-800) | 3.34 (2.36-4.59) | 310 (199-453) | 1.79 (1.16-2.64) |
| Madagascar | Male | 18112 (16868-19388) | 2383 (1818-2965) | 13.14 (10.16-16.25) | 1725 (1297-2173) | 9.55 (7.18-11.91) | 1040 (781-1362) | 5.72 (4.31-7.45) | 273 (183-387) | 1.50 (1.03-2.11) | 150 (90-228) | 0.83 (0.49-1.25) |
| Malawi | Female | 8380 (7770-8996) | 1522 (1228-1837) | 18.28 (14.77-21.69) | 1077 (850-1321) | 12.85 (10.24-15.49) | 648 (500-817) | 7.76 (5.97-9.55) | 373 (280-485) | 4.45 (3.36-5.81) | 271 (193-358) | 3.24 (2.34-4.24) |
| Malawi | Male | 9211 (8574-9736) | 1241 (956-1539) | 13.55 (10.55-16.58) | 892 (683-1124) | 9.67 (7.44-12.26) | 535 (403-691) | 5.81 (4.45-7.50) | 148 (105-203) | 1.61 (1.17-2.16) | 104 (74-144) | 1.14 (0.82-1.56) |
| Malaysia | Female | 27643 (26211-29083) | 4827 (3963-5726) | 17.47 (14.52-20.50) | 2991 (2433-3615) | 10.81 (8.88-12.97) | 2389 (1912-2890) | 8.59 (6.97-10.39) | 1259 (987-1574) | 4.56 (3.61-5.60) | 1150 (904-1459) | 4.15 (3.33-5.24) |
| Malaysia | Male | 37999 (36381-39528) | 8094 (6765-9245) | 21.35 (17.78-24.20) | 5053 (4122-6001) | 13.28 (10.89-15.67) | 3991 (3249-4869) | 10.49 (8.57-12.70) | 1666 (1322-2071) | 4.40 (3.45-5.43) | 1528 (1189-1906) | 4.02 (3.14-5.00) |
| Maldives | Female | 188 (178-198) | 32 (26-38) | 16.94 (14.04-20.08) | 20 (16-24) | 10.66 (8.72-12.84) | 16 (12-19) | 8.26 (6.62-10.16) | 7 (5-9) | 3.70 (2.71-4.85) | 6 (4-8) | 2.98 (2.09-4.00) |
| Maldives | Male | 313 (296-329) | 61 (51-70) | 19.43 (16.30-22.26) | 39 (32-46) | 12.29 (10.23-14.49) | 30 (24-36) | 9.54 (7.77-11.38) | 9 (7-12) | 2.99 (2.24-3.86) | 7 (5-10) | 2.31 (1.72-3.12) |
| Mali | Female | 9305 (8603-10079) | 1034 (736-1326) | 11.10 (7.96-14.08) | 743 (532-971) | 8.03 (5.78-10.25) | 453 (325-593) | 4.83 (3.54-6.22) | 247 (169-346) | 2.66 (1.83-3.68) | 179 (115-258) | 1.92 (1.26-2.73) |
| Mali | Male | 6923 (6443-7438) | 732 (527-934) | 10.59 (7.68-13.30) | 526 (378-673) | 7.59 (5.50-9.60) | 316 (229-411) | 4.58 (3.29-5.93) | 125 (82-181) | 1.81 (1.19-2.57) | 91 (58-136) | 1.32 (0.83-1.95) |
| Malta | Female | 704 (673-739) | 61 (42-82) | 8.66 (6.10-11.49) | 29 (20-40) | 4.05 (2.87-5.57) | 42 (30-57) | 5.97 (4.31-8.02) | 15 (10-21) | 2.09 (1.46-2.95) | 14 (10-21) | 2.04 (1.41-2.90) |
| Malta | Male | 570 (552-589) | 74 (58-89) | 12.95 (10.20-15.43) | 33 (25-43) | 5.84 (4.44-7.48) | 50 (39-61) | 8.70 (6.83-10.66) | 17 (12-22) | 2.92 (2.18-3.84) | 16 (12-22) | 2.85 (2.15-3.78) |
| Marshall Islands | Female | 60 (56-64) | 8 (6-10) | 13.51 (10.04-16.80) | 5 (4-7) | 9.10 (6.67-11.56) | 4 (3-5) | 6.42 (4.52-8.22) | 2 (1-2) | 2.51 (1.73-3.30) | 1 (1-2) | 1.85 (1.21-2.51) |
| Marshall Islands | Male | 95 (88-101) | 14 (10-18) | 15.03 (11.11-18.64) | 10 (7-12) | 10.18 (7.42-12.78) | 7 (5-9) | 7.14 (5.20-9.22) | 2 (1-3) | 2.05 (1.43-2.74) | 1 (1-2) | 1.46 (1.00-2.01) |
| Mauritania | Female | 2991 (2756-3218) | 339 (249-422) | 11.31 (8.37-13.98) | 241 (174-307) | 8.04 (5.87-10.03) | 149 (111-196) | 4.97 (3.76-6.44) | 75 (49-104) | 2.49 (1.70-3.43) | 48 (30-74) | 1.62 (1.01-2.43) |
| Mauritania | Male | 2449 (2255-2652) | 267 (201-330) | 10.95 (8.35-13.34) | 189 (141-237) | 7.73 (5.88-9.61) | 117 (88-152) | 4.81 (3.64-6.10) | 40 (27-56) | 1.66 (1.09-2.33) | 28 (17-43) | 1.14 (0.69-1.73) |
| Mauritius | Female | 1612 (1570-1655) | 294 (248-348) | 18.26 (15.26-21.61) | 190 (156-228) | 11.79 (9.65-14.21) | 144 (115-177) | 8.93 (7.16-11.00) | 93 (74-114) | 5.75 (4.62-7.09) | 90 (72-111) | 5.60 (4.50-6.89) |
| Mauritius | Male | 1866 (1823-1906) | 398 (325-462) | 21.37 (17.40-24.65) | 257 (206-311) | 13.80 (11.04-16.57) | 195 (154-235) | 10.45 (8.23-12.65) | 101 (79-126) | 5.42 (4.26-6.72) | 96 (75-121) | 5.14 (4.03-6.50) |
| Mexico | Female | 85699 (83189-88426) | 9531 (7401-11565) | 11.13 (8.61-13.42) | 6097 (4606-7668) | 7.13 (5.41-8.84) | 4762 (3589-5962) | 5.55 (4.20-6.88) | 2679 (1960-3376) | 3.12 (2.30-3.94) | 2454 (1798-3170) | 2.87 (2.10-3.67) |
| Mexico | Male | 98779 (95101-102704) | 13778 (11302-16269) | 13.93 (11.35-16.47) | 8873 (7126-10824) | 9.00 (7.15-10.87) | 6929 (5487-8427) | 7.01 (5.53-8.51) | 2574 (1967-3259) | 2.60 (1.99-3.28) | 2314 (1766-2923) | 2.34 (1.78-2.95) |
| Moldova | Female | 11597 (11230-11977) | 472 (210-777) | 4.06 (1.83-6.75) | 409 (193-650) | 3.53 (1.66-5.54) | 315 (163-484) | 2.72 (1.41-4.15) | 224 (99-359) | 1.93 (0.86-3.11) | 173 (74-285) | 1.50 (0.63-2.46) |
| Moldova | Male | 9978 (9715-10233) | 1181 (903-1443) | 11.84 (9.05-14.46) | 805 (611-1005) | 8.05 (6.16-10.16) | 545 (414-703) | 5.46 (4.14-7.01) | 330 (245-441) | 3.30 (2.46-4.45) | 207 (144-282) | 2.07 (1.44-2.84) |
| Mongolia | Female | 2870 (2741-3005) | 334 (259-410) | 11.62 (9.10-14.09) | 232 (176-287) | 8.08 (6.20-9.93) | 156 (118-197) | 5.42 (4.11-6.79) | 116 (89-149) | 4.07 (3.09-5.20) | 106 (80-141) | 3.69 (2.80-4.85) |
| Mongolia | Male | 3783 (3614-3953) | 672 (555-792) | 17.74 (14.78-20.97) | 453 (366-547) | 11.94 (9.77-14.47) | 302 (241-374) | 7.99 (6.42-9.80) | 183 (143-228) | 4.82 (3.85-6.04) | 138 (108-176) | 3.65 (2.86-4.59) |
| Montenegro | Female | 2246 (2140-2363) | 432 (342-527) | 19.19 (15.27-23.34) | 272 (209-348) | 12.14 (9.41-15.43) | 207 (158-267) | 9.19 (7.04-11.96) | 160 (122-211) | 7.12 (5.37-9.40) | 143 (106-197) | 6.39 (4.69-8.87) |
| Montenegro | Male | 1995 (1912-2079) | 464 (386-534) | 23.35 (19.60-26.65) | 306 (248-373) | 15.40 (12.48-18.39) | 237 (188-292) | 11.84 (9.40-14.52) | 148 (112-191) | 7.40 (5.71-9.45) | 126 (94-167) | 6.31 (4.69-8.28) |
| Morocco | Female | 65983 (61713-70177) | 263 (45-902) | 0.40 (0.07-1.38) | 251 (37-837) | 0.38 (0.06-1.27) | 232 (27-708) | 0.35 (0.04-1.09) | 63 (1-263) | 0.09 (0.00-0.41) | 47 (0-221) | 0.07 (0.00-0.32) |
| Morocco | Male | 65199 (61188-69747) | 3656 (2210-5287) | 5.59 (3.38-8.18) | 2864 (1800-4121) | 4.40 (2.78-6.35) | 1986 (1259-2827) | 3.06 (1.92-4.32) | 639 (340-1015) | 0.98 (0.53-1.54) | 430 (226-721) | 0.66 (0.35-1.10) |
| Mozambique | Female | 14140 (12892-15341) | 2768 (2226-3388) | 19.68 (15.88-23.76) | 1963 (1536-2459) | 13.95 (10.91-17.15) | 1188 (898-1519) | 8.40 (6.37-10.58) | 456 (322-621) | 3.21 (2.32-4.44) | 243 (158-355) | 1.73 (1.13-2.53) |
| Mozambique | Male | 18277 (16985-19477) | 2596 (2016-3216) | 14.29 (11.18-17.46) | 1889 (1443-2362) | 10.32 (7.92-12.90) | 1131 (824-1460) | 6.19 (4.62-7.95) | 242 (165-344) | 1.32 (0.91-1.89) | 123 (74-190) | 0.68 (0.41-1.01) |
| Myanmar | Female | 74512 (69632-78984) | 13491 (10981-15919) | 18.13 (14.92-21.19) | 9325 (7446-11324) | 12.51 (10.04-14.98) | 5712 (4487-7096) | 7.68 (6.08-9.30) | 3473 (2697-4403) | 4.67 (3.65-5.80) | 2251 (1681-2922) | 3.01 (2.27-3.95) |
| Myanmar | Male | 74880 (70453-79957) | 16220 (13093-19093) | 21.71 (17.55-25.16) | 11318 (8912-13707) | 15.10 (11.85-18.17) | 6958 (5445-8673) | 9.28 (7.32-11.50) | 3157 (2380-4061) | 4.22 (3.16-5.41) | 1841 (1342-2466) | 2.47 (1.78-3.28) |
| Namibia | Female | 1891 (1755-2028) | 140 (86-198) | 7.45 (4.59-10.54) | 109 (69-149) | 5.77 (3.63-7.93) | 77 (50-105) | 4.09 (2.56-5.54) | 39 (23-58) | 2.09 (1.24-3.05) | 35 (19-51) | 1.87 (1.02-2.71) |
| Namibia | Male | 1760 (1650-1860) | 134 (85-186) | 7.59 (4.85-10.46) | 101 (64-139) | 5.73 (3.65-7.96) | 70 (47-99) | 3.99 (2.71-5.62) | 27 (16-41) | 1.56 (0.90-2.35) | 24 (14-38) | 1.39 (0.78-2.15) |
| Nauru | Female | 16 (15-17) | 2 (2-3) | 15.02 (12.14-17.75) | 2 (1-2) | 9.53 (7.50-11.68) | 1 (1-2) | 7.72 (6.05-9.45) | 1 (0-1) | 3.29 (2.47-4.34) | 0 (0-0) | 1.99 (1.38-2.81) |
| Nauru | Male | 21 (20-23) | 3 (3-4) | 16.09 (12.89-19.15) | 2 (2-3) | 10.25 (8.13-12.39) | 2 (1-2) | 8.29 (6.45-10.27) | 1 (0-1) | 3.01 (2.30-3.84) | 0 (0-1) | 2.00 (1.45-2.63) |
| Nepal | Female | 24315 (22625-25976) | 2472 (1580-3340) | 10.22 (6.48-13.65) | 1783 (1149-2448) | 7.32 (4.73-10.06) | 1095 (706-1539) | 4.52 (2.90-6.29) | 378 (234-541) | 1.57 (0.97-2.19) | 276 (169-391) | 1.14 (0.71-1.62) |
| Nepal | Male | 29854 (27672-31915) | 3944 (3093-4794) | 13.24 (10.36-15.94) | 2802 (2164-3484) | 9.40 (7.27-11.52) | 1716 (1277-2199) | 5.76 (4.37-7.25) | 495 (370-658) | 1.66 (1.25-2.18) | 294 (214-394) | 0.99 (0.72-1.31) |
| Netherlands | Female | 19949 (19192-20696) | 1183 (774-1647) | 5.93 (3.89-8.23) | 354 (215-565) | 1.78 (1.09-2.86) | 1033 (690-1420) | 5.17 (3.46-7.16) | 160 (92-267) | 0.80 (0.46-1.35) | 134 (77-218) | 0.67 (0.39-1.11) |
| Netherlands | Male | 18496 (18019-18946) | 1873 (1494-2291) | 10.14 (8.00-12.31) | 494 (327-712) | 2.67 (1.77-3.83) | 1544 (1185-1925) | 8.33 (6.40-10.40) | 210 (134-317) | 1.13 (0.72-1.73) | 197 (124-297) | 1.06 (0.66-1.61) |
| New Zealand | Female | 5174 (4969-5389) | 248 (146-368) | 4.77 (2.79-7.13) | 104 (62-154) | 2.02 (1.22-2.93) | 213 (125-317) | 4.14 (2.43-6.11) | 51 (29-78) | 0.98 (0.56-1.49) | 46 (25-70) | 0.89 (0.49-1.34) |
| New Zealand | Male | 5473 (5340-5606) | 568 (451-691) | 10.39 (8.25-12.59) | 192 (138-255) | 3.50 (2.52-4.67) | 441 (349-544) | 8.06 (6.39-9.93) | 77 (53-106) | 1.40 (0.98-1.92) | 69 (48-96) | 1.26 (0.88-1.74) |
| Nicaragua | Female | 2804 (2652-2955) | 392 (320-463) | 14.00 (11.51-16.29) | 274 (218-331) | 9.78 (7.88-11.75) | 170 (135-212) | 6.09 (4.84-7.47) | 131 (100-165) | 4.68 (3.56-5.88) | 121 (92-154) | 4.33 (3.25-5.48) |
| Nicaragua | Male | 2738 (2585-2896) | 495 (407-594) | 18.08 (15.00-21.48) | 341 (279-414) | 12.43 (10.18-14.97) | 214 (173-264) | 7.81 (6.26-9.52) | 116 (89-151) | 4.21 (3.18-5.47) | 105 (78-139) | 3.83 (2.86-5.04) |
| Niger | Female | 9073 (8253-9914) | 1054 (756-1334) | 11.57 (8.44-14.48) | 759 (530-981) | 8.34 (6.01-10.63) | 453 (320-605) | 4.97 (3.57-6.55) | 161 (100-233) | 1.76 (1.12-2.48) | 75 (43-120) | 0.82 (0.48-1.32) |
| Niger | Male | 7045 (6318-7681) | 779 (573-964) | 11.10 (8.38-13.61) | 558 (409-703) | 7.96 (5.94-10.01) | 336 (235-436) | 4.77 (3.50-6.07) | 73 (46-108) | 1.04 (0.66-1.53) | 49 (28-78) | 0.69 (0.40-1.09) |
| Nigeria | Female | 86767 (82068-92407) | 7755 (5451-10094) | 8.95 (6.24-11.52) | 5729 (4249-7464) | 6.60 (4.87-8.47) | 3653 (2694-4774) | 4.21 (3.10-5.46) | 1660 (1167-2228) | 1.91 (1.34-2.54) | 934 (630-1273) | 1.08 (0.73-1.46) |
| Nigeria | Male | 90727 (85259-96272) | 8747 (6363-10932) | 9.64 (7.10-12.00) | 6301 (4681-8057) | 6.94 (5.17-8.82) | 3958 (2885-5020) | 4.36 (3.24-5.50) | 1327 (930-1804) | 1.46 (1.04-1.98) | 939 (659-1300) | 1.03 (0.73-1.44) |
| Norway | Female | 5567 (5358-5790) | 188 (80-317) | 3.38 (1.43-5.63) | 27 (13-62) | 0.47 (0.23-1.10) | 185 (80-314) | 3.34 (1.42-5.54) | 13 (6-32) | 0.23 (0.10-0.57) | 9 (4-21) | 0.15 (0.07-0.39) |
| Norway | Male | 5044 (4938-5144) | 548 (426-655) | 10.87 (8.48-13.06) | 49 (31-89) | 0.97 (0.62-1.77) | 516 (400-625) | 10.21 (7.92-12.41) | 23 (14-45) | 0.46 (0.28-0.89) | 21 (13-41) | 0.41 (0.25-0.81) |
| Oman | Female | 1590 (1495-1683) | 8 (2-29) | 0.51 (0.10-1.84) | 7 (1-26) | 0.45 (0.08-1.63) | 7 (1-25) | 0.45 (0.07-1.57) | 2 (0-10) | 0.12 (0.00-0.62) | 2 (0-9) | 0.11 (0.00-0.56) |
| Oman | Male | 2717 (2566-2867) | 196 (130-265) | 7.25 (4.80-9.64) | 129 (87-172) | 4.76 (3.20-6.36) | 128 (85-173) | 4.71 (3.16-6.27) | 31 (18-47) | 1.15 (0.66-1.70) | 27 (16-40) | 1.01 (0.59-1.49) |
| Pakistan | Female | 162322 (152608-171830) | 23325 (18833-27164) | 14.41 (11.84-16.54) | 16429 (12922-19817) | 10.14 (8.11-12.01) | 10101 (7810-12320) | 6.24 (4.87-7.58) | 5443 (4156-6949) | 3.36 (2.60-4.25) | 4549 (3396-5981) | 2.80 (2.12-3.63) |
| Pakistan | Male | 180501 (169844-191478) | 26742 (21970-31864) | 14.82 (12.25-17.22) | 18885 (15269-22958) | 10.43 (8.52-12.60) | 11588 (9125-14409) | 6.41 (5.11-7.99) | 3409 (2566-4491) | 1.88 (1.44-2.46) | 2589 (1891-3497) | 1.44 (1.06-1.93) |
| Palau | Female | 26 (25-28) | 4 (3-5) | 15.64 (12.38-18.95) | 3 (2-3) | 9.61 (7.43-11.95) | 2 (2-3) | 8.02 (6.25-10.03) | 1 (1-1) | 4.12 (3.12-5.29) | 1 (1-1) | 3.58 (2.66-4.73) |
| Palau | Male | 45 (42-48) | 7 (6-9) | 16.33 (12.71-19.52) | 5 (3-6) | 10.13 (7.85-12.39) | 4 (3-5) | 8.47 (6.43-10.63) | 1 (1-2) | 3.17 (2.38-4.08) | 1 (1-1) | 2.33 (1.69-3.05) |
| Panama | Female | 2392 (2299-2488) | 310 (256-368) | 12.90 (10.73-15.34) | 187 (149-232) | 7.82 (6.23-9.65) | 167 (132-207) | 7.00 (5.50-8.69) | 91 (71-117) | 3.80 (2.97-4.89) | 82 (62-105) | 3.42 (2.59-4.40) |
| Panama | Male | 3056 (2919-3195) | 519 (430-611) | 16.99 (14.22-19.97) | 309 (252-370) | 10.07 (8.28-12.04) | 274 (222-333) | 8.96 (7.36-10.82) | 120 (94-151) | 3.93 (3.09-4.95) | 102 (80-132) | 3.33 (2.62-4.31) |
| Papua New Guinea | Female | 6581 (6155-7043) | 857 (592-1070) | 13.04 (8.90-16.09) | 604 (410-766) | 9.18 (6.21-11.51) | 385 (265-504) | 5.85 (4.06-7.56) | 115 (70-166) | 1.75 (1.04-2.50) | 70 (38-112) | 1.07 (0.58-1.69) |
| Papua New Guinea | Male | 8500 (7904-9110) | 1189 (814-1491) | 14.00 (9.91-17.50) | 838 (582-1068) | 9.90 (6.95-12.43) | 540 (359-700) | 6.32 (4.19-8.19) | 103 (60-155) | 1.21 (0.73-1.84) | 69 (35-109) | 0.82 (0.41-1.28) |
| Paraguay | Female | 4604 (4343-4849) | 636 (517-748) | 13.85 (11.43-16.14) | 429 (335-515) | 9.30 (7.42-11.11) | 296 (231-369) | 6.44 (5.06-7.96) | 232 (178-288) | 5.04 (3.96-6.23) | 191 (145-245) | 4.16 (3.17-5.32) |
| Paraguay | Male | 5762 (5448-6103) | 954 (765-1133) | 16.57 (13.37-19.58) | 637 (499-769) | 11.04 (8.67-13.26) | 444 (348-546) | 7.68 (6.09-9.39) | 280 (212-351) | 4.85 (3.67-6.09) | 230 (170-296) | 3.97 (2.97-5.10) |
| Peru | Female | 13545 (12684-14447) | 1654 (1322-1994) | 12.18 (9.73-14.46) | 1099 (847-1358) | 8.10 (6.24-9.86) | 777 (582-987) | 5.72 (4.36-7.19) | 310 (234-397) | 2.27 (1.74-2.89) | 283 (212-362) | 2.09 (1.59-2.66) |
| Peru | Male | 15552 (14567-16476) | 2399 (1942-2872) | 15.49 (12.70-18.41) | 1585 (1270-1941) | 10.19 (8.21-12.40) | 1117 (887-1362) | 7.18 (5.76-8.71) | 327 (256-421) | 2.11 (1.65-2.70) | 283 (218-365) | 1.82 (1.41-2.31) |
| Philippines | Female | 98479 (95029-101885) | 17212 (13654-20608) | 17.46 (13.83-20.89) | 11674 (9246-14328) | 11.89 (9.46-14.38) | 7636 (5888-9423) | 7.76 (6.02-9.52) | 3904 (2985-4932) | 3.95 (3.04-4.96) | 2912 (2166-3752) | 2.96 (2.22-3.81) |
| Philippines | Male | 123373 (118225-128326) | 26538 (19975-31030) | 21.49 (16.15-25.13) | 18210 (13768-22049) | 14.74 (11.05-17.80) | 11854 (8776-14739) | 9.66 (7.18-11.96) | 4395 (3113-5522) | 3.56 (2.54-4.50) | 3040 (2167-3970) | 2.46 (1.77-3.21) |
| Poland | Female | 89903 (86999-92762) | 14264 (10835-17715) | 15.87 (12.13-19.62) | 8168 (6181-10449) | 9.11 (6.88-11.57) | 7818 (5765-10075) | 8.71 (6.51-11.16) | 5495 (4121-7062) | 6.14 (4.59-7.93) | 5332 (4016-6904) | 5.93 (4.48-7.60) |
| Poland | Male | 75141 (73481-76873) | 15018 (12376-17716) | 19.98 (16.41-23.50) | 8707 (7044-10735) | 11.59 (9.32-14.21) | 8333 (6705-10390) | 11.06 (8.93-13.73) | 4643 (3680-5803) | 6.18 (4.92-7.70) | 4285 (3388-5421) | 5.71 (4.50-7.18) |
| Portugal | Female | 17565 (16835-18280) | 768 (403-1249) | 4.38 (2.31-7.10) | 532 (277-828) | 3.04 (1.60-4.69) | 581 (304-899) | 3.32 (1.72-5.16) | 307 (151-495) | 1.76 (0.86-2.83) | 300 (150-482) | 1.71 (0.85-2.75) |
| Portugal | Male | 14302 (13930-14662) | 1518 (1195-1869) | 10.62 (8.34-13.00) | 818 (635-1018) | 5.72 (4.44-7.10) | 920 (729-1150) | 6.41 (5.10-8.04) | 402 (297-522) | 2.80 (2.08-3.66) | 380 (284-495) | 2.66 (1.97-3.47) |
| Puerto Rico | Female | 3794 (3620-3970) | 397 (288-507) | 10.48 (7.74-13.41) | 192 (137-257) | 5.06 (3.64-6.79) | 267 (193-347) | 7.05 (5.08-9.01) | 100 (70-144) | 2.65 (1.85-3.80) | 86 (57-122) | 2.26 (1.50-3.23) |
| Puerto Rico | Male | 4159 (3991-4333) | 562 (456-673) | 13.50 (11.02-15.96) | 266 (205-339) | 6.40 (4.95-8.12) | 370 (295-457) | 8.94 (7.09-10.92) | 108 (80-146) | 2.60 (1.94-3.52) | 93 (65-130) | 2.24 (1.57-3.12) |
| Qatar | Female | 256 (241-273) | 1 (0-4) | 0.37 (-0.00-1.51) | 0 (0-2) | 0.17 (-0.00-0.71) | 1 (0-4) | 0.36 (-0.00-1.51) | 0 (0-1) | 0.06 (0.00-0.36) | 0 (0-1) | 0.06 (0.00-0.34) |
| Qatar | Male | 750 (698-802) | 56 (37-78) | 7.52 (4.89-10.35) | 14 (8-22) | 1.85 (1.10-2.91) | 52 (33-71) | 6.88 (4.42-9.43) | 4 (2-6) | 0.47 (0.26-0.77) | 3 (2-5) | 0.42 (0.22-0.70) |
| Romania | Female | 77134 (74806-79428) | 15368 (12279-18711) | 19.94 (16.03-24.15) | 9273 (7021-11742) | 12.00 (9.16-15.14) | 8118 (6243-10431) | 10.53 (8.13-13.61) | 5338 (3987-6899) | 6.92 (5.18-8.94) | 4679 (3497-6113) | 6.07 (4.53-7.90) |
| Romania | Male | 64657 (63174-66161) | 15725 (13231-17847) | 24.32 (20.49-27.68) | 9899 (7945-11883) | 15.30 (12.41-18.46) | 8699 (6984-10566) | 13.46 (10.87-16.37) | 4955 (3882-6081) | 7.66 (6.05-9.38) | 4433 (3489-5496) | 6.85 (5.38-8.47) |
| Russia | Female | 516512 (502998-529802) | 50318 (37450-63082) | 9.73 (7.24-12.31) | 31518 (23675-40778) | 6.12 (4.61-7.88) | 25752 (19173-34001) | 4.99 (3.72-6.57) | 18002 (12877-23696) | 3.49 (2.52-4.59) | 12610 (8893-16823) | 2.44 (1.73-3.26) |
| Russia | Male | 379353 (370021-389133) | 64103 (52089-74925) | 16.85 (13.70-19.66) | 39546 (30949-46934) | 10.39 (8.23-12.42) | 32171 (25378-39180) | 8.50 (6.74-10.35) | 16871 (12931-20698) | 4.45 (3.42-5.47) | 12012 (8968-14987) | 3.16 (2.37-3.93) |
| Rwanda | Female | 6753 (6128-7360) | 1221 (939-1522) | 18.10 (13.90-21.97) | 861 (649-1093) | 12.76 (9.69-15.79) | 521 (385-675) | 7.70 (5.77-9.83) | 137 (92-201) | 2.03 (1.36-2.94) | 67 (38-109) | 0.99 (0.56-1.58) |
| Rwanda | Male | 5171 (4763-5613) | 677 (503-846) | 13.04 (10.04-16.24) | 484 (353-613) | 9.32 (7.01-11.73) | 293 (213-381) | 5.63 (4.21-7.27) | 50 (32-74) | 0.96 (0.62-1.41) | 31 (18-49) | 0.60 (0.36-0.95) |
| Saint Kitts and Nevis | Female | 68 (66-71) | 7 (5-8) | 9.77 (7.25-12.12) | 4 (3-5) | 5.91 (4.42-7.61) | 4 (3-5) | 5.64 (4.15-7.13) | 2 (1-3) | 3.14 (2.16-4.10) | 2 (1-3) | 2.85 (1.99-3.83) |
| Saint Kitts and Nevis | Male | 76 (72-80) | 11 (8-13) | 14.32 (11.02-17.01) | 6 (5-8) | 8.38 (6.44-10.46) | 6 (4-8) | 7.99 (5.95-9.91) | 2 (2-3) | 3.08 (2.32-4.08) | 2 (1-3) | 2.54 (1.80-3.38) |
| Saint Lucia | Female | 221 (214-228) | 23 (17-30) | 10.43 (7.84-13.37) | 15 (11-20) | 6.76 (5.06-8.81) | 12 (9-16) | 5.40 (3.96-7.10) | 8 (6-12) | 3.83 (2.78-5.23) | 7 (5-10) | 3.22 (2.30-4.54) |
| Saint Lucia | Male | 221 (214-229) | 30 (24-36) | 13.50 (10.94-16.11) | 19 (15-24) | 8.63 (6.83-10.67) | 15 (12-19) | 6.84 (5.41-8.64) | 7 (6-10) | 3.33 (2.52-4.30) | 6 (4-8) | 2.53 (1.87-3.39) |
| Saint Vincent and the Grenadines | Female | 157 (152-161) | 17 (13-23) | 11.04 (8.14-14.34) | 12 (8-15) | 7.37 (5.39-9.75) | 9 (6-12) | 5.51 (4.01-7.31) | 6 (4-8) | 3.76 (2.59-5.21) | 5 (3-7) | 3.18 (2.17-4.51) |
| Saint Vincent and the Grenadines | Male | 174 (170-178) | 25 (20-30) | 14.64 (11.56-17.37) | 17 (13-21) | 9.61 (7.43-11.89) | 12 (9-16) | 7.13 (5.50-9.07) | 6 (4-8) | 3.54 (2.55-4.67) | 5 (4-7) | 2.85 (2.08-3.87) |
| Samoa | Female | 249 (234-264) | 18 (13-23) | 7.17 (5.32-9.25) | 13 (9-17) | 5.12 (3.63-6.76) | 9 (6-12) | 3.45 (2.39-4.71) | 3 (2-5) | 1.29 (0.81-1.96) | 1 (1-2) | 0.48 (0.22-0.81) |
| Samoa | Male | 290 (275-307) | 19 (14-25) | 6.50 (4.65-8.77) | 14 (10-19) | 4.73 (3.30-6.54) | 9 (6-13) | 3.24 (2.23-4.42) | 3 (2-4) | 1.03 (0.65-1.53) | 2 (1-3) | 0.67 (0.40-1.10) |
| Sao Tome and Principe | Female | 134 (127-142) | 15 (11-18) | 10.84 (8.26-13.33) | 10 (8-13) | 7.69 (5.88-9.65) | 7 (5-8) | 4.84 (3.62-6.10) | 4 (3-5) | 2.86 (2.12-3.71) | 3 (2-3) | 1.88 (1.37-2.54) |
| Sao Tome and Principe | Male | 114 (108-121) | 13 (10-16) | 11.31 (8.83-13.70) | 9 (7-11) | 8.00 (6.27-9.92) | 6 (4-7) | 5.01 (3.82-6.30) | 2 (1-3) | 1.78 (1.30-2.35) | 1 (1-2) | 1.07 (0.74-1.46) |
| Saudi Arabia | Female | 17813 (16754-18835) | 100 (3-270) | 0.56 (0.02-1.52) | 83 (3-225) | 0.47 (0.02-1.24) | 86 (3-240) | 0.48 (0.02-1.34) | 14 (0-67) | 0.08 (0.00-0.37) | 12 (0-54) | 0.06 (0.00-0.31) |
| Saudi Arabia | Male | 35331 (33285-37396) | 2787 (1753-3905) | 7.91 (4.95-10.93) | 1701 (1084-2330) | 4.85 (3.05-6.59) | 1882 (1232-2558) | 5.35 (3.47-7.17) | 352 (198-501) | 0.99 (0.56-1.42) | 281 (167-416) | 0.79 (0.48-1.17) |
| Senegal | Female | 10344 (9644-11061) | 1160 (868-1433) | 11.22 (8.49-13.74) | 824 (621-1031) | 7.98 (6.03-10.07) | 511 (382-658) | 4.92 (3.73-6.34) | 242 (169-330) | 2.34 (1.60-3.14) | 128 (80-187) | 1.23 (0.78-1.81) |
| Senegal | Male | 10381 (9743-10970) | 1165 (871-1460) | 11.25 (8.55-13.91) | 827 (622-1061) | 7.98 (6.01-10.14) | 506 (381-657) | 4.87 (3.68-6.30) | 164 (109-236) | 1.58 (1.08-2.26) | 94 (61-145) | 0.91 (0.61-1.37) |
| Serbia | Female | 37399 (35613-39186) | 7468 (6004-8968) | 19.94 (16.16-23.75) | 4718 (3648-5891) | 12.61 (9.79-15.67) | 3550 (2774-4536) | 9.54 (7.50-12.05) | 3039 (2319-3848) | 8.10 (6.22-10.27) | 2854 (2176-3653) | 7.64 (5.82-9.80) |
| Serbia | Male | 29844 (28535-31150) | 6985 (5869-8064) | 23.41 (19.83-26.86) | 4642 (3728-5594) | 15.53 (12.68-18.49) | 3545 (2842-4346) | 11.88 (9.48-14.51) | 2415 (1874-2963) | 8.07 (6.31-9.93) | 2192 (1717-2765) | 7.33 (5.81-9.17) |
| Seychelles | Female | 116 (110-121) | 22 (17-27) | 18.81 (14.68-23.04) | 13 (10-17) | 11.65 (8.95-14.64) | 11 (8-15) | 9.83 (7.49-12.56) | 7 (5-10) | 6.44 (4.69-8.30) | 7 (5-9) | 5.96 (4.33-7.71) |
| Seychelles | Male | 123 (117-129) | 25 (20-30) | 20.57 (16.67-24.29) | 16 (12-19) | 12.72 (10.05-15.56) | 13 (10-16) | 10.67 (8.38-13.29) | 7 (5-9) | 5.35 (4.13-6.96) | 6 (5-8) | 4.79 (3.67-6.23) |
| Sierra Leone | Female | 5391 (5015-5780) | 602 (441-757) | 11.18 (8.33-13.85) | 435 (323-554) | 8.11 (6.06-10.11) | 260 (193-332) | 4.82 (3.63-6.06) | 119 (83-164) | 2.20 (1.55-3.03) | 71 (46-104) | 1.32 (0.85-1.92) |
| Sierra Leone | Male | 5098 (4766-5467) | 585 (454-733) | 11.52 (8.99-14.10) | 420 (324-534) | 8.29 (6.44-10.30) | 250 (191-318) | 4.92 (3.79-6.20) | 73 (50-102) | 1.44 (0.98-1.98) | 42 (27-65) | 0.83 (0.53-1.24) |
| Singapore | Female | 2629 (2525-2740) | 391 (296-479) | 14.85 (11.44-18.39) | 57 (31-96) | 2.19 (1.19-3.64) | 355 (269-439) | 13.48 (10.33-16.76) | 30 (16-52) | 1.14 (0.60-1.96) | 29 (15-50) | 1.09 (0.58-1.90) |
| Singapore | Male | 3528 (3447-3612) | 634 (469-773) | 17.97 (13.40-21.63) | 96 (51-152) | 2.73 (1.47-4.28) | 575 (422-706) | 16.27 (11.86-19.88) | 43 (22-70) | 1.22 (0.63-1.99) | 41 (21-66) | 1.15 (0.60-1.88) |
| Slovakia | Female | 14594 (13947-15257) | 2820 (2249-3418) | 19.33 (15.46-23.31) | 1527 (1163-1933) | 10.46 (8.03-13.22) | 1608 (1236-2059) | 11.00 (8.48-14.12) | 941 (714-1225) | 6.45 (4.88-8.41) | 876 (653-1149) | 6.01 (4.49-7.83) |
| Slovakia | Male | 11812 (11326-12283) | 2853 (2373-3284) | 24.20 (20.21-27.67) | 1635 (1307-1999) | 13.87 (11.13-16.86) | 1715 (1354-2069) | 14.52 (11.54-17.48) | 880 (678-1108) | 7.45 (5.76-9.38) | 820 (623-1047) | 6.93 (5.25-8.84) |
| Slovenia | Female | 3961 (3794-4112) | 779 (584-967) | 19.65 (14.85-24.40) | 389 (276-528) | 9.82 (7.00-13.26) | 494 (359-651) | 12.43 (9.05-16.47) | 238 (164-328) | 6.00 (4.16-8.27) | 215 (147-303) | 5.42 (3.75-7.71) |
| Slovenia | Male | 3054 (2977-3135) | 692 (576-796) | 22.57 (18.83-26.12) | 363 (282-451) | 11.87 (9.30-14.71) | 455 (365-557) | 14.89 (11.90-18.15) | 177 (132-232) | 5.78 (4.39-7.62) | 154 (111-210) | 5.01 (3.63-6.84) |
| Solomon Islands | Female | 646 (609-687) | 89 (66-110) | 13.83 (10.20-16.89) | 62 (46-78) | 9.64 (6.94-11.89) | 39 (28-50) | 6.08 (4.31-7.70) | 17 (12-23) | 2.60 (1.81-3.48) | 7 (4-10) | 1.06 (0.63-1.57) |
| Solomon Islands | Male | 916 (855-977) | 127 (87-162) | 13.85 (9.58-17.47) | 89 (60-116) | 9.71 (6.61-12.66) | 56 (38-74) | 6.17 (4.25-8.05) | 11 (7-15) | 1.16 (0.73-1.67) | 5 (3-8) | 0.59 (0.35-0.93) |
| Somalia | Female | 7143 (6398-7895) | 1337 (974-1696) | 18.62 (13.81-22.99) | 946 (668-1226) | 13.21 (9.65-16.54) | 569 (404-745) | 7.96 (5.69-10.15) | 269 (182-379) | 3.75 (2.61-5.24) | 194 (127-291) | 2.71 (1.75-4.01) |
| Somalia | Male | 6567 (5936-7209) | 869 (624-1122) | 13.22 (9.54-16.91) | 629 (442-819) | 9.61 (6.88-12.37) | 381 (264-510) | 5.79 (4.11-7.66) | 119 (75-176) | 1.81 (1.17-2.65) | 86 (52-131) | 1.31 (0.81-1.99) |
| South Africa | Female | 54390 (52846-55942) | 3631 (2328-5153) | 6.67 (4.29-9.41) | 2884 (1882-4014) | 5.32 (3.46-7.35) | 2189 (1480-2975) | 4.02 (2.71-5.51) | 1300 (796-1864) | 2.39 (1.44-3.43) | 1208 (730-1751) | 2.23 (1.33-3.22) |
| South Africa | Male | 41829 (40934-42773) | 2467 (1511-3580) | 5.94 (3.61-8.54) | 1949 (1206-2820) | 4.66 (2.89-6.74) | 1488 (943-2101) | 3.55 (2.24-4.99) | 540 (279-857) | 1.29 (0.67-2.05) | 494 (261-794) | 1.18 (0.62-1.90) |
| South Korea | Female | 41515 (38794-44113) | 7097 (5328-8847) | 17.06 (12.96-20.97) | 3247 (2315-4414) | 7.81 (5.63-10.49) | 4784 (3499-6276) | 11.58 (8.62-14.86) | 1854 (1307-2557) | 4.49 (3.16-6.11) | 1856 (1310-2562) | 4.47 (3.19-6.09) |
| South Korea | Male | 34603 (33097-36338) | 6104 (4479-7325) | 17.63 (13.16-21.01) | 2829 (2066-3608) | 8.19 (6.00-10.48) | 4152 (3016-5090) | 11.98 (8.68-14.61) | 1335 (958-1741) | 3.85 (2.78-5.00) | 1330 (963-1740) | 3.84 (2.79-5.01) |
| Spain | Female | 58949 (56194-61630) | 939 (278-2383) | 1.58 (0.47-4.02) | 752 (181-1758) | 1.27 (0.30-3.01) | 834 (217-2123) | 1.41 (0.36-3.60) | 385 (56-974) | 0.65 (0.10-1.65) | 345 (48-877) | 0.58 (0.08-1.48) |
| Spain | Male | 51448 (50237-52740) | 2013 (1350-2920) | 3.91 (2.65-5.67) | 1154 (705-1773) | 2.25 (1.38-3.44) | 1451 (929-2210) | 2.83 (1.81-4.28) | 430 (223-732) | 0.83 (0.44-1.43) | 390 (196-675) | 0.76 (0.38-1.31) |
| Sri Lanka | Female | 26053 (24180-27869) | 4404 (3540-5247) | 16.95 (13.68-20.05) | 2951 (2332-3620) | 11.33 (9.05-13.81) | 1946 (1531-2412) | 7.47 (5.90-9.19) | 1138 (859-1471) | 4.37 (3.31-5.60) | 993 (728-1297) | 3.82 (2.83-4.92) |
| Sri Lanka | Male | 27939 (25781-30064) | 5617 (4594-6742) | 20.11 (16.75-23.54) | 3784 (3038-4650) | 13.59 (11.13-16.37) | 2498 (1982-3208) | 8.95 (7.13-11.33) | 942 (708-1269) | 3.37 (2.58-4.47) | 778 (572-1064) | 2.80 (2.08-3.73) |
| Sudan | Female | 31097 (28896-33429) | 220 (68-548) | 0.71 (0.22-1.79) | 197 (62-508) | 0.63 (0.20-1.63) | 156 (44-384) | 0.50 (0.14-1.23) | 30 (3-121) | 0.10 (0.01-0.38) | 21 (1-100) | 0.07 (0.00-0.32) |
| Sudan | Male | 39347 (36515-42187) | 2555 (1668-3487) | 6.50 (4.29-8.83) | 2022 (1350-2772) | 5.16 (3.48-6.96) | 1287 (860-1739) | 3.27 (2.24-4.42) | 293 (161-463) | 0.74 (0.42-1.18) | 220 (116-357) | 0.56 (0.29-0.91) |
| Suriname | Female | 616 (579-651) | 58 (41-73) | 9.45 (6.82-11.79) | 41 (30-52) | 6.65 (4.82-8.38) | 28 (20-36) | 4.52 (3.23-5.78) | 21 (15-27) | 3.38 (2.41-4.40) | 18 (12-23) | 2.86 (1.99-3.83) |
| Suriname | Male | 689 (648-734) | 91 (70-111) | 13.21 (10.12-15.94) | 62 (46-78) | 9.01 (6.73-11.13) | 42 (32-54) | 6.10 (4.63-7.77) | 24 (17-31) | 3.42 (2.52-4.43) | 20 (14-26) | 2.86 (2.01-3.79) |
| Swaziland | Female | 813 (745-889) | 66 (42-93) | 8.24 (5.20-11.47) | 52 (33-71) | 6.39 (4.12-8.58) | 36 (23-48) | 4.39 (2.84-5.98) | 20 (12-29) | 2.48 (1.49-3.57) | 16 (9-23) | 1.93 (1.07-2.83) |
| Swaziland | Male | 739 (668-799) | 67 (43-94) | 9.18 (5.85-12.50) | 50 (32-71) | 6.83 (4.32-9.47) | 34 (23-48) | 4.60 (3.09-6.43) | 9 (5-14) | 1.23 (0.69-1.88) | 7 (4-11) | 0.93 (0.49-1.47) |
| Sweden | Female | 14871 (14221-15625) | 1064 (671-1466) | 7.18 (4.52-9.81) | 282 (155-455) | 1.90 (1.06-3.01) | 936 (603-1286) | 6.29 (4.05-8.54) | 132 (67-222) | 0.89 (0.45-1.48) | 109 (55-181) | 0.74 (0.38-1.21) |
| Sweden | Male | 13891 (13436-14348) | 1613 (1263-1963) | 11.62 (9.14-14.05) | 390 (253-578) | 2.79 (1.83-4.15) | 1352 (1043-1667) | 9.74 (7.53-12.00) | 175 (109-270) | 1.25 (0.78-1.95) | 156 (97-239) | 1.12 (0.70-1.72) |
| Switzerland | Female | 10503 (9970-11038) | 795 (482-1147) | 7.60 (4.56-10.95) | 77 (42-189) | 0.73 (0.40-1.82) | 765 (473-1105) | 7.24 (4.46-10.47) | 34 (18-83) | 0.32 (0.17-0.82) | 28 (15-73) | 0.27 (0.15-0.70) |
| Switzerland | Male | 9106 (8818-9377) | 1032 (801-1285) | 11.35 (8.79-14.00) | 93 (60-179) | 1.02 (0.65-1.97) | 975 (756-1222) | 10.74 (8.26-13.33) | 46 (29-95) | 0.50 (0.32-1.04) | 41 (25-84) | 0.45 (0.28-0.92) |
| Syria | Female | 20463 (19249-21671) | 77 (9-294) | 0.38 (0.04-1.41) | 74 (7-283) | 0.36 (0.04-1.36) | 63 (5-219) | 0.31 (0.03-1.09) | 21 (0-123) | 0.10 (0.00-0.60) | 18 (0-108) | 0.09 (0.00-0.53) |
| Syria | Male | 29596 (27634-31562) | 1711 (990-2450) | 5.78 (3.39-8.34) | 1362 (838-1908) | 4.60 (2.84-6.50) | 864 (558-1208) | 2.91 (1.90-4.08) | 435 (243-685) | 1.48 (0.82-2.30) | 369 (205-581) | 1.25 (0.68-1.98) |
| Taiwan (Province of China) | Female | 16122 (15562-16705) | 2748 (2110-3351) | 17.01 (13.10-20.79) | 1283 (946-1673) | 7.95 (5.90-10.31) | 1794 (1338-2282) | 11.10 (8.32-14.04) | 503 (358-703) | 3.13 (2.22-4.39) | 502 (357-692) | 3.12 (2.20-4.30) |
| Taiwan (Province of China) | Male | 22165 (21646-22703) | 3600 (2932-4266) | 16.24 (13.31-19.22) | 1683 (1320-2109) | 7.59 (6.00-9.45) | 2374 (1903-2865) | 10.74 (8.66-12.89) | 623 (483-813) | 2.80 (2.18-3.68) | 608 (473-788) | 2.74 (2.13-3.55) |
| Tajikistan | Female | 8436 (8037-8828) | 975 (752-1218) | 11.56 (8.93-14.19) | 710 (538-893) | 8.44 (6.45-10.54) | 432 (327-550) | 5.14 (3.89-6.49) | 295 (214-392) | 3.52 (2.57-4.59) | 256 (181-339) | 3.03 (2.17-4.05) |
| Tajikistan | Male | 8747 (8297-9188) | 1515 (1234-1816) | 17.35 (14.25-20.62) | 1066 (857-1302) | 12.16 (9.91-14.69) | 647 (515-805) | 7.39 (5.89-9.09) | 359 (279-461) | 4.10 (3.19-5.26) | 227 (164-312) | 2.60 (1.90-3.56) |
| Tanzania | Female | 24927 (22878-26903) | 4914 (3797-6025) | 19.66 (15.48-23.62) | 3405 (2579-4245) | 13.64 (10.59-16.73) | 2070 (1578-2681) | 8.33 (6.44-10.53) | 841 (596-1142) | 3.39 (2.45-4.46) | 366 (240-540) | 1.47 (0.98-2.12) |
| Tanzania | Male | 29451 (26966-31723) | 5081 (3940-6136) | 17.30 (13.40-20.48) | 3516 (2667-4299) | 11.99 (9.03-14.59) | 2152 (1604-2682) | 7.34 (5.50-9.02) | 531 (367-710) | 1.82 (1.27-2.42) | 333 (222-473) | 1.14 (0.75-1.62) |
| Thailand | Female | 64351 (60163-68606) | 9213 (7179-11260) | 14.35 (11.20-17.38) | 6043 (4683-7476) | 9.37 (7.21-11.40) | 4334 (3252-5436) | 6.75 (5.07-8.35) | 2312 (1744-2898) | 3.58 (2.73-4.49) | 2090 (1559-2681) | 3.24 (2.39-4.12) |
| Thailand | Male | 70565 (66090-74797) | 13047 (10236-15509) | 18.46 (14.64-21.63) | 8607 (6693-10500) | 12.24 (9.58-14.75) | 6179 (4755-7556) | 8.76 (6.74-10.57) | 2394 (1821-3104) | 3.39 (2.56-4.36) | 2092 (1588-2788) | 2.97 (2.27-3.95) |
| The Bahamas | Female | 375 (362-387) | 42 (30-55) | 11.33 (8.06-14.63) | 23 (16-30) | 6.07 (4.37-7.93) | 28 (20-36) | 7.48 (5.43-9.73) | 11 (7-15) | 2.83 (1.95-3.81) | 9 (6-13) | 2.48 (1.69-3.43) |
| The Bahamas | Male | 404 (389-421) | 65 (50-78) | 16.10 (12.51-19.24) | 34 (25-43) | 8.32 (6.34-10.54) | 41 (32-52) | 10.22 (7.92-12.75) | 13 (9-17) | 3.21 (2.31-4.24) | 11 (8-15) | 2.64 (1.86-3.61) |
| The Gambia | Female | 1516 (1406-1625) | 167 (124-211) | 11.02 (8.25-13.71) | 120 (91-153) | 7.96 (5.99-10.00) | 73 (55-94) | 4.79 (3.68-6.15) | 40 (28-54) | 2.63 (1.86-3.53) | 26 (17-38) | 1.70 (1.14-2.44) |
| The Gambia | Male | 1539 (1446-1635) | 177 (135-220) | 11.55 (8.71-14.10) | 127 (96-163) | 8.27 (6.23-10.41) | 77 (56-99) | 4.99 (3.69-6.33) | 27 (19-39) | 1.78 (1.23-2.54) | 19 (13-28) | 1.24 (0.80-1.78) |
| Timor-Leste | Female | 1276 (1191-1362) | 234 (186-283) | 18.28 (14.65-21.96) | 158 (124-196) | 12.46 (9.85-15.24) | 102 (78-129) | 8.00 (6.17-10.01) | 32 (23-44) | 2.50 (1.83-3.45) | 22 (15-32) | 1.70 (1.20-2.47) |
| Timor-Leste | Male | 1441 (1338-1553) | 309 (249-367) | 21.44 (17.76-24.93) | 211 (168-256) | 14.63 (11.87-17.55) | 135 (106-170) | 9.35 (7.51-11.68) | 42 (31-57) | 2.94 (2.21-3.90) | 28 (19-38) | 1.92 (1.38-2.62) |
| Togo | Female | 5172 (4795-5562) | 582 (427-733) | 11.26 (8.47-14.14) | 419 (310-538) | 8.09 (6.00-10.30) | 255 (184-335) | 4.93 (3.59-6.48) | 116 (78-168) | 2.24 (1.54-3.18) | 64 (41-97) | 1.24 (0.80-1.86) |
| Togo | Male | 4801 (4462-5145) | 570 (422-703) | 11.87 (8.96-14.36) | 406 (299-514) | 8.49 (6.32-10.54) | 246 (179-321) | 5.15 (3.76-6.58) | 67 (44-96) | 1.41 (0.93-1.99) | 32 (21-49) | 0.67 (0.43-1.04) |
| Tonga | Female | 77 (72-82) | 12 (9-14) | 14.95 (12.17-17.69) | 8 (6-10) | 10.06 (8.05-12.12) | 5 (4-6) | 6.68 (5.27-8.24) | 3 (2-4) | 3.74 (2.86-4.86) | 2 (2-3) | 3.16 (2.28-4.13) |
| Tonga | Male | 107 (100-114) | 18 (15-21) | 16.99 (13.91-19.85) | 12 (10-15) | 11.40 (9.23-13.69) | 8 (6-10) | 7.58 (6.04-9.26) | 3 (2-3) | 2.50 (1.88-3.28) | 2 (2-3) | 2.05 (1.52-2.76) |
| Trinidad and Tobago | Female | 1793 (1718-1875) | 155 (111-201) | 8.65 (6.18-11.13) | 98 (74-128) | 5.46 (4.11-7.10) | 89 (66-114) | 4.96 (3.69-6.35) | 48 (33-64) | 2.65 (1.84-3.54) | 39 (27-54) | 2.17 (1.53-2.98) |
| Trinidad and Tobago | Male | 2113 (2001-2221) | 298 (246-348) | 14.16 (11.76-16.42) | 180 (146-218) | 8.52 (6.92-10.36) | 162 (127-197) | 7.67 (6.15-9.29) | 70 (54-90) | 3.34 (2.58-4.29) | 60 (45-78) | 2.85 (2.15-3.68) |
| Tunisia | Female | 15070 (13580-16365) | 36 (5-160) | 0.24 (0.04-1.07) | 34 (4-151) | 0.23 (0.03-0.98) | 31 (3-125) | 0.21 (0.02-0.84) | 9 (0-63) | 0.06 (0.00-0.42) | 8 (0-54) | 0.05 (0.00-0.36) |
| Tunisia | Male | 18891 (17403-20352) | 926 (517-1379) | 4.92 (2.66-7.32) | 729 (426-1084) | 3.87 (2.20-5.70) | 512 (296-757) | 2.72 (1.60-3.97) | 194 (87-316) | 1.02 (0.46-1.67) | 168 (76-279) | 0.88 (0.40-1.49) |
| Turkey | Female | 99014 (93629-104751) | 0 (0-75) | 0.00 (-0.00-0.07) | 0 (0-73) | 0.00 (-0.00-0.07) | 0 (0-73) | 0.00 (-0.00-0.07) | 0 (0-29) | 0.00 (0.00-0.03) | 0 (0-29) | 0.00 (0.00-0.03) |
| Turkey | Male | 101146 (95573-106837) | 2545 (1087-4690) | 2.51 (1.08-4.63) | 2181 (968-3813) | 2.15 (0.96-3.79) | 1849 (775-3209) | 1.83 (0.77-3.14) | 634 (194-1251) | 0.62 (0.20-1.21) | 618 (193-1236) | 0.61 (0.19-1.20) |
| Turkmenistan | Female | 8904 (8487-9326) | 984 (770-1244) | 11.06 (8.66-13.82) | 667 (516-839) | 7.47 (5.88-9.42) | 491 (375-615) | 5.53 (4.21-6.93) | 339 (257-437) | 3.81 (2.89-4.86) | 299 (225-392) | 3.36 (2.53-4.37) |
| Turkmenistan | Male | 9616 (9153-10048) | 1635 (1353-1913) | 17.08 (14.19-19.80) | 1068 (868-1271) | 11.12 (9.05-13.14) | 773 (619-947) | 8.08 (6.52-9.86) | 364 (286-450) | 3.78 (2.95-4.66) | 266 (203-340) | 2.78 (2.13-3.49) |
| Tuvalu | Female | 21 (19-22) | 3 (3-4) | 15.81 (12.89-18.72) | 2 (2-3) | 10.62 (8.43-12.64) | 2 (1-2) | 7.27 (5.76-8.94) | 1 (0-1) | 2.84 (1.98-3.82) | 0 (0-0) | 1.67 (1.12-2.37) |
| Tuvalu | Male | 24 (23-26) | 4 (3-5) | 17.07 (13.39-19.79) | 3 (2-3) | 11.50 (8.94-13.80) | 2 (1-2) | 7.91 (6.21-9.57) | 1 (0-1) | 2.46 (1.80-3.30) | 0 (0-1) | 1.70 (1.15-2.46) |
| Uganda | Female | 12158 (11088-13274) | 2152 (1649-2683) | 17.73 (14.18-21.49) | 1519 (1154-1932) | 12.50 (9.75-15.39) | 920 (682-1174) | 7.56 (5.79-9.59) | 310 (214-423) | 2.53 (1.81-3.46) | 189 (126-276) | 1.55 (1.05-2.26) |
| Uganda | Male | 13343 (12417-14387) | 1725 (1331-2157) | 12.97 (10.03-15.87) | 1237 (940-1575) | 9.25 (7.15-11.50) | 753 (567-962) | 5.64 (4.28-7.12) | 165 (113-230) | 1.24 (0.84-1.71) | 96 (62-144) | 0.72 (0.47-1.06) |
| Ukraine | Female | 232729 (210324-255122) | 8125 (3605-14507) | 3.50 (1.55-6.20) | 7133 (3249-12036) | 3.07 (1.42-5.12) | 5439 (2623-8599) | 2.37 (1.11-3.67) | 4105 (1804-7048) | 1.76 (0.75-2.99) | 3450 (1527-5862) | 1.50 (0.65-2.50) |
| Ukraine | Male | 167558 (153353-182018) | 18329 (13778-23001) | 10.89 (8.39-13.61) | 12509 (9410-16091) | 7.47 (5.76-9.35) | 8368 (6335-10928) | 5.01 (3.90-6.27) | 4631 (3261-6171) | 2.76 (1.96-3.64) | 3190 (2241-4400) | 1.91 (1.37-2.57) |
| United Arab Emirates | Female | 1592 (1485-1699) | 8 (1-39) | 0.48 (0.06-2.46) | 5 (1-29) | 0.34 (0.04-1.79) | 7 (1-37) | 0.46 (0.05-2.35) | 1 (0-11) | 0.08 (0.00-0.66) | 1 (0-10) | 0.07 (0.00-0.61) |
| United Arab Emirates | Male | 3346 (3135-3570) | 265 (156-369) | 7.94 (4.64-11.11) | 118 (71-163) | 3.55 (2.13-4.91) | 214 (130-297) | 6.40 (3.89-8.86) | 25 (14-36) | 0.74 (0.41-1.07) | 22 (13-32) | 0.66 (0.37-0.97) |
| United Kingdom | Female | 73906 (71628-76188) | 4420 (2813-6104) | 6.00 (3.81-8.31) | 1790 (1146-2559) | 2.42 (1.56-3.45) | 3571 (2312-4929) | 4.83 (3.13-6.71) | 767 (481-1110) | 1.04 (0.65-1.49) | 701 (443-1002) | 0.95 (0.60-1.35) |
| United Kingdom | Male | 78971 (77762-80173) | 7656 (5713-9419) | 9.70 (7.28-11.93) | 2778 (1953-3757) | 3.52 (2.47-4.74) | 5781 (4253-7280) | 7.33 (5.42-9.26) | 1154 (778-1593) | 1.46 (0.99-2.02) | 1053 (718-1457) | 1.34 (0.91-1.84) |
| United States | Female | 422555 (406917-438324) | 34174 (24690-45114) | 8.10 (5.88-10.58) | 6804 (4070-11280) | 1.62 (0.96-2.66) | 31047 (22273-40979) | 7.34 (5.33-9.63) | 3537 (2013-5966) | 0.83 (0.48-1.41) | 3303 (1835-5555) | 0.78 (0.43-1.32) |
| United States | Male | 450375 (441797-458681) | 56668 (44297-67542) | 12.59 (9.76-14.95) | 10910 (6593-16018) | 2.42 (1.46-3.56) | 50350 (38367-60917) | 11.17 (8.51-13.48) | 4933 (2804-7274) | 1.09 (0.62-1.62) | 4620 (2647-6915) | 1.02 (0.58-1.53) |
| Uruguay | Female | 5136 (4979-5308) | 676 (542-804) | 13.13 (10.47-15.65) | 396 (302-491) | 7.73 (5.87-9.56) | 375 (281-467) | 7.29 (5.51-9.13) | 236 (176-299) | 4.60 (3.44-5.85) | 227 (171-289) | 4.42 (3.32-5.67) |
| Uruguay | Male | 4397 (4297-4494) | 684 (555-812) | 15.55 (12.64-18.32) | 395 (316-476) | 8.98 (7.18-10.85) | 375 (297-459) | 8.53 (6.78-10.49) | 198 (153-247) | 4.49 (3.47-5.62) | 175 (135-219) | 3.98 (3.07-4.98) |
| Uzbekistan | Female | 46061 (44571-47657) | 5145 (3954-6388) | 11.16 (8.65-13.90) | 3692 (2819-4603) | 8.02 (6.11-9.89) | 2311 (1762-2954) | 5.03 (3.84-6.42) | 1635 (1197-2112) | 3.56 (2.60-4.61) | 1400 (1012-1865) | 3.04 (2.19-4.02) |
| Uzbekistan | Male | 49898 (48150-51935) | 8295 (6689-9764) | 16.60 (13.35-19.52) | 5724 (4519-6909) | 11.46 (9.10-13.89) | 3570 (2741-4390) | 7.17 (5.54-8.79) | 1797 (1360-2294) | 3.60 (2.69-4.58) | 1419 (1046-1850) | 2.83 (2.11-3.67) |
| Vanuatu | Female | 297 (279-315) | 46 (38-55) | 15.59 (12.75-18.16) | 32 (25-38) | 10.70 (8.62-12.76) | 20 (16-25) | 6.91 (5.41-8.44) | 8 (6-11) | 2.73 (1.99-3.68) | 4 (2-6) | 1.30 (0.82-1.94) |
| Vanuatu | Male | 495 (468-522) | 81 (66-98) | 16.41 (13.44-19.64) | 56 (45-70) | 11.28 (9.03-13.90) | 36 (28-46) | 7.30 (5.76-9.23) | 8 (6-12) | 1.69 (1.18-2.38) | 4 (3-7) | 0.87 (0.56-1.30) |
| Venezuela | Female | 29000 (27429-30396) | 4026 (3283-4785) | 13.93 (11.40-16.44) | 2822 (2240-3461) | 9.73 (7.80-12.01) | 1766 (1364-2169) | 6.07 (4.72-7.48) | 1597 (1262-2018) | 5.48 (4.37-6.94) | 1507 (1167-1911) | 5.20 (4.06-6.63) |
| Venezuela | Male | 35825 (33822-38016) | 6722 (5444-7990) | 18.73 (15.24-22.02) | 4580 (3632-5626) | 12.74 (10.22-15.28) | 2880 (2230-3605) | 8.00 (6.30-9.91) | 1981 (1552-2490) | 5.56 (4.37-6.81) | 1835 (1433-2306) | 5.13 (4.03-6.37) |
| Vietnam | Female | 113845 (106800-121459) | 19644 (15885-23126) | 17.27 (14.03-20.18) | 13226 (10439-16101) | 11.60 (9.23-13.98) | 8703 (6774-10680) | 7.63 (6.06-9.24) | 4278 (3246-5562) | 3.74 (2.88-4.80) | 2979 (2151-3915) | 2.61 (1.90-3.44) |
| Vietnam | Male | 144924 (136380-153676) | 31271 (26000-37152) | 21.65 (18.09-25.13) | 21180 (17200-25606) | 14.63 (11.97-17.42) | 13836 (10960-17625) | 9.55 (7.74-11.91) | 5705 (4392-7402) | 3.93 (3.07-5.03) | 4062 (3059-5454) | 2.82 (2.15-3.69) |
| Yemen | Female | 26821 (24691-29087) | 121 (16-409) | 0.45 (0.06-1.56) | 112 (15-396) | 0.42 (0.06-1.46) | 94 (12-307) | 0.35 (0.04-1.14) | 17 (1-89) | 0.06 (0.00-0.34) | 13 (0-76) | 0.05 (0.00-0.28) |
| Yemen | Male | 29761 (27533-31928) | 1800 (1107-2555) | 6.06 (3.73-8.56) | 1436 (905-1987) | 4.83 (3.09-6.66) | 910 (584-1255) | 3.05 (2.01-4.21) | 274 (144-435) | 0.92 (0.50-1.45) | 237 (119-387) | 0.79 (0.42-1.28) |
| Zambia | Female | 8744 (8031-9396) | 1569 (1184-1922) | 17.97 (13.89-21.51) | 1103 (819-1380) | 12.63 (9.58-15.35) | 680 (510-869) | 7.76 (5.92-9.63) | 349 (249-473) | 4.00 (2.89-5.32) | 214 (147-299) | 2.46 (1.71-3.37) |
| Zambia | Male | 9811 (9199-10509) | 1288 (982-1601) | 13.15 (9.92-16.12) | 928 (676-1159) | 9.47 (6.94-11.83) | 568 (418-732) | 5.78 (4.31-7.37) | 176 (126-238) | 1.81 (1.29-2.41) | 107 (69-153) | 1.09 (0.71-1.55) |
| Zimbabwe | Female | 10735 (10049-11482) | 1257 (936-1587) | 11.73 (8.65-14.61) | 900 (679-1150) | 8.38 (6.21-10.69) | 561 (415-737) | 5.21 (3.88-6.83) | 316 (214-431) | 2.93 (2.00-4.03) | 259 (164-369) | 2.42 (1.55-3.41) |
| Zimbabwe | Male | 8188 (7700-8597) | 1062 (808-1297) | 13.04 (9.86-15.82) | 751 (561-948) | 9.21 (6.89-11.52) | 461 (343-601) | 5.64 (4.23-7.39) | 163 (108-232) | 1.99 (1.33-2.81) | 124 (81-188) | 1.51 (0.99-2.30) |

#### c. **Prevented CVD new cases**

| **Country** | **Sex** | **Baseline disease burden Median (95% UI)** | **Replacement of discretionary salt and non-discretionary salt** | | **Replacement of discretionary salt** | | **Replacement of non-discretionary salt** | | **Replacement of discretionary salt among those with diagnosed hypertension** | | **Replacement of discretionary salt among those with treated hypertension** | |
| --- | --- | --- | --- | --- | --- | --- | --- | --- | --- | --- | --- | --- |
|  |  |  | **No. of events averted Median (95% UI)** | **Percentage of national disease burden averted % (95% UI)** | **No. of events averted Median (95% UI)** | **Percentage of national disease burden averted % (95% UI)** | **No. of events averted Median (95% UI)** | **Percentage of national disease burden averted % (95% UI)** | **No. of events averted Median (95% UI)** | **Percentage of national disease burden averted % (95% UI)** | **No. of events averted Median (95% UI)** | **Percentage of national disease burden averted % (95% UI)** |
| **Global, millions** | **Female** | 28.30 (27.55-29.06) | 3.24 (2.93-3.53) | 11.45 (10.41-12.37) | 1.99 (1.78-2.21) | 7.05 (6.33-7.78) | 1.68 (1.50-1.87) | 5.94 (5.33-6.56) | 0.61 (0.55-0.68) | 2.15 (1.94-2.40) | 0.50 (0.45-0.56) | 1.77 (1.58-1.98) |
| **Global, millions** | **Male** | 30.22 (29.41-31.14) | 4.76 (4.38-5.12) | 15.74 (14.52-16.77) | 2.96 (2.67-3.24) | 9.81 (8.90-10.64) | 2.54 (2.30-2.80) | 8.42 (7.63-9.20) | 0.73 (0.66-0.81) | 2.41 (2.19-2.68) | 0.58 (0.53-0.65) | 1.93 (1.74-2.14) |
| Afghanistan | Female | 54243 (49792-58309) | 346 (60-846) | 0.64 (0.11-1.57) | 309 (57-782) | 0.57 (0.10-1.43) | 240 (41-618) | 0.45 (0.07-1.13) | 70 (8-233) | 0.13 (0.02-0.42) | 58 (7-200) | 0.11 (0.01-0.38) |
| Afghanistan | Male | 70511 (63817-76771) | 4775 (2703-7073) | 6.80 (3.85-9.94) | 3716 (2159-5452) | 5.28 (3.07-7.57) | 2291 (1331-3397) | 3.25 (1.89-4.76) | 615 (350-936) | 0.87 (0.51-1.32) | 491 (266-758) | 0.70 (0.39-1.06) |
| Albania | Female | 12877 (11997-13671) | 2246 (1837-2627) | 17.48 (14.56-20.08) | 1450 (1175-1743) | 11.30 (9.20-13.43) | 1017 (806-1248) | 7.88 (6.37-9.54) | 648 (507-832) | 5.02 (3.98-6.37) | 570 (434-739) | 4.45 (3.43-5.64) |
| Albania | Male | 13982 (12900-15073) | 3214 (2673-3771) | 22.96 (19.48-26.25) | 2160 (1737-2628) | 15.47 (12.50-18.38) | 1529 (1216-1916) | 10.86 (8.81-13.35) | 495 (362-649) | 3.54 (2.64-4.61) | 365 (253-495) | 2.61 (1.84-3.45) |
| Algeria | Female | 166207 (152222-180472) | 809 (97-2387) | 0.49 (0.06-1.47) | 743 (86-2214) | 0.44 (0.05-1.33) | 630 (63-1847) | 0.38 (0.04-1.11) | 172 (3-851) | 0.10 (0.00-0.50) | 142 (1-734) | 0.08 (0.00-0.44) |
| Algeria | Male | 230839 (212791-250735) | 14250 (8278-20147) | 6.14 (3.62-8.57) | 10878 (6637-15121) | 4.71 (2.88-6.44) | 7315 (4666-10236) | 3.20 (2.03-4.38) | 2366 (1291-3562) | 1.03 (0.57-1.51) | 1909 (1015-2931) | 0.83 (0.44-1.25) |
| American Samoa | Female | 159 (146-172) | 12 (8-15) | 7.43 (5.33-9.53) | 8 (5-10) | 4.72 (3.41-6.05) | 7 (5-9) | 4.35 (3.16-5.59) | 3 (2-4) | 1.63 (1.11-2.20) | 2 (1-3) | 1.27 (0.81-1.77) |
| American Samoa | Male | 182 (165-201) | 16 (12-21) | 8.88 (6.48-11.32) | 10 (7-13) | 5.57 (4.05-7.10) | 9 (7-12) | 5.12 (3.72-6.55) | 3 (2-4) | 1.73 (1.16-2.36) | 2 (1-3) | 1.28 (0.83-1.84) |
| Andorra | Female | 292 (272-311) | 10 (6-14) | 3.28 (1.96-4.75) | 5 (3-7) | 1.76 (1.06-2.46) | 8 (5-12) | 2.78 (1.66-3.95) | 2 (1-3) | 0.63 (0.33-1.00) | 2 (1-3) | 0.57 (0.31-0.91) |
| Andorra | Male | 405 (376-433) | 40 (31-50) | 10.00 (7.70-12.09) | 16 (12-21) | 3.98 (2.95-5.14) | 29 (22-37) | 7.26 (5.38-8.97) | 6 (4-8) | 1.43 (0.99-2.00) | 5 (4-7) | 1.30 (0.90-1.78) |
| Angola | Female | 38394 (35549-41329) | 2657 (1778-3632) | 6.92 (4.71-9.32) | 2039 (1368-2755) | 5.29 (3.56-7.17) | 1304 (874-1777) | 3.41 (2.33-4.60) | 546 (351-792) | 1.43 (0.91-2.05) | 259 (144-405) | 0.68 (0.38-1.04) |
| Angola | Male | 34433 (31560-36863) | 2864 (1991-3810) | 8.33 (5.88-10.98) | 2134 (1477-2881) | 6.23 (4.37-8.21) | 1356 (966-1825) | 3.96 (2.83-5.23) | 376 (237-561) | 1.10 (0.69-1.62) | 225 (127-353) | 0.65 (0.38-1.01) |
| Antigua and Barbuda | Female | 328 (299-358) | 25 (18-33) | 7.64 (5.59-9.80) | 16 (12-20) | 4.74 (3.53-6.05) | 14 (11-19) | 4.41 (3.32-5.67) | 7 (5-9) | 2.12 (1.51-2.78) | 6 (4-8) | 1.79 (1.26-2.46) |
| Antigua and Barbuda | Male | 332 (300-368) | 41 (31-50) | 12.25 (9.50-14.53) | 24 (18-30) | 7.17 (5.48-8.95) | 22 (16-28) | 6.67 (5.03-8.28) | 8 (5-10) | 2.31 (1.65-3.07) | 6 (4-9) | 1.85 (1.31-2.53) |
| Argentina | Female | 139243 (131488-147188) | 14308 (11546-16862) | 10.32 (8.35-11.92) | 8951 (7157-10853) | 6.42 (5.14-7.70) | 7004 (5437-8613) | 5.05 (3.94-6.14) | 4027 (3160-4977) | 2.88 (2.28-3.54) | 3492 (2715-4338) | 2.50 (1.97-3.10) |
| Argentina | Male | 152001 (141811-162930) | 22193 (17568-27038) | 14.64 (11.75-17.51) | 13938 (10950-17330) | 9.17 (7.25-11.28) | 10957 (8551-13863) | 7.22 (5.65-9.00) | 5544 (4321-7089) | 3.66 (2.85-4.58) | 4520 (3516-5758) | 2.97 (2.32-3.78) |
| Armenia | Female | 19110 (17383-20912) | 1892 (1428-2416) | 9.89 (7.48-12.50) | 1307 (992-1699) | 6.85 (5.23-8.77) | 885 (666-1161) | 4.63 (3.52-6.00) | 497 (346-699) | 2.61 (1.85-3.57) | 426 (303-609) | 2.23 (1.59-3.12) |
| Armenia | Male | 17712 (16107-19322) | 2824 (2274-3391) | 15.97 (12.91-18.66) | 1869 (1484-2278) | 10.55 (8.45-12.61) | 1257 (968-1589) | 7.09 (5.55-8.70) | 510 (376-679) | 2.90 (2.12-3.78) | 341 (250-458) | 1.93 (1.45-2.58) |
| Australia | Female | 107946 (100524-115250) | 2999 (1687-4827) | 2.78 (1.58-4.41) | 1133 (583-1851) | 1.05 (0.55-1.69) | 2826 (1559-4485) | 2.61 (1.45-4.09) | 373 (159-636) | 0.35 (0.15-0.59) | 328 (137-568) | 0.31 (0.13-0.52) |
| Australia | Male | 109000 (101185-116634) | 7408 (5042-9969) | 6.81 (4.69-9.08) | 2123 (1364-3088) | 1.95 (1.25-2.81) | 6499 (4400-8608) | 5.97 (4.12-7.77) | 639 (384-994) | 0.59 (0.35-0.89) | 569 (331-869) | 0.52 (0.30-0.79) |
| Austria | Female | 53256 (50688-55970) | 4885 (3946-5759) | 9.17 (7.48-10.79) | 1457 (998-1994) | 2.74 (1.90-3.69) | 3857 (3083-4710) | 7.23 (5.75-8.78) | 627 (405-888) | 1.17 (0.75-1.65) | 553 (357-812) | 1.04 (0.67-1.51) |
| Austria | Male | 56785 (54270-59400) | 7874 (6332-9308) | 13.84 (11.24-16.16) | 2326 (1715-3077) | 4.08 (3.02-5.38) | 6161 (4902-7452) | 10.86 (8.64-12.90) | 930 (660-1327) | 1.64 (1.14-2.33) | 898 (621-1262) | 1.58 (1.11-2.22) |
| Azerbaijan | Female | 50501 (47454-53694) | 5299 (4031-6508) | 10.49 (8.04-12.90) | 3651 (2776-4553) | 7.21 (5.52-8.96) | 2490 (1871-3162) | 4.92 (3.77-6.26) | 1565 (1132-2034) | 3.11 (2.29-4.04) | 1315 (940-1766) | 2.60 (1.89-3.47) |
| Azerbaijan | Male | 51784 (48488-55655) | 8506 (6794-10176) | 16.43 (13.31-19.38) | 5634 (4361-6885) | 10.82 (8.53-13.14) | 3813 (3001-4787) | 7.36 (5.89-9.14) | 1620 (1194-2112) | 3.13 (2.36-4.02) | 1162 (845-1540) | 2.24 (1.66-2.92) |
| Bahrain | Female | 3714 (3243-4163) | 21 (4-62) | 0.57 (0.10-1.66) | 18 (3-53) | 0.47 (0.08-1.40) | 19 (3-58) | 0.50 (0.09-1.52) | 4 (0-18) | 0.11 (0.00-0.49) | 3 (0-15) | 0.09 (0.00-0.42) |
| Bahrain | Male | 8591 (7365-9784) | 621 (366-938) | 7.26 (4.46-10.45) | 365 (224-531) | 4.31 (2.68-5.94) | 428 (265-642) | 5.05 (3.19-7.14) | 81 (46-131) | 0.95 (0.54-1.48) | 68 (36-111) | 0.80 (0.42-1.27) |
| Bangladesh | Female | 424029 (395527-452599) | 54067 (43125-64233) | 12.79 (10.36-14.92) | 37471 (29198-46138) | 8.82 (6.93-10.68) | 23498 (18144-29324) | 5.55 (4.33-6.83) | 8812 (6680-11132) | 2.09 (1.60-2.60) | 7038 (5315-8910) | 1.66 (1.26-2.08) |
| Bangladesh | Male | 604124 (560251-644816) | 82982 (65031-99494) | 13.77 (10.89-15.99) | 58040 (45012-70413) | 9.58 (7.54-11.49) | 36245 (27425-44751) | 6.00 (4.58-7.34) | 8723 (6515-11055) | 1.44 (1.08-1.82) | 6754 (5002-8809) | 1.12 (0.84-1.43) |
| Barbados | Female | 1690 (1531-1845) | 82 (49-122) | 4.87 (2.91-7.12) | 60 (39-85) | 3.58 (2.29-4.91) | 57 (36-81) | 3.38 (2.16-4.72) | 31 (18-46) | 1.86 (1.12-2.63) | 29 (17-41) | 1.71 (1.03-2.39) |
| Barbados | Male | 1567 (1431-1714) | 152 (109-194) | 9.73 (7.11-12.04) | 89 (65-118) | 5.74 (4.20-7.27) | 84 (61-111) | 5.35 (3.94-6.90) | 35 (25-48) | 2.25 (1.61-2.98) | 31 (22-44) | 2.00 (1.40-2.73) |
| Belarus | Female | 82785 (78457-87514) | 2983 (1631-4654) | 3.62 (2.00-5.56) | 2560 (1432-3828) | 3.09 (1.73-4.61) | 2044 (1177-2979) | 2.47 (1.47-3.60) | 1461 (752-2258) | 1.76 (0.91-2.73) | 1175 (599-1886) | 1.42 (0.73-2.27) |
| Belarus | Male | 67101 (62646-71129) | 8072 (6318-9731) | 12.06 (9.51-14.47) | 5319 (4175-6632) | 7.94 (6.21-9.79) | 3806 (2944-4820) | 5.67 (4.43-7.20) | 2257 (1671-2924) | 3.38 (2.50-4.32) | 1546 (1110-2083) | 2.31 (1.68-3.06) |
| Belgium | Female | 53959 (50615-56996) | 4415 (3536-5287) | 8.20 (6.59-9.81) | 1389 (991-1838) | 2.58 (1.85-3.44) | 3456 (2692-4259) | 6.39 (5.05-7.82) | 615 (421-841) | 1.15 (0.80-1.56) | 585 (399-803) | 1.09 (0.75-1.50) |
| Belgium | Male | 59373 (55610-63178) | 7655 (6251-9253) | 12.86 (10.69-15.34) | 2399 (1771-3223) | 4.05 (3.00-5.31) | 5927 (4765-7230) | 9.98 (8.02-12.12) | 919 (652-1291) | 1.55 (1.11-2.15) | 902 (629-1256) | 1.52 (1.08-2.11) |
| Belize | Female | 823 (745-903) | 60 (42-78) | 7.20 (5.15-9.42) | 42 (30-55) | 5.09 (3.69-6.66) | 29 (21-39) | 3.56 (2.59-4.70) | 16 (11-22) | 1.95 (1.36-2.61) | 13 (9-18) | 1.59 (1.07-2.17) |
| Belize | Male | 1000 (915-1095) | 119 (91-149) | 11.97 (9.22-14.47) | 80 (60-101) | 8.00 (6.06-9.84) | 55 (41-72) | 5.53 (4.14-7.01) | 21 (15-28) | 2.08 (1.48-2.69) | 16 (11-23) | 1.65 (1.15-2.22) |
| Benin | Female | 16277 (14969-17569) | 2037 (1619-2410) | 12.47 (10.10-14.46) | 1424 (1085-1722) | 8.74 (6.74-10.32) | 857 (648-1065) | 5.28 (4.09-6.52) | 305 (220-386) | 1.88 (1.37-2.36) | 208 (145-272) | 1.29 (0.91-1.65) |
| Benin | Male | 15748 (14384-17160) | 1647 (1230-2070) | 10.46 (7.79-12.77) | 1170 (873-1489) | 7.45 (5.51-9.30) | 716 (518-936) | 4.56 (3.32-5.87) | 133 (91-182) | 0.85 (0.59-1.13) | 95 (64-132) | 0.60 (0.42-0.81) |
| Bermuda | Female | 422 (379-467) | 32 (23-42) | 7.73 (5.65-9.67) | 2 (2-4) | 0.55 (0.38-0.93) | 31 (23-41) | 7.46 (5.46-9.32) | 1 (1-2) | 0.26 (0.17-0.46) | 1 (1-2) | 0.22 (0.15-0.40) |
| Bermuda | Male | 387 (352-421) | 45 (35-55) | 11.73 (9.24-13.88) | 3 (2-5) | 0.79 (0.58-1.34) | 43 (34-53) | 11.24 (8.77-13.38) | 1 (1-2) | 0.27 (0.19-0.48) | 1 (1-2) | 0.23 (0.15-0.42) |
| Bhutan | Female | 2002 (1783-2215) | 246 (186-300) | 12.32 (9.53-14.66) | 168 (125-214) | 8.45 (6.33-10.31) | 109 (80-141) | 5.44 (4.07-6.87) | 49 (35-65) | 2.46 (1.80-3.15) | 37 (26-50) | 1.84 (1.33-2.41) |
| Bhutan | Male | 2903 (2552-3215) | 403 (308-493) | 14.00 (11.01-16.48) | 276 (208-346) | 9.58 (7.39-11.73) | 178 (130-229) | 6.19 (4.64-7.72) | 62 (45-81) | 2.14 (1.60-2.73) | 40 (29-53) | 1.37 (0.99-1.81) |
| Bolivia | Female | 19642 (18220-21122) | 1871 (1316-2365) | 9.46 (6.78-11.81) | 1278 (893-1653) | 6.48 (4.64-8.30) | 819 (562-1074) | 4.16 (2.90-5.42) | 435 (298-583) | 2.21 (1.51-2.96) | 383 (256-543) | 1.95 (1.32-2.68) |
| Bolivia | Male | 22044 (20214-23986) | 2963 (2139-3765) | 13.39 (9.88-16.52) | 2022 (1458-2603) | 9.14 (6.64-11.50) | 1302 (922-1690) | 5.90 (4.16-7.53) | 481 (323-664) | 2.19 (1.49-2.94) | 389 (261-561) | 1.76 (1.16-2.49) |
| Bosnia and Herzegovina | Female | 21367 (19855-22860) | 3672 (3031-4321) | 17.24 (14.45-19.84) | 2348 (1888-2820) | 10.99 (8.94-12.94) | 1677 (1356-2082) | 7.86 (6.37-9.59) | 1245 (969-1552) | 5.83 (4.57-7.15) | 1149 (885-1437) | 5.36 (4.20-6.61) |
| Bosnia and Herzegovina | Male | 19828 (18210-21494) | 4490 (3764-5224) | 22.71 (19.19-25.68) | 2973 (2412-3598) | 15.02 (12.43-17.92) | 2148 (1723-2669) | 10.85 (8.74-13.13) | 1156 (897-1459) | 5.85 (4.57-7.35) | 1060 (818-1378) | 5.37 (4.16-6.92) |
| Botswana | Female | 5581 (5144-6023) | 415 (293-547) | 7.43 (5.17-9.67) | 298 (214-395) | 5.35 (3.85-6.99) | 218 (157-287) | 3.93 (2.79-5.09) | 103 (70-148) | 1.86 (1.25-2.58) | 84 (56-122) | 1.52 (1.01-2.15) |
| Botswana | Male | 4805 (4338-5236) | 406 (281-535) | 8.51 (5.96-10.95) | 288 (201-381) | 6.00 (4.28-7.75) | 210 (148-288) | 4.41 (3.15-5.84) | 63 (39-93) | 1.32 (0.83-1.87) | 43 (25-66) | 0.91 (0.54-1.35) |
| Brazil | Female | 504422 (469164-534721) | 51233 (40471-61695) | 10.21 (8.18-12.10) | 33557 (26141-41676) | 6.65 (5.21-8.15) | 24281 (18640-30201) | 4.80 (3.72-5.97) | 16405 (12469-20656) | 3.25 (2.51-4.08) | 15866 (12207-19951) | 3.15 (2.45-3.92) |
| Brazil | Male | 565725 (524886-606361) | 85146 (68097-101330) | 15.01 (12.24-17.76) | 55276 (43897-66833) | 9.76 (7.89-11.64) | 39899 (31440-49597) | 7.05 (5.66-8.66) | 22036 (17454-27135) | 3.92 (3.13-4.72) | 20442 (16158-25310) | 3.64 (2.89-4.42) |
| Brunei | Female | 676 (634-719) | 91 (70-112) | 13.43 (10.33-16.34) | 42 (31-54) | 6.29 (4.64-7.91) | 57 (43-72) | 8.48 (6.37-10.56) | 18 (13-24) | 2.70 (1.92-3.47) | 17 (12-22) | 2.50 (1.80-3.25) |
| Brunei | Male | 803 (756-853) | 157 (122-186) | 19.55 (15.53-22.70) | 74 (56-93) | 9.15 (7.10-11.39) | 100 (76-123) | 12.48 (9.64-15.01) | 26 (19-34) | 3.25 (2.42-4.20) | 24 (17-30) | 2.92 (2.21-3.76) |
| Bulgaria | Female | 58448 (55244-61435) | 10472 (8614-12227) | 17.92 (14.91-20.75) | 6268 (5123-7641) | 10.76 (8.71-12.96) | 5153 (4085-6349) | 8.82 (7.05-10.73) | 3378 (2661-4351) | 5.79 (4.58-7.40) | 3019 (2283-3944) | 5.20 (3.93-6.73) |
| Bulgaria | Male | 52292 (49473-55163) | 12356 (10188-14035) | 23.60 (19.70-26.52) | 7780 (6201-9297) | 14.85 (11.88-17.47) | 6340 (5075-7751) | 12.11 (9.74-14.62) | 3321 (2500-4250) | 6.36 (4.76-8.04) | 2852 (2075-3717) | 5.44 (4.02-7.14) |
| Burkina Faso | Female | 25140 (23083-27109) | 2214 (1576-2788) | 8.83 (6.41-11.05) | 1588 (1133-2047) | 6.34 (4.58-8.08) | 953 (671-1267) | 3.80 (2.69-5.06) | 319 (214-439) | 1.27 (0.86-1.74) | 214 (136-309) | 0.86 (0.55-1.23) |
| Burkina Faso | Male | 25925 (23668-28095) | 2610 (1883-3336) | 10.11 (7.35-12.64) | 1850 (1330-2434) | 7.17 (5.17-9.26) | 1111 (812-1509) | 4.29 (3.16-5.70) | 248 (163-358) | 0.95 (0.65-1.36) | 128 (80-202) | 0.49 (0.32-0.76) |
| Burundi | Female | 12792 (11829-13739) | 1896 (1516-2272) | 14.83 (12.16-17.44) | 1325 (1046-1619) | 10.38 (8.38-12.45) | 784 (604-970) | 6.13 (4.80-7.49) | 317 (227-431) | 2.48 (1.78-3.33) | 213 (140-301) | 1.66 (1.09-2.31) |
| Burundi | Male | 15352 (13942-16606) | 1725 (1258-2163) | 11.28 (8.50-13.72) | 1249 (911-1562) | 8.12 (6.02-10.03) | 742 (529-940) | 4.86 (3.55-6.11) | 212 (135-302) | 1.39 (0.91-1.94) | 143 (84-222) | 0.94 (0.54-1.42) |
| Cambodia | Female | 38028 (35908-40332) | 4796 (3491-6094) | 12.61 (9.25-16.02) | 3278 (2382-4257) | 8.60 (6.32-11.10) | 1998 (1412-2650) | 5.28 (3.70-6.87) | 833 (584-1140) | 2.19 (1.56-2.97) | 651 (444-937) | 1.72 (1.18-2.44) |
| Cambodia | Male | 32687 (30761-34648) | 5402 (3698-6779) | 16.62 (11.62-20.40) | 3735 (2546-4789) | 11.47 (7.92-14.54) | 2312 (1557-3032) | 7.11 (4.80-9.22) | 685 (439-943) | 2.09 (1.37-2.89) | 492 (299-711) | 1.51 (0.93-2.17) |
| Cameroon | Female | 37106 (34435-39685) | 3630 (2638-4462) | 9.81 (7.24-11.90) | 2577 (1892-3224) | 6.97 (5.21-8.60) | 1582 (1182-1994) | 4.29 (3.20-5.31) | 577 (398-772) | 1.55 (1.10-2.08) | 323 (212-459) | 0.87 (0.58-1.22) |
| Cameroon | Male | 39196 (36007-42075) | 4321 (3227-5406) | 11.03 (8.31-13.60) | 3051 (2297-3869) | 7.80 (5.96-9.73) | 1865 (1396-2452) | 4.76 (3.63-6.20) | 443 (298-607) | 1.13 (0.77-1.55) | 264 (173-381) | 0.68 (0.45-0.95) |
| Canada | Female | 182511 (171370-192694) | 12677 (9385-15779) | 6.95 (5.13-8.53) | 4014 (2782-5514) | 2.21 (1.52-3.04) | 10117 (7450-12828) | 5.53 (4.07-7.02) | 1407 (955-1986) | 0.77 (0.52-1.09) | 1367 (930-1963) | 0.75 (0.51-1.07) |
| Canada | Male | 215416 (202596-228480) | 24243 (18355-29783) | 11.31 (8.51-13.70) | 7556 (5134-10451) | 3.50 (2.40-4.84) | 19085 (13966-23838) | 8.86 (6.40-10.95) | 2941 (1935-4218) | 1.36 (0.91-1.93) | 2909 (1914-4159) | 1.34 (0.89-1.91) |
| Cape Verde | Female | 1520 (1374-1674) | 147 (112-185) | 9.63 (7.53-11.80) | 101 (79-130) | 6.67 (5.28-8.35) | 65 (50-85) | 4.28 (3.30-5.46) | 40 (30-54) | 2.65 (2.00-3.43) | 34 (25-45) | 2.21 (1.65-2.91) |
| Cape Verde | Male | 1327 (1213-1457) | 147 (112-183) | 11.08 (8.58-13.33) | 102 (77-129) | 7.68 (5.97-9.41) | 66 (48-85) | 4.91 (3.77-6.28) | 26 (19-34) | 1.96 (1.42-2.54) | 19 (13-26) | 1.42 (1.00-1.88) |
| Central African Republic | Female | 7417 (6779-8031) | 516 (348-709) | 6.99 (4.69-9.60) | 403 (269-543) | 5.44 (3.63-7.32) | 250 (170-338) | 3.38 (2.33-4.51) | 91 (54-128) | 1.22 (0.75-1.72) | 48 (28-73) | 0.65 (0.37-0.97) |
| Central African Republic | Male | 7139 (6485-7841) | 588 (374-814) | 8.32 (5.33-11.18) | 447 (278-610) | 6.29 (3.98-8.40) | 277 (178-379) | 3.90 (2.50-5.21) | 58 (33-87) | 0.82 (0.48-1.21) | 33 (18-50) | 0.46 (0.27-0.70) |
| Chad | Female | 16153 (14879-17434) | 1472 (1034-1901) | 9.12 (6.47-11.62) | 1062 (747-1393) | 6.61 (4.68-8.43) | 638 (443-858) | 3.96 (2.78-5.16) | 245 (164-354) | 1.52 (1.01-2.13) | 156 (98-233) | 0.97 (0.61-1.40) |
| Chad | Male | 20541 (18711-22160) | 2141 (1539-2672) | 10.51 (7.55-12.93) | 1532 (1098-1973) | 7.54 (5.45-9.52) | 919 (661-1199) | 4.50 (3.22-5.84) | 257 (165-377) | 1.25 (0.83-1.83) | 170 (106-260) | 0.82 (0.53-1.24) |
| Chile | Female | 51567 (48469-54488) | 4955 (3921-5939) | 9.66 (7.67-11.33) | 2907 (2254-3595) | 5.65 (4.43-6.91) | 2655 (2046-3273) | 5.15 (3.99-6.30) | 1346 (1032-1710) | 2.62 (2.00-3.27) | 1232 (934-1544) | 2.39 (1.84-2.99) |
| Chile | Male | 48300 (45909-50571) | 6846 (5484-8188) | 14.16 (11.49-16.71) | 3996 (3167-4891) | 8.28 (6.59-10.03) | 3631 (2834-4441) | 7.53 (5.92-9.18) | 1548 (1194-1927) | 3.20 (2.52-4.00) | 1315 (998-1648) | 2.72 (2.11-3.43) |
| China | Female | 7442522 (6920386-8033397) | 1318069 (1043020-1566751) | 17.76 (14.12-20.44) | 819458 (631355-1016945) | 10.98 (8.61-13.42) | 676197 (516811-841355) | 9.13 (7.06-11.09) | 226343 (174262-291672) | 3.04 (2.39-3.80) | 187600 (138739-244584) | 2.53 (1.90-3.24) |
| China | Male | 7281356 (6722116-7866937) | 1688639 (1372911-1967348) | 23.28 (18.78-26.50) | 1109861 (870116-1345599) | 15.29 (12.04-18.11) | 929770 (711293-1138166) | 12.79 (9.96-15.50) | 268934 (206858-342166) | 3.70 (2.87-4.68) | 211581 (161728-270117) | 2.91 (2.25-3.69) |
| Colombia | Female | 138789 (129302-148062) | 17656 (14133-21149) | 12.74 (10.25-14.89) | 11664 (9144-14088) | 8.40 (6.66-9.93) | 8090 (6357-9886) | 5.83 (4.65-7.08) | 5028 (3912-6225) | 3.62 (2.85-4.42) | 4350 (3420-5406) | 3.14 (2.49-3.80) |
| Colombia | Male | 138580 (128283-149227) | 28210 (22963-33012) | 20.44 (16.89-23.33) | 18685 (14805-22664) | 13.54 (10.86-16.10) | 13089 (10225-15984) | 9.43 (7.53-11.44) | 5970 (4740-7456) | 4.30 (3.44-5.37) | 4809 (3726-6219) | 3.48 (2.72-4.42) |
| Comoros | Female | 1491 (1363-1614) | 218 (178-266) | 14.67 (11.96-17.52) | 151 (121-188) | 10.12 (8.07-12.19) | 92 (72-116) | 6.21 (4.86-7.68) | 39 (29-53) | 2.62 (1.93-3.52) | 25 (18-36) | 1.69 (1.21-2.37) |
| Comoros | Male | 1382 (1245-1511) | 157 (121-195) | 11.42 (8.93-13.88) | 111 (85-141) | 8.06 (6.25-9.98) | 69 (51-90) | 5.00 (3.81-6.28) | 19 (13-27) | 1.38 (0.96-1.93) | 13 (8-19) | 0.91 (0.62-1.31) |
| Congo | Female | 9181 (8456-9846) | 656 (452-875) | 7.19 (4.92-9.45) | 503 (348-665) | 5.50 (3.81-7.18) | 329 (222-431) | 3.58 (2.49-4.69) | 119 (73-177) | 1.30 (0.81-1.91) | 71 (39-109) | 0.77 (0.43-1.18) |
| Congo | Male | 8982 (8136-9922) | 742 (510-1001) | 8.29 (5.62-10.94) | 555 (380-738) | 6.16 (4.19-8.09) | 354 (240-477) | 3.95 (2.71-5.18) | 89 (57-132) | 0.99 (0.62-1.44) | 64 (39-101) | 0.71 (0.44-1.12) |
| Costa Rica | Female | 15438 (14149-16836) | 1859 (1500-2207) | 12.04 (9.81-13.84) | 1147 (912-1418) | 7.42 (5.91-8.93) | 942 (729-1170) | 6.12 (4.78-7.40) | 570 (441-712) | 3.70 (2.90-4.55) | 552 (425-690) | 3.57 (2.79-4.39) |
| Costa Rica | Male | 16189 (14726-17449) | 2808 (2257-3326) | 17.33 (14.29-20.33) | 1691 (1341-2058) | 10.52 (8.49-12.51) | 1387 (1085-1714) | 8.57 (6.82-10.33) | 659 (515-829) | 4.08 (3.26-5.07) | 620 (480-777) | 3.82 (3.03-4.74) |
| Cote d'Ivoire | Female | 32270 (29965-34824) | 3516 (2680-4278) | 10.88 (8.43-13.14) | 2445 (1825-3046) | 7.55 (5.80-9.34) | 1532 (1141-1955) | 4.72 (3.61-6.00) | 542 (381-736) | 1.67 (1.18-2.27) | 346 (232-511) | 1.07 (0.73-1.55) |
| Cote d'Ivoire | Male | 40830 (37844-44276) | 4800 (3644-5968) | 11.76 (9.16-14.12) | 3344 (2548-4233) | 8.19 (6.35-10.18) | 2115 (1599-2694) | 5.15 (3.97-6.52) | 524 (351-738) | 1.28 (0.87-1.77) | 358 (226-529) | 0.87 (0.57-1.28) |
| Croatia | Female | 24565 (23191-25788) | 4123 (3417-4959) | 16.77 (13.99-19.81) | 2315 (1869-2869) | 9.40 (7.64-11.56) | 2206 (1760-2759) | 8.96 (7.17-11.02) | 1414 (1125-1794) | 5.75 (4.58-7.22) | 1235 (962-1585) | 5.03 (3.96-6.44) |
| Croatia | Male | 23449 (22142-24651) | 5250 (4468-5979) | 22.53 (19.18-25.04) | 3086 (2514-3721) | 13.20 (10.79-15.74) | 2922 (2377-3549) | 12.52 (10.21-14.92) | 1638 (1292-2018) | 6.98 (5.57-8.58) | 1181 (914-1465) | 5.02 (3.90-6.29) |
| Cuba | Female | 57223 (52895-61220) | 4105 (3022-5373) | 7.21 (5.30-9.29) | 2749 (2018-3634) | 4.81 (3.61-6.22) | 2104 (1557-2773) | 3.68 (2.76-4.75) | 1302 (928-1757) | 2.29 (1.67-3.01) | 1200 (853-1626) | 2.10 (1.53-2.82) |
| Cuba | Male | 55995 (51839-60067) | 6291 (4695-7748) | 11.27 (8.53-13.38) | 4056 (2991-5050) | 7.24 (5.41-8.89) | 3054 (2237-3870) | 5.47 (4.05-6.86) | 1456 (1049-1866) | 2.59 (1.90-3.30) | 1294 (936-1672) | 2.32 (1.69-2.96) |
| Cyprus | Female | 4327 (4042-4617) | 225 (156-295) | 5.23 (3.63-6.80) | 114 (79-150) | 2.64 (1.85-3.42) | 158 (112-208) | 3.67 (2.60-4.74) | 45 (29-63) | 1.04 (0.67-1.46) | 41 (26-59) | 0.94 (0.61-1.35) |
| Cyprus | Male | 4632 (4340-4954) | 505 (406-603) | 10.92 (8.88-12.89) | 235 (178-299) | 5.06 (3.89-6.35) | 329 (256-405) | 7.12 (5.59-8.66) | 90 (65-121) | 1.93 (1.43-2.60) | 82 (58-114) | 1.75 (1.25-2.43) |
| Czech Republic | Female | 75061 (70641-79820) | 12678 (10313-14872) | 16.89 (13.92-19.57) | 6338 (5088-7778) | 8.44 (6.83-10.29) | 7564 (6079-9227) | 10.11 (8.15-12.08) | 3285 (2517-4147) | 4.37 (3.40-5.44) | 3060 (2328-3877) | 4.07 (3.14-5.11) |
| Czech Republic | Male | 71974 (67127-76446) | 16135 (13378-18548) | 22.46 (19.01-25.47) | 8476 (6757-10342) | 11.84 (9.53-14.22) | 10053 (8030-12213) | 13.94 (11.45-16.89) | 4339 (3322-5478) | 6.06 (4.65-7.46) | 4055 (3173-5127) | 5.69 (4.43-7.08) |
| Democratic Republic of the Congo | Female | 112876 (105416-120362) | 1920 (635-3736) | 1.70 (0.55-3.27) | 1853 (615-3430) | 1.65 (0.54-3.02) | 1460 (503-2613) | 1.29 (0.45-2.26) | 383 (99-878) | 0.34 (0.09-0.77) | 221 (54-527) | 0.20 (0.05-0.47) |
| Democratic Republic of the Congo | Male | 105439 (97888-113402) | 3579 (1653-6024) | 3.38 (1.52-5.70) | 3187 (1510-5190) | 3.01 (1.41-4.87) | 2236 (1103-3552) | 2.11 (1.03-3.32) | 462 (169-886) | 0.44 (0.16-0.84) | 346 (118-672) | 0.33 (0.12-0.64) |
| Denmark | Female | 27990 (25995-29931) | 1615 (1149-2083) | 5.77 (4.14-7.42) | 320 (196-505) | 1.13 (0.71-1.83) | 1454 (1045-1863) | 5.19 (3.72-6.66) | 112 (63-189) | 0.40 (0.22-0.68) | 71 (39-124) | 0.25 (0.14-0.43) |
| Denmark | Male | 29864 (27805-31734) | 3337 (2695-3980) | 11.19 (9.10-13.12) | 627 (380-948) | 2.10 (1.28-3.16) | 2914 (2335-3542) | 9.81 (7.88-11.72) | 241 (142-386) | 0.81 (0.48-1.28) | 155 (86-255) | 0.52 (0.29-0.84) |
| Djibouti | Female | 1730 (1583-1874) | 255 (199-308) | 14.71 (11.65-17.36) | 174 (133-215) | 10.05 (7.76-12.00) | 110 (82-139) | 6.39 (4.92-7.89) | 41 (29-55) | 2.39 (1.68-3.14) | 27 (17-38) | 1.56 (1.02-2.18) |
| Djibouti | Male | 2295 (2083-2521) | 253 (186-324) | 11.02 (8.32-13.71) | 177 (129-232) | 7.79 (5.70-9.78) | 116 (81-153) | 5.04 (3.60-6.56) | 30 (20-45) | 1.32 (0.85-1.91) | 20 (12-31) | 0.88 (0.52-1.34) |
| Dominica | Female | 258 (235-282) | 20 (14-26) | 7.76 (5.48-9.92) | 14 (10-18) | 5.27 (3.82-6.80) | 10 (7-13) | 3.87 (2.75-5.02) | 7 (5-9) | 2.57 (1.81-3.40) | 6 (4-8) | 2.23 (1.54-2.97) |
| Dominica | Male | 258 (229-285) | 32 (24-39) | 12.42 (9.64-14.76) | 21 (16-27) | 8.11 (6.26-10.02) | 15 (11-19) | 5.84 (4.39-7.41) | 7 (5-9) | 2.54 (1.85-3.44) | 5 (4-8) | 2.09 (1.53-2.90) |
| Dominican Republic | Female | 31410 (29133-33547) | 2456 (1760-3117) | 7.81 (5.63-9.98) | 1680 (1241-2147) | 5.35 (3.94-6.79) | 1244 (923-1605) | 3.97 (2.97-5.09) | 871 (625-1152) | 2.78 (1.99-3.60) | 737 (520-988) | 2.36 (1.68-3.12) |
| Dominican Republic | Male | 34666 (32240-36933) | 4344 (3377-5245) | 12.53 (9.89-14.86) | 2809 (2165-3509) | 8.16 (6.39-10.02) | 2075 (1588-2613) | 6.02 (4.59-7.46) | 1126 (863-1458) | 3.28 (2.49-4.15) | 956 (723-1239) | 2.76 (2.05-3.55) |
| Ecuador | Female | 36135 (33669-38770) | 3555 (2661-4380) | 9.80 (7.46-12.00) | 2383 (1770-3000) | 6.56 (4.88-8.16) | 1633 (1187-2054) | 4.51 (3.28-5.69) | 785 (567-1001) | 2.17 (1.57-2.74) | 703 (501-911) | 1.94 (1.40-2.48) |
| Ecuador | Male | 40030 (36769-43346) | 5523 (4159-6816) | 13.75 (10.61-16.58) | 3657 (2719-4582) | 9.14 (6.86-11.19) | 2526 (1855-3200) | 6.32 (4.68-7.82) | 864 (625-1124) | 2.17 (1.59-2.78) | 677 (488-902) | 1.70 (1.23-2.19) |
| Egypt | Female | 318747 (293768-342158) | 2026 (492-5988) | 0.64 (0.16-1.88) | 1819 (405-5539) | 0.57 (0.13-1.72) | 1551 (300-4453) | 0.49 (0.09-1.38) | 450 (22-2090) | 0.14 (0.01-0.64) | 384 (15-1789) | 0.12 (0.00-0.56) |
| Egypt | Male | 512266 (471579-555750) | 34332 (20963-49810) | 6.73 (4.13-9.64) | 26306 (16318-37360) | 5.13 (3.17-7.20) | 17890 (11637-25445) | 3.51 (2.26-4.90) | 5624 (3217-8451) | 1.09 (0.62-1.64) | 4580 (2561-6968) | 0.89 (0.50-1.37) |
| El Salvador | Female | 17572 (16238-18909) | 2027 (1641-2413) | 11.55 (9.42-13.49) | 1365 (1086-1676) | 7.75 (6.25-9.39) | 906 (712-1134) | 5.17 (4.11-6.30) | 606 (462-779) | 3.45 (2.66-4.35) | 549 (420-707) | 3.14 (2.42-3.99) |
| El Salvador | Male | 15336 (13992-16595) | 2564 (2099-3076) | 16.71 (13.91-19.51) | 1683 (1359-2044) | 11.02 (9.02-13.08) | 1126 (882-1402) | 7.35 (5.82-8.97) | 495 (388-633) | 3.24 (2.52-4.06) | 437 (337-556) | 2.85 (2.24-3.55) |
| Equatorial Guinea | Female | 1752 (1599-1890) | 127 (88-168) | 7.29 (5.11-9.50) | 92 (65-120) | 5.31 (3.72-6.88) | 68 (47-89) | 3.89 (2.72-5.01) | 23 (14-34) | 1.29 (0.81-1.89) | 14 (8-22) | 0.83 (0.46-1.26) |
| Equatorial Guinea | Male | 1473 (1331-1633) | 129 (91-169) | 8.80 (6.22-11.19) | 91 (64-117) | 6.17 (4.36-7.84) | 66 (47-87) | 4.49 (3.19-5.72) | 14 (9-21) | 0.98 (0.64-1.42) | 10 (6-16) | 0.68 (0.42-1.07) |
| Eritrea | Female | 8671 (7977-9379) | 1178 (878-1457) | 13.64 (10.27-16.54) | 829 (609-1037) | 9.57 (6.99-11.75) | 492 (356-631) | 5.72 (4.09-7.15) | 137 (93-188) | 1.58 (1.08-2.15) | 92 (59-131) | 1.06 (0.68-1.51) |
| Eritrea | Male | 7277 (6635-7870) | 723 (521-927) | 9.94 (7.25-12.71) | 532 (371-693) | 7.31 (5.19-9.36) | 321 (224-426) | 4.39 (3.12-5.79) | 57 (38-82) | 0.78 (0.52-1.12) | 40 (25-60) | 0.55 (0.34-0.82) |
| Estonia | Female | 12933 (11655-14108) | 25 (0-132) | 0.19 (0.00-1.02) | 25 (0-122) | 0.19 (0.00-0.91) | 25 (0-125) | 0.19 (0.00-0.97) | 7 (0-41) | 0.06 (0.00-0.31) | 5 (0-33) | 0.04 (0.00-0.25) |
| Estonia | Male | 9373 (8524-10333) | 790 (562-1032) | 8.49 (6.12-10.82) | 423 (297-558) | 4.52 (3.22-5.80) | 523 (379-677) | 5.56 (4.05-7.09) | 111 (74-155) | 1.18 (0.78-1.62) | 86 (56-121) | 0.91 (0.60-1.27) |
| Ethiopia | Female | 104836 (96572-112973) | 13802 (10385-16683) | 13.17 (10.07-15.80) | 9669 (7140-11853) | 9.22 (6.99-11.20) | 5851 (4269-7394) | 5.58 (4.15-6.95) | 1558 (1129-2071) | 1.49 (1.07-1.96) | 753 (491-1072) | 0.72 (0.48-1.03) |
| Ethiopia | Male | 114748 (104663-124394) | 11781 (8747-15093) | 10.28 (7.71-12.95) | 8466 (6118-11078) | 7.41 (5.43-9.41) | 5133 (3763-6819) | 4.50 (3.31-5.81) | 1126 (762-1595) | 0.98 (0.68-1.37) | 723 (495-1067) | 0.64 (0.43-0.92) |
| Federated States of Micronesia | Female | 267 (248-285) | 30 (22-38) | 11.36 (8.08-14.19) | 21 (14-27) | 7.76 (5.35-9.82) | 13 (9-18) | 5.05 (3.59-6.57) | 6 (4-8) | 2.24 (1.51-3.01) | 4 (2-5) | 1.37 (0.87-1.91) |
| Federated States of Micronesia | Male | 287 (261-311) | 41 (28-52) | 14.20 (10.20-17.51) | 28 (19-36) | 9.72 (6.89-12.28) | 18 (12-24) | 6.39 (4.41-8.18) | 6 (4-8) | 2.15 (1.43-2.85) | 4 (2-5) | 1.25 (0.76-1.76) |
| Fiji | Female | 2762 (2537-2978) | 365 (292-450) | 13.25 (10.85-16.04) | 244 (192-307) | 8.82 (7.11-11.13) | 162 (126-208) | 5.90 (4.62-7.40) | 100 (74-132) | 3.60 (2.72-4.64) | 75 (53-102) | 2.72 (1.93-3.67) |
| Fiji | Male | 2916 (2643-3182) | 459 (350-562) | 15.75 (12.38-18.79) | 306 (231-392) | 10.54 (8.09-13.00) | 204 (152-267) | 7.01 (5.36-8.87) | 80 (56-111) | 2.73 (1.94-3.71) | 55 (36-80) | 1.89 (1.26-2.75) |
| Finland | Female | 35915 (33924-38108) | 1822 (1225-2483) | 5.05 (3.45-6.85) | 666 (436-921) | 1.84 (1.22-2.54) | 1568 (1077-2097) | 4.36 (2.99-5.78) | 303 (186-435) | 0.84 (0.51-1.21) | 258 (159-376) | 0.72 (0.44-1.05) |
| Finland | Male | 37543 (35030-39960) | 4673 (3797-5512) | 12.45 (10.36-14.41) | 1419 (1021-1875) | 3.81 (2.75-4.99) | 3668 (2906-4447) | 9.74 (7.95-11.70) | 598 (411-807) | 1.60 (1.11-2.17) | 530 (366-725) | 1.42 (0.98-1.93) |
| France | Female | 315469 (296554-332740) | 16573 (11206-21949) | 5.28 (3.63-7.04) | 7079 (4922-9434) | 2.24 (1.57-2.97) | 13148 (9261-16901) | 4.17 (2.98-5.34) | 2830 (1906-3896) | 0.90 (0.61-1.24) | 2548 (1674-3474) | 0.81 (0.53-1.11) |
| France | Male | 343983 (322755-365726) | 36217 (28656-43862) | 10.55 (8.39-12.46) | 13809 (10324-18094) | 4.03 (3.02-5.23) | 26559 (20413-32782) | 7.72 (6.01-9.44) | 5780 (4195-7914) | 1.69 (1.24-2.30) | 5240 (3748-7121) | 1.52 (1.11-2.05) |
| Gabon | Female | 3309 (3018-3608) | 232 (161-310) | 7.02 (4.91-9.24) | 165 (118-219) | 4.99 (3.62-6.51) | 125 (90-170) | 3.77 (2.71-5.06) | 45 (29-65) | 1.35 (0.91-1.91) | 32 (20-48) | 0.97 (0.60-1.44) |
| Gabon | Male | 3290 (2996-3615) | 270 (183-364) | 8.22 (5.52-10.90) | 190 (128-249) | 5.79 (3.91-7.47) | 142 (97-188) | 4.33 (3.00-5.76) | 33 (20-49) | 1.01 (0.60-1.46) | 24 (14-38) | 0.73 (0.42-1.13) |
| Georgia | Female | 31005 (28656-33303) | 3184 (2456-3937) | 10.30 (8.08-12.59) | 2206 (1661-2716) | 7.09 (5.47-8.67) | 1479 (1137-1886) | 4.79 (3.70-6.05) | 1130 (840-1432) | 3.64 (2.77-4.57) | 1013 (745-1301) | 3.26 (2.45-4.15) |
| Georgia | Male | 24880 (23041-26890) | 4046 (3300-4782) | 16.25 (13.39-18.80) | 2677 (2153-3255) | 10.79 (8.74-12.73) | 1806 (1436-2238) | 7.25 (5.85-8.80) | 1045 (808-1315) | 4.22 (3.26-5.13) | 812 (616-1041) | 3.24 (2.50-4.09) |
| Germany | Female | 526831 (499411-554976) | 30464 (21565-39843) | 5.82 (4.10-7.50) | 11016 (7578-14977) | 2.09 (1.44-2.84) | 25257 (18253-32650) | 4.80 (3.48-6.20) | 4594 (2863-6530) | 0.87 (0.55-1.24) | 4358 (2738-6243) | 0.82 (0.52-1.17) |
| Germany | Male | 619681 (589230-649495) | 76949 (62260-90379) | 12.40 (10.08-14.48) | 24441 (18252-32000) | 3.96 (2.92-5.10) | 59406 (47153-71536) | 9.64 (7.57-11.51) | 10380 (7364-13950) | 1.68 (1.19-2.24) | 9552 (6807-12991) | 1.55 (1.10-2.08) |
| Ghana | Female | 61129 (56988-65488) | 7850 (6222-9475) | 12.87 (10.39-15.15) | 5457 (4230-6773) | 8.95 (6.97-10.91) | 3399 (2630-4341) | 5.54 (4.33-6.93) | 1650 (1217-2133) | 2.70 (2.05-3.42) | 1312 (964-1699) | 2.13 (1.58-2.75) |
| Ghana | Male | 56633 (52524-60189) | 7865 (6139-9413) | 13.95 (11.05-16.47) | 5510 (4204-6758) | 9.77 (7.55-11.80) | 3428 (2602-4353) | 6.11 (4.64-7.59) | 901 (652-1194) | 1.59 (1.16-2.08) | 679 (487-915) | 1.21 (0.88-1.61) |
| Greece | Female | 59703 (56122-63059) | 3308 (2230-4236) | 5.53 (3.65-7.14) | 2007 (1362-2544) | 3.35 (2.31-4.28) | 2010 (1386-2570) | 3.37 (2.34-4.31) | 988 (656-1310) | 1.65 (1.10-2.20) | 973 (641-1283) | 1.62 (1.06-2.15) |
| Greece | Male | 55360 (52873-58466) | 5828 (4549-6961) | 10.55 (8.26-12.56) | 3274 (2504-4009) | 5.91 (4.54-7.24) | 3302 (2523-4078) | 5.97 (4.57-7.38) | 1437 (1070-1815) | 2.60 (1.96-3.25) | 1406 (1040-1775) | 2.54 (1.90-3.19) |
| Greenland | Female | 164 (152-175) | 12 (8-16) | 7.42 (5.08-9.51) | 3 (2-5) | 2.06 (1.23-3.08) | 10 (7-13) | 6.08 (4.15-7.82) | 1 (1-2) | 0.62 (0.36-0.96) | 1 (0-1) | 0.50 (0.29-0.79) |
| Greenland | Male | 275 (252-298) | 32 (22-40) | 11.57 (7.89-14.27) | 9 (5-13) | 3.25 (1.94-4.64) | 26 (17-33) | 9.39 (6.24-11.92) | 3 (2-4) | 0.96 (0.55-1.48) | 2 (1-3) | 0.80 (0.44-1.24) |
| Grenada | Female | 369 (337-404) | 29 (21-37) | 7.75 (5.63-9.85) | 19 (14-25) | 5.25 (3.93-6.71) | 15 (11-19) | 3.97 (2.91-5.13) | 9 (6-13) | 2.49 (1.74-3.37) | 8 (6-11) | 2.17 (1.54-2.99) |
| Grenada | Male | 369 (330-406) | 45 (34-56) | 12.26 (9.32-14.73) | 29 (22-37) | 7.89 (5.93-9.85) | 22 (16-29) | 5.93 (4.37-7.59) | 10 (7-13) | 2.63 (1.84-3.52) | 7 (5-10) | 1.92 (1.34-2.62) |
| Guatemala | Female | 27224 (25189-29078) | 3285 (2689-3878) | 12.11 (10.02-14.03) | 2225 (1753-2678) | 8.20 (6.53-9.76) | 1494 (1161-1833) | 5.50 (4.34-6.71) | 720 (552-943) | 2.66 (2.04-3.37) | 500 (369-660) | 1.84 (1.36-2.41) |
| Guatemala | Male | 28798 (26458-31359) | 4912 (3973-5887) | 17.10 (13.90-19.85) | 3243 (2584-3914) | 11.26 (9.06-13.41) | 2188 (1720-2705) | 7.58 (6.06-9.20) | 737 (567-941) | 2.56 (1.99-3.22) | 511 (384-682) | 1.78 (1.33-2.36) |
| Guinea | Female | 17196 (15942-18586) | 1663 (1226-2072) | 9.69 (7.31-11.95) | 1191 (889-1516) | 6.95 (5.25-8.71) | 726 (538-935) | 4.23 (3.18-5.33) | 366 (257-490) | 2.13 (1.50-2.84) | 180 (116-263) | 1.05 (0.67-1.51) |
| Guinea | Male | 19205 (17632-20578) | 2026 (1526-2520) | 10.60 (8.11-12.89) | 1443 (1070-1849) | 7.55 (5.70-9.45) | 868 (647-1132) | 4.53 (3.39-5.84) | 268 (185-385) | 1.40 (0.97-1.96) | 155 (97-235) | 0.81 (0.51-1.21) |
| Guinea-Bissau | Female | 2520 (2324-2734) | 251 (183-315) | 9.95 (7.31-12.25) | 180 (131-231) | 7.12 (5.34-8.96) | 109 (79-140) | 4.29 (3.18-5.52) | 46 (31-64) | 1.80 (1.25-2.53) | 29 (19-45) | 1.16 (0.77-1.76) |
| Guinea-Bissau | Male | 2433 (2230-2663) | 267 (192-337) | 10.95 (8.09-13.56) | 191 (138-248) | 7.87 (5.77-9.98) | 115 (84-151) | 4.73 (3.53-6.12) | 31 (20-46) | 1.29 (0.83-1.85) | 20 (12-32) | 0.84 (0.49-1.29) |
| Guyana | Female | 2178 (1991-2353) | 166 (118-213) | 7.66 (5.46-9.77) | 113 (81-147) | 5.20 (3.78-6.65) | 88 (63-115) | 4.05 (2.94-5.22) | 47 (33-63) | 2.18 (1.53-2.85) | 37 (25-51) | 1.71 (1.18-2.33) |
| Guyana | Male | 2277 (2064-2483) | 282 (216-350) | 12.41 (9.53-15.00) | 180 (135-233) | 7.93 (5.98-9.97) | 138 (105-182) | 6.07 (4.57-7.76) | 52 (38-71) | 2.30 (1.70-3.01) | 42 (30-57) | 1.85 (1.33-2.46) |
| Haiti | Female | 24827 (23074-26516) | 2036 (1510-2612) | 8.22 (6.00-10.48) | 1523 (1133-1952) | 6.14 (4.56-7.82) | 962 (709-1244) | 3.87 (2.92-4.94) | 558 (399-756) | 2.24 (1.62-3.01) | 299 (204-411) | 1.21 (0.84-1.65) |
| Haiti | Male | 25042 (23038-26908) | 3251 (2514-3897) | 12.98 (10.19-15.21) | 2281 (1748-2803) | 9.14 (7.03-11.00) | 1404 (1027-1748) | 5.60 (4.24-6.94) | 491 (361-624) | 1.96 (1.46-2.48) | 267 (191-356) | 1.07 (0.77-1.42) |
| Honduras | Female | 18074 (16737-19333) | 2095 (1635-2525) | 11.61 (9.33-13.75) | 1438 (1113-1805) | 8.00 (6.28-9.88) | 909 (679-1171) | 5.04 (3.83-6.27) | 606 (449-782) | 3.35 (2.50-4.28) | 564 (415-747) | 3.13 (2.29-4.04) |
| Honduras | Male | 19807 (18333-21264) | 3450 (2771-4149) | 17.48 (14.06-20.46) | 2317 (1820-2879) | 11.70 (9.30-14.07) | 1468 (1133-1828) | 7.41 (5.79-9.09) | 642 (475-849) | 3.26 (2.41-4.26) | 572 (415-764) | 2.90 (2.15-3.85) |
| Hungary | Female | 72196 (67750-76977) | 13634 (11413-15970) | 18.92 (16.05-21.78) | 7594 (6136-9240) | 10.48 (8.64-12.57) | 7341 (5918-9003) | 10.21 (8.28-12.24) | 4055 (3110-5122) | 5.60 (4.35-7.04) | 3622 (2743-4688) | 5.01 (3.83-6.46) |
| Hungary | Male | 60037 (55749-64111) | 15206 (12938-17320) | 25.38 (22.07-28.32) | 9177 (7481-11090) | 15.34 (12.59-18.24) | 8891 (7160-10844) | 14.82 (12.19-17.91) | 4063 (3097-5238) | 6.79 (5.26-8.65) | 3692 (2853-4786) | 6.18 (4.77-7.90) |
| Iceland | Female | 1228 (1140-1309) | 70 (49-91) | 5.73 (4.06-7.36) | 15 (9-22) | 1.20 (0.73-1.79) | 63 (45-82) | 5.16 (3.63-6.63) | 6 (4-10) | 0.52 (0.29-0.83) | 6 (3-9) | 0.48 (0.26-0.74) |
| Iceland | Male | 1727 (1602-1857) | 220 (176-271) | 12.74 (10.35-15.26) | 42 (26-63) | 2.45 (1.49-3.64) | 192 (151-237) | 11.10 (8.87-13.44) | 19 (11-30) | 1.11 (0.67-1.73) | 18 (10-29) | 1.04 (0.62-1.63) |
| India | Female | 4262998 (3893238-4625705) | 482241 (373643-574108) | 11.32 (9.03-13.23) | 334523 (254370-413176) | 7.84 (6.06-9.48) | 208048 (157824-265658) | 4.89 (3.79-6.12) | 67093 (49226-88113) | 1.58 (1.19-2.03) | 59141 (43390-77084) | 1.39 (1.04-1.75) |
| India | Male | 5378990 (4829371-5895547) | 831440 (634699-1000553) | 15.44 (11.81-18.18) | 568450 (427504-701178) | 10.64 (8.06-12.77) | 357274 (259932-450576) | 6.64 (4.94-8.25) | 86819 (63816-114090) | 1.62 (1.21-2.05) | 71827 (52501-95328) | 1.33 (1.00-1.72) |
| Indonesia | Female | 791816 (736582-839154) | 125961 (102209-148588) | 15.96 (13.05-18.68) | 83188 (66419-101226) | 10.55 (8.42-12.71) | 54814 (43344-67643) | 6.93 (5.56-8.47) | 20826 (15995-26866) | 2.63 (2.04-3.34) | 11247 (8380-14773) | 1.42 (1.06-1.85) |
| Indonesia | Male | 789429 (733543-845979) | 160561 (127905-190119) | 20.42 (16.63-23.66) | 107478 (82145-131922) | 13.58 (10.67-16.26) | 71368 (54999-88836) | 8.99 (7.14-10.99) | 17756 (13113-23067) | 2.26 (1.67-2.88) | 10105 (7392-13579) | 1.29 (0.94-1.69) |
| Iran | Female | 369813 (329050-409362) | 1684 (128-5369) | 0.46 (0.03-1.51) | 1583 (128-5120) | 0.43 (0.03-1.43) | 1343 (111-4297) | 0.36 (0.03-1.17) | 349 (2-1807) | 0.09 (0.00-0.50) | 300 (2-1534) | 0.08 (0.00-0.44) |
| Iran | Male | 489739 (434162-539959) | 29291 (18003-42013) | 5.99 (3.62-8.39) | 22276 (14149-31406) | 4.57 (2.90-6.34) | 15339 (9528-21960) | 3.15 (2.02-4.36) | 4538 (2581-6793) | 0.93 (0.54-1.37) | 3620 (2086-5444) | 0.74 (0.42-1.10) |
| Iraq | Female | 127829 (117679-138428) | 790 (188-2086) | 0.61 (0.15-1.59) | 708 (158-1926) | 0.56 (0.13-1.49) | 605 (115-1566) | 0.48 (0.09-1.24) | 188 (15-770) | 0.15 (0.01-0.61) | 136 (8-601) | 0.11 (0.01-0.47) |
| Iraq | Male | 191960 (175040-206173) | 13741 (8761-19484) | 7.21 (4.66-9.96) | 10440 (6737-14243) | 5.45 (3.59-7.31) | 7244 (4887-9954) | 3.80 (2.63-5.09) | 3381 (2039-4988) | 1.78 (1.06-2.59) | 2497 (1508-3711) | 1.31 (0.79-1.92) |
| Ireland | Female | 16305 (15144-17406) | 398 (195-643) | 2.44 (1.18-3.97) | 49 (27-98) | 0.31 (0.16-0.60) | 394 (194-633) | 2.42 (1.17-3.90) | 16 (7-34) | 0.10 (0.05-0.21) | 14 (6-29) | 0.08 (0.04-0.18) |
| Ireland | Male | 20019 (18667-21605) | 1966 (1524-2395) | 9.85 (7.62-11.69) | 152 (104-255) | 0.76 (0.52-1.27) | 1876 (1444-2294) | 9.38 (7.28-11.22) | 43 (29-82) | 0.22 (0.14-0.41) | 38 (24-75) | 0.19 (0.13-0.37) |
| Israel | Female | 28057 (26139-29907) | 1573 (1135-2022) | 5.61 (4.01-7.19) | 540 (361-744) | 1.93 (1.31-2.60) | 1299 (940-1696) | 4.64 (3.33-5.98) | 209 (129-297) | 0.75 (0.48-1.05) | 187 (118-269) | 0.67 (0.42-0.94) |
| Israel | Male | 31325 (29116-33590) | 3609 (2853-4291) | 11.50 (9.27-13.53) | 1142 (819-1485) | 3.64 (2.61-4.67) | 2816 (2194-3438) | 8.99 (7.06-10.84) | 393 (272-542) | 1.25 (0.87-1.73) | 352 (234-493) | 1.12 (0.76-1.55) |
| Italy | Female | 336975 (315862-356605) | 24089 (17775-30122) | 7.14 (5.31-8.98) | 10604 (7472-13992) | 3.16 (2.26-4.14) | 16600 (12373-21194) | 4.94 (3.72-6.27) | 4736 (3259-6341) | 1.40 (0.98-1.87) | 4366 (3002-5962) | 1.29 (0.89-1.75) |
| Italy | Male | 341484 (319948-363657) | 44008 (34002-54045) | 12.93 (10.11-15.43) | 18949 (14046-23990) | 5.54 (4.19-7.03) | 29700 (22600-37123) | 8.67 (6.69-10.69) | 7709 (5682-9940) | 2.27 (1.69-2.90) | 7250 (5312-9560) | 2.14 (1.57-2.77) |
| Jamaica | Female | 10110 (9250-10892) | 806 (596-1024) | 8.01 (6.01-9.93) | 569 (424-726) | 5.65 (4.27-6.99) | 387 (289-496) | 3.84 (2.87-4.86) | 295 (215-386) | 2.92 (2.15-3.74) | 254 (183-341) | 2.53 (1.84-3.27) |
| Jamaica | Male | 9998 (9122-11017) | 1206 (951-1470) | 12.06 (9.80-14.29) | 814 (630-1016) | 8.15 (6.48-9.93) | 548 (425-703) | 5.46 (4.34-6.79) | 276 (209-357) | 2.77 (2.15-3.48) | 221 (166-292) | 2.21 (1.68-2.87) |
| Japan | Female | 682546 (637055-728372) | 73756 (60367-87510) | 10.83 (9.02-12.62) | 29820 (22517-37840) | 4.38 (3.34-5.47) | 51663 (41164-62606) | 7.55 (6.10-9.06) | 12347 (8966-16076) | 1.81 (1.32-2.31) | 11140 (8133-14558) | 1.64 (1.20-2.11) |
| Japan | Male | 541197 (509015-575767) | 88452 (72203-103914) | 16.34 (13.45-18.73) | 34643 (26959-43115) | 6.40 (5.01-7.91) | 60172 (48187-72621) | 11.15 (9.06-13.14) | 15565 (11741-20218) | 2.88 (2.22-3.68) | 13228 (10041-17148) | 2.45 (1.88-3.11) |
| Jordan | Female | 35625 (31572-39420) | 179 (30-534) | 0.50 (0.08-1.50) | 160 (24-499) | 0.45 (0.07-1.39) | 138 (16-393) | 0.39 (0.05-1.10) | 39 (1-192) | 0.11 (0.00-0.54) | 37 (1-183) | 0.10 (0.00-0.51) |
| Jordan | Male | 56042 (49781-62752) | 3733 (2325-5434) | 6.64 (4.21-9.38) | 2850 (1785-4044) | 5.08 (3.21-6.99) | 1950 (1258-2768) | 3.48 (2.26-4.75) | 841 (476-1244) | 1.50 (0.86-2.16) | 757 (432-1130) | 1.36 (0.78-1.98) |
| Kazakhstan | Female | 85069 (79655-90567) | 8761 (6850-10851) | 10.24 (8.13-12.63) | 5703 (4468-7113) | 6.72 (5.28-8.28) | 4465 (3467-5643) | 5.25 (4.10-6.55) | 3050 (2321-3888) | 3.57 (2.72-4.52) | 2791 (2103-3617) | 3.27 (2.48-4.20) |
| Kazakhstan | Male | 73924 (68634-78996) | 12426 (10262-15021) | 16.88 (14.10-19.84) | 7821 (6218-9626) | 10.62 (8.57-12.63) | 6042 (4833-7679) | 8.21 (6.60-10.14) | 3230 (2509-4020) | 4.38 (3.45-5.35) | 2723 (2132-3429) | 3.70 (2.89-4.58) |
| Kenya | Female | 69594 (63787-75198) | 10055 (8086-12237) | 14.46 (11.91-17.07) | 6890 (5482-8486) | 9.91 (8.03-11.97) | 4270 (3320-5328) | 6.16 (4.85-7.52) | 1670 (1237-2198) | 2.40 (1.79-3.11) | 803 (552-1105) | 1.16 (0.81-1.54) |
| Kenya | Male | 66528 (59770-72538) | 63 (0-539) | 0.09 (0.00-0.79) | 63 (0-534) | 0.09 (0.00-0.78) | 62 (0-409) | 0.09 (0.00-0.62) | 8 (0-91) | 0.01 (0.00-0.13) | 3 (0-38) | 0.00 (0.00-0.06) |
| Kiribati | Female | 304 (281-325) | 32 (22-42) | 10.54 (7.17-13.57) | 22 (15-30) | 7.37 (5.01-9.60) | 14 (9-19) | 4.54 (3.08-6.04) | 5 (3-7) | 1.53 (0.99-2.16) | 2 (1-4) | 0.79 (0.44-1.22) |
| Kiribati | Male | 311 (288-333) | 44 (32-55) | 14.17 (10.40-17.27) | 31 (22-39) | 9.99 (7.24-12.39) | 19 (14-25) | 6.19 (4.49-7.84) | 5 (4-7) | 1.70 (1.15-2.30) | 3 (2-4) | 0.98 (0.60-1.44) |
| Kuwait | Female | 14170 (12532-15745) | 780 (444-1179) | 5.49 (3.17-7.94) | 441 (271-640) | 3.13 (1.96-4.48) | 588 (351-867) | 4.15 (2.50-5.91) | 147 (88-213) | 1.04 (0.63-1.48) | 137 (82-201) | 0.97 (0.58-1.38) |
| Kuwait | Male | 27172 (23558-30650) | 3508 (2516-4474) | 12.98 (9.51-15.79) | 1671 (1158-2317) | 6.22 (4.37-8.27) | 2313 (1605-3036) | 8.53 (6.08-10.86) | 513 (351-723) | 1.90 (1.33-2.58) | 465 (320-662) | 1.73 (1.21-2.33) |
| Kyrgyzstan | Female | 19836 (18303-21200) | 1875 (1394-2380) | 9.46 (7.20-11.78) | 1354 (1006-1710) | 6.83 (5.19-8.53) | 834 (612-1066) | 4.20 (3.15-5.37) | 598 (438-802) | 3.03 (2.21-4.01) | 546 (393-726) | 2.76 (1.99-3.64) |
| Kyrgyzstan | Male | 19720 (18192-21279) | 3102 (2335-3785) | 15.76 (11.76-18.84) | 2152 (1608-2660) | 10.88 (8.16-13.31) | 1307 (954-1668) | 6.64 (4.87-8.31) | 565 (408-741) | 2.88 (2.10-3.70) | 406 (290-543) | 2.07 (1.48-2.72) |
| Laos | Female | 12992 (12185-13800) | 1862 (1472-2263) | 14.35 (11.35-17.37) | 1260 (975-1549) | 9.71 (7.51-11.88) | 794 (614-1000) | 6.11 (4.72-7.67) | 273 (198-358) | 2.09 (1.50-2.74) | 197 (134-267) | 1.52 (1.03-2.06) |
| Laos | Male | 14251 (13424-15156) | 2715 (2195-3205) | 19.01 (15.35-22.23) | 1848 (1452-2225) | 12.98 (10.24-15.55) | 1170 (907-1454) | 8.18 (6.41-10.07) | 306 (220-407) | 2.15 (1.55-2.84) | 217 (149-303) | 1.52 (1.04-2.09) |
| Latvia | Female | 16941 (15865-17993) | 617 (308-989) | 3.66 (1.86-5.96) | 476 (265-705) | 2.82 (1.58-4.13) | 485 (268-740) | 2.85 (1.58-4.32) | 250 (137-388) | 1.48 (0.80-2.29) | 209 (108-323) | 1.23 (0.64-1.91) |
| Latvia | Male | 12122 (11212-13091) | 1804 (1448-2140) | 14.89 (12.17-17.26) | 995 (770-1222) | 8.23 (6.42-9.89) | 1023 (811-1269) | 8.42 (6.78-10.12) | 403 (297-523) | 3.33 (2.51-4.17) | 323 (235-421) | 2.67 (1.95-3.40) |
| Lebanon | Female | 32565 (29354-35814) | 135 (29-429) | 0.42 (0.09-1.32) | 124 (26-399) | 0.38 (0.08-1.23) | 104 (20-334) | 0.32 (0.06-1.04) | 27 (1-152) | 0.08 (0.00-0.48) | 25 (0-143) | 0.08 (0.00-0.46) |
| Lebanon | Male | 39801 (36292-43178) | 2359 (1506-3316) | 5.97 (3.78-8.27) | 1816 (1199-2474) | 4.59 (3.01-6.20) | 1255 (821-1722) | 3.17 (2.09-4.26) | 526 (307-784) | 1.33 (0.78-1.95) | 498 (290-730) | 1.25 (0.73-1.82) |
| Lesotho | Female | 4162 (3810-4503) | 292 (196-402) | 7.06 (4.74-9.59) | 229 (154-307) | 5.48 (3.70-7.26) | 143 (98-193) | 3.43 (2.36-4.59) | 78 (49-112) | 1.89 (1.18-2.66) | 72 (44-104) | 1.73 (1.07-2.46) |
| Lesotho | Male | 2807 (2545-3028) | 218 (142-302) | 7.83 (5.13-10.69) | 165 (109-231) | 5.94 (3.98-8.09) | 104 (69-144) | 3.73 (2.55-5.02) | 25 (14-40) | 0.91 (0.51-1.40) | 18 (10-28) | 0.65 (0.37-1.01) |
| Liberia | Female | 6317 (5804-6852) | 610 (461-759) | 9.66 (7.40-11.78) | 441 (336-555) | 6.99 (5.30-8.63) | 265 (200-339) | 4.19 (3.19-5.34) | 110 (79-147) | 1.73 (1.26-2.31) | 68 (45-98) | 1.08 (0.72-1.51) |
| Liberia | Male | 7428 (6733-8156) | 810 (603-1016) | 10.95 (8.20-13.31) | 580 (423-737) | 7.80 (5.79-9.73) | 348 (256-449) | 4.71 (3.43-5.92) | 106 (73-148) | 1.44 (0.98-1.94) | 72 (48-104) | 0.97 (0.66-1.38) |
| Libya | Female | 25934 (23366-28459) | 150 (44-403) | 0.58 (0.17-1.54) | 135 (35-366) | 0.52 (0.14-1.40) | 112 (29-314) | 0.44 (0.11-1.18) | 26 (1-130) | 0.10 (0.00-0.50) | 20 (0-104) | 0.08 (0.00-0.39) |
| Libya | Male | 36059 (32043-40320) | 2481 (1566-3517) | 6.91 (4.39-9.49) | 1862 (1203-2598) | 5.19 (3.41-7.02) | 1339 (886-1873) | 3.72 (2.51-5.04) | 430 (253-652) | 1.20 (0.70-1.80) | 324 (179-500) | 0.90 (0.51-1.37) |
| Lithuania | Female | 25718 (24277-27145) | 2573 (1960-3202) | 10.01 (7.68-12.39) | 1355 (1013-1740) | 5.28 (4.01-6.72) | 1517 (1132-1941) | 5.88 (4.40-7.48) | 717 (518-955) | 2.79 (2.05-3.70) | 583 (402-806) | 2.26 (1.58-3.11) |
| Lithuania | Male | 18668 (17346-20002) | 2204 (1750-2668) | 11.85 (9.54-14.18) | 1170 (905-1471) | 6.28 (4.91-7.84) | 1306 (1021-1653) | 6.98 (5.53-8.77) | 526 (391-689) | 2.82 (2.13-3.65) | 370 (263-503) | 1.98 (1.44-2.66) |
| Luxembourg | Female | 2194 (2048-2338) | 117 (82-153) | 5.31 (3.79-6.87) | 8 (6-11) | 0.35 (0.26-0.53) | 114 (80-149) | 5.18 (3.73-6.69) | 3 (2-4) | 0.12 (0.09-0.19) | 2 (2-4) | 0.11 (0.08-0.17) |
| Luxembourg | Male | 2514 (2344-2685) | 283 (232-343) | 11.32 (9.26-13.38) | 17 (13-25) | 0.68 (0.54-0.99) | 273 (223-331) | 10.91 (8.94-12.95) | 6 (5-9) | 0.24 (0.19-0.36) | 6 (4-9) | 0.23 (0.17-0.34) |
| Macedonia | Female | 12081 (11241-12885) | 2065 (1700-2466) | 17.21 (14.38-20.01) | 1328 (1064-1621) | 11.06 (9.00-13.20) | 942 (736-1187) | 7.83 (6.22-9.66) | 685 (515-874) | 5.69 (4.32-7.16) | 608 (456-801) | 5.05 (3.80-6.54) |
| Macedonia | Male | 12891 (11941-13908) | 3020 (2455-3530) | 23.48 (19.56-26.52) | 2020 (1592-2439) | 15.65 (12.66-18.56) | 1443 (1124-1777) | 11.20 (8.84-13.53) | 845 (633-1106) | 6.57 (5.02-8.50) | 720 (528-965) | 5.62 (4.17-7.31) |
| Madagascar | Female | 36627 (34277-39211) | 5406 (4112-6505) | 14.74 (11.47-17.67) | 3785 (2881-4694) | 10.35 (8.02-12.62) | 2255 (1682-2816) | 6.17 (4.66-7.72) | 799 (559-1061) | 2.18 (1.56-2.90) | 409 (267-607) | 1.13 (0.73-1.64) |
| Madagascar | Male | 37894 (35314-40879) | 4202 (3179-5371) | 11.13 (8.39-13.88) | 3054 (2255-3897) | 8.03 (6.00-10.18) | 1839 (1354-2432) | 4.84 (3.56-6.28) | 435 (286-629) | 1.15 (0.78-1.61) | 232 (139-357) | 0.61 (0.37-0.93) |
| Malawi | Female | 23023 (21113-24829) | 3316 (2633-4006) | 14.43 (11.75-17.08) | 2318 (1810-2820) | 10.06 (8.00-12.12) | 1382 (1053-1763) | 6.01 (4.69-7.41) | 645 (497-825) | 2.81 (2.17-3.49) | 450 (331-595) | 1.97 (1.46-2.54) |
| Malawi | Male | 21467 (19603-23366) | 2382 (1823-3039) | 11.14 (8.59-13.76) | 1719 (1278-2214) | 8.01 (6.06-10.10) | 1034 (773-1377) | 4.82 (3.60-6.20) | 245 (174-333) | 1.13 (0.84-1.54) | 168 (118-240) | 0.79 (0.56-1.10) |
| Malaysia | Female | 80945 (75827-85405) | 12777 (10541-15142) | 15.85 (13.20-18.50) | 7824 (6397-9549) | 9.68 (7.94-11.56) | 6201 (5010-7544) | 7.67 (6.25-9.26) | 2837 (2223-3586) | 3.52 (2.76-4.40) | 2579 (2023-3271) | 3.20 (2.50-3.99) |
| Malaysia | Male | 98136 (91812-104137) | 19917 (16443-22982) | 20.35 (16.98-22.91) | 12334 (9856-14732) | 12.51 (10.24-14.81) | 9720 (7845-11884) | 9.92 (8.03-12.00) | 3713 (2832-4653) | 3.79 (2.92-4.72) | 3388 (2617-4337) | 3.45 (2.68-4.32) |
| Maldives | Female | 692 (645-738) | 99 (81-118) | 14.33 (11.98-16.78) | 62 (50-74) | 8.85 (7.35-10.73) | 47 (38-58) | 6.84 (5.53-8.30) | 17 (13-22) | 2.51 (1.91-3.21) | 14 (10-18) | 1.96 (1.41-2.57) |
| Maldives | Male | 1071 (1001-1141) | 195 (157-230) | 18.24 (14.80-21.08) | 123 (97-150) | 11.46 (9.15-13.74) | 95 (75-116) | 8.84 (7.02-10.66) | 24 (18-32) | 2.22 (1.67-2.90) | 18 (13-24) | 1.67 (1.22-2.26) |
| Mali | Female | 22437 (20530-24335) | 2056 (1429-2583) | 9.17 (6.49-11.38) | 1466 (1040-1899) | 6.60 (4.68-8.40) | 885 (610-1144) | 3.95 (2.80-5.05) | 443 (290-610) | 1.97 (1.33-2.70) | 311 (196-445) | 1.39 (0.90-1.95) |
| Mali | Male | 26866 (24277-29256) | 2590 (1756-3338) | 9.63 (6.72-12.08) | 1849 (1252-2413) | 6.87 (4.79-8.81) | 1101 (758-1480) | 4.13 (2.90-5.35) | 389 (249-549) | 1.45 (0.95-2.01) | 278 (171-409) | 1.05 (0.65-1.48) |
| Malta | Female | 1887 (1755-2007) | 136 (103-168) | 7.19 (5.45-8.79) | 62 (45-79) | 3.28 (2.42-4.20) | 92 (70-117) | 4.88 (3.66-6.10) | 25 (18-35) | 1.34 (0.95-1.83) | 24 (17-33) | 1.31 (0.93-1.75) |
| Malta | Male | 2040 (1896-2187) | 253 (203-305) | 12.39 (10.06-14.83) | 112 (86-144) | 5.50 (4.27-6.91) | 167 (130-206) | 8.20 (6.41-9.99) | 48 (35-64) | 2.32 (1.73-3.09) | 46 (34-62) | 2.27 (1.69-3.02) |
| Marshall Islands | Female | 115 (107-124) | 12 (9-16) | 10.82 (7.82-13.68) | 8 (6-11) | 7.18 (5.14-9.31) | 6 (4-8) | 5.05 (3.52-6.60) | 2 (1-3) | 1.91 (1.30-2.58) | 2 (1-2) | 1.40 (0.92-1.96) |
| Marshall Islands | Male | 140 (129-151) | 19 (14-24) | 13.82 (10.30-16.98) | 13 (10-16) | 9.27 (6.92-11.63) | 9 (6-12) | 6.50 (4.68-8.31) | 3 (2-3) | 1.83 (1.26-2.45) | 2 (1-3) | 1.31 (0.89-1.82) |
| Mauritania | Female | 6673 (6097-7258) | 641 (470-807) | 9.59 (7.23-11.86) | 453 (329-577) | 6.82 (5.00-8.44) | 281 (200-367) | 4.20 (3.12-5.43) | 124 (83-169) | 1.86 (1.26-2.49) | 80 (48-116) | 1.20 (0.73-1.72) |
| Mauritania | Male | 7014 (6328-7745) | 727 (546-919) | 10.43 (7.77-12.74) | 510 (382-654) | 7.30 (5.50-9.12) | 318 (233-416) | 4.54 (3.34-5.81) | 93 (62-136) | 1.34 (0.89-1.90) | 63 (37-98) | 0.90 (0.54-1.34) |
| Mauritius | Female | 4961 (4615-5315) | 716 (595-863) | 14.53 (12.18-17.10) | 449 (366-548) | 9.02 (7.47-10.94) | 336 (269-418) | 6.81 (5.47-8.27) | 189 (152-239) | 3.84 (3.12-4.74) | 184 (146-230) | 3.72 (2.97-4.58) |
| Mauritius | Male | 4619 (4214-4992) | 862 (675-1032) | 18.72 (15.05-21.70) | 543 (420-680) | 11.84 (9.38-14.49) | 409 (311-517) | 8.90 (6.91-10.94) | 199 (149-260) | 4.30 (3.30-5.53) | 188 (140-248) | 4.07 (3.11-5.24) |
| Mexico | Female | 353054 (322604-380954) | 34272 (25989-42602) | 9.74 (7.36-11.81) | 21727 (16225-27512) | 6.17 (4.63-7.65) | 16868 (12424-21801) | 4.79 (3.59-6.00) | 7699 (5638-9969) | 2.19 (1.62-2.80) | 7004 (5110-9043) | 2.00 (1.48-2.52) |
| Mexico | Male | 363466 (330443-395238) | 48905 (38338-59290) | 13.46 (10.78-15.77) | 31048 (24069-38527) | 8.59 (6.75-10.45) | 24302 (18598-30249) | 6.71 (5.20-8.19) | 7306 (5454-9258) | 2.01 (1.58-2.53) | 6439 (4749-8281) | 1.78 (1.35-2.25) |
| Moldova | Female | 26162 (24607-28021) | 956 (495-1496) | 3.68 (1.90-5.74) | 829 (451-1252) | 3.15 (1.72-4.71) | 629 (366-924) | 2.40 (1.39-3.54) | 393 (195-619) | 1.51 (0.75-2.39) | 304 (146-489) | 1.16 (0.54-1.86) |
| Moldova | Male | 22435 (20835-24064) | 2658 (2084-3203) | 11.90 (9.43-14.11) | 1792 (1382-2225) | 8.01 (6.26-9.84) | 1218 (922-1533) | 5.41 (4.20-6.79) | 649 (482-840) | 2.88 (2.17-3.74) | 404 (279-537) | 1.79 (1.26-2.40) |
| Mongolia | Female | 10615 (9687-11435) | 1100 (838-1360) | 10.37 (8.01-12.59) | 769 (579-968) | 7.29 (5.53-8.97) | 516 (383-650) | 4.88 (3.80-6.06) | 327 (240-426) | 3.08 (2.27-3.94) | 291 (213-378) | 2.74 (2.06-3.52) |
| Mongolia | Male | 11496 (10524-12537) | 2086 (1669-2525) | 18.16 (14.82-21.36) | 1398 (1106-1726) | 12.12 (9.80-14.70) | 931 (726-1165) | 8.10 (6.41-9.98) | 489 (375-624) | 4.26 (3.34-5.31) | 364 (274-466) | 3.16 (2.44-4.01) |
| Montenegro | Female | 3582 (3326-3863) | 623 (507-746) | 17.42 (14.28-20.46) | 391 (304-481) | 10.88 (8.64-13.24) | 297 (230-369) | 8.24 (6.59-10.16) | 203 (154-261) | 5.66 (4.32-7.21) | 182 (134-242) | 5.06 (3.77-6.61) |
| Montenegro | Male | 3503 (3171-3830) | 813 (672-957) | 23.32 (19.76-26.26) | 528 (420-641) | 15.08 (12.30-17.91) | 404 (319-502) | 11.56 (9.29-13.98) | 223 (170-289) | 6.39 (4.92-8.13) | 189 (139-258) | 5.44 (3.99-7.20) |
| Morocco | Female | 176390 (162954-188738) | 916 (209-2731) | 0.52 (0.11-1.54) | 848 (166-2533) | 0.48 (0.09-1.44) | 731 (126-2098) | 0.42 (0.07-1.19) | 171 (5-726) | 0.10 (0.00-0.40) | 129 (2-615) | 0.07 (0.00-0.34) |
| Morocco | Male | 251235 (230663-273571) | 15737 (9939-22361) | 6.31 (3.96-8.92) | 12210 (7841-16868) | 4.86 (3.11-6.66) | 8228 (5408-11462) | 3.29 (2.17-4.47) | 2261 (1323-3399) | 0.90 (0.53-1.34) | 1487 (849-2317) | 0.59 (0.34-0.91) |
| Mozambique | Female | 36978 (34075-40068) | 5699 (4559-6771) | 15.33 (12.61-17.99) | 3971 (3131-4803) | 10.72 (8.61-12.77) | 2352 (1833-2905) | 6.38 (5.02-7.75) | 788 (584-1028) | 2.13 (1.56-2.78) | 418 (287-592) | 1.14 (0.79-1.58) |
| Mozambique | Male | 36517 (33846-39023) | 4229 (3163-5322) | 11.56 (8.98-14.40) | 3082 (2295-3938) | 8.47 (6.34-10.69) | 1850 (1369-2419) | 5.05 (3.78-6.55) | 353 (241-506) | 0.97 (0.67-1.36) | 175 (109-264) | 0.48 (0.30-0.72) |
| Myanmar | Female | 147489 (139296-155539) | 22108 (17597-26063) | 14.94 (12.09-17.50) | 15082 (11933-18167) | 10.20 (8.12-12.25) | 9108 (7104-11116) | 6.19 (4.87-7.53) | 5197 (4003-6479) | 3.52 (2.72-4.41) | 3352 (2520-4435) | 2.27 (1.69-2.98) |
| Myanmar | Male | 132330 (125332-139754) | 25592 (20467-29928) | 19.35 (15.59-22.50) | 17560 (13607-21430) | 13.28 (10.31-16.12) | 10730 (8384-13504) | 8.13 (6.37-10.11) | 4433 (3309-5738) | 3.36 (2.52-4.29) | 2560 (1844-3461) | 1.94 (1.41-2.59) |
| Namibia | Female | 4753 (4350-5149) | 329 (216-439) | 6.92 (4.56-9.16) | 244 (163-319) | 5.15 (3.50-6.69) | 169 (115-225) | 3.57 (2.47-4.68) | 75 (48-104) | 1.59 (1.02-2.17) | 66 (41-93) | 1.39 (0.87-1.94) |
| Namibia | Male | 3911 (3544-4269) | 316 (214-435) | 8.10 (5.50-10.87) | 232 (156-314) | 5.92 (3.97-7.97) | 159 (109-221) | 4.06 (2.82-5.54) | 52 (30-79) | 1.33 (0.79-1.97) | 46 (26-70) | 1.18 (0.69-1.78) |
| Nauru | Female | 23 (21-24) | 3 (2-4) | 12.55 (10.15-15.07) | 2 (1-2) | 7.91 (6.25-9.73) | 1 (1-2) | 6.42 (4.99-7.95) | 1 (0-1) | 2.58 (1.92-3.42) | 0 (0-0) | 1.54 (1.07-2.16) |
| Nauru | Male | 22 (20-24) | 3 (3-4) | 14.76 (11.78-17.66) | 2 (2-3) | 9.33 (7.35-11.29) | 2 (1-2) | 7.54 (5.83-9.40) | 1 (0-1) | 2.64 (1.98-3.41) | 0 (0-1) | 1.76 (1.25-2.32) |
| Nepal | Female | 75477 (69340-81377) | 7113 (4528-9726) | 9.42 (6.07-12.47) | 5046 (3180-7035) | 6.69 (4.20-9.19) | 3036 (1929-4407) | 4.06 (2.57-5.63) | 920 (561-1305) | 1.22 (0.75-1.72) | 645 (391-929) | 0.86 (0.53-1.23) |
| Nepal | Male | 95399 (87157-103174) | 12772 (9775-15873) | 13.49 (10.47-16.22) | 8984 (6694-11522) | 9.51 (7.28-11.69) | 5472 (4011-7101) | 5.73 (4.23-7.26) | 1441 (1039-1908) | 1.52 (1.12-1.97) | 839 (589-1124) | 0.88 (0.63-1.17) |
| Netherlands | Female | 76053 (71562-80656) | 4019 (2829-5386) | 5.28 (3.71-6.94) | 1204 (782-1721) | 1.58 (1.04-2.24) | 3511 (2487-4544) | 4.62 (3.27-5.98) | 416 (246-623) | 0.54 (0.33-0.81) | 345 (207-528) | 0.45 (0.27-0.70) |
| Netherlands | Male | 88317 (81506-94719) | 9597 (7549-11624) | 10.92 (8.78-12.92) | 2540 (1717-3579) | 2.89 (1.95-3.98) | 7874 (6121-9753) | 8.92 (7.11-10.90) | 817 (526-1187) | 0.93 (0.60-1.36) | 729 (460-1091) | 0.84 (0.53-1.23) |
| New Zealand | Female | 20985 (19665-22210) | 1229 (852-1589) | 5.88 (4.09-7.53) | 446 (298-619) | 2.14 (1.44-2.92) | 990 (692-1303) | 4.73 (3.29-6.15) | 172 (107-245) | 0.82 (0.52-1.17) | 151 (96-215) | 0.72 (0.46-1.03) |
| New Zealand | Male | 24141 (22520-25889) | 2777 (2219-3316) | 11.48 (9.26-13.46) | 923 (668-1235) | 3.82 (2.82-5.06) | 2146 (1665-2634) | 8.88 (6.93-10.72) | 313 (220-439) | 1.30 (0.93-1.81) | 273 (190-382) | 1.13 (0.79-1.57) |
| Nicaragua | Female | 13142 (12054-14191) | 1615 (1310-1908) | 12.26 (10.25-14.31) | 1115 (884-1364) | 8.50 (6.91-10.27) | 692 (546-864) | 5.25 (4.19-6.53) | 450 (348-575) | 3.41 (2.66-4.30) | 413 (312-538) | 3.15 (2.41-4.05) |
| Nicaragua | Male | 13570 (12538-14681) | 2421 (1971-2893) | 17.84 (14.74-20.93) | 1645 (1321-1988) | 12.06 (9.87-14.38) | 1021 (800-1267) | 7.47 (5.98-9.18) | 443 (330-579) | 3.28 (2.45-4.15) | 403 (299-535) | 2.97 (2.17-3.85) |
| Niger | Female | 23673 (21716-25551) | 2289 (1650-2880) | 9.66 (7.15-11.99) | 1645 (1156-2098) | 6.96 (4.91-8.66) | 980 (691-1282) | 4.15 (3.03-5.36) | 307 (193-428) | 1.30 (0.84-1.80) | 144 (82-220) | 0.61 (0.35-0.93) |
| Niger | Male | 24026 (21941-26235) | 2555 (1897-3209) | 10.64 (7.89-13.01) | 1825 (1343-2331) | 7.56 (5.57-9.50) | 1085 (783-1427) | 4.55 (3.27-5.88) | 204 (129-306) | 0.85 (0.54-1.24) | 134 (76-207) | 0.55 (0.32-0.85) |
| Nigeria | Female | 298357 (273390-324465) | 23133 (15933-30964) | 7.78 (5.40-10.11) | 17196 (12317-22933) | 5.77 (4.17-7.47) | 10898 (7772-14710) | 3.67 (2.67-4.83) | 4393 (2984-6122) | 1.48 (1.02-2.00) | 2535 (1691-3504) | 0.85 (0.57-1.15) |
| Nigeria | Male | 272783 (245997-295271) | 25366 (18495-32527) | 9.36 (6.92-11.72) | 18311 (13474-23820) | 6.74 (4.98-8.62) | 11479 (8307-14975) | 4.22 (3.14-5.42) | 3384 (2346-4617) | 1.24 (0.87-1.67) | 2336 (1615-3281) | 0.86 (0.61-1.19) |
| Norway | Female | 23317 (21701-24712) | 766 (457-1115) | 3.29 (1.95-4.75) | 101 (59-186) | 0.43 (0.25-0.78) | 751 (448-1092) | 3.24 (1.91-4.68) | 38 (20-74) | 0.16 (0.08-0.32) | 26 (13-56) | 0.11 (0.06-0.23) |
| Norway | Male | 27412 (25388-29335) | 3069 (2428-3654) | 11.18 (9.08-13.09) | 273 (176-478) | 1.00 (0.65-1.74) | 2900 (2292-3467) | 10.56 (8.50-12.44) | 103 (63-200) | 0.38 (0.23-0.71) | 89 (54-174) | 0.32 (0.20-0.63) |
| Oman | Female | 8926 (8050-9865) | 67 (18-174) | 0.76 (0.19-1.90) | 56 (13-143) | 0.63 (0.15-1.58) | 57 (13-141) | 0.64 (0.14-1.59) | 9 (0-41) | 0.10 (0.00-0.47) | 8 (0-38) | 0.09 (0.00-0.42) |
| Oman | Male | 18325 (16107-20367) | 1476 (970-1995) | 8.10 (5.36-10.63) | 937 (618-1276) | 5.13 (3.42-6.85) | 931 (597-1272) | 5.11 (3.41-6.82) | 169 (102-251) | 0.93 (0.56-1.36) | 148 (89-218) | 0.81 (0.48-1.17) |
| Pakistan | Female | 473636 (433436-513059) | 64466 (50563-77460) | 13.58 (10.99-15.69) | 45033 (34987-55686) | 9.51 (7.47-11.33) | 27468 (20993-34564) | 5.81 (4.51-7.18) | 13452 (9853-17324) | 2.85 (2.15-3.53) | 11158 (8182-14789) | 2.35 (1.74-3.03) |
| Pakistan | Male | 695521 (625436-767076) | 102178 (80288-125854) | 14.77 (11.74-17.32) | 71621 (55118-91179) | 10.33 (8.11-12.68) | 43872 (33742-57041) | 6.32 (4.88-7.97) | 12065 (8622-16124) | 1.74 (1.25-2.31) | 9149 (6414-12568) | 1.31 (0.94-1.78) |
| Palau | Female | 70 (64-76) | 9 (7-11) | 13.02 (10.26-15.72) | 6 (4-7) | 8.03 (6.13-9.99) | 5 (3-6) | 6.66 (5.11-8.31) | 2 (2-3) | 3.13 (2.33-4.03) | 2 (1-2) | 2.66 (1.94-3.50) |
| Palau | Male | 92 (83-100) | 14 (11-18) | 15.61 (12.18-18.48) | 9 (7-11) | 9.57 (7.34-11.63) | 7 (5-9) | 7.98 (6.06-9.92) | 3 (2-4) | 2.99 (2.23-3.91) | 2 (1-3) | 2.23 (1.61-2.97) |
| Panama | Female | 11277 (10330-12114) | 1342 (1088-1601) | 11.95 (9.82-13.92) | 803 (629-985) | 7.14 (5.67-8.60) | 713 (559-878) | 6.35 (5.06-7.73) | 331 (254-427) | 2.94 (2.29-3.72) | 293 (220-378) | 2.61 (2.02-3.30) |
| Panama | Male | 12851 (11742-14076) | 2191 (1763-2638) | 17.06 (14.14-19.95) | 1278 (1012-1572) | 9.94 (8.06-11.97) | 1136 (906-1429) | 8.86 (7.20-10.73) | 408 (316-521) | 3.17 (2.50-3.97) | 339 (263-438) | 2.65 (2.08-3.37) |
| Papua New Guinea | Female | 12304 (11307-13338) | 1267 (868-1620) | 10.33 (7.02-12.88) | 886 (597-1142) | 7.22 (4.83-9.07) | 562 (382-739) | 4.58 (3.10-5.94) | 161 (96-235) | 1.31 (0.78-1.89) | 98 (51-155) | 0.80 (0.43-1.26) |
| Papua New Guinea | Male | 17702 (16232-19094) | 2288 (1576-2883) | 12.94 (8.99-16.27) | 1596 (1096-2066) | 9.03 (6.31-11.55) | 1015 (683-1336) | 5.76 (3.86-7.56) | 188 (107-289) | 1.07 (0.61-1.61) | 126 (64-209) | 0.72 (0.37-1.14) |
| Paraguay | Female | 11441 (10769-12174) | 1282 (1032-1482) | 11.14 (9.11-12.84) | 845 (667-1012) | 7.38 (5.86-8.85) | 582 (453-709) | 5.07 (4.06-6.16) | 392 (302-482) | 3.42 (2.65-4.17) | 315 (238-404) | 2.75 (2.09-3.52) |
| Paraguay | Male | 14624 (13592-15785) | 2217 (1745-2679) | 15.13 (12.07-17.93) | 1465 (1115-1801) | 9.98 (7.73-12.06) | 1012 (777-1258) | 6.93 (5.36-8.54) | 524 (384-674) | 3.60 (2.69-4.56) | 400 (287-536) | 2.74 (1.98-3.57) |
| Peru | Female | 66491 (61491-71422) | 6511 (4788-7911) | 9.78 (7.31-11.80) | 4287 (3064-5398) | 6.47 (4.63-8.00) | 3016 (2178-3854) | 4.55 (3.27-5.75) | 974 (710-1224) | 1.46 (1.08-1.82) | 881 (640-1114) | 1.32 (0.98-1.66) |
| Peru | Male | 76614 (70261-82815) | 10661 (8126-13203) | 13.93 (10.67-16.88) | 7031 (5224-8848) | 9.18 (6.91-11.28) | 4903 (3721-6284) | 6.41 (4.94-8.00) | 1060 (818-1355) | 1.38 (1.08-1.74) | 894 (692-1135) | 1.17 (0.91-1.48) |
| Philippines | Female | 214510 (200623-226456) | 28329 (21813-34419) | 13.20 (10.27-15.90) | 18909 (14332-23488) | 8.83 (6.78-10.80) | 12230 (9216-15387) | 5.70 (4.37-7.10) | 5610 (4162-7102) | 2.63 (1.97-3.28) | 4160 (3098-5397) | 1.94 (1.46-2.49) |
| Philippines | Male | 196711 (183253-209200) | 36192 (26750-43059) | 18.37 (13.87-21.55) | 24576 (17986-29874) | 12.47 (9.36-15.17) | 15880 (11543-19978) | 8.11 (5.89-10.00) | 5451 (3842-6935) | 2.76 (1.96-3.52) | 3696 (2530-4908) | 1.88 (1.30-2.45) |
| Poland | Female | 177295 (166378-188256) | 22535 (18120-26899) | 12.75 (10.40-14.95) | 12845 (10031-15723) | 7.28 (5.72-8.76) | 12225 (9630-15092) | 6.92 (5.51-8.43) | 7418 (5711-9240) | 4.20 (3.27-5.18) | 7132 (5495-8919) | 4.02 (3.14-4.96) |
| Poland | Male | 180342 (168612-192736) | 34944 (27951-41137) | 19.35 (15.60-22.56) | 19780 (15464-24414) | 10.96 (8.63-13.33) | 18799 (14914-23020) | 10.45 (8.37-12.73) | 9471 (7307-11939) | 5.25 (4.05-6.53) | 8688 (6609-11088) | 4.83 (3.72-6.08) |
| Portugal | Female | 49133 (46142-52101) | 2020 (1355-2792) | 4.12 (2.71-5.66) | 1258 (836-1714) | 2.56 (1.69-3.48) | 1404 (932-1853) | 2.86 (1.90-3.80) | 591 (363-842) | 1.20 (0.73-1.72) | 571 (341-821) | 1.16 (0.70-1.69) |
| Portugal | Male | 41221 (38792-43523) | 4344 (3514-5181) | 10.56 (8.62-12.49) | 2294 (1800-2849) | 5.58 (4.37-6.83) | 2588 (2059-3178) | 6.26 (5.03-7.66) | 952 (698-1202) | 2.31 (1.72-2.90) | 892 (669-1135) | 2.18 (1.63-2.75) |
| Puerto Rico | Female | 21772 (19664-23676) | 1717 (1279-2154) | 7.94 (6.02-9.83) | 842 (619-1079) | 3.88 (2.88-4.94) | 1176 (863-1501) | 5.40 (4.02-6.79) | 397 (280-542) | 1.83 (1.32-2.46) | 338 (231-468) | 1.55 (1.09-2.12) |
| Puerto Rico | Male | 18320 (16725-20000) | 2154 (1707-2619) | 11.77 (9.47-13.91) | 1008 (764-1295) | 5.51 (4.25-6.88) | 1415 (1086-1755) | 7.73 (6.05-9.41) | 357 (252-489) | 1.93 (1.43-2.61) | 295 (208-413) | 1.61 (1.14-2.22) |
| Qatar | Female | 3391 (3031-3737) | 17 (0-55) | 0.50 (0.00-1.60) | 7 (0-24) | 0.21 (0.00-0.70) | 17 (0-53) | 0.49 (0.00-1.54) | 2 (0-9) | 0.05 (0.00-0.25) | 2 (0-8) | 0.05 (0.00-0.23) |
| Qatar | Male | 12252 (10566-13921) | 974 (574-1418) | 7.99 (4.81-11.44) | 231 (120-393) | 1.89 (1.02-3.14) | 882 (519-1270) | 7.28 (4.41-10.21) | 48 (24-88) | 0.39 (0.20-0.69) | 43 (21-77) | 0.35 (0.17-0.61) |
| Romania | Female | 140278 (132575-146922) | 24590 (20621-29040) | 17.48 (14.74-20.84) | 14352 (11637-17552) | 10.24 (8.32-12.50) | 12563 (10064-15497) | 8.96 (7.26-11.06) | 7481 (5870-9419) | 5.35 (4.22-6.72) | 6606 (5210-8281) | 4.71 (3.73-5.89) |
| Romania | Male | 121444 (115297-128074) | 28246 (23702-32212) | 23.25 (19.66-26.17) | 17218 (13805-20535) | 14.18 (11.52-16.72) | 15116 (12260-18542) | 12.45 (10.16-14.88) | 7799 (6063-9597) | 6.43 (5.06-7.79) | 6988 (5487-8520) | 5.76 (4.51-7.00) |
| Russia | Female | 1517552 (1370144-1665889) | 158945 (124181-195849) | 10.53 (8.30-12.76) | 99092 (75209-125249) | 6.53 (5.00-8.12) | 81105 (61774-103189) | 5.33 (4.10-6.70) | 51485 (37731-66737) | 3.39 (2.49-4.38) | 36839 (26568-49052) | 2.43 (1.77-3.18) |
| Russia | Male | 1066707 (963023-1155574) | 191251 (153820-230418) | 18.00 (14.61-20.95) | 116715 (91066-143348) | 10.96 (8.67-13.29) | 95463 (73943-120194) | 9.02 (7.07-11.02) | 46515 (35156-59098) | 4.38 (3.36-5.44) | 32633 (24427-41839) | 3.09 (2.31-3.87) |
| Rwanda | Female | 19532 (17966-21068) | 2717 (2070-3360) | 13.95 (10.83-16.82) | 1894 (1431-2356) | 9.73 (7.35-11.88) | 1128 (852-1455) | 5.81 (4.42-7.27) | 262 (178-363) | 1.34 (0.92-1.84) | 120 (71-186) | 0.61 (0.37-0.94) |
| Rwanda | Male | 15496 (14123-16675) | 1676 (1226-2139) | 10.84 (8.11-13.52) | 1201 (866-1535) | 7.79 (5.77-9.80) | 727 (520-961) | 4.70 (3.44-6.15) | 107 (68-162) | 0.70 (0.45-1.03) | 65 (39-101) | 0.42 (0.25-0.65) |
| Saint Kitts and Nevis | Female | 220 (201-241) | 17 (12-22) | 7.74 (5.38-9.94) | 11 (8-14) | 4.79 (3.43-6.23) | 10 (7-13) | 4.55 (3.26-5.86) | 5 (3-7) | 2.18 (1.51-2.95) | 4 (3-6) | 1.97 (1.35-2.74) |
| Saint Kitts and Nevis | Male | 233 (210-256) | 30 (23-37) | 13.05 (10.16-15.29) | 17 (13-22) | 7.54 (5.78-9.37) | 17 (12-21) | 7.18 (5.44-8.95) | 5 (4-7) | 2.30 (1.66-3.02) | 4 (3-6) | 1.82 (1.26-2.46) |
| Saint Lucia | Female | 781 (710-849) | 60 (43-77) | 7.68 (5.67-9.63) | 40 (29-51) | 5.09 (3.82-6.43) | 31 (23-41) | 4.04 (2.99-5.09) | 20 (14-26) | 2.51 (1.83-3.30) | 17 (12-22) | 2.14 (1.55-2.81) |
| Saint Lucia | Male | 790 (714-868) | 94 (72-117) | 11.98 (9.33-14.40) | 60 (45-79) | 7.55 (5.78-9.47) | 47 (36-63) | 5.97 (4.56-7.63) | 19 (14-25) | 2.41 (1.78-3.15) | 14 (10-19) | 1.74 (1.26-2.35) |
| Saint Vincent and the Grenadines | Female | 411 (374-450) | 31 (22-41) | 7.56 (5.39-9.76) | 21 (15-28) | 5.16 (3.81-6.65) | 16 (12-21) | 3.84 (2.79-4.99) | 10 (7-14) | 2.34 (1.63-3.23) | 8 (6-11) | 1.96 (1.36-2.70) |
| Saint Vincent and the Grenadines | Male | 478 (430-524) | 57 (44-70) | 12.00 (9.26-14.38) | 37 (28-47) | 7.80 (5.98-9.61) | 27 (20-36) | 5.72 (4.32-7.23) | 11 (8-15) | 2.32 (1.68-3.03) | 8 (6-12) | 1.78 (1.30-2.38) |
| Samoa | Female | 525 (484-564) | 25 (16-34) | 4.76 (3.18-6.40) | 18 (12-25) | 3.43 (2.23-4.72) | 12 (8-17) | 2.33 (1.50-3.29) | 4 (2-7) | 0.82 (0.47-1.28) | 2 (1-3) | 0.31 (0.13-0.54) |
| Samoa | Male | 562 (512-618) | 29 (19-40) | 5.18 (3.48-7.17) | 21 (14-31) | 3.79 (2.48-5.37) | 15 (9-21) | 2.60 (1.69-3.63) | 4 (3-7) | 0.77 (0.47-1.19) | 3 (2-5) | 0.51 (0.30-0.85) |
| Sao Tome and Principe | Female | 390 (359-425) | 39 (29-49) | 10.12 (7.74-12.30) | 28 (21-35) | 7.14 (5.44-8.84) | 17 (13-23) | 4.48 (3.43-5.70) | 9 (7-12) | 2.41 (1.76-3.13) | 6 (4-8) | 1.56 (1.12-2.09) |
| Sao Tome and Principe | Male | 373 (338-404) | 41 (31-52) | 11.12 (8.59-13.50) | 29 (22-37) | 7.86 (6.04-9.90) | 18 (14-24) | 4.88 (3.72-6.22) | 6 (4-8) | 1.54 (1.12-2.06) | 3 (2-5) | 0.89 (0.61-1.22) |
| Saudi Arabia | Female | 78423 (70154-86169) | 442 (16-1183) | 0.56 (0.02-1.51) | 363 (16-973) | 0.47 (0.02-1.25) | 377 (16-1016) | 0.48 (0.02-1.33) | 51 (0-238) | 0.06 (0.00-0.31) | 42 (0-204) | 0.05 (0.00-0.26) |
| Saudi Arabia | Male | 175894 (158559-193868) | 12733 (7825-18699) | 7.25 (4.42-10.39) | 7741 (4848-10972) | 4.40 (2.75-6.10) | 8586 (5449-12188) | 4.90 (3.16-6.73) | 1468 (848-2177) | 0.84 (0.48-1.23) | 1165 (659-1759) | 0.67 (0.38-0.99) |
| Senegal | Female | 24987 (22889-26983) | 2412 (1793-2991) | 9.67 (7.27-11.81) | 1714 (1274-2175) | 6.87 (5.13-8.56) | 1055 (768-1375) | 4.21 (3.10-5.48) | 445 (310-599) | 1.78 (1.27-2.37) | 241 (154-355) | 0.97 (0.62-1.39) |
| Senegal | Male | 24886 (22580-27117) | 2613 (1937-3337) | 10.50 (7.83-13.09) | 1846 (1346-2419) | 7.43 (5.52-9.46) | 1127 (826-1515) | 4.54 (3.44-5.93) | 310 (206-446) | 1.24 (0.86-1.76) | 172 (112-268) | 0.69 (0.45-1.05) |
| Serbia | Female | 75484 (71238-79777) | 13529 (11360-16061) | 18.00 (15.06-20.87) | 8493 (6883-10272) | 11.21 (9.15-13.53) | 6385 (5143-7694) | 8.48 (6.91-10.26) | 4956 (3926-6072) | 6.55 (5.19-8.00) | 4643 (3642-5734) | 6.12 (4.83-7.56) |
| Serbia | Male | 58508 (54735-62245) | 13563 (11475-15575) | 23.22 (19.76-26.14) | 8852 (7241-10459) | 15.10 (12.49-17.68) | 6699 (5371-8194) | 11.47 (9.28-13.94) | 3984 (3071-4886) | 6.79 (5.39-8.30) | 3598 (2826-4492) | 6.15 (4.91-7.58) |
| Seychelles | Female | 302 (280-323) | 38 (30-46) | 12.71 (10.23-15.11) | 23 (18-28) | 7.63 (6.03-9.37) | 19 (15-24) | 6.37 (4.97-7.78) | 11 (9-14) | 3.79 (2.96-4.72) | 10 (8-13) | 3.46 (2.65-4.31) |
| Seychelles | Male | 315 (291-339) | 51 (40-62) | 16.28 (12.98-19.00) | 31 (24-39) | 9.87 (7.66-11.96) | 26 (20-33) | 8.19 (6.33-10.14) | 12 (9-16) | 3.80 (2.89-4.88) | 11 (8-14) | 3.37 (2.48-4.41) |
| Sierra Leone | Female | 12028 (11122-12918) | 1186 (873-1462) | 9.92 (7.36-12.05) | 856 (626-1065) | 7.13 (5.30-8.69) | 508 (379-641) | 4.25 (3.19-5.24) | 207 (142-282) | 1.72 (1.20-2.34) | 121 (79-173) | 1.01 (0.67-1.40) |
| Sierra Leone | Male | 12994 (11818-14099) | 1431 (1071-1793) | 11.01 (8.45-13.49) | 1025 (770-1298) | 7.88 (6.03-9.79) | 606 (455-795) | 4.67 (3.60-6.00) | 157 (104-219) | 1.22 (0.81-1.67) | 89 (56-132) | 0.69 (0.44-1.00) |
| Singapore | Female | 14698 (13674-15631) | 1620 (1103-2051) | 11.08 (7.59-13.87) | 231 (123-384) | 1.57 (0.84-2.63) | 1459 (1007-1889) | 9.95 (6.89-12.78) | 99 (50-168) | 0.68 (0.35-1.16) | 95 (48-162) | 0.65 (0.33-1.12) |
| Singapore | Male | 16770 (15490-18159) | 2659 (1881-3388) | 15.96 (11.29-19.66) | 393 (205-648) | 2.33 (1.24-3.81) | 2418 (1666-3098) | 14.46 (10.10-17.99) | 147 (72-256) | 0.87 (0.44-1.51) | 138 (67-242) | 0.83 (0.41-1.45) |
| Slovakia | Female | 34038 (32016-35925) | 5923 (4895-6917) | 17.48 (14.52-20.23) | 3157 (2520-3858) | 9.29 (7.53-11.31) | 3325 (2661-4061) | 9.75 (7.89-11.89) | 1739 (1342-2211) | 5.12 (4.00-6.41) | 1618 (1250-2055) | 4.78 (3.70-5.99) |
| Slovakia | Male | 29411 (27514-31175) | 6814 (5751-7848) | 23.24 (19.90-26.33) | 3812 (3084-4657) | 13.06 (10.66-15.62) | 4011 (3254-4839) | 13.66 (11.07-16.41) | 1835 (1425-2317) | 6.24 (4.91-7.73) | 1682 (1294-2161) | 5.74 (4.46-7.27) |
| Slovenia | Female | 12746 (11785-13710) | 2105 (1730-2505) | 16.39 (13.84-19.60) | 1009 (793-1255) | 7.91 (6.30-9.72) | 1286 (1029-1594) | 10.08 (8.27-12.45) | 530 (401-686) | 4.14 (3.19-5.28) | 473 (352-626) | 3.72 (2.76-4.89) |
| Slovenia | Male | 12417 (11391-13510) | 2733 (2222-3236) | 22.05 (18.23-25.08) | 1369 (1056-1715) | 11.08 (8.59-13.41) | 1746 (1374-2153) | 14.01 (11.29-16.83) | 574 (419-768) | 4.63 (3.42-5.96) | 494 (348-672) | 3.98 (2.84-5.26) |
| Solomon Islands | Female | 1423 (1341-1513) | 161 (117-203) | 11.28 (8.08-14.03) | 112 (80-143) | 7.82 (5.59-9.91) | 70 (49-92) | 4.95 (3.49-6.34) | 28 (19-38) | 1.93 (1.33-2.64) | 11 (7-17) | 0.80 (0.48-1.16) |
| Solomon Islands | Male | 1650 (1523-1768) | 210 (141-273) | 12.82 (8.68-16.26) | 146 (98-194) | 8.89 (6.02-11.66) | 93 (61-124) | 5.65 (3.78-7.41) | 17 (10-24) | 1.01 (0.61-1.46) | 8 (5-14) | 0.51 (0.29-0.81) |
| Somalia | Female | 19608 (17959-21183) | 2762 (2061-3458) | 14.11 (10.57-17.22) | 1940 (1426-2434) | 9.90 (7.33-12.31) | 1147 (841-1500) | 5.88 (4.33-7.45) | 474 (334-643) | 2.43 (1.71-3.22) | 334 (221-472) | 1.70 (1.14-2.36) |
| Somalia | Male | 17682 (16234-18944) | 1852 (1294-2421) | 10.50 (7.56-13.46) | 1351 (944-1762) | 7.64 (5.50-9.87) | 819 (553-1103) | 4.63 (3.21-6.20) | 224 (147-327) | 1.27 (0.83-1.83) | 160 (99-235) | 0.90 (0.56-1.31) |
| South Africa | Female | 173143 (158882-187207) | 9896 (6447-13597) | 5.71 (3.74-7.82) | 7672 (5090-10313) | 4.45 (2.96-5.88) | 5756 (3944-7618) | 3.33 (2.30-4.38) | 2962 (1868-4121) | 1.71 (1.11-2.36) | 2694 (1694-3805) | 1.56 (1.00-2.16) |
| South Africa | Male | 131764 (119538-145022) | 7853 (4838-11211) | 5.96 (3.74-8.39) | 5973 (3745-8476) | 4.52 (2.87-6.30) | 4501 (2872-6272) | 3.42 (2.20-4.70) | 1376 (740-2172) | 1.04 (0.57-1.62) | 1242 (655-1983) | 0.94 (0.50-1.45) |
| South Korea | Female | 177240 (166787-187559) | 23397 (17655-28348) | 13.24 (10.02-15.78) | 10317 (7473-13161) | 5.83 (4.26-7.36) | 15405 (11453-19038) | 8.74 (6.46-10.70) | 4612 (3297-5928) | 2.60 (1.87-3.30) | 4599 (3304-5889) | 2.60 (1.88-3.31) |
| South Korea | Male | 159506 (149840-169185) | 26446 (19097-32078) | 16.65 (11.88-20.02) | 12022 (8328-15366) | 7.53 (5.29-9.59) | 17676 (12884-21907) | 11.11 (8.05-13.66) | 4583 (3163-6010) | 2.87 (1.98-3.74) | 4570 (3142-5933) | 2.86 (1.99-3.73) |
| Spain | Female | 210230 (198730-221340) | 4736 (2640-7106) | 2.25 (1.24-3.34) | 2981 (1522-4631) | 1.41 (0.72-2.23) | 3615 (1905-5526) | 1.72 (0.90-2.65) | 957 (358-1738) | 0.45 (0.17-0.83) | 816 (303-1504) | 0.39 (0.14-0.72) |
| Spain | Male | 226070 (214350-237876) | 16978 (12199-21724) | 7.50 (5.44-9.63) | 8723 (6118-11499) | 3.85 (2.74-5.04) | 11399 (7981-14882) | 5.01 (3.55-6.55) | 2545 (1631-3583) | 1.12 (0.73-1.58) | 2212 (1395-3165) | 0.98 (0.62-1.40) |
| Sri Lanka | Female | 75466 (70641-80142) | 11063 (9144-13122) | 14.67 (12.07-17.17) | 7289 (5894-8897) | 9.68 (7.87-11.53) | 4768 (3803-5868) | 6.32 (5.04-7.72) | 2420 (1875-3086) | 3.21 (2.51-4.06) | 2065 (1566-2718) | 2.75 (2.10-3.57) |
| Sri Lanka | Male | 68208 (63425-72868) | 12760 (10381-15051) | 18.74 (15.45-21.74) | 8512 (6843-10495) | 12.47 (10.18-14.98) | 5575 (4393-6966) | 8.14 (6.50-10.18) | 1893 (1408-2524) | 2.79 (2.08-3.68) | 1554 (1142-2116) | 2.27 (1.68-3.04) |
| Sudan | Female | 93026 (85163-100715) | 727 (246-1668) | 0.78 (0.26-1.80) | 659 (221-1495) | 0.70 (0.24-1.59) | 505 (154-1112) | 0.54 (0.17-1.19) | 84 (12-316) | 0.09 (0.01-0.33) | 58 (5-246) | 0.06 (0.01-0.26) |
| Sudan | Male | 149410 (136458-161009) | 10162 (6471-14143) | 6.79 (4.32-9.35) | 7932 (5130-11002) | 5.32 (3.48-7.20) | 4962 (3338-6805) | 3.34 (2.26-4.49) | 1000 (582-1566) | 0.67 (0.39-1.04) | 713 (387-1128) | 0.48 (0.26-0.74) |
| Suriname | Female | 2179 (1991-2366) | 160 (111-207) | 7.33 (5.18-9.33) | 113 (78-148) | 5.19 (3.68-6.67) | 76 (54-100) | 3.51 (2.55-4.57) | 52 (35-71) | 2.40 (1.63-3.22) | 44 (30-62) | 2.02 (1.37-2.77) |
| Suriname | Male | 2151 (1966-2372) | 251 (187-311) | 11.68 (8.80-14.10) | 170 (124-219) | 7.88 (5.86-9.76) | 114 (84-149) | 5.29 (3.98-6.79) | 57 (40-76) | 2.64 (1.90-3.48) | 47 (33-65) | 2.18 (1.55-2.89) |
| Swaziland | Female | 2020 (1839-2192) | 151 (101-199) | 7.47 (5.10-9.79) | 113 (77-146) | 5.60 (3.93-7.14) | 76 (53-101) | 3.80 (2.64-4.95) | 38 (25-53) | 1.89 (1.24-2.55) | 29 (17-41) | 1.42 (0.84-1.97) |
| Swaziland | Male | 1591 (1439-1742) | 137 (91-185) | 8.59 (5.86-11.45) | 100 (68-136) | 6.29 (4.25-8.39) | 67 (46-92) | 4.19 (2.88-5.73) | 16 (9-24) | 0.97 (0.56-1.47) | 12 (6-18) | 0.73 (0.39-1.13) |
| Sweden | Female | 62326 (57994-67116) | 3615 (2484-4676) | 5.81 (3.98-7.38) | 941 (593-1374) | 1.50 (0.94-2.18) | 3127 (2196-4104) | 5.04 (3.55-6.48) | 356 (211-541) | 0.57 (0.34-0.87) | 290 (172-453) | 0.47 (0.27-0.72) |
| Sweden | Male | 70299 (64953-75712) | 8112 (6486-9619) | 11.54 (9.28-13.37) | 1963 (1329-2723) | 2.79 (1.90-3.85) | 6748 (5312-8183) | 9.65 (7.71-11.43) | 709 (466-1038) | 1.01 (0.67-1.48) | 610 (391-899) | 0.86 (0.57-1.27) |
| Switzerland | Female | 36076 (33705-38237) | 1790 (1226-2361) | 4.96 (3.38-6.46) | 178 (105-331) | 0.49 (0.30-0.92) | 1718 (1169-2275) | 4.75 (3.22-6.23) | 62 (36-121) | 0.17 (0.10-0.33) | 51 (29-100) | 0.14 (0.08-0.28) |
| Switzerland | Male | 40676 (38109-43543) | 4452 (3465-5420) | 10.99 (8.61-13.08) | 402 (255-718) | 0.98 (0.63-1.79) | 4205 (3220-5142) | 10.37 (8.08-12.44) | 151 (92-287) | 0.37 (0.23-0.69) | 127 (78-242) | 0.31 (0.19-0.60) |
| Syria | Female | 67597 (61870-73438) | 282 (34-987) | 0.42 (0.05-1.46) | 270 (30-957) | 0.40 (0.04-1.42) | 229 (23-780) | 0.34 (0.04-1.14) | 74 (1-397) | 0.11 (0.00-0.58) | 60 (1-367) | 0.09 (0.00-0.53) |
| Syria | Male | 105760 (95835-115346) | 6637 (3813-9867) | 6.24 (3.67-9.14) | 5230 (3111-7387) | 4.92 (3.02-6.92) | 3254 (2063-4703) | 3.07 (1.98-4.40) | 1545 (836-2347) | 1.47 (0.81-2.20) | 1302 (700-2029) | 1.23 (0.66-1.91) |
| Taiwan (Province of China) | Female | 129813 (119259-139114) | 16442 (12815-19888) | 12.69 (10.00-15.01) | 7501 (5623-9432) | 5.78 (4.40-7.21) | 10657 (8090-13233) | 8.21 (6.32-10.05) | 2406 (1761-3194) | 1.86 (1.37-2.44) | 2396 (1770-3154) | 1.85 (1.38-2.42) |
| Taiwan (Province of China) | Male | 126596 (116873-136558) | 17139 (13325-20888) | 13.57 (10.81-16.24) | 7871 (5842-10106) | 6.22 (4.78-7.83) | 11207 (8505-14005) | 8.86 (6.80-10.87) | 2572 (1850-3393) | 2.03 (1.51-2.66) | 2492 (1790-3285) | 1.97 (1.45-2.56) |
| Tajikistan | Female | 24585 (22871-26436) | 2468 (1869-3088) | 10.02 (7.66-12.46) | 1782 (1355-2264) | 7.26 (5.59-9.11) | 1082 (833-1415) | 4.39 (3.42-5.63) | 643 (464-857) | 2.62 (1.92-3.43) | 544 (388-725) | 2.21 (1.58-2.94) |
| Tajikistan | Male | 31056 (28909-33403) | 5200 (4225-6215) | 16.76 (13.77-19.85) | 3608 (2866-4384) | 11.63 (9.34-13.91) | 2171 (1713-2714) | 6.98 (5.53-8.64) | 1047 (813-1331) | 3.36 (2.59-4.26) | 655 (477-888) | 2.11 (1.53-2.79) |
| Tanzania | Female | 76438 (70735-81876) | 11616 (9116-13830) | 15.17 (12.09-17.67) | 7995 (6177-9668) | 10.42 (8.14-12.37) | 4815 (3634-5982) | 6.30 (4.89-7.77) | 1673 (1205-2128) | 2.19 (1.62-2.76) | 722 (487-967) | 0.95 (0.66-1.27) |
| Tanzania | Male | 77538 (71657-83516) | 12278 (9273-14859) | 15.77 (12.27-18.64) | 8477 (6278-10445) | 10.90 (8.25-13.17) | 5158 (3771-6556) | 6.64 (5.00-8.25) | 1086 (762-1463) | 1.40 (0.99-1.87) | 652 (426-916) | 0.84 (0.57-1.17) |
| Thailand | Female | 249901 (235167-265720) | 31354 (22997-38886) | 12.54 (9.36-15.44) | 20129 (14749-25371) | 8.09 (5.99-10.07) | 14353 (10457-18288) | 5.76 (4.21-7.33) | 6563 (4840-8513) | 2.64 (1.94-3.37) | 5983 (4261-7768) | 2.40 (1.71-3.06) |
| Thailand | Male | 229148 (213272-243947) | 39426 (30191-47288) | 17.30 (13.41-20.33) | 25625 (19476-31630) | 11.24 (8.54-13.77) | 18365 (13668-23210) | 8.04 (6.01-9.93) | 6506 (4733-8812) | 2.84 (2.09-3.78) | 5645 (4126-7748) | 2.46 (1.81-3.28) |
| The Bahamas | Female | 1317 (1191-1435) | 101 (71-131) | 7.63 (5.35-9.78) | 54 (39-71) | 4.12 (3.00-5.33) | 67 (47-88) | 5.06 (3.72-6.53) | 23 (16-31) | 1.73 (1.22-2.27) | 20 (14-27) | 1.53 (1.07-2.08) |
| The Bahamas | Male | 1282 (1146-1400) | 160 (122-196) | 12.56 (9.72-14.98) | 81 (60-104) | 6.36 (4.82-7.98) | 100 (74-129) | 7.86 (5.98-9.82) | 27 (20-37) | 2.15 (1.59-2.82) | 22 (16-29) | 1.71 (1.25-2.28) |
| The Gambia | Female | 3289 (3018-3568) | 315 (243-395) | 9.63 (7.49-11.77) | 227 (175-285) | 6.93 (5.37-8.50) | 136 (104-173) | 4.15 (3.20-5.23) | 66 (48-88) | 2.00 (1.44-2.64) | 42 (28-60) | 1.29 (0.86-1.77) |
| The Gambia | Male | 3379 (3085-3707) | 369 (277-459) | 10.90 (8.34-13.23) | 263 (194-334) | 7.78 (5.74-9.68) | 158 (116-204) | 4.68 (3.45-5.91) | 48 (33-67) | 1.43 (0.98-1.97) | 32 (21-47) | 0.95 (0.63-1.37) |
| Timor-Leste | Female | 2202 (2063-2342) | 324 (257-388) | 14.69 (11.90-17.33) | 216 (170-264) | 9.82 (7.90-11.84) | 137 (105-173) | 6.22 (4.90-7.69) | 40 (29-55) | 1.83 (1.33-2.46) | 27 (20-39) | 1.24 (0.88-1.72) |
| Timor-Leste | Male | 2422 (2248-2605) | 471 (385-556) | 19.44 (16.10-22.57) | 317 (255-389) | 13.04 (10.67-15.86) | 202 (158-255) | 8.32 (6.70-10.42) | 59 (42-79) | 2.42 (1.79-3.20) | 38 (27-52) | 1.56 (1.12-2.14) |
| Togo | Female | 12921 (11813-13974) | 1215 (903-1540) | 9.38 (7.00-11.65) | 873 (646-1125) | 6.74 (5.01-8.56) | 524 (385-694) | 4.09 (3.00-5.27) | 220 (152-312) | 1.70 (1.19-2.39) | 118 (77-183) | 0.92 (0.61-1.39) |
| Togo | Male | 11773 (10650-12777) | 1279 (949-1609) | 11.01 (8.07-13.44) | 916 (682-1170) | 7.85 (5.80-9.79) | 555 (396-731) | 4.72 (3.42-6.12) | 131 (84-193) | 1.12 (0.73-1.63) | 62 (38-96) | 0.53 (0.33-0.80) |
| Tonga | Female | 291 (266-315) | 38 (30-46) | 13.12 (10.71-15.43) | 25 (20-31) | 8.77 (7.00-10.52) | 17 (13-21) | 5.81 (4.59-7.15) | 9 (6-12) | 2.98 (2.26-3.85) | 7 (5-10) | 2.47 (1.83-3.24) |
| Tonga | Male | 277 (249-306) | 43 (34-53) | 15.72 (12.67-18.35) | 29 (22-36) | 10.51 (8.28-12.70) | 19 (15-25) | 6.91 (5.44-8.62) | 6 (4-8) | 2.18 (1.64-2.86) | 5 (4-7) | 1.77 (1.29-2.41) |
| Trinidad and Tobago | Female | 6416 (5848-7076) | 489 (347-641) | 7.63 (5.52-9.82) | 307 (226-398) | 4.78 (3.57-6.09) | 278 (199-361) | 4.33 (3.16-5.53) | 132 (91-179) | 2.06 (1.45-2.72) | 108 (73-152) | 1.69 (1.17-2.32) |
| Trinidad and Tobago | Male | 6652 (6000-7253) | 872 (693-1036) | 13.14 (10.63-15.41) | 517 (397-644) | 7.81 (6.22-9.58) | 465 (355-583) | 6.98 (5.43-8.61) | 175 (131-231) | 2.63 (2.02-3.39) | 146 (104-197) | 2.18 (1.61-2.89) |
| Tunisia | Female | 62129 (55975-67941) | 246 (41-822) | 0.40 (0.06-1.33) | 225 (38-787) | 0.37 (0.06-1.24) | 196 (27-605) | 0.32 (0.04-0.97) | 49 (0-277) | 0.08 (0.00-0.45) | 39 (0-227) | 0.06 (0.00-0.37) |
| Tunisia | Male | 81927 (74680-90113) | 4692 (2828-7048) | 5.76 (3.43-8.42) | 3656 (2160-5249) | 4.43 (2.64-6.31) | 2494 (1543-3531) | 3.04 (1.91-4.21) | 797 (397-1239) | 0.97 (0.47-1.48) | 673 (337-1060) | 0.82 (0.42-1.29) |
| Turkey | Female | 367953 (337841-398060) | 6 (0-777) | 0.00 (0.00-0.22) | 6 (0-777) | 0.00 (0.00-0.22) | 6 (0-763) | 0.00 (0.00-0.21) | 0 (0-238) | 0.00 (0.00-0.06) | 0 (0-213) | 0.00 (0.00-0.06) |
| Turkey | Male | 468093 (427384-506051) | 15578 (7331-27648) | 3.33 (1.55-5.75) | 12683 (6285-21088) | 2.74 (1.35-4.47) | 10490 (5138-17731) | 2.26 (1.12-3.75) | 2963 (1150-5430) | 0.63 (0.24-1.15) | 2879 (1083-5335) | 0.61 (0.23-1.13) |
| Turkmenistan | Female | 19251 (17798-20617) | 1941 (1496-2430) | 10.09 (7.84-12.55) | 1309 (994-1648) | 6.82 (5.24-8.53) | 960 (733-1214) | 5.01 (3.87-6.26) | 562 (416-736) | 2.93 (2.20-3.74) | 488 (360-645) | 2.54 (1.90-3.29) |
| Turkmenistan | Male | 20919 (19233-22476) | 3578 (2848-4274) | 17.22 (13.90-19.97) | 2327 (1825-2813) | 11.11 (8.86-13.25) | 1682 (1282-2072) | 8.05 (6.26-9.81) | 661 (496-829) | 3.17 (2.44-3.92) | 484 (361-623) | 2.32 (1.76-2.92) |
| Tuvalu | Female | 38 (35-41) | 5 (4-6) | 13.20 (10.77-15.73) | 3 (3-4) | 8.82 (6.93-10.60) | 2 (2-3) | 5.98 (4.72-7.47) | 1 (1-1) | 2.26 (1.58-3.04) | 1 (0-1) | 1.34 (0.90-1.90) |
| Tuvalu | Male | 37 (34-41) | 6 (5-7) | 15.70 (12.34-18.44) | 4 (3-5) | 10.44 (8.06-12.74) | 3 (2-3) | 7.16 (5.63-8.87) | 1 (1-1) | 2.16 (1.56-2.91) | 1 (0-1) | 1.52 (1.00-2.22) |
| Uganda | Female | 44078 (40865-47154) | 6161 (4814-7393) | 14.07 (11.07-16.49) | 4296 (3302-5195) | 9.80 (7.66-11.71) | 2581 (1932-3144) | 5.87 (4.51-7.11) | 788 (567-1037) | 1.79 (1.30-2.31) | 469 (321-659) | 1.06 (0.74-1.45) |
| Uganda | Male | 38304 (34946-41304) | 4104 (2984-5192) | 10.72 (7.98-13.37) | 2959 (2170-3765) | 7.74 (5.70-9.67) | 1792 (1288-2373) | 4.68 (3.44-6.04) | 355 (241-493) | 0.93 (0.63-1.27) | 201 (129-294) | 0.53 (0.34-0.75) |
| Ukraine | Female | 486258 (434036-534021) | 16706 (8479-27548) | 3.46 (1.75-5.74) | 14560 (7702-23125) | 3.03 (1.58-4.72) | 11127 (6135-16658) | 2.32 (1.29-3.41) | 7348 (3524-12251) | 1.53 (0.72-2.50) | 6257 (3009-10561) | 1.31 (0.62-2.14) |
| Ukraine | Male | 363930 (326907-400309) | 43237 (33192-53747) | 11.91 (9.37-14.37) | 29576 (22475-37454) | 8.13 (6.34-9.95) | 19602 (15144-25259) | 5.41 (4.23-6.82) | 9737 (6855-13014) | 2.68 (1.97-3.49) | 6779 (4750-9259) | 1.87 (1.36-2.50) |
| United Arab Emirates | Female | 13245 (11754-14735) | 95 (12-255) | 0.73 (0.09-1.96) | 64 (7-164) | 0.48 (0.06-1.23) | 89 (11-234) | 0.67 (0.08-1.76) | 9 (0-32) | 0.07 (0.00-0.24) | 7 (0-29) | 0.05 (0.00-0.21) |
| United Arab Emirates | Male | 67110 (58934-75866) | 5123 (2836-7736) | 7.70 (4.26-10.96) | 2232 (1281-3428) | 3.33 (1.93-4.98) | 4108 (2336-6123) | 6.18 (3.43-8.80) | 382 (197-603) | 0.58 (0.30-0.86) | 331 (178-524) | 0.50 (0.26-0.75) |
| United Kingdom | Female | 279354 (260630-296037) | 12113 (7452-16993) | 4.34 (2.74-6.06) | 5048 (3320-6903) | 1.81 (1.17-2.47) | 9935 (6351-13709) | 3.56 (2.29-4.93) | 1751 (1100-2473) | 0.63 (0.40-0.88) | 1580 (994-2198) | 0.56 (0.36-0.78) |
| United Kingdom | Male | 297247 (278061-316389) | 26022 (18995-32095) | 8.75 (6.47-10.73) | 9357 (6552-12442) | 3.16 (2.23-4.13) | 19594 (14359-25150) | 6.58 (4.86-8.20) | 3204 (2152-4395) | 1.08 (0.73-1.45) | 2842 (1906-3935) | 0.96 (0.65-1.31) |
| United States | Female | 1597354 (1512288-1681889) | 108075 (77831-133818) | 6.75 (4.88-8.35) | 19909 (11591-31772) | 1.25 (0.72-1.99) | 95523 (68812-121211) | 5.99 (4.33-7.56) | 8755 (4806-14386) | 0.55 (0.30-0.91) | 8117 (4491-13193) | 0.51 (0.28-0.83) |
| United States | Male | 1751085 (1664951-1835902) | 186440 (139826-225072) | 10.63 (8.07-12.75) | 34681 (20490-53982) | 1.98 (1.17-3.08) | 163637 (121880-199961) | 9.36 (6.92-11.35) | 14046 (7923-22791) | 0.80 (0.45-1.29) | 13022 (7231-21439) | 0.74 (0.41-1.20) |
| Uruguay | Female | 14562 (13588-15413) | 1530 (1240-1800) | 10.52 (8.56-12.28) | 876 (689-1085) | 6.03 (4.79-7.33) | 832 (651-1025) | 5.73 (4.52-6.95) | 457 (348-577) | 3.14 (2.41-3.92) | 434 (331-549) | 2.99 (2.29-3.73) |
| Uruguay | Male | 13388 (12300-14391) | 1971 (1567-2383) | 14.76 (11.90-17.43) | 1125 (886-1400) | 8.41 (6.76-10.25) | 1072 (832-1328) | 8.03 (6.26-9.74) | 486 (367-621) | 3.64 (2.85-4.57) | 420 (310-539) | 3.15 (2.42-4.03) |
| Uzbekistan | Female | 170530 (162424-178459) | 19071 (15103-23460) | 11.17 (8.90-13.72) | 13478 (10552-16612) | 7.91 (6.15-9.70) | 8379 (6458-10453) | 4.90 (3.79-6.07) | 5421 (4023-7209) | 3.19 (2.35-4.18) | 4667 (3341-6189) | 2.74 (1.98-3.61) |
| Uzbekistan | Male | 169431 (160095-179126) | 28598 (23161-33852) | 16.90 (13.69-19.68) | 19520 (15510-23907) | 11.53 (9.16-13.79) | 12099 (9394-14815) | 7.14 (5.57-8.72) | 5540 (4138-7136) | 3.28 (2.46-4.16) | 4345 (3217-5670) | 2.56 (1.92-3.30) |
| Vanuatu | Female | 655 (606-700) | 85 (67-103) | 13.10 (10.67-15.39) | 58 (46-72) | 8.92 (7.22-10.70) | 37 (28-47) | 5.73 (4.44-7.07) | 14 (10-19) | 2.12 (1.50-2.86) | 7 (4-10) | 1.00 (0.63-1.50) |
| Vanuatu | Male | 761 (694-828) | 118 (92-145) | 15.50 (12.46-18.61) | 81 (61-103) | 10.57 (8.31-13.11) | 52 (39-68) | 6.81 (5.33-8.71) | 12 (8-17) | 1.54 (1.06-2.19) | 6 (4-9) | 0.80 (0.49-1.20) |
| Venezuela | Female | 83240 (78088-88762) | 9639 (7640-11673) | 11.62 (9.39-13.78) | 6674 (5096-8296) | 8.00 (6.30-9.89) | 4124 (3168-5224) | 4.96 (3.86-6.13) | 3261 (2509-4161) | 3.92 (3.04-4.87) | 3054 (2369-3904) | 3.68 (2.86-4.61) |
| Venezuela | Male | 91423 (84966-97506) | 15889 (12494-19177) | 17.40 (13.94-20.60) | 10737 (8437-13128) | 11.78 (9.30-14.12) | 6673 (5082-8414) | 7.31 (5.58-9.01) | 3940 (3063-4966) | 4.34 (3.37-5.39) | 3591 (2737-4515) | 3.93 (3.02-4.88) |
| Vietnam | Female | 279166 (262497-295280) | 39754 (32361-47431) | 14.24 (11.54-16.85) | 26351 (20950-31912) | 9.44 (7.59-11.32) | 17158 (13417-21019) | 6.17 (4.81-7.45) | 7053 (5477-8996) | 2.53 (1.97-3.22) | 4795 (3579-6332) | 1.72 (1.30-2.23) |
| Vietnam | Male | 308188 (291946-323813) | 61950 (50207-72881) | 20.12 (16.53-23.43) | 41582 (33198-50441) | 13.47 (10.93-16.27) | 27027 (21192-33700) | 8.81 (6.85-10.79) | 9588 (7282-12344) | 3.12 (2.39-3.98) | 6640 (4955-8689) | 2.15 (1.62-2.82) |
| Yemen | Female | 70045 (64438-75805) | 413 (65-1081) | 0.59 (0.09-1.53) | 371 (62-985) | 0.54 (0.09-1.39) | 295 (42-758) | 0.42 (0.06-1.07) | 42 (2-191) | 0.06 (0.00-0.27) | 33 (2-162) | 0.05 (0.00-0.23) |
| Yemen | Male | 98138 (89285-106028) | 6224 (3836-8643) | 6.30 (3.92-8.87) | 4864 (3074-6681) | 4.96 (3.20-6.82) | 3015 (1918-4222) | 3.07 (2.00-4.26) | 793 (431-1287) | 0.81 (0.44-1.28) | 671 (354-1066) | 0.68 (0.37-1.07) |
| Zambia | Female | 20737 (19336-22237) | 2920 (2282-3533) | 14.05 (11.24-16.80) | 2036 (1559-2527) | 9.81 (7.70-12.04) | 1231 (931-1537) | 5.94 (4.60-7.34) | 541 (403-693) | 2.61 (1.96-3.35) | 322 (231-447) | 1.56 (1.12-2.10) |
| Zambia | Male | 21483 (19951-23283) | 2291 (1681-2907) | 10.71 (7.78-13.27) | 1651 (1194-2102) | 7.64 (5.61-9.71) | 1012 (722-1343) | 4.72 (3.42-6.08) | 273 (189-372) | 1.28 (0.90-1.71) | 161 (107-227) | 0.75 (0.50-1.05) |
| Zimbabwe | Female | 24962 (22961-26910) | 2564 (1897-3194) | 10.29 (7.80-12.56) | 1819 (1350-2287) | 7.27 (5.51-9.03) | 1115 (833-1451) | 4.48 (3.32-5.82) | 548 (377-746) | 2.20 (1.53-2.95) | 438 (281-631) | 1.76 (1.13-2.50) |
| Zimbabwe | Male | 19324 (17571-21085) | 2329 (1729-2924) | 12.06 (9.21-14.67) | 1641 (1193-2115) | 8.49 (6.25-10.75) | 998 (731-1336) | 5.18 (3.83-6.70) | 299 (200-431) | 1.56 (1.05-2.21) | 226 (145-348) | 1.17 (0.76-1.76) |

#### d. **Prevented CVD DALYs**

| **Country** | **Sex** | **Baseline disease burden Median (95% UI)** | **Replacement of discretionary salt and non-discretionary salt** | | **Replacement of discretionary salt** | | **Replacement of non-discretionary salt** | | **Replacement of discretionary salt among those with diagnosed hypertension** | | **Replacement of discretionary salt among those with treated hypertension** | |
| --- | --- | --- | --- | --- | --- | --- | --- | --- | --- | --- | --- | --- |
|  |  |  | **No. of events averted Median (95% UI)** | **Percentage of national disease burden averted % (95% UI)** | **No. of events averted Median (95% UI)** | **Percentage of national disease burden averted % (95% UI)** | **No. of events averted Median (95% UI)** | **Percentage of national disease burden averted % (95% UI)** | **No. of events averted Median (95% UI)** | **Percentage of national disease burden averted % (95% UI)** | **No. of events averted Median (95% UI)** | **Percentage of national disease burden averted % (95% UI)** |
| **Global, millions** | **Female** | 162.82 (160.68-165.01) | 23.70 (21.77-25.31) | 14.56 (13.41-15.54) | 15.06 (13.71-16.41) | 9.25 (8.42-10.08) | 12.08 (10.96-13.19) | 7.42 (6.72-8.08) | 4.63 (4.21-5.08) | 2.84 (2.59-3.12) | 3.75 (3.39-4.14) | 2.31 (2.09-2.53) |
| **Global, millions** | **Male** | 218.01 (214.90-221.01) | 41.25 (38.02-43.99) | 18.90 (17.44-20.10) | 26.43 (24.05-28.71) | 12.13 (11.04-13.13) | 21.63 (19.58-23.59) | 9.92 (9.01-10.77) | 6.29 (5.71-6.97) | 2.89 (2.63-3.19) | 4.97 (4.45-5.51) | 2.28 (2.04-2.52) |
| Afghanistan | Female | 565558 (512623-619034) | 5667 (986-13501) | 1.01 (0.18-2.36) | 5045 (978-12061) | 0.90 (0.18-2.13) | 3740 (745-9270) | 0.66 (0.13-1.62) | 1053 (148-3241) | 0.19 (0.03-0.57) | 860 (126-2687) | 0.15 (0.02-0.47) |
| Afghanistan | Male | 636832 (587326-692706) | 53772 (31583-77338) | 8.47 (4.98-12.02) | 41627 (24201-59970) | 6.53 (3.83-9.29) | 25517 (15347-37563) | 4.00 (2.39-5.76) | 6528 (3741-9678) | 1.02 (0.59-1.52) | 5166 (2917-7780) | 0.81 (0.46-1.21) |
| Albania | Female | 100885 (95906-105800) | 20982 (17235-24662) | 20.80 (17.09-24.07) | 13594 (10883-16470) | 13.45 (11.00-16.31) | 9534 (7520-11709) | 9.44 (7.49-11.60) | 6466 (4959-8449) | 6.42 (4.99-8.35) | 5755 (4304-7478) | 5.70 (4.31-7.32) |
| Albania | Male | 116142 (110511-122242) | 29814 (25054-34498) | 25.70 (21.82-29.31) | 20210 (16378-24295) | 17.42 (14.28-20.63) | 14351 (11497-17452) | 12.34 (9.89-15.10) | 4841 (3565-6391) | 4.19 (3.07-5.45) | 3580 (2471-4997) | 3.07 (2.16-4.30) |
| Algeria | Female | 903038 (843202-962211) | 5808 (741-15093) | 0.64 (0.08-1.65) | 5247 (725-14042) | 0.58 (0.08-1.52) | 4373 (525-11452) | 0.48 (0.06-1.26) | 1034 (27-4482) | 0.11 (0.00-0.50) | 825 (13-3976) | 0.09 (0.00-0.45) |
| Algeria | Male | 988251 (929244-1045488) | 67337 (42417-93567) | 6.80 (4.28-9.42) | 52013 (33584-71690) | 5.27 (3.38-7.19) | 35443 (23508-49120) | 3.59 (2.39-4.93) | 10792 (6055-16540) | 1.10 (0.62-1.66) | 8857 (4762-13841) | 0.89 (0.48-1.39) |
| American Samoa | Female | 1282 (1211-1350) | 110 (79-144) | 8.61 (6.14-11.26) | 73 (51-94) | 5.69 (4.00-7.34) | 67 (48-88) | 5.24 (3.69-6.75) | 24 (16-32) | 1.88 (1.25-2.53) | 18 (12-26) | 1.43 (0.92-2.02) |
| American Samoa | Male | 2098 (1985-2210) | 192 (129-256) | 9.22 (6.23-12.12) | 127 (86-167) | 6.07 (4.12-7.90) | 117 (79-155) | 5.58 (3.82-7.35) | 35 (23-48) | 1.68 (1.09-2.30) | 25 (16-36) | 1.19 (0.77-1.69) |
| Andorra | Female | 1171 (1097-1247) | 55 (31-82) | 4.69 (2.60-7.01) | 31 (18-45) | 2.63 (1.51-3.84) | 47 (27-71) | 4.05 (2.35-5.98) | 13 (6-21) | 1.09 (0.54-1.75) | 12 (6-19) | 0.99 (0.49-1.59) |
| Andorra | Male | 1184 (1101-1265) | 137 (109-166) | 11.50 (9.08-13.82) | 54 (42-70) | 4.61 (3.52-5.81) | 99 (77-123) | 8.38 (6.53-10.23) | 21 (15-29) | 1.78 (1.28-2.47) | 19 (13-27) | 1.63 (1.15-2.21) |
| Angola | Female | 358265 (333527-384367) | 33718 (22336-46295) | 9.39 (6.20-12.85) | 25995 (17463-35517) | 7.26 (4.90-9.72) | 16867 (11417-23068) | 4.71 (3.15-6.31) | 7431 (4601-10923) | 2.07 (1.27-3.03) | 3474 (1930-5632) | 0.97 (0.53-1.58) |
| Angola | Male | 380643 (352472-408724) | 36830 (25872-49412) | 9.70 (6.78-12.88) | 27867 (19669-37196) | 7.30 (5.16-9.66) | 17686 (12573-24087) | 4.68 (3.32-6.19) | 5018 (3109-7518) | 1.33 (0.82-1.98) | 3008 (1712-4782) | 0.79 (0.45-1.26) |
| Antigua and Barbuda | Female | 1720 (1685-1755) | 192 (144-247) | 11.15 (8.27-14.35) | 120 (89-154) | 6.97 (5.17-8.88) | 112 (84-145) | 6.52 (4.91-8.39) | 56 (40-74) | 3.25 (2.33-4.29) | 47 (32-65) | 2.73 (1.87-3.75) |
| Antigua and Barbuda | Male | 2025 (1956-2092) | 309 (240-374) | 15.28 (11.82-18.34) | 184 (137-231) | 9.04 (6.77-11.39) | 171 (128-216) | 8.46 (6.32-10.56) | 65 (46-90) | 3.20 (2.26-4.40) | 54 (36-74) | 2.65 (1.78-3.68) |
| Argentina | Female | 600701 (585866-614417) | 86551 (71172-100501) | 14.39 (11.91-16.70) | 54671 (44439-65902) | 9.13 (7.40-10.93) | 43110 (34217-52857) | 7.18 (5.68-8.80) | 25236 (19984-31716) | 4.21 (3.34-5.25) | 22017 (17267-28258) | 3.67 (2.88-4.67) |
| Argentina | Male | 800064 (782982-816741) | 139893 (115456-166136) | 17.54 (14.42-20.73) | 88414 (71438-107219) | 11.06 (8.99-13.39) | 69752 (55531-86387) | 8.74 (6.98-10.71) | 36452 (28489-45783) | 4.57 (3.55-5.72) | 30247 (24009-38083) | 3.78 (3.00-4.77) |
| Armenia | Female | 104064 (100859-106935) | 12552 (9841-15765) | 12.08 (9.46-15.07) | 8603 (6521-10951) | 8.28 (6.29-10.48) | 5862 (4459-7560) | 5.64 (4.27-7.21) | 3466 (2406-4757) | 3.33 (2.34-4.61) | 3055 (2145-4262) | 2.94 (2.06-4.12) |
| Armenia | Male | 137741 (133824-141563) | 23617 (19424-27571) | 17.16 (14.10-19.91) | 15727 (12618-18766) | 11.40 (9.23-13.54) | 10596 (8428-13068) | 7.68 (6.10-9.42) | 4279 (3185-5615) | 3.11 (2.31-4.05) | 2844 (2143-3794) | 2.06 (1.56-2.74) |
| Australia | Female | 297511 (286575-307293) | 9417 (4815-15297) | 3.16 (1.63-5.14) | 3687 (1771-6173) | 1.23 (0.59-2.06) | 8831 (4430-14565) | 2.97 (1.51-4.87) | 1355 (525-2432) | 0.45 (0.18-0.82) | 1201 (463-2163) | 0.41 (0.16-0.72) |
| Australia | Male | 407363 (398536-416556) | 29622 (21123-39299) | 7.28 (5.22-9.64) | 8677 (5563-12405) | 2.13 (1.37-3.03) | 26070 (18448-34005) | 6.41 (4.53-8.40) | 2733 (1628-4214) | 0.67 (0.40-1.03) | 2452 (1436-3762) | 0.60 (0.35-0.92) |
| Austria | Female | 195719 (188796-202336) | 25707 (20394-30959) | 13.16 (10.48-15.70) | 7896 (5187-10829) | 4.04 (2.64-5.51) | 20324 (15596-25234) | 10.39 (7.99-12.80) | 3650 (2266-5418) | 1.87 (1.16-2.75) | 3284 (2030-4976) | 1.68 (1.04-2.52) |
| Austria | Male | 218730 (213657-223806) | 36711 (30313-43128) | 16.80 (13.93-19.75) | 11040 (8039-14574) | 5.05 (3.69-6.68) | 28777 (23410-34548) | 13.17 (10.71-15.84) | 4844 (3347-6802) | 2.21 (1.53-3.12) | 4685 (3276-6515) | 2.15 (1.51-3.00) |
| Azerbaijan | Female | 328319 (314188-342442) | 37435 (28344-47030) | 11.46 (8.59-14.33) | 26157 (19656-32874) | 7.99 (6.03-9.96) | 17886 (13361-23063) | 5.46 (4.09-6.98) | 11302 (8106-14607) | 3.44 (2.48-4.48) | 9444 (6688-12812) | 2.87 (2.03-3.90) |
| Azerbaijan | Male | 402857 (385286-419895) | 71908 (57631-84912) | 17.83 (14.37-20.94) | 47870 (37438-58086) | 11.87 (9.30-14.30) | 32680 (26161-40723) | 8.09 (6.42-10.07) | 13520 (10023-17565) | 3.36 (2.51-4.34) | 9694 (7134-12862) | 2.41 (1.79-3.19) |
| Bahrain | Female | 11521 (10924-12103) | 84 (18-238) | 0.73 (0.15-2.05) | 68 (14-192) | 0.59 (0.12-1.67) | 74 (16-210) | 0.63 (0.14-1.82) | 15 (0-66) | 0.13 (0.00-0.57) | 12 (0-61) | 0.11 (0.00-0.53) |
| Bahrain | Male | 23908 (22791-25162) | 2128 (1417-2873) | 8.88 (5.92-11.97) | 1236 (821-1632) | 5.16 (3.43-6.83) | 1448 (965-1945) | 6.06 (4.08-8.12) | 243 (146-350) | 1.02 (0.61-1.47) | 201 (115-300) | 0.84 (0.48-1.25) |
| Bangladesh | Female | 3141413 (2941648-3335031) | 479944 (388923-563812) | 15.32 (12.54-17.77) | 335018 (264244-409781) | 10.69 (8.46-12.86) | 209869 (165383-259365) | 6.69 (5.30-8.22) | 79597 (60928-100310) | 2.54 (1.96-3.17) | 63791 (48414-80845) | 2.02 (1.55-2.55) |
| Bangladesh | Male | 4542770 (4243860-4836136) | 706365 (560700-825840) | 15.53 (12.58-18.02) | 493341 (385958-595186) | 10.91 (8.66-12.94) | 311412 (236318-378491) | 6.86 (5.27-8.41) | 70979 (54097-89004) | 1.56 (1.19-1.95) | 55552 (41852-70413) | 1.22 (0.93-1.54) |
| Barbados | Female | 7663 (7383-7935) | 455 (277-668) | 5.92 (3.63-8.67) | 334 (216-456) | 4.37 (2.80-5.92) | 315 (205-437) | 4.13 (2.67-5.67) | 178 (107-249) | 2.33 (1.41-3.26) | 165 (101-233) | 2.15 (1.32-3.05) |
| Barbados | Male | 7913 (7562-8277) | 894 (666-1106) | 11.28 (8.45-13.93) | 530 (395-674) | 6.68 (5.02-8.49) | 494 (367-634) | 6.26 (4.69-8.06) | 218 (160-292) | 2.76 (2.02-3.69) | 195 (139-263) | 2.46 (1.77-3.32) |
| Belarus | Female | 632333 (607769-657711) | 25073 (13395-39589) | 3.97 (2.11-6.20) | 21403 (11549-32097) | 3.39 (1.85-5.04) | 16940 (9910-25048) | 2.69 (1.55-3.93) | 12212 (6352-19383) | 1.93 (1.00-3.02) | 9834 (5119-15814) | 1.56 (0.81-2.50) |
| Belarus | Male | 736140 (700409-769450) | 94681 (72672-115934) | 12.83 (9.90-15.54) | 62351 (48060-78204) | 8.46 (6.59-10.59) | 44493 (34334-57079) | 6.06 (4.70-7.78) | 26222 (19187-34387) | 3.56 (2.60-4.61) | 17887 (12835-24270) | 2.44 (1.74-3.28) |
| Belgium | Female | 165993 (160293-171763) | 18497 (14876-22018) | 11.12 (9.04-13.14) | 5838 (4197-7935) | 3.51 (2.53-4.76) | 14499 (11422-17708) | 8.73 (6.88-10.58) | 2715 (1869-3842) | 1.65 (1.14-2.29) | 2602 (1791-3626) | 1.57 (1.08-2.18) |
| Belgium | Male | 202419 (197605-207359) | 30020 (24999-35308) | 14.82 (12.43-17.36) | 9506 (7068-12268) | 4.69 (3.48-6.11) | 23321 (19075-27782) | 11.51 (9.46-13.68) | 3876 (2790-5260) | 1.92 (1.38-2.59) | 3813 (2762-5118) | 1.88 (1.36-2.54) |
| Belize | Female | 4061 (3946-4177) | 403 (281-512) | 9.93 (7.00-12.67) | 286 (203-364) | 7.04 (4.98-9.02) | 201 (142-264) | 4.96 (3.51-6.50) | 111 (76-144) | 2.73 (1.85-3.54) | 90 (60-123) | 2.20 (1.50-3.02) |
| Belize | Male | 5281 (5117-5455) | 798 (618-962) | 15.10 (11.75-18.21) | 538 (412-666) | 10.17 (7.84-12.59) | 376 (283-478) | 7.11 (5.39-9.01) | 142 (102-185) | 2.70 (1.94-3.51) | 112 (80-151) | 2.12 (1.51-2.87) |
| Benin | Female | 122794 (115248-129967) | 19010 (15209-22185) | 15.49 (12.66-17.79) | 13341 (10367-16157) | 10.85 (8.50-12.95) | 8087 (6216-10103) | 6.61 (5.17-8.12) | 2961 (2128-3815) | 2.41 (1.76-3.07) | 2054 (1411-2714) | 1.68 (1.15-2.19) |
| Benin | Male | 131321 (123163-139180) | 16383 (12165-20311) | 12.55 (9.26-15.27) | 11717 (8545-14825) | 8.95 (6.59-11.16) | 7206 (5122-9263) | 5.49 (3.96-6.97) | 1342 (909-1804) | 1.02 (0.71-1.37) | 960 (651-1310) | 0.73 (0.51-0.98) |
| Bermuda | Female | 1185 (1135-1233) | 113 (84-140) | 9.53 (7.21-11.78) | 8 (6-14) | 0.67 (0.48-1.14) | 109 (81-135) | 9.17 (6.93-11.35) | 4 (3-7) | 0.33 (0.22-0.59) | 3 (2-6) | 0.28 (0.18-0.52) |
| Bermuda | Male | 2062 (1989-2133) | 275 (219-325) | 13.28 (10.60-15.72) | 19 (14-31) | 0.91 (0.68-1.52) | 262 (208-313) | 12.76 (10.17-15.12) | 7 (5-12) | 0.32 (0.23-0.59) | 6 (4-10) | 0.28 (0.18-0.50) |
| Bhutan | Female | 10295 (9598-11055) | 1515 (1176-1817) | 14.76 (11.56-17.41) | 1052 (793-1296) | 10.20 (7.83-12.43) | 678 (512-870) | 6.57 (4.96-8.33) | 308 (228-397) | 2.99 (2.24-3.82) | 231 (166-308) | 2.25 (1.63-2.94) |
| Bhutan | Male | 14863 (13826-15901) | 2306 (1833-2740) | 15.55 (12.37-18.13) | 1590 (1258-1941) | 10.73 (8.46-12.96) | 1034 (789-1289) | 6.97 (5.32-8.57) | 348 (264-445) | 2.34 (1.77-2.97) | 222 (165-295) | 1.50 (1.10-1.96) |
| Bolivia | Female | 145573 (133324-157360) | 17680 (12866-22243) | 12.15 (8.98-14.94) | 12165 (8703-15716) | 8.38 (6.05-10.53) | 7885 (5411-10272) | 5.40 (3.84-6.96) | 4224 (2903-5747) | 2.92 (2.02-3.90) | 3761 (2482-5223) | 2.58 (1.73-3.54) |
| Bolivia | Male | 152643 (140393-164381) | 24507 (17863-30861) | 16.05 (11.95-19.81) | 16807 (12149-21416) | 11.04 (8.11-13.73) | 10911 (7867-14115) | 7.16 (5.18-9.08) | 4180 (2871-5829) | 2.76 (1.90-3.81) | 3443 (2292-4925) | 2.26 (1.49-3.16) |
| Bosnia and Herzegovina | Female | 149501 (142345-156511) | 30703 (25339-35983) | 20.57 (17.14-23.98) | 19713 (15793-23923) | 13.19 (10.62-15.91) | 14248 (11201-17590) | 9.54 (7.47-11.79) | 11165 (8654-13863) | 7.46 (5.80-9.31) | 10281 (7920-12953) | 6.87 (5.35-8.65) |
| Bosnia and Herzegovina | Male | 156160 (147590-164216) | 39794 (33631-45494) | 25.47 (21.70-28.66) | 26689 (22069-31635) | 17.07 (14.18-20.17) | 19231 (15596-23298) | 12.30 (10.01-14.86) | 10934 (8536-13784) | 6.99 (5.52-8.74) | 10098 (7809-12985) | 6.45 (4.98-8.26) |
| Botswana | Female | 30385 (28508-32470) | 2936 (2007-3926) | 9.66 (6.69-12.75) | 2156 (1510-2853) | 7.08 (4.90-9.29) | 1586 (1120-2105) | 5.25 (3.71-6.90) | 777 (514-1093) | 2.55 (1.70-3.63) | 642 (417-939) | 2.12 (1.40-3.06) |
| Botswana | Male | 34211 (31993-36547) | 3426 (2409-4521) | 10.03 (7.02-13.00) | 2467 (1733-3264) | 7.22 (5.14-9.39) | 1809 (1273-2436) | 5.27 (3.73-7.05) | 561 (340-812) | 1.64 (0.99-2.35) | 385 (223-581) | 1.13 (0.65-1.69) |
| Brazil | Female | 3332610 (3283152-3381390) | 461895 (367541-548637) | 13.85 (11.08-16.48) | 303807 (237521-375314) | 9.11 (7.13-11.22) | 221388 (169899-273603) | 6.63 (5.09-8.23) | 146005 (113131-183183) | 4.38 (3.40-5.49) | 141382 (109827-177969) | 4.24 (3.31-5.33) |
| Brazil | Male | 4387537 (4330427-4435838) | 789585 (650076-923959) | 18.03 (14.72-21.18) | 520125 (418333-617831) | 11.85 (9.57-14.07) | 376645 (303282-454807) | 8.58 (6.91-10.38) | 204718 (162564-248254) | 4.67 (3.71-5.67) | 190174 (150765-230979) | 4.33 (3.44-5.28) |
| Brunei | Female | 4407 (4209-4606) | 829 (642-1005) | 18.75 (14.64-22.49) | 395 (293-492) | 8.97 (6.67-11.21) | 530 (401-659) | 12.07 (9.16-14.89) | 167 (123-214) | 3.79 (2.77-4.81) | 155 (114-198) | 3.54 (2.61-4.47) |
| Brunei | Male | 8208 (7796-8594) | 1935 (1526-2299) | 23.63 (18.64-27.56) | 926 (713-1144) | 11.34 (8.57-13.92) | 1249 (961-1523) | 15.27 (11.78-18.49) | 295 (222-380) | 3.61 (2.71-4.61) | 260 (198-331) | 3.18 (2.41-4.06) |
| Bulgaria | Female | 591593 (574403-608827) | 136226 (112567-162057) | 23.07 (19.05-27.28) | 84087 (67406-103742) | 14.24 (11.44-17.58) | 69462 (54438-86866) | 11.77 (9.28-14.66) | 46388 (35685-60582) | 7.85 (5.96-10.28) | 41647 (30892-55968) | 7.05 (5.22-9.41) |
| Bulgaria | Male | 682527 (663183-702422) | 193200 (161071-218843) | 28.31 (23.62-32.04) | 124144 (98840-147689) | 18.26 (14.53-21.55) | 102381 (82197-123874) | 15.01 (12.10-18.20) | 53892 (40176-67784) | 7.91 (5.86-9.95) | 45835 (33467-60160) | 6.73 (4.93-8.83) |
| Burkina Faso | Female | 202620 (186008-219418) | 24293 (17014-31168) | 12.02 (8.48-15.14) | 17545 (12092-22842) | 8.61 (6.15-11.17) | 10727 (7387-14248) | 5.27 (3.69-6.92) | 3658 (2379-5234) | 1.82 (1.19-2.55) | 2507 (1511-3690) | 1.23 (0.76-1.84) |
| Burkina Faso | Male | 256142 (237524-274527) | 31585 (22818-40046) | 12.35 (8.86-15.63) | 22592 (16176-29633) | 8.81 (6.27-11.44) | 13537 (9721-18160) | 5.31 (3.84-7.07) | 3027 (2011-4388) | 1.18 (0.79-1.72) | 1551 (991-2424) | 0.61 (0.39-0.94) |
| Burundi | Female | 120044 (111936-128792) | 23107 (18641-27751) | 19.33 (15.90-22.87) | 16418 (12932-19983) | 13.68 (11.03-16.51) | 9837 (7637-12243) | 8.19 (6.45-9.92) | 4033 (2877-5493) | 3.37 (2.44-4.52) | 2723 (1799-3870) | 2.27 (1.51-3.22) |
| Burundi | Male | 153117 (141314-164320) | 20062 (14845-25012) | 13.08 (9.86-15.99) | 14551 (10796-18285) | 9.51 (7.02-11.88) | 8715 (6309-11195) | 5.71 (4.15-7.23) | 2448 (1560-3533) | 1.60 (1.04-2.26) | 1632 (927-2518) | 1.06 (0.61-1.65) |
| Cambodia | Female | 395713 (370131-420496) | 66934 (47950-85328) | 16.99 (12.05-21.39) | 46366 (32836-60484) | 11.76 (8.36-14.97) | 28634 (19762-38283) | 7.25 (5.00-9.56) | 11714 (8103-16110) | 2.98 (2.07-4.03) | 9251 (6197-13325) | 2.34 (1.58-3.34) |
| Cambodia | Male | 384352 (359250-410018) | 77335 (52746-97005) | 20.17 (13.81-24.68) | 54238 (36839-69592) | 14.07 (9.52-17.95) | 33739 (22271-43905) | 8.75 (5.80-11.33) | 9498 (6042-13055) | 2.49 (1.59-3.36) | 6802 (4181-9738) | 1.77 (1.09-2.52) |
| Cameroon | Female | 338440 (311851-364189) | 42307 (31173-51932) | 12.49 (9.28-15.28) | 30104 (22302-38180) | 8.91 (6.67-11.16) | 18638 (13945-23516) | 5.55 (4.14-6.93) | 6863 (4742-9319) | 2.02 (1.43-2.74) | 3862 (2541-5464) | 1.14 (0.76-1.60) |
| Cameroon | Male | 406337 (374038-439332) | 53434 (40610-67330) | 13.21 (10.20-16.36) | 38239 (28764-49295) | 9.43 (7.13-11.91) | 23408 (17677-30884) | 5.78 (4.36-7.48) | 5436 (3664-7466) | 1.35 (0.91-1.83) | 3202 (2097-4581) | 0.80 (0.52-1.12) |
| Canada | Female | 527354 (512860-542534) | 46737 (35142-57569) | 8.89 (6.67-10.86) | 15106 (10800-20386) | 2.87 (2.05-3.86) | 37595 (27974-46722) | 7.15 (5.28-8.80) | 5582 (3894-7917) | 1.06 (0.74-1.49) | 5439 (3804-7719) | 1.03 (0.72-1.45) |
| Canada | Male | 710871 (694894-727264) | 96395 (74128-114704) | 13.57 (10.44-16.12) | 30290 (21656-40394) | 4.26 (3.04-5.68) | 75828 (57258-91681) | 10.69 (8.05-12.89) | 11851 (8246-16130) | 1.67 (1.16-2.27) | 11725 (8063-15933) | 1.65 (1.15-2.26) |
| Cape Verde | Female | 9348 (8790-9852) | 1075 (847-1314) | 11.54 (9.03-14.03) | 755 (591-931) | 8.06 (6.31-10.02) | 486 (376-608) | 5.19 (4.01-6.47) | 309 (230-399) | 3.31 (2.48-4.22) | 260 (192-341) | 2.78 (2.06-3.63) |
| Cape Verde | Male | 12349 (11630-13058) | 1559 (1217-1909) | 12.65 (9.78-15.17) | 1088 (838-1345) | 8.83 (6.80-10.76) | 702 (529-891) | 5.68 (4.37-7.18) | 282 (203-369) | 2.29 (1.66-2.95) | 203 (144-272) | 1.65 (1.18-2.20) |
| Central African Republic | Female | 84686 (76645-93054) | 7849 (5148-10985) | 9.30 (6.07-12.68) | 6155 (4080-8400) | 7.29 (4.81-9.87) | 3849 (2682-5278) | 4.59 (3.10-6.15) | 1473 (848-2195) | 1.75 (1.02-2.54) | 786 (428-1210) | 0.93 (0.51-1.43) |
| Central African Republic | Male | 107926 (97341-118233) | 10449 (6407-14464) | 9.69 (5.96-13.29) | 7992 (5004-10982) | 7.44 (4.54-10.06) | 4936 (3012-6875) | 4.62 (2.82-6.28) | 1010 (561-1524) | 0.94 (0.52-1.41) | 571 (320-887) | 0.53 (0.29-0.80) |
| Chad | Female | 159519 (145974-172990) | 19203 (13560-24735) | 12.05 (8.61-15.31) | 13909 (9707-18242) | 8.70 (6.19-11.30) | 8432 (5788-11133) | 5.31 (3.64-6.90) | 3279 (2137-4755) | 2.05 (1.35-2.95) | 2103 (1279-3206) | 1.31 (0.80-1.98) |
| Chad | Male | 208207 (192368-222642) | 25683 (18686-32401) | 12.40 (9.11-15.35) | 18525 (13359-23560) | 8.93 (6.34-11.26) | 11065 (7806-14560) | 5.36 (3.81-6.90) | 3063 (1975-4436) | 1.48 (0.94-2.14) | 1991 (1240-3062) | 0.96 (0.60-1.46) |
| Chile | Female | 210670 (205411-215237) | 30853 (25353-36024) | 14.64 (12.01-17.01) | 18403 (14866-22207) | 8.74 (7.09-10.54) | 16695 (13176-20156) | 7.94 (6.27-9.55) | 8920 (7038-10997) | 4.24 (3.32-5.22) | 8142 (6368-10153) | 3.86 (3.02-4.82) |
| Chile | Male | 285962 (279733-292624) | 51621 (42236-60379) | 18.10 (14.81-20.99) | 30466 (24602-36561) | 10.65 (8.62-12.82) | 27843 (22091-33767) | 9.76 (7.78-11.78) | 12172 (9577-15112) | 4.27 (3.34-5.27) | 10428 (8251-13036) | 3.65 (2.89-4.55) |
| China | Female | 38944122 (37295692-40461171) | 8343265 (6742215-9633047) | 21.42 (17.47-24.52) | 5223515 (4119720-6315887) | 13.37 (10.60-16.11) | 4313977 (3421686-5266441) | 11.08 (8.79-13.48) | 1513726 (1159447-1904270) | 3.90 (2.99-4.87) | 1266616 (957057-1611900) | 3.26 (2.48-4.11) |
| China | Male | 57081760 (54686483-59429593) | 14865970 (12162752-16973375) | 26.10 (21.40-29.46) | 9826005 (7771194-11682209) | 17.24 (13.65-20.36) | 8241510 (6448663-10042703) | 14.50 (11.33-17.39) | 2372691 (1871923-2986567) | 4.16 (3.27-5.22) | 1881369 (1440850-2375608) | 3.29 (2.54-4.15) |
| Colombia | Female | 601056 (581899-621364) | 94184 (77751-110668) | 15.65 (12.83-18.31) | 62644 (50711-74472) | 10.41 (8.46-12.36) | 43625 (34969-53709) | 7.26 (5.82-8.91) | 28020 (22582-34326) | 4.65 (3.70-5.73) | 24313 (19436-29973) | 4.04 (3.23-5.00) |
| Colombia | Male | 772336 (740655-804559) | 182810 (153179-208081) | 23.65 (19.76-26.78) | 121777 (98357-144722) | 15.73 (12.73-18.62) | 85530 (68693-102633) | 11.05 (8.94-13.25) | 41218 (32246-51556) | 5.35 (4.24-6.61) | 33473 (26209-43290) | 4.32 (3.39-5.59) |
| Comoros | Female | 11995 (11123-12917) | 2299 (1829-2851) | 19.21 (15.51-23.07) | 1608 (1256-2020) | 13.45 (10.60-16.59) | 994 (756-1272) | 8.32 (6.38-10.42) | 430 (314-585) | 3.59 (2.65-4.89) | 279 (193-412) | 2.33 (1.62-3.36) |
| Comoros | Male | 10697 (9924-11497) | 1440 (1119-1800) | 13.45 (10.39-16.37) | 1030 (795-1300) | 9.62 (7.45-11.96) | 642 (487-825) | 6.03 (4.62-7.59) | 174 (120-243) | 1.64 (1.14-2.27) | 114 (78-165) | 1.06 (0.73-1.53) |
| Congo | Female | 103690 (96074-111867) | 9736 (6620-13308) | 9.44 (6.40-12.65) | 7533 (5098-10096) | 7.28 (4.94-9.66) | 4947 (3359-6642) | 4.77 (3.31-6.31) | 1826 (1145-2805) | 1.77 (1.12-2.66) | 1097 (588-1786) | 1.05 (0.58-1.68) |
| Congo | Male | 96639 (88748-104400) | 9529 (6476-12885) | 9.91 (6.79-13.19) | 7095 (4886-9511) | 7.37 (5.05-9.68) | 4589 (3171-6149) | 4.76 (3.28-6.29) | 1145 (726-1697) | 1.19 (0.76-1.75) | 821 (501-1308) | 0.85 (0.53-1.35) |
| Costa Rica | Female | 46832 (45595-48063) | 6962 (5825-8111) | 14.85 (12.43-17.30) | 4345 (3532-5216) | 9.26 (7.56-11.09) | 3590 (2852-4303) | 7.69 (6.12-9.21) | 2219 (1758-2734) | 4.73 (3.78-5.86) | 2145 (1700-2635) | 4.59 (3.65-5.66) |
| Costa Rica | Male | 68014 (66026-69886) | 13581 (11242-15866) | 20.02 (16.53-23.25) | 8269 (6768-9873) | 12.20 (9.98-14.50) | 6830 (5524-8185) | 10.00 (8.09-11.98) | 3273 (2648-4014) | 4.81 (3.87-5.91) | 3072 (2466-3778) | 4.51 (3.60-5.56) |
| Cote d'Ivoire | Female | 263570 (244336-283537) | 36600 (28096-44752) | 13.86 (10.71-16.75) | 25585 (19297-31833) | 9.76 (7.36-11.88) | 16229 (11937-20825) | 6.15 (4.65-7.77) | 5752 (4018-7895) | 2.19 (1.56-3.00) | 3714 (2449-5408) | 1.40 (0.94-2.07) |
| Cote d'Ivoire | Male | 425158 (394277-456630) | 59797 (44266-72522) | 14.07 (10.87-17.10) | 41888 (31504-52238) | 9.84 (7.51-12.31) | 26508 (20168-33604) | 6.23 (4.79-7.86) | 6368 (4380-8863) | 1.50 (1.03-2.08) | 4299 (2782-6259) | 1.01 (0.66-1.48) |
| Croatia | Female | 170668 (165460-175837) | 35077 (29015-41532) | 20.50 (16.88-24.29) | 19870 (16002-24865) | 11.61 (9.30-14.52) | 18931 (14709-23778) | 11.13 (8.59-13.83) | 12583 (10013-16079) | 7.39 (5.89-9.42) | 11061 (8666-14332) | 6.48 (5.07-8.36) |
| Croatia | Male | 176011 (171523-180570) | 45130 (38694-50653) | 25.66 (21.98-28.66) | 26842 (21908-32012) | 15.23 (12.46-18.12) | 25425 (20901-30502) | 14.45 (11.90-17.27) | 14766 (11707-17984) | 8.37 (6.68-10.19) | 10645 (8179-13256) | 6.04 (4.64-7.56) |
| Cuba | Female | 315201 (305732-323809) | 29069 (21328-37163) | 9.17 (6.86-11.71) | 19334 (14593-24948) | 6.15 (4.66-7.89) | 14776 (11166-19011) | 4.70 (3.55-6.02) | 9386 (6867-12414) | 2.98 (2.18-3.92) | 8667 (6338-11584) | 2.75 (2.03-3.68) |
| Cuba | Male | 404149 (391458-416815) | 53552 (40816-63409) | 13.25 (10.09-15.65) | 34577 (25836-42468) | 8.54 (6.38-10.49) | 26092 (19568-32458) | 6.47 (4.80-8.07) | 12866 (9303-16386) | 3.18 (2.30-4.06) | 11517 (8417-14715) | 2.85 (2.07-3.66) |
| Cyprus | Female | 18819 (17900-19779) | 1538 (1057-2034) | 8.16 (5.66-10.77) | 782 (564-1049) | 4.16 (3.00-5.53) | 1087 (763-1419) | 5.77 (4.06-7.57) | 338 (220-495) | 1.79 (1.18-2.61) | 311 (201-457) | 1.65 (1.06-2.44) |
| Cyprus | Male | 28443 (27013-29784) | 3963 (3201-4682) | 13.97 (11.36-16.38) | 1855 (1446-2335) | 6.51 (5.10-8.19) | 2602 (2065-3150) | 9.14 (7.34-11.07) | 726 (539-964) | 2.54 (1.89-3.37) | 660 (473-897) | 2.31 (1.69-3.16) |
| Czech Republic | Female | 334933 (324962-344871) | 67248 (55226-78246) | 20.06 (16.54-23.29) | 33746 (26785-41853) | 10.05 (8.00-12.46) | 40661 (32043-49407) | 12.11 (9.66-14.68) | 18436 (14169-23681) | 5.51 (4.20-7.05) | 17330 (13357-22311) | 5.16 (3.98-6.66) |
| Czech Republic | Male | 405166 (394034-416773) | 102158 (87100-116480) | 25.19 (21.47-28.63) | 54297 (43201-65042) | 13.40 (10.68-16.09) | 64175 (52354-77862) | 15.81 (13.02-19.21) | 29268 (22241-36464) | 7.21 (5.53-8.98) | 27565 (21483-34561) | 6.79 (5.33-8.47) |
| Democratic Republic of the Congo | Female | 1081881 (982486-1183894) | 25632 (7673-51469) | 2.36 (0.72-4.68) | 24571 (7597-47923) | 2.29 (0.71-4.44) | 19509 (6149-36882) | 1.81 (0.58-3.41) | 5494 (1282-13049) | 0.51 (0.12-1.18) | 3164 (711-8174) | 0.29 (0.07-0.74) |
| Democratic Republic of the Congo | Male | 1126695 (1032412-1231325) | 44269 (19360-76040) | 3.96 (1.72-6.87) | 39539 (17997-65463) | 3.55 (1.57-5.83) | 28054 (13119-44280) | 2.50 (1.19-3.92) | 6148 (2117-12048) | 0.54 (0.19-1.06) | 4599 (1453-9021) | 0.41 (0.13-0.80) |
| Denmark | Female | 85323 (82813-87698) | 6596 (4794-8505) | 7.76 (5.60-9.90) | 1337 (828-2102) | 1.57 (0.97-2.45) | 5939 (4325-7557) | 6.96 (5.08-8.85) | 505 (280-842) | 0.59 (0.33-0.98) | 327 (183-571) | 0.39 (0.22-0.66) |
| Denmark | Male | 114938 (112292-117351) | 15047 (12424-17680) | 13.11 (10.78-15.32) | 2830 (1786-4216) | 2.46 (1.55-3.66) | 13167 (10624-15638) | 11.49 (9.23-13.61) | 1174 (707-1840) | 1.02 (0.61-1.59) | 779 (449-1274) | 0.68 (0.39-1.10) |
| Djibouti | Female | 12122 (11226-12992) | 2398 (1867-2885) | 19.86 (15.80-23.32) | 1659 (1270-2032) | 13.70 (10.66-16.47) | 1063 (814-1339) | 8.82 (6.76-10.89) | 414 (285-566) | 3.41 (2.36-4.59) | 275 (175-400) | 2.26 (1.44-3.25) |
| Djibouti | Male | 21565 (19878-23202) | 2783 (2057-3531) | 12.97 (9.66-16.15) | 1988 (1440-2552) | 9.26 (6.69-11.75) | 1296 (899-1705) | 6.02 (4.32-7.78) | 333 (210-487) | 1.55 (1.03-2.24) | 224 (129-344) | 1.04 (0.59-1.57) |
| Dominica | Female | 1763 (1676-1852) | 187 (135-239) | 10.59 (7.73-13.44) | 129 (96-165) | 7.28 (5.40-9.38) | 94 (69-123) | 5.34 (3.93-6.98) | 65 (46-86) | 3.67 (2.61-4.88) | 56 (39-75) | 3.15 (2.19-4.29) |
| Dominica | Male | 2004 (1881-2123) | 314 (241-383) | 15.69 (12.21-18.80) | 208 (156-264) | 10.39 (7.78-12.97) | 151 (111-193) | 7.55 (5.63-9.49) | 69 (49-93) | 3.45 (2.44-4.61) | 57 (40-79) | 2.86 (2.04-3.93) |
| Dominican Republic | Female | 212787 (202444-223482) | 19766 (14413-25309) | 9.35 (6.78-11.85) | 13773 (10087-17617) | 6.48 (4.74-8.21) | 10274 (7650-13118) | 4.82 (3.60-6.17) | 7000 (5067-9078) | 3.29 (2.38-4.22) | 5916 (4173-7791) | 2.79 (1.95-3.69) |
| Dominican Republic | Male | 314798 (295522-333605) | 45545 (36012-55196) | 14.52 (11.39-17.39) | 29905 (23188-37142) | 9.54 (7.37-11.76) | 22158 (16974-28028) | 7.06 (5.39-8.86) | 11639 (8795-15032) | 3.71 (2.78-4.82) | 9744 (7263-12823) | 3.10 (2.30-4.04) |
| Ecuador | Female | 179299 (172883-185737) | 22965 (17840-27562) | 12.85 (10.04-15.36) | 15403 (11786-18965) | 8.61 (6.58-10.52) | 10672 (8159-13370) | 5.95 (4.56-7.43) | 5279 (3926-6677) | 2.95 (2.22-3.73) | 4766 (3527-6137) | 2.67 (2.00-3.41) |
| Ecuador | Male | 237430 (226226-248561) | 38603 (30082-46589) | 16.28 (12.78-19.55) | 25876 (19716-31565) | 10.87 (8.34-13.29) | 18010 (13653-22280) | 7.57 (5.80-9.35) | 6267 (4567-8007) | 2.64 (1.95-3.35) | 4978 (3552-6401) | 2.10 (1.52-2.68) |
| Egypt | Female | 2675481 (2519762-2828624) | 24841 (7083-64336) | 0.93 (0.26-2.40) | 22102 (6122-58269) | 0.83 (0.23-2.19) | 17862 (4403-45116) | 0.66 (0.16-1.70) | 4737 (301-20833) | 0.18 (0.01-0.78) | 4005 (221-18210) | 0.15 (0.01-0.68) |
| Egypt | Male | 3823922 (3615242-4026629) | 315125 (210484-439684) | 8.27 (5.46-11.37) | 239297 (160821-323260) | 6.24 (4.19-8.44) | 162222 (113573-221600) | 4.26 (2.95-5.75) | 45292 (26629-67774) | 1.19 (0.71-1.76) | 37072 (21030-57404) | 0.97 (0.56-1.49) |
| El Salvador | Female | 85840 (81378-90215) | 11902 (9687-14061) | 13.91 (11.38-16.28) | 8069 (6405-9843) | 9.40 (7.52-11.34) | 5385 (4230-6663) | 6.29 (4.97-7.71) | 3591 (2762-4542) | 4.19 (3.28-5.25) | 3255 (2517-4164) | 3.79 (2.98-4.85) |
| El Salvador | Male | 95675 (90637-100992) | 18592 (15253-21920) | 19.44 (16.25-22.69) | 12332 (10004-14780) | 12.92 (10.61-15.28) | 8299 (6522-10143) | 8.67 (6.89-10.45) | 3480 (2692-4357) | 3.63 (2.82-4.54) | 3054 (2373-3879) | 3.18 (2.48-3.98) |
| Equatorial Guinea | Female | 14576 (13290-15970) | 1381 (948-1873) | 9.46 (6.49-12.56) | 1007 (704-1344) | 6.93 (4.83-9.12) | 745 (510-1000) | 5.12 (3.59-6.72) | 259 (162-397) | 1.79 (1.14-2.67) | 168 (91-274) | 1.15 (0.63-1.82) |
| Equatorial Guinea | Male | 14397 (13119-15626) | 1488 (1072-1965) | 10.35 (7.43-13.43) | 1047 (767-1375) | 7.33 (5.22-9.40) | 764 (552-1008) | 5.33 (3.85-6.92) | 170 (111-250) | 1.19 (0.77-1.72) | 117 (72-184) | 0.82 (0.50-1.30) |
| Eritrea | Female | 90214 (83303-97285) | 16677 (12192-20700) | 18.47 (13.66-22.46) | 11873 (8433-14821) | 13.17 (9.52-16.18) | 7153 (4988-9221) | 7.93 (5.61-10.01) | 2046 (1354-2887) | 2.27 (1.52-3.15) | 1371 (842-2042) | 1.52 (0.95-2.24) |
| Eritrea | Male | 90485 (83157-98103) | 10268 (7198-13552) | 11.33 (8.06-14.53) | 7608 (5228-9996) | 8.38 (5.74-10.75) | 4609 (3139-6201) | 5.08 (3.55-6.81) | 791 (522-1157) | 0.87 (0.58-1.25) | 544 (341-809) | 0.60 (0.38-0.89) |
| Estonia | Female | 53871 (52068-55731) | 126 (0-921) | 0.24 (0.00-1.70) | 124 (0-853) | 0.23 (0.00-1.59) | 126 (0-911) | 0.24 (0.00-1.68) | 39 (0-290) | 0.07 (0.00-0.53) | 28 (0-212) | 0.05 (0.00-0.39) |
| Estonia | Male | 50880 (49574-52255) | 5381 (3796-7014) | 10.59 (7.41-13.75) | 2908 (2055-3804) | 5.73 (4.04-7.48) | 3581 (2573-4648) | 7.03 (5.09-9.12) | 790 (520-1091) | 1.55 (1.02-2.14) | 604 (393-869) | 1.19 (0.78-1.71) |
| Ethiopia | Female | 727940 (690270-766228) | 132155 (99884-160408) | 18.14 (13.81-21.92) | 93539 (69746-115093) | 12.83 (9.61-15.84) | 57440 (41588-72304) | 7.88 (5.67-9.83) | 15587 (10766-21620) | 2.15 (1.51-2.91) | 7634 (4760-11174) | 1.05 (0.66-1.50) |
| Ethiopia | Male | 820696 (777410-862024) | 101535 (73421-128233) | 12.36 (8.99-15.55) | 72938 (52612-95476) | 8.88 (6.44-11.51) | 44447 (32613-58045) | 5.41 (3.96-7.05) | 9633 (6411-13518) | 1.17 (0.79-1.66) | 6163 (4032-9009) | 0.75 (0.50-1.10) |
| Federated States of Micronesia | Female | 3422 (3183-3672) | 457 (318-580) | 13.34 (9.24-16.78) | 315 (214-410) | 9.21 (6.23-11.89) | 206 (144-275) | 6.07 (4.26-7.95) | 89 (57-120) | 2.59 (1.73-3.49) | 54 (34-77) | 1.58 (1.00-2.21) |
| Federated States of Micronesia | Male | 5814 (5373-6224) | 852 (589-1085) | 14.69 (10.40-18.52) | 596 (414-768) | 10.28 (7.18-13.17) | 392 (265-515) | 6.77 (4.59-8.74) | 119 (79-160) | 2.04 (1.32-2.76) | 68 (42-97) | 1.18 (0.71-1.69) |
| Fiji | Female | 26470 (24713-28131) | 4296 (3510-5272) | 16.25 (13.42-19.40) | 2913 (2352-3643) | 11.03 (8.98-13.62) | 1961 (1515-2467) | 7.41 (5.75-9.28) | 1151 (873-1496) | 4.35 (3.33-5.59) | 862 (610-1156) | 3.26 (2.38-4.35) |
| Fiji | Male | 45936 (42644-49363) | 7719 (5998-9382) | 16.80 (13.23-20.08) | 5237 (4012-6558) | 11.42 (8.72-14.18) | 3515 (2677-4469) | 7.61 (5.82-9.62) | 1266 (917-1669) | 2.77 (1.98-3.65) | 864 (582-1213) | 1.88 (1.27-2.64) |
| Finland | Female | 146532 (141088-151817) | 10106 (6595-14033) | 6.89 (4.52-9.53) | 3731 (2321-5390) | 2.54 (1.57-3.69) | 8790 (5942-12097) | 5.99 (4.04-8.21) | 1810 (1062-2788) | 1.24 (0.72-1.90) | 1571 (926-2401) | 1.07 (0.63-1.63) |
| Finland | Male | 182434 (178558-186552) | 26371 (22025-30672) | 14.45 (12.13-16.81) | 8149 (5887-10491) | 4.46 (3.18-5.78) | 20832 (16899-24863) | 11.43 (9.31-13.69) | 3560 (2468-4785) | 1.95 (1.36-2.64) | 3202 (2218-4359) | 1.76 (1.23-2.39) |
| France | Female | 938076 (907009-970894) | 71139 (46624-95723) | 7.58 (4.93-10.18) | 29870 (20682-41490) | 3.18 (2.21-4.36) | 55416 (38453-73484) | 5.89 (4.11-7.78) | 12819 (8357-18873) | 1.37 (0.89-2.03) | 11784 (7494-17170) | 1.25 (0.80-1.84) |
| France | Male | 1063537 (1036851-1090567) | 129850 (105230-152026) | 12.19 (9.88-14.30) | 49763 (38189-62862) | 4.67 (3.60-5.93) | 94806 (75669-113948) | 8.92 (7.18-10.64) | 21961 (16402-29361) | 2.06 (1.55-2.75) | 20031 (14745-26740) | 1.88 (1.39-2.51) |
| Gabon | Female | 27010 (24630-29177) | 2593 (1744-3456) | 9.65 (6.48-12.71) | 1846 (1237-2440) | 6.89 (4.66-9.05) | 1408 (970-1919) | 5.22 (3.68-7.01) | 539 (344-781) | 2.00 (1.28-2.85) | 392 (228-594) | 1.45 (0.86-2.18) |
| Gabon | Male | 34139 (31428-36861) | 3355 (2294-4509) | 9.81 (6.72-13.06) | 2372 (1583-3149) | 6.98 (4.60-9.05) | 1786 (1248-2417) | 5.25 (3.64-7.00) | 420 (253-626) | 1.24 (0.74-1.82) | 303 (178-483) | 0.89 (0.53-1.40) |
| Georgia | Female | 176825 (172548-181179) | 23094 (18298-28418) | 13.02 (10.24-15.98) | 15956 (12285-19956) | 9.04 (6.97-11.25) | 10847 (8232-13826) | 6.13 (4.71-7.80) | 8506 (6348-11027) | 4.80 (3.60-6.29) | 7681 (5624-10220) | 4.35 (3.19-5.77) |
| Georgia | Male | 218391 (214040-222772) | 40872 (34273-47365) | 18.70 (15.76-21.65) | 27392 (22326-32611) | 12.52 (10.27-14.91) | 18486 (14751-22445) | 8.48 (6.80-10.22) | 10817 (8436-13264) | 4.95 (3.89-6.12) | 8341 (6466-10522) | 3.81 (2.97-4.82) |
| Germany | Female | 2053856 (1983150-2135381) | 158710 (104590-217415) | 7.73 (5.07-10.59) | 58869 (38594-85373) | 2.86 (1.89-4.14) | 132758 (91891-180611) | 6.45 (4.49-8.77) | 26997 (16338-40689) | 1.32 (0.80-1.97) | 25818 (15706-39187) | 1.26 (0.76-1.90) |
| Germany | Male | 2498914 (2440923-2551964) | 352636 (288341-414156) | 14.11 (11.49-16.53) | 113309 (84215-145677) | 4.54 (3.37-5.79) | 274272 (219874-329283) | 10.96 (8.77-13.13) | 51462 (36296-69046) | 2.07 (1.45-2.77) | 48112 (33905-64707) | 1.92 (1.35-2.59) |
| Ghana | Female | 517514 (482423-552870) | 82753 (66168-98513) | 16.04 (12.77-18.73) | 58105 (44792-71720) | 11.25 (8.66-13.65) | 36318 (27775-46263) | 7.05 (5.36-8.75) | 18042 (13584-23089) | 3.47 (2.63-4.43) | 14291 (10432-18803) | 2.76 (2.01-3.60) |
| Ghana | Male | 601800 (563489-638444) | 97850 (76242-116534) | 16.34 (12.79-19.24) | 68660 (52592-84570) | 11.44 (8.72-13.97) | 43240 (32429-54467) | 7.20 (5.39-9.02) | 10960 (7916-14510) | 1.83 (1.33-2.43) | 8267 (5962-11234) | 1.37 (0.98-1.87) |
| Greece | Female | 315722 (305153-326226) | 25440 (16334-32977) | 8.05 (5.17-10.39) | 15237 (10752-19955) | 4.82 (3.42-6.24) | 15427 (10748-20341) | 4.89 (3.40-6.42) | 7909 (5297-10716) | 2.49 (1.66-3.39) | 7822 (5142-10633) | 2.47 (1.63-3.36) |
| Greece | Male | 377607 (370072-385120) | 49856 (39261-59689) | 13.21 (10.38-15.75) | 28170 (21875-34298) | 7.45 (5.83-9.07) | 28518 (21949-34984) | 7.55 (5.82-9.25) | 12268 (9341-15323) | 3.25 (2.46-4.06) | 12030 (8972-14967) | 3.19 (2.38-3.96) |
| Greenland | Female | 747 (711-783) | 77 (54-100) | 10.32 (7.20-13.16) | 22 (14-32) | 2.94 (1.80-4.29) | 64 (44-83) | 8.56 (5.92-10.99) | 6 (4-10) | 0.86 (0.52-1.29) | 5 (3-8) | 0.70 (0.42-1.05) |
| Greenland | Male | 1517 (1431-1596) | 224 (155-281) | 14.85 (10.18-18.26) | 64 (38-91) | 4.24 (2.53-6.01) | 184 (123-234) | 12.14 (8.15-15.29) | 18 (10-27) | 1.20 (0.69-1.82) | 15 (8-23) | 0.99 (0.55-1.52) |
| Grenada | Female | 2164 (2110-2223) | 218 (157-279) | 10.12 (7.29-12.86) | 149 (108-192) | 6.90 (5.03-8.76) | 113 (83-147) | 5.22 (3.82-6.71) | 73 (50-98) | 3.36 (2.31-4.51) | 63 (44-87) | 2.93 (2.04-4.00) |
| Grenada | Male | 2859 (2764-2952) | 423 (323-508) | 14.79 (11.23-17.72) | 274 (206-344) | 9.62 (7.20-11.99) | 206 (153-261) | 7.21 (5.34-9.15) | 96 (66-127) | 3.35 (2.29-4.53) | 71 (49-101) | 2.50 (1.72-3.49) |
| Guatemala | Female | 138962 (134365-143228) | 20886 (17191-24310) | 15.06 (12.40-17.50) | 14253 (11285-16933) | 10.26 (8.16-12.17) | 9579 (7538-11713) | 6.89 (5.46-8.41) | 4432 (3420-5607) | 3.19 (2.49-4.02) | 3077 (2307-3998) | 2.21 (1.66-2.86) |
| Guatemala | Male | 153395 (147788-158929) | 30338 (25350-35078) | 19.81 (16.49-23.00) | 20192 (16572-23751) | 13.18 (10.80-15.46) | 13665 (11155-16257) | 8.91 (7.31-10.69) | 4444 (3447-5583) | 2.90 (2.26-3.63) | 3082 (2309-4080) | 2.01 (1.52-2.62) |
| Guinea | Female | 175323 (162718-187303) | 21813 (15941-27164) | 12.43 (9.08-15.24) | 15758 (11663-20107) | 9.00 (6.78-11.25) | 9625 (6982-12473) | 5.52 (4.06-6.94) | 4893 (3363-6588) | 2.79 (1.97-3.74) | 2424 (1498-3629) | 1.38 (0.87-2.05) |
| Guinea | Male | 181947 (169123-194228) | 22591 (16729-28228) | 12.41 (9.20-15.27) | 16173 (11865-20663) | 8.90 (6.64-11.24) | 9750 (7171-12849) | 5.38 (3.99-6.88) | 2948 (1986-4202) | 1.62 (1.09-2.28) | 1697 (1012-2539) | 0.93 (0.57-1.39) |
| Guinea-Bissau | Female | 33418 (30850-36085) | 4273 (3103-5402) | 12.78 (9.39-15.75) | 3097 (2250-3983) | 9.24 (6.72-11.54) | 1874 (1355-2469) | 5.61 (4.16-7.23) | 779 (525-1110) | 2.35 (1.59-3.32) | 503 (319-763) | 1.50 (0.97-2.29) |
| Guinea-Bissau | Male | 35686 (33126-38210) | 4650 (3257-5830) | 13.01 (9.43-16.26) | 3350 (2347-4301) | 9.41 (6.73-11.97) | 2024 (1469-2638) | 5.67 (4.12-7.33) | 524 (344-752) | 1.47 (0.95-2.10) | 338 (193-520) | 0.95 (0.55-1.44) |
| Guyana | Female | 20932 (20027-21880) | 2133 (1524-2782) | 10.25 (7.29-13.24) | 1472 (1051-1925) | 7.03 (5.08-9.15) | 1157 (813-1494) | 5.51 (3.90-7.05) | 619 (430-827) | 2.95 (2.05-3.99) | 487 (328-665) | 2.33 (1.57-3.20) |
| Guyana | Male | 26859 (25382-28292) | 4178 (3150-5092) | 15.57 (11.75-18.96) | 2689 (1988-3442) | 10.00 (7.39-12.71) | 2068 (1542-2710) | 7.73 (5.74-9.91) | 773 (564-1048) | 2.88 (2.10-3.89) | 611 (431-840) | 2.29 (1.62-3.10) |
| Haiti | Female | 350870 (323371-377596) | 35302 (25454-46177) | 10.10 (7.34-13.05) | 26546 (19301-34507) | 7.58 (5.53-9.75) | 16864 (12551-21659) | 4.82 (3.61-6.13) | 9732 (6867-13168) | 2.77 (1.98-3.69) | 5234 (3572-7399) | 1.49 (1.03-2.08) |
| Haiti | Male | 301228 (277701-325349) | 45510 (35282-53656) | 15.01 (11.94-17.59) | 31974 (24391-38837) | 10.58 (8.21-12.90) | 19691 (14507-24698) | 6.51 (4.91-8.13) | 7117 (5205-9061) | 2.36 (1.72-2.99) | 3892 (2742-5225) | 1.29 (0.91-1.71) |
| Honduras | Female | 181732 (169536-192898) | 26808 (21338-32214) | 14.84 (11.89-17.56) | 18624 (14579-23173) | 10.27 (8.09-12.57) | 11844 (8881-15133) | 6.54 (4.85-8.18) | 7778 (5722-10140) | 4.32 (3.27-5.53) | 7288 (5330-9583) | 4.02 (3.00-5.22) |
| Honduras | Male | 186106 (174741-197693) | 38281 (30929-45532) | 20.62 (16.56-24.27) | 26038 (20711-31653) | 13.95 (11.15-16.81) | 16509 (12866-20465) | 8.88 (6.92-10.99) | 7700 (5728-10134) | 4.16 (3.08-5.41) | 6901 (5016-9158) | 3.72 (2.71-4.95) |
| Hungary | Female | 450389 (437996-462532) | 102854 (86422-118189) | 22.91 (19.28-26.26) | 58140 (47376-71126) | 12.95 (10.43-15.77) | 56891 (45131-68549) | 12.62 (10.01-15.12) | 32088 (24962-41297) | 7.14 (5.48-9.10) | 28826 (21704-37566) | 6.42 (4.82-8.38) |
| Hungary | Male | 483129 (470478-495649) | 140388 (122417-157332) | 29.12 (25.40-32.47) | 86373 (70824-102343) | 17.88 (14.67-21.01) | 83569 (68596-100417) | 17.29 (14.24-20.85) | 38888 (29658-49047) | 8.07 (6.18-10.08) | 35426 (27535-45031) | 7.32 (5.69-9.31) |
| Iceland | Female | 4498 (4330-4673) | 331 (228-440) | 7.35 (5.07-9.68) | 70 (42-109) | 1.57 (0.95-2.42) | 302 (208-393) | 6.70 (4.69-8.68) | 35 (19-57) | 0.77 (0.43-1.26) | 32 (17-52) | 0.70 (0.38-1.14) |
| Iceland | Male | 6076 (5910-6254) | 897 (744-1055) | 14.76 (12.31-17.43) | 174 (112-251) | 2.87 (1.84-4.14) | 782 (642-932) | 12.86 (10.61-15.25) | 83 (51-124) | 1.36 (0.85-2.05) | 78 (47-118) | 1.28 (0.77-1.96) |
| India | Female | 25881124 (25016533-26693925) | 3603946 (2816417-4159932) | 13.96 (10.88-16.11) | 2520600 (1909740-3034481) | 9.75 (7.41-11.70) | 1583991 (1189904-1984158) | 6.12 (4.61-7.58) | 496202 (367388-633986) | 1.92 (1.42-2.46) | 436193 (320666-551405) | 1.68 (1.24-2.12) |
| India | Male | 38640741 (37227421-39928945) | 6659995 (5017044-7914203) | 17.27 (13.11-20.40) | 4595749 (3489379-5587828) | 11.87 (8.99-14.46) | 2871848 (2165844-3559422) | 7.42 (5.61-9.17) | 657749 (500248-846022) | 1.71 (1.29-2.16) | 543464 (404913-690013) | 1.41 (1.05-1.79) |
| Indonesia | Female | 8563301 (8149796-8964706) | 1841664 (1480083-2186287) | 21.54 (17.43-25.21) | 1230643 (982956-1498078) | 14.36 (11.52-17.41) | 820319 (650690-1006030) | 9.60 (7.60-11.65) | 303521 (235508-395452) | 3.56 (2.74-4.49) | 165482 (122690-219316) | 1.93 (1.43-2.54) |
| Indonesia | Male | 10734003 (10207275-11277601) | 2598492 (2107078-3031476) | 24.31 (19.71-28.08) | 1755086 (1377441-2130131) | 16.33 (12.87-19.74) | 1173238 (927745-1440517) | 10.93 (8.62-13.33) | 276889 (201154-352498) | 2.58 (1.88-3.31) | 155924 (113192-204760) | 1.45 (1.05-1.91) |
| Iran | Female | 1408583 (1374488-1443704) | 7875 (571-23875) | 0.56 (0.04-1.69) | 7088 (556-22632) | 0.51 (0.04-1.62) | 6068 (507-18181) | 0.43 (0.04-1.30) | 1473 (15-7880) | 0.10 (0.00-0.56) | 1274 (11-6849) | 0.09 (0.00-0.48) |
| Iran | Male | 1933855 (1898131-1968711) | 133947 (86235-183528) | 6.93 (4.45-9.49) | 102035 (67204-137303) | 5.29 (3.48-7.14) | 70431 (47663-95881) | 3.64 (2.46-4.93) | 19462 (11264-28258) | 1.00 (0.58-1.47) | 15471 (8957-22849) | 0.80 (0.46-1.18) |
| Iraq | Female | 789818 (747421-833288) | 5925 (1472-14919) | 0.75 (0.19-1.89) | 5322 (1237-13699) | 0.67 (0.15-1.71) | 4488 (862-11552) | 0.57 (0.11-1.46) | 1400 (118-5623) | 0.18 (0.01-0.71) | 1023 (59-4282) | 0.13 (0.01-0.54) |
| Iraq | Male | 1188461 (1111799-1266061) | 93519 (61766-127891) | 7.82 (5.23-10.81) | 70874 (48128-95680) | 5.97 (4.03-8.01) | 49597 (34193-66615) | 4.17 (2.92-5.54) | 23626 (14297-34277) | 2.00 (1.21-2.88) | 17481 (10406-26277) | 1.47 (0.88-2.20) |
| Ireland | Female | 52010 (50115-53886) | 1629 (760-2615) | 3.13 (1.50-5.01) | 212 (110-411) | 0.41 (0.21-0.79) | 1612 (758-2563) | 3.10 (1.49-4.91) | 72 (32-155) | 0.14 (0.06-0.30) | 64 (27-140) | 0.12 (0.05-0.27) |
| Ireland | Male | 76117 (74181-77954) | 8670 (6837-10296) | 11.36 (8.97-13.55) | 678 (461-1113) | 0.89 (0.60-1.46) | 8269 (6436-9871) | 10.85 (8.54-12.91) | 200 (132-374) | 0.26 (0.17-0.49) | 177 (112-336) | 0.23 (0.15-0.44) |
| Israel | Female | 71501 (69122-73927) | 5394 (4037-7011) | 7.56 (5.61-9.71) | 1871 (1305-2502) | 2.62 (1.83-3.52) | 4450 (3387-5744) | 6.21 (4.75-8.01) | 807 (527-1140) | 1.13 (0.73-1.60) | 730 (476-1029) | 1.02 (0.67-1.44) |
| Israel | Male | 97085 (94826-99464) | 12827 (10409-15019) | 13.21 (10.84-15.31) | 4100 (3046-5268) | 4.22 (3.14-5.44) | 10063 (8023-11994) | 10.35 (8.25-12.34) | 1508 (1068-2019) | 1.56 (1.10-2.07) | 1370 (964-1828) | 1.41 (1.00-1.89) |
| Italy | Female | 1386951 (1336155-1439710) | 147392 (103401-196002) | 10.66 (7.59-13.98) | 66687 (46480-91142) | 4.81 (3.32-6.66) | 102728 (74661-136369) | 7.41 (5.40-9.81) | 34255 (22533-48501) | 2.48 (1.63-3.53) | 31798 (20476-45910) | 2.30 (1.49-3.29) |
| Italy | Male | 1440983 (1415244-1466635) | 217415 (176610-257131) | 15.09 (12.28-17.95) | 95493 (73332-120229) | 6.62 (5.10-8.32) | 147378 (117355-180777) | 10.25 (8.20-12.53) | 43027 (32022-55250) | 2.99 (2.22-3.84) | 41154 (30830-53152) | 2.86 (2.13-3.66) |
| Jamaica | Female | 54646 (52761-56610) | 5763 (4302-7250) | 10.52 (7.93-13.17) | 4097 (3088-5115) | 7.48 (5.66-9.33) | 2794 (2113-3541) | 5.12 (3.85-6.49) | 2154 (1572-2813) | 3.94 (2.87-5.15) | 1865 (1345-2473) | 3.41 (2.46-4.52) |
| Jamaica | Male | 58445 (55549-60974) | 8923 (7057-10527) | 15.26 (12.16-18.18) | 6037 (4750-7444) | 10.38 (8.14-12.69) | 4115 (3188-5142) | 7.03 (5.50-8.83) | 2160 (1661-2774) | 3.70 (2.83-4.72) | 1738 (1286-2244) | 2.97 (2.22-3.88) |
| Japan | Female | 2406213 (2316122-2501606) | 344358 (279923-403610) | 14.29 (11.72-16.78) | 141505 (105874-177519) | 5.87 (4.42-7.40) | 242534 (189623-296156) | 10.06 (7.99-12.16) | 61378 (44193-79758) | 2.55 (1.84-3.31) | 56366 (40469-74086) | 2.33 (1.68-3.05) |
| Japan | Male | 3016946 (2968168-3064633) | 553793 (465120-629988) | 18.38 (15.35-21.03) | 219838 (172909-266207) | 7.28 (5.75-8.88) | 378521 (313557-445767) | 12.56 (10.38-14.75) | 98598 (75941-124558) | 3.26 (2.52-4.11) | 84220 (64800-106113) | 2.80 (2.15-3.52) |
| Jordan | Female | 109811 (104008-115326) | 684 (125-2057) | 0.62 (0.11-1.87) | 610 (111-1923) | 0.55 (0.10-1.75) | 529 (91-1485) | 0.48 (0.08-1.36) | 145 (4-746) | 0.13 (0.00-0.68) | 138 (3-718) | 0.13 (0.00-0.65) |
| Jordan | Male | 167856 (159147-176446) | 13763 (9206-18767) | 8.20 (5.47-11.05) | 10398 (6962-13801) | 6.18 (4.16-8.34) | 7140 (4857-9500) | 4.26 (2.91-5.59) | 2819 (1671-4049) | 1.69 (1.02-2.43) | 2554 (1551-3673) | 1.52 (0.91-2.17) |
| Kazakhstan | Female | 561090 (546813-574762) | 65830 (51980-81243) | 11.74 (9.31-14.41) | 42958 (33687-53113) | 7.67 (6.03-9.48) | 33831 (26450-42424) | 6.02 (4.71-7.57) | 23892 (18383-30500) | 4.25 (3.27-5.39) | 21947 (16576-28155) | 3.92 (2.97-5.04) |
| Kazakhstan | Male | 625873 (607314-646162) | 112935 (94506-132261) | 18.04 (15.07-21.07) | 71459 (57395-85225) | 11.45 (9.13-13.54) | 55226 (44147-68368) | 8.83 (7.02-10.85) | 30754 (24010-37471) | 4.92 (3.83-5.99) | 26185 (20210-32122) | 4.18 (3.23-5.13) |
| Kenya | Female | 433544 (409253-459206) | 82740 (67135-98993) | 19.16 (15.77-22.61) | 57810 (46033-70673) | 13.34 (10.76-16.13) | 36259 (28578-44770) | 8.40 (6.60-10.16) | 14397 (10723-18925) | 3.34 (2.53-4.32) | 7054 (4957-9693) | 1.63 (1.15-2.23) |
| Kenya | Male | 447862 (424889-470097) | 505 (0-4532) | 0.11 (0.00-1.01) | 505 (0-4532) | 0.11 (0.00-1.01) | 497 (0-3591) | 0.11 (0.00-0.81) | 61 (0-747) | 0.01 (0.00-0.16) | 24 (0-332) | 0.01 (0.00-0.07) |
| Kiribati | Female | 2574 (2398-2733) | 319 (214-412) | 12.42 (8.38-15.84) | 225 (149-295) | 8.76 (5.91-11.31) | 141 (93-192) | 5.48 (3.63-7.34) | 47 (29-66) | 1.83 (1.13-2.53) | 24 (13-37) | 0.92 (0.52-1.40) |
| Kiribati | Male | 5320 (4946-5697) | 769 (554-971) | 14.46 (10.16-18.19) | 548 (383-704) | 10.35 (7.13-13.10) | 345 (239-452) | 6.49 (4.50-8.45) | 87 (57-118) | 1.63 (1.06-2.18) | 48 (30-70) | 0.91 (0.55-1.32) |
| Kuwait | Female | 20853 (20265-21441) | 1408 (826-1948) | 6.76 (3.93-9.38) | 792 (508-1097) | 3.80 (2.43-5.25) | 1056 (653-1454) | 5.07 (3.12-6.91) | 258 (157-354) | 1.24 (0.76-1.71) | 241 (147-338) | 1.15 (0.70-1.61) |
| Kuwait | Male | 89700 (85288-94406) | 13308 (9817-16069) | 14.82 (10.98-17.70) | 6451 (4510-8247) | 7.17 (5.06-9.23) | 8817 (6450-10870) | 9.82 (7.17-11.98) | 1738 (1241-2278) | 1.94 (1.39-2.54) | 1559 (1120-2035) | 1.74 (1.25-2.27) |
| Kyrgyzstan | Female | 140118 (134975-144885) | 15358 (11727-18988) | 10.99 (8.33-13.53) | 11092 (8495-13864) | 7.93 (6.06-9.83) | 6848 (5210-8818) | 4.90 (3.73-6.24) | 4967 (3672-6604) | 3.54 (2.65-4.69) | 4525 (3335-5976) | 3.24 (2.38-4.27) |
| Kyrgyzstan | Male | 191582 (184964-199688) | 33634 (24899-39862) | 17.51 (13.13-20.70) | 23379 (17216-28439) | 12.23 (9.08-14.77) | 14229 (10425-17684) | 7.45 (5.50-9.23) | 6017 (4365-7784) | 3.14 (2.28-4.06) | 4342 (3072-5680) | 2.26 (1.59-2.97) |
| Laos | Female | 166714 (156199-177277) | 32149 (25004-39460) | 19.35 (15.02-23.42) | 22023 (16783-27353) | 13.23 (10.02-16.25) | 13983 (10491-17796) | 8.43 (6.37-10.59) | 4694 (3272-6142) | 2.82 (1.99-3.65) | 3389 (2293-4561) | 2.04 (1.40-2.73) |
| Laos | Male | 221070 (206942-235546) | 50851 (40726-60125) | 23.03 (18.41-26.89) | 35269 (27332-42483) | 15.89 (12.40-19.06) | 22434 (17343-28004) | 10.14 (7.92-12.57) | 5324 (3831-7121) | 2.41 (1.74-3.18) | 3737 (2623-5219) | 1.70 (1.18-2.30) |
| Latvia | Female | 116483 (112815-120082) | 4976 (2501-8182) | 4.27 (2.15-7.03) | 3843 (2093-5755) | 3.32 (1.80-4.95) | 3885 (2116-5910) | 3.33 (1.83-5.06) | 2057 (1073-3180) | 1.77 (0.92-2.77) | 1727 (859-2631) | 1.48 (0.74-2.25) |
| Latvia | Male | 108374 (105454-111188) | 17904 (14648-20931) | 16.52 (13.59-19.19) | 9998 (7878-11999) | 9.23 (7.23-11.02) | 10169 (8253-12316) | 9.42 (7.60-11.28) | 4119 (3082-5126) | 3.80 (2.87-4.75) | 3313 (2443-4185) | 3.06 (2.26-3.84) |
| Lebanon | Female | 76049 (71915-80003) | 378 (99-1004) | 0.50 (0.13-1.33) | 333 (87-907) | 0.44 (0.12-1.21) | 271 (72-745) | 0.36 (0.09-0.97) | 57 (3-323) | 0.07 (0.00-0.42) | 52 (2-303) | 0.07 (0.00-0.40) |
| Lebanon | Male | 109535 (104737-114626) | 7147 (4930-9617) | 6.53 (4.54-8.66) | 5480 (3838-7289) | 5.01 (3.48-6.59) | 3798 (2699-5044) | 3.45 (2.43-4.56) | 1432 (878-2102) | 1.31 (0.79-1.93) | 1336 (781-1967) | 1.22 (0.72-1.79) |
| Lesotho | Female | 45872 (42153-49831) | 4395 (2920-6036) | 9.62 (6.32-12.98) | 3445 (2321-4663) | 7.52 (5.14-10.06) | 2181 (1478-2973) | 4.77 (3.31-6.40) | 1208 (746-1775) | 2.64 (1.65-3.80) | 1108 (684-1602) | 2.41 (1.49-3.48) |
| Lesotho | Male | 38769 (34628-42926) | 3940 (2567-5384) | 10.19 (6.78-13.75) | 2988 (2005-4153) | 7.76 (5.27-10.67) | 1875 (1251-2629) | 4.89 (3.24-6.75) | 456 (260-711) | 1.18 (0.69-1.83) | 332 (186-532) | 0.85 (0.49-1.35) |
| Liberia | Female | 65843 (61115-70534) | 8290 (6310-10452) | 12.65 (9.75-15.62) | 6036 (4547-7729) | 9.16 (6.99-11.44) | 3653 (2748-4666) | 5.56 (4.19-7.14) | 1514 (1070-2071) | 2.30 (1.62-3.09) | 947 (615-1353) | 1.44 (0.96-2.03) |
| Liberia | Male | 68518 (63481-73520) | 9107 (6799-11383) | 13.37 (10.12-16.29) | 6577 (4820-8418) | 9.61 (7.22-11.98) | 3971 (2879-5181) | 5.82 (4.28-7.43) | 1164 (789-1622) | 1.70 (1.18-2.32) | 780 (528-1128) | 1.15 (0.77-1.65) |
| Libya | Female | 144536 (134303-153938) | 1559 (513-3446) | 1.07 (0.35-2.34) | 1345 (425-2980) | 0.92 (0.30-2.05) | 1083 (349-2440) | 0.75 (0.24-1.68) | 184 (8-819) | 0.12 (0.01-0.57) | 140 (4-669) | 0.10 (0.00-0.46) |
| Libya | Male | 181296 (169300-194963) | 15936 (10781-21151) | 8.84 (5.99-11.58) | 11815 (8235-15775) | 6.55 (4.66-8.55) | 8459 (5927-11323) | 4.65 (3.32-6.17) | 2338 (1463-3462) | 1.29 (0.79-1.88) | 1735 (1004-2715) | 0.96 (0.55-1.47) |
| Lithuania | Female | 153617 (148596-159360) | 17234 (13014-21477) | 11.23 (8.57-13.98) | 9145 (6839-11879) | 5.94 (4.44-7.72) | 10151 (7606-13024) | 6.62 (4.96-8.46) | 4885 (3588-6661) | 3.18 (2.34-4.34) | 3993 (2742-5529) | 2.60 (1.80-3.61) |
| Lithuania | Male | 153642 (149716-158160) | 19941 (15912-23926) | 12.98 (10.35-15.59) | 10664 (8295-13364) | 6.94 (5.40-8.70) | 11860 (9314-14814) | 7.70 (6.05-9.64) | 4795 (3598-6236) | 3.12 (2.33-4.03) | 3370 (2411-4528) | 2.20 (1.58-2.95) |
| Luxembourg | Female | 7860 (7617-8089) | 628 (446-814) | 7.99 (5.66-10.33) | 42 (31-60) | 0.54 (0.40-0.76) | 613 (436-790) | 7.80 (5.58-10.06) | 16 (11-24) | 0.21 (0.14-0.31) | 15 (10-22) | 0.19 (0.13-0.29) |
| Luxembourg | Male | 10095 (9841-10360) | 1353 (1118-1598) | 13.38 (11.14-15.80) | 83 (67-118) | 0.83 (0.66-1.17) | 1305 (1080-1545) | 12.91 (10.76-15.29) | 32 (25-47) | 0.32 (0.25-0.47) | 31 (23-44) | 0.30 (0.23-0.43) |
| Macedonia | Female | 113649 (108357-118670) | 23625 (19350-27938) | 20.77 (17.16-24.46) | 15358 (12180-18649) | 13.49 (10.83-16.32) | 10874 (8552-13627) | 9.58 (7.56-12.00) | 8289 (6267-10684) | 7.29 (5.55-9.42) | 7385 (5472-9741) | 6.51 (4.87-8.60) |
| Macedonia | Male | 118609 (112366-124431) | 31317 (26226-35599) | 26.48 (22.21-29.86) | 21087 (17091-25091) | 17.81 (14.50-21.02) | 15168 (12062-18443) | 12.78 (10.27-15.53) | 9284 (6959-12086) | 7.84 (5.94-10.14) | 7966 (5862-10342) | 6.73 (4.94-8.77) |
| Madagascar | Female | 453381 (421530-487942) | 90553 (68775-111793) | 19.97 (15.33-24.12) | 64567 (47714-80253) | 14.17 (10.79-17.44) | 38850 (28360-49917) | 8.61 (6.38-10.72) | 13943 (9534-19258) | 3.07 (2.14-4.21) | 7190 (4624-10711) | 1.58 (1.01-2.37) |
| Madagascar | Male | 513843 (471913-552723) | 64985 (47492-83053) | 12.67 (9.23-16.10) | 47860 (34045-60980) | 9.30 (6.71-11.77) | 28995 (20768-37479) | 5.67 (4.05-7.32) | 6525 (4265-9257) | 1.27 (0.84-1.81) | 3399 (1989-5338) | 0.66 (0.40-1.04) |
| Malawi | Female | 191850 (179176-205755) | 36252 (28531-43085) | 18.86 (15.11-22.24) | 25450 (19691-30987) | 13.29 (10.39-16.08) | 15493 (11644-19161) | 8.02 (6.18-9.90) | 7380 (5569-9550) | 3.84 (2.89-4.89) | 5214 (3702-6860) | 2.72 (1.92-3.60) |
| Malawi | Male | 251747 (234342-268508) | 32535 (24689-40757) | 12.95 (9.96-16.17) | 23609 (17726-30691) | 9.41 (7.02-12.08) | 14292 (10579-18728) | 5.69 (4.23-7.42) | 3214 (2320-4383) | 1.28 (0.92-1.74) | 2209 (1500-3074) | 0.88 (0.61-1.24) |
| Malaysia | Female | 588651 (560422-615771) | 116287 (96620-135815) | 19.79 (16.52-22.96) | 71688 (57938-85413) | 12.17 (10.00-14.52) | 56991 (46794-68299) | 9.70 (7.89-11.52) | 25710 (20303-32039) | 4.37 (3.47-5.43) | 23283 (18319-29498) | 3.97 (3.14-4.95) |
| Malaysia | Male | 973312 (933089-1011756) | 227866 (187824-259702) | 23.43 (19.59-26.52) | 141896 (114849-167367) | 14.55 (11.79-17.11) | 112370 (89906-134977) | 11.56 (9.23-13.89) | 39652 (30348-48875) | 4.06 (3.13-5.00) | 35546 (27218-44542) | 3.65 (2.82-4.57) |
| Maldives | Female | 3718 (3539-3886) | 711 (599-832) | 19.14 (16.13-22.15) | 446 (371-538) | 12.01 (9.99-14.39) | 346 (281-421) | 9.28 (7.49-11.18) | 129 (98-165) | 3.45 (2.64-4.46) | 101 (73-134) | 2.72 (1.98-3.57) |
| Maldives | Male | 7901 (7506-8273) | 1715 (1380-2011) | 21.67 (17.37-25.23) | 1087 (862-1316) | 13.75 (10.92-16.55) | 846 (655-1034) | 10.75 (8.26-13.01) | 192 (145-247) | 2.44 (1.82-3.11) | 144 (105-189) | 1.83 (1.34-2.40) |
| Mali | Female | 229508 (211606-246546) | 27286 (19521-34455) | 11.96 (8.47-15.03) | 19794 (14218-25595) | 8.66 (6.27-11.09) | 11950 (8543-15730) | 5.24 (3.79-6.80) | 5930 (4037-8214) | 2.58 (1.75-3.55) | 4157 (2764-5985) | 1.82 (1.19-2.57) |
| Mali | Male | 167970 (155603-180457) | 19372 (13407-25094) | 11.59 (8.09-14.59) | 13985 (9599-18023) | 8.33 (5.77-10.51) | 8350 (5878-11189) | 5.01 (3.52-6.63) | 2852 (1835-4031) | 1.70 (1.11-2.39) | 2049 (1247-3000) | 1.22 (0.73-1.76) |
| Malta | Female | 9668 (9302-10034) | 974 (740-1225) | 10.06 (7.70-12.51) | 449 (342-591) | 4.65 (3.56-6.13) | 671 (500-842) | 6.94 (5.17-8.69) | 199 (146-280) | 2.06 (1.51-2.84) | 194 (140-271) | 2.01 (1.46-2.80) |
| Malta | Male | 10170 (9905-10454) | 1543 (1278-1818) | 15.16 (12.55-17.93) | 694 (547-862) | 6.81 (5.38-8.51) | 1031 (828-1234) | 10.15 (8.14-12.12) | 298 (223-390) | 2.94 (2.20-3.83) | 291 (219-378) | 2.87 (2.16-3.70) |
| Marshall Islands | Female | 1709 (1568-1841) | 215 (150-282) | 12.60 (8.85-16.14) | 145 (98-192) | 8.48 (5.78-11.12) | 102 (70-139) | 6.01 (4.12-8.00) | 37 (24-50) | 2.16 (1.44-2.93) | 27 (18-38) | 1.59 (1.04-2.22) |
| Marshall Islands | Male | 2984 (2748-3216) | 417 (295-532) | 14.04 (10.01-17.53) | 285 (200-369) | 9.60 (6.76-12.25) | 202 (142-268) | 6.76 (4.81-8.90) | 51 (35-68) | 1.71 (1.17-2.27) | 35 (24-48) | 1.18 (0.78-1.61) |
| Mauritania | Female | 65573 (60989-70142) | 8056 (6025-10114) | 12.31 (9.37-15.31) | 5752 (4213-7348) | 8.82 (6.43-11.17) | 3578 (2680-4721) | 5.46 (4.08-7.03) | 1617 (1078-2302) | 2.47 (1.64-3.43) | 1038 (615-1544) | 1.59 (0.97-2.35) |
| Mauritania | Male | 52610 (48563-56599) | 6394 (4809-7968) | 12.20 (9.27-14.99) | 4507 (3397-5746) | 8.60 (6.45-10.76) | 2827 (2088-3630) | 5.37 (3.95-6.84) | 847 (546-1205) | 1.60 (1.05-2.26) | 570 (351-859) | 1.09 (0.66-1.60) |
| Mauritius | Female | 30810 (30118-31558) | 6268 (5262-7319) | 20.37 (17.08-23.79) | 3990 (3335-4736) | 12.95 (10.84-15.35) | 3032 (2463-3660) | 9.83 (8.01-11.86) | 1653 (1336-2026) | 5.36 (4.36-6.54) | 1591 (1278-1963) | 5.17 (4.17-6.32) |
| Mauritius | Male | 45591 (44515-46566) | 10702 (8697-12397) | 23.55 (19.07-27.13) | 6878 (5483-8253) | 15.12 (12.04-18.14) | 5236 (4068-6362) | 11.48 (9.01-13.95) | 2248 (1733-2822) | 4.94 (3.80-6.18) | 2093 (1598-2645) | 4.59 (3.53-5.83) |
| Mexico | Female | 1576870 (1532630-1621943) | 190373 (147355-227528) | 12.10 (9.40-14.42) | 121105 (92793-148591) | 7.68 (5.84-9.39) | 94049 (71502-116429) | 5.96 (4.55-7.38) | 44667 (33439-55935) | 2.83 (2.12-3.56) | 40616 (30898-51232) | 2.59 (1.96-3.25) |
| Mexico | Male | 2237996 (2148577-2326938) | 345962 (279274-404085) | 15.51 (12.45-18.09) | 221681 (177447-266884) | 9.93 (7.85-11.99) | 173195 (136179-211404) | 7.78 (6.11-9.40) | 50969 (40067-63835) | 2.28 (1.79-2.84) | 45021 (34481-56151) | 2.01 (1.53-2.52) |
| Moldova | Female | 196618 (190962-201853) | 8577 (4263-13877) | 4.37 (2.15-7.11) | 7421 (3792-11457) | 3.79 (1.92-5.84) | 5721 (3066-8714) | 2.90 (1.57-4.41) | 3547 (1615-5828) | 1.81 (0.83-2.92) | 2758 (1203-4677) | 1.40 (0.62-2.35) |
| Moldova | Male | 217914 (211932-223837) | 29194 (22435-34901) | 13.39 (10.36-15.95) | 19731 (15057-24761) | 9.03 (6.87-11.32) | 13377 (10164-16908) | 6.14 (4.67-7.75) | 7274 (5296-9708) | 3.33 (2.42-4.44) | 4531 (3100-6245) | 2.08 (1.42-2.86) |
| Mongolia | Female | 59190 (56465-61680) | 7296 (5740-8763) | 12.27 (9.63-14.82) | 5090 (3943-6223) | 8.58 (6.69-10.46) | 3417 (2646-4233) | 5.76 (4.46-7.14) | 2148 (1610-2716) | 3.63 (2.71-4.58) | 1909 (1445-2440) | 3.22 (2.42-4.10) |
| Mongolia | Male | 98282 (93400-102795) | 19576 (15975-23271) | 20.00 (16.30-23.30) | 13183 (10492-16060) | 13.42 (10.71-16.23) | 8800 (6938-10958) | 8.99 (7.06-11.02) | 4583 (3586-5779) | 4.64 (3.67-5.83) | 3384 (2626-4317) | 3.45 (2.69-4.40) |
| Montenegro | Female | 33372 (31890-34845) | 6853 (5571-8157) | 20.62 (16.93-24.14) | 4304 (3389-5360) | 12.92 (10.19-15.90) | 3264 (2544-4099) | 9.78 (7.73-12.13) | 2336 (1787-3018) | 7.02 (5.35-9.05) | 2089 (1530-2819) | 6.26 (4.61-8.38) |
| Montenegro | Male | 36630 (35111-38238) | 9389 (7957-10716) | 25.64 (21.83-28.96) | 6114 (4992-7288) | 16.66 (13.70-19.79) | 4696 (3770-5720) | 12.88 (10.32-15.48) | 2690 (2043-3386) | 7.33 (5.65-9.14) | 2278 (1685-3014) | 6.21 (4.62-8.12) |
| Morocco | Female | 1400920 (1310431-1488416) | 9650 (2551-25030) | 0.69 (0.18-1.81) | 8867 (1962-23566) | 0.64 (0.14-1.70) | 7481 (1544-19857) | 0.54 (0.11-1.41) | 1603 (60-6256) | 0.12 (0.00-0.45) | 1163 (25-5336) | 0.08 (0.00-0.39) |
| Morocco | Male | 1424566 (1325676-1512300) | 100035 (63560-142754) | 7.05 (4.48-9.90) | 77174 (50015-105593) | 5.45 (3.56-7.45) | 52789 (34449-71798) | 3.70 (2.45-5.04) | 14612 (8732-22216) | 1.03 (0.61-1.54) | 9627 (5299-15175) | 0.67 (0.38-1.06) |
| Mozambique | Female | 320026 (292999-346813) | 66428 (51450-80920) | 20.81 (16.85-24.58) | 47169 (36223-58148) | 14.75 (11.72-17.93) | 28412 (21341-35904) | 8.89 (6.93-10.99) | 9947 (7116-13478) | 3.11 (2.25-4.10) | 5336 (3433-7720) | 1.67 (1.10-2.40) |
| Mozambique | Male | 515082 (476787-555527) | 71589 (53086-91901) | 13.85 (10.54-17.53) | 52483 (38662-68359) | 10.19 (7.60-13.03) | 31651 (23062-41869) | 6.13 (4.52-8.06) | 5963 (4030-8756) | 1.16 (0.79-1.68) | 2941 (1794-4422) | 0.57 (0.35-0.85) |
| Myanmar | Female | 1614734 (1513572-1717992) | 320023 (254900-380077) | 19.85 (15.98-23.23) | 221618 (173390-266405) | 13.70 (10.82-16.40) | 135970 (103594-168483) | 8.43 (6.60-10.24) | 74912 (56536-94244) | 4.65 (3.56-5.86) | 48440 (35651-63808) | 3.00 (2.20-3.95) |
| Myanmar | Male | 1897077 (1775764-2015946) | 447853 (351374-525146) | 23.66 (18.72-27.50) | 311976 (239705-377720) | 16.44 (12.71-19.86) | 191981 (149761-239068) | 10.10 (7.87-12.62) | 76307 (56291-97890) | 4.03 (2.92-5.17) | 43960 (31260-58711) | 2.31 (1.65-3.09) |
| Namibia | Female | 38369 (35647-41152) | 3485 (2244-4722) | 9.09 (5.95-12.25) | 2625 (1734-3514) | 6.85 (4.52-9.06) | 1850 (1244-2455) | 4.81 (3.26-6.37) | 851 (532-1198) | 2.21 (1.41-3.12) | 754 (454-1062) | 1.96 (1.18-2.77) |
| Namibia | Male | 43856 (40875-46760) | 4277 (2816-5796) | 9.72 (6.45-13.08) | 3121 (2067-4315) | 7.13 (4.78-9.75) | 2153 (1479-2990) | 4.90 (3.40-6.76) | 723 (428-1087) | 1.65 (0.98-2.44) | 636 (363-976) | 1.45 (0.81-2.20) |
| Nauru | Female | 446 (417-476) | 64 (50-77) | 14.33 (11.30-17.02) | 41 (31-51) | 9.23 (7.09-11.32) | 33 (26-42) | 7.47 (5.79-9.22) | 13 (9-17) | 2.86 (2.12-3.75) | 8 (5-10) | 1.68 (1.16-2.31) |
| Nauru | Male | 693 (643-749) | 104 (79-128) | 14.95 (11.69-18.31) | 67 (50-85) | 9.68 (7.26-12.03) | 54 (39-70) | 7.84 (5.79-10.02) | 17 (13-23) | 2.50 (1.85-3.29) | 11 (8-15) | 1.59 (1.12-2.13) |
| Nepal | Female | 496762 (459425-531903) | 57456 (37536-77029) | 11.55 (7.48-15.29) | 40844 (25679-55836) | 8.23 (5.28-11.14) | 24886 (15663-34794) | 5.04 (3.16-6.91) | 7469 (4632-10602) | 1.51 (0.94-2.09) | 5284 (3259-7684) | 1.07 (0.66-1.52) |
| Nepal | Male | 667903 (623579-713866) | 101170 (80005-121269) | 15.21 (12.11-18.06) | 71496 (55187-88109) | 10.72 (8.37-12.96) | 43533 (32516-54343) | 6.52 (4.91-8.16) | 11156 (8267-14273) | 1.67 (1.25-2.14) | 6477 (4632-8492) | 0.97 (0.70-1.28) |
| Netherlands | Female | 256327 (248849-264320) | 17346 (12139-22893) | 6.77 (4.73-8.89) | 5171 (3349-7621) | 2.01 (1.30-2.96) | 15156 (10755-19791) | 5.93 (4.21-7.69) | 1971 (1173-3159) | 0.77 (0.46-1.23) | 1650 (1003-2614) | 0.64 (0.40-1.02) |
| Netherlands | Male | 306055 (299992-313244) | 36049 (29464-42850) | 11.81 (9.58-14.03) | 9613 (6577-12970) | 3.14 (2.13-4.22) | 29622 (23616-35815) | 9.67 (7.77-11.73) | 3433 (2212-4999) | 1.12 (0.72-1.62) | 3136 (2032-4514) | 1.03 (0.66-1.48) |
| New Zealand | Female | 70351 (68253-72481) | 4340 (2919-5867) | 6.21 (4.20-8.30) | 1690 (1153-2327) | 2.41 (1.65-3.30) | 3634 (2472-4832) | 5.19 (3.51-6.86) | 696 (452-1026) | 0.99 (0.64-1.44) | 623 (393-898) | 0.88 (0.56-1.26) |
| New Zealand | Male | 95625 (93782-97555) | 11755 (9536-13694) | 12.32 (10.03-14.29) | 3938 (2930-5110) | 4.11 (3.08-5.33) | 9102 (7265-10802) | 9.50 (7.60-11.28) | 1358 (989-1825) | 1.42 (1.04-1.91) | 1201 (863-1604) | 1.26 (0.90-1.68) |
| Nicaragua | Female | 57042 (53950-60026) | 8825 (7216-10398) | 15.55 (12.86-17.97) | 6194 (4981-7415) | 10.85 (8.71-12.91) | 3838 (3030-4705) | 6.75 (5.36-8.24) | 2499 (1887-3142) | 4.39 (3.36-5.41) | 2308 (1709-2935) | 4.05 (3.02-5.13) |
| Nicaragua | Male | 62514 (58838-66261) | 13061 (10900-15387) | 20.93 (17.42-24.31) | 8944 (7349-10658) | 14.29 (11.79-16.78) | 5599 (4547-6815) | 8.95 (7.30-10.83) | 2444 (1848-3116) | 3.89 (3.00-5.00) | 2201 (1648-2862) | 3.53 (2.67-4.60) |
| Niger | Female | 215763 (197653-234125) | 27049 (19519-34409) | 12.57 (9.17-15.60) | 19516 (13796-25374) | 9.14 (6.47-11.56) | 11716 (8380-15650) | 5.45 (3.93-7.04) | 3769 (2340-5516) | 1.75 (1.09-2.50) | 1773 (970-2793) | 0.82 (0.46-1.29) |
| Niger | Male | 165814 (151192-180511) | 20735 (14938-26057) | 12.52 (9.20-15.41) | 14885 (10715-19175) | 9.00 (6.67-11.37) | 8928 (6300-11743) | 5.39 (3.88-6.92) | 1693 (1034-2501) | 1.03 (0.64-1.51) | 1111 (656-1706) | 0.67 (0.39-1.02) |
| Nigeria | Female | 1927047 (1808229-2053310) | 190074 (135365-247308) | 9.86 (7.07-12.82) | 141212 (102183-184047) | 7.31 (5.34-9.48) | 90280 (65670-117999) | 4.67 (3.42-6.05) | 37171 (25546-50603) | 1.93 (1.33-2.63) | 21427 (14341-29585) | 1.11 (0.73-1.51) |
| Nigeria | Male | 2099421 (1969348-2224653) | 227989 (167580-286051) | 10.90 (8.04-13.64) | 165136 (120305-209889) | 7.89 (5.82-9.96) | 103776 (76615-132247) | 4.93 (3.66-6.31) | 30183 (20901-40938) | 1.45 (1.01-1.92) | 20898 (14344-28808) | 1.00 (0.69-1.37) |
| Norway | Female | 72560 (70487-74616) | 2900 (1582-4380) | 3.99 (2.19-5.95) | 396 (219-782) | 0.55 (0.30-1.07) | 2861 (1558-4259) | 3.93 (2.16-5.86) | 165 (85-357) | 0.23 (0.12-0.49) | 113 (56-245) | 0.15 (0.08-0.34) |
| Norway | Male | 89823 (88249-91460) | 11053 (8968-12951) | 12.30 (10.08-14.38) | 996 (657-1674) | 1.11 (0.73-1.86) | 10435 (8379-12277) | 11.64 (9.35-13.71) | 407 (248-761) | 0.45 (0.28-0.85) | 354 (218-667) | 0.39 (0.24-0.74) |
| Oman | Female | 35750 (33680-37727) | 370 (87-891) | 1.05 (0.25-2.49) | 301 (72-715) | 0.84 (0.20-2.01) | 301 (69-745) | 0.84 (0.20-2.07) | 49 (2-223) | 0.14 (0.01-0.63) | 45 (1-203) | 0.12 (0.00-0.56) |
| Oman | Male | 74155 (69814-78248) | 6958 (4855-9012) | 9.40 (6.54-12.04) | 4422 (3041-5749) | 6.00 (4.18-7.70) | 4384 (3007-5844) | 5.91 (4.11-7.72) | 809 (501-1159) | 1.09 (0.69-1.56) | 695 (426-997) | 0.94 (0.57-1.37) |
| Pakistan | Female | 3780619 (3513364-4015890) | 611384 (493266-721916) | 16.18 (13.23-18.73) | 430351 (339115-523339) | 11.43 (9.08-13.70) | 264740 (203281-327743) | 6.99 (5.51-8.61) | 125639 (94178-159961) | 3.32 (2.53-4.19) | 104677 (77879-135078) | 2.76 (2.07-3.52) |
| Pakistan | Male | 4618323 (4318530-4922909) | 772034 (625990-912928) | 16.74 (13.72-19.61) | 544796 (437625-664841) | 11.81 (9.49-14.22) | 333039 (263139-420971) | 7.26 (5.68-9.10) | 86705 (63870-115428) | 1.88 (1.41-2.47) | 65097 (47617-88242) | 1.41 (1.03-1.91) |
| Palau | Female | 585 (548-621) | 93 (73-113) | 15.84 (12.61-19.08) | 57 (44-72) | 9.73 (7.50-12.11) | 48 (36-60) | 8.11 (6.32-10.21) | 23 (17-30) | 3.88 (2.85-5.02) | 19 (14-26) | 3.30 (2.38-4.37) |
| Palau | Male | 1362 (1272-1452) | 214 (160-263) | 15.76 (11.76-19.27) | 134 (99-170) | 9.86 (7.57-12.33) | 112 (82-144) | 8.23 (6.09-10.59) | 38 (27-50) | 2.78 (2.02-3.66) | 26 (19-36) | 1.95 (1.37-2.63) |
| Panama | Female | 41827 (40440-43198) | 6066 (5056-7077) | 14.51 (12.05-16.85) | 3657 (2960-4430) | 8.74 (7.07-10.54) | 3262 (2616-3934) | 7.77 (6.20-9.43) | 1512 (1196-1906) | 3.61 (2.85-4.56) | 1354 (1046-1740) | 3.23 (2.53-4.10) |
| Panama | Male | 60649 (57934-63061) | 11880 (9874-13856) | 19.60 (16.37-22.76) | 7008 (5680-8377) | 11.53 (9.39-13.74) | 6235 (5042-7431) | 10.29 (8.39-12.32) | 2300 (1797-2872) | 3.80 (2.99-4.72) | 1919 (1490-2449) | 3.16 (2.49-4.06) |
| Papua New Guinea | Female | 173538 (161152-184890) | 22134 (14695-28271) | 12.73 (8.52-16.06) | 15699 (10283-20274) | 9.07 (5.95-11.71) | 10031 (6695-13394) | 5.79 (3.87-7.66) | 2669 (1579-4010) | 1.55 (0.92-2.30) | 1609 (841-2581) | 0.93 (0.49-1.48) |
| Papua New Guinea | Male | 238311 (220477-256740) | 32830 (22519-42403) | 13.76 (9.53-17.49) | 23208 (15674-30520) | 9.71 (6.72-12.56) | 14880 (10132-19948) | 6.25 (4.17-8.30) | 2513 (1466-3814) | 1.05 (0.61-1.57) | 1683 (859-2635) | 0.70 (0.36-1.10) |
| Paraguay | Female | 87785 (83228-92264) | 13234 (10789-15400) | 15.08 (12.34-17.27) | 8870 (7000-10527) | 10.09 (7.98-11.94) | 6122 (4804-7501) | 6.96 (5.50-8.48) | 4197 (3205-5158) | 4.80 (3.69-5.85) | 3392 (2558-4326) | 3.85 (2.93-4.90) |
| Paraguay | Male | 129695 (121771-137267) | 23621 (18522-28051) | 18.22 (14.52-21.50) | 15684 (12017-18899) | 12.12 (9.28-14.55) | 10945 (8294-13454) | 8.43 (6.55-10.32) | 5915 (4398-7508) | 4.58 (3.37-5.76) | 4590 (3308-5982) | 3.53 (2.59-4.64) |
| Peru | Female | 268798 (254069-283706) | 32985 (25109-39917) | 12.28 (9.39-14.78) | 21991 (16284-27264) | 8.16 (6.01-10.01) | 15503 (11183-19504) | 5.76 (4.22-7.28) | 5079 (3828-6405) | 1.90 (1.42-2.35) | 4628 (3457-5822) | 1.72 (1.30-2.14) |
| Peru | Male | 357330 (335496-377675) | 59773 (47744-71768) | 16.80 (13.48-20.01) | 39851 (30896-48307) | 11.16 (8.69-13.39) | 27977 (21689-34164) | 7.81 (6.14-9.69) | 6020 (4651-7616) | 1.69 (1.31-2.12) | 5096 (3891-6526) | 1.43 (1.10-1.81) |
| Philippines | Female | 2239939 (2161806-2318358) | 418183 (319829-502627) | 18.69 (14.31-22.46) | 283241 (215053-350724) | 12.70 (9.63-15.66) | 185667 (138424-230644) | 8.28 (6.21-10.30) | 82338 (62452-103880) | 3.68 (2.79-4.65) | 61470 (45163-78214) | 2.75 (2.01-3.49) |
| Philippines | Male | 3517579 (3356865-3664562) | 798105 (587374-943182) | 22.67 (16.93-26.70) | 547450 (404376-671923) | 15.60 (11.52-18.98) | 356879 (262972-447769) | 10.18 (7.41-12.74) | 114655 (80459-146631) | 3.26 (2.27-4.16) | 76733 (53486-100356) | 2.19 (1.52-2.88) |
| Poland | Female | 1215643 (1186399-1250854) | 206079 (168226-243920) | 16.94 (13.85-20.02) | 117505 (93576-144239) | 9.67 (7.71-11.78) | 112267 (88379-137597) | 9.22 (7.28-11.28) | 72195 (56350-90021) | 5.94 (4.62-7.39) | 69707 (54703-87156) | 5.73 (4.49-7.14) |
| Poland | Male | 1457690 (1425094-1490608) | 320030 (257276-369196) | 21.98 (17.78-25.34) | 184094 (145661-220283) | 12.62 (10.04-15.16) | 174724 (141181-210399) | 12.00 (9.74-14.46) | 90087 (70946-111072) | 6.18 (4.83-7.65) | 82802 (64390-102526) | 5.67 (4.44-7.08) |
| Portugal | Female | 222753 (215850-230584) | 12290 (7534-17795) | 5.50 (3.39-7.93) | 8020 (4872-11246) | 3.60 (2.22-5.06) | 8903 (5525-12377) | 3.97 (2.49-5.61) | 3947 (2187-5967) | 1.77 (1.00-2.66) | 3840 (2143-5739) | 1.72 (0.96-2.57) |
| Portugal | Male | 244285 (239428-249690) | 32124 (26482-38111) | 13.15 (10.83-15.59) | 17206 (13599-20986) | 7.03 (5.58-8.60) | 19244 (15612-23720) | 7.90 (6.43-9.67) | 7221 (5460-9130) | 2.96 (2.24-3.74) | 6820 (5206-8636) | 2.79 (2.13-3.53) |
| Puerto Rico | Female | 58395 (56521-60391) | 5974 (4562-7453) | 10.24 (7.71-12.72) | 2916 (2168-3790) | 4.99 (3.75-6.42) | 4062 (3060-5109) | 6.97 (5.25-8.68) | 1421 (1024-1934) | 2.43 (1.77-3.30) | 1204 (861-1658) | 2.07 (1.47-2.81) |
| Puerto Rico | Male | 79980 (76920-83063) | 11422 (9154-13407) | 14.28 (11.53-16.76) | 5418 (4194-6706) | 6.77 (5.24-8.44) | 7541 (5991-9148) | 9.44 (7.51-11.42) | 1909 (1395-2572) | 2.39 (1.75-3.22) | 1604 (1110-2179) | 2.00 (1.41-2.71) |
| Qatar | Female | 6419 (6056-6767) | 41 (0-129) | 0.64 (0.00-1.99) | 18 (0-52) | 0.28 (0.00-0.81) | 40 (0-124) | 0.61 (0.00-1.90) | 5 (0-21) | 0.07 (0.00-0.33) | 4 (0-20) | 0.06 (0.00-0.31) |
| Qatar | Male | 26081 (23882-27967) | 2418 (1488-3396) | 9.32 (5.80-12.85) | 570 (316-939) | 2.20 (1.25-3.53) | 2186 (1353-3060) | 8.42 (5.15-11.60) | 110 (60-183) | 0.42 (0.23-0.69) | 96 (51-162) | 0.37 (0.20-0.62) |
| Romania | Female | 1085613 (1057593-1113991) | 233565 (191821-277330) | 21.53 (17.67-25.49) | 138705 (108249-171089) | 12.77 (9.98-15.81) | 121242 (96595-150952) | 11.19 (8.92-13.94) | 74813 (57007-94479) | 6.89 (5.25-8.67) | 65775 (49872-83839) | 6.06 (4.60-7.74) |
| Romania | Male | 1176744 (1148756-1204107) | 315462 (268557-356775) | 26.87 (22.93-30.15) | 196759 (158396-231348) | 16.69 (13.55-19.62) | 172282 (141728-206646) | 14.67 (12.04-17.48) | 90485 (70657-109590) | 7.69 (6.05-9.30) | 81193 (63539-99005) | 6.87 (5.41-8.38) |
| Russia | Female | 8019517 (7832813-8202479) | 915944 (718371-1110501) | 11.40 (8.94-13.82) | 570821 (444212-708529) | 7.11 (5.55-8.83) | 468090 (364725-583989) | 5.82 (4.55-7.32) | 298254 (223443-379032) | 3.73 (2.78-4.73) | 213460 (156840-274106) | 2.66 (1.96-3.43) |
| Russia | Male | 8441040 (8235599-8652467) | 1615958 (1302753-1883733) | 19.14 (15.46-22.20) | 986776 (775119-1196185) | 11.70 (9.19-14.10) | 810264 (630285-1002332) | 9.63 (7.49-11.80) | 396119 (299299-491792) | 4.68 (3.57-5.85) | 278577 (206966-347758) | 3.30 (2.44-4.15) |
| Rwanda | Female | 150790 (138574-164091) | 28148 (21378-35141) | 18.76 (14.35-22.48) | 19910 (14872-24739) | 13.20 (10.00-16.11) | 12046 (8897-15470) | 7.98 (5.96-10.06) | 2877 (1951-4090) | 1.90 (1.30-2.69) | 1337 (795-2091) | 0.89 (0.52-1.38) |
| Rwanda | Male | 132545 (120864-145004) | 17024 (12495-21760) | 12.83 (9.54-16.32) | 12319 (8895-15965) | 9.28 (6.66-11.96) | 7444 (5324-9835) | 5.63 (4.03-7.38) | 1097 (684-1630) | 0.83 (0.52-1.25) | 658 (379-1059) | 0.50 (0.29-0.78) |
| Saint Kitts and Nevis | Female | 1231 (1190-1268) | 121 (87-154) | 9.88 (7.13-12.51) | 75 (54-96) | 6.10 (4.45-7.81) | 71 (52-91) | 5.82 (4.25-7.37) | 36 (25-48) | 2.94 (2.06-3.87) | 33 (23-45) | 2.66 (1.83-3.62) |
| Saint Kitts and Nevis | Male | 1812 (1715-1911) | 281 (214-336) | 15.52 (11.91-18.38) | 164 (123-208) | 9.09 (6.70-11.42) | 156 (116-200) | 8.64 (6.35-10.82) | 53 (38-72) | 2.94 (2.09-3.95) | 43 (29-58) | 2.37 (1.61-3.20) |
| Saint Lucia | Female | 3508 (3402-3611) | 366 (279-461) | 10.46 (8.00-13.12) | 244 (183-307) | 6.95 (5.25-8.82) | 194 (146-248) | 5.53 (4.19-7.06) | 124 (92-163) | 3.54 (2.63-4.62) | 105 (77-144) | 2.99 (2.20-4.08) |
| Saint Lucia | Male | 4379 (4220-4534) | 651 (515-785) | 14.89 (11.76-17.86) | 417 (322-514) | 9.53 (7.43-11.72) | 329 (258-416) | 7.52 (5.89-9.45) | 137 (103-179) | 3.14 (2.35-4.06) | 101 (74-135) | 2.30 (1.70-3.11) |
| Saint Vincent and the Grenadines | Female | 2801 (2726-2878) | 302 (217-388) | 10.76 (7.80-13.84) | 208 (153-266) | 7.42 (5.52-9.50) | 155 (114-201) | 5.53 (4.07-7.12) | 94 (67-128) | 3.38 (2.40-4.58) | 79 (55-109) | 2.83 (1.98-3.91) |
| Saint Vincent and the Grenadines | Male | 3675 (3575-3773) | 574 (447-686) | 15.63 (12.22-18.48) | 377 (289-464) | 10.25 (7.84-12.62) | 279 (214-351) | 7.61 (5.82-9.52) | 115 (85-153) | 3.14 (2.32-4.12) | 89 (65-119) | 2.42 (1.76-3.25) |
| Samoa | Female | 5573 (5256-5879) | 276 (188-379) | 4.96 (3.40-6.75) | 202 (131-287) | 3.63 (2.37-5.10) | 140 (90-203) | 2.51 (1.60-3.64) | 49 (28-77) | 0.87 (0.50-1.37) | 19 (8-33) | 0.34 (0.15-0.59) |
| Samoa | Male | 7729 (7263-8201) | 348 (226-506) | 4.50 (2.88-6.53) | 265 (164-401) | 3.42 (2.09-5.22) | 187 (111-282) | 2.41 (1.43-3.65) | 52 (30-80) | 0.67 (0.39-1.04) | 34 (19-57) | 0.44 (0.25-0.75) |
| Sao Tome and Principe | Female | 3011 (2841-3173) | 373 (277-459) | 12.37 (9.36-15.10) | 265 (197-333) | 8.79 (6.67-10.89) | 166 (123-214) | 5.53 (4.14-6.96) | 91 (64-118) | 3.01 (2.16-3.92) | 59 (40-79) | 1.95 (1.38-2.62) |
| Sao Tome and Principe | Male | 2861 (2679-3038) | 366 (279-451) | 12.78 (9.71-15.52) | 258 (196-329) | 9.03 (6.88-11.30) | 161 (121-208) | 5.64 (4.31-7.21) | 50 (36-68) | 1.76 (1.28-2.35) | 29 (20-40) | 1.01 (0.69-1.40) |
| Saudi Arabia | Female | 529245 (492938-564883) | 4334 (116-11051) | 0.82 (0.02-2.05) | 3510 (116-8609) | 0.66 (0.02-1.69) | 3614 (116-9285) | 0.68 (0.02-1.75) | 402 (7-1765) | 0.08 (0.00-0.33) | 334 (4-1445) | 0.06 (0.00-0.27) |
| Saudi Arabia | Male | 1149832 (1067622-1233676) | 105587 (63473-149890) | 9.22 (5.53-12.94) | 63800 (39001-87483) | 5.56 (3.43-7.60) | 70401 (44560-98206) | 6.12 (3.87-8.33) | 10273 (6155-14745) | 0.89 (0.53-1.30) | 8102 (4892-11910) | 0.71 (0.41-1.04) |
| Senegal | Female | 226255 (211601-241901) | 27787 (20546-34377) | 12.27 (9.16-14.97) | 19796 (14491-24893) | 8.75 (6.51-10.92) | 12332 (8815-15804) | 5.42 (3.96-7.01) | 5311 (3666-7142) | 2.34 (1.62-3.11) | 2845 (1763-4158) | 1.26 (0.78-1.84) |
| Senegal | Male | 238352 (223560-253784) | 29614 (22096-36946) | 12.42 (9.23-15.37) | 21021 (15687-26869) | 8.80 (6.57-11.25) | 12872 (9634-16934) | 5.41 (4.04-7.08) | 3556 (2402-5040) | 1.49 (1.00-2.11) | 1974 (1289-3016) | 0.83 (0.54-1.27) |
| Serbia | Female | 531262 (509922-554230) | 110828 (91085-130870) | 20.80 (17.25-24.43) | 69820 (55630-85800) | 13.16 (10.43-16.16) | 52705 (41716-64972) | 9.94 (7.90-12.23) | 42400 (32537-52912) | 7.98 (6.20-10.01) | 40036 (30508-50611) | 7.53 (5.77-9.45) |
| Serbia | Male | 526293 (505099-549310) | 134834 (115077-152942) | 25.66 (21.98-29.03) | 88778 (72964-105334) | 16.88 (13.92-19.77) | 67700 (54698-81447) | 12.85 (10.33-15.45) | 41886 (32861-51502) | 7.96 (6.25-9.73) | 38076 (30087-47116) | 7.21 (5.79-8.94) |
| Seychelles | Female | 2188 (2096-2277) | 419 (335-504) | 19.13 (15.37-22.88) | 259 (201-319) | 11.82 (9.26-14.48) | 217 (169-272) | 9.93 (7.74-12.41) | 130 (98-167) | 5.96 (4.50-7.62) | 120 (88-155) | 5.46 (4.09-7.02) |
| Seychelles | Male | 3141 (2999-3279) | 643 (517-768) | 20.55 (16.33-24.31) | 398 (309-498) | 12.71 (9.80-15.66) | 333 (259-421) | 10.62 (8.25-13.26) | 152 (116-195) | 4.82 (3.65-6.15) | 133 (100-173) | 4.24 (3.19-5.44) |
| Sierra Leone | Female | 125213 (115840-134102) | 15498 (11410-18921) | 12.41 (9.24-15.08) | 11200 (8339-14049) | 8.97 (6.69-11.17) | 6686 (5000-8398) | 5.39 (4.01-6.70) | 2743 (1883-3755) | 2.19 (1.55-2.96) | 1606 (1048-2284) | 1.28 (0.84-1.82) |
| Sierra Leone | Male | 124718 (115459-133957) | 16049 (12325-19921) | 12.87 (9.99-15.77) | 11509 (8812-14649) | 9.26 (7.11-11.53) | 6861 (5244-8926) | 5.51 (4.22-7.08) | 1747 (1162-2406) | 1.40 (0.94-1.91) | 988 (624-1434) | 0.79 (0.50-1.13) |
| Singapore | Female | 48728 (47442-50030) | 7453 (5405-9218) | 15.33 (11.09-18.87) | 1099 (605-1735) | 2.26 (1.24-3.59) | 6756 (4864-8465) | 13.89 (10.06-17.37) | 489 (257-824) | 1.00 (0.53-1.68) | 469 (243-783) | 0.96 (0.50-1.59) |
| Singapore | Male | 85758 (84088-87539) | 16450 (11685-20068) | 19.17 (13.62-23.38) | 2447 (1336-3955) | 2.86 (1.54-4.61) | 14883 (10460-18484) | 17.38 (12.22-21.50) | 923 (475-1554) | 1.07 (0.56-1.81) | 874 (437-1476) | 1.02 (0.51-1.72) |
| Slovakia | Female | 221091 (211252-229918) | 45088 (37173-52775) | 20.42 (16.80-23.81) | 24158 (19011-29615) | 10.96 (8.65-13.42) | 25452 (20028-31396) | 11.52 (9.12-14.12) | 13754 (10484-17509) | 6.23 (4.78-7.86) | 12879 (9691-16372) | 5.81 (4.39-7.41) |
| Slovakia | Male | 240015 (229525-249517) | 62294 (52906-71253) | 25.95 (22.15-29.38) | 35265 (28509-42595) | 14.70 (12.05-17.72) | 36870 (29883-44516) | 15.40 (12.47-18.44) | 17397 (13541-21758) | 7.25 (5.69-9.03) | 16105 (12343-20529) | 6.68 (5.18-8.50) |
| Slovenia | Female | 45819 (44311-47382) | 9839 (7906-11845) | 21.45 (17.42-25.99) | 4852 (3708-6270) | 10.60 (8.10-13.72) | 6149 (4816-7814) | 13.46 (10.57-16.95) | 2741 (2011-3694) | 6.01 (4.41-8.08) | 2491 (1759-3397) | 5.43 (3.87-7.40) |
| Slovenia | Male | 50177 (48827-51458) | 12580 (10732-14227) | 25.12 (21.45-28.22) | 6500 (5169-7778) | 12.94 (10.44-15.54) | 8116 (6763-9742) | 16.24 (13.47-19.35) | 2901 (2210-3693) | 5.79 (4.41-7.36) | 2491 (1820-3258) | 4.97 (3.64-6.48) |
| Solomon Islands | Female | 16861 (15752-17961) | 2184 (1546-2741) | 13.01 (9.29-16.27) | 1534 (1073-1966) | 9.12 (6.39-11.65) | 967 (661-1256) | 5.77 (3.93-7.46) | 370 (248-507) | 2.20 (1.50-3.00) | 152 (91-227) | 0.90 (0.55-1.35) |
| Solomon Islands | Male | 26671 (24659-28461) | 3406 (2240-4535) | 12.88 (8.29-16.87) | 2411 (1581-3269) | 9.10 (5.95-12.25) | 1545 (977-2106) | 5.79 (3.66-7.89) | 245 (149-366) | 0.92 (0.56-1.36) | 122 (69-194) | 0.46 (0.26-0.74) |
| Somalia | Female | 184056 (165977-203795) | 34981 (26191-44690) | 19.09 (14.33-23.42) | 24902 (18144-31994) | 13.58 (10.04-16.76) | 15002 (10866-19651) | 8.18 (5.85-10.44) | 6375 (4455-8981) | 3.47 (2.45-4.71) | 4498 (2954-6582) | 2.46 (1.62-3.57) |
| Somalia | Male | 194437 (173741-216468) | 24655 (17050-32581) | 12.75 (8.77-16.57) | 18195 (12714-23993) | 9.36 (6.48-12.38) | 11055 (7542-15148) | 5.69 (3.83-7.64) | 2997 (1910-4390) | 1.54 (0.99-2.26) | 2110 (1268-3217) | 1.08 (0.67-1.66) |
| South Africa | Female | 1073945 (1045930-1102161) | 85677 (57977-116930) | 7.96 (5.41-10.95) | 67061 (46513-89744) | 6.25 (4.36-8.36) | 50606 (36251-66322) | 4.73 (3.36-6.16) | 25779 (16990-35847) | 2.40 (1.59-3.36) | 23500 (15292-33012) | 2.19 (1.43-3.06) |
| South Africa | Male | 1023145 (996445-1048041) | 78603 (49747-109286) | 7.67 (4.89-10.67) | 59742 (38908-83035) | 5.84 (3.79-8.09) | 45264 (29709-62520) | 4.42 (2.92-6.06) | 13480 (7487-20752) | 1.32 (0.73-2.02) | 12122 (6564-18520) | 1.18 (0.64-1.79) |
| South Korea | Female | 638582 (608916-672872) | 114855 (90458-138720) | 18.01 (14.00-21.61) | 52000 (37955-65818) | 8.11 (6.03-10.29) | 76723 (58423-96209) | 12.02 (9.16-14.87) | 25634 (18280-33359) | 4.01 (2.91-5.20) | 25582 (18289-33444) | 4.00 (2.89-5.18) |
| South Korea | Male | 757925 (728855-787137) | 146959 (111302-175750) | 19.43 (14.59-23.15) | 67312 (48444-85150) | 8.87 (6.40-11.20) | 98969 (74070-120910) | 13.03 (9.73-15.87) | 26664 (19212-34705) | 3.52 (2.55-4.57) | 26660 (19281-34760) | 3.52 (2.55-4.58) |
| Spain | Female | 717215 (691144-742991) | 17810 (8909-32386) | 2.49 (1.24-4.52) | 12347 (5363-22919) | 1.73 (0.75-3.17) | 14573 (6634-27591) | 2.02 (0.93-3.84) | 4797 (1352-10717) | 0.67 (0.19-1.50) | 4193 (1135-9221) | 0.58 (0.16-1.31) |
| Spain | Male | 876014 (858427-895942) | 62055 (45437-79710) | 7.10 (5.17-9.09) | 33111 (22764-44337) | 3.76 (2.63-5.04) | 42349 (30063-56328) | 4.84 (3.43-6.47) | 10156 (6321-14878) | 1.16 (0.72-1.69) | 8939 (5392-13271) | 1.02 (0.61-1.51) |
| Sri Lanka | Female | 484386 (452248-514220) | 92220 (74642-109492) | 19.09 (15.71-22.19) | 61485 (49370-74069) | 12.68 (10.37-15.06) | 40410 (32144-49443) | 8.37 (6.67-10.15) | 20913 (15909-26583) | 4.31 (3.31-5.46) | 17970 (13450-23382) | 3.71 (2.79-4.79) |
| Sri Lanka | Male | 626172 (577914-675841) | 140360 (116373-165716) | 22.37 (18.70-25.86) | 94346 (76432-113468) | 15.12 (12.40-18.04) | 62170 (50010-77613) | 9.95 (7.95-12.26) | 20483 (15331-27010) | 3.25 (2.44-4.30) | 16594 (12110-22329) | 2.65 (1.96-3.49) |
| Sudan | Female | 756570 (699961-813402) | 10098 (3436-19865) | 1.33 (0.46-2.62) | 8815 (3252-17966) | 1.17 (0.43-2.37) | 6469 (2393-12917) | 0.85 (0.32-1.69) | 926 (177-3077) | 0.12 (0.02-0.41) | 589 (77-2365) | 0.08 (0.01-0.32) |
| Sudan | Male | 976424 (905576-1049304) | 82323 (56112-111654) | 8.48 (5.84-11.27) | 63586 (44672-84681) | 6.57 (4.64-8.61) | 40285 (28048-52325) | 4.10 (2.93-5.35) | 7255 (4336-11117) | 0.74 (0.45-1.13) | 5135 (2900-8132) | 0.53 (0.30-0.83) |
| Suriname | Female | 12445 (11794-13128) | 1140 (803-1452) | 9.15 (6.47-11.66) | 815 (577-1052) | 6.53 (4.64-8.41) | 555 (390-730) | 4.46 (3.18-5.82) | 372 (250-487) | 3.00 (2.01-3.91) | 313 (208-424) | 2.51 (1.71-3.39) |
| Suriname | Male | 17176 (16006-18300) | 2391 (1706-2935) | 13.93 (10.27-17.12) | 1620 (1159-2064) | 9.43 (6.88-12.03) | 1098 (787-1437) | 6.39 (4.63-8.33) | 532 (367-697) | 3.09 (2.13-4.10) | 433 (298-585) | 2.52 (1.74-3.41) |
| Swaziland | Female | 18328 (16707-19951) | 1830 (1240-2449) | 10.02 (6.76-13.21) | 1384 (946-1820) | 7.54 (5.22-9.88) | 953 (656-1252) | 5.17 (3.59-6.74) | 476 (302-671) | 2.61 (1.68-3.61) | 360 (213-512) | 1.98 (1.17-2.79) |
| Swaziland | Male | 21067 (19038-23002) | 2362 (1593-3263) | 11.35 (7.53-15.23) | 1731 (1166-2397) | 8.25 (5.53-11.30) | 1166 (782-1623) | 5.54 (3.77-7.66) | 264 (151-405) | 1.26 (0.72-1.90) | 192 (109-305) | 0.91 (0.51-1.43) |
| Sweden | Female | 198569 (191687-205988) | 15280 (10512-19922) | 7.71 (5.32-9.96) | 4015 (2462-5835) | 2.04 (1.24-2.93) | 13325 (9355-17246) | 6.70 (4.72-8.68) | 1670 (954-2561) | 0.84 (0.48-1.29) | 1379 (764-2161) | 0.69 (0.39-1.08) |
| Sweden | Male | 233269 (225678-240631) | 29940 (24185-34941) | 12.86 (10.45-14.88) | 7206 (5027-10061) | 3.10 (2.16-4.29) | 25004 (19968-29916) | 10.75 (8.61-12.80) | 2846 (1861-4185) | 1.22 (0.80-1.81) | 2499 (1624-3719) | 1.08 (0.70-1.59) |
| Switzerland | Female | 120358 (115433-125233) | 9772 (6535-13169) | 8.10 (5.38-10.92) | 956 (579-2065) | 0.79 (0.48-1.73) | 9350 (6256-12552) | 7.75 (5.21-10.47) | 374 (211-858) | 0.31 (0.18-0.71) | 307 (175-724) | 0.26 (0.15-0.60) |
| Switzerland | Male | 139988 (136417-143544) | 18159 (14404-21679) | 12.96 (10.37-15.48) | 1644 (1049-2837) | 1.17 (0.75-2.02) | 17139 (13667-20512) | 12.24 (9.78-14.65) | 690 (432-1291) | 0.49 (0.31-0.91) | 604 (370-1127) | 0.43 (0.27-0.80) |
| Syria | Female | 445985 (418461-474265) | 2799 (459-8033) | 0.62 (0.10-1.76) | 2589 (432-7639) | 0.58 (0.10-1.69) | 2082 (333-5913) | 0.47 (0.07-1.32) | 563 (18-2882) | 0.13 (0.00-0.64) | 458 (12-2628) | 0.10 (0.00-0.58) |
| Syria | Male | 698714 (647936-751969) | 49803 (30730-71598) | 7.15 (4.35-10.03) | 39010 (24672-54562) | 5.58 (3.41-7.67) | 24278 (15862-34301) | 3.47 (2.29-4.88) | 10936 (6005-16570) | 1.56 (0.86-2.33) | 9086 (5002-13823) | 1.30 (0.71-1.98) |
| Taiwan (Province of China) | Female | 291158 (283564-298764) | 49011 (39277-58383) | 16.86 (13.49-19.95) | 22682 (17448-28380) | 7.78 (5.95-9.70) | 32066 (24609-39155) | 11.00 (8.53-13.42) | 7768 (5814-10317) | 2.67 (2.00-3.55) | 7742 (5793-10109) | 2.65 (1.98-3.47) |
| Taiwan (Province of China) | Male | 494183 (483276-505171) | 80739 (64517-94782) | 16.29 (13.08-19.20) | 37668 (28848-46774) | 7.62 (5.82-9.49) | 53400 (41199-64124) | 10.80 (8.33-13.01) | 11749 (8831-15282) | 2.37 (1.77-3.08) | 11312 (8550-14472) | 2.29 (1.74-2.93) |
| Tajikistan | Female | 185843 (176261-195681) | 21586 (16161-26945) | 11.60 (8.73-14.45) | 15668 (11833-19834) | 8.48 (6.38-10.59) | 9602 (7272-12328) | 5.18 (3.96-6.61) | 5550 (3952-7247) | 2.98 (2.17-3.85) | 4645 (3258-6185) | 2.50 (1.76-3.32) |
| Tajikistan | Male | 208127 (196478-219346) | 39308 (32263-46822) | 18.93 (15.61-22.03) | 27522 (22141-32857) | 13.23 (10.72-15.72) | 16698 (13251-20613) | 8.02 (6.38-9.73) | 7708 (6009-9926) | 3.71 (2.85-4.71) | 4788 (3478-6307) | 2.31 (1.70-3.06) |
| Tanzania | Female | 540396 (503149-577398) | 108561 (85895-131299) | 20.09 (15.91-23.56) | 75950 (58955-92838) | 14.01 (11.00-16.86) | 46173 (35686-58165) | 8.53 (6.67-10.54) | 16864 (12281-22571) | 3.12 (2.28-4.07) | 7356 (4931-10432) | 1.36 (0.92-1.91) |
| Tanzania | Male | 723980 (671127-774913) | 129808 (100921-158352) | 18.01 (14.01-21.49) | 90535 (67513-111939) | 12.52 (9.47-15.26) | 55303 (41024-69858) | 7.67 (5.78-9.47) | 11767 (8138-15890) | 1.63 (1.12-2.16) | 7100 (4657-9992) | 0.99 (0.65-1.36) |
| Thailand | Female | 1222827 (1158155-1292388) | 196068 (146340-236859) | 16.04 (12.09-19.40) | 127560 (94713-157026) | 10.40 (7.81-12.74) | 91096 (66790-113569) | 7.45 (5.45-9.35) | 41094 (30273-52402) | 3.37 (2.48-4.29) | 37428 (26951-48036) | 3.06 (2.22-3.93) |
| Thailand | Male | 1745890 (1641931-1845622) | 363235 (279053-434414) | 20.94 (16.15-24.53) | 239381 (182076-292597) | 13.69 (10.53-16.68) | 172343 (129297-215198) | 9.90 (7.44-12.22) | 52601 (38450-69992) | 3.04 (2.24-3.96) | 44984 (32987-59748) | 2.58 (1.89-3.44) |
| The Bahamas | Female | 7550 (7291-7817) | 838 (596-1092) | 11.13 (7.85-14.40) | 461 (333-603) | 6.12 (4.40-8.00) | 564 (406-737) | 7.48 (5.39-9.77) | 194 (134-261) | 2.57 (1.78-3.46) | 170 (115-233) | 2.26 (1.52-3.10) |
| The Bahamas | Male | 9995 (9528-10400) | 1711 (1333-2067) | 17.20 (13.24-20.62) | 887 (667-1134) | 8.90 (6.58-11.32) | 1089 (825-1392) | 10.92 (8.17-13.86) | 294 (209-395) | 2.96 (2.10-3.96) | 233 (165-322) | 2.33 (1.64-3.19) |
| The Gambia | Female | 32835 (30616-35145) | 4012 (3097-5022) | 12.23 (9.54-15.01) | 2899 (2191-3689) | 8.84 (6.76-10.97) | 1751 (1334-2260) | 5.33 (4.08-6.80) | 854 (605-1154) | 2.60 (1.83-3.44) | 546 (362-763) | 1.66 (1.13-2.31) |
| The Gambia | Male | 37707 (35063-40168) | 4849 (3608-6006) | 12.87 (9.74-15.69) | 3484 (2561-4443) | 9.26 (7.05-11.50) | 2109 (1550-2738) | 5.61 (4.18-7.12) | 626 (426-881) | 1.67 (1.15-2.33) | 415 (270-603) | 1.10 (0.73-1.59) |
| Timor-Leste | Female | 28024 (26179-29801) | 5594 (4417-6722) | 19.94 (16.10-23.87) | 3796 (2988-4680) | 13.57 (10.71-16.41) | 2433 (1883-3069) | 8.68 (6.74-10.77) | 716 (512-983) | 2.55 (1.84-3.45) | 488 (337-687) | 1.75 (1.21-2.46) |
| Timor-Leste | Male | 34608 (32108-37226) | 8171 (6849-9692) | 23.69 (19.87-27.41) | 5565 (4580-6820) | 16.15 (13.36-19.30) | 3551 (2874-4517) | 10.31 (8.41-12.68) | 976 (723-1320) | 2.82 (2.13-3.73) | 632 (458-880) | 1.84 (1.33-2.50) |
| Togo | Female | 123207 (113965-132831) | 15061 (10939-19243) | 12.21 (9.15-15.29) | 10873 (7925-14059) | 8.82 (6.52-11.36) | 6582 (4761-8803) | 5.38 (3.94-7.01) | 2778 (1873-3932) | 2.24 (1.54-3.24) | 1500 (963-2287) | 1.22 (0.78-1.85) |
| Togo | Male | 129672 (120263-139457) | 16840 (12500-21081) | 13.03 (9.61-15.89) | 12106 (8797-15399) | 9.34 (6.90-11.71) | 7313 (5274-9632) | 5.66 (4.07-7.21) | 1719 (1120-2490) | 1.33 (0.85-1.92) | 804 (485-1253) | 0.62 (0.37-0.95) |
| Tonga | Female | 1505 (1410-1591) | 232 (190-274) | 15.47 (12.69-18.10) | 157 (126-191) | 10.48 (8.42-12.55) | 105 (83-128) | 6.97 (5.54-8.47) | 52 (40-67) | 3.43 (2.65-4.44) | 43 (31-56) | 2.85 (2.15-3.69) |
| Tonga | Male | 2615 (2428-2788) | 438 (348-521) | 16.76 (13.51-19.78) | 297 (231-363) | 11.37 (9.03-13.75) | 197 (153-245) | 7.54 (5.97-9.32) | 58 (42-76) | 2.22 (1.66-2.91) | 46 (34-62) | 1.76 (1.29-2.34) |
| Trinidad and Tobago | Female | 33112 (31827-34465) | 3113 (2289-3933) | 9.39 (6.89-11.92) | 1972 (1476-2512) | 5.94 (4.49-7.52) | 1792 (1310-2270) | 5.41 (3.98-6.82) | 844 (598-1123) | 2.54 (1.82-3.38) | 687 (482-948) | 2.07 (1.46-2.84) |
| Trinidad and Tobago | Male | 48378 (45691-50816) | 7469 (6023-8859) | 15.53 (12.65-18.08) | 4488 (3530-5518) | 9.31 (7.42-11.40) | 4046 (3186-4967) | 8.35 (6.60-10.20) | 1493 (1148-1931) | 3.08 (2.38-3.98) | 1239 (897-1618) | 2.56 (1.88-3.32) |
| Tunisia | Female | 274226 (250127-297984) | 1434 (298-4462) | 0.52 (0.11-1.62) | 1296 (234-4169) | 0.47 (0.09-1.53) | 1087 (183-3278) | 0.40 (0.07-1.21) | 243 (3-1470) | 0.09 (0.00-0.53) | 195 (2-1237) | 0.07 (0.00-0.44) |
| Tunisia | Male | 391140 (361165-419687) | 25393 (15726-36444) | 6.51 (3.97-9.20) | 19538 (12266-27477) | 5.04 (3.15-6.93) | 13485 (8701-18546) | 3.45 (2.24-4.76) | 4213 (2031-6407) | 1.07 (0.53-1.65) | 3598 (1795-5654) | 0.92 (0.46-1.43) |
| Turkey | Female | 1640901 (1552451-1721965) | 25 (0-2819) | 0.00 (0.00-0.17) | 25 (0-2702) | 0.00 (0.00-0.17) | 25 (0-2623) | 0.00 (0.00-0.16) | 1 (0-901) | 0.00 (0.00-0.05) | 1 (0-792) | 0.00 (0.00-0.05) |
| Turkey | Male | 2161894 (2046370-2282865) | 76668 (36498-130696) | 3.54 (1.70-5.99) | 63109 (31219-103094) | 2.92 (1.43-4.72) | 52063 (25844-85079) | 2.42 (1.21-3.93) | 14958 (5385-27322) | 0.69 (0.25-1.26) | 14423 (5266-26613) | 0.67 (0.24-1.23) |
| Turkmenistan | Female | 180344 (171522-188936) | 20525 (15789-25763) | 11.36 (8.74-14.20) | 13964 (10596-17452) | 7.76 (5.88-9.67) | 10285 (7729-12822) | 5.70 (4.37-7.09) | 5899 (4426-7697) | 3.28 (2.45-4.24) | 5123 (3835-6727) | 2.84 (2.12-3.72) |
| Turkmenistan | Male | 238473 (225702-251628) | 45020 (36233-52837) | 18.87 (15.41-21.89) | 29332 (23059-35279) | 12.26 (9.80-14.67) | 21357 (16587-26135) | 8.94 (6.93-10.95) | 8207 (6257-10132) | 3.43 (2.65-4.25) | 6010 (4552-7564) | 2.52 (1.92-3.17) |
| Tuvalu | Female | 465 (438-494) | 74 (59-89) | 15.89 (12.72-18.84) | 50 (39-61) | 10.73 (8.40-12.90) | 34 (27-43) | 7.35 (5.71-9.01) | 13 (9-17) | 2.71 (1.93-3.65) | 8 (5-11) | 1.62 (1.10-2.28) |
| Tuvalu | Male | 675 (629-720) | 111 (83-131) | 16.47 (12.58-19.23) | 75 (55-91) | 11.15 (8.33-13.57) | 52 (39-64) | 7.67 (5.87-9.52) | 14 (10-19) | 2.15 (1.55-2.88) | 10 (7-14) | 1.47 (0.99-2.11) |
| Uganda | Female | 269346 (247176-289815) | 49340 (38874-60007) | 18.31 (14.68-21.88) | 34860 (27107-43388) | 12.93 (10.21-15.75) | 21091 (16113-26352) | 7.83 (6.13-9.57) | 6589 (4718-8845) | 2.46 (1.78-3.23) | 3980 (2698-5616) | 1.47 (1.00-2.07) |
| Uganda | Male | 357862 (328937-386446) | 44767 (33060-56695) | 12.52 (9.24-15.58) | 32492 (23390-41364) | 9.07 (6.67-11.44) | 19774 (14201-25961) | 5.53 (4.01-7.20) | 3857 (2602-5338) | 1.07 (0.73-1.46) | 2177 (1379-3128) | 0.61 (0.39-0.87) |
| Ukraine | Female | 3539519 (3245186-3836863) | 133798 (66139-225923) | 3.83 (1.91-6.38) | 119614 (60745-188383) | 3.36 (1.69-5.26) | 90453 (48124-135528) | 2.56 (1.36-3.75) | 60342 (28069-100428) | 1.72 (0.80-2.83) | 51285 (24202-87640) | 1.46 (0.68-2.42) |
| Ukraine | Male | 3465826 (3156380-3762938) | 436131 (333649-528129) | 12.63 (9.86-15.02) | 296432 (227789-371808) | 8.58 (6.60-10.48) | 198841 (153907-251465) | 5.75 (4.47-7.13) | 97733 (71527-129137) | 2.82 (2.05-3.66) | 68374 (48565-91512) | 1.98 (1.41-2.64) |
| United Arab Emirates | Female | 42246 (39437-44872) | 360 (60-1074) | 0.86 (0.14-2.49) | 239 (33-699) | 0.57 (0.08-1.68) | 331 (52-1001) | 0.79 (0.13-2.34) | 37 (1-220) | 0.09 (0.00-0.52) | 32 (1-199) | 0.08 (0.00-0.47) |
| United Arab Emirates | Male | 120162 (111518-128774) | 10997 (6495-15409) | 9.17 (5.37-12.74) | 4792 (2912-6855) | 3.98 (2.40-5.79) | 8776 (5294-12399) | 7.33 (4.39-10.21) | 819 (449-1208) | 0.68 (0.38-0.99) | 710 (394-1074) | 0.60 (0.32-0.88) |
| United Kingdom | Female | 1031248 (1008047-1055796) | 64933 (42053-88850) | 6.30 (4.09-8.58) | 26510 (17626-36123) | 2.57 (1.71-3.51) | 52729 (35461-71109) | 5.11 (3.44-6.91) | 9782 (6286-13724) | 0.95 (0.61-1.32) | 8926 (5607-12487) | 0.87 (0.55-1.20) |
| United Kingdom | Male | 1410418 (1391706-1427745) | 150059 (112814-182494) | 10.65 (8.03-12.89) | 54421 (39054-70537) | 3.86 (2.78-5.00) | 112902 (85396-139958) | 8.00 (6.07-9.90) | 19270 (13319-25859) | 1.37 (0.94-1.84) | 17255 (11830-23309) | 1.22 (0.84-1.65) |
| United States | Female | 6691507 (6537326-6847676) | 649345 (489497-804894) | 9.70 (7.34-12.08) | 125902 (77615-190753) | 1.88 (1.16-2.84) | 577397 (434086-720774) | 8.62 (6.50-10.85) | 54986 (32380-86085) | 0.82 (0.48-1.29) | 50844 (29931-80109) | 0.76 (0.45-1.18) |
| United States | Male | 8991320 (8875684-9103683) | 1334404 (1022061-1583741) | 14.84 (11.32-17.64) | 252967 (154090-375684) | 2.82 (1.72-4.17) | 1176474 (890064-1407859) | 13.09 (9.89-15.67) | 98906 (58995-152821) | 1.10 (0.65-1.70) | 90130 (53526-141424) | 1.00 (0.60-1.58) |
| Uruguay | Female | 73051 (71104-74741) | 10384 (8672-12036) | 14.24 (11.81-16.42) | 6019 (4898-7261) | 8.28 (6.70-9.92) | 5717 (4584-6914) | 7.84 (6.28-9.43) | 3209 (2501-3939) | 4.41 (3.43-5.39) | 3044 (2384-3826) | 4.18 (3.29-5.22) |
| Uruguay | Male | 82549 (80777-84404) | 14283 (11645-16702) | 17.28 (14.14-20.19) | 8226 (6671-9912) | 9.94 (8.10-12.02) | 7855 (6211-9493) | 9.51 (7.49-11.46) | 3604 (2788-4496) | 4.37 (3.38-5.46) | 3112 (2410-3919) | 3.77 (2.92-4.74) |
| Uzbekistan | Female | 931090 (900924-961706) | 105971 (80807-131945) | 11.39 (8.73-14.12) | 76519 (58443-95659) | 8.21 (6.29-10.17) | 48043 (36870-59789) | 5.15 (3.97-6.44) | 29318 (21502-38192) | 3.14 (2.30-4.07) | 25176 (18206-32955) | 2.70 (1.94-3.51) |
| Uzbekistan | Male | 1223516 (1173238-1268613) | 224938 (180618-265479) | 18.39 (14.89-21.46) | 155604 (122902-187633) | 12.69 (10.11-15.20) | 96519 (74329-120291) | 7.88 (6.15-9.73) | 41723 (31284-52835) | 3.41 (2.56-4.31) | 32563 (24049-41801) | 2.66 (2.00-3.41) |
| Vanuatu | Female | 7601 (7120-8028) | 1160 (931-1379) | 15.29 (12.30-18.07) | 806 (640-984) | 10.61 (8.43-12.75) | 523 (402-652) | 6.91 (5.32-8.61) | 184 (130-249) | 2.41 (1.73-3.29) | 87 (55-131) | 1.15 (0.71-1.73) |
| Vanuatu | Male | 14968 (14054-15795) | 2318 (1827-2835) | 15.48 (12.37-18.85) | 1611 (1257-2026) | 10.80 (8.41-13.55) | 1044 (813-1333) | 7.02 (5.45-9.06) | 208 (143-298) | 1.39 (0.96-1.97) | 104 (66-159) | 0.70 (0.44-1.06) |
| Venezuela | Female | 557654 (530561-584085) | 81851 (66339-98313) | 14.68 (11.83-17.40) | 57041 (45027-70326) | 10.24 (8.07-12.54) | 35584 (27844-44099) | 6.38 (5.01-7.92) | 27827 (21697-34973) | 4.99 (3.89-6.21) | 26081 (20269-33324) | 4.68 (3.63-5.96) |
| Venezuela | Male | 823396 (773664-877988) | 167961 (132647-199397) | 20.48 (16.25-24.17) | 114629 (88804-139345) | 13.92 (10.89-16.76) | 71613 (54615-89159) | 8.71 (6.63-10.74) | 42581 (32558-54135) | 5.17 (3.90-6.48) | 38413 (29197-48837) | 4.67 (3.58-5.84) |
| Vietnam | Female | 2111958 (1991516-2227446) | 404301 (324141-479657) | 19.14 (15.47-22.45) | 272422 (214961-327863) | 12.86 (10.25-15.35) | 178142 (139748-215995) | 8.44 (6.70-10.21) | 75491 (58148-95291) | 3.57 (2.74-4.49) | 51661 (38429-67683) | 2.44 (1.82-3.16) |
| Vietnam | Male | 3504297 (3288049-3721327) | 831137 (684317-973281) | 23.70 (19.59-27.45) | 563233 (451707-678476) | 16.09 (12.93-19.27) | 369761 (292232-456645) | 10.55 (8.27-12.95) | 130026 (98911-166154) | 3.71 (2.83-4.72) | 89470 (67168-117897) | 2.56 (1.91-3.35) |
| Yemen | Female | 634126 (582373-682462) | 5020 (788-12644) | 0.79 (0.13-1.99) | 4473 (722-11446) | 0.70 (0.12-1.82) | 3456 (612-8708) | 0.55 (0.10-1.35) | 469 (33-2183) | 0.07 (0.01-0.34) | 362 (22-1860) | 0.06 (0.00-0.30) |
| Yemen | Male | 757835 (703702-814616) | 56409 (35029-77893) | 7.41 (4.66-10.29) | 43979 (28723-60066) | 5.84 (3.86-7.93) | 27303 (17736-37526) | 3.62 (2.37-4.94) | 6718 (3868-10550) | 0.89 (0.51-1.38) | 5689 (3157-9123) | 0.75 (0.42-1.20) |
| Zambia | Female | 200126 (185719-215241) | 37406 (28703-45449) | 18.64 (14.48-22.22) | 26401 (19929-32640) | 13.11 (10.04-15.97) | 16121 (11991-20513) | 8.05 (6.11-10.01) | 7324 (5284-9594) | 3.67 (2.67-4.75) | 4420 (3082-6185) | 2.20 (1.54-3.05) |
| Zambia | Male | 255496 (236263-274491) | 32567 (23606-41507) | 12.81 (9.33-16.12) | 23692 (16684-30158) | 9.25 (6.59-11.74) | 14655 (10463-18889) | 5.74 (4.10-7.40) | 3916 (2658-5268) | 1.54 (1.05-2.05) | 2282 (1486-3232) | 0.89 (0.58-1.28) |
| Zimbabwe | Female | 253095 (235619-270623) | 34262 (25844-42892) | 13.50 (10.28-16.69) | 24378 (18221-30901) | 9.65 (7.25-12.10) | 15260 (11118-19551) | 6.04 (4.44-7.73) | 7635 (5204-10552) | 3.01 (2.10-4.09) | 6093 (3850-8756) | 2.41 (1.55-3.47) |
| Zimbabwe | Male | 208998 (195584-222214) | 31217 (23598-38645) | 14.97 (11.33-18.33) | 22222 (16040-28023) | 10.56 (7.81-13.26) | 13523 (10003-17690) | 6.49 (4.77-8.50) | 4019 (2669-5768) | 1.92 (1.28-2.74) | 3020 (1906-4580) | 1.44 (0.92-2.18) |

#### e. **Prevented CKD deaths**

| **Country** | **Sex** | **Baseline disease burden Median (95% UI)** | **Replacement of discretionary salt and non-discretionary salt** | | **Replacement of discretionary salt** | | **Replacement of non-discretionary salt** | | **Replacement of discretionary salt among those with diagnosed hypertension** | | **Replacement of discretionary salt among those with treated hypertension** | |
| --- | --- | --- | --- | --- | --- | --- | --- | --- | --- | --- | --- | --- |
|  |  |  | **No. of events averted Median (95% UI)** | **Percentage of national disease burden averted % (95% UI)** | **No. of events averted Median (95% UI)** | **Percentage of national disease burden averted % (95% UI)** | **No. of events averted Median (95% UI)** | **Percentage of national disease burden averted % (95% UI)** | **No. of events averted Median (95% UI)** | **Percentage of national disease burden averted % (95% UI)** | **No. of events averted Median (95% UI)** | **Percentage of national disease burden averted % (95% UI)** |
| **Global, millions** | **Female** | 0.71 (0.70-0.73) | 0.07 (0.07-0.08) | 10.43 (9.75-11.04) | 0.04 (0.04-0.05) | 6.01 (5.60-6.40) | 0.04 (0.04-0.04) | 5.68 (5.25-6.10) | 0.02 (0.01-0.02) | 2.14 (1.98-2.30) | 0.01 (0.01-0.01) | 1.82 (1.68-1.96) |
| **Global, millions** | **Male** | 0.77 (0.75-0.78) | 0.10 (0.09-0.10) | 12.89 (12.20-13.57) | 0.06 (0.05-0.06) | 7.54 (7.06-8.02) | 0.05 (0.05-0.06) | 6.96 (6.48-7.46) | 0.02 (0.02-0.02) | 2.17 (2.02-2.34) | 0.01 (0.01-0.02) | 1.82 (1.69-1.96) |
| Afghanistan | Female | 3096 (2639-3519) | 12 (2-36) | 0.40 (0.06-1.16) | 11 (2-36) | 0.36 (0.06-1.12) | 9 (1-27) | 0.28 (0.04-0.87) | 3 (0-12) | 0.09 (0.01-0.37) | 2 (0-10) | 0.07 (0.01-0.31) |
| Afghanistan | Male | 1389 (1108-1636) | 59 (33-93) | 4.30 (2.49-6.60) | 47 (27-72) | 3.40 (2.01-5.12) | 30 (17-45) | 2.15 (1.30-3.14) | 9 (4-16) | 0.67 (0.34-1.12) | 8 (4-14) | 0.55 (0.29-0.97) |
| Albania | Female | 202 (181-223) | 38 (31-46) | 19.04 (15.75-22.38) | 25 (19-31) | 12.20 (9.93-14.99) | 17 (13-22) | 8.49 (6.71-10.56) | 12 (9-16) | 5.90 (4.60-7.65) | 11 (8-14) | 5.23 (3.98-6.98) |
| Albania | Male | 215 (189-241) | 49 (39-59) | 22.84 (19.31-26.24) | 33 (26-42) | 15.42 (12.49-18.68) | 23 (18-30) | 10.81 (8.65-13.52) | 8 (6-12) | 3.92 (2.85-5.20) | 6 (4-9) | 2.90 (2.02-4.15) |
| Algeria | Female | 5240 (4676-5785) | 13 (1-46) | 0.25 (0.02-0.86) | 12 (1-43) | 0.23 (0.02-0.80) | 11 (1-34) | 0.21 (0.02-0.65) | 3 (0-18) | 0.06 (0.00-0.35) | 3 (0-16) | 0.06 (0.00-0.32) |
| Algeria | Male | 4370 (3901-4819) | 164 (87-253) | 3.77 (2.05-5.72) | 130 (72-204) | 2.97 (1.67-4.56) | 91 (51-142) | 2.07 (1.17-3.20) | 36 (17-64) | 0.83 (0.38-1.43) | 30 (13-54) | 0.70 (0.30-1.19) |
| American Samoa | Female | 15 (14-17) | 1 (1-2) | 7.81 (5.60-9.75) | 1 (1-1) | 4.85 (3.49-6.17) | 1 (0-1) | 4.47 (3.18-5.69) | 0 (0-0) | 1.72 (1.16-2.33) | 0 (0-0) | 1.32 (0.83-1.89) |
| American Samoa | Male | 15 (14-17) | 1 (1-2) | 9.14 (6.82-11.32) | 1 (1-1) | 5.53 (4.13-6.95) | 1 (1-1) | 5.07 (3.85-6.41) | 0 (0-0) | 1.82 (1.25-2.46) | 0 (0-0) | 1.39 (0.92-2.02) |
| Andorra | Female | 9 (8-11) | 0 (0-1) | 3.82 (1.62-6.65) | 0 (0-0) | 2.28 (1.03-3.74) | 0 (0-1) | 3.44 (1.49-5.81) | 0 (0-0) | 1.11 (0.46-1.96) | 0 (0-0) | 1.01 (0.41-1.73) |
| Andorra | Male | 7 (7-8) | 1 (1-1) | 10.33 (7.42-13.11) | 0 (0-0) | 4.08 (2.84-5.46) | 1 (0-1) | 7.46 (5.48-9.59) | 0 (0-0) | 1.98 (1.30-2.81) | 0 (0-0) | 1.88 (1.22-2.61) |
| Angola | Female | 1318 (1192-1452) | 84 (61-111) | 6.43 (4.66-8.30) | 62 (46-84) | 4.73 (3.50-6.12) | 39 (29-52) | 3.00 (2.20-3.85) | 18 (12-25) | 1.39 (0.92-1.90) | 8 (5-13) | 0.64 (0.38-0.98) |
| Angola | Male | 2291 (2070-2514) | 158 (112-205) | 6.94 (4.95-8.75) | 115 (84-152) | 5.03 (3.67-6.45) | 72 (52-95) | 3.17 (2.33-4.07) | 24 (15-35) | 1.07 (0.68-1.51) | 15 (9-23) | 0.66 (0.40-1.00) |
| Antigua and Barbuda | Female | 21 (20-23) | 2 (1-2) | 7.97 (5.97-9.93) | 1 (1-1) | 4.83 (3.66-6.05) | 1 (1-1) | 4.48 (3.45-5.70) | 0 (0-1) | 2.35 (1.69-3.05) | 0 (0-1) | 1.99 (1.41-2.69) |
| Antigua and Barbuda | Male | 24 (22-26) | 3 (2-3) | 11.33 (8.87-13.43) | 2 (1-2) | 6.57 (5.02-8.12) | 1 (1-2) | 6.11 (4.61-7.60) | 1 (0-1) | 2.52 (1.83-3.42) | 1 (0-1) | 2.14 (1.50-2.93) |
| Argentina | Female | 7665 (7049-8231) | 960 (791-1135) | 12.50 (10.45-14.51) | 598 (481-726) | 7.79 (6.28-9.38) | 465 (366-577) | 6.10 (4.83-7.52) | 300 (237-376) | 3.91 (3.11-4.82) | 267 (208-340) | 3.47 (2.75-4.38) |
| Argentina | Male | 7479 (6859-8054) | 1070 (869-1280) | 14.33 (11.88-16.85) | 663 (528-813) | 8.94 (7.18-10.72) | 520 (412-654) | 7.00 (5.55-8.57) | 323 (252-401) | 4.32 (3.41-5.30) | 287 (224-363) | 3.86 (3.02-4.76) |
| Armenia | Female | 228 (211-246) | 25 (19-32) | 11.01 (8.58-13.68) | 17 (13-22) | 7.46 (5.68-9.47) | 11 (9-15) | 5.01 (3.76-6.46) | 7 (5-10) | 3.03 (2.12-4.14) | 6 (4-8) | 2.67 (1.86-3.65) |
| Armenia | Male | 199 (182-215) | 28 (23-34) | 14.19 (11.62-16.79) | 19 (15-23) | 9.31 (7.49-11.27) | 12 (10-16) | 6.19 (4.92-7.81) | 6 (4-7) | 2.80 (2.11-3.71) | 4 (3-5) | 1.87 (1.40-2.56) |
| Australia | Female | 2559 (2330-2812) | 73 (25-138) | 2.85 (0.98-5.51) | 31 (11-59) | 1.21 (0.44-2.27) | 71 (24-132) | 2.76 (0.96-5.13) | 13 (4-26) | 0.52 (0.17-1.02) | 12 (4-23) | 0.46 (0.15-0.92) |
| Australia | Male | 2431 (2218-2648) | 133 (76-208) | 5.52 (3.13-8.36) | 42 (22-69) | 1.73 (0.89-2.76) | 122 (69-184) | 5.02 (2.86-7.44) | 17 (9-29) | 0.71 (0.36-1.18) | 16 (8-28) | 0.67 (0.32-1.12) |
| Austria | Female | 2090 (1904-2257) | 278 (218-339) | 13.40 (10.44-15.95) | 83 (51-122) | 4.00 (2.45-5.87) | 217 (168-277) | 10.41 (7.94-13.09) | 41 (23-63) | 1.94 (1.14-3.06) | 36 (20-58) | 1.74 (1.01-2.75) |
| Austria | Male | 1321 (1239-1400) | 221 (173-269) | 16.69 (13.43-19.97) | 64 (44-92) | 4.89 (3.33-6.88) | 170 (132-212) | 12.95 (10.06-16.13) | 32 (21-48) | 2.41 (1.58-3.63) | 31 (20-47) | 2.39 (1.52-3.51) |
| Azerbaijan | Female | 446 (399-491) | 39 (30-50) | 8.82 (6.81-10.96) | 27 (21-35) | 6.09 (4.71-7.59) | 18 (14-24) | 4.14 (3.17-5.26) | 11 (8-15) | 2.55 (1.87-3.30) | 9 (7-13) | 2.13 (1.54-2.82) |
| Azerbaijan | Male | 499 (443-558) | 64 (51-78) | 12.79 (10.50-15.09) | 42 (32-52) | 8.40 (6.73-10.22) | 28 (22-37) | 5.68 (4.51-7.06) | 12 (9-16) | 2.38 (1.80-3.03) | 9 (6-11) | 1.71 (1.27-2.28) |
| Bahrain | Female | 117 (106-128) | 0 (0-2) | 0.32 (0.03-1.34) | 0 (0-1) | 0.29 (0.03-1.16) | 0 (0-1) | 0.30 (0.03-1.22) | 0 (0-1) | 0.09 (0.00-0.48) | 0 (0-1) | 0.07 (0.00-0.42) |
| Bahrain | Male | 137 (122-151) | 6 (4-10) | 4.77 (2.81-7.01) | 4 (2-6) | 2.87 (1.75-4.08) | 5 (3-7) | 3.35 (2.06-4.85) | 1 (1-2) | 0.82 (0.43-1.30) | 1 (0-2) | 0.71 (0.35-1.14) |
| Bangladesh | Female | 6450 (5870-7128) | 753 (617-902) | 11.73 (9.75-13.46) | 517 (416-639) | 8.05 (6.56-9.65) | 321 (251-411) | 5.00 (3.97-6.11) | 129 (99-168) | 2.01 (1.57-2.52) | 103 (79-135) | 1.60 (1.26-2.02) |
| Bangladesh | Male | 7593 (6721-8444) | 856 (674-1042) | 11.30 (9.05-13.24) | 589 (454-733) | 7.82 (6.18-9.37) | 366 (282-465) | 4.82 (3.73-6.00) | 96 (69-125) | 1.27 (0.94-1.61) | 75 (55-99) | 0.99 (0.74-1.28) |
| Barbados | Female | 76 (69-82) | 4 (2-6) | 5.02 (2.96-7.34) | 3 (2-4) | 3.66 (2.31-5.00) | 3 (2-4) | 3.42 (2.21-4.81) | 2 (1-2) | 2.02 (1.24-2.88) | 1 (1-2) | 1.86 (1.12-2.65) |
| Barbados | Male | 72 (65-78) | 7 (5-8) | 9.28 (6.86-11.30) | 4 (3-5) | 5.42 (4.03-6.90) | 4 (3-5) | 5.06 (3.76-6.37) | 2 (1-2) | 2.48 (1.79-3.25) | 2 (1-2) | 2.28 (1.61-3.02) |
| Belarus | Female | 182 (168-197) | 6 (3-10) | 3.37 (1.85-5.21) | 5 (3-8) | 2.88 (1.62-4.33) | 4 (2-6) | 2.30 (1.32-3.38) | 3 (1-4) | 1.59 (0.83-2.48) | 2 (1-4) | 1.28 (0.66-2.03) |
| Belarus | Male | 174 (161-188) | 17 (13-21) | 9.87 (7.66-11.86) | 11 (9-14) | 6.50 (5.04-7.97) | 8 (6-10) | 4.62 (3.62-5.75) | 5 (3-6) | 2.73 (2.00-3.48) | 3 (2-4) | 1.85 (1.34-2.44) |
| Belgium | Female | 1611 (1427-1788) | 206 (154-254) | 12.86 (9.80-15.36) | 64 (41-92) | 3.98 (2.57-5.69) | 160 (115-204) | 9.94 (7.38-12.43) | 33 (21-49) | 2.03 (1.31-2.94) | 31 (20-47) | 1.95 (1.23-2.85) |
| Belgium | Male | 1202 (1088-1313) | 174 (142-215) | 14.51 (11.94-17.44) | 54 (37-76) | 4.52 (3.05-6.18) | 135 (106-169) | 11.26 (8.90-13.83) | 26 (17-37) | 2.19 (1.44-3.07) | 26 (18-37) | 2.19 (1.45-3.06) |
| Belize | Female | 64 (60-69) | 4 (3-5) | 6.85 (5.00-8.63) | 3 (2-4) | 4.73 (3.51-5.97) | 2 (2-3) | 3.28 (2.46-4.27) | 1 (1-2) | 1.93 (1.36-2.50) | 1 (1-1) | 1.57 (1.08-2.11) |
| Belize | Male | 66 (61-70) | 7 (5-8) | 10.51 (8.39-12.43) | 5 (4-6) | 6.98 (5.51-8.44) | 3 (2-4) | 4.81 (3.71-5.94) | 1 (1-2) | 2.13 (1.61-2.82) | 1 (1-2) | 1.74 (1.26-2.38) |
| Benin | Female | 696 (635-762) | 77 (61-91) | 11.01 (9.07-12.62) | 53 (41-64) | 7.63 (6.10-8.98) | 32 (24-39) | 4.59 (3.61-5.56) | 12 (9-16) | 1.79 (1.33-2.29) | 9 (6-12) | 1.28 (0.91-1.72) |
| Benin | Male | 988 (901-1074) | 84 (62-104) | 8.49 (6.41-10.38) | 59 (44-75) | 5.99 (4.57-7.40) | 36 (27-46) | 3.63 (2.73-4.57) | 7 (5-10) | 0.75 (0.52-1.04) | 5 (4-8) | 0.55 (0.38-0.78) |
| Bermuda | Female | 10 (9-11) | 1 (1-1) | 9.39 (7.02-11.53) | 0 (0-0) | 0.64 (0.45-1.19) | 1 (1-1) | 9.03 (6.79-11.19) | 0 (0-0) | 0.32 (0.22-0.63) | 0 (0-0) | 0.27 (0.18-0.55) |
| Bermuda | Male | 12 (11-13) | 1 (1-2) | 12.09 (9.74-14.55) | 0 (0-0) | 0.80 (0.60-1.44) | 1 (1-2) | 11.56 (9.28-13.90) | 0 (0-0) | 0.32 (0.22-0.64) | 0 (0-0) | 0.28 (0.18-0.54) |
| Bhutan | Female | 54 (48-60) | 6 (5-7) | 11.12 (8.93-12.98) | 4 (3-5) | 7.60 (5.90-9.17) | 3 (2-3) | 4.86 (3.81-5.93) | 1 (1-2) | 2.28 (1.71-2.89) | 1 (1-1) | 1.77 (1.32-2.32) |
| Bhutan | Male | 63 (56-69) | 8 (6-9) | 11.99 (9.79-14.02) | 5 (4-6) | 8.14 (6.54-9.79) | 3 (3-4) | 5.27 (4.05-6.45) | 1 (1-2) | 1.93 (1.45-2.45) | 1 (1-1) | 1.34 (1.00-1.77) |
| Bolivia | Female | 2616 (2363-2876) | 268 (200-332) | 10.30 (7.74-12.22) | 183 (133-230) | 6.98 (5.11-8.59) | 117 (84-151) | 4.47 (3.29-5.70) | 70 (48-95) | 2.70 (1.86-3.60) | 63 (42-89) | 2.41 (1.65-3.32) |
| Bolivia | Male | 2016 (1786-2224) | 253 (188-316) | 12.62 (9.46-15.36) | 171 (126-217) | 8.54 (6.30-10.51) | 110 (78-144) | 5.47 (3.97-6.91) | 53 (36-74) | 2.62 (1.76-3.64) | 44 (29-63) | 2.20 (1.47-3.12) |
| Bosnia and Herzegovina | Female | 350 (317-382) | 67 (53-81) | 19.29 (15.85-22.56) | 43 (33-54) | 12.22 (9.68-15.00) | 31 (23-39) | 8.75 (6.87-11.01) | 25 (19-32) | 7.09 (5.50-8.89) | 23 (17-30) | 6.52 (5.02-8.40) |
| Bosnia and Herzegovina | Male | 332 (304-362) | 75 (63-87) | 22.69 (19.44-25.50) | 50 (41-60) | 15.07 (12.32-17.87) | 36 (29-44) | 10.90 (8.87-13.12) | 22 (17-28) | 6.75 (5.20-8.34) | 21 (16-27) | 6.21 (4.75-7.87) |
| Botswana | Female | 152 (139-167) | 9 (6-12) | 5.87 (3.90-7.89) | 7 (5-9) | 4.35 (3.07-5.74) | 5 (3-6) | 3.20 (2.27-4.20) | 2 (2-4) | 1.63 (1.05-2.32) | 2 (1-3) | 1.43 (0.93-2.03) |
| Botswana | Male | 185 (166-205) | 10 (7-14) | 5.52 (3.68-7.39) | 7 (5-10) | 4.04 (2.71-5.36) | 5 (4-8) | 2.99 (2.04-4.06) | 2 (1-3) | 1.02 (0.59-1.51) | 1 (1-2) | 0.74 (0.41-1.14) |
| Brazil | Female | 21811 (20354-23267) | 2505 (2029-3034) | 11.50 (9.40-13.55) | 1624 (1277-2022) | 7.45 (5.95-9.11) | 1177 (913-1460) | 5.40 (4.25-6.63) | 902 (698-1146) | 4.14 (3.25-5.14) | 878 (683-1117) | 4.03 (3.18-4.99) |
| Brazil | Male | 23316 (21836-24749) | 3243 (2710-3877) | 13.99 (11.73-16.25) | 2089 (1721-2509) | 8.98 (7.36-10.67) | 1500 (1221-1857) | 6.46 (5.29-7.84) | 975 (788-1206) | 4.20 (3.39-5.06) | 922 (747-1145) | 3.99 (3.22-4.86) |
| Brunei | Female | 42 (39-46) | 6 (5-8) | 14.97 (12.11-17.77) | 3 (2-4) | 6.93 (5.30-8.59) | 4 (3-5) | 9.38 (7.37-11.54) | 1 (1-2) | 3.28 (2.45-4.30) | 1 (1-2) | 3.12 (2.30-4.06) |
| Brunei | Male | 43 (39-46) | 7 (6-9) | 16.95 (13.88-19.69) | 3 (3-4) | 7.77 (6.12-9.58) | 5 (4-6) | 10.66 (8.55-12.82) | 1 (1-2) | 3.27 (2.44-4.19) | 1 (1-2) | 3.06 (2.31-3.91) |
| Bulgaria | Female | 1134 (1036-1226) | 209 (172-250) | 18.42 (15.45-21.47) | 125 (101-156) | 11.11 (9.06-13.57) | 102 (82-128) | 9.08 (7.37-11.02) | 69 (54-90) | 6.08 (4.85-7.80) | 61 (47-82) | 5.44 (4.19-7.08) |
| Bulgaria | Male | 1287 (1178-1391) | 285 (238-331) | 22.25 (18.72-25.36) | 180 (140-220) | 13.98 (11.19-16.78) | 146 (117-180) | 11.39 (9.16-13.94) | 81 (60-106) | 6.34 (4.72-8.25) | 71 (51-94) | 5.49 (3.98-7.30) |
| Burkina Faso | Female | 1581 (1435-1724) | 124 (91-160) | 7.84 (5.86-9.86) | 88 (66-114) | 5.60 (4.20-7.02) | 53 (39-70) | 3.37 (2.52-4.34) | 19 (13-27) | 1.22 (0.83-1.66) | 14 (9-20) | 0.86 (0.56-1.26) |
| Burkina Faso | Male | 1688 (1535-1841) | 141 (103-180) | 8.36 (6.25-10.51) | 100 (73-130) | 5.91 (4.43-7.53) | 59 (43-78) | 3.52 (2.59-4.59) | 15 (10-23) | 0.91 (0.62-1.32) | 8 (5-13) | 0.50 (0.32-0.79) |
| Burundi | Female | 509 (461-558) | 66 (54-79) | 12.91 (10.90-15.12) | 46 (37-55) | 8.97 (7.37-10.72) | 27 (21-33) | 5.23 (4.26-6.40) | 12 (9-17) | 2.44 (1.77-3.25) | 8 (6-12) | 1.68 (1.13-2.37) |
| Burundi | Male | 852 (747-953) | 89 (66-110) | 10.41 (8.03-12.63) | 63 (47-79) | 7.38 (5.63-9.00) | 37 (27-48) | 4.31 (3.19-5.50) | 12 (8-18) | 1.47 (0.97-2.10) | 9 (5-14) | 1.04 (0.62-1.57) |
| Cambodia | Female | 1396 (1247-1540) | 165 (118-218) | 11.82 (8.66-14.95) | 112 (80-150) | 8.04 (5.84-10.28) | 68 (47-94) | 4.91 (3.53-6.45) | 29 (20-41) | 2.10 (1.48-2.84) | 23 (15-34) | 1.65 (1.11-2.33) |
| Cambodia | Male | 1150 (1019-1283) | 156 (111-196) | 13.60 (9.89-16.61) | 107 (76-136) | 9.33 (6.63-11.64) | 66 (46-85) | 5.73 (4.04-7.23) | 20 (13-28) | 1.78 (1.18-2.38) | 15 (9-21) | 1.29 (0.86-1.84) |
| Cameroon | Female | 2208 (1979-2431) | 181 (133-227) | 8.24 (6.19-10.01) | 128 (96-162) | 5.83 (4.50-7.19) | 78 (58-100) | 3.55 (2.70-4.40) | 31 (21-43) | 1.42 (1.00-1.92) | 18 (12-27) | 0.83 (0.56-1.18) |
| Cameroon | Male | 3068 (2754-3350) | 263 (199-330) | 8.58 (6.72-10.56) | 184 (141-235) | 6.03 (4.72-7.52) | 113 (86-145) | 3.68 (2.82-4.62) | 30 (20-43) | 0.98 (0.67-1.37) | 19 (12-29) | 0.62 (0.40-0.93) |
| Canada | Female | 4118 (3746-4478) | 364 (250-476) | 8.86 (6.10-11.44) | 119 (78-169) | 2.90 (1.89-4.04) | 294 (205-382) | 7.17 (5.05-9.23) | 50 (31-73) | 1.21 (0.77-1.72) | 49 (31-71) | 1.18 (0.77-1.70) |
| Canada | Male | 3646 (3347-3932) | 454 (360-558) | 12.50 (9.96-14.84) | 139 (95-197) | 3.85 (2.60-5.27) | 355 (273-438) | 9.81 (7.38-11.86) | 68 (44-97) | 1.86 (1.25-2.63) | 67 (45-97) | 1.85 (1.22-2.61) |
| Cape Verde | Female | 54 (48-60) | 5 (4-6) | 9.25 (6.91-11.68) | 3 (3-5) | 6.38 (4.94-8.16) | 2 (2-3) | 4.08 (3.07-5.40) | 2 (1-2) | 2.84 (2.05-3.87) | 1 (1-2) | 2.51 (1.73-3.47) |
| Cape Verde | Male | 67 (60-74) | 6 (5-8) | 9.47 (7.22-11.33) | 4 (3-5) | 6.48 (5.06-7.99) | 3 (2-4) | 4.15 (3.17-5.22) | 1 (1-2) | 1.89 (1.39-2.52) | 1 (1-1) | 1.50 (1.06-2.05) |
| Central African Republic | Female | 303 (274-335) | 18 (13-24) | 5.97 (4.22-7.72) | 14 (10-18) | 4.50 (3.15-5.79) | 8 (6-11) | 2.75 (1.99-3.51) | 3 (2-5) | 1.13 (0.70-1.53) | 2 (1-3) | 0.59 (0.35-0.89) |
| Central African Republic | Male | 546 (484-608) | 34 (22-45) | 6.23 (4.08-8.17) | 25 (17-34) | 4.66 (3.10-6.05) | 15 (10-21) | 2.83 (1.92-3.70) | 4 (2-6) | 0.75 (0.44-1.08) | 2 (1-4) | 0.43 (0.25-0.65) |
| Chad | Female | 505 (454-554) | 39 (28-49) | 7.67 (5.74-9.60) | 28 (20-36) | 5.49 (4.08-6.88) | 16 (12-21) | 3.27 (2.41-4.21) | 7 (5-9) | 1.34 (0.90-1.82) | 4 (3-6) | 0.87 (0.55-1.25) |
| Chad | Male | 995 (882-1113) | 84 (61-109) | 8.47 (6.23-10.33) | 60 (43-78) | 6.00 (4.38-7.51) | 35 (25-47) | 3.55 (2.59-4.54) | 11 (7-17) | 1.13 (0.74-1.67) | 8 (5-12) | 0.76 (0.48-1.19) |
| Chile | Female | 2818 (2580-3050) | 370 (303-443) | 13.19 (10.84-15.47) | 217 (172-268) | 7.73 (6.16-9.34) | 196 (151-245) | 6.99 (5.37-8.48) | 114 (88-145) | 4.08 (3.17-5.05) | 106 (81-135) | 3.75 (2.90-4.74) |
| Chile | Male | 2369 (2197-2546) | 358 (291-428) | 15.14 (12.54-17.75) | 207 (165-253) | 8.75 (7.12-10.51) | 187 (148-232) | 7.92 (6.39-9.66) | 104 (79-129) | 4.38 (3.42-5.35) | 92 (70-114) | 3.88 (3.01-4.80) |
| China | Female | 92005 (84834-99738) | 16031 (12673-19274) | 17.43 (14.18-20.27) | 9785 (7660-11995) | 10.67 (8.42-12.87) | 8044 (6207-9990) | 8.76 (6.85-10.67) | 3024 (2285-3810) | 3.30 (2.52-4.05) | 2526 (1899-3234) | 2.76 (2.07-3.44) |
| China | Male | 110341 (100263-119294) | 21944 (17585-25644) | 19.93 (16.44-22.75) | 13889 (10882-16677) | 12.57 (10.02-14.90) | 11480 (8838-14158) | 10.43 (8.20-12.66) | 3643 (2786-4624) | 3.32 (2.56-4.13) | 2926 (2206-3745) | 2.67 (2.06-3.42) |
| Colombia | Female | 3832 (3541-4125) | 522 (430-623) | 13.65 (11.48-15.97) | 343 (276-412) | 8.97 (7.29-10.56) | 238 (191-294) | 6.21 (5.07-7.58) | 167 (130-208) | 4.38 (3.51-5.36) | 145 (113-185) | 3.79 (3.03-4.74) |
| Colombia | Male | 4701 (4352-5091) | 963 (803-1100) | 20.52 (17.39-22.98) | 631 (515-750) | 13.42 (11.09-15.66) | 440 (355-533) | 9.38 (7.63-11.18) | 254 (200-318) | 5.40 (4.27-6.66) | 210 (162-266) | 4.46 (3.52-5.60) |
| Comoros | Female | 84 (76-93) | 11 (9-14) | 13.41 (11.32-15.76) | 8 (6-10) | 9.15 (7.48-10.87) | 5 (4-6) | 5.56 (4.54-6.79) | 2 (2-3) | 2.59 (1.92-3.44) | 1 (1-2) | 1.70 (1.19-2.34) |
| Comoros | Male | 88 (80-96) | 10 (8-12) | 11.06 (8.76-13.23) | 7 (5-8) | 7.69 (6.03-9.37) | 4 (3-5) | 4.67 (3.62-5.90) | 1 (1-2) | 1.54 (1.07-2.17) | 1 (1-1) | 1.06 (0.72-1.55) |
| Congo | Female | 532 (486-585) | 34 (25-44) | 6.44 (4.71-8.10) | 25 (18-33) | 4.74 (3.44-5.99) | 16 (12-21) | 3.04 (2.26-3.91) | 6 (4-9) | 1.22 (0.80-1.79) | 4 (2-6) | 0.73 (0.42-1.12) |
| Congo | Male | 640 (579-704) | 45 (33-59) | 7.02 (5.24-9.06) | 33 (24-43) | 5.10 (3.80-6.51) | 21 (15-27) | 3.25 (2.43-4.17) | 7 (4-10) | 1.02 (0.67-1.47) | 5 (3-8) | 0.77 (0.48-1.21) |
| Costa Rica | Female | 693 (642-740) | 85 (70-101) | 12.31 (10.33-14.08) | 52 (42-63) | 7.52 (6.14-9.09) | 43 (34-53) | 6.20 (4.94-7.48) | 29 (23-36) | 4.21 (3.40-5.14) | 28 (23-35) | 4.09 (3.31-5.04) |
| Costa Rica | Male | 898 (833-966) | 143 (119-168) | 15.91 (13.47-18.37) | 85 (70-103) | 9.55 (7.93-11.35) | 70 (57-84) | 7.79 (6.42-9.24) | 40 (32-50) | 4.48 (3.66-5.47) | 39 (31-48) | 4.30 (3.50-5.22) |
| Cote d'Ivoire | Female | 1284 (1168-1408) | 117 (92-145) | 9.18 (7.19-11.01) | 81 (62-102) | 6.32 (4.88-7.72) | 50 (38-65) | 3.94 (3.01-4.93) | 20 (14-27) | 1.58 (1.12-2.13) | 13 (9-20) | 1.05 (0.70-1.51) |
| Cote d'Ivoire | Male | 2321 (2132-2537) | 211 (166-259) | 9.07 (7.12-10.95) | 146 (113-182) | 6.29 (4.98-7.64) | 91 (70-116) | 3.91 (3.05-4.94) | 26 (18-37) | 1.13 (0.76-1.56) | 19 (12-29) | 0.82 (0.52-1.20) |
| Croatia | Female | 799 (729-871) | 163 (132-196) | 20.40 (16.57-23.90) | 91 (70-115) | 11.42 (8.91-14.07) | 87 (66-110) | 10.82 (8.39-13.58) | 59 (44-76) | 7.38 (5.71-9.29) | 52 (39-68) | 6.46 (4.90-8.37) |
| Croatia | Male | 534 (486-580) | 127 (105-148) | 23.85 (20.08-27.11) | 75 (60-93) | 14.10 (11.22-17.10) | 71 (56-90) | 13.39 (10.65-16.35) | 44 (34-56) | 8.30 (6.50-10.36) | 32 (24-42) | 6.01 (4.52-7.76) |
| Cuba | Female | 1604 (1489-1721) | 128 (95-164) | 8.04 (5.98-10.23) | 85 (63-110) | 5.27 (4.00-6.79) | 64 (48-85) | 4.02 (3.03-5.20) | 42 (31-56) | 2.63 (1.92-3.45) | 39 (28-52) | 2.43 (1.78-3.22) |
| Cuba | Male | 1589 (1476-1705) | 179 (142-216) | 11.21 (8.99-13.55) | 114 (89-143) | 7.20 (5.66-8.89) | 86 (67-110) | 5.37 (4.23-6.80) | 46 (34-60) | 2.89 (2.15-3.72) | 42 (31-56) | 2.64 (1.96-3.43) |
| Cyprus | Female | 172 (152-193) | 14 (8-19) | 7.95 (4.83-10.79) | 7 (5-10) | 3.98 (2.65-5.48) | 9 (6-13) | 5.52 (3.65-7.51) | 3 (2-5) | 1.94 (1.23-2.88) | 3 (2-5) | 1.78 (1.06-2.66) |
| Cyprus | Male | 169 (150-189) | 20 (15-26) | 11.93 (8.86-14.57) | 9 (7-13) | 5.42 (3.92-7.30) | 13 (10-17) | 7.72 (5.69-10.17) | 4 (3-6) | 2.64 (1.86-3.82) | 4 (3-6) | 2.48 (1.72-3.56) |
| Czech Republic | Female | 881 (814-961) | 174 (141-206) | 19.87 (16.33-22.79) | 87 (68-109) | 9.86 (7.78-12.08) | 104 (82-129) | 11.80 (9.35-14.32) | 49 (37-63) | 5.50 (4.31-6.99) | 46 (35-59) | 5.19 (4.00-6.58) |
| Czech Republic | Male | 673 (613-733) | 157 (131-185) | 23.45 (19.95-26.71) | 83 (64-103) | 12.34 (9.71-15.21) | 98 (78-122) | 14.59 (11.76-17.59) | 47 (36-61) | 7.03 (5.42-8.94) | 45 (34-58) | 6.77 (5.17-8.62) |
| Democratic Republic of the Congo | Female | 5310 (4822-5811) | 79 (29-158) | 1.49 (0.52-2.89) | 76 (27-143) | 1.43 (0.51-2.64) | 60 (23-105) | 1.12 (0.44-1.96) | 18 (5-39) | 0.34 (0.10-0.74) | 11 (3-25) | 0.20 (0.05-0.47) |
| Democratic Republic of the Congo | Male | 7229 (6515-8004) | 186 (89-326) | 2.60 (1.26-4.36) | 165 (84-283) | 2.31 (1.14-3.74) | 117 (58-190) | 1.61 (0.80-2.51) | 30 (12-59) | 0.42 (0.17-0.78) | 23 (9-46) | 0.32 (0.12-0.62) |
| Denmark | Female | 810 (748-871) | 69 (42-93) | 8.52 (5.38-11.39) | 14 (7-25) | 1.67 (0.88-3.01) | 62 (39-84) | 7.62 (4.85-10.28) | 6 (3-10) | 0.71 (0.35-1.29) | 4 (2-7) | 0.48 (0.24-0.91) |
| Denmark | Male | 889 (834-941) | 110 (85-132) | 12.42 (9.68-14.78) | 20 (11-34) | 2.31 (1.26-3.75) | 96 (72-117) | 10.82 (8.29-13.15) | 10 (5-17) | 1.13 (0.60-1.89) | 7 (4-13) | 0.83 (0.44-1.44) |
| Djibouti | Female | 75 (68-83) | 10 (8-12) | 13.09 (10.59-15.18) | 7 (5-8) | 8.86 (7.02-10.42) | 4 (3-5) | 5.60 (4.38-6.71) | 2 (1-2) | 2.40 (1.73-3.16) | 1 (1-2) | 1.62 (1.07-2.29) |
| Djibouti | Male | 124 (111-137) | 12 (10-16) | 10.08 (7.89-12.32) | 9 (6-11) | 6.97 (5.25-8.67) | 5 (4-7) | 4.45 (3.28-5.65) | 2 (1-2) | 1.38 (0.89-1.97) | 1 (1-2) | 0.96 (0.59-1.45) |
| Dominica | Female | 19 (17-20) | 2 (1-2) | 8.32 (6.19-10.36) | 1 (1-1) | 5.60 (4.22-7.14) | 1 (1-1) | 4.03 (2.99-5.24) | 1 (0-1) | 2.89 (2.07-3.79) | 0 (0-1) | 2.50 (1.75-3.35) |
| Dominica | Male | 20 (18-22) | 2 (2-3) | 11.03 (8.69-13.04) | 1 (1-2) | 7.20 (5.52-8.86) | 1 (1-1) | 5.15 (3.91-6.43) | 1 (0-1) | 2.62 (1.93-3.43) | 0 (0-1) | 2.23 (1.62-3.07) |
| Dominican Republic | Female | 1014 (922-1104) | 78 (57-98) | 7.69 (5.80-9.47) | 52 (39-67) | 5.17 (3.98-6.46) | 39 (28-50) | 3.82 (2.90-4.80) | 29 (20-38) | 2.85 (2.08-3.63) | 25 (17-33) | 2.43 (1.76-3.16) |
| Dominican Republic | Male | 1293 (1158-1435) | 141 (111-171) | 10.94 (8.88-12.85) | 91 (72-113) | 7.04 (5.66-8.59) | 67 (51-85) | 5.21 (4.02-6.38) | 41 (31-53) | 3.19 (2.45-3.95) | 36 (27-47) | 2.78 (2.14-3.54) |
| Ecuador | Female | 3127 (2838-3425) | 334 (258-408) | 10.72 (8.36-12.67) | 222 (167-276) | 7.08 (5.46-8.64) | 152 (114-192) | 4.88 (3.71-6.13) | 84 (61-108) | 2.69 (1.97-3.42) | 77 (56-99) | 2.46 (1.81-3.13) |
| Ecuador | Male | 3287 (2966-3586) | 425 (321-519) | 13.00 (9.97-15.48) | 280 (212-349) | 8.52 (6.43-10.38) | 193 (141-245) | 5.87 (4.31-7.33) | 86 (62-112) | 2.61 (1.91-3.38) | 70 (50-92) | 2.13 (1.54-2.77) |
| Egypt | Female | 15845 (14276-17284) | 61 (9-236) | 0.38 (0.06-1.49) | 57 (7-228) | 0.36 (0.04-1.43) | 51 (5-176) | 0.32 (0.03-1.14) | 19 (0-96) | 0.12 (0.00-0.60) | 16 (0-83) | 0.10 (0.00-0.52) |
| Egypt | Male | 16560 (15007-18135) | 761 (447-1115) | 4.61 (2.67-6.63) | 589 (359-835) | 3.55 (2.17-5.05) | 405 (253-587) | 2.45 (1.55-3.46) | 152 (82-233) | 0.92 (0.51-1.41) | 126 (68-197) | 0.76 (0.42-1.17) |
| El Salvador | Female | 1895 (1751-2049) | 218 (178-261) | 11.58 (9.58-13.33) | 146 (117-179) | 7.74 (6.32-9.25) | 97 (77-122) | 5.12 (4.10-6.26) | 70 (54-89) | 3.69 (2.85-4.57) | 64 (49-81) | 3.37 (2.62-4.14) |
| El Salvador | Male | 2466 (2266-2670) | 371 (305-442) | 15.11 (12.50-17.59) | 244 (195-295) | 9.87 (8.00-11.79) | 161 (129-202) | 6.55 (5.34-8.01) | 77 (59-98) | 3.13 (2.47-3.94) | 69 (53-88) | 2.80 (2.20-3.52) |
| Equatorial Guinea | Female | 103 (93-114) | 7 (5-9) | 7.01 (5.20-8.67) | 5 (4-6) | 4.88 (3.50-6.06) | 4 (3-5) | 3.53 (2.65-4.47) | 1 (1-2) | 1.32 (0.87-1.92) | 1 (0-1) | 0.85 (0.49-1.32) |
| Equatorial Guinea | Male | 125 (112-138) | 9 (7-12) | 7.39 (5.61-9.31) | 6 (5-8) | 5.08 (3.92-6.41) | 5 (3-6) | 3.68 (2.80-4.64) | 1 (1-2) | 1.02 (0.66-1.48) | 1 (1-1) | 0.73 (0.44-1.17) |
| Eritrea | Female | 390 (348-432) | 47 (36-59) | 12.16 (9.48-14.57) | 33 (25-41) | 8.42 (6.51-10.32) | 19 (14-25) | 5.00 (3.78-6.18) | 6 (4-9) | 1.58 (1.09-2.12) | 4 (3-6) | 1.08 (0.74-1.57) |
| Eritrea | Male | 433 (369-490) | 38 (28-48) | 8.68 (6.55-10.68) | 27 (19-35) | 6.19 (4.69-7.77) | 16 (11-21) | 3.70 (2.73-4.72) | 3 (2-5) | 0.78 (0.53-1.10) | 2 (1-4) | 0.56 (0.35-0.84) |
| Estonia | Female | 358 (331-385) | 1 (0-4) | 0.16 (0.00-1.24) | 1 (0-4) | 0.15 (0.00-1.15) | 1 (0-4) | 0.15 (0.00-1.22) | 0 (0-1) | 0.05 (0.00-0.36) | 0 (0-1) | 0.03 (0.00-0.28) |
| Estonia | Male | 166 (156-175) | 12 (8-16) | 7.40 (4.98-9.88) | 7 (5-9) | 4.00 (2.80-5.28) | 8 (5-11) | 4.92 (3.35-6.39) | 2 (1-3) | 1.06 (0.70-1.52) | 1 (1-2) | 0.79 (0.51-1.13) |
| Ethiopia | Female | 6858 (6339-7316) | 849 (671-1029) | 12.41 (9.81-14.82) | 589 (457-719) | 8.59 (6.75-10.33) | 351 (269-442) | 5.17 (3.99-6.37) | 106 (75-143) | 1.54 (1.11-2.06) | 54 (34-78) | 0.78 (0.50-1.14) |
| Ethiopia | Male | 8627 (7966-9319) | 876 (656-1093) | 10.23 (7.63-12.47) | 621 (454-791) | 7.20 (5.31-8.95) | 372 (271-490) | 4.32 (3.17-5.63) | 97 (64-139) | 1.12 (0.74-1.60) | 68 (43-102) | 0.78 (0.51-1.16) |
| Federated States of Micronesia | Female | 21 (18-24) | 2 (2-3) | 10.19 (7.39-12.99) | 1 (1-2) | 6.95 (4.92-8.93) | 1 (1-1) | 4.50 (3.25-5.91) | 0 (0-1) | 2.03 (1.35-2.76) | 0 (0-0) | 1.24 (0.78-1.74) |
| Federated States of Micronesia | Male | 15 (13-18) | 2 (1-2) | 11.61 (8.45-14.73) | 1 (1-2) | 7.87 (5.64-10.16) | 1 (1-1) | 5.09 (3.58-6.65) | 0 (0-0) | 1.80 (1.18-2.44) | 0 (0-0) | 1.06 (0.67-1.51) |
| Fiji | Female | 126 (112-139) | 16 (13-20) | 12.58 (10.19-15.27) | 10 (8-14) | 8.34 (6.70-10.39) | 7 (5-9) | 5.55 (4.37-7.07) | 4 (3-6) | 3.46 (2.62-4.49) | 3 (2-5) | 2.61 (1.88-3.57) |
| Fiji | Male | 178 (157-199) | 25 (19-31) | 14.13 (11.28-16.93) | 17 (13-21) | 9.36 (7.37-11.52) | 11 (8-14) | 6.21 (4.75-7.75) | 5 (3-6) | 2.56 (1.86-3.48) | 3 (2-5) | 1.83 (1.26-2.58) |
| Finland | Female | 495 (448-544) | 32 (19-48) | 6.55 (3.83-9.52) | 12 (7-19) | 2.39 (1.42-3.74) | 28 (17-40) | 5.69 (3.46-8.07) | 6 (3-10) | 1.26 (0.70-1.98) | 5 (3-9) | 1.10 (0.61-1.72) |
| Finland | Male | 321 (301-343) | 42 (32-51) | 12.94 (10.32-15.49) | 12 (8-18) | 3.86 (2.50-5.46) | 33 (25-41) | 10.13 (7.78-12.50) | 6 (4-9) | 1.89 (1.20-2.77) | 6 (4-8) | 1.80 (1.12-2.58) |
| France | Female | 8375 (7656-9080) | 693 (404-970) | 8.31 (4.84-11.42) | 284 (181-416) | 3.38 (2.11-4.98) | 532 (325-732) | 6.34 (3.93-8.67) | 130 (78-197) | 1.55 (0.93-2.35) | 119 (73-182) | 1.43 (0.86-2.18) |
| France | Male | 6740 (6314-7147) | 807 (622-997) | 12.08 (9.22-14.67) | 303 (214-415) | 4.51 (3.19-6.17) | 585 (438-749) | 8.71 (6.46-11.05) | 161 (112-226) | 2.40 (1.67-3.32) | 152 (106-215) | 2.26 (1.58-3.15) |
| Gabon | Female | 266 (238-297) | 20 (14-26) | 7.33 (5.60-9.42) | 13 (10-17) | 5.00 (3.79-6.40) | 10 (7-13) | 3.73 (2.83-4.84) | 4 (3-6) | 1.53 (1.04-2.18) | 3 (2-5) | 1.12 (0.69-1.70) |
| Gabon | Male | 324 (288-361) | 24 (16-30) | 7.30 (5.17-9.32) | 16 (11-21) | 4.98 (3.57-6.35) | 12 (9-16) | 3.72 (2.69-4.76) | 3 (2-5) | 1.07 (0.70-1.53) | 3 (1-4) | 0.80 (0.48-1.23) |
| Georgia | Female | 350 (315-381) | 38 (28-47) | 10.87 (8.30-13.15) | 26 (19-33) | 7.39 (5.48-9.15) | 17 (13-22) | 4.95 (3.73-6.25) | 13 (10-18) | 3.87 (2.82-4.98) | 12 (9-17) | 3.50 (2.54-4.65) |
| Georgia | Male | 353 (325-382) | 51 (41-61) | 14.44 (12.03-16.99) | 33 (27-41) | 9.48 (7.65-11.43) | 22 (18-28) | 6.32 (5.04-7.86) | 14 (11-18) | 3.94 (3.03-5.00) | 11 (8-15) | 3.11 (2.34-4.13) |
| Germany | Female | 23487 (20853-26149) | 1713 (975-2551) | 7.33 (4.26-10.36) | 634 (368-1000) | 2.72 (1.59-4.09) | 1431 (848-2106) | 6.14 (3.66-8.61) | 322 (174-507) | 1.36 (0.76-2.15) | 304 (165-490) | 1.30 (0.74-2.06) |
| Germany | Male | 15950 (14416-17621) | 2033 (1564-2502) | 12.74 (9.83-15.55) | 641 (404-913) | 4.02 (2.55-5.72) | 1579 (1173-2001) | 9.85 (7.43-12.44) | 342 (210-494) | 2.14 (1.31-3.12) | 328 (202-483) | 2.07 (1.27-3.02) |
| Ghana | Female | 2410 (2195-2646) | 263 (214-315) | 10.93 (9.00-12.72) | 181 (142-226) | 7.52 (6.02-9.07) | 112 (88-143) | 4.64 (3.69-5.72) | 62 (46-80) | 2.57 (1.98-3.24) | 50 (37-67) | 2.08 (1.58-2.66) |
| Ghana | Male | 4025 (3669-4392) | 435 (344-522) | 10.81 (8.71-12.65) | 298 (237-372) | 7.46 (5.89-8.94) | 187 (142-232) | 4.65 (3.61-5.67) | 58 (42-76) | 1.44 (1.05-1.86) | 45 (32-62) | 1.11 (0.81-1.51) |
| Greece | Female | 3672 (3400-3941) | 296 (177-407) | 8.06 (4.90-11.03) | 177 (117-244) | 4.82 (3.21-6.63) | 178 (116-248) | 4.82 (3.14-6.70) | 100 (64-143) | 2.73 (1.75-3.83) | 99 (64-140) | 2.69 (1.74-3.77) |
| Greece | Male | 3377 (3175-3564) | 399 (292-484) | 11.78 (8.74-14.21) | 221 (161-283) | 6.55 (4.78-8.44) | 222 (158-288) | 6.56 (4.76-8.48) | 123 (86-165) | 3.64 (2.56-4.78) | 121 (85-163) | 3.60 (2.55-4.81) |
| Greenland | Female | 3 (3-3) | 0 (0-0) | 8.36 (5.92-10.54) | 0 (0-0) | 2.36 (1.52-3.33) | 0 (0-0) | 6.94 (5.07-8.91) | 0 (0-0) | 0.89 (0.54-1.32) | 0 (0-0) | 0.72 (0.43-1.11) |
| Greenland | Male | 5 (4-5) | 1 (0-1) | 11.15 (8.35-13.55) | 0 (0-0) | 3.09 (2.02-4.41) | 0 (0-1) | 9.05 (6.66-11.13) | 0 (0-0) | 1.10 (0.69-1.64) | 0 (0-0) | 0.96 (0.60-1.47) |
| Grenada | Female | 27 (25-29) | 2 (2-3) | 7.82 (5.82-9.77) | 1 (1-2) | 5.21 (3.95-6.57) | 1 (1-1) | 3.92 (2.93-5.04) | 1 (0-1) | 2.61 (1.86-3.48) | 1 (0-1) | 2.30 (1.62-3.08) |
| Grenada | Male | 34 (31-37) | 4 (3-4) | 10.60 (7.99-12.98) | 2 (2-3) | 6.79 (4.96-8.53) | 2 (1-2) | 5.03 (3.72-6.52) | 1 (1-1) | 2.59 (1.77-3.59) | 1 (0-1) | 2.02 (1.32-2.89) |
| Guatemala | Female | 2684 (2500-2881) | 301 (244-360) | 11.28 (9.25-13.11) | 203 (161-249) | 7.56 (6.07-9.07) | 135 (104-169) | 5.06 (3.95-6.21) | 72 (54-94) | 2.69 (2.06-3.43) | 51 (37-68) | 1.90 (1.40-2.52) |
| Guatemala | Male | 2553 (2373-2739) | 379 (309-452) | 14.86 (12.20-17.29) | 247 (201-302) | 9.71 (7.94-11.56) | 166 (133-206) | 6.49 (5.31-7.86) | 65 (49-83) | 2.54 (1.93-3.20) | 46 (35-62) | 1.82 (1.37-2.39) |
| Guinea | Female | 684 (621-742) | 56 (42-71) | 8.23 (6.31-10.15) | 40 (30-51) | 5.86 (4.55-7.32) | 24 (18-31) | 3.55 (2.71-4.48) | 14 (10-19) | 2.01 (1.44-2.68) | 7 (4-10) | 0.97 (0.63-1.45) |
| Guinea | Male | 976 (869-1090) | 85 (64-106) | 8.70 (6.73-10.50) | 60 (46-77) | 6.18 (4.71-7.67) | 36 (27-47) | 3.70 (2.78-4.66) | 13 (9-19) | 1.35 (0.92-1.89) | 8 (5-12) | 0.82 (0.50-1.21) |
| Guinea-Bissau | Female | 116 (105-126) | 9 (7-11) | 7.70 (5.85-9.42) | 6 (5-8) | 5.50 (4.11-6.87) | 4 (3-5) | 3.30 (2.49-4.16) | 2 (1-2) | 1.50 (1.05-2.07) | 1 (1-2) | 0.99 (0.63-1.45) |
| Guinea-Bissau | Male | 162 (148-177) | 13 (10-16) | 7.98 (6.06-9.84) | 9 (7-12) | 5.71 (4.30-7.13) | 5 (4-7) | 3.40 (2.59-4.35) | 2 (1-2) | 1.02 (0.66-1.46) | 1 (1-2) | 0.70 (0.40-1.07) |
| Guyana | Female | 170 (155-182) | 12 (8-15) | 6.87 (4.95-8.76) | 8 (6-10) | 4.61 (3.36-5.85) | 6 (4-8) | 3.58 (2.60-4.60) | 3 (2-5) | 2.02 (1.43-2.65) | 3 (2-4) | 1.61 (1.09-2.14) |
| Guyana | Male | 172 (156-188) | 17 (13-22) | 10.22 (7.91-12.34) | 11 (8-14) | 6.49 (4.97-8.04) | 8 (6-11) | 4.96 (3.72-6.29) | 4 (3-5) | 2.13 (1.57-2.80) | 3 (2-4) | 1.75 (1.30-2.31) |
| Haiti | Female | 999 (797-1193) | 65 (44-90) | 6.63 (4.73-8.58) | 48 (33-66) | 4.84 (3.60-6.29) | 30 (21-42) | 3.05 (2.25-3.95) | 19 (12-27) | 1.89 (1.33-2.60) | 10 (6-15) | 1.02 (0.69-1.47) |
| Haiti | Male | 1006 (777-1221) | 104 (73-136) | 10.44 (8.27-12.44) | 72 (51-97) | 7.24 (5.67-8.80) | 44 (31-60) | 4.38 (3.39-5.56) | 19 (12-27) | 1.88 (1.33-2.52) | 11 (7-17) | 1.09 (0.76-1.57) |
| Honduras | Female | 807 (728-887) | 89 (69-107) | 11.00 (8.78-12.95) | 61 (46-76) | 7.53 (5.90-9.16) | 38 (29-49) | 4.72 (3.61-5.99) | 27 (20-36) | 3.41 (2.53-4.37) | 26 (19-34) | 3.21 (2.36-4.14) |
| Honduras | Male | 605 (538-667) | 93 (72-113) | 15.34 (12.28-18.02) | 62 (48-77) | 10.21 (8.09-12.35) | 39 (29-49) | 6.41 (5.00-7.89) | 21 (15-27) | 3.39 (2.52-4.41) | 18 (13-25) | 3.04 (2.21-4.05) |
| Hungary | Female | 1425 (1318-1550) | 299 (245-356) | 21.07 (17.48-24.29) | 166 (132-208) | 11.67 (9.33-14.29) | 161 (126-200) | 11.29 (8.94-13.85) | 94 (72-124) | 6.64 (5.15-8.56) | 85 (63-115) | 5.93 (4.44-7.90) |
| Hungary | Male | 738 (678-803) | 182 (154-213) | 24.77 (21.37-28.05) | 109 (86-133) | 14.73 (11.82-17.78) | 104 (84-129) | 14.17 (11.58-17.10) | 53 (40-69) | 7.19 (5.45-9.22) | 48 (36-62) | 6.50 (4.98-8.31) |
| Iceland | Female | 22 (20-24) | 2 (1-2) | 7.68 (4.80-10.40) | 0 (0-1) | 1.63 (0.84-2.75) | 2 (1-2) | 7.00 (4.46-9.31) | 0 (0-0) | 0.88 (0.43-1.49) | 0 (0-0) | 0.81 (0.39-1.37) |
| Iceland | Male | 18 (17-20) | 3 (2-3) | 14.21 (11.54-17.08) | 1 (0-1) | 2.71 (1.56-4.30) | 2 (2-3) | 12.35 (9.82-15.10) | 0 (0-0) | 1.57 (0.86-2.50) | 0 (0-0) | 1.49 (0.84-2.42) |
| India | Female | 70318 (65175-75592) | 6708 (5306-7864) | 9.52 (7.62-11.01) | 4589 (3591-5560) | 6.54 (5.12-7.83) | 2855 (2187-3528) | 4.07 (3.15-5.03) | 1004 (738-1280) | 1.42 (1.06-1.79) | 879 (655-1129) | 1.25 (0.94-1.59) |
| India | Male | 102104 (94798-109659) | 12340 (9707-14296) | 12.08 (9.68-13.88) | 8419 (6525-10052) | 8.24 (6.55-9.66) | 5209 (4002-6337) | 5.13 (3.96-6.12) | 1496 (1132-1873) | 1.47 (1.13-1.81) | 1252 (964-1589) | 1.23 (0.95-1.54) |
| Indonesia | Female | 20270 (18618-22028) | 2929 (2338-3552) | 14.42 (11.80-16.99) | 1927 (1499-2347) | 9.47 (7.66-11.40) | 1260 (985-1585) | 6.22 (5.00-7.56) | 482 (361-617) | 2.36 (1.82-2.94) | 259 (191-345) | 1.28 (0.96-1.66) |
| Indonesia | Male | 28584 (25924-30908) | 4686 (3758-5537) | 16.35 (13.44-19.15) | 3082 (2413-3764) | 10.82 (8.58-13.00) | 2030 (1579-2513) | 7.08 (5.66-8.74) | 533 (398-693) | 1.88 (1.41-2.40) | 310 (227-410) | 1.09 (0.80-1.43) |
| Iran | Female | 6650 (6139-7117) | 16 (1-69) | 0.23 (0.01-1.03) | 15 (1-64) | 0.22 (0.01-0.99) | 14 (1-53) | 0.21 (0.01-0.82) | 4 (0-26) | 0.07 (0.00-0.40) | 4 (0-22) | 0.06 (0.00-0.34) |
| Iran | Male | 6786 (6283-7305) | 248 (143-381) | 3.67 (2.10-5.57) | 196 (117-295) | 2.89 (1.70-4.36) | 138 (84-210) | 2.04 (1.23-3.08) | 51 (26-83) | 0.75 (0.38-1.23) | 41 (21-68) | 0.61 (0.31-1.00) |
| Iraq | Female | 3998 (3603-4385) | 13 (2-46) | 0.33 (0.04-1.18) | 13 (2-43) | 0.32 (0.04-1.07) | 11 (1-37) | 0.29 (0.03-0.91) | 4 (0-21) | 0.11 (0.00-0.50) | 3 (0-16) | 0.08 (0.00-0.41) |
| Iraq | Male | 4063 (3583-4549) | 188 (108-278) | 4.64 (2.74-6.77) | 147 (87-211) | 3.61 (2.18-5.12) | 104 (64-147) | 2.56 (1.61-3.55) | 56 (30-87) | 1.40 (0.75-2.11) | 43 (22-69) | 1.06 (0.56-1.68) |
| Ireland | Female | 356 (320-393) | 10 (3-19) | 2.81 (0.87-5.22) | 1 (0-3) | 0.40 (0.14-0.92) | 10 (3-18) | 2.80 (0.87-5.15) | 1 (0-1) | 0.16 (0.05-0.38) | 1 (0-1) | 0.14 (0.04-0.36) |
| Ireland | Male | 345 (312-376) | 36 (25-45) | 10.36 (7.36-12.99) | 3 (2-5) | 0.76 (0.51-1.55) | 34 (24-43) | 9.87 (7.06-12.39) | 1 (1-2) | 0.28 (0.17-0.60) | 1 (1-2) | 0.26 (0.16-0.55) |
| Israel | Female | 1375 (1226-1517) | 111 (73-152) | 8.11 (5.39-10.78) | 38 (24-56) | 2.76 (1.74-4.04) | 91 (61-125) | 6.70 (4.58-8.85) | 18 (11-28) | 1.34 (0.80-2.01) | 17 (10-25) | 1.22 (0.74-1.81) |
| Israel | Male | 1405 (1276-1532) | 169 (133-205) | 12.13 (9.63-14.34) | 53 (36-72) | 3.75 (2.64-5.10) | 132 (100-164) | 9.38 (7.29-11.56) | 24 (16-34) | 1.70 (1.16-2.40) | 22 (15-31) | 1.59 (1.07-2.23) |
| Italy | Female | 10615 (9401-11784) | 1031 (687-1382) | 9.73 (6.45-12.75) | 454 (304-647) | 4.30 (2.93-6.02) | 712 (485-982) | 6.71 (4.56-9.04) | 249 (160-365) | 2.36 (1.55-3.33) | 230 (149-341) | 2.18 (1.43-3.15) |
| Italy | Male | 8475 (7729-9283) | 1104 (829-1374) | 13.01 (9.77-15.87) | 478 (340-643) | 5.63 (4.02-7.49) | 750 (545-967) | 8.81 (6.48-11.16) | 250 (174-342) | 2.95 (2.05-3.94) | 243 (168-334) | 2.86 (1.97-3.85) |
| Jamaica | Female | 430 (396-464) | 34 (25-43) | 7.87 (5.91-9.69) | 24 (18-30) | 5.47 (4.19-6.81) | 16 (12-20) | 3.73 (2.85-4.61) | 13 (9-17) | 2.92 (2.16-3.77) | 11 (8-15) | 2.54 (1.86-3.33) |
| Jamaica | Male | 494 (452-536) | 55 (44-67) | 11.13 (9.16-13.07) | 37 (30-45) | 7.45 (6.04-8.97) | 25 (19-31) | 5.02 (3.99-6.16) | 14 (11-19) | 2.90 (2.23-3.67) | 12 (9-16) | 2.42 (1.84-3.15) |
| Japan | Female | 26524 (23194-29714) | 4092 (3210-5210) | 15.55 (12.32-19.17) | 1665 (1155-2326) | 6.31 (4.43-8.42) | 2850 (2123-3690) | 10.79 (8.29-13.77) | 792 (544-1115) | 2.98 (2.07-4.14) | 737 (496-1047) | 2.80 (1.90-3.88) |
| Japan | Male | 26161 (24052-28255) | 4612 (3711-5581) | 17.63 (14.26-20.66) | 1806 (1364-2345) | 6.90 (5.20-8.87) | 3146 (2462-3923) | 12.05 (9.48-14.64) | 925 (688-1245) | 3.55 (2.64-4.67) | 832 (615-1105) | 3.18 (2.38-4.18) |
| Jordan | Female | 953 (868-1036) | 2 (0-10) | 0.25 (0.03-1.07) | 2 (0-10) | 0.23 (0.02-1.05) | 2 (0-8) | 0.20 (0.02-0.84) | 1 (0-5) | 0.08 (0.00-0.50) | 1 (0-5) | 0.07 (0.00-0.48) |
| Jordan | Male | 869 (782-944) | 37 (22-56) | 4.31 (2.62-6.36) | 29 (18-43) | 3.35 (2.08-4.80) | 20 (13-29) | 2.34 (1.47-3.33) | 10 (6-16) | 1.18 (0.63-1.80) | 9 (5-15) | 1.10 (0.58-1.69) |
| Kazakhstan | Female | 833 (767-897) | 82 (64-103) | 9.94 (7.92-12.24) | 53 (41-67) | 6.42 (5.02-7.98) | 42 (32-53) | 4.98 (3.94-6.34) | 30 (22-38) | 3.55 (2.70-4.59) | 27 (20-36) | 3.27 (2.45-4.28) |
| Kazakhstan | Male | 1101 (1015-1181) | 151 (124-180) | 13.69 (11.42-15.97) | 93 (74-114) | 8.47 (6.84-10.17) | 72 (57-91) | 6.56 (5.26-8.03) | 43 (32-53) | 3.88 (2.98-4.78) | 36 (27-47) | 3.30 (2.53-4.14) |
| Kenya | Female | 3263 (2995-3552) | 429 (352-507) | 13.12 (10.98-15.36) | 292 (238-353) | 8.93 (7.39-10.73) | 180 (145-223) | 5.50 (4.48-6.70) | 76 (57-101) | 2.34 (1.77-3.04) | 38 (27-53) | 1.18 (0.83-1.60) |
| Kenya | Male | 3591 (3245-3925) | 3 (0-29) | 0.10 (0.00-0.80) | 3 (0-29) | 0.10 (0.00-0.80) | 3 (0-23) | 0.09 (0.00-0.63) | 1 (0-5) | 0.01 (0.00-0.15) | 0 (0-2) | 0.01 (0.00-0.07) |
| Kiribati | Female | 14 (13-16) | 1 (1-2) | 9.89 (6.62-12.81) | 1 (1-1) | 6.83 (4.61-8.96) | 1 (0-1) | 4.21 (2.83-5.66) | 0 (0-0) | 1.43 (0.92-2.05) | 0 (0-0) | 0.74 (0.43-1.17) |
| Kiribati | Male | 13 (10-15) | 1 (1-2) | 11.40 (8.57-14.16) | 1 (1-1) | 7.87 (5.79-9.93) | 1 (0-1) | 4.86 (3.54-6.31) | 0 (0-0) | 1.41 (0.96-1.98) | 0 (0-0) | 0.84 (0.53-1.26) |
| Kuwait | Female | 151 (139-164) | 5 (3-8) | 3.62 (2.20-5.22) | 3 (2-5) | 2.21 (1.34-3.17) | 4 (3-6) | 2.86 (1.81-4.09) | 1 (1-2) | 0.97 (0.53-1.47) | 1 (1-2) | 0.93 (0.49-1.42) |
| Kuwait | Male | 215 (196-234) | 20 (16-26) | 9.54 (7.38-11.61) | 10 (7-12) | 4.59 (3.52-5.70) | 13 (10-17) | 6.28 (4.92-7.71) | 4 (3-5) | 1.90 (1.37-2.50) | 4 (3-5) | 1.74 (1.26-2.28) |
| Kyrgyzstan | Female | 209 (195-224) | 16 (12-20) | 7.64 (5.80-9.52) | 12 (9-15) | 5.52 (4.20-6.92) | 7 (5-9) | 3.38 (2.52-4.35) | 5 (4-6) | 2.35 (1.76-3.05) | 4 (3-6) | 2.12 (1.58-2.79) |
| Kyrgyzstan | Male | 257 (239-275) | 30 (24-37) | 11.86 (9.39-14.13) | 21 (16-26) | 8.14 (6.26-9.93) | 13 (10-16) | 4.94 (3.75-6.09) | 5 (4-7) | 2.02 (1.49-2.60) | 4 (3-5) | 1.46 (1.06-1.92) |
| Laos | Female | 815 (719-910) | 109 (84-135) | 13.40 (10.51-16.17) | 73 (56-91) | 9.03 (6.99-10.95) | 46 (35-58) | 5.64 (4.36-7.03) | 16 (12-22) | 2.02 (1.45-2.64) | 12 (8-16) | 1.46 (0.96-1.98) |
| Laos | Male | 868 (765-980) | 137 (109-168) | 15.87 (12.82-18.61) | 92 (73-116) | 10.68 (8.55-12.79) | 58 (44-75) | 6.72 (5.25-8.35) | 17 (12-23) | 1.91 (1.43-2.59) | 12 (8-17) | 1.36 (0.96-1.91) |
| Latvia | Female | 219 (201-239) | 9 (4-14) | 3.96 (1.91-6.39) | 7 (4-10) | 3.04 (1.61-4.54) | 7 (4-10) | 3.09 (1.64-4.62) | 4 (2-6) | 1.63 (0.80-2.55) | 3 (1-5) | 1.35 (0.63-2.11) |
| Latvia | Male | 102 (94-109) | 14 (11-16) | 13.40 (11.18-15.46) | 7 (6-9) | 7.35 (5.93-8.85) | 8 (6-9) | 7.51 (6.08-9.03) | 3 (2-4) | 3.08 (2.36-3.90) | 3 (2-3) | 2.47 (1.83-3.19) |
| Lebanon | Female | 1000 (893-1105) | 1 (0-7) | 0.15 (0.02-0.70) | 1 (0-7) | 0.14 (0.02-0.65) | 1 (0-6) | 0.13 (0.02-0.57) | 0 (0-3) | 0.04 (0.00-0.29) | 0 (0-3) | 0.04 (0.00-0.27) |
| Lebanon | Male | 1084 (973-1205) | 34 (18-57) | 3.15 (1.70-5.29) | 28 (15-46) | 2.55 (1.35-4.32) | 20 (11-34) | 1.85 (0.98-3.05) | 10 (5-18) | 0.90 (0.43-1.64) | 9 (5-17) | 0.86 (0.39-1.54) |
| Lesotho | Female | 227 (200-254) | 13 (8-18) | 5.80 (3.82-8.03) | 10 (7-14) | 4.59 (3.04-6.09) | 7 (4-9) | 2.89 (1.94-3.88) | 4 (2-6) | 1.68 (1.03-2.39) | 4 (2-5) | 1.58 (0.96-2.31) |
| Lesotho | Male | 201 (179-224) | 10 (7-15) | 5.21 (3.27-7.30) | 8 (5-11) | 4.03 (2.60-5.63) | 5 (3-7) | 2.55 (1.66-3.53) | 1 (1-2) | 0.69 (0.36-1.11) | 1 (1-2) | 0.49 (0.27-0.80) |
| Liberia | Female | 356 (321-388) | 28 (22-36) | 8.02 (6.25-9.79) | 20 (16-26) | 5.76 (4.48-7.00) | 12 (9-15) | 3.40 (2.69-4.28) | 6 (4-8) | 1.56 (1.13-2.06) | 4 (2-5) | 0.99 (0.66-1.40) |
| Liberia | Male | 491 (449-534) | 42 (32-52) | 8.61 (6.64-10.37) | 30 (23-37) | 6.10 (4.68-7.56) | 18 (13-23) | 3.64 (2.77-4.63) | 6 (4-9) | 1.30 (0.89-1.80) | 5 (3-7) | 0.92 (0.61-1.30) |
| Libya | Female | 1073 (967-1173) | 4 (1-12) | 0.35 (0.07-1.11) | 3 (1-11) | 0.32 (0.06-1.08) | 3 (1-9) | 0.28 (0.05-0.88) | 1 (0-5) | 0.08 (0.00-0.44) | 1 (0-4) | 0.07 (0.00-0.36) |
| Libya | Male | 1016 (902-1119) | 44 (27-63) | 4.32 (2.70-6.22) | 34 (21-48) | 3.30 (2.11-4.68) | 24 (15-35) | 2.42 (1.53-3.45) | 9 (5-15) | 0.93 (0.51-1.47) | 8 (4-13) | 0.75 (0.39-1.23) |
| Lithuania | Female | 195 (179-211) | 20 (16-25) | 10.28 (8.11-12.51) | 11 (8-14) | 5.43 (4.16-6.91) | 12 (9-15) | 6.07 (4.65-7.61) | 6 (4-7) | 2.87 (2.11-3.74) | 5 (3-6) | 2.31 (1.65-3.14) |
| Lithuania | Male | 135 (124-146) | 14 (11-17) | 10.64 (8.58-12.62) | 8 (6-10) | 5.62 (4.43-6.96) | 8 (7-11) | 6.24 (4.96-7.78) | 3 (3-5) | 2.58 (1.92-3.34) | 2 (2-3) | 1.84 (1.32-2.46) |
| Luxembourg | Female | 97 (88-108) | 8 (5-11) | 8.22 (5.22-11.15) | 1 (0-1) | 0.53 (0.38-0.78) | 8 (5-11) | 7.99 (5.14-10.78) | 0 (0-0) | 0.23 (0.16-0.36) | 0 (0-0) | 0.22 (0.14-0.33) |
| Luxembourg | Male | 61 (55-66) | 7 (5-9) | 11.94 (8.98-14.60) | 0 (0-1) | 0.72 (0.53-1.09) | 7 (5-9) | 11.49 (8.68-14.05) | 0 (0-0) | 0.35 (0.25-0.53) | 0 (0-0) | 0.33 (0.24-0.52) |
| Macedonia | Female | 167 (148-186) | 30 (24-37) | 18.20 (15.01-21.33) | 19 (15-24) | 11.66 (9.29-14.22) | 14 (10-17) | 8.20 (6.44-10.22) | 11 (8-14) | 6.41 (4.85-8.34) | 10 (7-13) | 5.73 (4.22-7.69) |
| Macedonia | Male | 200 (179-223) | 45 (36-53) | 22.42 (18.70-25.43) | 30 (23-37) | 14.90 (11.84-17.87) | 21 (16-27) | 10.66 (8.43-12.96) | 14 (10-18) | 6.92 (5.05-8.97) | 12 (8-16) | 5.88 (4.32-7.87) |
| Madagascar | Female | 1114 (1009-1215) | 134 (108-164) | 12.13 (9.77-14.46) | 93 (75-115) | 8.44 (6.73-10.14) | 55 (43-70) | 4.99 (3.88-6.13) | 22 (16-30) | 1.98 (1.47-2.68) | 12 (8-18) | 1.08 (0.71-1.56) |
| Madagascar | Male | 1241 (1108-1367) | 123 (95-152) | 9.92 (7.95-12.09) | 87 (65-110) | 7.05 (5.43-8.79) | 52 (39-67) | 4.17 (3.21-5.38) | 15 (10-21) | 1.19 (0.79-1.66) | 8 (5-13) | 0.67 (0.38-1.03) |
| Malawi | Female | 1248 (1133-1354) | 165 (132-200) | 13.22 (10.96-15.47) | 114 (89-138) | 9.13 (7.37-10.85) | 67 (52-84) | 5.40 (4.31-6.59) | 38 (29-50) | 3.09 (2.42-3.87) | 28 (20-37) | 2.24 (1.65-2.87) |
| Malawi | Male | 1546 (1409-1685) | 154 (119-191) | 10.00 (7.90-12.16) | 109 (83-137) | 7.08 (5.55-8.73) | 65 (49-85) | 4.20 (3.21-5.34) | 19 (14-26) | 1.25 (0.91-1.69) | 14 (10-20) | 0.90 (0.64-1.25) |
| Malaysia | Female | 3695 (3348-4023) | 576 (475-681) | 15.54 (13.01-18.30) | 349 (284-428) | 9.44 (7.78-11.48) | 276 (222-343) | 7.47 (6.13-9.07) | 140 (110-175) | 3.76 (2.99-4.69) | 127 (99-159) | 3.44 (2.73-4.31) |
| Malaysia | Male | 3772 (3423-4122) | 664 (547-779) | 17.61 (14.73-20.27) | 405 (322-491) | 10.72 (8.88-12.80) | 319 (257-395) | 8.45 (6.95-10.35) | 135 (104-172) | 3.58 (2.82-4.51) | 125 (96-160) | 3.31 (2.62-4.15) |
| Maldives | Female | 33 (30-36) | 5 (4-6) | 15.01 (12.56-17.81) | 3 (2-4) | 9.30 (7.64-11.20) | 2 (2-3) | 7.14 (5.84-8.80) | 1 (1-1) | 3.02 (2.25-3.92) | 1 (1-1) | 2.43 (1.72-3.32) |
| Maldives | Male | 53 (48-57) | 8 (7-10) | 16.06 (13.37-18.46) | 5 (4-6) | 9.95 (8.19-11.79) | 4 (3-5) | 7.62 (6.18-9.13) | 1 (1-2) | 2.48 (1.85-3.26) | 1 (1-1) | 1.95 (1.43-2.63) |
| Mali | Female | 1110 (1007-1210) | 86 (62-109) | 7.77 (5.71-9.60) | 61 (46-79) | 5.54 (4.15-6.97) | 37 (27-47) | 3.32 (2.44-4.16) | 20 (13-27) | 1.80 (1.25-2.39) | 14 (9-20) | 1.28 (0.85-1.80) |
| Mali | Male | 1501 (1347-1636) | 123 (89-157) | 8.18 (6.06-10.38) | 87 (63-114) | 5.78 (4.28-7.43) | 52 (38-67) | 3.47 (2.53-4.47) | 21 (13-30) | 1.39 (0.91-1.99) | 15 (10-23) | 1.02 (0.65-1.55) |
| Malta | Female | 84 (75-92) | 8 (5-10) | 9.35 (6.66-11.95) | 4 (3-5) | 4.31 (3.15-5.77) | 5 (4-7) | 6.37 (4.74-8.35) | 2 (1-3) | 2.12 (1.48-2.92) | 2 (1-2) | 2.08 (1.46-2.88) |
| Malta | Male | 57 (51-62) | 7 (6-9) | 12.80 (10.16-15.18) | 3 (2-4) | 5.65 (4.22-7.25) | 5 (4-6) | 8.49 (6.58-10.39) | 2 (1-2) | 2.89 (2.14-3.82) | 2 (1-2) | 2.84 (2.09-3.78) |
| Marshall Islands | Female | 8 (5-11) | 1 (0-1) | 9.18 (5.70-12.15) | 0 (0-1) | 6.06 (3.64-8.09) | 0 (0-1) | 4.20 (2.61-5.70) | 0 (0-0) | 1.59 (0.92-2.27) | 0 (0-0) | 1.15 (0.63-1.72) |
| Marshall Islands | Male | 9 (5-12) | 1 (0-2) | 11.31 (6.78-15.31) | 1 (0-1) | 7.47 (4.52-10.19) | 0 (0-1) | 5.20 (3.16-7.38) | 0 (0-0) | 1.52 (0.78-2.47) | 0 (0-0) | 1.10 (0.49-1.83) |
| Mauritania | Female | 358 (318-400) | 30 (22-39) | 8.53 (6.49-10.38) | 21 (16-28) | 5.99 (4.43-7.42) | 13 (10-17) | 3.69 (2.76-4.60) | 6 (4-9) | 1.79 (1.21-2.40) | 4 (3-6) | 1.16 (0.74-1.73) |
| Mauritania | Male | 369 (327-409) | 34 (25-43) | 9.19 (6.79-11.22) | 23 (17-30) | 6.38 (4.86-8.00) | 14 (11-19) | 3.92 (2.97-4.95) | 5 (3-7) | 1.34 (0.88-1.91) | 3 (2-6) | 0.94 (0.57-1.46) |
| Mauritius | Female | 679 (633-722) | 105 (87-125) | 15.53 (12.97-18.35) | 65 (53-80) | 9.69 (7.98-11.49) | 49 (40-60) | 7.23 (5.87-8.84) | 29 (24-36) | 4.35 (3.54-5.30) | 29 (23-35) | 4.23 (3.42-5.13) |
| Mauritius | Male | 722 (675-772) | 124 (100-148) | 17.22 (14.03-20.07) | 78 (61-96) | 10.75 (8.52-13.18) | 58 (45-73) | 8.06 (6.28-10.01) | 30 (23-38) | 4.17 (3.23-5.33) | 29 (22-37) | 3.93 (3.03-5.06) |
| Mexico | Female | 33084 (30659-35360) | 3131 (2403-3850) | 9.51 (7.39-11.45) | 1972 (1509-2462) | 5.95 (4.54-7.33) | 1520 (1164-1906) | 4.62 (3.56-5.70) | 761 (576-968) | 2.30 (1.73-2.88) | 696 (524-882) | 2.10 (1.59-2.63) |
| Mexico | Male | 33007 (30762-35433) | 3903 (3149-4578) | 11.83 (9.70-13.74) | 2464 (1944-2970) | 7.46 (5.97-8.95) | 1919 (1500-2350) | 5.81 (4.60-7.02) | 665 (515-831) | 2.01 (1.56-2.47) | 596 (459-749) | 1.80 (1.41-2.25) |
| Moldova | Female | 102 (93-112) | 3 (2-6) | 3.26 (1.59-5.41) | 3 (1-5) | 2.80 (1.41-4.50) | 2 (1-3) | 2.15 (1.10-3.22) | 1 (1-2) | 1.29 (0.58-2.16) | 1 (0-2) | 0.97 (0.43-1.66) |
| Moldova | Male | 101 (93-109) | 10 (8-12) | 9.78 (7.55-11.65) | 7 (5-8) | 6.58 (5.02-8.09) | 4 (3-6) | 4.42 (3.34-5.53) | 2 (2-3) | 2.41 (1.76-3.13) | 2 (1-2) | 1.49 (1.04-2.07) |
| Mongolia | Female | 130 (119-141) | 12 (9-14) | 9.11 (7.29-10.93) | 8 (6-10) | 6.26 (4.95-7.64) | 5 (4-7) | 4.17 (3.29-5.17) | 4 (3-5) | 2.75 (2.10-3.46) | 3 (2-4) | 2.48 (1.89-3.18) |
| Mongolia | Male | 151 (136-164) | 20 (16-24) | 13.42 (11.10-15.61) | 13 (11-16) | 8.87 (7.27-10.58) | 9 (7-11) | 5.85 (4.72-7.15) | 5 (4-6) | 3.17 (2.56-3.98) | 4 (3-5) | 2.37 (1.86-3.00) |
| Montenegro | Female | 75 (67-83) | 14 (11-17) | 19.05 (15.56-22.05) | 9 (7-11) | 11.80 (9.38-14.46) | 7 (5-9) | 9.00 (7.02-11.06) | 5 (4-6) | 6.55 (4.99-8.34) | 4 (3-6) | 5.88 (4.32-7.87) |
| Montenegro | Male | 89 (80-99) | 20 (17-24) | 22.62 (19.49-25.45) | 13 (10-16) | 14.65 (12.15-17.49) | 10 (8-13) | 11.19 (9.08-13.69) | 6 (5-8) | 6.78 (5.14-8.63) | 5 (4-7) | 5.76 (4.26-7.63) |
| Morocco | Female | 6214 (5643-6861) | 19 (3-71) | 0.31 (0.04-1.12) | 18 (2-66) | 0.30 (0.03-1.07) | 16 (2-57) | 0.26 (0.03-0.92) | 4 (0-22) | 0.07 (0.00-0.36) | 3 (0-18) | 0.05 (0.00-0.29) |
| Morocco | Male | 5530 (4876-6217) | 228 (127-355) | 4.13 (2.36-6.22) | 182 (103-275) | 3.28 (1.91-4.80) | 127 (73-189) | 2.27 (1.33-3.29) | 42 (22-70) | 0.77 (0.39-1.24) | 29 (14-50) | 0.53 (0.25-0.89) |
| Mozambique | Female | 1338 (1210-1466) | 179 (145-217) | 13.40 (11.29-15.67) | 124 (100-152) | 9.29 (7.70-11.07) | 73 (58-91) | 5.49 (4.35-6.66) | 27 (20-36) | 2.04 (1.50-2.61) | 15 (10-21) | 1.08 (0.75-1.53) |
| Mozambique | Male | 1725 (1532-1906) | 165 (128-205) | 9.60 (7.74-11.49) | 118 (90-149) | 6.85 (5.40-8.42) | 70 (52-90) | 4.05 (3.15-5.06) | 16 (11-22) | 0.91 (0.64-1.25) | 8 (5-12) | 0.48 (0.30-0.69) |
| Myanmar | Female | 5589 (5069-6115) | 784 (638-941) | 14.02 (11.65-16.57) | 533 (427-656) | 9.56 (7.72-11.45) | 321 (253-401) | 5.76 (4.57-7.08) | 184 (142-235) | 3.29 (2.58-4.11) | 120 (88-154) | 2.14 (1.63-2.74) |
| Myanmar | Male | 5003 (4538-5475) | 805 (654-956) | 16.15 (13.31-18.73) | 548 (431-673) | 10.99 (8.86-13.17) | 331 (262-417) | 6.66 (5.30-8.27) | 147 (111-192) | 2.95 (2.26-3.75) | 87 (63-117) | 1.73 (1.26-2.30) |
| Namibia | Female | 121 (108-134) | 7 (4-10) | 5.62 (3.53-7.91) | 5 (3-7) | 4.33 (2.86-5.75) | 4 (2-5) | 3.02 (2.01-3.97) | 2 (1-3) | 1.47 (0.90-2.09) | 2 (1-2) | 1.31 (0.77-1.87) |
| Namibia | Male | 183 (165-201) | 10 (6-14) | 5.40 (3.50-7.48) | 7 (5-10) | 4.03 (2.67-5.58) | 5 (3-7) | 2.84 (1.88-3.91) | 2 (1-3) | 1.07 (0.62-1.63) | 2 (1-3) | 0.96 (0.54-1.51) |
| Nauru | Female | 2 (1-2) | 0 (0-0) | 11.06 (8.92-13.51) | 0 (0-0) | 6.86 (5.38-8.64) | 0 (0-0) | 5.55 (4.34-6.94) | 0 (0-0) | 2.31 (1.73-3.11) | 0 (0-0) | 1.40 (0.96-2.02) |
| Nauru | Male | 2 (1-2) | 0 (0-0) | 12.64 (9.99-15.28) | 0 (0-0) | 7.80 (6.09-9.73) | 0 (0-0) | 6.27 (4.75-7.85) | 0 (0-0) | 2.39 (1.75-3.20) | 0 (0-0) | 1.65 (1.16-2.35) |
| Nepal | Female | 1670 (1516-1829) | 138 (89-183) | 8.31 (5.37-10.74) | 97 (63-133) | 5.85 (3.70-7.70) | 59 (37-81) | 3.53 (2.28-4.84) | 19 (11-26) | 1.12 (0.68-1.54) | 13 (8-19) | 0.80 (0.49-1.13) |
| Nepal | Male | 2254 (2008-2476) | 256 (196-310) | 11.34 (9.11-13.46) | 179 (135-222) | 7.93 (6.17-9.65) | 108 (79-137) | 4.79 (3.67-5.95) | 30 (22-40) | 1.34 (1.01-1.73) | 18 (13-24) | 0.79 (0.58-1.04) |
| Netherlands | Female | 2459 (2268-2655) | 184 (117-256) | 7.51 (4.83-10.28) | 53 (32-86) | 2.18 (1.31-3.49) | 161 (104-219) | 6.53 (4.23-8.81) | 23 (13-39) | 0.95 (0.53-1.54) | 19 (11-33) | 0.79 (0.45-1.32) |
| Netherlands | Male | 1962 (1832-2102) | 229 (176-288) | 11.74 (9.07-14.48) | 59 (35-90) | 3.02 (1.81-4.54) | 188 (140-241) | 9.55 (7.14-12.20) | 26 (15-40) | 1.30 (0.77-2.08) | 24 (14-38) | 1.23 (0.73-1.94) |
| New Zealand | Female | 468 (429-507) | 27 (17-38) | 5.85 (3.74-8.19) | 11 (7-16) | 2.38 (1.54-3.33) | 23 (15-33) | 5.00 (3.15-6.94) | 5 (3-7) | 1.06 (0.64-1.53) | 4 (3-7) | 0.95 (0.58-1.37) |
| New Zealand | Male | 481 (444-517) | 52 (41-64) | 10.88 (8.59-13.04) | 17 (12-23) | 3.58 (2.55-4.78) | 40 (31-50) | 8.36 (6.53-10.24) | 7 (5-10) | 1.43 (0.98-1.98) | 6 (4-9) | 1.30 (0.89-1.81) |
| Nicaragua | Female | 1017 (934-1107) | 117 (96-138) | 11.54 (9.52-13.21) | 81 (64-98) | 7.96 (6.46-9.47) | 49 (39-62) | 4.86 (3.88-6.04) | 35 (27-45) | 3.45 (2.66-4.27) | 32 (24-42) | 3.20 (2.41-4.08) |
| Nicaragua | Male | 1605 (1480-1732) | 241 (199-288) | 15.10 (12.41-17.63) | 161 (131-196) | 10.14 (8.30-11.96) | 100 (79-123) | 6.25 (4.98-7.60) | 47 (36-62) | 2.93 (2.29-3.79) | 43 (32-57) | 2.66 (1.98-3.49) |
| Niger | Female | 630 (562-699) | 51 (37-65) | 8.14 (6.10-9.97) | 37 (26-47) | 5.80 (4.28-7.21) | 22 (16-28) | 3.44 (2.54-4.30) | 7 (5-11) | 1.17 (0.77-1.61) | 3 (2-5) | 0.55 (0.33-0.87) |
| Niger | Male | 1067 (921-1214) | 92 (65-117) | 8.62 (6.28-10.62) | 65 (46-84) | 6.12 (4.56-7.69) | 39 (28-51) | 3.64 (2.65-4.61) | 8 (5-13) | 0.78 (0.48-1.16) | 6 (3-9) | 0.53 (0.31-0.84) |
| Nigeria | Female | 8755 (8019-9518) | 601 (430-789) | 6.90 (5.00-8.76) | 443 (330-570) | 5.09 (3.78-6.37) | 279 (210-367) | 3.19 (2.40-4.09) | 120 (85-160) | 1.38 (0.99-1.79) | 67 (46-90) | 0.77 (0.53-1.03) |
| Nigeria | Male | 13346 (12271-14546) | 1038 (763-1317) | 7.79 (5.71-9.67) | 741 (546-954) | 5.57 (4.17-7.02) | 462 (348-589) | 3.46 (2.63-4.35) | 150 (105-207) | 1.13 (0.79-1.52) | 107 (74-150) | 0.80 (0.57-1.11) |
| Norway | Female | 392 (353-433) | 16 (6-28) | 4.12 (1.60-7.08) | 2 (1-6) | 0.57 (0.26-1.36) | 16 (6-27) | 4.06 (1.60-6.92) | 1 (0-3) | 0.26 (0.11-0.66) | 1 (0-2) | 0.18 (0.08-0.47) |
| Norway | Male | 412 (377-446) | 51 (39-63) | 12.25 (9.52-14.85) | 4 (3-9) | 1.04 (0.65-2.18) | 48 (36-59) | 11.54 (8.91-14.14) | 2 (1-5) | 0.51 (0.30-1.12) | 2 (1-4) | 0.47 (0.27-1.00) |
| Oman | Female | 249 (225-273) | 1 (0-4) | 0.38 (0.06-1.48) | 1 (0-3) | 0.34 (0.05-1.26) | 1 (0-3) | 0.34 (0.05-1.23) | 0 (0-1) | 0.09 (0.00-0.49) | 0 (0-1) | 0.08 (0.00-0.46) |
| Oman | Male | 353 (312-392) | 17 (11-24) | 4.92 (3.07-6.74) | 11 (7-16) | 3.20 (2.08-4.42) | 11 (7-16) | 3.18 (2.07-4.43) | 3 (2-5) | 0.85 (0.49-1.27) | 3 (1-4) | 0.75 (0.42-1.15) |
| Pakistan | Female | 14253 (13088-15460) | 1556 (1281-1834) | 10.97 (9.11-12.68) | 1081 (873-1300) | 7.60 (6.18-9.02) | 654 (525-813) | 4.63 (3.71-5.60) | 336 (260-426) | 2.37 (1.87-2.98) | 282 (213-368) | 1.98 (1.51-2.57) |
| Pakistan | Male | 14720 (13502-15886) | 1687 (1397-2008) | 11.53 (9.56-13.35) | 1173 (952-1440) | 7.99 (6.61-9.54) | 714 (568-897) | 4.86 (3.92-5.97) | 207 (156-274) | 1.41 (1.08-1.82) | 159 (116-212) | 1.08 (0.80-1.43) |
| Palau | Female | 4 (4-5) | 1 (0-1) | 13.31 (10.59-16.27) | 0 (0-0) | 8.04 (6.25-10.22) | 0 (0-0) | 6.68 (5.12-8.52) | 0 (0-0) | 3.29 (2.46-4.36) | 0 (0-0) | 2.85 (2.10-3.86) |
| Palau | Male | 4 (4-5) | 1 (0-1) | 12.85 (10.04-15.59) | 0 (0-0) | 7.80 (6.11-9.72) | 0 (0-0) | 6.49 (4.96-8.18) | 0 (0-0) | 2.49 (1.88-3.21) | 0 (0-0) | 1.89 (1.40-2.51) |
| Panama | Female | 545 (505-588) | 65 (54-78) | 12.02 (10.12-13.92) | 39 (31-48) | 7.12 (5.79-8.63) | 35 (27-43) | 6.35 (5.06-7.62) | 18 (13-22) | 3.21 (2.51-4.04) | 16 (12-20) | 2.87 (2.23-3.64) |
| Panama | Male | 817 (754-880) | 128 (106-153) | 15.74 (13.33-18.31) | 74 (61-90) | 9.11 (7.57-10.78) | 66 (54-81) | 8.05 (6.63-9.66) | 28 (22-35) | 3.42 (2.68-4.27) | 24 (18-31) | 2.91 (2.29-3.74) |
| Papua New Guinea | Female | 226 (201-250) | 18 (12-25) | 8.20 (5.46-10.53) | 13 (8-17) | 5.73 (3.79-7.50) | 8 (5-11) | 3.64 (2.38-4.85) | 2 (1-3) | 1.02 (0.59-1.51) | 1 (1-2) | 0.62 (0.34-0.98) |
| Papua New Guinea | Male | 405 (359-452) | 40 (28-52) | 9.85 (6.93-12.55) | 28 (19-37) | 6.83 (4.78-8.90) | 18 (12-24) | 4.35 (2.97-5.70) | 3 (2-5) | 0.82 (0.49-1.25) | 2 (1-4) | 0.56 (0.29-0.88) |
| Paraguay | Female | 552 (502-603) | 65 (54-78) | 11.85 (9.91-13.76) | 43 (34-53) | 7.82 (6.29-9.31) | 30 (23-37) | 5.37 (4.30-6.59) | 22 (17-28) | 4.05 (3.15-4.97) | 18 (14-24) | 3.35 (2.58-4.21) |
| Paraguay | Male | 808 (734-886) | 113 (91-136) | 14.01 (11.55-16.62) | 74 (59-91) | 9.18 (7.36-11.01) | 51 (40-64) | 6.30 (5.08-7.76) | 32 (24-42) | 4.01 (3.09-5.04) | 27 (20-36) | 3.33 (2.50-4.36) |
| Peru | Female | 5278 (4753-5767) | 589 (467-709) | 11.17 (9.02-13.07) | 387 (297-475) | 7.32 (5.77-8.79) | 270 (205-346) | 5.14 (3.98-6.32) | 106 (80-135) | 2.02 (1.55-2.50) | 97 (72-123) | 1.84 (1.40-2.30) |
| Peru | Male | 4922 (4386-5473) | 657 (529-809) | 13.40 (10.90-16.00) | 427 (335-541) | 8.72 (6.95-10.52) | 299 (230-383) | 6.10 (4.80-7.50) | 97 (73-128) | 1.96 (1.51-2.50) | 83 (63-110) | 1.69 (1.30-2.14) |
| Philippines | Female | 14587 (13503-15633) | 1931 (1516-2347) | 13.27 (10.56-15.92) | 1298 (997-1593) | 8.84 (7.00-10.78) | 828 (642-1037) | 5.69 (4.49-7.01) | 403 (306-510) | 2.76 (2.13-3.45) | 299 (224-387) | 2.06 (1.55-2.64) |
| Philippines | Male | 17010 (15625-18277) | 2565 (1943-3055) | 15.06 (11.67-17.79) | 1717 (1305-2106) | 10.12 (7.67-12.34) | 1106 (807-1384) | 6.51 (4.78-8.14) | 404 (293-514) | 2.36 (1.72-3.00) | 282 (201-372) | 1.66 (1.19-2.16) |
| Poland | Female | 2697 (2497-2928) | 445 (347-539) | 16.51 (13.17-19.57) | 250 (190-315) | 9.31 (7.19-11.36) | 238 (182-298) | 8.85 (6.85-10.84) | 157 (120-199) | 5.87 (4.49-7.29) | 153 (116-194) | 5.66 (4.32-7.04) |
| Poland | Male | 2251 (2078-2420) | 430 (350-512) | 19.15 (15.71-22.34) | 246 (195-304) | 10.87 (8.74-13.40) | 234 (185-291) | 10.40 (8.37-12.82) | 128 (100-163) | 5.68 (4.45-7.12) | 118 (90-153) | 5.25 (4.12-6.65) |
| Portugal | Female | 2718 (2432-3011) | 124 (56-216) | 4.55 (2.06-7.83) | 86 (41-137) | 3.21 (1.49-4.95) | 95 (45-154) | 3.52 (1.64-5.53) | 49 (22-80) | 1.82 (0.78-2.90) | 48 (22-78) | 1.76 (0.81-2.85) |
| Portugal | Male | 2002 (1811-2207) | 203 (147-264) | 10.20 (7.33-12.95) | 108 (78-146) | 5.43 (3.91-7.16) | 121 (88-164) | 6.09 (4.42-8.01) | 57 (39-79) | 2.85 (1.95-3.88) | 54 (38-75) | 2.72 (1.87-3.62) |
| Puerto Rico | Female | 1172 (1100-1243) | 113 (84-141) | 9.58 (7.35-11.94) | 54 (39-68) | 4.55 (3.34-5.81) | 74 (55-95) | 6.34 (4.77-8.04) | 27 (19-37) | 2.29 (1.63-3.13) | 23 (16-32) | 1.95 (1.33-2.71) |
| Puerto Rico | Male | 1358 (1283-1430) | 164 (135-196) | 12.16 (9.89-14.36) | 76 (60-97) | 5.62 (4.36-7.15) | 107 (85-131) | 7.91 (6.32-9.55) | 30 (22-41) | 2.24 (1.66-3.06) | 26 (19-37) | 1.94 (1.40-2.74) |
| Qatar | Female | 73 (67-81) | 0 (0-1) | 0.29 (0.00-1.28) | 0 (0-0) | 0.14 (0.00-0.62) | 0 (0-1) | 0.28 (0.00-1.28) | 0 (0-0) | 0.05 (0.00-0.31) | 0 (0-0) | 0.05 (0.00-0.30) |
| Qatar | Male | 120 (107-134) | 6 (4-8) | 4.83 (3.13-6.65) | 1 (1-2) | 1.18 (0.69-1.81) | 5 (3-8) | 4.40 (2.94-6.05) | 0 (0-1) | 0.35 (0.18-0.59) | 0 (0-1) | 0.32 (0.16-0.54) |
| Romania | Female | 1679 (1571-1786) | 320 (260-377) | 19.00 (15.59-22.34) | 186 (145-228) | 11.08 (8.69-13.47) | 163 (128-202) | 9.69 (7.62-12.10) | 100 (76-126) | 5.98 (4.54-7.51) | 88 (66-111) | 5.27 (4.01-6.53) |
| Romania | Male | 1632 (1545-1720) | 367 (304-427) | 22.62 (18.98-25.92) | 225 (179-273) | 13.78 (11.02-16.61) | 197 (157-241) | 12.07 (9.66-14.60) | 107 (83-135) | 6.60 (5.13-8.21) | 96 (74-122) | 5.88 (4.56-7.38) |
| Russia | Female | 8928 (8297-9626) | 901 (688-1115) | 10.13 (7.94-12.18) | 560 (438-711) | 6.28 (4.97-7.86) | 457 (346-580) | 5.13 (3.94-6.49) | 299 (222-390) | 3.36 (2.50-4.31) | 214 (156-278) | 2.40 (1.75-3.10) |
| Russia | Male | 5458 (5077-5825) | 783 (635-930) | 14.44 (11.76-16.80) | 473 (376-579) | 8.71 (6.94-10.41) | 387 (302-479) | 7.12 (5.59-8.71) | 198 (153-247) | 3.63 (2.82-4.48) | 140 (106-179) | 2.57 (1.95-3.23) |
| Rwanda | Female | 955 (852-1066) | 124 (97-152) | 12.99 (10.56-15.36) | 85 (66-107) | 8.98 (7.21-10.79) | 51 (39-65) | 5.33 (4.22-6.56) | 13 (9-19) | 1.39 (0.94-1.88) | 6 (4-10) | 0.66 (0.38-1.04) |
| Rwanda | Male | 956 (856-1047) | 96 (73-120) | 10.10 (7.84-12.27) | 68 (52-87) | 7.12 (5.48-8.73) | 41 (30-53) | 4.29 (3.23-5.40) | 7 (5-11) | 0.76 (0.48-1.09) | 5 (3-7) | 0.48 (0.28-0.73) |
| Saint Kitts and Nevis | Female | 15 (14-16) | 1 (1-2) | 8.01 (5.79-9.92) | 1 (1-1) | 4.80 (3.51-6.04) | 1 (0-1) | 4.57 (3.29-5.77) | 0 (0-0) | 2.42 (1.66-3.14) | 0 (0-0) | 2.20 (1.50-2.97) |
| Saint Kitts and Nevis | Male | 19 (17-20) | 2 (2-2) | 11.02 (8.55-12.97) | 1 (1-1) | 6.31 (4.87-7.77) | 1 (1-1) | 6.00 (4.47-7.43) | 0 (0-1) | 2.31 (1.70-3.04) | 0 (0-0) | 1.91 (1.38-2.58) |
| Saint Lucia | Female | 42 (39-45) | 3 (3-4) | 8.30 (6.45-10.19) | 2 (2-3) | 5.38 (4.17-6.64) | 2 (1-2) | 4.26 (3.30-5.31) | 1 (1-2) | 2.84 (2.16-3.66) | 1 (1-1) | 2.39 (1.80-3.17) |
| Saint Lucia | Male | 49 (46-53) | 5 (4-7) | 11.17 (9.08-13.05) | 3 (3-4) | 6.97 (5.59-8.54) | 3 (2-3) | 5.49 (4.36-6.81) | 1 (1-2) | 2.56 (1.94-3.28) | 1 (1-1) | 1.95 (1.43-2.57) |
| Saint Vincent and the Grenadines | Female | 24 (22-25) | 2 (1-2) | 7.69 (5.68-9.61) | 1 (1-2) | 5.15 (3.92-6.52) | 1 (1-1) | 3.82 (2.90-4.87) | 1 (0-1) | 2.43 (1.75-3.24) | 0 (0-1) | 2.06 (1.45-2.75) |
| Saint Vincent and the Grenadines | Male | 30 (28-32) | 3 (3-4) | 11.06 (8.80-13.10) | 2 (2-3) | 7.14 (5.50-8.62) | 2 (1-2) | 5.21 (4.10-6.61) | 1 (1-1) | 2.51 (1.82-3.25) | 1 (0-1) | 2.02 (1.48-2.73) |
| Samoa | Female | 31 (28-34) | 2 (1-2) | 5.16 (3.70-6.98) | 1 (1-2) | 3.67 (2.53-5.06) | 1 (1-1) | 2.48 (1.69-3.53) | 0 (0-0) | 0.87 (0.52-1.43) | 0 (0-0) | 0.33 (0.15-0.57) |
| Samoa | Male | 25 (23-28) | 2 (1-2) | 6.46 (4.84-8.49) | 1 (1-2) | 4.56 (3.35-6.03) | 1 (1-1) | 3.08 (2.24-4.09) | 0 (0-0) | 0.98 (0.63-1.45) | 0 (0-0) | 0.65 (0.39-1.06) |
| Sao Tome and Principe | Female | 37 (34-40) | 3 (2-4) | 8.64 (6.69-10.65) | 2 (2-3) | 6.06 (4.71-7.56) | 1 (1-2) | 3.78 (2.90-4.76) | 1 (1-1) | 2.16 (1.62-2.83) | 1 (0-1) | 1.44 (1.03-2.00) |
| Sao Tome and Principe | Male | 29 (26-32) | 3 (2-3) | 9.00 (7.20-10.87) | 2 (1-2) | 6.30 (5.03-7.79) | 1 (1-1) | 3.89 (3.03-4.84) | 0 (0-1) | 1.41 (1.04-1.88) | 0 (0-0) | 0.87 (0.59-1.18) |
| Saudi Arabia | Female | 5324 (4865-5792) | 19 (1-54) | 0.36 (0.01-1.02) | 15 (1-45) | 0.29 (0.01-0.84) | 16 (1-49) | 0.30 (0.01-0.89) | 3 (0-14) | 0.05 (0.00-0.27) | 2 (0-12) | 0.04 (0.00-0.23) |
| Saudi Arabia | Male | 6702 (6053-7313) | 315 (200-453) | 4.73 (3.04-6.71) | 194 (125-272) | 2.91 (1.91-3.97) | 216 (140-301) | 3.21 (2.13-4.45) | 46 (26-72) | 0.70 (0.39-1.03) | 37 (21-59) | 0.56 (0.32-0.87) |
| Senegal | Female | 1213 (1096-1322) | 103 (77-132) | 8.53 (6.55-10.61) | 73 (55-93) | 6.04 (4.68-7.44) | 45 (33-58) | 3.70 (2.84-4.69) | 21 (14-29) | 1.71 (1.21-2.31) | 11 (7-16) | 0.89 (0.57-1.31) |
| Senegal | Male | 1583 (1429-1741) | 139 (103-177) | 8.77 (6.62-10.90) | 98 (73-127) | 6.16 (4.68-7.79) | 59 (44-77) | 3.75 (2.83-4.81) | 19 (13-28) | 1.21 (0.83-1.72) | 11 (7-18) | 0.71 (0.47-1.09) |
| Serbia | Female | 1263 (1153-1368) | 244 (197-288) | 19.42 (15.92-22.50) | 153 (120-188) | 12.05 (9.63-14.48) | 114 (89-141) | 9.06 (7.26-11.01) | 94 (71-117) | 7.42 (5.72-9.06) | 88 (67-112) | 6.96 (5.38-8.70) |
| Serbia | Male | 1444 (1324-1561) | 333 (276-386) | 23.06 (19.66-26.18) | 216 (174-263) | 14.99 (12.24-17.94) | 163 (129-204) | 11.39 (8.98-13.88) | 108 (84-137) | 7.51 (5.88-9.27) | 99 (77-126) | 6.83 (5.32-8.60) |
| Seychelles | Female | 24 (22-26) | 3 (3-4) | 14.19 (11.77-16.84) | 2 (2-3) | 8.55 (6.91-10.41) | 2 (1-2) | 7.08 (5.72-8.77) | 1 (1-1) | 4.46 (3.51-5.56) | 1 (1-1) | 4.08 (3.15-5.18) |
| Seychelles | Male | 26 (23-28) | 4 (3-5) | 15.22 (12.42-17.74) | 2 (2-3) | 9.16 (7.12-11.01) | 2 (2-2) | 7.58 (5.97-9.26) | 1 (1-1) | 3.73 (2.92-4.73) | 1 (1-1) | 3.39 (2.56-4.31) |
| Sierra Leone | Female | 382 (346-419) | 32 (23-40) | 8.34 (6.09-10.23) | 23 (17-29) | 5.98 (4.55-7.37) | 13 (10-17) | 3.53 (2.66-4.36) | 6 (4-8) | 1.55 (1.05-2.11) | 4 (2-5) | 0.93 (0.59-1.35) |
| Sierra Leone | Male | 541 (484-591) | 48 (36-60) | 8.88 (6.81-10.84) | 34 (26-43) | 6.29 (4.89-7.87) | 20 (15-26) | 3.70 (2.83-4.68) | 6 (4-8) | 1.07 (0.72-1.54) | 3 (2-5) | 0.64 (0.38-0.97) |
| Singapore | Female | 452 (418-485) | 67 (51-82) | 14.73 (11.46-17.90) | 9 (5-16) | 2.06 (1.09-3.48) | 60 (46-75) | 13.23 (10.34-16.45) | 5 (2-9) | 1.07 (0.54-1.87) | 5 (2-8) | 1.03 (0.53-1.78) |
| Singapore | Male | 380 (350-408) | 62 (47-76) | 16.31 (12.53-19.61) | 9 (5-14) | 2.33 (1.27-3.77) | 55 (42-69) | 14.67 (11.18-17.72) | 4 (2-7) | 1.12 (0.60-1.89) | 4 (2-7) | 1.07 (0.57-1.81) |
| Slovakia | Female | 440 (395-488) | 83 (66-101) | 18.92 (15.68-22.14) | 45 (34-56) | 10.08 (8.15-12.26) | 47 (36-59) | 10.59 (8.31-13.08) | 26 (19-33) | 5.82 (4.52-7.38) | 24 (18-31) | 5.43 (4.19-6.88) |
| Slovakia | Male | 387 (349-421) | 86 (72-100) | 22.39 (18.97-25.28) | 48 (38-59) | 12.55 (10.18-15.01) | 51 (40-62) | 13.15 (10.56-15.66) | 25 (19-32) | 6.53 (5.01-8.14) | 23 (18-30) | 6.05 (4.62-7.67) |
| Slovenia | Female | 209 (187-234) | 43 (34-52) | 20.64 (16.56-24.21) | 21 (15-27) | 9.89 (7.47-12.43) | 26 (20-34) | 12.60 (9.69-15.89) | 12 (8-16) | 5.63 (4.11-7.34) | 11 (7-14) | 5.07 (3.61-6.69) |
| Slovenia | Male | 118 (108-128) | 28 (23-33) | 23.84 (19.96-27.19) | 14 (11-18) | 12.08 (9.25-15.16) | 18 (14-23) | 15.32 (12.28-18.81) | 7 (5-9) | 5.71 (4.09-7.70) | 6 (4-8) | 4.86 (3.46-6.83) |
| Solomon Islands | Female | 39 (35-44) | 3 (2-4) | 8.53 (6.04-10.81) | 2 (2-3) | 5.92 (4.13-7.55) | 1 (1-2) | 3.69 (2.59-4.74) | 1 (0-1) | 1.48 (1.01-2.00) | 0 (0-0) | 0.60 (0.36-0.89) |
| Solomon Islands | Male | 45 (40-51) | 4 (3-5) | 8.94 (6.28-11.50) | 3 (2-4) | 6.17 (4.36-8.18) | 2 (1-2) | 3.90 (2.70-5.16) | 0 (0-0) | 0.70 (0.44-1.04) | 0 (0-0) | 0.36 (0.21-0.56) |
| Somalia | Female | 1116 (1002-1240) | 135 (102-169) | 12.13 (9.47-14.76) | 94 (70-119) | 8.44 (6.42-10.44) | 55 (41-73) | 4.97 (3.76-6.31) | 26 (18-35) | 2.33 (1.66-3.08) | 18 (12-26) | 1.67 (1.12-2.35) |
| Somalia | Male | 1294 (1101-1468) | 113 (82-151) | 8.77 (6.53-11.06) | 81 (58-109) | 6.25 (4.63-7.98) | 48 (34-65) | 3.74 (2.67-4.82) | 16 (10-23) | 1.22 (0.79-1.72) | 11 (7-18) | 0.88 (0.56-1.33) |
| South Africa | Female | 6817 (6373-7240) | 345 (218-488) | 5.05 (3.25-7.03) | 273 (178-370) | 4.00 (2.64-5.37) | 204 (139-272) | 2.99 (2.08-3.95) | 115 (69-161) | 1.70 (1.04-2.35) | 107 (65-151) | 1.58 (0.97-2.21) |
| South Africa | Male | 6435 (6031-6860) | 269 (162-390) | 4.19 (2.51-5.98) | 209 (127-300) | 3.26 (1.97-4.69) | 158 (97-226) | 2.48 (1.52-3.46) | 58 (29-92) | 0.89 (0.45-1.42) | 53 (27-86) | 0.82 (0.42-1.34) |
| South Korea | Female | 5103 (4516-5687) | 937 (713-1178) | 18.46 (14.13-22.28) | 415 (297-557) | 8.16 (5.95-10.67) | 620 (459-813) | 12.14 (9.06-15.43) | 227 (159-308) | 4.44 (3.19-5.87) | 227 (158-308) | 4.44 (3.18-5.89) |
| South Korea | Male | 4715 (4262-5200) | 835 (612-1027) | 17.82 (13.06-21.15) | 377 (273-498) | 8.03 (5.77-10.41) | 561 (400-700) | 11.93 (8.72-14.66) | 183 (130-247) | 3.91 (2.78-5.14) | 182 (130-246) | 3.89 (2.76-5.13) |
| Spain | Female | 8826 (7807-9769) | 141 (23-386) | 1.60 (0.25-4.30) | 116 (16-288) | 1.35 (0.18-3.22) | 133 (19-345) | 1.49 (0.21-3.88) | 60 (6-154) | 0.68 (0.06-1.74) | 54 (5-134) | 0.62 (0.06-1.56) |
| Spain | Male | 5977 (5438-6524) | 116 (50-258) | 1.94 (0.84-4.28) | 81 (27-185) | 1.37 (0.45-3.05) | 94 (35-220) | 1.59 (0.58-3.62) | 36 (9-88) | 0.60 (0.16-1.48) | 34 (8-82) | 0.57 (0.14-1.39) |
| Sri Lanka | Female | 2311 (2056-2573) | 357 (286-437) | 15.57 (12.75-18.29) | 234 (185-292) | 10.21 (8.26-12.06) | 153 (118-194) | 6.67 (5.22-8.12) | 84 (63-111) | 3.63 (2.74-4.60) | 73 (53-96) | 3.16 (2.34-4.04) |
| Sri Lanka | Male | 2852 (2521-3206) | 487 (391-591) | 17.06 (14.32-19.87) | 321 (254-405) | 11.26 (9.26-13.44) | 210 (164-268) | 7.35 (5.90-9.09) | 76 (56-103) | 2.68 (2.05-3.56) | 63 (45-89) | 2.21 (1.65-3.01) |
| Sudan | Female | 2389 (2141-2622) | 12 (3-30) | 0.49 (0.14-1.30) | 11 (3-28) | 0.44 (0.13-1.21) | 8 (2-22) | 0.35 (0.09-0.90) | 2 (0-7) | 0.07 (0.01-0.29) | 1 (0-6) | 0.05 (0.00-0.24) |
| Sudan | Male | 2837 (2497-3183) | 126 (75-182) | 4.49 (2.74-6.34) | 101 (63-145) | 3.55 (2.24-4.99) | 64 (41-91) | 2.26 (1.52-3.14) | 16 (8-27) | 0.55 (0.30-0.91) | 12 (6-21) | 0.43 (0.20-0.73) |
| Suriname | Female | 117 (106-127) | 8 (6-11) | 7.20 (5.16-9.06) | 6 (4-8) | 5.02 (3.63-6.41) | 4 (3-5) | 3.40 (2.47-4.35) | 3 (2-4) | 2.43 (1.69-3.20) | 2 (2-3) | 2.06 (1.43-2.80) |
| Suriname | Male | 131 (118-144) | 13 (10-16) | 10.24 (7.98-12.21) | 9 (7-11) | 6.87 (5.22-8.48) | 6 (5-8) | 4.58 (3.50-5.85) | 3 (2-4) | 2.60 (1.89-3.34) | 3 (2-4) | 2.20 (1.58-2.96) |
| Swaziland | Female | 112 (100-124) | 7 (4-9) | 5.97 (3.89-8.16) | 5 (3-7) | 4.55 (3.04-5.92) | 3 (2-5) | 3.11 (2.06-3.99) | 2 (1-3) | 1.65 (1.01-2.31) | 1 (1-2) | 1.27 (0.74-1.82) |
| Swaziland | Male | 149 (134-166) | 9 (6-12) | 5.72 (3.74-7.72) | 6 (4-9) | 4.23 (2.80-5.71) | 4 (3-6) | 2.85 (1.90-3.92) | 1 (1-2) | 0.72 (0.42-1.12) | 1 (0-1) | 0.55 (0.30-0.89) |
| Sweden | Female | 1312 (1169-1450) | 107 (64-149) | 8.22 (5.14-11.11) | 28 (15-44) | 2.12 (1.14-3.35) | 94 (60-129) | 7.16 (4.62-9.58) | 12 (6-21) | 0.94 (0.47-1.56) | 10 (5-17) | 0.77 (0.39-1.31) |
| Sweden | Male | 1200 (1087-1316) | 154 (118-190) | 12.82 (9.88-15.58) | 36 (22-57) | 2.99 (1.82-4.68) | 128 (95-165) | 10.71 (8.03-13.33) | 16 (10-26) | 1.34 (0.81-2.17) | 14 (9-23) | 1.20 (0.71-1.92) |
| Switzerland | Female | 1543 (1385-1697) | 126 (78-179) | 8.16 (5.19-11.26) | 12 (7-27) | 0.76 (0.43-1.77) | 120 (74-169) | 7.79 (5.03-10.71) | 5 (3-12) | 0.31 (0.17-0.79) | 4 (2-10) | 0.26 (0.14-0.67) |
| Switzerland | Male | 1046 (972-1122) | 126 (96-159) | 12.14 (9.21-14.94) | 11 (7-22) | 1.01 (0.64-2.08) | 120 (89-151) | 11.44 (8.61-14.13) | 5 (3-11) | 0.52 (0.32-1.08) | 5 (3-10) | 0.47 (0.28-0.96) |
| Syria | Female | 2302 (2063-2525) | 6 (1-27) | 0.27 (0.03-1.15) | 6 (1-27) | 0.26 (0.02-1.13) | 5 (0-21) | 0.23 (0.02-0.91) | 2 (0-12) | 0.08 (0.00-0.51) | 2 (0-11) | 0.07 (0.00-0.46) |
| Syria | Male | 2423 (2137-2662) | 102 (60-156) | 4.23 (2.46-6.45) | 81 (48-120) | 3.37 (2.02-4.94) | 52 (32-75) | 2.16 (1.35-3.15) | 27 (14-46) | 1.13 (0.59-1.88) | 23 (12-41) | 0.96 (0.50-1.66) |
| Taiwan (Province of China) | Female | 4615 (4200-4994) | 761 (589-933) | 16.46 (13.02-19.94) | 343 (254-442) | 7.39 (5.54-9.40) | 485 (365-614) | 10.52 (8.10-13.09) | 130 (92-177) | 2.82 (2.02-3.78) | 131 (91-176) | 2.82 (2.04-3.76) |
| Taiwan (Province of China) | Male | 4183 (3855-4511) | 647 (525-783) | 15.49 (12.71-18.52) | 292 (228-375) | 7.00 (5.48-8.91) | 416 (328-517) | 9.96 (7.91-12.23) | 112 (86-150) | 2.69 (2.08-3.55) | 110 (85-149) | 2.65 (2.02-3.51) |
| Tajikistan | Female | 67 (60-74) | 5 (4-6) | 7.60 (5.72-9.61) | 4 (3-5) | 5.47 (4.13-6.97) | 2 (2-3) | 3.31 (2.47-4.28) | 1 (1-2) | 1.71 (1.26-2.24) | 1 (1-1) | 1.43 (1.02-1.90) |
| Tajikistan | Male | 66 (57-75) | 8 (7-10) | 12.93 (10.35-15.21) | 6 (5-7) | 8.92 (7.00-10.56) | 3 (3-4) | 5.34 (4.13-6.44) | 1 (1-2) | 2.20 (1.67-2.87) | 1 (1-1) | 1.37 (0.98-1.85) |
| Tanzania | Female | 3954 (3591-4326) | 565 (451-688) | 14.28 (11.66-16.75) | 385 (297-469) | 9.75 (7.65-11.53) | 231 (177-286) | 5.82 (4.62-7.05) | 90 (66-118) | 2.28 (1.69-2.88) | 40 (26-55) | 1.00 (0.67-1.39) |
| Tanzania | Male | 4073 (3683-4450) | 552 (405-676) | 13.56 (10.43-16.34) | 375 (279-468) | 9.25 (6.96-11.26) | 228 (163-288) | 5.58 (4.08-6.95) | 58 (39-79) | 1.41 (0.98-1.91) | 37 (24-52) | 0.90 (0.59-1.27) |
| Thailand | Female | 17226 (15545-18850) | 2445 (1868-3048) | 14.21 (11.04-17.10) | 1576 (1184-2005) | 9.17 (7.01-11.12) | 1114 (840-1445) | 6.48 (4.92-8.20) | 567 (418-736) | 3.29 (2.49-4.11) | 515 (373-670) | 2.99 (2.20-3.83) |
| Thailand | Male | 14280 (12873-15696) | 2310 (1809-2812) | 16.19 (12.92-19.21) | 1488 (1148-1862) | 10.45 (8.19-12.65) | 1064 (801-1322) | 7.44 (5.72-9.12) | 437 (322-581) | 3.05 (2.31-3.99) | 389 (286-526) | 2.72 (2.02-3.62) |
| The Bahamas | Female | 68 (63-73) | 5 (4-6) | 7.30 (5.45-9.14) | 3 (2-3) | 3.87 (2.90-4.93) | 3 (2-4) | 4.77 (3.62-5.95) | 1 (1-2) | 1.70 (1.21-2.23) | 1 (1-1) | 1.50 (1.05-2.00) |
| The Bahamas | Male | 73 (67-79) | 8 (6-9) | 10.65 (8.48-12.51) | 4 (3-5) | 5.28 (4.19-6.58) | 5 (4-6) | 6.61 (5.19-8.14) | 1 (1-2) | 2.04 (1.50-2.61) | 1 (1-2) | 1.67 (1.24-2.22) |
| The Gambia | Female | 141 (127-154) | 12 (9-15) | 8.42 (6.34-10.43) | 8 (6-11) | 6.01 (4.61-7.43) | 5 (4-6) | 3.58 (2.78-4.53) | 3 (2-4) | 1.90 (1.35-2.56) | 2 (1-3) | 1.25 (0.83-1.79) |
| The Gambia | Male | 222 (202-242) | 19 (14-24) | 8.67 (6.56-10.50) | 14 (10-17) | 6.13 (4.62-7.68) | 8 (6-11) | 3.65 (2.75-4.67) | 3 (2-4) | 1.31 (0.90-1.85) | 2 (1-3) | 0.93 (0.60-1.36) |
| Timor-Leste | Female | 107 (94-120) | 15 (12-19) | 14.33 (11.45-17.18) | 10 (8-13) | 9.54 (7.57-11.62) | 6 (5-8) | 6.05 (4.67-7.52) | 2 (1-3) | 1.82 (1.35-2.48) | 1 (1-2) | 1.23 (0.86-1.79) |
| Timor-Leste | Male | 117 (100-135) | 20 (16-25) | 16.99 (14.20-19.95) | 13 (10-17) | 11.32 (9.28-13.80) | 8 (6-11) | 7.18 (5.76-9.06) | 3 (2-4) | 2.22 (1.64-2.99) | 2 (1-2) | 1.46 (1.02-2.04) |
| Togo | Female | 459 (417-505) | 36 (27-46) | 7.94 (5.95-9.82) | 26 (19-33) | 5.64 (4.30-7.08) | 16 (11-20) | 3.40 (2.53-4.33) | 7 (5-10) | 1.49 (1.05-2.07) | 4 (2-6) | 0.83 (0.52-1.22) |
| Togo | Male | 579 (526-636) | 48 (37-60) | 8.34 (6.41-10.14) | 34 (25-43) | 5.90 (4.46-7.36) | 20 (15-27) | 3.53 (2.66-4.56) | 6 (4-8) | 0.97 (0.65-1.38) | 3 (2-4) | 0.48 (0.30-0.73) |
| Tonga | Female | 15 (13-16) | 2 (2-2) | 13.33 (10.96-15.80) | 1 (1-2) | 8.87 (7.21-10.82) | 1 (1-1) | 5.86 (4.68-7.22) | 0 (0-1) | 3.09 (2.36-4.06) | 0 (0-1) | 2.59 (1.94-3.46) |
| Tonga | Male | 1 (1-1) | 0 (0-0) | 14.86 (12.21-17.69) | 0 (0-0) | 9.79 (7.90-11.96) | 0 (0-0) | 6.48 (5.19-8.10) | 0 (0-0) | 2.14 (1.58-2.82) | 0 (0-0) | 1.79 (1.30-2.54) |
| Trinidad and Tobago | Female | 316 (289-340) | 23 (17-30) | 7.33 (5.38-9.33) | 14 (11-19) | 4.55 (3.39-5.82) | 13 (9-17) | 4.14 (3.04-5.26) | 7 (5-9) | 2.08 (1.48-2.76) | 5 (4-7) | 1.68 (1.16-2.33) |
| Trinidad and Tobago | Male | 432 (391-473) | 50 (40-61) | 11.59 (9.58-13.46) | 30 (23-37) | 6.84 (5.51-8.35) | 26 (21-33) | 6.13 (4.88-7.53) | 11 (9-15) | 2.64 (2.06-3.47) | 10 (7-13) | 2.28 (1.72-3.00) |
| Tunisia | Female | 1680 (1491-1851) | 3 (0-16) | 0.21 (0.03-0.93) | 3 (0-15) | 0.19 (0.02-0.89) | 3 (0-13) | 0.18 (0.02-0.74) | 1 (0-6) | 0.06 (0.00-0.37) | 1 (0-5) | 0.04 (0.00-0.31) |
| Tunisia | Male | 2063 (1799-2307) | 75 (39-121) | 3.64 (1.89-5.76) | 59 (31-96) | 2.91 (1.55-4.54) | 42 (23-66) | 2.05 (1.10-3.14) | 16 (7-29) | 0.81 (0.35-1.38) | 14 (6-26) | 0.70 (0.30-1.24) |
| Turkey | Female | 12871 (11563-14215) | 0 (0-9) | 0.00 (0.00-0.07) | 0 (0-9) | 0.00 (0.00-0.07) | 0 (0-9) | 0.00 (0.00-0.07) | 0 (0-4) | 0.00 (0.00-0.03) | 0 (0-3) | 0.00 (0.00-0.02) |
| Turkey | Male | 11711 (10515-12891) | 210 (75-421) | 1.78 (0.64-3.53) | 183 (63-359) | 1.57 (0.55-3.02) | 157 (53-294) | 1.34 (0.43-2.47) | 57 (13-126) | 0.49 (0.11-1.07) | 55 (13-125) | 0.47 (0.11-1.06) |
| Turkmenistan | Female | 285 (265-307) | 24 (19-29) | 8.41 (6.64-10.27) | 16 (12-20) | 5.63 (4.36-6.97) | 12 (9-15) | 4.12 (3.21-5.07) | 6 (5-8) | 2.28 (1.70-2.90) | 6 (4-7) | 1.97 (1.47-2.54) |
| Turkmenistan | Male | 393 (362-424) | 51 (41-60) | 12.93 (10.67-14.99) | 33 (26-39) | 8.29 (6.64-9.82) | 23 (19-29) | 5.96 (4.76-7.29) | 9 (7-11) | 2.28 (1.77-2.81) | 7 (5-8) | 1.67 (1.27-2.11) |
| Tuvalu | Female | 2 (2-2) | 0 (0-0) | 12.57 (9.98-15.15) | 0 (0-0) | 8.28 (6.43-10.28) | 0 (0-0) | 5.64 (4.29-7.01) | 0 (0-0) | 2.13 (1.47-2.93) | 0 (0-0) | 1.26 (0.82-1.79) |
| Tuvalu | Male | 2 (1-2) | 0 (0-0) | 13.77 (10.98-16.45) | 0 (0-0) | 9.05 (7.13-11.26) | 0 (0-0) | 6.16 (4.87-7.68) | 0 (0-0) | 1.93 (1.39-2.58) | 0 (0-0) | 1.36 (0.90-1.93) |
| Uganda | Female | 2105 (1904-2301) | 275 (223-332) | 13.08 (10.69-15.29) | 190 (152-232) | 9.04 (7.34-10.84) | 113 (88-141) | 5.36 (4.23-6.50) | 37 (27-49) | 1.75 (1.30-2.32) | 23 (15-32) | 1.07 (0.71-1.50) |
| Uganda | Male | 2571 (2319-2800) | 256 (201-316) | 10.00 (7.96-12.21) | 179 (141-228) | 7.02 (5.50-8.75) | 109 (83-138) | 4.24 (3.22-5.32) | 25 (17-35) | 0.96 (0.67-1.37) | 15 (9-22) | 0.57 (0.36-0.87) |
| Ukraine | Female | 793 (725-871) | 26 (14-39) | 3.22 (1.81-4.96) | 22 (13-33) | 2.78 (1.59-4.17) | 17 (10-24) | 2.09 (1.24-3.00) | 10 (5-16) | 1.20 (0.63-1.95) | 8 (4-13) | 1.01 (0.52-1.67) |
| Ukraine | Male | 783 (709-853) | 75 (60-93) | 9.69 (7.74-11.46) | 51 (40-64) | 6.53 (5.24-7.98) | 34 (26-43) | 4.34 (3.46-5.34) | 15 (11-20) | 1.90 (1.44-2.44) | 10 (7-14) | 1.32 (0.97-1.72) |
| United Arab Emirates | Female | 292 (248-335) | 1 (0-6) | 0.35 (0.04-1.98) | 1 (0-4) | 0.26 (0.02-1.36) | 1 (0-6) | 0.32 (0.04-1.93) | 0 (0-2) | 0.06 (0.00-0.51) | 0 (0-1) | 0.05 (0.00-0.46) |
| United Arab Emirates | Male | 368 (329-408) | 18 (11-27) | 5.02 (3.00-7.13) | 8 (5-11) | 2.21 (1.42-3.09) | 15 (9-21) | 4.04 (2.53-5.64) | 2 (1-3) | 0.50 (0.29-0.74) | 2 (1-3) | 0.45 (0.27-0.69) |
| United Kingdom | Female | 5362 (4970-5800) | 406 (256-561) | 7.56 (4.80-10.46) | 157 (98-228) | 2.94 (1.84-4.19) | 321 (208-449) | 6.02 (3.87-8.27) | 65 (39-95) | 1.21 (0.72-1.75) | 59 (36-87) | 1.11 (0.67-1.62) |
| United Kingdom | Male | 4274 (3977-4595) | 470 (345-591) | 11.05 (8.16-13.61) | 166 (113-236) | 3.89 (2.64-5.41) | 352 (253-456) | 8.23 (6.10-10.51) | 70 (46-100) | 1.65 (1.09-2.33) | 65 (42-94) | 1.52 (1.01-2.15) |
| United States | Female | 67739 (63979-71490) | 5640 (4136-7135) | 8.33 (6.13-10.49) | 1090 (637-1767) | 1.61 (0.94-2.56) | 5036 (3779-6509) | 7.46 (5.59-9.56) | 538 (302-891) | 0.79 (0.45-1.30) | 503 (282-849) | 0.74 (0.42-1.23) |
| United States | Male | 67605 (64595-70556) | 7905 (6337-9505) | 11.69 (9.41-13.90) | 1455 (842-2168) | 2.16 (1.27-3.22) | 6926 (5424-8394) | 10.24 (8.08-12.34) | 675 (377-1033) | 1.00 (0.55-1.51) | 640 (354-985) | 0.95 (0.52-1.45) |
| Uruguay | Female | 595 (542-646) | 80 (65-96) | 13.44 (10.98-15.73) | 46 (36-57) | 7.68 (6.13-9.42) | 43 (33-55) | 7.29 (5.62-8.99) | 26 (20-34) | 4.44 (3.43-5.56) | 25 (19-33) | 4.26 (3.28-5.42) |
| Uruguay | Male | 571 (526-614) | 86 (69-105) | 15.17 (12.35-18.03) | 49 (39-61) | 8.59 (6.83-10.42) | 46 (36-57) | 8.14 (6.48-9.96) | 25 (19-32) | 4.44 (3.36-5.50) | 23 (17-29) | 4.00 (3.04-5.05) |
| Uzbekistan | Female | 1576 (1473-1686) | 136 (105-166) | 8.63 (6.76-10.40) | 97 (74-119) | 6.14 (4.77-7.45) | 60 (46-76) | 3.83 (2.97-4.75) | 36 (26-46) | 2.29 (1.68-2.91) | 31 (22-41) | 1.97 (1.44-2.56) |
| Uzbekistan | Male | 1791 (1663-1919) | 235 (193-276) | 13.13 (10.86-15.27) | 160 (128-191) | 8.93 (7.22-10.63) | 98 (78-122) | 5.50 (4.39-6.73) | 43 (32-54) | 2.38 (1.81-3.00) | 33 (25-43) | 1.87 (1.40-2.39) |
| Vanuatu | Female | 26 (22-29) | 3 (2-4) | 11.39 (9.13-13.64) | 2 (1-2) | 7.72 (6.13-9.50) | 1 (1-2) | 4.94 (3.83-6.17) | 0 (0-1) | 1.87 (1.34-2.60) | 0 (0-0) | 0.89 (0.54-1.39) |
| Vanuatu | Male | 27 (23-32) | 3 (3-4) | 12.56 (10.10-15.22) | 2 (2-3) | 8.45 (6.74-10.58) | 1 (1-2) | 5.42 (4.26-7.04) | 0 (0-1) | 1.28 (0.89-1.84) | 0 (0-0) | 0.69 (0.42-1.04) |
| Venezuela | Female | 5482 (5045-5926) | 637 (509-765) | 11.61 (9.39-13.50) | 438 (345-545) | 7.98 (6.36-9.72) | 270 (209-342) | 4.92 (3.81-6.02) | 231 (178-292) | 4.24 (3.28-5.21) | 218 (168-278) | 3.99 (3.10-4.94) |
| Venezuela | Male | 6118 (5597-6650) | 946 (768-1136) | 15.51 (12.66-18.20) | 633 (507-775) | 10.38 (8.39-12.43) | 392 (306-494) | 6.39 (5.06-7.94) | 266 (205-332) | 4.36 (3.45-5.42) | 246 (190-312) | 4.03 (3.18-5.03) |
| Vietnam | Female | 12885 (11637-14093) | 1891 (1529-2270) | 14.75 (12.06-17.30) | 1252 (993-1536) | 9.73 (7.87-11.61) | 814 (633-1018) | 6.32 (5.00-7.70) | 373 (285-490) | 2.90 (2.24-3.71) | 259 (189-343) | 2.01 (1.49-2.62) |
| Vietnam | Male | 10403 (9302-11561) | 1772 (1430-2129) | 17.04 (14.27-19.73) | 1173 (932-1469) | 11.24 (9.24-13.41) | 760 (596-953) | 7.29 (5.90-8.97) | 316 (236-417) | 3.03 (2.36-3.89) | 230 (168-316) | 2.20 (1.67-2.93) |
| Yemen | Female | 1083 (948-1211) | 3 (0-12) | 0.31 (0.04-1.07) | 3 (0-11) | 0.29 (0.03-1.05) | 3 (0-9) | 0.25 (0.02-0.83) | 0 (0-3) | 0.04 (0.00-0.24) | 0 (0-2) | 0.04 (0.00-0.21) |
| Yemen | Male | 1268 (1085-1461) | 54 (30-82) | 4.29 (2.44-6.21) | 43 (25-64) | 3.40 (2.03-4.90) | 27 (16-41) | 2.15 (1.37-3.06) | 9 (4-15) | 0.70 (0.36-1.17) | 8 (4-13) | 0.61 (0.29-1.01) |
| Zambia | Female | 1230 (1114-1343) | 149 (120-185) | 12.21 (9.85-14.67) | 103 (83-129) | 8.44 (6.67-10.27) | 62 (47-79) | 5.05 (3.90-6.29) | 31 (23-41) | 2.53 (1.91-3.22) | 19 (14-26) | 1.55 (1.11-2.08) |
| Zambia | Male | 1842 (1631-2036) | 169 (127-213) | 9.18 (7.02-11.34) | 120 (88-153) | 6.55 (4.82-8.23) | 73 (52-94) | 3.97 (2.94-5.08) | 23 (16-31) | 1.24 (0.86-1.66) | 14 (9-20) | 0.76 (0.49-1.10) |
| Zimbabwe | Female | 1186 (1070-1305) | 100 (75-129) | 8.42 (6.36-10.42) | 71 (52-91) | 5.96 (4.54-7.38) | 44 (32-56) | 3.67 (2.79-4.58) | 23 (15-32) | 1.94 (1.32-2.65) | 19 (12-28) | 1.59 (1.03-2.25) |
| Zimbabwe | Male | 1159 (1049-1277) | 106 (80-133) | 9.21 (7.03-11.08) | 74 (55-95) | 6.41 (4.88-7.93) | 45 (33-59) | 3.88 (2.97-5.01) | 16 (10-23) | 1.34 (0.91-1.92) | 12 (8-18) | 1.04 (0.69-1.58) |

#### f. **Prevented CKD new cases**

| **Country** | **Sex** | **Baseline disease burden Median (95% UI)** | **Replacement of discretionary salt and non-discretionary salt** | | **Replacement of discretionary salt** | | **Replacement of non-discretionary salt** | | **Replacement of discretionary salt among those with diagnosed hypertension** | | **Replacement of discretionary salt among those with treated hypertension** | |
| --- | --- | --- | --- | --- | --- | --- | --- | --- | --- | --- | --- | --- |
|  |  |  | **No. of events averted Median (95% UI)** | **Percentage of national disease burden averted % (95% UI)** | **No. of events averted Median (95% UI)** | **Percentage of national disease burden averted % (95% UI)** | **No. of events averted Median (95% UI)** | **Percentage of national disease burden averted % (95% UI)** | **No. of events averted Median (95% UI)** | **Percentage of national disease burden averted % (95% UI)** | **Replacement of discretionary salt among those with treated hypertension** | **Percentage of national disease burden averted % (95% UI)** |
| **Global, millions** | **Female** | 10.64 (10.38-10.92) | 1.09 (1.00-1.18) | 10.20 (9.38-11.00) | 0.61 (0.55-0.67) | 5.75 (5.24-6.30) | 0.61 (0.55-0.67) | 5.73 (5.21-6.30) | 0.20 (0.19-0.22) | 1.91 (1.75-2.11) | 0.17 (0.16-0.19) | 1.64 (1.49-1.81) |
| **Global, millions** | **Male** | 8.32 (8.12-8.51) | 1.09 (1.02-1.18) | 13.12 (12.24-14.08) | 0.61 (0.56-0.67) | 7.38 (6.76-8.03) | 0.62 (0.57-0.68) | 7.48 (6.86-8.18) | 0.18 (0.16-0.20) | 2.13 (1.96-2.34) | 0.15 (0.14-0.16) | 1.78 (1.64-1.97) |
| Afghanistan | Female | 21989 (20068-24187) | 104 (17-282) | 0.47 (0.08-1.30) | 95 (14-261) | 0.43 (0.07-1.19) | 73 (11-207) | 0.33 (0.05-0.94) | 22 (2-81) | 0.10 (0.01-0.37) | 18 (2-71) | 0.08 (0.01-0.32) |
| Afghanistan | Male | 14616 (13282-15942) | 690 (422-1022) | 4.75 (2.89-6.96) | 536 (328-780) | 3.69 (2.25-5.31) | 330 (209-484) | 2.27 (1.41-3.27) | 95 (52-150) | 0.65 (0.37-1.01) | 76 (43-122) | 0.52 (0.30-0.84) |
| Albania | Female | 4333 (3897-4781) | 697 (551-862) | 16.11 (12.99-19.33) | 448 (344-569) | 10.35 (8.18-12.79) | 313 (236-407) | 7.25 (5.57-9.26) | 190 (139-255) | 4.39 (3.28-5.74) | 166 (117-226) | 3.84 (2.82-5.15) |
| Albania | Male | 4041 (3614-4460) | 847 (679-1028) | 21.04 (17.07-24.59) | 566 (440-715) | 14.07 (11.02-17.39) | 397 (300-518) | 9.89 (7.63-12.52) | 133 (93-186) | 3.30 (2.38-4.47) | 98 (66-143) | 2.43 (1.65-3.47) |
| Algeria | Female | 83478 (75205-92322) | 327 (27-1213) | 0.40 (0.03-1.43) | 305 (23-1114) | 0.37 (0.03-1.32) | 272 (16-943) | 0.32 (0.02-1.10) | 83 (1-441) | 0.10 (0.00-0.52) | 72 (0-382) | 0.09 (0.00-0.46) |
| Algeria | Male | 61789 (56421-67793) | 3005 (1749-4340) | 4.87 (2.87-6.98) | 2306 (1417-3289) | 3.74 (2.32-5.21) | 1556 (1016-2238) | 2.51 (1.66-3.55) | 559 (291-892) | 0.91 (0.47-1.40) | 458 (234-756) | 0.74 (0.38-1.19) |
| American Samoa | Female | 70 (62-78) | 4 (2-5) | 5.27 (3.34-7.34) | 2 (2-3) | 3.55 (2.25-4.80) | 2 (1-3) | 3.28 (2.11-4.44) | 1 (0-1) | 1.15 (0.72-1.63) | 1 (0-1) | 0.87 (0.51-1.33) |
| American Samoa | Male | 74 (67-83) | 5 (4-7) | 7.00 (4.87-9.28) | 3 (2-4) | 4.38 (3.07-5.77) | 3 (2-4) | 4.04 (2.85-5.33) | 1 (1-1) | 1.36 (0.85-1.92) | 1 (0-1) | 1.00 (0.62-1.46) |
| Andorra | Female | 190 (172-211) | 7 (3-11) | 3.62 (1.83-5.81) | 4 (2-6) | 2.07 (1.09-3.20) | 6 (3-10) | 3.16 (1.63-5.03) | 2 (1-3) | 0.84 (0.39-1.43) | 1 (1-2) | 0.77 (0.36-1.33) |
| Andorra | Male | 145 (131-159) | 14 (10-18) | 9.77 (7.16-12.44) | 6 (4-8) | 3.87 (2.80-5.30) | 10 (7-13) | 7.04 (5.19-9.16) | 2 (2-4) | 1.70 (1.13-2.47) | 2 (2-3) | 1.58 (1.04-2.29) |
| Angola | Female | 8269 (7395-9166) | 448 (295-626) | 5.40 (3.56-7.39) | 340 (231-476) | 4.15 (2.77-5.60) | 219 (148-302) | 2.66 (1.81-3.59) | 94 (57-138) | 1.15 (0.70-1.67) | 44 (24-70) | 0.54 (0.29-0.84) |
| Angola | Male | 6220 (5615-6821) | 378 (251-512) | 6.08 (4.01-8.13) | 279 (186-382) | 4.50 (3.06-6.11) | 177 (119-240) | 2.86 (1.93-3.83) | 54 (31-83) | 0.87 (0.51-1.33) | 32 (17-55) | 0.52 (0.28-0.87) |
| Antigua and Barbuda | Female | 194 (170-215) | 12 (8-17) | 6.28 (4.22-8.47) | 8 (5-10) | 3.95 (2.73-5.22) | 7 (5-10) | 3.70 (2.54-4.99) | 3 (2-5) | 1.71 (1.16-2.37) | 3 (2-4) | 1.44 (0.93-2.06) |
| Antigua and Barbuda | Male | 168 (150-184) | 17 (12-21) | 9.94 (7.50-12.31) | 10 (7-13) | 5.79 (4.15-7.45) | 9 (6-12) | 5.34 (3.99-6.88) | 3 (2-4) | 1.85 (1.29-2.56) | 2 (2-4) | 1.48 (0.98-2.11) |
| Argentina | Female | 85206 (77386-93887) | 9819 (7587-12028) | 11.55 (9.25-13.75) | 6119 (4664-7843) | 7.19 (5.65-8.93) | 4782 (3626-6285) | 5.62 (4.36-7.26) | 2909 (2163-3841) | 3.43 (2.62-4.35) | 2549 (1893-3393) | 3.00 (2.28-3.90) |
| Argentina | Male | 62807 (56152-69650) | 8568 (6660-10713) | 13.67 (10.84-16.53) | 5327 (4100-6791) | 8.50 (6.60-10.65) | 4164 (3111-5476) | 6.66 (5.14-8.46) | 2454 (1823-3222) | 3.94 (2.98-5.07) | 2138 (1587-2808) | 3.43 (2.57-4.39) |
| Armenia | Female | 4018 (3562-4470) | 303 (210-408) | 7.61 (5.29-10.00) | 215 (149-285) | 5.37 (3.77-7.06) | 146 (99-197) | 3.66 (2.53-4.83) | 74 (48-102) | 1.83 (1.19-2.54) | 61 (40-89) | 1.51 (1.02-2.16) |
| Armenia | Male | 3180 (2877-3489) | 404 (318-504) | 12.71 (10.13-15.68) | 267 (206-340) | 8.41 (6.55-10.57) | 178 (135-231) | 5.59 (4.32-7.18) | 70 (50-96) | 2.21 (1.63-2.99) | 47 (34-67) | 1.47 (1.06-2.03) |
| Australia | Female | 74851 (66415-82484) | 1956 (878-3701) | 2.65 (1.18-4.91) | 808 (324-1491) | 1.08 (0.45-1.99) | 1873 (804-3452) | 2.50 (1.08-4.63) | 293 (97-564) | 0.39 (0.13-0.76) | 257 (82-504) | 0.34 (0.11-0.67) |
| Australia | Male | 59291 (53441-65166) | 3277 (1794-5073) | 5.52 (3.07-8.32) | 978 (522-1668) | 1.67 (0.88-2.76) | 2919 (1653-4451) | 4.94 (2.81-7.35) | 373 (178-667) | 0.63 (0.30-1.12) | 345 (166-616) | 0.59 (0.28-1.01) |
| Austria | Female | 30183 (26578-33882) | 3454 (2550-4403) | 11.47 (8.90-14.05) | 1026 (634-1548) | 3.40 (2.15-4.94) | 2722 (1979-3550) | 9.07 (6.90-11.50) | 463 (269-730) | 1.52 (0.91-2.33) | 407 (239-658) | 1.35 (0.79-2.18) |
| Austria | Male | 20170 (17893-22184) | 3052 (2373-3793) | 15.18 (11.98-18.33) | 890 (617-1270) | 4.43 (3.12-6.33) | 2371 (1823-3014) | 11.85 (9.20-14.67) | 405 (262-639) | 2.01 (1.32-3.11) | 395 (256-609) | 1.95 (1.27-2.96) |
| Azerbaijan | Female | 11400 (10117-12622) | 823 (557-1082) | 7.26 (4.92-9.33) | 575 (389-758) | 5.07 (3.53-6.65) | 393 (268-539) | 3.47 (2.35-4.65) | 207 (133-283) | 1.82 (1.20-2.47) | 168 (106-237) | 1.48 (0.96-2.05) |
| Azerbaijan | Male | 8750 (7901-9784) | 1060 (810-1353) | 12.11 (9.48-14.78) | 697 (517-909) | 8.00 (5.94-10.08) | 472 (353-628) | 5.38 (4.09-6.90) | 178 (125-247) | 2.05 (1.47-2.72) | 128 (89-176) | 1.46 (1.03-1.98) |
| Bahrain | Female | 2093 (1845-2330) | 8 (1-32) | 0.40 (0.04-1.52) | 7 (1-27) | 0.35 (0.03-1.31) | 8 (1-29) | 0.37 (0.03-1.41) | 2 (0-11) | 0.10 (0.00-0.51) | 2 (0-9) | 0.08 (0.00-0.43) |
| Bahrain | Male | 2632 (2343-2932) | 136 (80-201) | 5.23 (3.10-7.52) | 80 (50-112) | 3.06 (1.89-4.21) | 94 (56-137) | 3.58 (2.20-5.10) | 20 (11-32) | 0.76 (0.41-1.16) | 17 (9-27) | 0.64 (0.33-1.03) |
| Bangladesh | Female | 85076 (76618-93861) | 9109 (7102-11081) | 10.65 (8.48-12.69) | 6247 (4775-7823) | 7.35 (5.69-9.15) | 3900 (2947-5053) | 4.58 (3.48-5.77) | 1473 (1093-1918) | 1.73 (1.31-2.23) | 1179 (863-1554) | 1.39 (1.02-1.78) |
| Bangladesh | Male | 88429 (80788-96900) | 9566 (7421-11670) | 10.80 (8.50-12.95) | 6605 (5009-8310) | 7.46 (5.75-9.14) | 4093 (3037-5265) | 4.63 (3.45-5.87) | 1016 (729-1345) | 1.14 (0.84-1.49) | 790 (565-1043) | 0.89 (0.65-1.16) |
| Barbados | Female | 816 (721-907) | 35 (20-54) | 4.37 (2.50-6.58) | 26 (15-37) | 3.22 (1.89-4.55) | 25 (14-36) | 3.03 (1.80-4.36) | 13 (7-20) | 1.62 (0.90-2.38) | 12 (7-18) | 1.49 (0.81-2.18) |
| Barbados | Male | 622 (558-680) | 51 (36-67) | 8.24 (5.75-10.64) | 30 (21-41) | 4.84 (3.39-6.40) | 28 (19-38) | 4.54 (3.14-5.97) | 12 (8-17) | 1.89 (1.30-2.62) | 10 (7-15) | 1.68 (1.12-2.34) |
| Belarus | Female | 11399 (10169-12625) | 372 (199-592) | 3.29 (1.80-5.07) | 318 (175-486) | 2.79 (1.61-4.24) | 252 (145-376) | 2.21 (1.29-3.23) | 160 (81-261) | 1.41 (0.74-2.22) | 129 (63-215) | 1.14 (0.57-1.86) |
| Belarus | Male | 7920 (7102-8672) | 770 (591-973) | 9.81 (7.60-11.93) | 507 (384-653) | 6.46 (4.94-8.08) | 363 (280-469) | 4.61 (3.58-5.80) | 208 (146-279) | 2.65 (1.85-3.44) | 142 (100-194) | 1.79 (1.27-2.40) |
| Belgium | Female | 35040 (30993-38663) | 3791 (2851-4787) | 10.78 (8.31-13.30) | 1185 (770-1715) | 3.40 (2.24-4.85) | 2945 (2174-3899) | 8.42 (6.40-10.73) | 559 (347-835) | 1.61 (0.99-2.35) | 531 (326-798) | 1.52 (0.96-2.25) |
| Belgium | Male | 23193 (20915-25489) | 3192 (2518-4026) | 13.80 (11.12-17.00) | 994 (689-1408) | 4.27 (3.01-6.03) | 2467 (1896-3147) | 10.67 (8.33-13.28) | 444 (296-650) | 1.91 (1.30-2.73) | 438 (296-653) | 1.89 (1.30-2.74) |
| Belize | Female | 491 (442-542) | 27 (17-37) | 5.54 (3.40-7.48) | 19 (12-27) | 3.94 (2.52-5.36) | 14 (9-19) | 2.75 (1.78-3.85) | 7 (4-10) | 1.44 (0.91-2.05) | 6 (4-9) | 1.18 (0.73-1.75) |
| Belize | Male | 454 (415-494) | 41 (31-53) | 9.10 (6.85-11.42) | 28 (20-36) | 6.08 (4.56-7.72) | 19 (14-25) | 4.19 (3.08-5.45) | 7 (5-10) | 1.53 (1.09-2.05) | 5 (4-8) | 1.20 (0.80-1.67) |
| Benin | Female | 5314 (4796-5812) | 523 (406-629) | 9.89 (7.79-11.59) | 365 (275-450) | 6.88 (5.25-8.35) | 219 (161-281) | 4.14 (3.08-5.13) | 77 (54-103) | 1.46 (1.03-1.88) | 52 (37-72) | 0.99 (0.69-1.31) |
| Benin | Male | 3563 (3239-3917) | 291 (209-375) | 8.16 (6.04-10.46) | 206 (148-269) | 5.77 (4.17-7.47) | 125 (90-169) | 3.50 (2.57-4.69) | 26 (17-37) | 0.72 (0.46-1.03) | 18 (11-27) | 0.52 (0.32-0.75) |
| Bermuda | Female | 209 (188-236) | 14 (9-19) | 6.85 (4.60-8.96) | 1 (1-2) | 0.49 (0.33-0.92) | 14 (9-19) | 6.64 (4.44-8.76) | 0 (0-1) | 0.23 (0.15-0.43) | 0 (0-1) | 0.20 (0.12-0.40) |
| Bermuda | Male | 179 (161-196) | 18 (13-23) | 10.19 (7.54-12.57) | 1 (1-2) | 0.68 (0.48-1.19) | 17 (13-22) | 9.77 (7.13-11.99) | 0 (0-1) | 0.23 (0.15-0.42) | 0 (0-1) | 0.19 (0.12-0.38) |
| Bhutan | Female | 559 (504-616) | 54 (40-67) | 9.76 (7.28-11.71) | 37 (27-48) | 6.66 (4.78-8.31) | 24 (17-31) | 4.27 (3.15-5.49) | 10 (7-14) | 1.86 (1.32-2.39) | 8 (5-10) | 1.35 (0.95-1.81) |
| Bhutan | Male | 533 (484-582) | 58 (45-70) | 10.89 (8.49-13.00) | 40 (30-50) | 7.42 (5.69-9.15) | 25 (19-32) | 4.78 (3.59-5.95) | 9 (6-11) | 1.62 (1.20-2.10) | 5 (4-7) | 1.03 (0.76-1.40) |
| Bolivia | Female | 12248 (11094-13421) | 1099 (768-1399) | 9.00 (6.33-11.29) | 754 (513-992) | 6.15 (4.22-7.97) | 479 (320-653) | 3.91 (2.66-5.19) | 269 (172-373) | 2.19 (1.42-3.01) | 236 (150-347) | 1.95 (1.24-2.82) |
| Bolivia | Male | 10753 (9739-11622) | 1241 (877-1604) | 11.56 (8.23-14.50) | 840 (587-1094) | 7.84 (5.56-9.93) | 536 (366-706) | 5.00 (3.54-6.56) | 223 (143-325) | 2.07 (1.36-2.93) | 184 (113-270) | 1.71 (1.07-2.45) |
| Bosnia and Herzegovina | Female | 7526 (6726-8337) | 1240 (981-1530) | 16.57 (13.30-19.87) | 788 (617-988) | 10.48 (8.26-12.95) | 563 (427-727) | 7.53 (5.76-9.45) | 405 (301-527) | 5.41 (4.10-6.92) | 373 (277-488) | 4.96 (3.77-6.41) |
| Bosnia and Herzegovina | Male | 5816 (5280-6397) | 1210 (986-1444) | 20.86 (17.44-24.13) | 801 (630-994) | 13.77 (11.07-16.67) | 574 (448-731) | 9.85 (7.80-12.25) | 328 (244-430) | 5.61 (4.25-7.20) | 302 (222-400) | 5.17 (3.93-6.85) |
| Botswana | Female | 1640 (1473-1792) | 99 (66-135) | 6.08 (3.99-8.12) | 71 (48-95) | 4.38 (2.99-5.76) | 52 (35-69) | 3.18 (2.19-4.20) | 25 (16-35) | 1.52 (0.96-2.11) | 20 (13-30) | 1.23 (0.78-1.79) |
| Botswana | Male | 1608 (1466-1753) | 97 (67-131) | 6.04 (4.23-8.15) | 69 (48-93) | 4.30 (2.92-5.76) | 50 (35-69) | 3.14 (2.19-4.26) | 15 (9-23) | 0.96 (0.55-1.42) | 11 (6-17) | 0.67 (0.36-1.04) |
| Brazil | Female | 356045 (326119-381225) | 36551 (28035-44880) | 10.33 (8.04-12.59) | 23896 (17749-30062) | 6.69 (5.04-8.52) | 17159 (12319-22052) | 4.83 (3.54-6.08) | 11968 (8692-15423) | 3.36 (2.50-4.32) | 11627 (8387-14926) | 3.25 (2.42-4.18) |
| Brazil | Male | 270258 (251791-289251) | 35012 (27657-42756) | 12.92 (10.29-15.60) | 22581 (17460-27863) | 8.36 (6.58-10.22) | 16234 (12588-20306) | 5.98 (4.67-7.46) | 9308 (7062-11712) | 3.45 (2.67-4.28) | 8702 (6647-10977) | 3.22 (2.50-4.04) |
| Brunei | Female | 543 (480-600) | 75 (56-95) | 13.91 (10.67-17.26) | 35 (25-46) | 6.44 (4.70-8.27) | 47 (34-61) | 8.68 (6.56-11.12) | 16 (11-21) | 2.87 (2.02-3.78) | 15 (10-19) | 2.71 (1.89-3.56) |
| Brunei | Male | 540 (482-594) | 90 (70-109) | 16.67 (13.13-19.82) | 41 (32-54) | 7.65 (5.86-9.70) | 56 (43-71) | 10.52 (8.07-12.83) | 17 (12-23) | 3.08 (2.29-4.12) | 15 (11-21) | 2.85 (2.08-3.78) |
| Bulgaria | Female | 15743 (14016-17495) | 2592 (2001-3213) | 16.50 (13.16-19.66) | 1561 (1181-1994) | 9.94 (7.72-12.36) | 1277 (959-1637) | 8.12 (6.28-10.10) | 797 (588-1088) | 5.05 (3.82-6.79) | 713 (505-1002) | 4.51 (3.32-6.18) |
| Bulgaria | Male | 13380 (11996-14753) | 2787 (2244-3370) | 20.89 (16.87-24.36) | 1743 (1338-2203) | 13.07 (10.05-15.97) | 1423 (1091-1826) | 10.69 (8.31-13.38) | 755 (531-1008) | 5.66 (3.97-7.42) | 636 (449-914) | 4.82 (3.40-6.68) |
| Burkina Faso | Female | 8940 (8130-9784) | 635 (439-832) | 7.10 (5.00-9.13) | 452 (313-599) | 5.06 (3.58-6.70) | 272 (186-370) | 3.06 (2.08-4.06) | 92 (60-134) | 1.04 (0.68-1.46) | 62 (37-93) | 0.69 (0.43-1.04) |
| Burkina Faso | Male | 5680 (5131-6208) | 457 (322-602) | 8.05 (5.68-10.34) | 321 (226-435) | 5.68 (3.99-7.56) | 192 (135-265) | 3.38 (2.38-4.64) | 48 (30-75) | 0.84 (0.54-1.31) | 25 (14-42) | 0.44 (0.26-0.76) |
| Burundi | Female | 2472 (2222-2724) | 297 (231-366) | 11.97 (9.67-14.59) | 206 (158-259) | 8.35 (6.64-10.46) | 120 (92-156) | 4.89 (3.83-6.10) | 51 (36-72) | 2.08 (1.45-2.88) | 35 (22-51) | 1.42 (0.91-2.03) |
| Burundi | Male | 2561 (2342-2819) | 246 (178-316) | 9.56 (7.05-12.08) | 175 (125-228) | 6.79 (4.89-8.63) | 102 (73-138) | 4.00 (2.87-5.25) | 32 (20-49) | 1.26 (0.79-1.87) | 22 (12-35) | 0.87 (0.47-1.36) |
| Cambodia | Female | 14182 (12733-15595) | 1614 (1027-2126) | 11.44 (7.49-14.80) | 1102 (704-1476) | 7.77 (5.11-10.24) | 672 (436-936) | 4.75 (3.08-6.43) | 273 (177-392) | 1.94 (1.29-2.75) | 215 (136-317) | 1.52 (0.97-2.22) |
| Cambodia | Male | 7924 (7153-8689) | 1018 (668-1326) | 12.87 (8.46-16.39) | 700 (449-929) | 8.83 (5.85-11.65) | 432 (277-579) | 5.44 (3.51-7.22) | 121 (74-178) | 1.53 (0.97-2.18) | 87 (51-128) | 1.10 (0.63-1.62) |
| Cameroon | Female | 17478 (15879-19259) | 1322 (938-1702) | 7.54 (5.32-9.53) | 935 (675-1233) | 5.36 (3.88-6.92) | 573 (410-761) | 3.27 (2.38-4.27) | 204 (139-289) | 1.17 (0.79-1.65) | 114 (72-172) | 0.66 (0.41-0.96) |
| Cameroon | Male | 12275 (10992-13446) | 1048 (763-1363) | 8.57 (6.38-10.92) | 736 (534-980) | 6.00 (4.39-7.86) | 448 (329-597) | 3.66 (2.69-4.81) | 119 (78-177) | 0.97 (0.64-1.42) | 73 (44-111) | 0.59 (0.36-0.90) |
| Canada | Female | 100955 (89213-110976) | 8875 (6352-11718) | 8.85 (6.44-11.16) | 2798 (1758-4126) | 2.78 (1.77-3.98) | 7038 (5006-9466) | 6.99 (5.04-9.14) | 1060 (645-1624) | 1.06 (0.64-1.56) | 1029 (633-1579) | 1.03 (0.64-1.52) |
| Canada | Male | 81853 (72711-90537) | 9856 (7158-12593) | 12.08 (8.77-15.16) | 3050 (2006-4424) | 3.72 (2.46-5.35) | 7736 (5422-10013) | 9.45 (6.62-12.10) | 1361 (872-2043) | 1.67 (1.07-2.46) | 1355 (861-2050) | 1.66 (1.05-2.45) |
| Cape Verde | Female | 437 (395-481) | 36 (27-46) | 8.34 (6.33-10.33) | 25 (19-33) | 5.75 (4.38-7.29) | 16 (12-21) | 3.67 (2.79-4.80) | 10 (7-13) | 2.27 (1.68-3.01) | 8 (6-11) | 1.88 (1.36-2.55) |
| Cape Verde | Male | 297 (268-327) | 26 (19-33) | 8.71 (6.53-10.88) | 18 (13-24) | 6.00 (4.51-7.68) | 11 (8-15) | 3.84 (2.84-5.03) | 5 (3-7) | 1.59 (1.11-2.17) | 3 (2-5) | 1.17 (0.79-1.67) |
| Central African Republic | Female | 1373 (1232-1503) | 73 (48-102) | 5.30 (3.50-7.16) | 56 (36-77) | 4.10 (2.71-5.48) | 34 (23-47) | 2.51 (1.72-3.37) | 12 (7-18) | 0.91 (0.54-1.29) | 7 (4-10) | 0.49 (0.28-0.74) |
| Central African Republic | Male | 1129 (1018-1250) | 66 (42-91) | 5.80 (3.67-8.00) | 50 (32-68) | 4.38 (2.78-5.94) | 30 (20-41) | 2.69 (1.78-3.55) | 7 (4-11) | 0.61 (0.33-0.94) | 4 (2-6) | 0.35 (0.19-0.55) |
| Chad | Female | 4596 (4192-5007) | 310 (211-415) | 6.77 (4.67-8.89) | 223 (151-298) | 4.86 (3.27-6.46) | 134 (91-184) | 2.92 (2.00-3.94) | 52 (33-75) | 1.12 (0.73-1.62) | 33 (19-50) | 0.71 (0.43-1.09) |
| Chad | Male | 3991 (3608-4363) | 333 (234-432) | 8.33 (5.89-10.49) | 235 (165-316) | 5.91 (4.17-7.68) | 140 (95-185) | 3.51 (2.45-4.63) | 44 (27-67) | 1.10 (0.68-1.67) | 29 (17-48) | 0.73 (0.42-1.21) |
| Chile | Female | 45747 (40307-51377) | 5311 (3963-6716) | 11.62 (8.93-14.00) | 3105 (2292-4019) | 6.80 (5.04-8.49) | 2808 (2072-3662) | 6.19 (4.55-7.86) | 1523 (1112-2024) | 3.32 (2.47-4.27) | 1395 (1014-1875) | 3.05 (2.20-3.90) |
| Chile | Male | 33478 (29938-36649) | 4752 (3653-5939) | 14.22 (11.19-17.20) | 2754 (2051-3449) | 8.24 (6.41-10.25) | 2501 (1866-3228) | 7.51 (5.68-9.37) | 1251 (938-1622) | 3.77 (2.84-4.80) | 1103 (813-1447) | 3.29 (2.48-4.28) |
| China | Female | 1849012 (1716587-1992324) | 323478 (252270-394023) | 17.57 (13.74-20.78) | 199573 (149801-248476) | 10.79 (8.22-13.26) | 164047 (124048-209175) | 8.91 (6.75-11.24) | 56799 (41755-73181) | 3.08 (2.30-3.90) | 47040 (34195-61973) | 2.55 (1.85-3.32) |
| China | Male | 1408199 (1308160-1514413) | 284764 (225413-341521) | 20.25 (16.16-23.68) | 183742 (141459-227376) | 13.10 (10.15-15.83) | 152584 (116796-195115) | 10.83 (8.37-13.49) | 46516 (34648-61166) | 3.32 (2.49-4.31) | 36961 (27221-48389) | 2.63 (1.98-3.43) |
| Colombia | Female | 89230 (81187-97227) | 10997 (8696-13650) | 12.34 (9.80-14.97) | 7277 (5591-9087) | 8.18 (6.40-9.98) | 5025 (3788-6462) | 5.65 (4.30-7.05) | 3129 (2380-4060) | 3.52 (2.69-4.44) | 2705 (2060-3500) | 3.05 (2.34-3.89) |
| Colombia | Male | 76530 (69333-83741) | 13719 (10823-16339) | 17.99 (14.33-20.97) | 9046 (6889-11143) | 11.78 (9.23-14.32) | 6257 (4713-7966) | 8.20 (6.29-10.13) | 2928 (2163-3854) | 3.82 (2.89-4.88) | 2351 (1745-3161) | 3.06 (2.31-4.07) |
| Comoros | Female | 340 (305-373) | 42 (33-52) | 12.49 (9.91-15.12) | 29 (22-36) | 8.58 (6.74-10.48) | 18 (13-22) | 5.20 (4.01-6.57) | 8 (5-10) | 2.27 (1.63-3.03) | 5 (3-7) | 1.47 (1.02-2.07) |
| Comoros | Male | 233 (212-253) | 23 (17-29) | 9.78 (7.31-12.38) | 16 (12-21) | 6.86 (5.11-8.82) | 10 (7-13) | 4.22 (3.09-5.52) | 3 (2-4) | 1.25 (0.83-1.84) | 2 (1-3) | 0.83 (0.54-1.24) |
| Congo | Female | 2231 (1997-2439) | 123 (80-166) | 5.53 (3.68-7.43) | 93 (61-123) | 4.19 (2.77-5.63) | 60 (41-82) | 2.72 (1.85-3.62) | 22 (13-33) | 0.98 (0.60-1.48) | 13 (7-21) | 0.57 (0.31-0.90) |
| Congo | Male | 1650 (1496-1813) | 100 (67-139) | 6.07 (4.06-8.30) | 74 (49-102) | 4.53 (3.04-6.07) | 47 (32-66) | 2.90 (1.94-3.90) | 13 (8-21) | 0.80 (0.47-1.22) | 10 (5-16) | 0.58 (0.33-0.95) |
| Costa Rica | Female | 14121 (13140-15095) | 1556 (1145-1936) | 11.04 (8.21-13.53) | 955 (687-1249) | 6.77 (4.85-8.67) | 781 (558-1043) | 5.51 (3.98-7.28) | 449 (319-601) | 3.17 (2.25-4.18) | 431 (303-581) | 3.04 (2.16-4.05) |
| Costa Rica | Male | 10121 (9430-10817) | 1485 (1181-1817) | 14.74 (11.66-17.73) | 887 (693-1126) | 8.81 (6.94-11.03) | 727 (560-917) | 7.20 (5.61-8.97) | 340 (258-442) | 3.36 (2.61-4.36) | 318 (238-411) | 3.15 (2.36-4.06) |
| Cote d'Ivoire | Female | 11750 (10562-12860) | 962 (706-1214) | 8.22 (6.04-10.12) | 668 (475-856) | 5.70 (4.18-7.15) | 415 (303-547) | 3.55 (2.62-4.61) | 146 (97-205) | 1.24 (0.84-1.73) | 93 (59-140) | 0.79 (0.51-1.18) |
| Cote d'Ivoire | Male | 9395 (8477-10354) | 842 (613-1072) | 8.98 (6.65-11.21) | 581 (421-760) | 6.20 (4.56-7.97) | 363 (265-485) | 3.85 (2.86-5.08) | 104 (67-153) | 1.10 (0.72-1.61) | 72 (43-114) | 0.77 (0.46-1.23) |
| Croatia | Female | 12231 (10859-13529) | 2094 (1605-2608) | 17.11 (13.83-20.60) | 1177 (878-1549) | 9.60 (7.41-12.16) | 1112 (838-1431) | 9.09 (7.11-11.50) | 703 (512-939) | 5.76 (4.34-7.49) | 610 (442-834) | 5.01 (3.70-6.60) |
| Croatia | Male | 9455 (8434-10444) | 2044 (1644-2448) | 21.65 (17.89-25.26) | 1196 (921-1510) | 12.63 (9.88-15.71) | 1131 (875-1440) | 11.94 (9.36-15.03) | 657 (493-847) | 6.96 (5.32-8.82) | 477 (348-631) | 5.05 (3.72-6.56) |
| Cuba | Female | 26933 (24013-29999) | 1683 (1089-2354) | 6.21 (4.16-8.54) | 1140 (775-1573) | 4.23 (2.92-5.83) | 877 (582-1217) | 3.25 (2.26-4.47) | 525 (348-733) | 1.94 (1.30-2.71) | 485 (315-677) | 1.80 (1.19-2.51) |
| Cuba | Male | 21259 (19406-23203) | 2051 (1501-2586) | 9.62 (7.05-11.89) | 1309 (927-1695) | 6.16 (4.43-7.77) | 986 (709-1303) | 4.65 (3.31-6.08) | 466 (323-629) | 2.19 (1.52-2.90) | 416 (285-561) | 1.95 (1.37-2.60) |
| Cyprus | Female | 3520 (3118-3933) | 234 (144-339) | 6.65 (4.15-9.43) | 122 (77-170) | 3.46 (2.19-4.79) | 168 (107-239) | 4.77 (3.04-6.60) | 51 (30-79) | 1.46 (0.84-2.23) | 47 (27-73) | 1.34 (0.78-2.02) |
| Cyprus | Male | 2863 (2534-3197) | 331 (248-422) | 11.62 (8.77-14.28) | 153 (108-210) | 5.32 (3.84-7.16) | 215 (158-289) | 7.52 (5.61-9.69) | 67 (46-97) | 2.33 (1.63-3.28) | 62 (41-92) | 2.15 (1.49-3.17) |
| Czech Republic | Female | 24537 (21775-27347) | 4080 (3153-5118) | 16.84 (13.10-20.16) | 2042 (1532-2696) | 8.35 (6.31-10.58) | 2456 (1827-3170) | 10.01 (7.63-12.55) | 1050 (747-1436) | 4.29 (3.16-5.63) | 979 (695-1349) | 4.00 (2.88-5.27) |
| Czech Republic | Male | 19765 (17474-21776) | 4186 (3337-5128) | 21.25 (17.35-24.76) | 2194 (1658-2804) | 11.16 (8.58-13.99) | 2593 (1994-3295) | 13.19 (10.28-16.20) | 1177 (856-1569) | 5.99 (4.44-7.83) | 1113 (815-1490) | 5.66 (4.23-7.33) |
| Democratic Republic of the Congo | Female | 26257 (23864-28895) | 346 (111-735) | 1.33 (0.41-2.73) | 333 (105-671) | 1.27 (0.40-2.54) | 266 (93-507) | 1.01 (0.36-1.89) | 72 (18-175) | 0.27 (0.07-0.66) | 41 (10-107) | 0.16 (0.04-0.41) |
| Democratic Republic of the Congo | Male | 18076 (16400-19772) | 439 (182-778) | 2.42 (1.02-4.35) | 393 (172-670) | 2.18 (0.93-3.73) | 277 (122-452) | 1.53 (0.67-2.51) | 64 (21-134) | 0.35 (0.12-0.75) | 48 (17-101) | 0.27 (0.09-0.56) |
| Denmark | Female | 16727 (14756-18578) | 1227 (782-1748) | 7.39 (4.87-10.17) | 248 (136-420) | 1.48 (0.82-2.53) | 1111 (712-1589) | 6.67 (4.45-9.12) | 92 (48-169) | 0.56 (0.29-1.01) | 60 (30-112) | 0.36 (0.17-0.66) |
| Denmark | Male | 12697 (11389-14066) | 1507 (1133-1941) | 11.93 (9.12-14.53) | 279 (158-467) | 2.19 (1.23-3.63) | 1315 (974-1710) | 10.39 (7.81-13.03) | 126 (66-222) | 0.98 (0.53-1.72) | 86 (45-160) | 0.68 (0.35-1.26) |
| Djibouti | Female | 391 (350-430) | 46 (36-57) | 11.80 (9.24-14.11) | 31 (24-39) | 8.00 (6.14-9.71) | 20 (15-25) | 5.05 (3.82-6.30) | 8 (5-11) | 1.95 (1.33-2.61) | 5 (3-8) | 1.27 (0.81-1.87) |
| Djibouti | Male | 388 (349-430) | 36 (26-47) | 9.24 (6.76-11.85) | 25 (18-33) | 6.42 (4.55-8.18) | 16 (11-22) | 4.11 (2.95-5.44) | 5 (3-7) | 1.19 (0.74-1.82) | 3 (2-5) | 0.81 (0.44-1.27) |
| Dominica | Female | 129 (116-143) | 8 (5-11) | 6.49 (4.31-8.66) | 6 (4-8) | 4.50 (3.05-5.96) | 4 (3-6) | 3.31 (2.24-4.43) | 3 (2-4) | 2.12 (1.40-2.95) | 2 (2-3) | 1.83 (1.19-2.54) |
| Dominica | Male | 120 (109-132) | 12 (9-15) | 10.05 (7.64-12.29) | 8 (6-10) | 6.51 (4.85-8.21) | 6 (4-7) | 4.69 (3.47-6.03) | 2 (2-3) | 2.04 (1.43-2.80) | 2 (1-3) | 1.68 (1.15-2.36) |
| Dominican Republic | Female | 12149 (11055-13350) | 772 (528-1033) | 6.36 (4.36-8.38) | 534 (371-717) | 4.41 (3.07-5.76) | 399 (281-534) | 3.28 (2.36-4.30) | 273 (182-375) | 2.25 (1.51-3.06) | 232 (153-323) | 1.92 (1.28-2.61) |
| Dominican Republic | Male | 11034 (10026-12012) | 1075 (806-1328) | 9.69 (7.44-11.73) | 695 (515-885) | 6.32 (4.78-7.90) | 511 (372-654) | 4.66 (3.44-5.84) | 274 (199-358) | 2.49 (1.83-3.25) | 230 (167-308) | 2.08 (1.51-2.79) |
| Ecuador | Female | 31377 (28085-34636) | 2861 (1990-3654) | 9.13 (6.34-11.46) | 1915 (1285-2538) | 6.12 (4.14-7.87) | 1310 (890-1766) | 4.16 (2.78-5.50) | 653 (428-873) | 2.08 (1.38-2.75) | 585 (383-810) | 1.87 (1.23-2.56) |
| Ecuador | Male | 23652 (21211-25781) | 2789 (1992-3538) | 11.83 (8.65-14.61) | 1838 (1298-2360) | 7.81 (5.69-9.73) | 1268 (886-1652) | 5.38 (3.86-6.88) | 485 (324-661) | 2.06 (1.40-2.70) | 386 (258-526) | 1.64 (1.11-2.19) |
| Egypt | Female | 158251 (139909-177432) | 739 (127-2574) | 0.47 (0.08-1.59) | 683 (111-2412) | 0.44 (0.07-1.52) | 582 (78-1926) | 0.37 (0.05-1.20) | 190 (5-943) | 0.12 (0.00-0.60) | 168 (4-829) | 0.11 (0.00-0.53) |
| Egypt | Male | 122734 (110750-135543) | 6214 (3748-8972) | 5.05 (3.08-7.24) | 4698 (2923-6725) | 3.84 (2.44-5.44) | 3205 (2047-4578) | 2.61 (1.68-3.66) | 1064 (596-1663) | 0.87 (0.49-1.33) | 882 (482-1374) | 0.72 (0.40-1.11) |
| El Salvador | Female | 11979 (10830-13089) | 1263 (971-1561) | 10.64 (8.21-12.74) | 856 (640-1077) | 7.15 (5.38-8.84) | 566 (410-727) | 4.73 (3.49-6.05) | 368 (270-475) | 3.06 (2.26-3.93) | 332 (241-433) | 2.78 (2.06-3.59) |
| El Salvador | Male | 11197 (10235-12226) | 1549 (1210-1910) | 13.84 (10.93-16.91) | 1015 (776-1276) | 9.08 (7.02-11.23) | 673 (513-867) | 6.03 (4.59-7.64) | 263 (195-348) | 2.35 (1.78-3.05) | 227 (169-299) | 2.03 (1.52-2.65) |
| Equatorial Guinea | Female | 513 (458-564) | 29 (19-39) | 5.72 (3.82-7.68) | 21 (14-28) | 4.12 (2.82-5.53) | 15 (11-21) | 3.04 (2.06-4.04) | 5 (3-8) | 1.03 (0.62-1.57) | 3 (2-5) | 0.65 (0.35-1.02) |
| Equatorial Guinea | Male | 320 (290-352) | 20 (14-28) | 6.35 (4.38-8.55) | 14 (10-19) | 4.45 (3.06-5.90) | 10 (7-14) | 3.22 (2.24-4.27) | 2 (1-4) | 0.78 (0.46-1.19) | 2 (1-3) | 0.54 (0.30-0.88) |
| Eritrea | Female | 1787 (1601-1961) | 194 (144-246) | 10.85 (8.23-13.28) | 135 (98-173) | 7.55 (5.55-9.38) | 80 (58-103) | 4.47 (3.28-5.72) | 23 (15-33) | 1.29 (0.87-1.79) | 15 (10-23) | 0.86 (0.57-1.26) |
| Eritrea | Male | 1313 (1193-1439) | 103 (74-134) | 7.88 (5.68-10.17) | 74 (52-99) | 5.68 (4.06-7.39) | 44 (30-61) | 3.36 (2.36-4.51) | 9 (6-13) | 0.66 (0.42-0.99) | 6 (4-9) | 0.47 (0.28-0.71) |
| Estonia | Female | 2807 (2455-3161) | 5 (0-28) | 0.17 (0.00-0.97) | 5 (0-26) | 0.17 (0.00-0.93) | 5 (0-27) | 0.17 (0.00-0.96) | 1 (0-9) | 0.05 (0.00-0.32) | 1 (0-7) | 0.04 (0.00-0.25) |
| Estonia | Male | 1994 (1776-2228) | 143 (98-195) | 7.15 (5.01-9.51) | 76 (53-104) | 3.82 (2.69-5.06) | 93 (66-127) | 4.67 (3.36-6.23) | 20 (13-30) | 1.02 (0.64-1.43) | 16 (10-24) | 0.78 (0.50-1.15) |
| Ethiopia | Female | 25432 (23258-27714) | 2867 (2139-3573) | 11.28 (8.50-13.90) | 1996 (1449-2493) | 7.83 (5.80-9.74) | 1195 (853-1539) | 4.69 (3.39-6.04) | 332 (233-459) | 1.31 (0.90-1.76) | 162 (104-245) | 0.64 (0.40-0.93) |
| Ethiopia | Male | 21367 (19493-23191) | 1931 (1361-2496) | 9.06 (6.32-11.69) | 1357 (965-1824) | 6.42 (4.52-8.40) | 822 (574-1107) | 3.86 (2.64-5.19) | 194 (127-293) | 0.91 (0.61-1.34) | 127 (80-196) | 0.59 (0.38-0.92) |
| Federated States of Micronesia | Female | 111 (98-124) | 9 (6-13) | 8.29 (5.46-11.01) | 6 (4-9) | 5.66 (3.66-7.80) | 4 (3-6) | 3.69 (2.38-5.05) | 2 (1-3) | 1.58 (0.98-2.24) | 1 (1-2) | 0.97 (0.57-1.42) |
| Federated States of Micronesia | Male | 106 (96-118) | 11 (8-15) | 10.66 (7.58-13.70) | 8 (5-10) | 7.29 (5.03-9.47) | 5 (3-7) | 4.71 (3.18-6.28) | 2 (1-2) | 1.58 (1.01-2.19) | 1 (1-1) | 0.92 (0.54-1.31) |
| Fiji | Female | 966 (865-1071) | 105 (78-131) | 10.87 (8.26-13.30) | 70 (52-91) | 7.27 (5.51-9.25) | 47 (34-61) | 4.81 (3.56-6.25) | 27 (19-36) | 2.78 (2.05-3.74) | 20 (13-28) | 2.08 (1.45-2.85) |
| Fiji | Male | 1007 (900-1107) | 131 (99-164) | 12.90 (10.06-16.10) | 86 (65-112) | 8.56 (6.47-10.92) | 58 (41-76) | 5.70 (4.24-7.35) | 22 (16-32) | 2.23 (1.59-3.10) | 16 (10-23) | 1.56 (1.03-2.29) |
| Finland | Female | 16529 (14668-18564) | 953 (551-1409) | 5.75 (3.31-8.44) | 353 (206-531) | 2.15 (1.27-3.14) | 834 (493-1223) | 5.05 (3.01-7.16) | 169 (92-263) | 1.03 (0.57-1.55) | 146 (80-234) | 0.89 (0.48-1.36) |
| Finland | Male | 9276 (8267-10232) | 1149 (897-1412) | 12.48 (9.74-14.89) | 346 (227-492) | 3.74 (2.44-5.24) | 904 (675-1131) | 9.75 (7.50-12.04) | 162 (103-240) | 1.76 (1.12-2.52) | 150 (94-220) | 1.62 (1.01-2.36) |
| France | Female | 196033 (175372-215047) | 12888 (8029-18178) | 6.57 (4.24-9.04) | 5569 (3598-7778) | 2.84 (1.88-3.98) | 10157 (6707-14285) | 5.20 (3.43-7.01) | 2331 (1455-3482) | 1.20 (0.76-1.74) | 2105 (1276-3114) | 1.08 (0.66-1.60) |
| France | Male | 129259 (116248-142162) | 14210 (10697-17855) | 11.03 (8.47-13.60) | 5330 (3856-7471) | 4.14 (3.00-5.73) | 10320 (7527-13162) | 8.03 (6.05-10.19) | 2604 (1817-3782) | 2.02 (1.42-2.89) | 2414 (1653-3510) | 1.86 (1.31-2.70) |
| Gabon | Female | 1029 (931-1135) | 59 (39-79) | 5.68 (3.87-7.59) | 41 (28-56) | 4.03 (2.83-5.38) | 31 (22-42) | 3.06 (2.15-4.12) | 11 (7-17) | 1.11 (0.73-1.59) | 8 (5-13) | 0.79 (0.48-1.24) |
| Gabon | Male | 787 (708-868) | 49 (31-69) | 6.19 (4.01-8.57) | 34 (22-48) | 4.31 (2.88-5.85) | 26 (17-36) | 3.25 (2.17-4.46) | 7 (4-10) | 0.82 (0.47-1.27) | 5 (3-8) | 0.60 (0.33-1.01) |
| Georgia | Female | 5041 (4464-5563) | 391 (279-516) | 7.79 (5.65-9.95) | 275 (194-357) | 5.50 (3.95-6.95) | 187 (134-244) | 3.73 (2.71-4.75) | 126 (84-167) | 2.50 (1.74-3.22) | 110 (74-148) | 2.18 (1.46-2.85) |
| Georgia | Male | 3718 (3351-4115) | 475 (375-591) | 12.78 (10.20-15.30) | 313 (241-392) | 8.47 (6.54-10.35) | 209 (161-269) | 5.65 (4.37-7.05) | 116 (85-152) | 3.13 (2.34-4.04) | 89 (65-117) | 2.40 (1.77-3.07) |
| Germany | Female | 311911 (283202-343774) | 21372 (13556-29387) | 6.84 (4.42-9.39) | 7714 (4868-11137) | 2.48 (1.55-3.56) | 17653 (11322-24464) | 5.68 (3.72-7.76) | 3339 (1947-5167) | 1.08 (0.63-1.64) | 3192 (1888-4844) | 1.03 (0.60-1.54) |
| Germany | Male | 212763 (193893-230852) | 26086 (20337-32413) | 12.28 (9.68-14.84) | 8281 (5676-11516) | 3.91 (2.70-5.41) | 20115 (15176-25766) | 9.53 (7.19-11.84) | 3980 (2600-5687) | 1.87 (1.21-2.67) | 3746 (2384-5422) | 1.77 (1.15-2.52) |
| Ghana | Female | 17945 (16190-19749) | 1789 (1393-2188) | 10.00 (7.86-11.95) | 1230 (924-1560) | 6.87 (5.25-8.44) | 762 (575-986) | 4.26 (3.26-5.36) | 377 (274-495) | 2.12 (1.55-2.72) | 299 (210-405) | 1.67 (1.20-2.20) |
| Ghana | Male | 9796 (8873-10711) | 1035 (784-1287) | 10.59 (8.22-12.79) | 714 (531-912) | 7.27 (5.56-9.22) | 444 (326-577) | 4.57 (3.34-5.78) | 134 (93-189) | 1.37 (0.96-1.88) | 103 (71-146) | 1.05 (0.74-1.49) |
| Greece | Female | 39261 (34909-44017) | 2658 (1592-3677) | 6.81 (4.10-9.23) | 1641 (1040-2267) | 4.21 (2.69-5.71) | 1651 (1050-2302) | 4.19 (2.75-5.84) | 817 (510-1165) | 2.10 (1.27-2.92) | 812 (491-1162) | 2.07 (1.25-2.88) |
| Greece | Male | 25763 (23104-28551) | 2886 (2123-3638) | 11.27 (8.44-13.68) | 1611 (1183-2086) | 6.28 (4.64-7.98) | 1624 (1167-2125) | 6.31 (4.62-8.09) | 812 (561-1082) | 3.15 (2.23-4.12) | 802 (556-1062) | 3.12 (2.21-4.09) |
| Greenland | Female | 73 (65-81) | 6 (4-8) | 8.10 (5.54-10.40) | 2 (1-3) | 2.23 (1.32-3.40) | 5 (3-6) | 6.64 (4.53-8.76) | 1 (0-1) | 0.74 (0.42-1.16) | 0 (0-1) | 0.60 (0.34-0.95) |
| Greenland | Male | 73 (66-80) | 8 (5-10) | 10.94 (7.53-13.80) | 2 (1-3) | 3.03 (1.80-4.53) | 7 (4-9) | 8.90 (5.97-11.41) | 1 (0-1) | 1.02 (0.61-1.57) | 1 (0-1) | 0.88 (0.49-1.39) |
| Grenada | Female | 189 (169-208) | 12 (8-16) | 6.36 (4.12-8.47) | 8 (6-11) | 4.39 (2.97-5.85) | 6 (4-9) | 3.30 (2.28-4.49) | 4 (2-5) | 2.02 (1.32-2.83) | 3 (2-5) | 1.75 (1.15-2.47) |
| Grenada | Male | 205 (184-226) | 20 (15-25) | 9.61 (7.15-12.10) | 13 (9-17) | 6.17 (4.49-8.08) | 9 (7-13) | 4.64 (3.34-6.07) | 4 (3-6) | 2.08 (1.39-2.88) | 3 (2-5) | 1.54 (1.04-2.19) |
| Guatemala | Female | 22239 (19917-24301) | 2391 (1836-2942) | 10.77 (8.42-12.86) | 1610 (1191-2020) | 7.25 (5.51-9.04) | 1076 (786-1385) | 4.86 (3.64-6.14) | 520 (370-695) | 2.34 (1.70-3.12) | 359 (243-494) | 1.61 (1.13-2.21) |
| Guatemala | Male | 19789 (18087-21521) | 2811 (2167-3447) | 14.19 (11.15-17.01) | 1829 (1413-2298) | 9.29 (7.29-11.43) | 1223 (932-1551) | 6.19 (4.83-7.72) | 410 (299-536) | 2.05 (1.53-2.68) | 281 (195-382) | 1.42 (1.02-1.91) |
| Guinea | Female | 5344 (4814-5835) | 401 (292-514) | 7.51 (5.53-9.31) | 287 (212-374) | 5.36 (3.96-6.81) | 173 (126-226) | 3.23 (2.42-4.16) | 88 (60-121) | 1.65 (1.12-2.21) | 43 (27-65) | 0.81 (0.51-1.20) |
| Guinea | Male | 3886 (3529-4235) | 329 (231-424) | 8.50 (6.04-10.70) | 232 (164-310) | 6.03 (4.29-7.81) | 139 (95-190) | 3.60 (2.49-4.75) | 48 (30-72) | 1.24 (0.79-1.86) | 28 (16-45) | 0.73 (0.41-1.15) |
| Guinea-Bissau | Female | 742 (669-812) | 52 (37-67) | 7.07 (5.03-8.78) | 38 (26-49) | 5.09 (3.60-6.49) | 23 (15-30) | 3.04 (2.16-3.94) | 9 (6-13) | 1.26 (0.82-1.80) | 6 (4-9) | 0.81 (0.50-1.24) |
| Guinea-Bissau | Male | 497 (449-549) | 40 (29-52) | 8.04 (5.72-10.36) | 28 (20-38) | 5.73 (4.08-7.44) | 17 (12-23) | 3.41 (2.46-4.62) | 5 (3-8) | 1.03 (0.64-1.53) | 3 (2-6) | 0.68 (0.37-1.14) |
| Guyana | Female | 1008 (908-1116) | 60 (40-82) | 5.96 (4.02-8.17) | 41 (28-56) | 4.09 (2.79-5.53) | 32 (22-43) | 3.18 (2.18-4.28) | 17 (11-23) | 1.66 (1.12-2.27) | 13 (8-19) | 1.31 (0.85-1.84) |
| Guyana | Male | 878 (793-959) | 82 (60-105) | 9.37 (6.97-11.63) | 52 (38-68) | 5.96 (4.29-7.69) | 40 (29-53) | 4.55 (3.32-6.02) | 15 (11-20) | 1.68 (1.21-2.29) | 12 (8-16) | 1.33 (0.94-1.82) |
| Haiti | Female | 9356 (8415-10304) | 572 (390-772) | 6.12 (4.25-8.16) | 425 (302-563) | 4.54 (3.30-6.07) | 269 (194-355) | 2.87 (2.10-3.79) | 158 (108-221) | 1.68 (1.16-2.35) | 85 (55-125) | 0.91 (0.59-1.32) |
| Haiti | Male | 7579 (6994-8288) | 730 (559-887) | 9.60 (7.47-11.46) | 507 (381-640) | 6.67 (5.11-8.24) | 310 (228-400) | 4.08 (3.04-5.20) | 111 (81-148) | 1.47 (1.07-1.92) | 61 (42-83) | 0.80 (0.57-1.09) |
| Honduras | Female | 11666 (10542-12806) | 1158 (844-1439) | 9.99 (7.48-12.07) | 797 (563-1016) | 6.84 (4.98-8.63) | 500 (348-661) | 4.24 (3.04-5.56) | 313 (221-416) | 2.71 (1.96-3.56) | 295 (203-390) | 2.54 (1.77-3.32) |
| Honduras | Male | 9050 (8231-9872) | 1266 (970-1565) | 14.01 (10.84-16.90) | 844 (649-1064) | 9.31 (7.18-11.65) | 531 (394-679) | 5.87 (4.42-7.36) | 221 (162-305) | 2.45 (1.79-3.27) | 196 (136-269) | 2.16 (1.54-2.97) |
| Hungary | Female | 25390 (22424-28538) | 4802 (3766-5970) | 18.98 (15.37-22.38) | 2651 (2025-3480) | 10.50 (8.16-13.15) | 2581 (1949-3317) | 10.16 (7.88-12.66) | 1400 (1011-1915) | 5.52 (4.13-7.26) | 1252 (879-1744) | 4.93 (3.52-6.57) |
| Hungary | Male | 16334 (14545-17845) | 3807 (3150-4556) | 23.41 (19.63-26.82) | 2285 (1766-2871) | 14.03 (11.03-17.17) | 2205 (1712-2778) | 13.58 (10.74-16.58) | 1040 (755-1388) | 6.40 (4.72-8.39) | 945 (699-1292) | 5.84 (4.34-7.72) |
| Iceland | Female | 642 (577-702) | 45 (30-62) | 7.08 (4.77-9.35) | 10 (5-16) | 1.50 (0.84-2.41) | 41 (27-55) | 6.37 (4.41-8.49) | 5 (2-8) | 0.72 (0.38-1.20) | 4 (2-7) | 0.65 (0.34-1.12) |
| Iceland | Male | 515 (466-564) | 68 (55-85) | 13.32 (10.63-16.35) | 13 (8-21) | 2.51 (1.49-3.96) | 59 (47-75) | 11.57 (9.04-14.34) | 7 (4-12) | 1.35 (0.77-2.22) | 7 (4-11) | 1.27 (0.72-2.12) |
| India | Female | 1058872 (965145-1153518) | 100351 (75894-122484) | 9.50 (7.28-11.33) | 68759 (51341-88272) | 6.54 (4.91-8.11) | 43120 (31692-55789) | 4.08 (2.97-5.18) | 13925 (9769-18564) | 1.31 (0.91-1.73) | 12334 (8519-16564) | 1.17 (0.81-1.52) |
| India | Male | 1024042 (941643-1101673) | 121052 (94002-147883) | 11.85 (9.21-14.22) | 82823 (62698-102767) | 8.13 (6.17-9.94) | 51275 (38116-65554) | 5.02 (3.73-6.26) | 13356 (9783-17775) | 1.31 (0.97-1.71) | 11159 (8011-14932) | 1.10 (0.80-1.43) |
| Indonesia | Female | 246115 (225223-267707) | 35626 (27722-43596) | 14.54 (11.59-17.32) | 23313 (17602-29357) | 9.50 (7.31-11.61) | 15396 (11730-19256) | 6.26 (4.83-7.75) | 5798 (4228-7612) | 2.35 (1.75-3.06) | 3141 (2266-4328) | 1.28 (0.91-1.72) |
| Indonesia | Male | 181951 (165259-197186) | 29072 (22404-35579) | 16.03 (12.67-19.13) | 19174 (14563-24040) | 10.56 (8.13-13.00) | 12618 (9470-16309) | 6.95 (5.34-8.78) | 3087 (2220-4178) | 1.70 (1.24-2.27) | 1746 (1227-2395) | 0.96 (0.69-1.30) |
| Iran | Female | 183445 (169117-200716) | 715 (49-2563) | 0.39 (0.03-1.42) | 674 (44-2475) | 0.36 (0.02-1.33) | 598 (40-2027) | 0.33 (0.02-1.10) | 171 (1-862) | 0.09 (0.00-0.49) | 145 (1-748) | 0.08 (0.00-0.41) |
| Iran | Male | 137157 (127120-148033) | 6468 (3910-9219) | 4.76 (2.84-6.62) | 4909 (3075-6874) | 3.60 (2.28-5.01) | 3356 (2113-4732) | 2.45 (1.54-3.41) | 997 (547-1482) | 0.73 (0.41-1.08) | 796 (436-1197) | 0.58 (0.32-0.87) |
| Iraq | Female | 61072 (54668-67412) | 286 (53-939) | 0.47 (0.09-1.47) | 261 (46-861) | 0.43 (0.07-1.38) | 233 (33-687) | 0.38 (0.05-1.13) | 81 (5-372) | 0.14 (0.01-0.59) | 60 (2-286) | 0.10 (0.00-0.48) |
| Iraq | Male | 47967 (43244-52538) | 2589 (1652-3680) | 5.43 (3.46-7.45) | 1965 (1257-2692) | 4.08 (2.65-5.51) | 1353 (903-1860) | 2.83 (1.93-3.81) | 648 (367-971) | 1.36 (0.79-1.97) | 479 (286-734) | 1.00 (0.60-1.53) |
| Ireland | Female | 10430 (9622-11275) | 306 (130-537) | 2.95 (1.23-5.13) | 39 (19-83) | 0.37 (0.18-0.78) | 303 (129-530) | 2.94 (1.23-5.02) | 13 (6-28) | 0.12 (0.05-0.26) | 11 (5-25) | 0.11 (0.05-0.24) |
| Ireland | Male | 8474 (7757-9183) | 834 (618-1058) | 9.93 (7.31-12.24) | 63 (42-119) | 0.74 (0.49-1.40) | 796 (590-1011) | 9.42 (6.94-11.69) | 20 (13-42) | 0.24 (0.15-0.49) | 18 (11-40) | 0.21 (0.13-0.47) |
| Israel | Female | 21645 (19039-24280) | 1454 (943-2047) | 6.76 (4.49-9.07) | 507 (315-755) | 2.35 (1.43-3.43) | 1215 (792-1688) | 5.64 (3.76-7.67) | 209 (119-324) | 0.96 (0.55-1.46) | 188 (106-290) | 0.86 (0.49-1.31) |
| Israel | Male | 15684 (14109-17408) | 1788 (1382-2239) | 11.44 (9.13-14.02) | 561 (381-774) | 3.58 (2.45-4.92) | 1392 (1045-1809) | 8.89 (6.89-11.12) | 226 (146-327) | 1.45 (0.94-2.07) | 208 (131-300) | 1.32 (0.84-1.88) |
| Italy | Female | 198271 (179524-215463) | 17963 (12666-23562) | 9.09 (6.43-11.81) | 8022 (5419-10984) | 4.02 (2.76-5.47) | 12524 (8856-16813) | 6.33 (4.57-8.19) | 3939 (2569-5600) | 1.99 (1.31-2.76) | 3655 (2322-5234) | 1.84 (1.20-2.60) |
| Italy | Male | 133431 (121758-145166) | 17030 (13043-21313) | 12.76 (10.03-15.64) | 7303 (5409-9739) | 5.49 (4.09-7.18) | 11431 (8606-14994) | 8.59 (6.55-10.92) | 3551 (2503-4792) | 2.66 (1.94-3.54) | 3418 (2425-4688) | 2.56 (1.85-3.43) |
| Jamaica | Female | 4410 (3966-4805) | 285 (196-377) | 6.48 (4.51-8.42) | 204 (145-268) | 4.63 (3.29-6.03) | 139 (100-186) | 3.19 (2.30-4.18) | 101 (69-135) | 2.28 (1.59-3.07) | 87 (58-118) | 1.98 (1.35-2.66) |
| Jamaica | Male | 4187 (3790-4591) | 410 (313-510) | 9.75 (7.59-11.97) | 275 (204-354) | 6.57 (4.97-8.33) | 183 (137-239) | 4.39 (3.35-5.65) | 91 (65-121) | 2.17 (1.57-2.83) | 72 (50-98) | 1.72 (1.23-2.32) |
| Japan | Female | 499332 (451910-547237) | 69494 (56284-85487) | 13.96 (11.43-16.75) | 28379 (20116-37287) | 5.71 (4.16-7.25) | 48933 (37659-61311) | 9.81 (7.79-12.18) | 12313 (8514-16856) | 2.48 (1.75-3.31) | 11230 (7738-15434) | 2.26 (1.61-3.02) |
| Japan | Male | 493406 (449794-531770) | 83111 (65543-100132) | 17.01 (13.53-20.05) | 32198 (24010-42858) | 6.54 (4.96-8.62) | 56490 (43207-71074) | 11.47 (8.90-14.01) | 15606 (11513-21521) | 3.17 (2.32-4.37) | 13603 (9885-18427) | 2.76 (2.00-3.76) |
| Jordan | Female | 19061 (16916-21226) | 66 (7-268) | 0.34 (0.03-1.37) | 61 (7-257) | 0.33 (0.03-1.28) | 55 (5-197) | 0.29 (0.03-1.03) | 18 (0-110) | 0.10 (0.00-0.58) | 18 (0-103) | 0.09 (0.00-0.54) |
| Jordan | Male | 16255 (14783-17800) | 805 (507-1203) | 4.97 (3.17-7.26) | 612 (392-886) | 3.79 (2.42-5.39) | 417 (267-593) | 2.56 (1.67-3.59) | 191 (106-295) | 1.19 (0.66-1.78) | 174 (99-274) | 1.08 (0.62-1.65) |
| Kazakhstan | Female | 17378 (15503-19210) | 1320 (951-1753) | 7.60 (5.56-9.88) | 879 (635-1181) | 5.04 (3.73-6.66) | 689 (499-903) | 3.96 (2.95-5.13) | 411 (290-566) | 2.36 (1.69-3.21) | 368 (254-510) | 2.11 (1.51-2.88) |
| Kazakhstan | Male | 14170 (12761-15622) | 1770 (1404-2191) | 12.49 (10.00-15.14) | 1102 (858-1375) | 7.83 (6.13-9.58) | 844 (656-1106) | 5.99 (4.63-7.69) | 434 (331-563) | 3.09 (2.32-3.87) | 369 (277-479) | 2.62 (1.96-3.32) |
| Kenya | Female | 14157 (12907-15409) | 1690 (1335-2074) | 11.92 (9.73-14.48) | 1146 (882-1441) | 8.15 (6.46-10.24) | 708 (545-903) | 5.02 (3.93-6.28) | 281 (201-377) | 1.99 (1.44-2.66) | 137 (94-192) | 0.97 (0.67-1.34) |
| Kenya | Male | 11849 (10940-12950) | 7 (0-98) | 0.06 (0.00-0.81) | 7 (0-96) | 0.06 (0.00-0.81) | 7 (0-71) | 0.06 (0.00-0.60) | 1 (0-15) | 0.01 (0.00-0.13) | 0 (0-6) | 0.00 (0.00-0.05) |
| Kiribati | Female | 85 (77-94) | 6 (4-9) | 7.32 (4.42-9.74) | 4 (3-6) | 5.17 (3.13-6.93) | 3 (2-4) | 3.19 (1.96-4.43) | 1 (1-1) | 1.05 (0.63-1.53) | 0 (0-1) | 0.53 (0.29-0.83) |
| Kiribati | Male | 71 (65-78) | 8 (5-10) | 10.49 (7.73-13.25) | 5 (4-7) | 7.23 (5.31-9.42) | 3 (2-4) | 4.49 (3.16-5.89) | 1 (1-1) | 1.27 (0.88-1.80) | 1 (0-1) | 0.73 (0.45-1.09) |
| Kuwait | Female | 6552 (5816-7265) | 277 (159-412) | 4.26 (2.49-6.13) | 156 (95-225) | 2.41 (1.46-3.42) | 211 (125-306) | 3.21 (1.92-4.56) | 56 (33-82) | 0.85 (0.49-1.24) | 52 (30-77) | 0.80 (0.46-1.15) |
| Kuwait | Male | 6142 (5517-6694) | 584 (427-735) | 9.56 (7.02-11.69) | 275 (195-368) | 4.52 (3.12-5.93) | 380 (281-490) | 6.21 (4.60-7.85) | 92 (64-125) | 1.49 (1.03-2.00) | 83 (58-113) | 1.35 (0.94-1.81) |
| Kyrgyzstan | Female | 3793 (3416-4198) | 239 (157-323) | 6.30 (4.24-8.39) | 174 (117-240) | 4.60 (3.08-6.19) | 107 (71-148) | 2.82 (1.88-3.83) | 62 (40-87) | 1.64 (1.08-2.30) | 53 (35-77) | 1.40 (0.91-1.99) |
| Kyrgyzstan | Male | 3186 (2878-3491) | 357 (259-452) | 11.25 (8.16-13.93) | 246 (174-314) | 7.73 (5.53-9.79) | 149 (106-195) | 4.66 (3.35-6.06) | 55 (38-75) | 1.73 (1.20-2.35) | 39 (26-54) | 1.23 (0.83-1.66) |
| Laos | Female | 6073 (5479-6685) | 783 (572-988) | 12.86 (9.52-16.08) | 523 (375-670) | 8.63 (6.28-10.90) | 328 (236-427) | 5.42 (3.95-7.05) | 112 (78-157) | 1.85 (1.30-2.53) | 81 (53-117) | 1.34 (0.88-1.92) |
| Laos | Male | 4461 (4072-4880) | 671 (530-831) | 15.02 (11.78-18.16) | 454 (345-582) | 10.14 (7.87-12.61) | 284 (215-369) | 6.34 (4.88-8.02) | 72 (50-99) | 1.61 (1.13-2.19) | 51 (34-74) | 1.13 (0.75-1.62) |
| Latvia | Female | 3210 (2898-3571) | 118 (59-185) | 3.65 (1.85-5.77) | 88 (48-131) | 2.75 (1.51-4.02) | 90 (49-134) | 2.80 (1.53-4.12) | 42 (21-66) | 1.31 (0.67-2.01) | 35 (17-55) | 1.08 (0.54-1.68) |
| Latvia | Male | 2079 (1852-2265) | 269 (211-326) | 12.96 (10.46-15.32) | 147 (112-186) | 7.09 (5.47-8.66) | 151 (117-192) | 7.29 (5.75-8.94) | 59 (43-78) | 2.84 (2.12-3.65) | 47 (33-63) | 2.28 (1.67-2.99) |
| Lebanon | Female | 14691 (13324-16331) | 51 (8-217) | 0.35 (0.06-1.52) | 49 (7-212) | 0.33 (0.05-1.43) | 44 (5-175) | 0.30 (0.04-1.18) | 13 (0-86) | 0.09 (0.00-0.59) | 13 (0-82) | 0.09 (0.00-0.56) |
| Lebanon | Male | 9891 (8985-10820) | 492 (292-718) | 5.04 (3.02-7.20) | 378 (234-533) | 3.86 (2.43-5.39) | 262 (167-370) | 2.66 (1.73-3.68) | 117 (64-181) | 1.18 (0.67-1.80) | 111 (59-172) | 1.14 (0.62-1.72) |
| Lesotho | Female | 1169 (1059-1278) | 69 (46-94) | 5.86 (3.89-8.03) | 53 (35-72) | 4.56 (3.07-6.11) | 33 (22-45) | 2.86 (1.95-3.84) | 18 (11-26) | 1.54 (0.93-2.20) | 16 (10-24) | 1.40 (0.83-2.06) |
| Lesotho | Male | 1021 (926-1113) | 58 (38-80) | 5.66 (3.78-7.66) | 44 (29-61) | 4.27 (2.88-5.82) | 27 (18-37) | 2.68 (1.79-3.55) | 6 (3-10) | 0.61 (0.34-0.96) | 4 (2-7) | 0.44 (0.24-0.71) |
| Liberia | Female | 2243 (2012-2466) | 163 (119-206) | 7.27 (5.42-9.09) | 117 (85-151) | 5.24 (3.79-6.61) | 70 (50-92) | 3.12 (2.30-4.07) | 29 (20-39) | 1.28 (0.88-1.75) | 18 (12-26) | 0.80 (0.51-1.15) |
| Liberia | Male | 1487 (1345-1622) | 126 (91-162) | 8.49 (6.19-10.69) | 90 (64-117) | 6.02 (4.38-7.67) | 53 (38-71) | 3.58 (2.60-4.71) | 18 (12-28) | 1.25 (0.82-1.80) | 13 (8-19) | 0.86 (0.52-1.29) |
| Libya | Female | 12860 (11488-14229) | 57 (13-173) | 0.45 (0.10-1.34) | 52 (10-162) | 0.40 (0.08-1.23) | 45 (7-138) | 0.35 (0.06-1.10) | 12 (0-62) | 0.10 (0.00-0.49) | 10 (0-51) | 0.07 (0.00-0.41) |
| Libya | Male | 9525 (8680-10418) | 501 (322-700) | 5.29 (3.41-7.32) | 376 (246-513) | 3.92 (2.63-5.40) | 267 (183-368) | 2.82 (1.95-3.80) | 91 (52-139) | 0.95 (0.54-1.43) | 70 (38-112) | 0.74 (0.41-1.16) |
| Lithuania | Female | 4017 (3566-4400) | 375 (288-475) | 9.39 (7.36-11.74) | 198 (148-264) | 4.95 (3.78-6.41) | 222 (166-287) | 5.56 (4.27-6.98) | 97 (71-132) | 2.44 (1.79-3.25) | 78 (54-109) | 1.95 (1.37-2.68) |
| Lithuania | Male | 2768 (2494-3012) | 282 (219-351) | 10.21 (8.01-12.32) | 148 (112-193) | 5.41 (4.16-6.83) | 166 (126-217) | 5.98 (4.65-7.62) | 66 (48-88) | 2.38 (1.76-3.12) | 46 (32-64) | 1.66 (1.17-2.28) |
| Luxembourg | Female | 1616 (1447-1786) | 111 (72-154) | 6.90 (4.48-9.32) | 8 (5-11) | 0.47 (0.33-0.70) | 109 (71-149) | 6.76 (4.37-9.05) | 3 (2-5) | 0.17 (0.12-0.27) | 3 (2-4) | 0.16 (0.10-0.25) |
| Luxembourg | Male | 1120 (1003-1234) | 129 (98-163) | 11.61 (9.04-14.26) | 8 (6-12) | 0.70 (0.52-1.02) | 124 (94-158) | 11.20 (8.70-13.72) | 3 (2-5) | 0.30 (0.22-0.46) | 3 (2-5) | 0.29 (0.20-0.43) |
| Macedonia | Female | 4434 (3928-4898) | 674 (528-848) | 15.27 (12.36-18.70) | 433 (330-558) | 9.84 (7.71-12.21) | 305 (228-398) | 6.91 (5.33-8.78) | 212 (155-284) | 4.80 (3.62-6.26) | 190 (136-257) | 4.28 (3.17-5.65) |
| Macedonia | Male | 4140 (3699-4585) | 851 (680-1028) | 20.72 (16.83-24.08) | 562 (441-718) | 13.59 (10.75-16.91) | 403 (302-514) | 9.80 (7.49-12.14) | 242 (178-336) | 5.89 (4.36-7.89) | 208 (147-291) | 5.03 (3.61-6.99) |
| Madagascar | Female | 6231 (5577-6846) | 703 (513-877) | 11.31 (8.51-13.97) | 489 (350-615) | 7.89 (5.83-9.82) | 288 (206-376) | 4.66 (3.40-5.88) | 109 (73-150) | 1.74 (1.20-2.40) | 56 (35-85) | 0.90 (0.56-1.37) |
| Madagascar | Male | 5168 (4690-5721) | 472 (349-614) | 9.15 (6.70-11.59) | 335 (240-443) | 6.49 (4.70-8.30) | 199 (142-276) | 3.86 (2.81-5.17) | 52 (34-79) | 1.01 (0.66-1.50) | 28 (16-46) | 0.55 (0.31-0.88) |
| Malawi | Female | 4756 (4304-5238) | 592 (462-733) | 12.44 (9.92-15.11) | 409 (309-514) | 8.55 (6.73-10.58) | 243 (182-309) | 5.10 (3.85-6.39) | 122 (91-162) | 2.56 (1.91-3.29) | 86 (61-117) | 1.81 (1.29-2.43) |
| Malawi | Male | 3793 (3416-4141) | 362 (274-467) | 9.53 (7.22-12.05) | 255 (189-334) | 6.76 (5.01-8.65) | 152 (109-205) | 4.03 (2.94-5.36) | 41 (28-60) | 1.08 (0.74-1.54) | 29 (19-42) | 0.75 (0.51-1.10) |
| Malaysia | Female | 44367 (39660-49088) | 6699 (5206-8334) | 15.16 (12.14-18.61) | 4073 (3090-5186) | 9.17 (7.21-11.70) | 3212 (2430-4142) | 7.28 (5.62-9.15) | 1445 (1064-1943) | 3.28 (2.44-4.30) | 1314 (948-1806) | 2.97 (2.21-3.97) |
| Malaysia | Male | 29196 (26558-32258) | 4947 (3952-6050) | 16.92 (13.77-19.93) | 3030 (2342-3852) | 10.32 (8.15-12.69) | 2376 (1841-3051) | 8.15 (6.38-10.30) | 891 (653-1182) | 3.04 (2.29-3.96) | 805 (593-1066) | 2.75 (2.08-3.55) |
| Maldives | Female | 513 (459-567) | 73 (56-91) | 14.37 (11.18-17.35) | 45 (34-58) | 8.82 (6.78-10.94) | 35 (25-45) | 6.73 (5.08-8.55) | 12 (9-17) | 2.42 (1.73-3.29) | 10 (6-13) | 1.86 (1.28-2.58) |
| Maldives | Male | 498 (451-546) | 76 (59-93) | 15.28 (12.12-18.29) | 47 (36-60) | 9.44 (7.35-11.80) | 36 (27-46) | 7.27 (5.58-9.12) | 9 (7-13) | 1.90 (1.36-2.61) | 7 (5-10) | 1.41 (0.97-2.03) |
| Mali | Female | 7907 (7186-8712) | 561 (384-732) | 7.11 (4.93-9.08) | 402 (281-532) | 5.08 (3.52-6.72) | 239 (167-324) | 3.04 (2.14-4.03) | 120 (78-171) | 1.52 (0.99-2.11) | 84 (53-128) | 1.07 (0.68-1.58) |
| Mali | Male | 5636 (5064-6153) | 443 (309-584) | 7.90 (5.46-10.39) | 312 (214-426) | 5.58 (3.84-7.44) | 189 (131-256) | 3.35 (2.35-4.51) | 73 (46-111) | 1.29 (0.81-1.94) | 53 (31-83) | 0.93 (0.56-1.46) |
| Malta | Female | 1552 (1372-1699) | 142 (99-186) | 9.21 (6.63-11.78) | 66 (46-91) | 4.24 (2.95-5.70) | 97 (68-132) | 6.28 (4.50-8.25) | 29 (19-41) | 1.84 (1.21-2.58) | 28 (18-41) | 1.79 (1.18-2.55) |
| Malta | Male | 1104 (982-1221) | 141 (109-177) | 12.82 (9.96-16.00) | 62 (46-85) | 5.66 (4.19-7.56) | 93 (68-122) | 8.48 (6.29-10.88) | 30 (21-42) | 2.72 (1.93-3.75) | 29 (20-41) | 2.66 (1.87-3.67) |
| Marshall Islands | Female | 45 (40-50) | 3 (2-5) | 7.66 (5.21-10.17) | 2 (1-3) | 5.09 (3.31-6.91) | 2 (1-2) | 3.57 (2.32-4.96) | 1 (0-1) | 1.28 (0.81-1.82) | 0 (0-1) | 0.94 (0.58-1.38) |
| Marshall Islands | Male | 44 (40-48) | 5 (3-6) | 10.61 (7.73-13.42) | 3 (2-4) | 7.01 (5.14-9.06) | 2 (2-3) | 4.90 (3.49-6.39) | 1 (0-1) | 1.40 (0.94-1.91) | 0 (0-1) | 1.00 (0.66-1.47) |
| Mauritania | Female | 2616 (2344-2863) | 198 (139-259) | 7.61 (5.49-9.57) | 140 (99-184) | 5.36 (3.84-6.89) | 86 (60-117) | 3.30 (2.35-4.27) | 38 (24-54) | 1.46 (0.94-2.06) | 24 (15-38) | 0.93 (0.56-1.40) |
| Mauritania | Male | 1555 (1409-1717) | 134 (96-174) | 8.63 (6.25-11.02) | 93 (66-122) | 6.00 (4.29-7.81) | 57 (41-78) | 3.70 (2.70-5.00) | 19 (11-29) | 1.20 (0.73-1.81) | 13 (7-21) | 0.82 (0.44-1.33) |
| Mauritius | Female | 4079 (3691-4485) | 595 (451-750) | 14.62 (11.22-18.19) | 370 (275-473) | 9.07 (6.91-11.65) | 278 (204-362) | 6.78 (5.07-8.78) | 143 (104-192) | 3.50 (2.64-4.61) | 136 (97-183) | 3.33 (2.48-4.42) |
| Mauritius | Male | 3148 (2830-3487) | 516 (382-640) | 16.37 (12.69-19.62) | 324 (238-421) | 10.30 (7.81-12.91) | 242 (176-320) | 7.67 (5.74-9.94) | 108 (77-145) | 3.45 (2.53-4.55) | 101 (71-137) | 3.21 (2.32-4.23) |
| Mexico | Female | 335425 (310930-360940) | 28889 (20525-37296) | 8.63 (6.20-10.99) | 18112 (12838-24470) | 5.43 (3.78-7.16) | 14003 (9946-19082) | 4.18 (2.95-5.64) | 6162 (4315-8355) | 1.84 (1.29-2.46) | 5594 (3889-7747) | 1.67 (1.16-2.28) |
| Mexico | Male | 247527 (230301-262860) | 26719 (19957-32955) | 10.82 (8.14-13.06) | 16967 (12548-21367) | 6.84 (5.08-8.60) | 13189 (9515-16819) | 5.33 (3.89-6.66) | 3744 (2767-4745) | 1.51 (1.13-1.94) | 3274 (2380-4224) | 1.32 (0.98-1.71) |
| Moldova | Female | 3696 (3342-4075) | 124 (66-203) | 3.36 (1.77-5.28) | 105 (57-166) | 2.84 (1.55-4.35) | 79 (45-121) | 2.16 (1.21-3.18) | 43 (21-74) | 1.16 (0.57-1.93) | 32 (15-55) | 0.86 (0.40-1.43) |
| Moldova | Male | 2614 (2373-2858) | 256 (195-315) | 9.81 (7.55-11.88) | 172 (130-217) | 6.61 (5.08-8.22) | 116 (87-150) | 4.43 (3.37-5.60) | 60 (43-81) | 2.30 (1.68-3.06) | 37 (25-52) | 1.41 (0.99-1.96) |
| Mongolia | Female | 2451 (2179-2724) | 179 (127-235) | 7.33 (5.26-9.47) | 126 (89-166) | 5.13 (3.70-6.74) | 84 (59-111) | 3.43 (2.42-4.54) | 46 (31-63) | 1.86 (1.30-2.54) | 40 (27-54) | 1.62 (1.11-2.21) |
| Mongolia | Male | 2025 (1829-2232) | 255 (199-313) | 12.62 (10.05-15.16) | 169 (130-211) | 8.32 (6.53-10.16) | 111 (85-144) | 5.51 (4.24-6.93) | 55 (42-71) | 2.71 (2.07-3.44) | 41 (31-53) | 1.99 (1.52-2.58) |
| Montenegro | Female | 1409 (1240-1570) | 222 (170-280) | 15.87 (12.50-19.33) | 139 (104-180) | 9.86 (7.54-12.53) | 105 (79-137) | 7.49 (5.69-9.47) | 69 (49-95) | 4.91 (3.59-6.60) | 62 (43-87) | 4.40 (3.15-6.02) |
| Montenegro | Male | 1174 (1030-1310) | 242 (190-295) | 20.61 (16.72-24.13) | 154 (118-199) | 13.26 (10.38-16.28) | 119 (89-151) | 10.09 (7.88-12.67) | 66 (48-91) | 5.69 (4.19-7.46) | 57 (39-78) | 4.87 (3.47-6.67) |
| Morocco | Female | 74253 (66767-81124) | 331 (52-1090) | 0.44 (0.07-1.48) | 309 (44-1035) | 0.42 (0.06-1.37) | 271 (33-871) | 0.37 (0.04-1.18) | 69 (1-327) | 0.09 (0.00-0.45) | 53 (1-258) | 0.07 (0.00-0.35) |
| Morocco | Male | 55208 (49793-60673) | 2783 (1712-4065) | 5.04 (3.08-7.28) | 2131 (1309-3043) | 3.86 (2.41-5.40) | 1447 (898-2006) | 2.62 (1.68-3.61) | 423 (234-654) | 0.77 (0.41-1.17) | 279 (151-452) | 0.51 (0.28-0.80) |
| Mozambique | Female | 6908 (6206-7586) | 869 (692-1064) | 12.57 (10.23-14.99) | 605 (471-756) | 8.72 (6.98-10.54) | 355 (273-446) | 5.15 (4.01-6.39) | 123 (89-167) | 1.80 (1.29-2.35) | 65 (44-95) | 0.95 (0.64-1.36) |
| Mozambique | Male | 5254 (4756-5760) | 498 (369-631) | 9.47 (7.16-11.91) | 356 (259-469) | 6.79 (4.97-8.78) | 208 (148-284) | 3.98 (2.90-5.23) | 45 (30-67) | 0.86 (0.58-1.27) | 23 (13-36) | 0.44 (0.25-0.68) |
| Myanmar | Female | 67709 (60753-74212) | 9417 (6874-11655) | 14.02 (10.61-16.99) | 6385 (4542-8069) | 9.47 (7.07-11.81) | 3851 (2760-4969) | 5.73 (4.18-7.21) | 2169 (1515-2829) | 3.22 (2.31-4.14) | 1395 (959-1912) | 2.07 (1.46-2.85) |
| Myanmar | Male | 36581 (33271-40429) | 5702 (4349-6993) | 15.58 (12.06-18.32) | 3884 (2889-4936) | 10.60 (8.04-13.09) | 2346 (1759-3070) | 6.41 (4.89-8.22) | 968 (681-1290) | 2.64 (1.91-3.44) | 556 (385-772) | 1.52 (1.06-2.07) |
| Namibia | Female | 1385 (1256-1519) | 79 (50-109) | 5.73 (3.66-7.85) | 59 (39-80) | 4.28 (2.84-5.72) | 41 (27-56) | 2.95 (1.97-3.99) | 18 (11-27) | 1.33 (0.85-1.90) | 16 (10-24) | 1.16 (0.70-1.68) |
| Namibia | Male | 1336 (1207-1455) | 78 (50-109) | 5.83 (3.77-8.04) | 57 (37-81) | 4.28 (2.77-5.98) | 39 (26-55) | 2.95 (1.96-4.10) | 13 (7-21) | 0.98 (0.54-1.51) | 12 (6-18) | 0.86 (0.47-1.37) |
| Nauru | Female | 8 (7-9) | 1 (1-1) | 9.62 (7.56-12.01) | 0 (0-1) | 6.03 (4.64-7.80) | 0 (0-1) | 4.88 (3.72-6.23) | 0 (0-0) | 1.92 (1.40-2.65) | 0 (0-0) | 1.14 (0.75-1.66) |
| Nauru | Male | 7 (7-8) | 1 (1-1) | 11.28 (8.76-13.71) | 1 (0-1) | 7.03 (5.39-8.71) | 0 (0-1) | 5.65 (4.24-7.14) | 0 (0-0) | 2.02 (1.51-2.71) | 0 (0-0) | 1.37 (0.94-1.85) |
| Nepal | Female | 26913 (24245-29618) | 1945 (1153-2708) | 7.28 (4.31-9.81) | 1371 (805-1952) | 5.13 (2.98-7.13) | 831 (489-1248) | 3.13 (1.81-4.50) | 235 (132-341) | 0.88 (0.49-1.26) | 163 (90-238) | 0.61 (0.34-0.88) |
| Nepal | Male | 22009 (19868-24058) | 2303 (1725-2879) | 10.46 (8.00-12.90) | 1610 (1182-2065) | 7.32 (5.48-9.29) | 974 (696-1261) | 4.41 (3.16-5.64) | 251 (177-341) | 1.14 (0.82-1.52) | 145 (99-199) | 0.66 (0.46-0.89) |
| Netherlands | Female | 50850 (45294-56792) | 3218 (2003-4662) | 6.32 (3.88-8.93) | 970 (559-1537) | 1.92 (1.12-2.92) | 2849 (1791-3996) | 5.59 (3.51-7.70) | 367 (192-608) | 0.72 (0.39-1.18) | 310 (162-518) | 0.61 (0.32-0.99) |
| Netherlands | Male | 37176 (33259-40673) | 4053 (2981-5081) | 10.97 (8.03-13.34) | 1060 (648-1605) | 2.86 (1.74-4.28) | 3318 (2403-4259) | 8.95 (6.48-11.30) | 415 (236-649) | 1.11 (0.64-1.75) | 383 (219-611) | 1.03 (0.59-1.61) |
| New Zealand | Female | 14044 (12569-15368) | 961 (617-1296) | 6.87 (4.50-9.15) | 342 (213-504) | 2.46 (1.52-3.54) | 769 (492-1049) | 5.51 (3.64-7.43) | 137 (78-210) | 0.98 (0.58-1.48) | 120 (70-188) | 0.86 (0.50-1.34) |
| New Zealand | Male | 11049 (9901-12167) | 1207 (906-1507) | 10.95 (8.36-13.51) | 394 (274-569) | 3.56 (2.46-5.10) | 926 (674-1187) | 8.38 (6.14-10.59) | 152 (102-225) | 1.37 (0.92-2.03) | 136 (89-208) | 1.23 (0.80-1.84) |
| Nicaragua | Female | 9484 (8649-10352) | 1021 (777-1246) | 10.77 (8.52-12.88) | 707 (525-888) | 7.44 (5.68-9.20) | 433 (324-561) | 4.57 (3.43-5.86) | 275 (199-368) | 2.90 (2.14-3.76) | 253 (183-340) | 2.68 (1.97-3.57) |
| Nicaragua | Male | 12572 (11431-13726) | 1759 (1351-2167) | 13.98 (11.11-16.95) | 1179 (891-1486) | 9.36 (7.39-11.70) | 727 (547-920) | 5.76 (4.45-7.22) | 264 (188-362) | 2.10 (1.53-2.79) | 238 (167-328) | 1.88 (1.35-2.55) |
| Niger | Female | 6942 (6266-7586) | 515 (360-675) | 7.44 (5.34-9.39) | 370 (261-485) | 5.31 (3.77-6.82) | 218 (152-293) | 3.15 (2.25-4.10) | 69 (42-101) | 1.00 (0.62-1.41) | 32 (18-53) | 0.46 (0.26-0.72) |
| Niger | Male | 5121 (4656-5639) | 427 (305-553) | 8.35 (6.04-10.74) | 303 (215-397) | 5.89 (4.23-7.67) | 179 (128-244) | 3.51 (2.52-4.70) | 37 (22-59) | 0.73 (0.43-1.15) | 25 (13-41) | 0.49 (0.25-0.80) |
| Nigeria | Female | 103881 (95364-113241) | 6325 (4309-8531) | 6.12 (4.18-8.10) | 4681 (3267-6326) | 4.50 (3.22-5.98) | 2953 (2059-4014) | 2.84 (1.99-3.85) | 1199 (794-1677) | 1.15 (0.77-1.63) | 688 (438-999) | 0.66 (0.42-0.94) |
| Nigeria | Male | 56775 (51924-61423) | 4263 (2984-5696) | 7.57 (5.27-9.80) | 3052 (2191-4108) | 5.38 (3.90-7.17) | 1888 (1349-2529) | 3.34 (2.39-4.44) | 621 (410-886) | 1.10 (0.72-1.54) | 438 (284-654) | 0.78 (0.51-1.13) |
| Norway | Female | 12262 (11158-13326) | 469 (236-741) | 3.80 (1.90-6.08) | 64 (34-130) | 0.52 (0.27-1.04) | 464 (232-727) | 3.77 (1.87-5.98) | 26 (13-58) | 0.22 (0.10-0.47) | 18 (8-41) | 0.15 (0.07-0.33) |
| Norway | Male | 9425 (8680-10112) | 1081 (823-1323) | 11.50 (8.72-13.90) | 92 (57-184) | 0.98 (0.61-1.95) | 1019 (763-1257) | 10.88 (8.10-13.21) | 42 (24-90) | 0.45 (0.26-0.95) | 37 (21-80) | 0.39 (0.23-0.85) |
| Oman | Female | 4085 (3689-4496) | 21 (4-66) | 0.51 (0.09-1.62) | 18 (3-56) | 0.43 (0.07-1.34) | 18 (3-55) | 0.44 (0.07-1.32) | 4 (0-19) | 0.10 (0.00-0.47) | 3 (0-18) | 0.09 (0.00-0.42) |
| Oman | Male | 4526 (4126-4935) | 251 (164-334) | 5.54 (3.62-7.35) | 159 (106-215) | 3.52 (2.36-4.69) | 158 (105-214) | 3.48 (2.30-4.68) | 33 (20-49) | 0.74 (0.44-1.08) | 29 (17-43) | 0.64 (0.38-0.95) |
| Pakistan | Female | 119242 (107862-130483) | 12187 (9516-14977) | 10.26 (8.14-12.19) | 8533 (6548-10647) | 7.16 (5.53-8.78) | 5154 (3905-6601) | 4.34 (3.30-5.45) | 2400 (1778-3163) | 2.01 (1.54-2.63) | 1986 (1463-2656) | 1.67 (1.23-2.24) |
| Pakistan | Male | 105106 (95801-114625) | 11473 (9070-13893) | 10.91 (8.89-12.93) | 7990 (6245-9945) | 7.61 (6.01-9.31) | 4854 (3703-6244) | 4.61 (3.54-5.82) | 1323 (955-1771) | 1.26 (0.93-1.66) | 996 (713-1378) | 0.96 (0.68-1.30) |
| Palau | Female | 34 (29-38) | 4 (3-5) | 10.57 (7.85-13.37) | 2 (2-3) | 6.47 (4.64-8.42) | 2 (1-2) | 5.37 (3.96-7.02) | 1 (1-1) | 2.43 (1.67-3.29) | 1 (0-1) | 2.02 (1.39-2.77) |
| Palau | Male | 45 (40-50) | 6 (4-7) | 12.52 (9.32-15.51) | 3 (2-5) | 7.59 (5.61-9.66) | 3 (2-4) | 6.31 (4.59-8.09) | 1 (1-1) | 2.36 (1.70-3.17) | 1 (1-1) | 1.74 (1.21-2.41) |
| Panama | Female | 7485 (6757-8119) | 829 (640-1014) | 11.16 (8.63-13.23) | 497 (373-628) | 6.60 (5.04-8.34) | 440 (321-561) | 5.89 (4.40-7.38) | 204 (148-272) | 2.73 (2.01-3.59) | 181 (129-242) | 2.40 (1.79-3.19) |
| Panama | Male | 7704 (7006-8442) | 1114 (855-1370) | 14.46 (11.40-17.48) | 643 (491-809) | 8.37 (6.55-10.33) | 570 (430-719) | 7.39 (5.82-9.23) | 201 (148-263) | 2.61 (1.95-3.37) | 167 (119-223) | 2.16 (1.61-2.86) |
| Papua New Guinea | Female | 3033 (2702-3339) | 218 (148-289) | 7.25 (4.81-9.46) | 154 (103-207) | 5.07 (3.36-6.76) | 97 (65-134) | 3.21 (2.15-4.37) | 26 (15-40) | 0.86 (0.49-1.28) | 15 (8-25) | 0.51 (0.26-0.82) |
| Papua New Guinea | Male | 3658 (3351-3982) | 335 (234-441) | 9.11 (6.44-11.85) | 233 (162-315) | 6.37 (4.44-8.46) | 147 (99-202) | 4.04 (2.75-5.41) | 27 (15-41) | 0.73 (0.42-1.11) | 18 (9-29) | 0.49 (0.25-0.78) |
| Paraguay | Female | 9655 (8669-10562) | 999 (752-1231) | 10.40 (8.07-12.49) | 662 (483-840) | 6.90 (5.21-8.52) | 452 (332-582) | 4.73 (3.53-5.90) | 308 (222-402) | 3.20 (2.36-4.15) | 246 (172-334) | 2.57 (1.83-3.44) |
| Paraguay | Male | 8132 (7366-8993) | 1027 (782-1290) | 12.67 (9.90-15.35) | 674 (500-860) | 8.31 (6.28-10.21) | 463 (344-600) | 5.69 (4.34-7.19) | 246 (177-327) | 3.02 (2.22-3.91) | 190 (133-254) | 2.31 (1.64-3.08) |
| Peru | Female | 54931 (49330-60512) | 5193 (3663-6520) | 9.52 (6.94-11.72) | 3423 (2398-4414) | 6.22 (4.48-7.97) | 2401 (1648-3160) | 4.39 (3.08-5.71) | 824 (571-1094) | 1.51 (1.06-1.98) | 744 (502-995) | 1.37 (0.94-1.79) |
| Peru | Male | 35475 (32386-38539) | 4303 (3260-5415) | 12.17 (9.36-15.06) | 2807 (2113-3587) | 7.94 (5.96-9.94) | 1968 (1471-2543) | 5.54 (4.18-7.13) | 514 (381-683) | 1.45 (1.09-1.90) | 437 (330-585) | 1.23 (0.94-1.62) |
| Philippines | Female | 127216 (116688-137645) | 16204 (11978-20790) | 12.78 (9.46-16.04) | 10805 (7855-14150) | 8.50 (6.15-11.04) | 6940 (4922-9078) | 5.46 (3.94-7.06) | 3189 (2268-4251) | 2.51 (1.77-3.36) | 2377 (1629-3231) | 1.87 (1.31-2.48) |
| Philippines | Male | 94664 (87919-102635) | 13648 (9992-16966) | 14.40 (10.57-17.67) | 9172 (6538-11742) | 9.70 (6.98-12.15) | 5908 (4190-7673) | 6.22 (4.44-8.05) | 2006 (1368-2681) | 2.11 (1.42-2.76) | 1363 (882-1827) | 1.44 (0.94-1.92) |
| Poland | Female | 84176 (75720-92628) | 11506 (8930-14306) | 13.64 (10.74-16.76) | 6523 (4876-8407) | 7.78 (6.00-9.84) | 6199 (4601-8038) | 7.37 (5.56-9.52) | 3817 (2804-5024) | 4.55 (3.38-5.88) | 3678 (2691-4850) | 4.39 (3.25-5.68) |
| Poland | Male | 51982 (47085-56962) | 9242 (7250-11506) | 17.83 (14.32-21.28) | 5227 (3971-6745) | 10.03 (7.79-12.61) | 4957 (3879-6351) | 9.54 (7.54-11.82) | 2599 (1942-3405) | 4.99 (3.84-6.41) | 2399 (1764-3161) | 4.59 (3.49-5.95) |
| Portugal | Female | 37305 (33276-41551) | 1983 (1144-2916) | 5.31 (3.07-7.73) | 1251 (730-1869) | 3.36 (1.99-4.92) | 1395 (798-2045) | 3.74 (2.16-5.35) | 618 (326-990) | 1.66 (0.88-2.55) | 600 (316-942) | 1.61 (0.84-2.51) |
| Portugal | Male | 22539 (20039-24912) | 2415 (1832-3033) | 10.79 (8.19-13.19) | 1283 (934-1645) | 5.73 (4.22-7.28) | 1448 (1070-1874) | 6.44 (4.83-8.08) | 612 (416-821) | 2.74 (1.88-3.58) | 580 (404-778) | 2.61 (1.83-3.39) |
| Puerto Rico | Female | 11644 (10234-12874) | 834 (564-1093) | 7.16 (4.93-9.32) | 416 (285-564) | 3.60 (2.47-4.76) | 573 (397-776) | 4.96 (3.47-6.56) | 193 (121-280) | 1.66 (1.09-2.38) | 165 (106-245) | 1.43 (0.92-2.06) |
| Puerto Rico | Male | 9217 (8248-10137) | 958 (720-1198) | 10.44 (8.03-12.74) | 447 (332-589) | 4.83 (3.64-6.28) | 628 (458-801) | 6.82 (5.07-8.64) | 154 (106-218) | 1.65 (1.16-2.35) | 126 (86-182) | 1.36 (0.93-1.98) |
| Qatar | Female | 2150 (1896-2409) | 7 (0-29) | 0.34 (0.00-1.31) | 3 (0-14) | 0.15 (0.00-0.62) | 7 (0-28) | 0.33 (0.00-1.31) | 1 (0-6) | 0.05 (0.00-0.26) | 1 (0-5) | 0.04 (0.00-0.24) |
| Qatar | Male | 3744 (3349-4134) | 192 (116-279) | 5.14 (3.13-7.41) | 45 (24-76) | 1.21 (0.66-1.99) | 173 (107-249) | 4.64 (2.85-6.63) | 11 (5-19) | 0.29 (0.15-0.52) | 10 (5-17) | 0.26 (0.13-0.46) |
| Romania | Female | 39727 (36071-43529) | 6692 (5132-8144) | 16.79 (13.44-20.23) | 3885 (2912-4972) | 9.74 (7.57-12.23) | 3404 (2531-4302) | 8.54 (6.58-10.80) | 1989 (1463-2619) | 5.00 (3.85-6.52) | 1776 (1309-2357) | 4.45 (3.36-5.82) |
| Romania | Male | 32187 (28893-34962) | 6774 (5364-8062) | 21.10 (17.25-24.49) | 4094 (3136-5130) | 12.77 (9.97-15.57) | 3585 (2783-4543) | 11.16 (8.75-13.73) | 1885 (1413-2448) | 5.90 (4.44-7.41) | 1687 (1242-2235) | 5.28 (4.01-6.77) |
| Russia | Female | 242006 (224167-264492) | 24636 (18744-30215) | 10.14 (7.79-12.31) | 15218 (11188-19362) | 6.27 (4.71-7.89) | 12393 (9258-15940) | 5.12 (3.80-6.51) | 7093 (5090-9160) | 2.93 (2.11-3.75) | 5167 (3681-6901) | 2.13 (1.53-2.81) |
| Russia | Male | 164084 (150263-178479) | 23378 (17971-28544) | 14.26 (11.11-17.02) | 13988 (10626-17624) | 8.60 (6.61-10.50) | 11489 (8669-14607) | 7.03 (5.32-8.75) | 5419 (4049-6951) | 3.31 (2.51-4.18) | 3794 (2782-5034) | 2.31 (1.70-3.00) |
| Rwanda | Female | 4278 (3869-4707) | 512 (391-641) | 12.05 (9.06-14.62) | 354 (263-451) | 8.31 (6.16-10.27) | 211 (155-276) | 4.95 (3.67-6.28) | 51 (33-72) | 1.19 (0.79-1.66) | 23 (13-38) | 0.55 (0.30-0.87) |
| Rwanda | Male | 2877 (2587-3152) | 267 (190-344) | 9.31 (6.71-11.77) | 189 (133-249) | 6.59 (4.71-8.50) | 114 (77-152) | 3.96 (2.70-5.24) | 18 (11-29) | 0.64 (0.39-0.98) | 11 (6-18) | 0.39 (0.22-0.64) |
| Saint Kitts and Nevis | Female | 118 (105-131) | 7 (5-10) | 6.27 (4.00-8.46) | 5 (3-6) | 3.90 (2.58-5.27) | 4 (3-6) | 3.72 (2.46-5.01) | 2 (1-3) | 1.73 (1.10-2.50) | 2 (1-3) | 1.55 (0.96-2.27) |
| Saint Kitts and Nevis | Male | 127 (113-141) | 13 (9-16) | 9.97 (7.25-12.18) | 7 (5-9) | 5.71 (4.09-7.33) | 7 (5-9) | 5.45 (3.84-7.04) | 2 (1-3) | 1.71 (1.17-2.33) | 2 (1-3) | 1.35 (0.90-1.95) |
| Saint Lucia | Female | 378 (340-419) | 24 (16-32) | 6.40 (4.41-8.45) | 16 (11-22) | 4.33 (3.03-5.66) | 13 (9-17) | 3.42 (2.40-4.55) | 8 (5-11) | 2.05 (1.41-2.82) | 7 (4-9) | 1.75 (1.16-2.46) |
| Saint Lucia | Male | 335 (302-368) | 33 (24-41) | 9.78 (7.39-11.94) | 21 (15-27) | 6.13 (4.64-7.85) | 16 (12-21) | 4.84 (3.61-6.20) | 6 (5-9) | 1.93 (1.38-2.59) | 5 (3-7) | 1.40 (0.98-1.90) |
| Saint Vincent and the Grenadines | Female | 201 (179-221) | 13 (8-17) | 6.37 (4.05-8.43) | 9 (6-12) | 4.40 (2.91-5.85) | 7 (4-9) | 3.25 (2.21-4.43) | 4 (2-5) | 1.96 (1.27-2.72) | 3 (2-5) | 1.62 (1.06-2.29) |
| Saint Vincent and the Grenadines | Male | 193 (175-211) | 19 (14-24) | 9.77 (7.46-12.09) | 12 (9-16) | 6.30 (4.72-8.09) | 9 (7-12) | 4.63 (3.47-6.06) | 4 (2-5) | 1.86 (1.29-2.51) | 3 (2-4) | 1.41 (0.99-1.98) |
| Samoa | Female | 191 (169-210) | 5 (3-7) | 2.54 (1.49-3.79) | 4 (2-6) | 1.90 (1.08-2.92) | 3 (1-4) | 1.35 (0.75-2.12) | 1 (0-1) | 0.45 (0.22-0.77) | 0 (0-1) | 0.18 (0.07-0.35) |
| Samoa | Male | 189 (170-207) | 7 (5-11) | 3.93 (2.50-5.64) | 5 (3-8) | 2.91 (1.80-4.25) | 4 (2-6) | 1.98 (1.23-2.93) | 1 (1-2) | 0.59 (0.34-0.93) | 1 (0-1) | 0.39 (0.21-0.67) |
| Sao Tome and Principe | Female | 166 (148-184) | 13 (10-17) | 7.82 (5.83-9.69) | 9 (7-12) | 5.50 (4.19-6.96) | 6 (4-8) | 3.43 (2.53-4.39) | 3 (2-4) | 1.85 (1.33-2.44) | 2 (1-3) | 1.18 (0.81-1.63) |
| Sao Tome and Principe | Male | 93 (84-102) | 8 (6-10) | 8.74 (6.45-10.95) | 6 (4-7) | 6.12 (4.51-7.84) | 3 (2-5) | 3.76 (2.68-4.96) | 1 (1-2) | 1.31 (0.87-1.84) | 1 (0-1) | 0.78 (0.48-1.12) |
| Saudi Arabia | Female | 54361 (48382-60498) | 215 (7-633) | 0.40 (0.01-1.14) | 179 (7-517) | 0.33 (0.01-0.96) | 188 (7-545) | 0.35 (0.01-1.00) | 28 (0-147) | 0.05 (0.00-0.27) | 23 (0-123) | 0.04 (0.00-0.23) |
| Saudi Arabia | Male | 62906 (56534-69134) | 3115 (1961-4657) | 4.96 (3.15-7.22) | 1884 (1190-2695) | 3.00 (1.92-4.23) | 2100 (1363-2987) | 3.34 (2.16-4.69) | 397 (228-612) | 0.64 (0.36-0.96) | 315 (184-510) | 0.51 (0.30-0.78) |
| Senegal | Female | 7225 (6574-7895) | 558 (401-713) | 7.75 (5.67-9.80) | 395 (283-516) | 5.49 (4.07-7.03) | 243 (176-321) | 3.34 (2.45-4.42) | 103 (70-145) | 1.43 (0.97-1.97) | 55 (34-85) | 0.77 (0.47-1.17) |
| Senegal | Male | 5346 (4862-5810) | 452 (322-587) | 8.45 (6.01-10.79) | 315 (224-423) | 5.92 (4.26-7.85) | 192 (136-261) | 3.59 (2.55-4.91) | 60 (38-91) | 1.12 (0.72-1.70) | 34 (20-56) | 0.63 (0.37-1.06) |
| Serbia | Female | 21221 (18842-23535) | 3495 (2760-4384) | 16.51 (13.27-20.13) | 2173 (1688-2863) | 10.27 (8.10-13.06) | 1645 (1248-2098) | 7.79 (5.98-9.80) | 1223 (911-1656) | 5.75 (4.42-7.49) | 1150 (840-1552) | 5.42 (4.10-7.06) |
| Serbia | Male | 17560 (15882-19292) | 3714 (3047-4517) | 21.22 (17.57-24.81) | 2405 (1897-3013) | 13.71 (10.96-16.80) | 1814 (1386-2350) | 10.37 (8.02-12.95) | 1128 (842-1484) | 6.43 (4.92-8.31) | 1026 (774-1349) | 5.86 (4.48-7.59) |
| Seychelles | Female | 203 (181-225) | 27 (20-33) | 13.23 (10.01-16.17) | 16 (12-20) | 7.90 (5.92-9.96) | 13 (10-18) | 6.55 (4.86-8.41) | 8 (6-10) | 3.82 (2.81-5.00) | 7 (5-9) | 3.47 (2.53-4.51) |
| Seychelles | Male | 162 (144-179) | 23 (17-29) | 14.22 (10.91-17.45) | 14 (10-18) | 8.51 (6.37-10.99) | 11 (8-15) | 7.09 (5.24-9.10) | 5 (4-7) | 3.24 (2.36-4.34) | 5 (3-6) | 2.85 (2.04-3.87) |
| Sierra Leone | Female | 3490 (3184-3820) | 269 (192-340) | 7.68 (5.60-9.63) | 192 (141-250) | 5.52 (4.09-7.04) | 113 (84-149) | 3.25 (2.42-4.18) | 46 (32-66) | 1.33 (0.91-1.85) | 27 (17-42) | 0.78 (0.49-1.16) |
| Sierra Leone | Male | 2425 (2179-2642) | 211 (155-274) | 8.78 (6.54-11.12) | 150 (109-201) | 6.23 (4.60-8.09) | 89 (65-118) | 3.68 (2.70-4.82) | 25 (16-39) | 1.05 (0.68-1.57) | 15 (9-23) | 0.61 (0.36-0.98) |
| Singapore | Female | 13353 (11941-14598) | 1675 (1138-2208) | 12.61 (8.59-16.35) | 237 (117-426) | 1.78 (0.88-3.14) | 1505 (1014-2028) | 11.35 (7.69-14.91) | 105 (48-196) | 0.79 (0.37-1.47) | 100 (46-190) | 0.76 (0.35-1.40) |
| Singapore | Male | 11706 (10587-12777) | 1750 (1219-2223) | 14.97 (10.51-18.76) | 249 (123-445) | 2.12 (1.07-3.71) | 1576 (1078-2045) | 13.45 (9.24-17.22) | 107 (52-195) | 0.92 (0.45-1.63) | 101 (49-185) | 0.87 (0.42-1.57) |
| Slovakia | Female | 11711 (10446-12928) | 1914 (1473-2375) | 16.34 (12.78-19.67) | 1013 (772-1316) | 8.69 (6.60-10.99) | 1071 (807-1371) | 9.15 (7.12-11.48) | 536 (399-716) | 4.59 (3.40-6.05) | 503 (366-676) | 4.30 (3.14-5.66) |
| Slovakia | Male | 8722 (7766-9628) | 1802 (1436-2224) | 20.76 (16.95-24.25) | 1000 (766-1303) | 11.50 (8.93-14.42) | 1048 (813-1365) | 12.04 (9.42-15.04) | 490 (360-659) | 5.63 (4.17-7.42) | 451 (326-615) | 5.17 (3.84-6.83) |
| Slovenia | Female | 4951 (4438-5492) | 866 (661-1063) | 17.43 (13.78-20.98) | 413 (308-537) | 8.32 (6.34-10.55) | 528 (399-678) | 10.67 (8.23-13.58) | 215 (156-302) | 4.38 (3.23-6.01) | 195 (136-269) | 3.94 (2.83-5.30) |
| Slovenia | Male | 4183 (3693-4603) | 903 (712-1093) | 21.72 (17.64-25.30) | 451 (339-593) | 10.86 (8.29-13.79) | 574 (435-735) | 13.78 (10.82-17.15) | 201 (145-279) | 4.86 (3.53-6.50) | 173 (119-250) | 4.18 (2.92-5.80) |
| Solomon Islands | Female | 314 (283-345) | 21 (14-27) | 6.64 (4.57-8.67) | 14 (10-19) | 4.63 (3.10-6.17) | 9 (6-13) | 2.91 (1.90-4.02) | 3 (2-5) | 1.08 (0.70-1.52) | 1 (1-2) | 0.44 (0.26-0.68) |
| Solomon Islands | Male | 311 (282-341) | 27 (18-35) | 8.71 (5.95-11.25) | 19 (13-25) | 6.05 (4.06-8.05) | 12 (8-16) | 3.81 (2.53-5.15) | 2 (1-3) | 0.66 (0.40-0.98) | 1 (1-2) | 0.33 (0.19-0.55) |
| Somalia | Female | 3803 (3409-4193) | 421 (315-534) | 11.13 (8.28-13.71) | 292 (212-377) | 7.78 (5.63-9.74) | 173 (122-228) | 4.56 (3.31-5.92) | 75 (51-105) | 1.99 (1.38-2.69) | 53 (34-78) | 1.40 (0.90-2.00) |
| Somalia | Male | 2760 (2467-3032) | 229 (163-295) | 8.31 (5.90-10.55) | 164 (116-216) | 5.94 (4.25-7.67) | 98 (67-130) | 3.58 (2.44-4.68) | 31 (19-45) | 1.11 (0.70-1.64) | 22 (13-35) | 0.80 (0.48-1.24) |
| South Africa | Female | 59036 (54213-63859) | 2913 (1876-4050) | 4.94 (3.19-6.87) | 2276 (1522-3081) | 3.87 (2.54-5.18) | 1706 (1160-2288) | 2.89 (1.95-3.85) | 885 (556-1234) | 1.50 (0.94-2.07) | 816 (498-1153) | 1.38 (0.85-1.95) |
| South Africa | Male | 51678 (47501-55344) | 2225 (1279-3366) | 4.31 (2.49-6.40) | 1708 (1008-2536) | 3.30 (1.96-4.92) | 1289 (786-1871) | 2.50 (1.55-3.62) | 424 (216-705) | 0.82 (0.41-1.35) | 386 (189-655) | 0.75 (0.37-1.24) |
| South Korea | Female | 130200 (117572-143489) | 21034 (15216-26490) | 16.34 (11.95-19.76) | 9271 (6538-12253) | 7.15 (5.08-9.22) | 13859 (9861-18064) | 10.72 (7.77-13.58) | 4450 (3076-5992) | 3.45 (2.35-4.54) | 4444 (3054-5990) | 3.44 (2.35-4.51) |
| South Korea | Male | 95092 (87805-102602) | 15593 (10781-19476) | 16.31 (11.32-20.21) | 6955 (4789-9221) | 7.30 (5.11-9.63) | 10289 (7048-13338) | 10.84 (7.55-13.76) | 3061 (2104-4158) | 3.25 (2.24-4.34) | 3058 (2099-4161) | 3.23 (2.22-4.32) |
| Spain | Female | 143887 (127055-159276) | 2874 (1265-5732) | 1.99 (0.87-3.99) | 2021 (761-4105) | 1.42 (0.52-2.80) | 2376 (941-4677) | 1.65 (0.65-3.31) | 799 (188-1858) | 0.56 (0.13-1.29) | 691 (159-1634) | 0.49 (0.11-1.13) |
| Spain | Male | 90952 (81795-100066) | 3609 (2183-5468) | 3.98 (2.45-6.00) | 2071 (1160-3264) | 2.29 (1.32-3.60) | 2606 (1503-4129) | 2.86 (1.66-4.40) | 786 (378-1399) | 0.87 (0.42-1.49) | 709 (332-1290) | 0.79 (0.37-1.41) |
| Sri Lanka | Female | 44051 (39488-48754) | 6671 (5222-8274) | 15.18 (12.24-18.32) | 4386 (3371-5461) | 9.95 (7.83-12.27) | 2841 (2198-3705) | 6.45 (5.01-8.27) | 1417 (1020-1875) | 3.22 (2.38-4.25) | 1207 (854-1608) | 2.74 (1.99-3.60) |
| Sri Lanka | Male | 30501 (27133-33854) | 5060 (3964-6238) | 16.64 (13.34-20.15) | 3339 (2582-4290) | 11.00 (8.73-13.70) | 2193 (1611-2833) | 7.18 (5.52-9.11) | 719 (513-1007) | 2.36 (1.72-3.21) | 578 (399-810) | 1.91 (1.34-2.55) |
| Sudan | Female | 35228 (31799-38550) | 197 (53-527) | 0.56 (0.16-1.54) | 181 (48-495) | 0.52 (0.14-1.39) | 145 (33-373) | 0.41 (0.09-1.05) | 27 (3-120) | 0.08 (0.01-0.33) | 20 (1-94) | 0.06 (0.00-0.27) |
| Sudan | Male | 30363 (27675-33073) | 1564 (968-2210) | 5.20 (3.15-7.17) | 1221 (771-1720) | 4.03 (2.60-5.56) | 755 (496-1060) | 2.51 (1.68-3.47) | 169 (92-265) | 0.55 (0.30-0.87) | 123 (62-206) | 0.41 (0.21-0.67) |
| Suriname | Female | 973 (873-1067) | 58 (36-79) | 5.98 (3.85-8.04) | 42 (27-56) | 4.27 (2.76-5.75) | 28 (18-39) | 2.90 (1.91-3.91) | 19 (12-26) | 1.93 (1.22-2.70) | 16 (10-23) | 1.62 (1.02-2.35) |
| Suriname | Male | 856 (778-947) | 78 (57-100) | 9.12 (6.59-11.43) | 52 (37-70) | 6.13 (4.37-7.93) | 35 (25-48) | 4.10 (2.94-5.40) | 17 (12-24) | 2.01 (1.41-2.72) | 14 (10-20) | 1.66 (1.13-2.28) |
| Swaziland | Female | 671 (609-737) | 41 (27-55) | 6.11 (4.15-8.25) | 31 (20-40) | 4.56 (3.13-5.95) | 21 (14-28) | 3.05 (2.08-4.05) | 10 (6-14) | 1.50 (0.96-2.09) | 8 (4-11) | 1.12 (0.66-1.61) |
| Swaziland | Male | 687 (623-747) | 42 (28-57) | 6.07 (4.08-8.14) | 30 (20-42) | 4.43 (2.96-6.05) | 20 (14-28) | 2.95 (1.98-4.03) | 4 (3-7) | 0.64 (0.37-1.01) | 3 (2-5) | 0.48 (0.26-0.78) |
| Sweden | Female | 21050 (18791-23371) | 1529 (993-2056) | 7.32 (4.83-9.61) | 393 (229-629) | 1.89 (1.13-2.91) | 1334 (894-1812) | 6.35 (4.24-8.40) | 156 (86-260) | 0.75 (0.42-1.20) | 127 (69-220) | 0.62 (0.34-1.02) |
| Sweden | Male | 17283 (15550-19001) | 2066 (1598-2613) | 12.05 (9.42-14.54) | 493 (313-743) | 2.86 (1.82-4.25) | 1728 (1301-2213) | 10.06 (7.64-12.44) | 204 (124-321) | 1.18 (0.72-1.87) | 179 (108-287) | 1.04 (0.64-1.66) |
| Switzerland | Female | 28202 (24941-31654) | 1741 (1017-2569) | 6.19 (3.72-8.96) | 171 (95-368) | 0.61 (0.33-1.33) | 1673 (977-2488) | 5.97 (3.59-8.49) | 62 (33-145) | 0.22 (0.12-0.51) | 51 (27-125) | 0.18 (0.10-0.44) |
| Switzerland | Male | 18944 (17016-20700) | 2149 (1537-2685) | 11.32 (8.32-13.93) | 185 (112-351) | 0.97 (0.60-1.89) | 2025 (1449-2559) | 10.67 (7.83-13.19) | 84 (49-174) | 0.44 (0.26-0.93) | 73 (43-153) | 0.39 (0.23-0.82) |
| Syria | Female | 31230 (28023-34536) | 105 (9-478) | 0.34 (0.03-1.49) | 102 (7-464) | 0.33 (0.02-1.48) | 90 (6-359) | 0.29 (0.02-1.16) | 31 (0-191) | 0.10 (0.00-0.62) | 27 (0-177) | 0.09 (0.00-0.57) |
| Syria | Male | 23087 (20664-25333) | 1155 (660-1721) | 5.04 (2.87-7.35) | 907 (547-1329) | 3.95 (2.39-5.65) | 561 (351-811) | 2.45 (1.52-3.49) | 279 (147-452) | 1.22 (0.64-1.93) | 235 (122-384) | 1.02 (0.53-1.64) |
| Taiwan (Province of China) | Female | 67866 (62906-72532) | 8606 (6395-10743) | 12.67 (9.55-15.69) | 3914 (2796-5123) | 5.77 (4.13-7.52) | 5534 (4127-7232) | 8.18 (6.08-10.45) | 1256 (864-1767) | 1.86 (1.27-2.56) | 1247 (853-1772) | 1.85 (1.27-2.56) |
| Taiwan (Province of China) | Male | 55052 (51272-58546) | 6983 (5302-8750) | 12.70 (9.77-15.82) | 3157 (2312-4223) | 5.76 (4.25-7.56) | 4527 (3353-5828) | 8.22 (6.02-10.45) | 1081 (774-1495) | 1.96 (1.40-2.70) | 1045 (745-1446) | 1.90 (1.36-2.61) |
| Tajikistan | Female | 3844 (3442-4244) | 272 (183-357) | 7.09 (4.87-9.19) | 199 (131-262) | 5.13 (3.49-6.72) | 120 (81-162) | 3.10 (2.17-4.17) | 59 (39-84) | 1.55 (1.05-2.12) | 48 (31-68) | 1.26 (0.84-1.74) |
| Tajikistan | Male | 3429 (3116-3750) | 425 (337-524) | 12.45 (10.00-15.09) | 294 (228-371) | 8.59 (6.75-10.56) | 176 (135-224) | 5.12 (4.01-6.43) | 78 (57-103) | 2.26 (1.71-2.93) | 48 (34-67) | 1.40 (1.00-1.91) |
| Tanzania | Female | 17573 (15819-19160) | 2272 (1742-2821) | 12.97 (10.08-15.37) | 1557 (1163-1930) | 8.90 (6.71-10.71) | 931 (699-1187) | 5.32 (4.05-6.64) | 341 (235-450) | 1.94 (1.38-2.50) | 146 (98-211) | 0.84 (0.56-1.19) |
| Tanzania | Male | 12976 (11815-14216) | 1701 (1234-2177) | 13.16 (9.71-16.43) | 1164 (830-1511) | 8.99 (6.48-11.25) | 702 (500-924) | 5.42 (3.88-7.05) | 166 (108-240) | 1.28 (0.86-1.84) | 102 (62-152) | 0.79 (0.48-1.13) |
| Thailand | Female | 175644 (158430-195476) | 23078 (15284-29978) | 13.16 (8.80-16.77) | 14793 (9670-19512) | 8.39 (5.56-10.95) | 10520 (6986-14164) | 6.00 (4.02-7.96) | 4785 (3088-6687) | 2.71 (1.80-3.70) | 4298 (2701-6042) | 2.44 (1.58-3.33) |
| Thailand | Male | 135587 (121664-149991) | 21599 (15475-27025) | 15.90 (11.72-19.24) | 13886 (9870-18168) | 10.25 (7.50-13.12) | 9935 (6877-13242) | 7.32 (5.15-9.43) | 3556 (2359-5145) | 2.64 (1.80-3.71) | 3083 (2029-4471) | 2.30 (1.52-3.25) |
| The Bahamas | Female | 654 (587-721) | 40 (26-55) | 6.12 (3.99-8.25) | 22 (15-30) | 3.34 (2.31-4.53) | 27 (18-36) | 4.10 (2.81-5.47) | 9 (6-13) | 1.38 (0.89-1.93) | 8 (5-11) | 1.21 (0.79-1.72) |
| The Bahamas | Male | 536 (484-592) | 52 (38-65) | 9.62 (7.27-11.78) | 26 (19-34) | 4.87 (3.55-6.25) | 32 (23-42) | 6.01 (4.40-7.62) | 9 (6-12) | 1.63 (1.15-2.23) | 7 (5-10) | 1.29 (0.91-1.80) |
| The Gambia | Female | 999 (909-1093) | 76 (57-95) | 7.56 (5.75-9.38) | 54 (40-69) | 5.42 (4.07-6.78) | 32 (24-42) | 3.24 (2.45-4.20) | 15 (11-21) | 1.54 (1.07-2.07) | 10 (6-14) | 0.99 (0.64-1.40) |
| The Gambia | Male | 678 (613-741) | 58 (41-75) | 8.57 (6.09-10.81) | 41 (29-54) | 6.08 (4.34-7.83) | 24 (17-33) | 3.61 (2.56-4.76) | 8 (5-13) | 1.25 (0.81-1.84) | 6 (4-9) | 0.86 (0.53-1.31) |
| Timor-Leste | Female | 979 (884-1083) | 135 (105-171) | 13.83 (10.89-17.01) | 90 (69-116) | 9.19 (7.20-11.54) | 57 (43-74) | 5.81 (4.43-7.38) | 17 (12-24) | 1.72 (1.21-2.38) | 11 (8-17) | 1.16 (0.79-1.69) |
| Timor-Leste | Male | 688 (625-754) | 113 (91-138) | 16.48 (13.44-19.49) | 76 (60-95) | 10.99 (8.77-13.50) | 48 (36-62) | 6.91 (5.37-8.79) | 13 (10-19) | 1.95 (1.43-2.68) | 9 (6-12) | 1.28 (0.87-1.77) |
| Togo | Female | 4079 (3662-4459) | 299 (209-385) | 7.33 (5.25-9.31) | 214 (150-280) | 5.24 (3.80-6.75) | 127 (89-172) | 3.15 (2.21-4.16) | 54 (36-75) | 1.32 (0.88-1.82) | 29 (18-45) | 0.71 (0.45-1.07) |
| Togo | Male | 2510 (2283-2732) | 208 (151-270) | 8.28 (6.11-10.52) | 146 (107-195) | 5.82 (4.29-7.62) | 87 (62-121) | 3.50 (2.51-4.70) | 24 (15-37) | 0.95 (0.58-1.43) | 11 (7-19) | 0.46 (0.27-0.74) |
| Tonga | Female | 104 (93-114) | 11 (9-14) | 10.94 (8.66-13.35) | 8 (6-10) | 7.36 (5.73-9.13) | 5 (4-7) | 4.88 (3.70-6.19) | 2 (2-3) | 2.31 (1.68-3.05) | 2 (1-3) | 1.88 (1.36-2.52) |
| Tonga | Male | 98 (88-107) | 13 (10-16) | 13.27 (10.44-16.00) | 9 (6-11) | 8.83 (6.74-10.89) | 6 (4-7) | 5.79 (4.49-7.32) | 2 (1-2) | 1.81 (1.28-2.38) | 1 (1-2) | 1.46 (1.03-2.00) |
| Trinidad and Tobago | Female | 2842 (2550-3140) | 193 (130-256) | 6.81 (4.57-8.81) | 121 (85-163) | 4.25 (3.01-5.59) | 109 (76-145) | 3.85 (2.74-5.10) | 50 (34-71) | 1.76 (1.21-2.44) | 41 (27-60) | 1.45 (0.98-2.06) |
| Trinidad and Tobago | Male | 2779 (2488-3066) | 298 (227-371) | 10.80 (8.48-12.90) | 177 (132-231) | 6.38 (4.83-8.13) | 158 (115-208) | 5.71 (4.26-7.24) | 60 (42-81) | 2.13 (1.57-2.86) | 49 (33-67) | 1.76 (1.24-2.36) |
| Tunisia | Female | 31041 (27696-34543) | 99 (11-450) | 0.32 (0.03-1.44) | 94 (9-434) | 0.30 (0.03-1.39) | 86 (7-359) | 0.28 (0.02-1.14) | 25 (0-168) | 0.08 (0.00-0.56) | 20 (0-148) | 0.07 (0.00-0.46) |
| Tunisia | Male | 22101 (19903-24440) | 1039 (577-1625) | 4.71 (2.65-7.18) | 802 (461-1218) | 3.65 (2.09-5.31) | 549 (325-806) | 2.49 (1.48-3.60) | 193 (87-316) | 0.88 (0.41-1.38) | 165 (74-281) | 0.75 (0.35-1.26) |
| Turkey | Female | 216768 (195933-237591) | 2 (0-333) | 0.00 (0.00-0.15) | 2 (0-333) | 0.00 (0.00-0.15) | 2 (0-333) | 0.00 (0.00-0.15) | 0 (0-131) | 0.00 (0.00-0.06) | 0 (0-126) | 0.00 (0.00-0.06) |
| Turkey | Male | 160197 (143198-176439) | 3875 (1720-7351) | 2.44 (1.08-4.47) | 3248 (1488-5894) | 2.03 (0.93-3.57) | 2686 (1182-4878) | 1.68 (0.75-2.98) | 833 (261-1678) | 0.52 (0.16-1.03) | 803 (261-1686) | 0.50 (0.17-1.00) |
| Turkmenistan | Female | 3481 (3086-3875) | 257 (178-339) | 7.35 (5.31-9.62) | 173 (119-228) | 4.98 (3.56-6.69) | 127 (90-170) | 3.65 (2.62-4.81) | 60 (41-83) | 1.72 (1.20-2.39) | 51 (34-71) | 1.46 (1.01-2.02) |
| Turkmenistan | Male | 3210 (2932-3536) | 395 (304-485) | 12.33 (9.66-14.84) | 253 (192-320) | 7.90 (6.11-9.68) | 183 (137-233) | 5.72 (4.36-7.12) | 63 (46-82) | 1.96 (1.45-2.49) | 46 (34-61) | 1.42 (1.06-1.86) |
| Tuvalu | Female | 12 (11-14) | 1 (1-2) | 10.92 (8.43-13.58) | 1 (1-1) | 7.28 (5.54-9.28) | 1 (0-1) | 4.91 (3.67-6.41) | 0 (0-0) | 1.82 (1.29-2.52) | 0 (0-0) | 1.10 (0.74-1.62) |
| Tuvalu | Male | 12 (11-13) | 1 (1-2) | 12.77 (9.90-15.81) | 1 (1-1) | 8.44 (6.41-10.84) | 1 (0-1) | 5.76 (4.43-7.39) | 0 (0-0) | 1.74 (1.23-2.43) | 0 (0-0) | 1.22 (0.79-1.84) |
| Uganda | Female | 8013 (7308-8791) | 972 (751-1200) | 12.13 (9.56-14.75) | 673 (516-845) | 8.41 (6.45-10.36) | 399 (302-510) | 5.01 (3.80-6.24) | 126 (88-171) | 1.57 (1.13-2.10) | 75 (51-111) | 0.94 (0.63-1.34) |
| Uganda | Male | 6565 (5897-7159) | 609 (441-787) | 9.33 (6.85-11.98) | 430 (314-566) | 6.62 (4.82-8.45) | 261 (183-342) | 3.95 (2.83-5.12) | 56 (37-83) | 0.86 (0.57-1.25) | 32 (20-50) | 0.50 (0.31-0.74) |
| Ukraine | Female | 46776 (42139-51169) | 1546 (852-2391) | 3.32 (1.82-5.09) | 1328 (752-1991) | 2.85 (1.60-4.23) | 1000 (604-1461) | 2.13 (1.30-3.10) | 600 (303-951) | 1.28 (0.66-2.05) | 506 (258-819) | 1.07 (0.54-1.74) |
| Ukraine | Male | 32505 (29191-35709) | 3138 (2408-3916) | 9.61 (7.65-11.75) | 2125 (1640-2684) | 6.53 (5.15-8.09) | 1411 (1068-1824) | 4.35 (3.38-5.49) | 672 (473-899) | 2.06 (1.50-2.70) | 468 (327-645) | 1.44 (1.02-1.97) |
| United Arab Emirates | Female | 8495 (7452-9476) | 35 (3-112) | 0.40 (0.04-1.28) | 24 (2-75) | 0.29 (0.02-0.87) | 33 (3-105) | 0.38 (0.03-1.20) | 4 (0-17) | 0.05 (0.00-0.19) | 3 (0-15) | 0.04 (0.00-0.18) |
| United Arab Emirates | Male | 22546 (19994-25136) | 1137 (646-1684) | 5.05 (2.98-7.35) | 485 (284-744) | 2.14 (1.28-3.20) | 902 (514-1323) | 4.00 (2.37-5.76) | 86 (47-136) | 0.38 (0.21-0.59) | 75 (42-122) | 0.34 (0.19-0.53) |
| United Kingdom | Female | 158583 (145358-172882) | 9254 (5239-13456) | 5.84 (3.37-8.46) | 3849 (2365-5584) | 2.42 (1.52-3.48) | 7572 (4332-10940) | 4.78 (2.82-6.94) | 1425 (809-2136) | 0.90 (0.52-1.35) | 1290 (727-1947) | 0.81 (0.47-1.23) |
| United Kingdom | Male | 129257 (118482-139912) | 12876 (8968-16710) | 10.01 (7.07-12.68) | 4592 (3004-6543) | 3.56 (2.34-4.94) | 9676 (6782-12808) | 7.53 (5.35-9.76) | 1822 (1172-2643) | 1.41 (0.93-2.00) | 1656 (1043-2420) | 1.28 (0.83-1.85) |
| United States | Female | 1042155 (960215-1140251) | 86722 (59777-110288) | 8.23 (5.82-10.53) | 16050 (8908-26992) | 1.54 (0.86-2.52) | 76246 (51695-99716) | 7.33 (5.11-9.46) | 7202 (3813-12489) | 0.69 (0.37-1.18) | 6676 (3584-11590) | 0.64 (0.34-1.09) |
| United States | Male | 775554 (713462-838166) | 90015 (66408-112438) | 11.62 (8.57-14.16) | 16635 (9194-26856) | 2.16 (1.18-3.50) | 78467 (58494-101215) | 10.20 (7.56-12.78) | 7417 (3865-12354) | 0.96 (0.50-1.58) | 6930 (3575-11668) | 0.90 (0.47-1.50) |
| Uruguay | Female | 7608 (6793-8421) | 939 (732-1147) | 12.34 (9.68-14.65) | 536 (403-675) | 7.05 (5.37-8.72) | 509 (381-651) | 6.68 (5.10-8.37) | 293 (219-378) | 3.85 (2.86-4.86) | 280 (206-366) | 3.68 (2.74-4.67) |
| Uruguay | Male | 5451 (4968-5890) | 776 (609-969) | 14.32 (11.24-17.37) | 438 (337-556) | 8.10 (6.39-10.11) | 419 (315-542) | 7.74 (5.91-9.64) | 215 (160-277) | 3.95 (2.94-5.13) | 190 (141-252) | 3.50 (2.61-4.58) |
| Uzbekistan | Female | 28228 (24985-31520) | 2029 (1428-2674) | 7.19 (5.17-9.47) | 1462 (1047-1934) | 5.24 (3.76-6.77) | 915 (643-1237) | 3.27 (2.30-4.35) | 492 (329-669) | 1.74 (1.18-2.37) | 415 (274-569) | 1.47 (0.98-2.00) |
| Uzbekistan | Male | 23936 (21402-26255) | 2954 (2295-3645) | 12.40 (9.83-14.94) | 2020 (1503-2568) | 8.44 (6.42-10.47) | 1239 (919-1619) | 5.18 (3.92-6.67) | 503 (359-679) | 2.11 (1.51-2.77) | 394 (269-536) | 1.65 (1.17-2.21) |
| Vanuatu | Female | 199 (178-220) | 19 (15-24) | 9.63 (7.52-11.95) | 13 (10-17) | 6.61 (5.09-8.30) | 8 (6-11) | 4.22 (3.19-5.39) | 3 (2-4) | 1.47 (1.04-2.06) | 1 (1-2) | 0.70 (0.42-1.08) |
| Vanuatu | Male | 180 (163-197) | 22 (17-27) | 12.08 (9.51-14.59) | 15 (12-19) | 8.15 (6.46-10.18) | 9 (7-12) | 5.21 (4.00-6.74) | 2 (1-3) | 1.20 (0.80-1.75) | 1 (1-2) | 0.63 (0.38-0.96) |
| Venezuela | Female | 57631 (51293-63615) | 6032 (4465-7660) | 10.52 (7.95-12.94) | 4160 (2999-5388) | 7.22 (5.33-9.32) | 2568 (1864-3364) | 4.47 (3.25-5.77) | 1998 (1422-2690) | 3.48 (2.54-4.55) | 1872 (1326-2506) | 3.25 (2.37-4.27) |
| Venezuela | Male | 52436 (47493-57323) | 7479 (5511-9320) | 14.23 (10.60-17.47) | 5000 (3645-6453) | 9.60 (7.12-11.91) | 3079 (2227-4056) | 5.89 (4.28-7.62) | 1786 (1283-2381) | 3.42 (2.53-4.39) | 1599 (1152-2111) | 3.06 (2.23-3.93) |
| Vietnam | Female | 131248 (117975-144623) | 18520 (14207-23334) | 14.15 (10.97-17.12) | 12252 (9325-15325) | 9.32 (7.15-11.58) | 7892 (5897-10301) | 6.02 (4.58-7.77) | 3243 (2369-4265) | 2.48 (1.82-3.29) | 2204 (1537-3017) | 1.69 (1.19-2.23) |
| Vietnam | Male | 73061 (65776-81069) | 12025 (9493-14734) | 16.47 (13.34-19.84) | 7956 (6235-10124) | 10.92 (8.70-13.54) | 5183 (3856-6671) | 7.05 (5.44-8.96) | 1875 (1378-2565) | 2.58 (1.93-3.39) | 1294 (909-1760) | 1.78 (1.27-2.34) |
| Yemen | Female | 24420 (22056-26795) | 99 (12-317) | 0.41 (0.05-1.29) | 91 (11-304) | 0.38 (0.04-1.23) | 76 (8-230) | 0.31 (0.03-0.93) | 12 (0-66) | 0.05 (0.00-0.27) | 9 (0-53) | 0.04 (0.00-0.21) |
| Yemen | Male | 18564 (16982-20155) | 872 (513-1267) | 4.72 (2.82-6.71) | 679 (426-970) | 3.69 (2.32-5.16) | 422 (271-605) | 2.27 (1.48-3.20) | 121 (64-193) | 0.65 (0.35-1.04) | 103 (52-169) | 0.55 (0.29-0.90) |
| Zambia | Female | 5046 (4542-5548) | 574 (421-710) | 11.37 (8.69-13.92) | 395 (285-498) | 7.83 (5.87-9.76) | 237 (173-308) | 4.73 (3.51-5.99) | 108 (77-144) | 2.16 (1.54-2.81) | 65 (44-91) | 1.29 (0.89-1.83) |
| Zambia | Male | 4054 (3663-4485) | 358 (254-463) | 8.90 (6.43-11.21) | 253 (176-328) | 6.26 (4.42-8.00) | 154 (108-209) | 3.78 (2.67-5.04) | 45 (31-65) | 1.13 (0.76-1.58) | 27 (17-40) | 0.67 (0.41-0.98) |
| Zimbabwe | Female | 7163 (6510-7848) | 595 (428-752) | 8.29 (6.20-10.37) | 420 (304-537) | 5.86 (4.35-7.44) | 258 (185-337) | 3.60 (2.60-4.66) | 126 (84-176) | 1.75 (1.18-2.45) | 101 (62-151) | 1.41 (0.87-2.10) |
| Zimbabwe | Male | 6101 (5545-6608) | 545 (403-693) | 9.04 (6.72-11.14) | 382 (280-496) | 6.27 (4.57-8.05) | 232 (168-309) | 3.81 (2.84-5.02) | 72 (47-108) | 1.18 (0.80-1.74) | 55 (34-86) | 0.90 (0.57-1.40) |

#### g. **Prevented CKD DALYs**

| **Country** | **Sex** | **Baseline disease burden Median (95% UI)** | **Replacement of discretionary salt and non-discretionary salt** | | **Replacement of discretionary salt** | | **Replacement of non-discretionary salt** | | **Replacement of discretionary salt among those with diagnosed hypertension** | | **Replacement of discretionary salt among those with treated hypertension** | |
| --- | --- | --- | --- | --- | --- | --- | --- | --- | --- | --- | --- | --- |
|  |  |  | **No. of events averted Median (95% UI)** | **Percentage of national disease burden averted % (95% UI)** | **No. of events averted Median (95% UI)** | **Percentage of national disease burden averted % (95% UI)** | **No. of events averted Median (95% UI)** | **Percentage of national disease burden averted % (95% UI)** | **No. of events averted Median (95% UI)** | **Percentage of national disease burden averted % (95% UI)** | **No. of events averted Median (95% UI)** | **Percentage of national disease burden averted % (95% UI)** |
| **Global, millions** | **Female** | 19.17 (18.86-19.49) | 1.90 (1.77-2.02) | 9.90 (9.24-10.49) | 1.15 (1.06-1.24) | 6.00 (5.54-6.45) | 0.99 (0.91-1.07) | 5.18 (4.78-5.59) | 0.34 (0.32-0.37) | 1.80 (1.67-1.93) | 0.29 (0.26-0.31) | 1.49 (1.38-1.60) |
| **Global, millions** | **Male** | 21.71 (21.32-22.07) | 2.69 (2.53-2.85) | 12.39 (11.66-13.06) | 1.64 (1.51-1.75) | 7.53 (7.01-8.02) | 1.41 (1.32-1.51) | 6.52 (6.06-6.98) | 0.39 (0.36-0.42) | 1.79 (1.67-1.94) | 0.32 (0.29-0.34) | 1.46 (1.35-1.58) |
| Afghanistan | Female | 96093 (82098-108290) | 567 (96-1382) | 0.60 (0.10-1.45) | 505 (84-1206) | 0.53 (0.09-1.28) | 371 (67-922) | 0.39 (0.07-0.93) | 105 (14-338) | 0.11 (0.02-0.35) | 84 (12-276) | 0.09 (0.01-0.29) |
| Afghanistan | Male | 42044 (35441-48252) | 2043 (1233-2968) | 4.86 (3.02-6.94) | 1573 (936-2237) | 3.74 (2.34-5.25) | 965 (582-1391) | 2.29 (1.45-3.30) | 258 (142-408) | 0.61 (0.34-0.94) | 205 (112-336) | 0.49 (0.28-0.77) |
| Albania | Female | 4967 (4612-5319) | 847 (692-994) | 17.01 (14.30-19.72) | 543 (444-657) | 10.94 (9.05-13.09) | 380 (301-468) | 7.62 (6.15-9.34) | 236 (185-303) | 4.76 (3.77-6.08) | 208 (160-271) | 4.19 (3.24-5.50) |
| Albania | Male | 5514 (5078-5922) | 1172 (971-1347) | 21.23 (18.17-24.06) | 780 (634-940) | 14.22 (11.71-16.80) | 551 (442-675) | 10.01 (8.12-12.08) | 179 (134-235) | 3.25 (2.43-4.25) | 132 (92-183) | 2.38 (1.71-3.31) |
| Algeria | Female | 123316 (113933-132748) | 584 (81-1500) | 0.48 (0.07-1.21) | 520 (67-1369) | 0.42 (0.05-1.10) | 424 (47-1062) | 0.34 (0.04-0.87) | 95 (3-426) | 0.08 (0.00-0.36) | 76 (1-377) | 0.06 (0.00-0.30) |
| Algeria | Male | 101626 (93506-110728) | 4637 (2913-6519) | 4.59 (2.84-6.23) | 3570 (2274-4917) | 3.52 (2.25-4.81) | 2435 (1585-3371) | 2.40 (1.54-3.27) | 807 (425-1265) | 0.79 (0.42-1.24) | 659 (337-1029) | 0.65 (0.33-1.01) |
| American Samoa | Female | 445 (401-488) | 25 (17-34) | 5.72 (3.81-7.46) | 17 (11-22) | 3.75 (2.56-4.91) | 15 (11-20) | 3.44 (2.34-4.51) | 5 (3-8) | 1.23 (0.80-1.70) | 4 (2-6) | 0.92 (0.57-1.33) |
| American Samoa | Male | 442 (401-486) | 30 (22-40) | 6.88 (4.95-8.87) | 19 (14-25) | 4.36 (3.15-5.58) | 18 (13-23) | 3.99 (2.94-5.12) | 6 (4-8) | 1.29 (0.87-1.76) | 4 (3-6) | 0.94 (0.62-1.36) |
| Andorra | Female | 178 (164-191) | 7 (4-11) | 3.97 (2.29-6.06) | 4 (2-6) | 2.21 (1.30-3.20) | 6 (4-9) | 3.44 (1.98-5.06) | 2 (1-3) | 0.92 (0.47-1.44) | 1 (1-2) | 0.83 (0.42-1.32) |
| Andorra | Male | 176 (164-187) | 18 (14-21) | 9.96 (7.94-12.14) | 7 (5-9) | 3.93 (3.05-4.97) | 13 (10-16) | 7.18 (5.70-8.78) | 3 (2-4) | 1.64 (1.15-2.18) | 3 (2-4) | 1.52 (1.08-2.09) |
| Angola | Female | 46685 (42872-50397) | 2577 (1779-3471) | 5.51 (3.75-7.39) | 1915 (1344-2642) | 4.12 (2.86-5.54) | 1221 (838-1669) | 2.62 (1.81-3.48) | 465 (310-661) | 0.99 (0.66-1.39) | 219 (129-332) | 0.47 (0.28-0.71) |
| Angola | Male | 73813 (67236-79705) | 4610 (3294-6116) | 6.23 (4.48-8.17) | 3409 (2396-4458) | 4.59 (3.28-6.01) | 2144 (1502-2803) | 2.91 (2.08-3.79) | 584 (361-842) | 0.79 (0.50-1.12) | 348 (202-537) | 0.47 (0.28-0.72) |
| Antigua and Barbuda | Female | 507 (477-540) | 34 (24-43) | 6.65 (4.79-8.46) | 21 (16-26) | 4.13 (3.08-5.18) | 19 (14-25) | 3.84 (2.84-4.96) | 9 (7-12) | 1.84 (1.32-2.41) | 8 (5-11) | 1.55 (1.07-2.09) |
| Antigua and Barbuda | Male | 608 (563-650) | 62 (48-76) | 10.35 (8.06-12.31) | 37 (28-46) | 6.03 (4.58-7.47) | 34 (26-42) | 5.59 (4.31-7.00) | 13 (9-17) | 2.07 (1.51-2.76) | 10 (7-14) | 1.70 (1.20-2.35) |
| Argentina | Female | 145716 (135925-154911) | 16470 (13596-19307) | 11.30 (9.45-13.06) | 10261 (8296-12391) | 7.05 (5.76-8.42) | 8037 (6420-9961) | 5.53 (4.42-6.73) | 4723 (3819-5956) | 3.26 (2.62-4.03) | 4143 (3247-5190) | 2.84 (2.27-3.52) |
| Argentina | Male | 159635 (149874-171142) | 21499 (17763-25641) | 13.43 (11.08-15.72) | 13385 (10756-16292) | 8.37 (6.78-10.11) | 10460 (8317-12884) | 6.56 (5.24-8.00) | 5880 (4523-7382) | 3.67 (2.86-4.52) | 5023 (3885-6267) | 3.15 (2.44-3.86) |
| Armenia | Female | 7654 (7190-8113) | 699 (552-878) | 9.19 (7.21-11.27) | 482 (375-604) | 6.32 (4.88-7.84) | 326 (246-414) | 4.25 (3.24-5.30) | 176 (124-240) | 2.29 (1.66-3.10) | 151 (105-203) | 1.98 (1.38-2.66) |
| Armenia | Male | 7049 (6592-7492) | 941 (777-1116) | 13.36 (11.08-15.61) | 619 (500-751) | 8.80 (7.15-10.47) | 414 (333-514) | 5.87 (4.72-7.25) | 162 (122-213) | 2.30 (1.75-3.01) | 107 (80-145) | 1.52 (1.15-2.05) |
| Australia | Female | 45435 (42703-48291) | 1350 (685-2255) | 2.99 (1.51-4.98) | 533 (256-901) | 1.17 (0.56-1.99) | 1283 (637-2125) | 2.83 (1.40-4.69) | 200 (80-365) | 0.44 (0.18-0.81) | 178 (69-325) | 0.39 (0.15-0.72) |
| Australia | Male | 45522 (42538-48426) | 2681 (1813-3784) | 5.89 (3.98-8.24) | 807 (497-1175) | 1.76 (1.08-2.60) | 2403 (1633-3317) | 5.29 (3.60-7.15) | 287 (160-457) | 0.64 (0.35-1.00) | 264 (143-419) | 0.58 (0.32-0.92) |
| Austria | Female | 30754 (28990-32595) | 3786 (3034-4527) | 12.34 (9.95-14.49) | 1139 (737-1561) | 3.72 (2.39-5.09) | 2971 (2333-3689) | 9.66 (7.69-11.80) | 533 (320-785) | 1.73 (1.06-2.54) | 473 (290-704) | 1.54 (0.94-2.31) |
| Austria | Male | 24938 (23811-26066) | 3850 (3175-4479) | 15.47 (12.77-17.93) | 1125 (810-1525) | 4.51 (3.28-6.09) | 2998 (2430-3598) | 12.04 (9.74-14.34) | 518 (351-740) | 2.08 (1.41-2.98) | 506 (343-724) | 2.03 (1.37-2.91) |
| Azerbaijan | Female | 20740 (19274-22220) | 1628 (1231-2022) | 7.86 (5.94-9.61) | 1126 (843-1392) | 5.43 (4.09-6.73) | 769 (571-985) | 3.71 (2.76-4.71) | 406 (295-533) | 1.96 (1.40-2.52) | 334 (242-440) | 1.61 (1.15-2.11) |
| Azerbaijan | Male | 22052 (20279-23889) | 2710 (2153-3308) | 12.29 (9.95-14.53) | 1783 (1389-2181) | 8.11 (6.33-9.80) | 1204 (932-1515) | 5.48 (4.33-6.73) | 414 (307-540) | 1.88 (1.42-2.39) | 294 (217-385) | 1.34 (0.99-1.74) |
| Bahrain | Female | 3044 (2813-3286) | 15 (3-45) | 0.51 (0.10-1.43) | 12 (2-36) | 0.41 (0.08-1.21) | 13 (3-41) | 0.44 (0.09-1.30) | 3 (0-13) | 0.09 (0.00-0.43) | 2 (0-11) | 0.07 (0.00-0.37) |
| Bahrain | Male | 4227 (3872-4560) | 228 (150-317) | 5.40 (3.55-7.41) | 131 (86-178) | 3.13 (2.07-4.14) | 154 (102-215) | 3.68 (2.50-5.07) | 30 (17-45) | 0.70 (0.40-1.06) | 25 (13-39) | 0.59 (0.31-0.92) |
| Bangladesh | Female | 200214 (185631-214527) | 20960 (17166-24587) | 10.48 (8.63-12.08) | 14496 (11478-17462) | 7.24 (5.85-8.61) | 8984 (7170-11150) | 4.51 (3.63-5.46) | 3343 (2596-4233) | 1.67 (1.31-2.07) | 2659 (2028-3354) | 1.33 (1.04-1.64) |
| Bangladesh | Male | 277670 (256708-298322) | 29524 (23250-35210) | 10.63 (8.48-12.51) | 20503 (15886-24810) | 7.38 (5.82-8.83) | 12659 (9625-15797) | 4.58 (3.53-5.59) | 2927 (2174-3676) | 1.06 (0.79-1.31) | 2281 (1657-2947) | 0.82 (0.60-1.04) |
| Barbados | Female | 1638 (1522-1762) | 74 (45-108) | 4.55 (2.66-6.67) | 54 (36-75) | 3.31 (2.13-4.51) | 51 (34-72) | 3.13 (2.00-4.31) | 28 (17-40) | 1.70 (1.01-2.40) | 26 (15-37) | 1.56 (0.95-2.22) |
| Barbados | Male | 1697 (1572-1824) | 144 (108-181) | 8.54 (6.41-10.53) | 85 (63-109) | 5.00 (3.76-6.33) | 79 (59-102) | 4.69 (3.48-5.87) | 34 (25-45) | 2.02 (1.47-2.66) | 31 (22-42) | 1.83 (1.30-2.42) |
| Belarus | Female | 12437 (11650-13235) | 433 (252-649) | 3.47 (2.04-5.14) | 365 (219-531) | 2.93 (1.74-4.30) | 291 (178-415) | 2.33 (1.44-3.31) | 194 (106-295) | 1.56 (0.86-2.34) | 154 (84-244) | 1.25 (0.67-1.95) |
| Belarus | Male | 8695 (8164-9227) | 858 (685-1033) | 9.89 (8.01-11.71) | 563 (448-693) | 6.48 (5.18-7.92) | 402 (318-498) | 4.61 (3.66-5.71) | 219 (162-281) | 2.52 (1.89-3.17) | 147 (110-196) | 1.70 (1.25-2.21) |
| Belgium | Female | 28783 (26785-30624) | 3269 (2618-3938) | 11.38 (9.19-13.45) | 1034 (717-1377) | 3.58 (2.54-4.80) | 2541 (1997-3162) | 8.82 (7.00-10.81) | 491 (333-677) | 1.70 (1.18-2.35) | 468 (313-654) | 1.62 (1.11-2.26) |
| Belgium | Male | 26221 (24673-27825) | 3598 (3019-4240) | 13.67 (11.60-16.09) | 1118 (835-1477) | 4.27 (3.19-5.63) | 2790 (2254-3350) | 10.63 (8.64-12.64) | 485 (344-669) | 1.86 (1.32-2.54) | 480 (337-654) | 1.83 (1.30-2.51) |
| Belize | Female | 1881 (1760-2000) | 101 (69-137) | 5.41 (3.68-7.23) | 72 (49-96) | 3.86 (2.64-5.07) | 51 (35-69) | 2.70 (1.87-3.66) | 26 (17-35) | 1.37 (0.93-1.84) | 21 (14-29) | 1.11 (0.72-1.54) |
| Belize | Male | 1956 (1826-2082) | 179 (137-221) | 9.13 (6.99-11.13) | 120 (90-150) | 6.12 (4.60-7.63) | 83 (62-106) | 4.22 (3.19-5.37) | 30 (22-40) | 1.54 (1.14-2.02) | 24 (16-33) | 1.22 (0.85-1.67) |
| Benin | Female | 20437 (18963-21875) | 1967 (1592-2324) | 9.64 (7.91-11.13) | 1368 (1081-1666) | 6.72 (5.28-8.00) | 826 (634-1012) | 4.05 (3.15-4.89) | 281 (207-359) | 1.38 (1.04-1.72) | 192 (139-256) | 0.95 (0.68-1.24) |
| Benin | Male | 29539 (27075-32050) | 2216 (1662-2750) | 7.51 (5.62-9.28) | 1574 (1187-1997) | 5.34 (4.01-6.69) | 957 (704-1225) | 3.26 (2.41-4.11) | 170 (117-230) | 0.58 (0.39-0.79) | 120 (82-167) | 0.41 (0.28-0.56) |
| Bermuda | Female | 201 (188-216) | 16 (12-20) | 7.75 (5.87-9.55) | 1 (1-2) | 0.55 (0.39-0.96) | 15 (11-19) | 7.48 (5.64-9.18) | 1 (0-1) | 0.26 (0.18-0.49) | 0 (0-1) | 0.22 (0.15-0.42) |
| Bermuda | Male | 273 (256-291) | 30 (24-36) | 10.87 (8.69-12.79) | 2 (1-3) | 0.73 (0.54-1.24) | 28 (23-34) | 10.41 (8.33-12.25) | 1 (1-1) | 0.26 (0.18-0.48) | 1 (0-1) | 0.23 (0.15-0.41) |
| Bhutan | Female | 1692 (1551-1834) | 164 (125-198) | 9.67 (7.58-11.51) | 112 (84-139) | 6.63 (5.05-8.10) | 71 (54-91) | 4.22 (3.23-5.29) | 31 (23-40) | 1.85 (1.38-2.34) | 23 (17-30) | 1.36 (1.00-1.76) |
| Bhutan | Male | 1893 (1743-2027) | 207 (164-246) | 10.95 (8.87-12.83) | 141 (111-173) | 7.45 (5.97-8.88) | 92 (69-112) | 4.82 (3.74-5.90) | 30 (23-38) | 1.58 (1.21-1.98) | 19 (15-25) | 1.01 (0.77-1.33) |
| Bolivia | Female | 62150 (57023-67277) | 5480 (3896-6792) | 8.78 (6.45-10.73) | 3724 (2623-4732) | 5.99 (4.26-7.51) | 2379 (1682-3134) | 3.83 (2.74-4.98) | 1319 (877-1765) | 2.11 (1.42-2.83) | 1158 (755-1627) | 1.88 (1.23-2.59) |
| Bolivia | Male | 50236 (45198-55219) | 5772 (4198-7279) | 11.47 (8.43-14.07) | 3901 (2798-4957) | 7.79 (5.71-9.65) | 2505 (1796-3265) | 4.99 (3.61-6.34) | 1041 (700-1453) | 2.07 (1.40-2.88) | 866 (559-1242) | 1.71 (1.13-2.43) |
| Bosnia and Herzegovina | Female | 8351 (7827-8895) | 1471 (1216-1725) | 17.66 (14.74-20.32) | 939 (764-1131) | 11.23 (9.12-13.31) | 671 (530-831) | 8.00 (6.42-9.79) | 503 (394-622) | 6.05 (4.76-7.40) | 463 (360-579) | 5.56 (4.32-6.86) |
| Bosnia and Herzegovina | Male | 8689 (8139-9243) | 1831 (1544-2112) | 21.14 (18.08-23.86) | 1216 (995-1452) | 13.98 (11.63-16.61) | 873 (710-1065) | 10.05 (8.20-11.99) | 497 (388-620) | 5.70 (4.50-7.07) | 455 (352-584) | 5.24 (4.10-6.71) |
| Botswana | Female | 4825 (4456-5188) | 298 (208-388) | 6.20 (4.36-7.92) | 213 (152-275) | 4.42 (3.17-5.64) | 155 (110-203) | 3.22 (2.29-4.11) | 70 (48-96) | 1.45 (0.99-1.95) | 57 (38-78) | 1.18 (0.79-1.61) |
| Botswana | Male | 5933 (5370-6503) | 372 (265-496) | 6.31 (4.53-8.14) | 262 (189-348) | 4.45 (3.21-5.71) | 191 (136-258) | 3.24 (2.36-4.26) | 55 (35-78) | 0.93 (0.57-1.31) | 38 (22-56) | 0.64 (0.37-0.92) |
| Brazil | Female | 585280 (554599-615005) | 59550 (48088-71301) | 10.17 (8.22-12.10) | 38739 (30312-47762) | 6.61 (5.22-8.14) | 27976 (21615-34720) | 4.77 (3.74-5.93) | 19182 (14832-24028) | 3.28 (2.54-4.06) | 18605 (14355-23267) | 3.18 (2.48-3.97) |
| Brazil | Male | 640870 (607973-670999) | 83830 (68953-99082) | 13.11 (10.89-15.30) | 53992 (44175-64525) | 8.43 (6.95-10.04) | 38856 (31300-47809) | 6.08 (4.91-7.42) | 22549 (17974-27313) | 3.53 (2.84-4.25) | 21097 (16833-25878) | 3.30 (2.64-4.04) |
| Brunei | Female | 1132 (1056-1208) | 150 (118-185) | 13.27 (10.41-15.96) | 70 (52-87) | 6.18 (4.65-7.66) | 94 (73-118) | 8.32 (6.42-10.30) | 30 (22-39) | 2.67 (1.96-3.39) | 28 (21-37) | 2.50 (1.85-3.21) |
| Brunei | Male | 1346 (1249-1439) | 211 (169-252) | 15.73 (12.67-18.45) | 97 (75-122) | 7.25 (5.66-8.92) | 133 (105-164) | 9.94 (7.82-11.92) | 35 (26-45) | 2.62 (1.96-3.35) | 32 (24-41) | 2.38 (1.82-3.03) |
| Bulgaria | Female | 26980 (24907-28933) | 4505 (3749-5329) | 16.76 (14.03-19.30) | 2730 (2206-3274) | 10.14 (8.26-12.14) | 2225 (1782-2732) | 8.28 (6.69-9.91) | 1395 (1097-1774) | 5.18 (4.14-6.60) | 1245 (952-1609) | 4.61 (3.59-5.96) |
| Bulgaria | Male | 31531 (29486-33655) | 6566 (5400-7589) | 20.81 (17.44-23.62) | 4110 (3251-5006) | 13.08 (10.50-15.52) | 3362 (2668-4089) | 10.63 (8.59-12.81) | 1740 (1263-2246) | 5.52 (4.07-7.06) | 1484 (1069-1979) | 4.71 (3.42-6.20) |
| Burkina Faso | Female | 45947 (42320-49795) | 3044 (2258-3897) | 6.67 (4.90-8.40) | 2192 (1597-2822) | 4.78 (3.46-6.01) | 1320 (939-1729) | 2.88 (2.04-3.75) | 428 (292-597) | 0.94 (0.64-1.27) | 289 (186-413) | 0.63 (0.41-0.90) |
| Burkina Faso | Male | 48443 (44429-52516) | 3585 (2654-4690) | 7.46 (5.56-9.34) | 2567 (1861-3352) | 5.30 (3.86-6.80) | 1526 (1106-2048) | 3.15 (2.32-4.13) | 334 (224-498) | 0.69 (0.47-1.01) | 177 (113-278) | 0.36 (0.24-0.57) |
| Burundi | Female | 15355 (14010-16612) | 1699 (1369-2048) | 11.08 (8.95-13.10) | 1185 (945-1460) | 7.73 (6.13-9.37) | 694 (544-877) | 4.53 (3.60-5.54) | 284 (204-391) | 1.85 (1.34-2.52) | 191 (123-271) | 1.24 (0.81-1.75) |
| Burundi | Male | 24916 (22340-27415) | 2097 (1556-2642) | 8.44 (6.36-10.40) | 1494 (1093-1931) | 6.05 (4.51-7.46) | 889 (631-1175) | 3.59 (2.60-4.57) | 263 (172-381) | 1.06 (0.70-1.51) | 180 (102-283) | 0.72 (0.43-1.10) |
| Cambodia | Female | 47907 (43670-52164) | 4779 (3199-6292) | 9.95 (6.70-12.86) | 3265 (2211-4403) | 6.81 (4.65-9.01) | 1992 (1327-2732) | 4.15 (2.79-5.61) | 772 (514-1062) | 1.62 (1.09-2.18) | 602 (398-868) | 1.25 (0.84-1.78) |
| Cambodia | Male | 41662 (37652-45429) | 4928 (3470-6268) | 11.89 (8.33-14.96) | 3411 (2318-4390) | 8.18 (5.64-10.51) | 2102 (1433-2736) | 5.05 (3.44-6.52) | 542 (352-750) | 1.31 (0.84-1.76) | 387 (233-556) | 0.93 (0.55-1.33) |
| Cameroon | Female | 69653 (63179-75362) | 5020 (3731-6198) | 7.23 (5.37-8.72) | 3594 (2650-4455) | 5.17 (3.95-6.37) | 2198 (1627-2757) | 3.18 (2.42-3.92) | 756 (535-1000) | 1.09 (0.78-1.44) | 427 (274-592) | 0.61 (0.41-0.85) |
| Cameroon | Male | 99876 (89923-108918) | 7631 (5777-9643) | 7.69 (5.90-9.49) | 5423 (4086-6849) | 5.46 (4.20-6.79) | 3292 (2481-4323) | 3.33 (2.58-4.26) | 728 (497-1006) | 0.74 (0.51-1.00) | 442 (285-641) | 0.44 (0.29-0.64) |
| Canada | Female | 72537 (68277-77319) | 5999 (4430-7520) | 8.27 (6.08-10.16) | 1931 (1346-2573) | 2.67 (1.88-3.53) | 4803 (3540-6031) | 6.62 (4.93-8.21) | 733 (491-1026) | 1.01 (0.68-1.40) | 716 (480-1005) | 0.98 (0.67-1.38) |
| Canada | Male | 75277 (70974-79442) | 8877 (7082-10531) | 11.79 (9.48-13.85) | 2755 (2017-3670) | 3.67 (2.68-4.85) | 6971 (5451-8347) | 9.27 (7.23-11.04) | 1157 (824-1600) | 1.55 (1.09-2.12) | 1153 (817-1589) | 1.54 (1.08-2.13) |
| Cape Verde | Female | 1158 (1077-1241) | 97 (76-120) | 8.40 (6.61-10.18) | 68 (52-84) | 5.82 (4.56-7.15) | 43 (34-55) | 3.73 (2.97-4.70) | 27 (20-35) | 2.36 (1.79-3.05) | 23 (17-31) | 2.01 (1.45-2.66) |
| Cape Verde | Male | 1760 (1596-1925) | 150 (114-185) | 8.47 (6.62-10.30) | 104 (80-130) | 5.90 (4.61-7.24) | 66 (50-86) | 3.77 (2.92-4.74) | 26 (19-35) | 1.49 (1.11-1.95) | 19 (14-27) | 1.11 (0.79-1.50) |
| Central African Republic | Female | 10964 (9937-11925) | 572 (385-781) | 5.23 (3.53-6.99) | 437 (291-595) | 3.99 (2.64-5.30) | 269 (181-369) | 2.47 (1.67-3.31) | 87 (54-122) | 0.79 (0.50-1.10) | 46 (26-68) | 0.42 (0.25-0.63) |
| Central African Republic | Male | 18973 (16971-20946) | 1099 (710-1515) | 5.84 (3.87-7.78) | 825 (532-1132) | 4.37 (2.88-5.80) | 504 (326-674) | 2.66 (1.78-3.48) | 102 (59-153) | 0.54 (0.31-0.79) | 58 (33-88) | 0.31 (0.18-0.45) |
| Chad | Female | 17170 (15782-18549) | 1111 (775-1444) | 6.48 (4.57-8.28) | 802 (564-1059) | 4.67 (3.30-6.02) | 483 (329-640) | 2.80 (1.94-3.68) | 179 (121-252) | 1.04 (0.71-1.46) | 114 (70-168) | 0.66 (0.42-0.97) |
| Chad | Male | 30445 (27182-33511) | 2296 (1672-2934) | 7.57 (5.58-9.25) | 1643 (1192-2101) | 5.45 (3.98-6.81) | 980 (704-1268) | 3.24 (2.31-4.13) | 262 (175-384) | 0.87 (0.57-1.26) | 174 (112-263) | 0.57 (0.37-0.86) |
| Chile | Female | 50885 (47505-54074) | 5991 (4937-7048) | 11.81 (9.73-13.59) | 3512 (2818-4264) | 6.90 (5.58-8.25) | 3182 (2522-3853) | 6.27 (4.99-7.46) | 1737 (1372-2153) | 3.41 (2.70-4.16) | 1585 (1254-1983) | 3.13 (2.46-3.85) |
| Chile | Male | 50863 (47874-53973) | 7178 (5921-8537) | 14.11 (11.74-16.39) | 4162 (3407-5075) | 8.15 (6.68-9.69) | 3774 (3026-4590) | 7.42 (5.99-8.87) | 1849 (1456-2282) | 3.64 (2.87-4.43) | 1618 (1266-2036) | 3.17 (2.50-3.90) |
| China | Female | 2785666 (2621046-2955008) | 475940 (379031-558715) | 17.07 (13.89-19.83) | 293960 (227258-358592) | 10.56 (8.29-12.74) | 243323 (184554-301243) | 8.72 (6.74-10.66) | 78964 (61077-98926) | 2.84 (2.17-3.51) | 65440 (49383-83195) | 2.35 (1.78-2.94) |
| China | Male | 3194503 (2992943-3411778) | 623407 (508547-724004) | 19.58 (16.16-22.29) | 407266 (319828-487755) | 12.77 (10.14-15.00) | 339157 (262085-416209) | 10.66 (8.25-12.80) | 93999 (70831-120319) | 2.94 (2.25-3.72) | 73817 (55607-93890) | 2.31 (1.76-2.93) |
| Colombia | Female | 98938 (93738-104488) | 12057 (9894-14344) | 12.14 (10.09-14.21) | 7943 (6402-9468) | 8.01 (6.46-9.43) | 5480 (4394-6714) | 5.53 (4.45-6.72) | 3515 (2810-4293) | 3.55 (2.81-4.30) | 3042 (2418-3761) | 3.08 (2.45-3.74) |
| Colombia | Male | 122517 (115362-130012) | 22974 (19065-26333) | 18.73 (15.74-21.24) | 15113 (12391-18007) | 12.29 (10.13-14.52) | 10484 (8512-12665) | 8.55 (6.93-10.19) | 5290 (4200-6614) | 4.31 (3.43-5.34) | 4301 (3375-5460) | 3.52 (2.74-4.43) |
| Comoros | Female | 2199 (2003-2404) | 260 (210-317) | 11.84 (9.71-14.12) | 179 (142-219) | 8.12 (6.58-9.75) | 109 (85-137) | 4.95 (3.93-6.08) | 46 (34-63) | 2.11 (1.57-2.78) | 30 (21-42) | 1.37 (0.98-1.90) |
| Comoros | Male | 2307 (2118-2517) | 209 (162-260) | 9.04 (7.22-11.05) | 147 (113-187) | 6.36 (4.98-7.88) | 90 (69-117) | 3.92 (3.01-4.98) | 26 (18-37) | 1.12 (0.77-1.61) | 17 (11-26) | 0.75 (0.50-1.10) |
| Congo | Female | 17776 (16106-19363) | 993 (690-1321) | 5.58 (3.88-7.27) | 743 (514-996) | 4.17 (2.90-5.49) | 478 (333-638) | 2.69 (1.91-3.54) | 154 (98-222) | 0.87 (0.55-1.24) | 91 (51-134) | 0.51 (0.29-0.75) |
| Congo | Male | 20004 (18144-21649) | 1257 (895-1651) | 6.30 (4.49-8.26) | 922 (650-1195) | 4.64 (3.26-6.04) | 590 (423-768) | 2.96 (2.12-3.83) | 144 (93-210) | 0.72 (0.47-1.07) | 106 (63-166) | 0.53 (0.32-0.82) |
| Costa Rica | Female | 17378 (16462-18376) | 1952 (1584-2267) | 11.25 (9.22-12.94) | 1197 (957-1473) | 6.89 (5.55-8.35) | 989 (776-1202) | 5.69 (4.48-6.87) | 599 (476-744) | 3.46 (2.75-4.25) | 579 (455-719) | 3.34 (2.62-4.13) |
| Costa Rica | Male | 23101 (21667-24511) | 3471 (2844-4082) | 15.02 (12.49-17.44) | 2075 (1692-2538) | 9.02 (7.36-10.76) | 1701 (1360-2055) | 7.36 (5.98-8.79) | 842 (678-1056) | 3.66 (2.98-4.55) | 797 (634-986) | 3.45 (2.75-4.24) |
| Cote d'Ivoire | Female | 40776 (37495-43998) | 3291 (2503-4048) | 8.09 (6.18-9.74) | 2281 (1703-2807) | 5.61 (4.22-6.87) | 1421 (1059-1792) | 3.49 (2.62-4.43) | 476 (342-636) | 1.17 (0.84-1.56) | 309 (206-442) | 0.76 (0.51-1.09) |
| Cote d'Ivoire | Male | 74960 (69049-81405) | 6210 (4773-7750) | 8.27 (6.49-10.00) | 4316 (3310-5492) | 5.78 (4.45-7.10) | 2692 (2053-3455) | 3.61 (2.81-4.49) | 628 (443-883) | 0.84 (0.58-1.18) | 435 (280-631) | 0.58 (0.38-0.84) |
| Croatia | Female | 14472 (13520-15523) | 2741 (2234-3213) | 18.89 (15.68-21.87) | 1540 (1199-1900) | 10.62 (8.45-12.93) | 1460 (1152-1808) | 10.06 (8.12-12.29) | 957 (729-1206) | 6.59 (5.14-8.29) | 839 (639-1076) | 5.78 (4.43-7.32) |
| Croatia | Male | 11723 (10901-12552) | 2625 (2209-3043) | 22.44 (19.26-25.29) | 1544 (1239-1881) | 13.19 (10.69-15.76) | 1464 (1173-1819) | 12.51 (10.20-15.17) | 861 (674-1088) | 7.35 (5.82-9.07) | 623 (472-810) | 5.32 (4.08-6.73) |
| Cuba | Female | 37151 (34807-39457) | 2484 (1766-3253) | 6.69 (4.83-8.70) | 1668 (1232-2207) | 4.47 (3.33-5.85) | 1267 (936-1681) | 3.42 (2.56-4.48) | 781 (562-1061) | 2.10 (1.53-2.81) | 722 (519-969) | 1.95 (1.39-2.61) |
| Cuba | Male | 40356 (37756-42857) | 4053 (3176-4888) | 10.10 (7.95-12.00) | 2608 (1986-3214) | 6.48 (4.96-7.97) | 1958 (1488-2470) | 4.86 (3.69-6.09) | 957 (708-1228) | 2.38 (1.76-3.00) | 863 (632-1115) | 2.14 (1.58-2.74) |
| Cyprus | Female | 3108 (2875-3350) | 227 (153-299) | 7.33 (5.04-9.54) | 114 (81-153) | 3.69 (2.66-4.84) | 158 (110-211) | 5.13 (3.64-6.67) | 50 (32-70) | 1.59 (1.05-2.26) | 45 (29-65) | 1.46 (0.96-2.09) |
| Cyprus | Male | 3586 (3284-3836) | 419 (327-500) | 11.73 (9.33-13.92) | 192 (145-246) | 5.36 (4.15-6.87) | 272 (213-339) | 7.61 (6.04-9.31) | 82 (60-113) | 2.30 (1.68-3.15) | 76 (54-108) | 2.12 (1.55-3.04) |
| Czech Republic | Female | 20013 (18891-21236) | 3597 (2965-4178) | 18.11 (14.88-20.51) | 1798 (1420-2189) | 8.98 (7.24-10.80) | 2151 (1717-2578) | 10.77 (8.65-12.75) | 947 (730-1198) | 4.74 (3.72-5.89) | 885 (682-1124) | 4.44 (3.43-5.58) |
| Czech Republic | Male | 18356 (17172-19578) | 3979 (3387-4565) | 21.70 (18.66-24.44) | 2083 (1674-2540) | 11.39 (9.17-13.65) | 2474 (2016-2971) | 13.46 (11.01-16.01) | 1107 (853-1404) | 6.06 (4.75-7.49) | 1048 (813-1317) | 5.73 (4.45-7.03) |
| Democratic Republic of the Congo | Female | 172782 (159801-186764) | 2349 (852-4462) | 1.36 (0.51-2.54) | 2230 (837-4101) | 1.29 (0.50-2.34) | 1728 (718-2967) | 1.00 (0.42-1.70) | 421 (121-916) | 0.24 (0.07-0.53) | 247 (71-543) | 0.14 (0.04-0.32) |
| Democratic Republic of the Congo | Male | 237164 (214322-258356) | 5972 (2929-9761) | 2.51 (1.22-4.06) | 5268 (2627-8422) | 2.22 (1.10-3.48) | 3626 (1877-5689) | 1.53 (0.79-2.33) | 744 (288-1450) | 0.32 (0.12-0.59) | 567 (220-1092) | 0.24 (0.09-0.45) |
| Denmark | Female | 14272 (13529-14994) | 1130 (787-1472) | 7.91 (5.57-10.18) | 226 (135-370) | 1.58 (0.94-2.57) | 1007 (709-1330) | 7.08 (5.00-9.16) | 87 (48-152) | 0.61 (0.34-1.05) | 58 (32-103) | 0.41 (0.22-0.72) |
| Denmark | Male | 17291 (16576-18079) | 2092 (1666-2459) | 12.10 (9.80-14.11) | 387 (233-606) | 2.23 (1.34-3.47) | 1826 (1426-2176) | 10.55 (8.32-12.46) | 173 (100-278) | 1.01 (0.57-1.61) | 122 (69-206) | 0.70 (0.40-1.19) |
| Djibouti | Female | 2226 (2019-2429) | 252 (197-307) | 11.37 (9.05-13.49) | 171 (132-210) | 7.69 (5.97-9.32) | 108 (83-135) | 4.90 (3.76-5.98) | 41 (29-56) | 1.86 (1.30-2.46) | 27 (18-39) | 1.22 (0.80-1.74) |
| Djibouti | Male | 3732 (3377-4080) | 306 (226-393) | 8.21 (6.20-10.33) | 215 (156-277) | 5.77 (4.25-7.30) | 138 (99-180) | 3.70 (2.67-4.75) | 37 (24-55) | 1.00 (0.65-1.46) | 25 (15-39) | 0.68 (0.40-1.02) |
| Dominica | Female | 422 (392-456) | 29 (21-37) | 6.84 (4.87-8.66) | 20 (14-26) | 4.67 (3.40-6.00) | 14 (10-19) | 3.42 (2.46-4.39) | 9 (7-13) | 2.25 (1.61-2.93) | 8 (6-11) | 1.93 (1.33-2.57) |
| Dominica | Male | 551 (497-603) | 56 (43-69) | 10.22 (8.01-12.09) | 37 (28-46) | 6.65 (5.12-8.27) | 26 (20-34) | 4.77 (3.60-5.93) | 12 (8-16) | 2.11 (1.53-2.80) | 10 (7-13) | 1.76 (1.28-2.39) |
| Dominican Republic | Female | 28034 (25917-30114) | 1676 (1229-2165) | 6.02 (4.39-7.69) | 1166 (859-1498) | 4.18 (3.07-5.36) | 860 (648-1117) | 3.10 (2.29-3.92) | 568 (409-751) | 2.03 (1.46-2.62) | 479 (336-636) | 1.71 (1.22-2.23) |
| Dominican Republic | Male | 38183 (34374-41842) | 3721 (2910-4550) | 9.71 (7.72-11.57) | 2413 (1870-3035) | 6.32 (4.92-7.73) | 1775 (1345-2255) | 4.64 (3.57-5.80) | 930 (698-1189) | 2.45 (1.88-3.07) | 779 (595-1021) | 2.05 (1.56-2.63) |
| Ecuador | Female | 70942 (64792-77658) | 6494 (4822-7991) | 9.19 (6.89-11.10) | 4346 (3169-5459) | 6.08 (4.49-7.50) | 2969 (2178-3846) | 4.20 (3.06-5.34) | 1473 (1041-1886) | 2.08 (1.46-2.63) | 1317 (940-1710) | 1.87 (1.33-2.37) |
| Ecuador | Male | 76916 (69686-84108) | 9150 (6876-11324) | 11.87 (9.11-14.31) | 6008 (4476-7522) | 7.85 (5.95-9.56) | 4160 (3042-5231) | 5.39 (4.04-6.77) | 1582 (1134-2063) | 2.07 (1.49-2.65) | 1271 (921-1686) | 1.65 (1.18-2.15) |
| Egypt | Female | 407841 (371754-444546) | 2314 (594-6605) | 0.56 (0.14-1.66) | 2059 (516-5950) | 0.50 (0.13-1.48) | 1722 (369-4704) | 0.42 (0.09-1.13) | 502 (26-2291) | 0.12 (0.01-0.57) | 423 (20-1962) | 0.10 (0.00-0.50) |
| Egypt | Male | 441463 (402241-480510) | 22271 (13979-32181) | 5.04 (3.27-7.06) | 16943 (10901-23694) | 3.83 (2.52-5.27) | 11491 (7774-16120) | 2.61 (1.76-3.58) | 3694 (2064-5608) | 0.84 (0.47-1.24) | 3063 (1655-4670) | 0.69 (0.39-1.02) |
| El Salvador | Female | 45399 (41927-49066) | 4728 (3747-5689) | 10.40 (8.33-12.14) | 3178 (2432-3912) | 7.00 (5.45-8.50) | 2109 (1627-2677) | 4.65 (3.58-5.78) | 1343 (1003-1722) | 2.96 (2.27-3.72) | 1217 (918-1557) | 2.69 (2.03-3.37) |
| El Salvador | Male | 65983 (60889-71637) | 9293 (7539-11322) | 14.12 (11.60-16.76) | 6072 (4834-7553) | 9.23 (7.46-11.15) | 4056 (3197-5065) | 6.17 (4.90-7.58) | 1648 (1261-2125) | 2.49 (1.93-3.14) | 1436 (1082-1854) | 2.17 (1.68-2.75) |
| Equatorial Guinea | Female | 3290 (2960-3619) | 197 (140-256) | 5.97 (4.30-7.61) | 139 (97-184) | 4.21 (2.98-5.46) | 101 (72-133) | 3.06 (2.19-3.97) | 30 (20-43) | 0.91 (0.60-1.30) | 19 (11-28) | 0.58 (0.33-0.85) |
| Equatorial Guinea | Male | 3968 (3538-4344) | 263 (190-338) | 6.67 (4.84-8.48) | 181 (129-235) | 4.60 (3.37-5.88) | 132 (96-170) | 3.34 (2.45-4.26) | 27 (18-39) | 0.68 (0.45-0.98) | 19 (11-30) | 0.48 (0.29-0.74) |
| Eritrea | Female | 11688 (10612-12744) | 1199 (889-1508) | 10.34 (7.73-12.73) | 837 (616-1067) | 7.21 (5.33-8.98) | 499 (355-645) | 4.26 (3.14-5.39) | 141 (95-195) | 1.21 (0.83-1.64) | 94 (62-139) | 0.81 (0.55-1.16) |
| Eritrea | Male | 14484 (12689-16349) | 976 (696-1298) | 6.72 (4.90-8.59) | 710 (498-947) | 4.92 (3.47-6.36) | 425 (290-582) | 2.95 (2.09-3.90) | 77 (51-114) | 0.54 (0.35-0.77) | 55 (34-84) | 0.38 (0.23-0.56) |
| Estonia | Female | 7337 (6967-7686) | 14 (0-74) | 0.18 (0.00-1.03) | 13 (0-68) | 0.18 (0.00-0.92) | 14 (0-72) | 0.18 (0.00-0.97) | 4 (0-22) | 0.05 (0.00-0.30) | 3 (0-17) | 0.04 (0.00-0.23) |
| Estonia | Male | 3886 (3727-4044) | 281 (202-359) | 7.23 (5.25-9.22) | 151 (109-190) | 3.86 (2.83-4.89) | 185 (135-233) | 4.76 (3.48-5.99) | 39 (26-52) | 1.00 (0.67-1.35) | 29 (20-41) | 0.75 (0.51-1.05) |
| Ethiopia | Female | 184634 (172995-196334) | 19591 (14941-23737) | 10.63 (8.24-12.76) | 13624 (10219-16657) | 7.39 (5.63-8.95) | 8197 (6055-10278) | 4.46 (3.31-5.52) | 2218 (1573-2965) | 1.21 (0.86-1.59) | 1083 (705-1520) | 0.59 (0.38-0.82) |
| Ethiopia | Male | 224022 (207481-239572) | 18524 (13696-23494) | 8.27 (6.20-10.28) | 13184 (9585-16822) | 5.89 (4.36-7.48) | 7990 (5803-10542) | 3.56 (2.59-4.61) | 1826 (1270-2647) | 0.81 (0.56-1.18) | 1222 (804-1821) | 0.54 (0.36-0.80) |
| Federated States of Micronesia | Female | 693 (612-768) | 56 (39-74) | 8.05 (5.62-10.36) | 38 (26-52) | 5.55 (3.72-7.37) | 25 (17-35) | 3.59 (2.51-4.84) | 11 (7-16) | 1.55 (1.00-2.17) | 6 (4-9) | 0.94 (0.57-1.34) |
| Federated States of Micronesia | Male | 541 (470-607) | 51 (35-67) | 9.48 (6.64-12.03) | 35 (24-47) | 6.49 (4.57-8.35) | 23 (15-31) | 4.24 (2.88-5.50) | 7 (5-10) | 1.35 (0.88-1.84) | 4 (2-6) | 0.79 (0.47-1.11) |
| Fiji | Female | 4078 (3675-4466) | 428 (330-534) | 10.44 (8.30-12.80) | 287 (219-364) | 7.03 (5.53-8.78) | 190 (145-247) | 4.68 (3.62-5.95) | 110 (81-147) | 2.71 (2.00-3.54) | 82 (56-113) | 2.01 (1.43-2.72) |
| Fiji | Male | 5482 (4819-6071) | 646 (501-805) | 11.85 (9.37-14.39) | 429 (328-548) | 7.93 (6.10-9.85) | 288 (217-374) | 5.26 (4.04-6.63) | 109 (78-150) | 2.00 (1.45-2.70) | 75 (51-107) | 1.38 (0.96-1.97) |
| Finland | Female | 10081 (9508-10680) | 633 (424-860) | 6.25 (4.22-8.47) | 231 (150-323) | 2.29 (1.51-3.22) | 547 (364-733) | 5.44 (3.64-7.29) | 111 (66-162) | 1.10 (0.65-1.62) | 95 (56-143) | 0.95 (0.57-1.42) |
| Finland | Male | 8139 (7707-8547) | 1025 (847-1189) | 12.62 (10.51-14.53) | 307 (218-410) | 3.78 (2.72-5.01) | 804 (648-957) | 9.87 (8.12-11.66) | 138 (94-191) | 1.69 (1.17-2.31) | 127 (86-176) | 1.56 (1.07-2.13) |
| France | Female | 135050 (127097-142842) | 10107 (6566-13524) | 7.48 (4.90-9.87) | 4192 (2881-5782) | 3.10 (2.12-4.26) | 7822 (5330-10355) | 5.77 (3.98-7.53) | 1829 (1174-2605) | 1.35 (0.87-1.92) | 1670 (1081-2390) | 1.23 (0.78-1.77) |
| France | Male | 127837 (121988-133574) | 14488 (11581-17178) | 11.34 (9.21-13.45) | 5439 (4150-7064) | 4.25 (3.25-5.53) | 10556 (8385-12771) | 8.23 (6.56-9.95) | 2645 (1940-3579) | 2.07 (1.52-2.79) | 2449 (1806-3323) | 1.92 (1.41-2.62) |
| Gabon | Female | 7213 (6600-7860) | 435 (315-568) | 6.01 (4.42-7.76) | 302 (216-393) | 4.20 (3.05-5.35) | 227 (167-297) | 3.16 (2.32-4.07) | 79 (53-107) | 1.09 (0.75-1.48) | 56 (35-83) | 0.78 (0.50-1.14) |
| Gabon | Male | 9437 (8413-10455) | 603 (420-809) | 6.42 (4.55-8.37) | 421 (297-551) | 4.47 (3.08-5.66) | 314 (222-421) | 3.33 (2.40-4.40) | 72 (45-108) | 0.77 (0.48-1.12) | 53 (31-85) | 0.57 (0.33-0.88) |
| Georgia | Female | 12219 (11432-13038) | 1150 (898-1403) | 9.41 (7.33-11.28) | 793 (604-970) | 6.50 (4.95-7.85) | 534 (408-667) | 4.35 (3.39-5.36) | 385 (281-488) | 3.15 (2.34-3.94) | 343 (254-448) | 2.80 (2.08-3.64) |
| Georgia | Male | 11765 (11094-12445) | 1580 (1308-1852) | 13.43 (11.16-15.60) | 1040 (847-1249) | 8.84 (7.26-10.51) | 695 (556-849) | 5.92 (4.74-7.13) | 385 (299-485) | 3.28 (2.55-4.08) | 295 (229-376) | 2.52 (1.94-3.18) |
| Germany | Female | 338513 (311637-364658) | 24175 (15607-33043) | 7.16 (4.59-9.66) | 8871 (5709-12779) | 2.63 (1.71-3.72) | 20129 (13465-27362) | 5.97 (3.95-8.04) | 4205 (2485-6309) | 1.24 (0.75-1.84) | 4005 (2363-6087) | 1.19 (0.72-1.79) |
| Germany | Male | 291423 (271822-308917) | 36386 (29244-43635) | 12.44 (10.09-14.80) | 11454 (8004-15467) | 3.96 (2.74-5.32) | 28116 (21886-34597) | 9.65 (7.62-11.72) | 5622 (3744-7852) | 1.92 (1.29-2.64) | 5350 (3574-7531) | 1.83 (1.24-2.57) |
| Ghana | Female | 69440 (63950-74781) | 6562 (5274-7947) | 9.52 (7.63-11.29) | 4539 (3542-5617) | 6.57 (5.15-8.00) | 2816 (2168-3548) | 4.07 (3.16-5.05) | 1347 (1021-1722) | 1.94 (1.46-2.44) | 1070 (790-1381) | 1.53 (1.13-1.97) |
| Ghana | Male | 118675 (107852-128637) | 11642 (9064-14180) | 9.85 (7.86-11.58) | 8057 (6147-9960) | 6.77 (5.40-8.20) | 5018 (3775-6396) | 4.22 (3.28-5.23) | 1297 (949-1700) | 1.09 (0.81-1.42) | 982 (706-1321) | 0.83 (0.61-1.10) |
| Greece | Female | 54411 (51457-57555) | 4081 (2645-5427) | 7.50 (4.83-9.91) | 2476 (1667-3284) | 4.54 (3.09-5.95) | 2495 (1689-3293) | 4.57 (3.15-6.12) | 1316 (849-1801) | 2.43 (1.59-3.33) | 1302 (827-1781) | 2.38 (1.54-3.25) |
| Greece | Male | 57485 (55004-60035) | 6560 (5077-7774) | 11.39 (8.87-13.48) | 3659 (2773-4547) | 6.37 (4.87-7.82) | 3678 (2802-4584) | 6.41 (4.90-7.94) | 1875 (1381-2413) | 3.26 (2.40-4.16) | 1850 (1357-2391) | 3.22 (2.34-4.13) |
| Greenland | Female | 78 (73-84) | 6 (4-8) | 7.71 (5.44-9.64) | 2 (1-2) | 2.14 (1.36-3.02) | 5 (3-6) | 6.34 (4.48-8.04) | 1 (0-1) | 0.69 (0.42-1.03) | 0 (0-1) | 0.57 (0.34-0.85) |
| Greenland | Male | 129 (118-141) | 14 (10-17) | 10.72 (7.77-13.08) | 4 (2-6) | 3.00 (1.86-4.24) | 11 (8-14) | 8.66 (6.19-10.91) | 1 (1-2) | 0.94 (0.58-1.41) | 1 (1-2) | 0.80 (0.47-1.22) |
| Grenada | Female | 661 (622-707) | 43 (31-56) | 6.53 (4.60-8.30) | 29 (21-38) | 4.44 (3.27-5.66) | 22 (16-29) | 3.34 (2.45-4.31) | 14 (9-18) | 2.04 (1.44-2.73) | 12 (8-16) | 1.78 (1.25-2.38) |
| Grenada | Male | 949 (879-1025) | 93 (71-115) | 9.83 (7.53-11.85) | 60 (45-76) | 6.33 (4.78-7.87) | 45 (33-58) | 4.70 (3.51-6.01) | 20 (14-28) | 2.16 (1.50-2.90) | 15 (10-22) | 1.62 (1.09-2.24) |
| Guatemala | Female | 75986 (70900-81204) | 8029 (6411-9510) | 10.58 (8.53-12.27) | 5409 (4175-6632) | 7.12 (5.56-8.65) | 3613 (2781-4514) | 4.74 (3.69-5.90) | 1628 (1231-2125) | 2.14 (1.65-2.76) | 1117 (800-1491) | 1.47 (1.06-1.95) |
| Guatemala | Male | 74116 (69343-78366) | 10445 (8416-12379) | 14.12 (11.60-16.58) | 6827 (5422-8249) | 9.21 (7.43-11.08) | 4568 (3601-5604) | 6.18 (4.93-7.56) | 1479 (1115-1892) | 2.00 (1.53-2.52) | 1017 (750-1371) | 1.38 (1.01-1.84) |
| Guinea | Female | 21058 (19501-22534) | 1516 (1134-1886) | 7.23 (5.47-8.91) | 1091 (827-1381) | 5.18 (3.97-6.53) | 661 (498-847) | 3.13 (2.39-3.98) | 320 (229-433) | 1.52 (1.10-2.02) | 157 (103-230) | 0.75 (0.49-1.07) |
| Guinea | Male | 27794 (25128-30701) | 2161 (1646-2708) | 7.81 (6.02-9.53) | 1541 (1166-1988) | 5.57 (4.22-6.95) | 933 (692-1210) | 3.36 (2.51-4.22) | 282 (191-391) | 1.01 (0.71-1.40) | 164 (101-241) | 0.59 (0.36-0.86) |
| Guinea-Bissau | Female | 4054 (3719-4395) | 271 (199-340) | 6.72 (4.90-8.32) | 196 (141-246) | 4.86 (3.51-6.05) | 118 (85-151) | 2.91 (2.09-3.72) | 46 (31-62) | 1.13 (0.78-1.54) | 29 (19-43) | 0.72 (0.47-1.07) |
| Guinea-Bissau | Male | 5610 (5088-6113) | 402 (297-515) | 7.18 (5.30-8.96) | 287 (207-376) | 5.16 (3.80-6.56) | 173 (127-227) | 3.09 (2.28-3.95) | 42 (28-63) | 0.76 (0.51-1.10) | 28 (16-43) | 0.49 (0.29-0.77) |
| Guyana | Female | 4942 (4568-5327) | 285 (194-378) | 5.80 (3.97-7.67) | 195 (137-261) | 3.95 (2.78-5.25) | 153 (106-202) | 3.10 (2.15-4.08) | 77 (52-105) | 1.56 (1.07-2.11) | 61 (40-84) | 1.23 (0.81-1.68) |
| Guyana | Male | 5200 (4738-5662) | 485 (366-610) | 9.40 (7.11-11.45) | 311 (226-397) | 5.98 (4.40-7.51) | 237 (176-314) | 4.55 (3.41-5.81) | 87 (63-119) | 1.68 (1.23-2.28) | 70 (49-95) | 1.34 (0.95-1.81) |
| Haiti | Female | 34184 (28136-39777) | 1956 (1382-2746) | 5.83 (4.12-7.59) | 1484 (1064-2023) | 4.38 (3.14-5.66) | 931 (663-1294) | 2.75 (2.02-3.57) | 510 (348-747) | 1.51 (1.06-2.08) | 270 (175-403) | 0.80 (0.55-1.14) |
| Haiti | Male | 33445 (27524-39426) | 3199 (2326-4053) | 9.64 (7.43-11.35) | 2228 (1609-2907) | 6.70 (5.08-8.15) | 1364 (965-1809) | 4.07 (3.03-5.14) | 480 (324-658) | 1.44 (1.04-1.88) | 264 (172-385) | 0.79 (0.55-1.10) |
| Honduras | Female | 24383 (22334-26534) | 2377 (1826-2924) | 9.79 (7.64-11.77) | 1639 (1219-2087) | 6.73 (5.11-8.33) | 1025 (749-1335) | 4.20 (3.10-5.41) | 658 (479-855) | 2.70 (1.99-3.45) | 615 (443-808) | 2.53 (1.87-3.24) |
| Honduras | Male | 19109 (17553-20570) | 2766 (2179-3292) | 14.48 (11.60-17.04) | 1836 (1442-2257) | 9.62 (7.57-11.61) | 1155 (882-1437) | 6.06 (4.69-7.42) | 529 (384-696) | 2.78 (2.07-3.61) | 469 (331-638) | 2.47 (1.76-3.27) |
| Hungary | Female | 26749 (25160-28365) | 5298 (4422-6108) | 19.82 (16.68-22.55) | 2929 (2394-3537) | 11.00 (8.94-13.07) | 2847 (2252-3424) | 10.65 (8.59-12.71) | 1593 (1219-2033) | 5.95 (4.67-7.47) | 1425 (1081-1877) | 5.34 (4.08-6.87) |
| Hungary | Male | 18694 (17535-19902) | 4397 (3783-4994) | 23.54 (20.56-26.22) | 2619 (2141-3140) | 14.06 (11.58-16.50) | 2533 (2060-3051) | 13.58 (11.17-16.14) | 1187 (912-1500) | 6.40 (4.92-7.96) | 1085 (834-1379) | 5.81 (4.49-7.22) |
| Iceland | Female | 436 (412-460) | 31 (22-40) | 7.13 (5.00-9.25) | 7 (4-10) | 1.53 (0.91-2.32) | 28 (20-37) | 6.47 (4.59-8.33) | 3 (2-5) | 0.74 (0.42-1.19) | 3 (2-5) | 0.67 (0.37-1.09) |
| Iceland | Male | 435 (413-456) | 57 (48-67) | 13.19 (11.07-15.43) | 11 (7-16) | 2.54 (1.64-3.66) | 50 (41-59) | 11.49 (9.41-13.51) | 6 (3-9) | 1.31 (0.80-1.94) | 5 (3-8) | 1.23 (0.75-1.88) |
| India | Female | 2646287 (2497645-2786353) | 242446 (187039-286433) | 9.20 (7.11-10.73) | 168957 (128133-205662) | 6.39 (4.85-7.75) | 104824 (79381-131900) | 3.97 (3.01-4.95) | 31406 (23092-39841) | 1.19 (0.88-1.50) | 27443 (20074-34749) | 1.03 (0.76-1.30) |
| India | Male | 3522632 (3328099-3718498) | 415098 (322433-492899) | 11.81 (9.21-13.90) | 281402 (215560-339428) | 8.00 (6.23-9.55) | 175525 (135645-214069) | 4.98 (3.84-6.06) | 41032 (31086-51310) | 1.17 (0.89-1.43) | 34009 (25411-42402) | 0.97 (0.73-1.20) |
| Indonesia | Female | 776435 (722108-832663) | 103189 (82899-123293) | 13.30 (10.75-15.77) | 68039 (52909-83454) | 8.78 (6.92-10.61) | 44513 (35016-54675) | 5.76 (4.56-6.99) | 15366 (11940-19780) | 1.98 (1.54-2.52) | 8317 (6116-10970) | 1.07 (0.79-1.40) |
| Indonesia | Male | 1022441 (944021-1093372) | 153926 (123900-184217) | 15.10 (12.24-17.71) | 101954 (79106-124703) | 9.99 (7.87-12.04) | 67255 (51687-83490) | 6.56 (5.21-8.11) | 15015 (10904-19396) | 1.47 (1.08-1.90) | 8466 (6078-11117) | 0.83 (0.61-1.08) |
| Iran | Female | 164284 (155972-173933) | 679 (51-2099) | 0.41 (0.03-1.24) | 621 (51-1879) | 0.38 (0.03-1.16) | 524 (46-1572) | 0.32 (0.03-0.95) | 124 (1-663) | 0.08 (0.00-0.41) | 105 (1-567) | 0.06 (0.00-0.35) |
| Iran | Male | 174124 (164020-183097) | 7818 (4926-10935) | 4.50 (2.87-6.19) | 5947 (3828-8199) | 3.42 (2.23-4.67) | 4110 (2758-5678) | 2.35 (1.58-3.24) | 1212 (679-1845) | 0.70 (0.39-1.06) | 976 (548-1486) | 0.57 (0.31-0.85) |
| Iraq | Female | 103378 (94407-112835) | 578 (146-1430) | 0.55 (0.14-1.37) | 517 (128-1265) | 0.50 (0.12-1.21) | 425 (94-1025) | 0.41 (0.09-0.97) | 130 (12-493) | 0.12 (0.01-0.48) | 92 (6-386) | 0.09 (0.01-0.37) |
| Iraq | Male | 109069 (98034-120043) | 5789 (3689-8010) | 5.31 (3.40-7.20) | 4363 (2769-5922) | 4.00 (2.59-5.34) | 3016 (2013-4099) | 2.78 (1.92-3.71) | 1445 (817-2133) | 1.34 (0.76-1.94) | 1071 (596-1638) | 0.98 (0.56-1.48) |
| Ireland | Female | 8746 (8243-9326) | 256 (119-414) | 2.93 (1.38-4.70) | 34 (17-68) | 0.38 (0.19-0.77) | 253 (119-407) | 2.91 (1.36-4.61) | 12 (5-25) | 0.13 (0.06-0.28) | 10 (4-23) | 0.12 (0.05-0.26) |
| Ireland | Male | 9853 (9291-10397) | 981 (762-1189) | 9.99 (7.78-11.92) | 75 (51-134) | 0.76 (0.52-1.34) | 937 (731-1145) | 9.53 (7.48-11.47) | 24 (15-47) | 0.24 (0.16-0.48) | 21 (14-44) | 0.22 (0.14-0.43) |
| Israel | Female | 23426 (21824-25031) | 1750 (1272-2271) | 7.47 (5.47-9.58) | 596 (412-810) | 2.57 (1.78-3.47) | 1446 (1055-1857) | 6.18 (4.53-7.88) | 261 (169-373) | 1.12 (0.71-1.59) | 235 (151-339) | 1.01 (0.64-1.43) |
| Israel | Male | 27567 (25880-29226) | 3212 (2608-3767) | 11.67 (9.62-13.57) | 1003 (734-1291) | 3.65 (2.68-4.66) | 2498 (2019-3029) | 9.06 (7.31-10.84) | 400 (279-541) | 1.45 (1.02-1.94) | 369 (255-505) | 1.34 (0.93-1.80) |
| Italy | Female | 166148 (154218-177375) | 15493 (11002-19946) | 9.35 (6.74-11.81) | 6911 (4917-9260) | 4.17 (3.02-5.57) | 10725 (7827-13961) | 6.48 (4.80-8.35) | 3462 (2352-4894) | 2.09 (1.44-2.90) | 3218 (2166-4550) | 1.93 (1.32-2.69) |
| Italy | Male | 158983 (149601-168672) | 20440 (16551-24570) | 12.85 (10.42-15.15) | 8832 (6764-11301) | 5.55 (4.28-6.97) | 13687 (10891-17116) | 8.66 (6.92-10.70) | 4143 (3050-5380) | 2.61 (1.93-3.38) | 3980 (2910-5248) | 2.50 (1.84-3.24) |
| Jamaica | Female | 11542 (10687-12376) | 732 (531-936) | 6.37 (4.62-8.03) | 522 (384-672) | 4.53 (3.31-5.76) | 357 (263-461) | 3.09 (2.33-3.98) | 252 (181-331) | 2.18 (1.58-2.84) | 217 (151-286) | 1.88 (1.32-2.45) |
| Jamaica | Male | 13108 (12086-14054) | 1312 (1050-1584) | 10.01 (8.11-11.90) | 878 (683-1086) | 6.70 (5.30-8.23) | 590 (461-737) | 4.51 (3.61-5.59) | 298 (227-380) | 2.27 (1.76-2.89) | 241 (181-316) | 1.83 (1.40-2.38) |
| Japan | Female | 417167 (387272-448918) | 60670 (49584-73217) | 14.58 (12.11-17.24) | 24708 (18325-31628) | 5.96 (4.43-7.53) | 42235 (33478-52734) | 10.16 (8.16-12.45) | 11107 (8049-14720) | 2.67 (1.93-3.48) | 10225 (7344-13428) | 2.45 (1.77-3.25) |
| Japan | Male | 501264 (473542-529261) | 85096 (69832-98099) | 17.02 (14.08-19.43) | 33136 (25689-41725) | 6.62 (5.21-8.23) | 57844 (46454-69173) | 11.54 (9.39-13.67) | 16109 (12229-20732) | 3.23 (2.47-4.12) | 14171 (10697-18343) | 2.83 (2.15-3.61) |
| Jordan | Female | 24023 (21997-25788) | 109 (22-306) | 0.45 (0.09-1.25) | 97 (19-275) | 0.40 (0.08-1.16) | 79 (13-221) | 0.33 (0.05-0.93) | 21 (1-115) | 0.09 (0.00-0.48) | 19 (0-108) | 0.08 (0.00-0.46) |
| Jordan | Male | 24932 (23075-26671) | 1253 (843-1747) | 5.02 (3.42-6.89) | 942 (632-1303) | 3.78 (2.56-5.07) | 644 (433-874) | 2.57 (1.76-3.44) | 270 (155-394) | 1.08 (0.62-1.58) | 245 (137-361) | 0.98 (0.57-1.43) |
| Kazakhstan | Female | 35563 (33644-37536) | 3024 (2388-3764) | 8.52 (6.73-10.36) | 1977 (1538-2496) | 5.57 (4.41-6.94) | 1547 (1200-1954) | 4.34 (3.41-5.39) | 959 (719-1233) | 2.71 (2.02-3.45) | 872 (640-1126) | 2.44 (1.81-3.13) |
| Kazakhstan | Male | 39550 (37226-41689) | 5107 (4234-6046) | 12.94 (10.79-15.17) | 3183 (2592-3799) | 8.07 (6.55-9.57) | 2441 (1967-3039) | 6.20 (5.03-7.57) | 1254 (969-1539) | 3.17 (2.49-3.84) | 1057 (822-1316) | 2.68 (2.09-3.29) |
| Kenya | Female | 89681 (82828-96002) | 10203 (8321-12194) | 11.35 (9.35-13.44) | 6967 (5587-8557) | 7.79 (6.27-9.41) | 4294 (3422-5362) | 4.80 (3.86-5.87) | 1666 (1255-2187) | 1.85 (1.41-2.41) | 804 (567-1093) | 0.90 (0.64-1.21) |
| Kenya | Male | 104069 (95720-112207) | 71 (0-604) | 0.07 (0.00-0.57) | 71 (0-604) | 0.07 (0.00-0.57) | 70 (0-516) | 0.07 (0.00-0.50) | 9 (0-107) | 0.01 (0.00-0.10) | 4 (0-47) | 0.00 (0.00-0.04) |
| Kiribati | Female | 503 (445-560) | 38 (24-52) | 7.56 (4.82-10.14) | 27 (17-37) | 5.33 (3.41-7.09) | 16 (10-23) | 3.27 (2.09-4.51) | 5 (3-8) | 1.08 (0.67-1.55) | 3 (1-4) | 0.55 (0.30-0.86) |
| Kiribati | Male | 468 (386-539) | 42 (29-56) | 9.08 (6.46-11.46) | 29 (20-40) | 6.37 (4.45-8.17) | 18 (13-25) | 3.94 (2.77-5.21) | 5 (3-7) | 1.05 (0.69-1.48) | 3 (2-4) | 0.59 (0.37-0.87) |
| Kuwait | Female | 4678 (4424-4977) | 185 (114-265) | 3.96 (2.41-5.62) | 104 (67-143) | 2.22 (1.43-3.02) | 138 (88-191) | 2.97 (1.89-4.06) | 35 (22-49) | 0.75 (0.46-1.03) | 33 (20-46) | 0.70 (0.42-0.98) |
| Kuwait | Male | 6134 (5725-6547) | 584 (454-705) | 9.53 (7.42-11.37) | 279 (209-355) | 4.53 (3.44-5.64) | 384 (294-470) | 6.27 (4.86-7.60) | 89 (66-114) | 1.44 (1.07-1.83) | 80 (60-104) | 1.30 (0.98-1.65) |
| Kyrgyzstan | Female | 10366 (9806-10929) | 681 (480-876) | 6.55 (4.60-8.39) | 493 (346-638) | 4.76 (3.36-6.12) | 303 (213-395) | 2.92 (2.04-3.79) | 184 (131-245) | 1.77 (1.27-2.33) | 161 (116-215) | 1.55 (1.12-2.08) |
| Kyrgyzstan | Male | 11874 (11120-12572) | 1305 (958-1621) | 11.02 (8.27-13.56) | 902 (647-1149) | 7.61 (5.51-9.52) | 547 (396-692) | 4.60 (3.37-5.86) | 179 (129-232) | 1.51 (1.09-1.95) | 125 (89-164) | 1.06 (0.76-1.38) |
| Laos | Female | 27010 (24403-29407) | 3221 (2444-3995) | 11.91 (9.07-14.63) | 2160 (1607-2755) | 8.02 (6.07-9.98) | 1358 (994-1753) | 5.04 (3.76-6.43) | 438 (303-578) | 1.62 (1.15-2.15) | 316 (207-444) | 1.17 (0.79-1.59) |
| Laos | Male | 28889 (25734-32006) | 4160 (3228-5157) | 14.43 (11.30-17.13) | 2811 (2154-3509) | 9.79 (7.61-11.84) | 1760 (1335-2265) | 6.14 (4.71-7.66) | 420 (300-584) | 1.46 (1.08-1.98) | 297 (202-424) | 1.03 (0.71-1.47) |
| Latvia | Female | 5990 (5655-6329) | 230 (119-349) | 3.82 (2.07-5.84) | 174 (100-251) | 2.92 (1.70-4.16) | 176 (100-256) | 2.93 (1.71-4.23) | 87 (47-132) | 1.46 (0.78-2.19) | 72 (38-109) | 1.20 (0.63-1.82) |
| Latvia | Male | 3273 (3091-3448) | 423 (350-493) | 12.95 (10.79-14.99) | 232 (185-278) | 7.12 (5.68-8.41) | 238 (194-283) | 7.24 (5.95-8.69) | 90 (69-113) | 2.77 (2.10-3.44) | 72 (53-91) | 2.19 (1.63-2.77) |
| Lebanon | Female | 17956 (16544-19359) | 65 (15-190) | 0.36 (0.09-1.06) | 58 (15-176) | 0.32 (0.08-0.98) | 48 (11-139) | 0.27 (0.06-0.78) | 11 (0-64) | 0.06 (0.00-0.36) | 10 (0-59) | 0.05 (0.00-0.33) |
| Lebanon | Male | 19960 (18284-21384) | 848 (537-1228) | 4.29 (2.70-6.07) | 655 (420-939) | 3.30 (2.12-4.66) | 462 (295-649) | 2.31 (1.49-3.30) | 198 (105-314) | 0.99 (0.54-1.58) | 185 (104-294) | 0.94 (0.52-1.49) |
| Lesotho | Female | 6588 (6006-7164) | 393 (271-534) | 6.02 (4.19-8.01) | 304 (208-405) | 4.62 (3.20-6.11) | 189 (132-253) | 2.87 (2.02-3.75) | 99 (63-141) | 1.51 (0.97-2.11) | 89 (57-127) | 1.36 (0.87-1.90) |
| Lesotho | Male | 6600 (5936-7211) | 385 (251-523) | 5.83 (3.86-7.87) | 288 (185-392) | 4.35 (2.91-5.82) | 181 (118-242) | 2.72 (1.82-3.62) | 41 (23-63) | 0.62 (0.36-0.95) | 29 (16-45) | 0.44 (0.25-0.69) |
| Liberia | Female | 11322 (10394-12301) | 786 (595-983) | 6.95 (5.34-8.58) | 570 (423-714) | 5.03 (3.84-6.22) | 338 (255-435) | 3.00 (2.28-3.86) | 133 (95-177) | 1.17 (0.86-1.56) | 83 (55-117) | 0.73 (0.50-1.01) |
| Liberia | Male | 15258 (13860-16690) | 1161 (878-1438) | 7.60 (5.84-9.39) | 834 (624-1042) | 5.45 (4.09-6.81) | 496 (369-635) | 3.24 (2.45-4.15) | 143 (98-194) | 0.93 (0.65-1.26) | 98 (64-139) | 0.64 (0.43-0.91) |
| Libya | Female | 28262 (25610-30855) | 165 (50-396) | 0.58 (0.18-1.38) | 143 (43-347) | 0.51 (0.15-1.24) | 117 (35-300) | 0.42 (0.12-1.05) | 23 (1-112) | 0.08 (0.00-0.40) | 18 (0-90) | 0.06 (0.00-0.32) |
| Libya | Male | 26445 (24039-29116) | 1370 (939-1909) | 5.20 (3.45-6.92) | 1020 (714-1379) | 3.84 (2.69-5.07) | 725 (517-999) | 2.74 (1.97-3.65) | 230 (135-341) | 0.87 (0.52-1.27) | 177 (100-272) | 0.67 (0.38-1.00) |
| Lithuania | Female | 6824 (6423-7269) | 666 (528-814) | 9.74 (7.80-11.74) | 352 (269-446) | 5.14 (4.02-6.47) | 393 (310-486) | 5.76 (4.51-7.05) | 177 (131-232) | 2.59 (1.94-3.37) | 143 (102-193) | 2.08 (1.50-2.81) |
| Lithuania | Male | 4585 (4315-4850) | 462 (377-551) | 10.14 (8.24-11.94) | 245 (194-305) | 5.37 (4.24-6.57) | 272 (218-340) | 5.93 (4.77-7.36) | 106 (80-136) | 2.31 (1.75-2.95) | 74 (54-98) | 1.60 (1.16-2.13) |
| Luxembourg | Female | 1611 (1501-1726) | 122 (84-159) | 7.56 (5.31-9.77) | 8 (6-12) | 0.50 (0.37-0.72) | 119 (82-154) | 7.37 (5.23-9.54) | 3 (2-5) | 0.20 (0.14-0.30) | 3 (2-4) | 0.18 (0.12-0.27) |
| Luxembourg | Male | 1355 (1270-1434) | 159 (128-190) | 11.72 (9.63-13.85) | 10 (8-13) | 0.71 (0.56-0.99) | 153 (123-184) | 11.28 (9.32-13.37) | 4 (3-6) | 0.29 (0.22-0.42) | 4 (3-5) | 0.28 (0.21-0.41) |
| Macedonia | Female | 4889 (4512-5257) | 787 (645-942) | 16.18 (13.49-18.89) | 509 (403-622) | 10.36 (8.39-12.54) | 358 (283-448) | 7.32 (5.90-8.96) | 256 (196-328) | 5.21 (4.04-6.65) | 228 (169-299) | 4.65 (3.50-6.07) |
| Macedonia | Male | 5673 (5138-6149) | 1170 (974-1374) | 20.78 (17.55-23.63) | 778 (626-949) | 13.75 (11.21-16.32) | 558 (444-687) | 9.90 (7.90-11.81) | 329 (247-429) | 5.84 (4.42-7.60) | 282 (208-374) | 4.97 (3.69-6.49) |
| Madagascar | Female | 36836 (33696-40000) | 3818 (2933-4617) | 10.35 (7.99-12.34) | 2678 (2001-3292) | 7.25 (5.53-8.75) | 1573 (1167-1996) | 4.26 (3.23-5.34) | 559 (387-758) | 1.50 (1.07-2.04) | 289 (187-424) | 0.78 (0.50-1.15) |
| Madagascar | Male | 39144 (35189-42889) | 3107 (2376-4036) | 7.95 (6.16-10.03) | 2232 (1670-2901) | 5.75 (4.34-7.24) | 1341 (1001-1791) | 3.42 (2.61-4.42) | 326 (224-480) | 0.84 (0.57-1.21) | 178 (105-275) | 0.45 (0.27-0.71) |
| Malawi | Female | 35131 (32378-38042) | 3940 (3134-4730) | 11.24 (9.05-13.27) | 2724 (2122-3328) | 7.81 (6.13-9.34) | 1628 (1232-2028) | 4.62 (3.56-5.67) | 771 (575-981) | 2.20 (1.68-2.75) | 541 (392-714) | 1.54 (1.14-1.99) |
| Malawi | Male | 47077 (43062-51465) | 3793 (2891-4834) | 8.04 (6.21-9.96) | 2701 (2037-3501) | 5.75 (4.40-7.29) | 1619 (1225-2157) | 3.46 (2.62-4.42) | 397 (283-551) | 0.84 (0.61-1.16) | 275 (193-395) | 0.59 (0.42-0.84) |
| Malaysia | Female | 108522 (100729-116153) | 15803 (13090-19020) | 14.63 (12.23-17.19) | 9692 (7811-11880) | 8.90 (7.23-10.74) | 7629 (6146-9401) | 7.05 (5.74-8.47) | 3330 (2627-4315) | 3.08 (2.44-3.87) | 3026 (2361-3937) | 2.80 (2.21-3.54) |
| Malaysia | Male | 119830 (110403-129335) | 19826 (16411-23045) | 16.63 (13.90-19.03) | 12097 (9680-14571) | 10.15 (8.23-12.08) | 9529 (7683-11733) | 7.99 (6.50-9.72) | 3501 (2695-4407) | 2.93 (2.26-3.63) | 3188 (2426-4010) | 2.66 (2.07-3.32) |
| Maldives | Female | 976 (908-1042) | 131 (107-156) | 13.43 (11.10-15.87) | 81 (66-99) | 8.30 (6.78-10.02) | 62 (49-77) | 6.38 (5.14-7.82) | 22 (17-29) | 2.29 (1.74-2.94) | 17 (13-23) | 1.79 (1.29-2.36) |
| Maldives | Male | 1659 (1532-1781) | 231 (186-273) | 13.88 (11.28-16.26) | 143 (114-174) | 8.63 (6.84-10.42) | 111 (87-136) | 6.69 (5.27-8.08) | 27 (21-35) | 1.64 (1.23-2.12) | 21 (15-27) | 1.24 (0.92-1.67) |
| Mali | Female | 36782 (33869-39382) | 2436 (1724-3071) | 6.64 (4.79-8.29) | 1752 (1262-2259) | 4.76 (3.47-6.04) | 1054 (747-1368) | 2.86 (2.04-3.68) | 505 (348-681) | 1.37 (0.95-1.85) | 355 (233-495) | 0.96 (0.64-1.35) |
| Mali | Male | 43949 (39341-48038) | 3088 (2201-3955) | 7.06 (5.02-8.90) | 2200 (1555-2828) | 5.05 (3.59-6.44) | 1313 (930-1744) | 3.01 (2.18-3.95) | 455 (296-655) | 1.04 (0.68-1.48) | 328 (205-488) | 0.75 (0.47-1.11) |
| Malta | Female | 1519 (1415-1619) | 141 (108-177) | 9.29 (7.13-11.49) | 65 (49-83) | 4.28 (3.26-5.43) | 96 (75-123) | 6.35 (4.91-8.01) | 29 (21-39) | 1.91 (1.38-2.54) | 28 (20-38) | 1.87 (1.35-2.49) |
| Malta | Male | 1266 (1193-1348) | 163 (133-193) | 12.88 (10.59-15.12) | 72 (56-91) | 5.68 (4.48-7.09) | 108 (86-130) | 8.50 (6.78-10.22) | 33 (25-43) | 2.60 (1.98-3.39) | 32 (24-42) | 2.53 (1.93-3.31) |
| Marshall Islands | Female | 308 (219-387) | 21 (12-33) | 6.99 (4.24-9.96) | 14 (8-23) | 4.65 (2.82-6.73) | 10 (5-16) | 3.27 (1.99-4.79) | 4 (2-6) | 1.16 (0.67-1.75) | 3 (1-4) | 0.84 (0.49-1.32) |
| Marshall Islands | Male | 307 (204-411) | 27 (13-44) | 8.97 (5.44-12.73) | 18 (9-29) | 5.99 (3.71-8.56) | 13 (6-21) | 4.16 (2.45-6.00) | 3 (1-6) | 1.11 (0.58-1.77) | 2 (1-4) | 0.79 (0.39-1.27) |
| Mauritania | Female | 9674 (8782-10533) | 730 (545-914) | 7.57 (5.70-9.31) | 515 (385-659) | 5.35 (3.98-6.65) | 318 (234-409) | 3.30 (2.45-4.18) | 140 (94-194) | 1.45 (0.99-1.97) | 90 (57-134) | 0.94 (0.58-1.38) |
| Mauritania | Male | 9395 (8491-10274) | 763 (572-949) | 8.13 (6.18-9.95) | 538 (404-676) | 5.71 (4.36-7.12) | 331 (247-426) | 3.53 (2.70-4.45) | 99 (65-140) | 1.05 (0.70-1.49) | 67 (41-103) | 0.72 (0.45-1.07) |
| Mauritius | Female | 17121 (16194-18222) | 2489 (2027-2974) | 14.49 (11.92-17.23) | 1550 (1262-1901) | 9.04 (7.36-10.98) | 1166 (918-1433) | 6.80 (5.36-8.35) | 617 (499-774) | 3.61 (2.90-4.48) | 593 (474-743) | 3.48 (2.76-4.30) |
| Mauritius | Male | 20388 (19063-21633) | 3281 (2633-3918) | 16.14 (13.06-19.01) | 2058 (1625-2568) | 10.12 (8.02-12.38) | 1546 (1183-1954) | 7.59 (5.91-9.44) | 697 (535-907) | 3.43 (2.64-4.40) | 653 (492-847) | 3.20 (2.43-4.12) |
| Mexico | Female | 898932 (831679-964352) | 76780 (57634-95230) | 8.57 (6.40-10.49) | 48532 (35690-61553) | 5.42 (4.02-6.77) | 37597 (27669-48138) | 4.18 (3.11-5.28) | 16371 (12279-21123) | 1.83 (1.36-2.33) | 14908 (11052-19223) | 1.67 (1.23-2.13) |
| Mexico | Male | 960249 (901261-1021949) | 105282 (83938-125582) | 11.01 (8.75-12.88) | 66507 (52311-82314) | 6.93 (5.44-8.41) | 52018 (40079-63869) | 5.43 (4.23-6.59) | 15120 (11620-19197) | 1.58 (1.22-1.95) | 13377 (10160-17149) | 1.40 (1.05-1.76) |
| Moldova | Female | 7166 (6695-7679) | 246 (136-385) | 3.43 (1.90-5.32) | 209 (122-313) | 2.92 (1.69-4.33) | 158 (92-232) | 2.21 (1.31-3.12) | 92 (48-146) | 1.28 (0.67-1.99) | 70 (34-111) | 0.97 (0.48-1.53) |
| Moldova | Male | 5034 (4742-5345) | 493 (389-582) | 9.80 (7.77-11.50) | 331 (261-402) | 6.58 (5.17-8.04) | 223 (172-276) | 4.43 (3.46-5.49) | 111 (82-146) | 2.22 (1.65-2.84) | 69 (49-93) | 1.36 (0.98-1.86) |
| Mongolia | Female | 5570 (5204-5902) | 434 (331-538) | 7.79 (6.00-9.60) | 301 (229-377) | 5.41 (4.10-6.73) | 201 (152-253) | 3.61 (2.73-4.54) | 111 (83-141) | 2.00 (1.50-2.51) | 97 (73-125) | 1.75 (1.31-2.23) |
| Mongolia | Male | 6466 (5974-6982) | 826 (664-998) | 12.79 (10.24-15.17) | 545 (435-669) | 8.42 (6.79-10.10) | 361 (280-453) | 5.56 (4.40-6.86) | 162 (127-203) | 2.51 (1.98-3.10) | 119 (92-149) | 1.83 (1.44-2.30) |
| Montenegro | Female | 1647 (1524-1773) | 279 (227-332) | 17.05 (14.08-19.82) | 175 (137-216) | 10.61 (8.54-12.79) | 133 (103-161) | 8.06 (6.47-9.75) | 90 (69-117) | 5.50 (4.24-6.93) | 81 (60-109) | 4.92 (3.65-6.45) |
| Montenegro | Male | 2081 (1906-2251) | 437 (371-508) | 21.01 (18.06-23.60) | 282 (231-339) | 13.57 (11.33-15.90) | 215 (174-269) | 10.37 (8.49-12.62) | 120 (93-155) | 5.79 (4.49-7.27) | 102 (76-135) | 4.91 (3.70-6.47) |
| Morocco | Female | 149872 (136259-164775) | 744 (201-2022) | 0.49 (0.13-1.32) | 682 (148-1897) | 0.45 (0.10-1.23) | 558 (114-1508) | 0.37 (0.07-1.02) | 117 (4-531) | 0.08 (0.00-0.35) | 88 (2-432) | 0.06 (0.00-0.29) |
| Morocco | Male | 125289 (111992-138184) | 5963 (3656-8707) | 4.81 (2.97-6.84) | 4592 (2938-6491) | 3.68 (2.35-5.12) | 3150 (1996-4378) | 2.52 (1.60-3.48) | 931 (509-1467) | 0.74 (0.41-1.15) | 620 (329-1008) | 0.50 (0.27-0.79) |
| Mozambique | Female | 38286 (35221-41279) | 4478 (3670-5404) | 11.68 (9.75-13.75) | 3103 (2493-3782) | 8.15 (6.65-9.71) | 1844 (1445-2272) | 4.84 (3.89-5.86) | 612 (455-801) | 1.60 (1.19-2.09) | 325 (227-459) | 0.85 (0.61-1.18) |
| Mozambique | Male | 56769 (51200-62496) | 4259 (3239-5486) | 7.53 (5.81-9.35) | 3118 (2333-4008) | 5.50 (4.18-6.91) | 1850 (1359-2437) | 3.28 (2.48-4.20) | 350 (244-491) | 0.61 (0.43-0.85) | 176 (110-263) | 0.31 (0.19-0.45) |
| Myanmar | Female | 190176 (175782-204620) | 23921 (19011-28955) | 12.60 (9.97-14.93) | 16290 (12755-19909) | 8.63 (6.75-10.35) | 9882 (7645-12351) | 5.19 (3.99-6.42) | 5131 (3924-6493) | 2.70 (2.10-3.41) | 3305 (2477-4349) | 1.75 (1.30-2.29) |
| Myanmar | Male | 166947 (153372-180884) | 24583 (19312-29718) | 14.82 (11.80-17.26) | 16822 (13066-20971) | 10.13 (8.00-12.15) | 10192 (7875-12840) | 6.13 (4.79-7.60) | 3936 (2945-5232) | 2.37 (1.77-3.06) | 2270 (1649-3049) | 1.36 (1.02-1.82) |
| Namibia | Female | 3816 (3511-4149) | 225 (153-303) | 5.90 (4.04-7.79) | 166 (114-217) | 4.37 (3.02-5.64) | 114 (79-150) | 2.99 (2.10-3.91) | 49 (31-69) | 1.28 (0.83-1.77) | 43 (26-60) | 1.12 (0.71-1.55) |
| Namibia | Male | 5722 (5213-6274) | 349 (239-473) | 6.14 (4.15-8.06) | 252 (171-349) | 4.44 (3.05-5.91) | 172 (119-237) | 3.03 (2.09-4.12) | 54 (32-82) | 0.95 (0.59-1.42) | 48 (27-73) | 0.83 (0.48-1.25) |
| Nauru | Female | 58 (52-64) | 5 (4-6) | 8.86 (6.86-10.81) | 3 (2-4) | 5.64 (4.32-7.01) | 3 (2-3) | 4.55 (3.49-5.69) | 1 (1-1) | 1.73 (1.26-2.35) | 1 (0-1) | 1.02 (0.69-1.49) |
| Nauru | Male | 55 (48-62) | 5 (4-7) | 10.03 (7.57-12.34) | 3 (2-5) | 6.29 (4.74-7.84) | 3 (2-4) | 5.07 (3.70-6.38) | 1 (1-1) | 1.73 (1.26-2.34) | 1 (0-1) | 1.15 (0.79-1.61) |
| Nepal | Female | 69116 (64028-74216) | 4875 (3113-6669) | 7.08 (4.52-9.48) | 3436 (2150-4797) | 5.01 (3.13-6.90) | 2097 (1298-2954) | 3.05 (1.91-4.21) | 597 (370-839) | 0.86 (0.54-1.18) | 412 (258-594) | 0.60 (0.37-0.84) |
| Nepal | Male | 78074 (72331-83738) | 8213 (6479-9917) | 10.55 (8.37-12.56) | 5761 (4460-7078) | 7.39 (5.73-9.02) | 3486 (2651-4376) | 4.46 (3.37-5.52) | 873 (646-1132) | 1.13 (0.84-1.44) | 503 (363-665) | 0.65 (0.47-0.85) |
| Netherlands | Female | 41147 (39039-43319) | 2859 (2011-3766) | 6.94 (4.88-9.11) | 846 (545-1247) | 2.06 (1.34-2.99) | 2488 (1740-3256) | 6.06 (4.28-7.86) | 333 (199-529) | 0.80 (0.49-1.26) | 277 (169-434) | 0.67 (0.41-1.07) |
| Netherlands | Male | 39672 (37734-41614) | 4477 (3565-5352) | 11.29 (9.11-13.31) | 1158 (776-1627) | 2.92 (1.96-4.10) | 3632 (2899-4403) | 9.19 (7.34-11.10) | 445 (282-671) | 1.12 (0.71-1.67) | 413 (259-620) | 1.04 (0.65-1.55) |
| New Zealand | Female | 9763 (9223-10278) | 600 (420-784) | 6.14 (4.24-7.93) | 227 (153-305) | 2.32 (1.57-3.09) | 493 (342-639) | 5.02 (3.51-6.51) | 89 (57-124) | 0.92 (0.59-1.28) | 79 (51-112) | 0.81 (0.53-1.14) |
| New Zealand | Male | 10077 (9500-10673) | 1077 (874-1270) | 10.71 (8.71-12.47) | 354 (267-470) | 3.51 (2.69-4.63) | 833 (652-1004) | 8.27 (6.55-9.85) | 128 (93-174) | 1.27 (0.93-1.73) | 114 (82-156) | 1.13 (0.82-1.55) |
| Nicaragua | Female | 28797 (26677-30862) | 3097 (2474-3665) | 10.77 (8.63-12.56) | 2132 (1667-2614) | 7.43 (5.85-9.00) | 1314 (1015-1667) | 4.56 (3.58-5.75) | 826 (616-1057) | 2.86 (2.17-3.63) | 764 (564-985) | 2.65 (1.99-3.40) |
| Nicaragua | Male | 51871 (47953-55697) | 7428 (5978-8943) | 14.34 (11.47-16.97) | 5001 (3941-6077) | 9.64 (7.69-11.51) | 3066 (2399-3847) | 5.93 (4.65-7.24) | 1210 (913-1572) | 2.34 (1.75-3.01) | 1089 (786-1452) | 2.10 (1.54-2.80) |
| Niger | Female | 21221 (19415-23002) | 1522 (1134-1919) | 7.17 (5.31-8.89) | 1085 (791-1402) | 5.14 (3.74-6.43) | 647 (476-840) | 3.06 (2.23-3.95) | 199 (127-276) | 0.94 (0.61-1.28) | 93 (55-145) | 0.44 (0.26-0.67) |
| Niger | Male | 32104 (28053-36087) | 2462 (1766-3106) | 7.68 (5.73-9.50) | 1764 (1269-2303) | 5.48 (4.04-6.95) | 1049 (746-1357) | 3.28 (2.41-4.19) | 193 (121-288) | 0.60 (0.38-0.89) | 127 (73-203) | 0.40 (0.23-0.61) |
| Nigeria | Female | 310704 (289964-330956) | 18368 (13110-23873) | 5.95 (4.29-7.58) | 13744 (10135-17626) | 4.42 (3.26-5.59) | 8727 (6329-11320) | 2.81 (2.04-3.62) | 3371 (2419-4517) | 1.09 (0.78-1.44) | 1928 (1328-2619) | 0.62 (0.43-0.84) |
| Nigeria | Male | 401202 (367193-434393) | 27460 (20243-34437) | 6.85 (4.98-8.50) | 19842 (14822-25182) | 4.97 (3.74-6.19) | 12404 (9281-15745) | 3.10 (2.33-3.87) | 3488 (2450-4624) | 0.87 (0.62-1.15) | 2419 (1721-3328) | 0.61 (0.42-0.82) |
| Norway | Female | 7634 (7155-8106) | 317 (177-480) | 4.13 (2.32-6.19) | 43 (24-84) | 0.56 (0.31-1.10) | 312 (174-466) | 4.07 (2.29-6.03) | 17 (9-38) | 0.23 (0.12-0.50) | 12 (6-27) | 0.16 (0.08-0.35) |
| Norway | Male | 9168 (8647-9682) | 1069 (861-1267) | 11.62 (9.49-13.60) | 94 (62-167) | 1.03 (0.68-1.83) | 1007 (804-1195) | 10.97 (8.87-12.93) | 40 (24-79) | 0.44 (0.27-0.85) | 36 (22-72) | 0.39 (0.24-0.78) |
| Oman | Female | 6741 (6209-7280) | 46 (11-112) | 0.69 (0.17-1.68) | 36 (8-92) | 0.54 (0.13-1.33) | 37 (8-90) | 0.55 (0.12-1.39) | 6 (0-28) | 0.09 (0.00-0.41) | 5 (0-26) | 0.08 (0.00-0.38) |
| Oman | Male | 10989 (9886-12136) | 617 (414-807) | 5.62 (3.84-7.38) | 387 (267-513) | 3.53 (2.48-4.61) | 384 (264-512) | 3.50 (2.41-4.58) | 77 (46-114) | 0.70 (0.43-1.01) | 66 (39-98) | 0.60 (0.37-0.89) |
| Pakistan | Female | 506475 (467015-544382) | 50729 (40338-60774) | 10.03 (8.05-11.85) | 35345 (27380-43274) | 7.02 (5.51-8.42) | 21566 (16368-27037) | 4.25 (3.25-5.24) | 9527 (7257-12215) | 1.89 (1.45-2.39) | 7947 (5890-10352) | 1.57 (1.17-2.01) |
| Pakistan | Male | 519803 (482759-556410) | 55825 (45632-67316) | 10.78 (8.86-12.65) | 38957 (31213-48145) | 7.50 (6.07-9.02) | 23604 (18576-29723) | 4.54 (3.58-5.64) | 6003 (4500-7851) | 1.15 (0.87-1.48) | 4528 (3341-5965) | 0.87 (0.65-1.15) |
| Palau | Female | 124 (111-137) | 14 (10-18) | 11.31 (8.78-13.87) | 9 (6-11) | 6.89 (5.18-8.75) | 7 (5-9) | 5.71 (4.32-7.32) | 3 (2-5) | 2.68 (1.99-3.57) | 3 (2-4) | 2.29 (1.66-3.11) |
| Palau | Male | 154 (136-172) | 16 (12-21) | 10.69 (8.16-13.08) | 10 (7-13) | 6.54 (4.89-8.24) | 8 (6-11) | 5.48 (4.06-6.93) | 3 (2-4) | 1.92 (1.40-2.52) | 2 (1-3) | 1.40 (0.97-1.88) |
| Panama | Female | 13785 (12912-14652) | 1513 (1233-1789) | 11.00 (9.08-12.82) | 903 (706-1098) | 6.54 (5.17-7.90) | 800 (628-980) | 5.80 (4.62-7.06) | 364 (277-461) | 2.63 (2.03-3.31) | 321 (240-409) | 2.33 (1.80-2.96) |
| Panama | Male | 20348 (19063-21601) | 3009 (2468-3573) | 14.85 (12.37-17.35) | 1742 (1405-2104) | 8.56 (6.98-10.23) | 1544 (1251-1884) | 7.60 (6.26-9.26) | 578 (449-723) | 2.84 (2.23-3.54) | 483 (369-629) | 2.39 (1.86-3.06) |
| Papua New Guinea | Female | 11153 (10167-12065) | 720 (484-937) | 6.51 (4.31-8.41) | 511 (330-681) | 4.57 (3.00-6.09) | 326 (217-442) | 2.94 (1.95-3.93) | 84 (50-124) | 0.75 (0.44-1.11) | 50 (27-81) | 0.45 (0.24-0.74) |
| Papua New Guinea | Male | 17084 (15458-18623) | 1342 (939-1772) | 7.88 (5.55-10.19) | 945 (650-1269) | 5.56 (3.87-7.23) | 603 (415-813) | 3.55 (2.38-4.64) | 101 (60-156) | 0.59 (0.35-0.90) | 66 (34-108) | 0.39 (0.20-0.62) |
| Paraguay | Female | 14867 (13737-15934) | 1570 (1288-1848) | 10.58 (8.69-12.19) | 1041 (830-1248) | 7.01 (5.62-8.35) | 714 (568-871) | 4.79 (3.79-5.84) | 481 (371-606) | 3.24 (2.51-4.00) | 387 (294-500) | 2.61 (2.02-3.33) |
| Paraguay | Male | 21215 (19289-23004) | 2743 (2196-3339) | 13.01 (10.60-15.49) | 1803 (1407-2223) | 8.54 (6.73-10.26) | 1242 (982-1562) | 5.89 (4.60-7.15) | 691 (526-907) | 3.28 (2.49-4.14) | 548 (410-720) | 2.59 (1.94-3.33) |
| Peru | Female | 117302 (108120-126197) | 10929 (8418-13214) | 9.36 (7.27-11.10) | 7230 (5388-8946) | 6.17 (4.66-7.51) | 5060 (3749-6414) | 4.34 (3.22-5.40) | 1753 (1297-2221) | 1.50 (1.13-1.86) | 1589 (1180-2020) | 1.36 (1.03-1.69) |
| Peru | Male | 112699 (101524-122511) | 13792 (10551-16796) | 12.20 (9.50-14.65) | 9007 (6828-11178) | 7.94 (6.14-9.72) | 6272 (4791-7902) | 5.56 (4.36-6.87) | 1633 (1222-2086) | 1.45 (1.11-1.83) | 1390 (1046-1782) | 1.23 (0.96-1.57) |
| Philippines | Female | 459577 (428616-489200) | 53206 (40103-66252) | 11.60 (8.81-14.22) | 35631 (26623-44710) | 7.75 (5.83-9.69) | 22878 (16949-29370) | 4.99 (3.72-6.30) | 9938 (7375-12952) | 2.18 (1.61-2.78) | 7353 (5380-9649) | 1.61 (1.17-2.09) |
| Philippines | Male | 576995 (534851-619128) | 79264 (59426-96246) | 13.76 (10.43-16.34) | 53283 (39761-66810) | 9.26 (6.91-11.38) | 34279 (25064-43097) | 5.95 (4.32-7.44) | 11023 (7845-14140) | 1.92 (1.35-2.40) | 7443 (5231-9871) | 1.30 (0.90-1.70) |
| Poland | Female | 70110 (66341-73998) | 10325 (8405-12109) | 14.75 (12.02-17.16) | 5853 (4620-7084) | 8.38 (6.66-10.02) | 5575 (4432-6791) | 7.93 (6.39-9.65) | 3479 (2700-4286) | 4.97 (3.89-6.09) | 3346 (2580-4159) | 4.78 (3.73-5.86) |
| Poland | Male | 63291 (59633-66798) | 11299 (9350-13029) | 17.88 (14.88-20.61) | 6397 (5142-7687) | 10.11 (8.18-12.32) | 6086 (4975-7409) | 9.62 (7.96-11.61) | 3109 (2467-3845) | 4.92 (3.90-6.05) | 2848 (2234-3562) | 4.50 (3.56-5.66) |
| Portugal | Female | 40656 (37819-43633) | 2020 (1196-3084) | 4.99 (3.00-7.45) | 1341 (820-1922) | 3.30 (2.02-4.70) | 1482 (882-2148) | 3.67 (2.19-5.28) | 688 (374-1055) | 1.70 (0.94-2.54) | 663 (361-1041) | 1.64 (0.91-2.53) |
| Portugal | Male | 35031 (32666-37400) | 3761 (2925-4620) | 10.72 (8.44-12.96) | 1995 (1534-2526) | 5.69 (4.42-7.06) | 2238 (1757-2792) | 6.36 (5.06-7.89) | 935 (672-1231) | 2.67 (1.94-3.48) | 886 (646-1161) | 2.53 (1.86-3.25) |
| Puerto Rico | Female | 21290 (20329-22365) | 1741 (1328-2136) | 8.15 (6.33-10.07) | 850 (631-1077) | 3.98 (2.99-5.03) | 1175 (890-1479) | 5.52 (4.27-6.91) | 406 (289-548) | 1.91 (1.37-2.55) | 343 (244-471) | 1.61 (1.15-2.20) |
| Puerto Rico | Male | 29047 (27643-30384) | 3203 (2552-3775) | 11.02 (8.86-12.92) | 1481 (1179-1855) | 5.11 (4.07-6.42) | 2097 (1651-2523) | 7.22 (5.70-8.64) | 534 (391-720) | 1.84 (1.36-2.47) | 449 (326-628) | 1.55 (1.13-2.16) |
| Qatar | Female | 2240 (2065-2405) | 9 (0-29) | 0.40 (0.00-1.29) | 4 (0-12) | 0.17 (0.00-0.55) | 9 (0-28) | 0.39 (0.00-1.25) | 1 (0-5) | 0.05 (0.00-0.24) | 1 (0-5) | 0.05 (0.00-0.22) |
| Qatar | Male | 4989 (4506-5449) | 266 (164-370) | 5.32 (3.33-7.56) | 61 (34-97) | 1.22 (0.69-1.94) | 240 (147-335) | 4.80 (3.06-6.63) | 13 (7-21) | 0.26 (0.14-0.42) | 11 (6-19) | 0.23 (0.12-0.37) |
| Romania | Female | 45055 (42808-47486) | 7725 (6419-9102) | 17.20 (14.17-20.25) | 4518 (3612-5484) | 10.06 (7.99-12.03) | 3965 (3172-4844) | 8.80 (7.11-10.80) | 2320 (1799-2899) | 5.17 (4.01-6.39) | 2055 (1596-2571) | 4.57 (3.56-5.67) |
| Romania | Male | 45472 (43392-47510) | 9581 (8059-10897) | 21.13 (17.74-23.87) | 5831 (4688-6915) | 12.84 (10.39-15.20) | 5107 (4170-6153) | 11.21 (9.11-13.46) | 2639 (2053-3240) | 5.83 (4.56-7.11) | 2369 (1837-2913) | 5.20 (4.08-6.41) |
| Russia | Female | 268046 (253376-283975) | 27090 (21145-32735) | 10.08 (7.90-12.02) | 16761 (13178-20751) | 6.24 (4.90-7.71) | 13708 (10547-17282) | 5.10 (3.99-6.34) | 8466 (6337-10707) | 3.16 (2.37-3.95) | 6095 (4444-7896) | 2.27 (1.68-2.91) |
| Russia | Male | 183089 (173468-192716) | 26045 (20921-30950) | 14.29 (11.53-16.56) | 15692 (12357-19012) | 8.58 (6.76-10.33) | 12837 (10040-15860) | 7.03 (5.54-8.56) | 6077 (4663-7543) | 3.32 (2.55-4.12) | 4255 (3210-5363) | 2.32 (1.78-2.90) |
| Rwanda | Female | 25905 (23434-28365) | 2896 (2241-3544) | 11.20 (8.77-13.39) | 2008 (1538-2491) | 7.78 (6.01-9.51) | 1190 (904-1525) | 4.61 (3.53-5.79) | 280 (187-394) | 1.08 (0.73-1.48) | 130 (77-197) | 0.50 (0.29-0.76) |
| Rwanda | Male | 27921 (25191-30326) | 2264 (1690-2919) | 8.14 (6.14-10.11) | 1621 (1188-2113) | 5.81 (4.39-7.31) | 978 (695-1293) | 3.50 (2.55-4.57) | 149 (94-220) | 0.53 (0.35-0.78) | 92 (54-142) | 0.33 (0.20-0.50) |
| Saint Kitts and Nevis | Female | 357 (331-382) | 24 (17-31) | 6.79 (4.72-8.63) | 15 (11-19) | 4.16 (3.01-5.37) | 14 (10-18) | 3.97 (2.83-5.09) | 7 (5-9) | 1.91 (1.32-2.55) | 6 (4-9) | 1.73 (1.17-2.35) |
| Saint Kitts and Nevis | Male | 522 (478-571) | 54 (41-65) | 10.29 (8.01-12.23) | 31 (23-39) | 5.88 (4.50-7.35) | 29 (21-37) | 5.63 (4.14-7.03) | 10 (7-14) | 1.91 (1.36-2.57) | 8 (6-11) | 1.53 (1.07-2.08) |
| Saint Lucia | Female | 995 (928-1061) | 66 (49-83) | 6.62 (4.99-8.26) | 44 (33-56) | 4.38 (3.32-5.49) | 35 (26-44) | 3.49 (2.64-4.36) | 21 (15-27) | 2.08 (1.54-2.70) | 18 (13-23) | 1.76 (1.30-2.35) |
| Saint Lucia | Male | 1290 (1205-1375) | 128 (101-155) | 9.93 (7.90-11.73) | 81 (63-101) | 6.24 (4.91-7.70) | 63 (50-80) | 4.93 (3.86-6.07) | 25 (19-33) | 1.97 (1.50-2.52) | 19 (14-25) | 1.44 (1.06-1.88) |
| Saint Vincent and the Grenadines | Female | 604 (569-646) | 37 (26-48) | 6.15 (4.38-7.82) | 26 (19-33) | 4.24 (3.16-5.42) | 19 (14-25) | 3.14 (2.31-4.07) | 11 (8-15) | 1.81 (1.29-2.42) | 9 (6-12) | 1.50 (1.06-2.00) |
| Saint Vincent and the Grenadines | Male | 781 (730-836) | 78 (60-94) | 9.98 (7.93-11.85) | 50 (38-63) | 6.47 (5.01-7.91) | 37 (28-47) | 4.72 (3.67-6.00) | 15 (11-20) | 1.91 (1.40-2.48) | 12 (8-15) | 1.48 (1.08-1.95) |
| Samoa | Female | 959 (865-1053) | 28 (18-40) | 2.92 (1.95-4.17) | 21 (13-30) | 2.15 (1.34-3.16) | 14 (8-21) | 1.50 (0.88-2.22) | 5 (3-8) | 0.51 (0.28-0.84) | 2 (1-3) | 0.20 (0.08-0.35) |
| Samoa | Male | 779 (706-847) | 30 (20-42) | 3.81 (2.63-5.29) | 22 (15-31) | 2.79 (1.87-3.93) | 15 (10-21) | 1.92 (1.27-2.73) | 4 (3-7) | 0.57 (0.36-0.86) | 3 (2-5) | 0.38 (0.22-0.62) |
| Sao Tome and Principe | Female | 983 (911-1051) | 76 (59-95) | 7.77 (6.08-9.53) | 54 (42-68) | 5.49 (4.32-6.85) | 34 (25-43) | 3.44 (2.63-4.27) | 18 (13-23) | 1.82 (1.37-2.34) | 11 (8-16) | 1.17 (0.86-1.57) |
| Sao Tome and Principe | Male | 809 (735-888) | 66 (51-82) | 8.18 (6.38-9.86) | 46 (36-59) | 5.73 (4.51-7.15) | 29 (22-37) | 3.57 (2.76-4.43) | 9 (7-12) | 1.12 (0.82-1.47) | 5 (4-7) | 0.66 (0.45-0.90) |
| Saudi Arabia | Female | 179271 (162993-194362) | 841 (28-2152) | 0.47 (0.02-1.19) | 658 (28-1693) | 0.37 (0.02-0.94) | 701 (28-1779) | 0.40 (0.02-0.99) | 81 (1-375) | 0.05 (0.00-0.21) | 66 (1-315) | 0.04 (0.00-0.17) |
| Saudi Arabia | Male | 219548 (199741-237607) | 11101 (6907-16031) | 5.07 (3.23-7.09) | 6636 (4207-9180) | 3.03 (1.96-4.13) | 7376 (4768-10232) | 3.37 (2.21-4.59) | 1241 (736-1852) | 0.57 (0.34-0.84) | 995 (578-1510) | 0.46 (0.27-0.69) |
| Senegal | Female | 32428 (29793-35222) | 2456 (1825-3075) | 7.57 (5.72-9.30) | 1744 (1309-2201) | 5.38 (4.09-6.66) | 1076 (781-1381) | 3.30 (2.45-4.21) | 443 (310-606) | 1.37 (0.97-1.83) | 235 (154-347) | 0.73 (0.47-1.05) |
| Senegal | Male | 43356 (39126-47635) | 3378 (2500-4377) | 7.84 (5.90-9.75) | 2395 (1787-3112) | 5.53 (4.20-7.04) | 1454 (1088-1920) | 3.36 (2.53-4.30) | 396 (272-576) | 0.91 (0.62-1.31) | 224 (146-344) | 0.52 (0.34-0.80) |
| Serbia | Female | 25034 (23204-26746) | 4460 (3687-5212) | 17.87 (14.83-20.49) | 2777 (2226-3371) | 11.14 (9.04-13.28) | 2092 (1670-2560) | 8.35 (6.78-10.04) | 1619 (1250-1996) | 6.49 (5.07-7.94) | 1521 (1170-1910) | 6.10 (4.77-7.50) |
| Serbia | Male | 32020 (29787-34170) | 6941 (5852-7988) | 21.76 (18.65-24.53) | 4488 (3699-5423) | 14.08 (11.66-16.58) | 3392 (2737-4165) | 10.64 (8.64-12.91) | 2098 (1648-2622) | 6.58 (5.23-8.08) | 1906 (1498-2422) | 5.98 (4.76-7.47) |
| Seychelles | Female | 613 (569-658) | 73 (58-88) | 11.95 (9.75-14.21) | 44 (34-55) | 7.18 (5.71-8.77) | 37 (29-46) | 5.96 (4.74-7.36) | 22 (17-27) | 3.53 (2.77-4.37) | 20 (15-25) | 3.23 (2.49-4.02) |
| Seychelles | Male | 776 (710-838) | 99 (78-120) | 12.81 (10.16-15.16) | 59 (46-74) | 7.69 (5.90-9.40) | 50 (38-62) | 6.39 (5.03-7.92) | 23 (17-29) | 2.93 (2.22-3.71) | 20 (15-26) | 2.61 (1.96-3.31) |
| Sierra Leone | Female | 12081 (11106-12954) | 888 (666-1097) | 7.42 (5.53-8.89) | 641 (479-790) | 5.33 (4.05-6.52) | 381 (286-473) | 3.18 (2.39-3.91) | 148 (104-198) | 1.23 (0.87-1.62) | 87 (55-121) | 0.72 (0.47-1.00) |
| Sierra Leone | Male | 16543 (14795-18108) | 1310 (1001-1641) | 7.97 (6.18-9.85) | 942 (704-1185) | 5.70 (4.40-7.03) | 553 (416-721) | 3.36 (2.59-4.27) | 137 (92-192) | 0.83 (0.57-1.15) | 78 (49-118) | 0.47 (0.30-0.70) |
| Singapore | Female | 9905 (9360-10450) | 1272 (911-1570) | 12.86 (9.31-15.74) | 182 (99-297) | 1.84 (0.99-2.98) | 1141 (826-1432) | 11.55 (8.46-14.39) | 85 (45-145) | 0.86 (0.43-1.48) | 82 (42-139) | 0.82 (0.42-1.39) |
| Singapore | Male | 10540 (9908-11139) | 1564 (1152-1894) | 14.88 (11.01-17.88) | 223 (122-365) | 2.14 (1.16-3.50) | 1416 (1034-1725) | 13.41 (9.73-16.35) | 95 (51-162) | 0.91 (0.48-1.55) | 91 (47-159) | 0.86 (0.45-1.49) |
| Slovakia | Female | 11039 (10269-11848) | 1887 (1547-2231) | 17.14 (14.38-19.72) | 1005 (808-1236) | 9.10 (7.41-11.00) | 1061 (845-1302) | 9.59 (7.74-11.61) | 542 (418-684) | 4.92 (3.86-6.14) | 506 (388-642) | 4.59 (3.58-5.71) |
| Slovakia | Male | 11084 (10319-11837) | 2305 (1924-2639) | 20.81 (17.83-23.50) | 1289 (1042-1547) | 11.60 (9.47-13.89) | 1347 (1069-1621) | 12.14 (9.83-14.41) | 612 (472-766) | 5.53 (4.29-6.85) | 562 (429-716) | 5.08 (3.97-6.41) |
| Slovenia | Female | 4182 (3905-4476) | 785 (646-930) | 18.86 (15.71-21.89) | 377 (294-469) | 9.02 (7.12-11.08) | 483 (386-594) | 11.56 (9.32-14.26) | 205 (152-263) | 4.90 (3.65-6.23) | 184 (135-241) | 4.40 (3.28-5.67) |
| Slovenia | Male | 3212 (3015-3394) | 702 (596-796) | 21.92 (18.74-24.65) | 355 (286-427) | 11.01 (8.90-13.23) | 448 (369-535) | 13.97 (11.66-16.59) | 155 (116-196) | 4.83 (3.68-6.22) | 132 (96-176) | 4.12 (3.01-5.43) |
| Solomon Islands | Female | 1670 (1514-1821) | 104 (71-138) | 6.23 (4.24-8.15) | 73 (50-98) | 4.38 (2.95-5.81) | 46 (30-62) | 2.76 (1.82-3.73) | 16 (11-23) | 0.98 (0.67-1.38) | 7 (4-10) | 0.40 (0.25-0.61) |
| Solomon Islands | Male | 1878 (1667-2069) | 127 (86-172) | 6.79 (4.63-9.02) | 90 (60-122) | 4.78 (3.24-6.50) | 57 (38-79) | 3.04 (2.01-4.20) | 9 (5-13) | 0.46 (0.29-0.69) | 4 (2-7) | 0.23 (0.13-0.36) |
| Somalia | Female | 35581 (32205-38912) | 3701 (2750-4605) | 10.41 (7.89-12.78) | 2574 (1892-3253) | 7.28 (5.38-9.06) | 1511 (1118-1965) | 4.27 (3.13-5.47) | 637 (441-887) | 1.79 (1.25-2.42) | 446 (291-642) | 1.26 (0.83-1.80) |
| Somalia | Male | 44378 (38290-50171) | 3133 (2164-4157) | 7.04 (5.02-9.16) | 2267 (1546-3017) | 5.11 (3.66-6.68) | 1370 (907-1865) | 3.07 (2.10-4.15) | 380 (244-556) | 0.86 (0.56-1.23) | 272 (168-418) | 0.62 (0.39-0.93) |
| South Africa | Female | 194050 (183594-204291) | 9596 (6577-12699) | 4.96 (3.41-6.53) | 7406 (5179-9574) | 3.81 (2.68-4.97) | 5506 (3974-7093) | 2.84 (2.03-3.66) | 2653 (1750-3599) | 1.37 (0.91-1.84) | 2427 (1579-3301) | 1.25 (0.82-1.68) |
| South Africa | Male | 198281 (186115-210933) | 9213 (5995-12628) | 4.65 (3.02-6.39) | 6887 (4552-9444) | 3.49 (2.31-4.75) | 5179 (3449-7065) | 2.61 (1.74-3.54) | 1486 (825-2253) | 0.75 (0.42-1.14) | 1330 (736-2058) | 0.67 (0.37-1.04) |
| South Korea | Female | 92187 (85586-98965) | 15230 (11628-18225) | 16.41 (12.84-19.40) | 6729 (4964-8540) | 7.29 (5.40-9.09) | 10066 (7517-12459) | 10.90 (8.27-13.44) | 3361 (2447-4315) | 3.66 (2.65-4.64) | 3346 (2443-4351) | 3.64 (2.65-4.64) |
| South Korea | Male | 106147 (98514-113993) | 17245 (12882-20978) | 16.30 (12.25-19.48) | 7773 (5665-10000) | 7.35 (5.41-9.34) | 11463 (8425-14305) | 10.82 (8.05-13.27) | 3428 (2424-4531) | 3.23 (2.35-4.20) | 3427 (2431-4533) | 3.23 (2.33-4.17) |
| Spain | Female | 130213 (120429-139652) | 2720 (1219-5422) | 2.10 (0.93-4.23) | 1970 (705-3877) | 1.51 (0.55-2.99) | 2284 (885-4640) | 1.75 (0.69-3.56) | 820 (176-1853) | 0.63 (0.13-1.43) | 724 (148-1660) | 0.55 (0.11-1.26) |
| Spain | Male | 109851 (103145-117008) | 4322 (2913-6463) | 3.93 (2.66-5.80) | 2489 (1508-3857) | 2.26 (1.38-3.48) | 3095 (1970-4771) | 2.82 (1.79-4.32) | 862 (429-1536) | 0.78 (0.39-1.39) | 788 (365-1436) | 0.71 (0.34-1.30) |
| Sri Lanka | Female | 69379 (63688-75586) | 10077 (8153-12081) | 14.60 (11.97-17.02) | 6615 (5267-8119) | 9.58 (7.77-11.49) | 4332 (3400-5347) | 6.25 (4.95-7.49) | 2106 (1610-2704) | 3.06 (2.36-3.85) | 1815 (1341-2360) | 2.62 (1.94-3.33) |
| Sri Lanka | Male | 85024 (77046-92928) | 13624 (11017-16469) | 16.13 (13.53-18.71) | 9040 (7216-11230) | 10.66 (8.81-12.86) | 5890 (4590-7569) | 6.96 (5.59-8.52) | 1898 (1409-2577) | 2.24 (1.68-2.93) | 1527 (1131-2098) | 1.80 (1.36-2.41) |
| Sudan | Female | 71123 (65469-76704) | 581 (198-1177) | 0.83 (0.28-1.63) | 513 (184-1025) | 0.72 (0.25-1.44) | 371 (140-744) | 0.53 (0.19-1.05) | 55 (11-193) | 0.08 (0.02-0.27) | 36 (4-151) | 0.05 (0.01-0.21) |
| Sudan | Male | 76183 (69311-83210) | 4015 (2675-5381) | 5.27 (3.59-6.87) | 3081 (2164-4122) | 4.07 (2.85-5.31) | 1927 (1345-2574) | 2.54 (1.81-3.30) | 378 (215-595) | 0.50 (0.29-0.78) | 280 (145-462) | 0.37 (0.19-0.59) |
| Suriname | Female | 3120 (2859-3368) | 179 (123-231) | 5.73 (3.99-7.44) | 127 (88-167) | 4.07 (2.84-5.31) | 86 (60-115) | 2.76 (1.93-3.61) | 56 (38-75) | 1.79 (1.21-2.40) | 47 (31-65) | 1.51 (1.02-2.05) |
| Suriname | Male | 3773 (3425-4118) | 344 (253-432) | 9.12 (6.82-11.19) | 230 (167-300) | 6.15 (4.53-7.76) | 155 (112-204) | 4.11 (3.01-5.29) | 75 (55-102) | 2.02 (1.48-2.65) | 62 (44-84) | 1.66 (1.17-2.20) |
| Swaziland | Female | 3485 (3173-3820) | 217 (151-287) | 6.22 (4.41-8.10) | 159 (113-209) | 4.59 (3.28-5.88) | 107 (75-141) | 3.08 (2.20-3.98) | 50 (32-69) | 1.43 (0.94-1.93) | 36 (23-52) | 1.05 (0.65-1.45) |
| Swaziland | Male | 5138 (4626-5692) | 325 (217-442) | 6.37 (4.30-8.33) | 233 (153-319) | 4.58 (3.07-6.07) | 155 (103-214) | 3.03 (2.05-4.09) | 32 (19-49) | 0.62 (0.37-0.94) | 23 (13-37) | 0.45 (0.26-0.71) |
| Sweden | Female | 21317 (19894-22772) | 1626 (1071-2125) | 7.64 (5.06-9.87) | 423 (251-639) | 1.99 (1.21-2.98) | 1402 (960-1824) | 6.60 (4.62-8.44) | 176 (100-278) | 0.83 (0.48-1.31) | 145 (80-235) | 0.68 (0.38-1.11) |
| Sweden | Male | 22345 (20870-23729) | 2729 (2198-3253) | 12.25 (9.90-14.44) | 653 (432-928) | 2.91 (1.97-4.12) | 2268 (1806-2803) | 10.19 (8.17-12.27) | 269 (169-405) | 1.21 (0.77-1.80) | 238 (148-359) | 1.07 (0.67-1.61) |
| Switzerland | Female | 25052 (23510-26573) | 1797 (1211-2415) | 7.21 (4.95-9.50) | 174 (104-353) | 0.70 (0.43-1.42) | 1729 (1159-2303) | 6.90 (4.81-9.11) | 67 (39-150) | 0.27 (0.16-0.59) | 55 (31-120) | 0.22 (0.13-0.49) |
| Switzerland | Male | 19413 (18374-20376) | 2251 (1774-2707) | 11.59 (9.26-13.82) | 198 (129-356) | 1.02 (0.66-1.82) | 2123 (1658-2559) | 10.91 (8.68-13.15) | 89 (56-169) | 0.46 (0.29-0.87) | 79 (49-150) | 0.41 (0.25-0.76) |
| Syria | Female | 57238 (51702-62362) | 237 (37-739) | 0.41 (0.06-1.29) | 219 (34-697) | 0.38 (0.06-1.22) | 176 (26-553) | 0.31 (0.04-0.96) | 51 (1-280) | 0.09 (0.00-0.48) | 44 (1-252) | 0.08 (0.00-0.43) |
| Syria | Male | 58999 (52888-64744) | 2838 (1692-4140) | 4.84 (2.88-6.94) | 2217 (1379-3158) | 3.75 (2.33-5.23) | 1380 (875-2004) | 2.35 (1.52-3.31) | 658 (352-1056) | 1.12 (0.61-1.75) | 556 (290-900) | 0.94 (0.50-1.52) |
| Taiwan (Province of China) | Female | 99979 (94455-106103) | 13918 (10912-16609) | 13.93 (11.20-16.45) | 6341 (4816-7802) | 6.35 (4.85-7.88) | 8966 (7052-10997) | 8.94 (7.09-10.99) | 2214 (1631-2918) | 2.22 (1.64-2.88) | 2199 (1632-2853) | 2.19 (1.63-2.85) |
| Taiwan (Province of China) | Male | 103058 (97007-108451) | 13523 (10861-15940) | 13.10 (10.68-15.46) | 6146 (4808-7764) | 5.97 (4.67-7.47) | 8759 (6832-10644) | 8.52 (6.69-10.37) | 2126 (1621-2791) | 2.07 (1.59-2.67) | 2066 (1577-2686) | 2.02 (1.54-2.61) |
| Tajikistan | Female | 6804 (6299-7279) | 493 (359-633) | 7.26 (5.28-9.28) | 356 (257-461) | 5.23 (3.78-6.80) | 216 (153-283) | 3.17 (2.30-4.12) | 101 (74-131) | 1.49 (1.11-1.94) | 84 (60-110) | 1.23 (0.88-1.64) |
| Tajikistan | Male | 6191 (5684-6711) | 783 (606-945) | 12.68 (9.91-14.97) | 539 (414-652) | 8.72 (6.75-10.37) | 322 (247-399) | 5.20 (3.99-6.29) | 123 (94-154) | 1.97 (1.53-2.50) | 74 (55-98) | 1.21 (0.89-1.58) |
| Tanzania | Female | 105573 (96680-114035) | 12777 (10022-15361) | 12.17 (9.76-14.31) | 8732 (6767-10703) | 8.34 (6.49-9.95) | 5268 (4096-6533) | 5.02 (3.91-6.11) | 1860 (1350-2400) | 1.76 (1.28-2.24) | 807 (546-1113) | 0.76 (0.53-1.05) |
| Tanzania | Male | 114806 (104897-125540) | 13432 (10384-16704) | 11.73 (9.11-14.20) | 9204 (6983-11585) | 8.05 (6.14-9.87) | 5592 (4126-7072) | 4.88 (3.62-6.08) | 1213 (824-1643) | 1.06 (0.73-1.42) | 732 (485-1057) | 0.64 (0.42-0.91) |
| Thailand | Female | 437112 (406226-471419) | 56414 (41836-69558) | 12.92 (9.69-15.73) | 36308 (26427-45623) | 8.30 (6.09-10.31) | 25809 (18529-32658) | 5.91 (4.30-7.49) | 12039 (8502-15533) | 2.76 (1.97-3.53) | 10866 (7650-14207) | 2.50 (1.79-3.24) |
| Thailand | Male | 410917 (377784-441644) | 61800 (47935-75219) | 15.15 (11.88-17.84) | 39999 (30897-50037) | 9.83 (7.62-11.92) | 28503 (21411-35935) | 6.99 (5.25-8.57) | 10169 (7563-13619) | 2.48 (1.85-3.27) | 8829 (6468-11889) | 2.16 (1.61-2.87) |
| The Bahamas | Female | 1898 (1765-2024) | 113 (80-147) | 5.96 (4.22-7.73) | 62 (45-81) | 3.25 (2.36-4.25) | 76 (55-98) | 4.02 (2.89-5.14) | 25 (17-33) | 1.29 (0.91-1.74) | 22 (15-29) | 1.14 (0.78-1.53) |
| The Bahamas | Male | 2194 (2032-2359) | 212 (166-258) | 9.62 (7.58-11.50) | 106 (80-136) | 4.85 (3.73-6.08) | 133 (100-167) | 6.01 (4.58-7.50) | 35 (26-45) | 1.59 (1.17-2.05) | 28 (20-37) | 1.26 (0.93-1.67) |
| The Gambia | Female | 4010 (3715-4332) | 296 (230-365) | 7.38 (5.77-9.01) | 213 (162-265) | 5.32 (4.18-6.50) | 127 (98-161) | 3.18 (2.47-4.03) | 60 (43-78) | 1.48 (1.08-1.92) | 38 (25-53) | 0.95 (0.65-1.32) |
| The Gambia | Male | 6600 (6009-7208) | 514 (386-636) | 7.76 (5.89-9.55) | 368 (276-460) | 5.56 (4.22-6.90) | 219 (162-279) | 3.31 (2.51-4.22) | 63 (43-88) | 0.96 (0.66-1.32) | 43 (28-62) | 0.65 (0.43-0.93) |
| Timor-Leste | Female | 3325 (3016-3653) | 438 (342-547) | 13.20 (10.48-15.85) | 291 (225-370) | 8.81 (6.93-10.85) | 185 (142-238) | 5.57 (4.36-6.91) | 53 (39-74) | 1.59 (1.17-2.21) | 36 (25-51) | 1.08 (0.76-1.52) |
| Timor-Leste | Male | 3568 (3161-3987) | 568 (453-694) | 15.97 (13.33-18.58) | 381 (300-478) | 10.67 (8.77-12.85) | 240 (187-309) | 6.75 (5.39-8.33) | 66 (48-90) | 1.85 (1.38-2.41) | 43 (29-59) | 1.19 (0.85-1.63) |
| Togo | Female | 14918 (13658-16153) | 1027 (748-1300) | 6.90 (5.02-8.50) | 735 (540-940) | 4.94 (3.65-6.19) | 442 (321-575) | 2.97 (2.15-3.82) | 178 (125-251) | 1.20 (0.84-1.65) | 97 (63-140) | 0.65 (0.43-0.94) |
| Togo | Male | 19077 (17475-20769) | 1437 (1065-1817) | 7.54 (5.66-9.34) | 1019 (766-1325) | 5.38 (4.06-6.78) | 616 (453-813) | 3.24 (2.38-4.14) | 139 (93-203) | 0.74 (0.49-1.06) | 67 (41-101) | 0.35 (0.22-0.53) |
| Tonga | Female | 416 (378-453) | 46 (37-55) | 11.01 (9.05-12.98) | 31 (24-38) | 7.42 (5.96-8.97) | 20 (16-25) | 4.89 (3.88-6.04) | 10 (7-13) | 2.36 (1.80-3.04) | 8 (6-10) | 1.95 (1.45-2.50) |
| Tonga | Male | 80 (74-85) | 10 (8-12) | 12.19 (9.94-14.43) | 6 (5-8) | 8.12 (6.55-9.84) | 4 (3-5) | 5.38 (4.30-6.64) | 1 (1-2) | 1.63 (1.21-2.09) | 1 (1-1) | 1.32 (0.98-1.78) |
| Trinidad and Tobago | Female | 8042 (7501-8597) | 534 (380-683) | 6.64 (4.71-8.49) | 335 (244-437) | 4.17 (3.07-5.37) | 304 (219-391) | 3.78 (2.75-4.86) | 137 (96-186) | 1.70 (1.19-2.29) | 111 (77-156) | 1.39 (0.94-1.93) |
| Trinidad and Tobago | Male | 11572 (10513-12617) | 1248 (994-1511) | 10.84 (8.71-12.75) | 739 (579-928) | 6.43 (5.09-7.86) | 663 (508-840) | 5.75 (4.53-7.07) | 249 (188-329) | 2.17 (1.64-2.81) | 208 (151-277) | 1.81 (1.32-2.36) |
| Tunisia | Female | 37686 (34073-40848) | 150 (31-450) | 0.40 (0.08-1.17) | 134 (27-414) | 0.36 (0.07-1.11) | 110 (21-331) | 0.30 (0.05-0.89) | 23 (0-145) | 0.06 (0.00-0.39) | 20 (0-120) | 0.05 (0.00-0.33) |
| Tunisia | Male | 45021 (40579-49129) | 1973 (1166-2969) | 4.39 (2.64-6.37) | 1525 (910-2218) | 3.40 (2.06-4.82) | 1052 (637-1502) | 2.33 (1.44-3.28) | 351 (167-554) | 0.78 (0.38-1.22) | 303 (142-506) | 0.68 (0.32-1.08) |
| Turkey | Female | 261519 (242601-282545) | 3 (0-341) | 0.00 (0.00-0.13) | 3 (0-341) | 0.00 (0.00-0.13) | 3 (0-316) | 0.00 (0.00-0.13) | 0 (0-124) | 0.00 (0.00-0.05) | 0 (0-103) | 0.00 (0.00-0.04) |
| Turkey | Male | 268925 (246244-290621) | 6132 (2892-10955) | 2.29 (1.09-4.01) | 5140 (2407-8714) | 1.91 (0.90-3.22) | 4286 (2005-7316) | 1.60 (0.73-2.67) | 1319 (408-2581) | 0.49 (0.16-0.94) | 1274 (376-2535) | 0.48 (0.14-0.93) |
| Turkmenistan | Female | 12233 (11477-12990) | 935 (706-1154) | 7.64 (5.77-9.45) | 625 (471-795) | 5.12 (3.89-6.47) | 457 (346-582) | 3.74 (2.84-4.71) | 210 (155-272) | 1.72 (1.29-2.23) | 179 (131-234) | 1.46 (1.09-1.90) |
| Turkmenistan | Male | 16249 (15073-17285) | 2009 (1585-2430) | 12.42 (9.83-14.64) | 1289 (1000-1575) | 7.94 (6.18-9.48) | 926 (718-1152) | 5.72 (4.42-7.02) | 274 (211-342) | 1.69 (1.31-2.11) | 199 (152-253) | 1.23 (0.94-1.54) |
| Tuvalu | Female | 62 (56-70) | 6 (5-8) | 10.44 (8.12-12.76) | 4 (3-6) | 6.97 (5.39-8.71) | 3 (2-4) | 4.75 (3.60-5.96) | 1 (1-2) | 1.74 (1.20-2.37) | 1 (0-1) | 1.03 (0.68-1.49) |
| Tuvalu | Male | 55 (49-62) | 6 (5-8) | 11.30 (8.87-13.54) | 4 (3-5) | 7.53 (5.78-9.36) | 3 (2-4) | 5.14 (3.99-6.51) | 1 (1-1) | 1.50 (1.07-2.01) | 1 (0-1) | 1.04 (0.68-1.49) |
| Uganda | Female | 57334 (52334-61918) | 6375 (5075-7629) | 11.15 (9.02-13.15) | 4413 (3483-5377) | 7.72 (6.17-9.31) | 2646 (2018-3246) | 4.62 (3.60-5.63) | 802 (587-1056) | 1.40 (1.03-1.83) | 482 (322-668) | 0.84 (0.57-1.16) |
| Uganda | Male | 76066 (68906-82847) | 6014 (4567-7687) | 7.89 (6.07-9.86) | 4316 (3179-5479) | 5.66 (4.29-7.21) | 2616 (1938-3390) | 3.45 (2.60-4.47) | 525 (356-728) | 0.69 (0.48-0.96) | 301 (189-450) | 0.40 (0.25-0.60) |
| Ukraine | Female | 63138 (58918-67315) | 2139 (1249-3189) | 3.40 (1.96-4.94) | 1851 (1095-2682) | 2.93 (1.75-4.21) | 1377 (869-1931) | 2.20 (1.38-3.05) | 820 (440-1275) | 1.29 (0.68-1.97) | 686 (370-1073) | 1.09 (0.59-1.70) |
| Ukraine | Male | 45530 (42485-48641) | 4405 (3563-5240) | 9.70 (7.93-11.42) | 2974 (2401-3627) | 6.53 (5.30-7.86) | 1986 (1568-2467) | 4.35 (3.50-5.39) | 833 (640-1069) | 1.84 (1.41-2.33) | 572 (429-757) | 1.26 (0.94-1.65) |
| United Arab Emirates | Female | 9674 (8669-10697) | 56 (8-160) | 0.58 (0.09-1.61) | 35 (5-109) | 0.37 (0.05-1.15) | 50 (7-148) | 0.52 (0.07-1.50) | 5 (0-32) | 0.06 (0.00-0.33) | 5 (0-28) | 0.05 (0.00-0.29) |
| United Arab Emirates | Male | 18810 (17062-20554) | 1006 (592-1403) | 5.38 (3.17-7.34) | 423 (258-604) | 2.25 (1.39-3.19) | 791 (459-1121) | 4.21 (2.51-5.83) | 69 (41-102) | 0.37 (0.22-0.53) | 61 (34-90) | 0.32 (0.19-0.48) |
| United Kingdom | Female | 126343 (120806-132356) | 7752 (5062-10436) | 6.11 (4.04-8.22) | 3111 (2122-4275) | 2.47 (1.68-3.37) | 6228 (4285-8280) | 4.96 (3.38-6.63) | 1161 (740-1658) | 0.92 (0.59-1.30) | 1056 (681-1484) | 0.84 (0.54-1.17) |
| United Kingdom | Male | 125267 (119500-131054) | 12212 (9436-14752) | 9.74 (7.62-11.74) | 4364 (3171-5734) | 3.49 (2.54-4.57) | 9207 (7047-11320) | 7.34 (5.67-9.05) | 1641 (1139-2199) | 1.31 (0.91-1.75) | 1489 (1026-2023) | 1.19 (0.82-1.62) |
| United States | Female | 1409789 (1357390-1467524) | 112621 (85463-140823) | 7.98 (6.13-9.89) | 21455 (12992-32679) | 1.52 (0.92-2.30) | 100507 (76406-126537) | 7.13 (5.40-8.91) | 9637 (5596-15356) | 0.68 (0.40-1.08) | 8949 (5149-14152) | 0.64 (0.37-1.00) |
| United States | Male | 1512258 (1459634-1562738) | 171912 (134718-205679) | 11.36 (8.91-13.47) | 32058 (19444-47347) | 2.12 (1.30-3.14) | 150825 (116285-181646) | 9.98 (7.72-11.88) | 13474 (7819-20302) | 0.89 (0.52-1.35) | 12551 (7207-18883) | 0.83 (0.48-1.26) |
| Uruguay | Female | 10054 (9391-10717) | 1242 (1025-1442) | 12.33 (10.27-14.24) | 711 (574-864) | 7.05 (5.74-8.43) | 673 (537-818) | 6.73 (5.36-7.96) | 381 (298-473) | 3.80 (3.01-4.61) | 362 (288-457) | 3.61 (2.87-4.48) |
| Uruguay | Male | 10978 (10284-11702) | 1572 (1294-1879) | 14.31 (11.86-16.78) | 889 (716-1086) | 8.10 (6.62-9.67) | 846 (677-1026) | 7.70 (6.24-9.27) | 420 (326-526) | 3.84 (2.99-4.73) | 374 (288-473) | 3.40 (2.63-4.21) |
| Uzbekistan | Female | 73587 (69221-77498) | 5608 (4219-6951) | 7.62 (5.71-9.41) | 4023 (2997-4988) | 5.49 (4.06-6.74) | 2488 (1872-3165) | 3.38 (2.56-4.27) | 1280 (954-1619) | 1.74 (1.29-2.20) | 1089 (797-1409) | 1.48 (1.08-1.91) |
| Uzbekistan | Male | 76347 (71615-80779) | 9599 (7757-11345) | 12.58 (10.26-14.66) | 6536 (5181-7840) | 8.57 (6.83-10.19) | 4014 (3155-4957) | 5.27 (4.15-6.46) | 1440 (1089-1800) | 1.89 (1.44-2.34) | 1112 (844-1416) | 1.46 (1.10-1.82) |
| Vanuatu | Female | 944 (840-1053) | 86 (65-109) | 9.13 (7.11-11.05) | 59 (44-76) | 6.28 (4.83-7.85) | 38 (28-50) | 4.05 (3.04-5.13) | 13 (9-19) | 1.38 (0.99-1.95) | 6 (4-9) | 0.65 (0.41-0.99) |
| Vanuatu | Male | 986 (847-1120) | 100 (75-130) | 10.13 (7.88-12.44) | 68 (50-91) | 6.93 (5.40-8.66) | 44 (32-60) | 4.47 (3.42-5.75) | 9 (6-14) | 0.93 (0.63-1.34) | 5 (3-8) | 0.48 (0.30-0.75) |
| Venezuela | Female | 142054 (132259-152577) | 14746 (11503-17897) | 10.39 (8.19-12.42) | 10174 (7811-12768) | 7.19 (5.49-8.93) | 6285 (4693-7883) | 4.43 (3.40-5.52) | 4879 (3680-6241) | 3.45 (2.63-4.31) | 4580 (3429-5876) | 3.23 (2.42-4.10) |
| Venezuela | Male | 167864 (154330-180786) | 24573 (19395-29885) | 14.65 (11.76-17.43) | 16452 (12811-20278) | 9.83 (7.74-11.86) | 10165 (7829-12852) | 6.07 (4.65-7.52) | 6175 (4625-7779) | 3.68 (2.85-4.61) | 5623 (4275-7121) | 3.35 (2.57-4.21) |
| Vietnam | Female | 339791 (313050-370183) | 45316 (35814-54849) | 13.28 (10.66-15.70) | 29948 (23265-36962) | 8.81 (6.93-10.60) | 19449 (15035-24125) | 5.70 (4.44-6.95) | 7623 (5779-9794) | 2.24 (1.75-2.83) | 5223 (3858-6824) | 1.54 (1.14-1.97) |
| Vietnam | Male | 299704 (270670-327166) | 47178 (37771-57044) | 15.88 (13.19-18.42) | 31392 (24745-39179) | 10.50 (8.63-12.64) | 20279 (15799-25915) | 6.80 (5.38-8.34) | 7123 (5433-9407) | 2.39 (1.84-3.06) | 4953 (3718-6771) | 1.66 (1.26-2.19) |
| Yemen | Female | 33701 (30655-36612) | 181 (33-437) | 0.54 (0.09-1.29) | 159 (29-401) | 0.47 (0.09-1.19) | 120 (23-294) | 0.36 (0.07-0.88) | 16 (1-77) | 0.05 (0.00-0.22) | 12 (1-62) | 0.04 (0.00-0.18) |
| Yemen | Male | 37380 (33023-41320) | 1771 (1091-2465) | 4.72 (2.98-6.41) | 1361 (891-1921) | 3.67 (2.39-5.01) | 847 (562-1185) | 2.28 (1.53-3.08) | 224 (119-363) | 0.60 (0.33-0.96) | 192 (100-317) | 0.52 (0.28-0.81) |
| Zambia | Female | 36791 (33459-40089) | 3781 (2973-4596) | 10.30 (8.13-12.29) | 2633 (2017-3233) | 7.15 (5.50-8.74) | 1587 (1185-2010) | 4.31 (3.30-5.38) | 682 (505-892) | 1.85 (1.41-2.39) | 409 (294-557) | 1.11 (0.80-1.50) |
| Zambia | Male | 57859 (50623-64552) | 4206 (3130-5488) | 7.30 (5.47-9.33) | 3007 (2174-3964) | 5.23 (3.79-6.69) | 1850 (1348-2482) | 3.20 (2.36-4.16) | 488 (344-682) | 0.85 (0.59-1.17) | 289 (187-417) | 0.50 (0.33-0.73) |
| Zimbabwe | Female | 39251 (35961-42737) | 3223 (2444-4048) | 8.20 (6.24-10.07) | 2267 (1688-2876) | 5.78 (4.33-7.18) | 1399 (1016-1801) | 3.56 (2.63-4.52) | 629 (439-863) | 1.61 (1.14-2.15) | 499 (326-716) | 1.28 (0.84-1.80) |
| Zimbabwe | Male | 38793 (35299-42529) | 3388 (2534-4294) | 8.76 (6.65-10.75) | 2381 (1740-3048) | 6.14 (4.49-7.70) | 1440 (1045-1929) | 3.73 (2.75-4.87) | 403 (275-592) | 1.04 (0.71-1.51) | 303 (200-458) | 0.79 (0.53-1.17) |

### **Supplementary table 5. Impact of prevented CVD deaths, hyperkalaemia-related CVD deaths, and its net effect in patients with advanced CKD for each country**

#### a. **Prevented CVD deaths in patients with CKD**

| **Country** | **Replacement of discretionary salt and non-discretionary salt Median (95% UI)** | **Replacement of discretionary salt Median (95% UI)** | **Replacement of non-discretionary salt  Median (95% UI)** |
| --- | --- | --- | --- |
| Afghanistan | 231 (128-363) | 188 (109-297) | 125 (71-196) |
| Albania | 1010 (870-1154) | 668 (556-787) | 469 (386-571) |
| Algeria | 421 (219-712) | 349 (171-597) | 253 (132-441) |
| American Samoa | 4 (3-5) | 2 (2-3) | 2 (2-3) |
| Andorra | 4 (3-6) | 2 (1-3) | 3 (2-5) |
| Angola | 912 (711-1140) | 669 (518-832) | 422 (326-525) |
| Antigua and Barbuda | 7 (5-8) | 4 (3-5) | 4 (3-4) |
| Argentina | 2860 (2440-3338) | 1801 (1499-2163) | 1414 (1150-1717) |
| Armenia | 962 (802-1143) | 639 (524-762) | 430 (347-532) |
| Australia | 716 (390-1134) | 264 (139-439) | 676 (362-1055) |
| Austria | 1927 (1594-2294) | 579 (419-781) | 1506 (1216-1851) |
| Azerbaijan | 2214 (1864-2615) | 1474 (1230-1764) | 1007 (833-1220) |
| Bahrain | 8 (5-12) | 5 (3-8) | 6 (4-9) |
| Bangladesh | 13488 (11501-15497) | 9320 (7813-10973) | 5827 (4778-6982) |
| Barbados | 19 (14-25) | 12 (9-16) | 12 (9-15) |
| Belarus | 1628 (1156-2239) | 1229 (858-1685) | 923 (657-1252) |
| Belgium | 1305 (1080-1522) | 408 (299-537) | 1010 (820-1207) |
| Belize | 13 (11-16) | 9 (7-11) | 6 (5-8) |
| Benin | 339 (285-392) | 238 (199-280) | 144 (117-173) |
| Bermuda | 7 (5-8) | 0 (0-1) | 6 (5-8) |
| Bhutan | 59 (51-69) | 41 (34-48) | 26 (22-32) |
| Bolivia | 436 (356-514) | 296 (238-359) | 191 (151-232) |
| Bosnia and Herzegovina | 1265 (1081-1466) | 826 (696-989) | 595 (494-726) |
| Botswana | 61 (43-82) | 47 (33-62) | 35 (25-46) |
| Brazil | 16730 (14625-19208) | 10912 (9282-12646) | 7878 (6682-9291) |
| Brunei | 25 (21-29) | 12 (10-14) | 16 (13-19) |
| Bulgaria | 5993 (5149-6868) | 3788 (3123-4498) | 3119 (2568-3765) |
| Burkina Faso | 547 (436-666) | 392 (309-477) | 236 (185-295) |
| Burundi | 198 (169-231) | 140 (118-165) | 83 (69-99) |
| Cambodia | 1357 (1131-1614) | 931 (771-1125) | 578 (467-707) |
| Cameroon | 1087 (875-1295) | 766 (622-927) | 470 (383-569) |
| Canada | 2683 (2124-3249) | 856 (622-1135) | 2145 (1666-2588) |
| Cape Verde | 33 (27-40) | 23 (18-28) | 15 (12-18) |
| Central African Republic | 189 (143-238) | 142 (109-177) | 87 (66-111) |
| Chad | 375 (303-449) | 267 (217-326) | 160 (127-197) |
| Chile | 1165 (965-1348) | 686 (559-820) | 621 (503-744) |
| China | 249071 (214636-287064) | 153483 (129884-181127) | 126581 (105090-150993) |
| Colombia | 5460 (4746-6257) | 3592 (3092-4198) | 2526 (2110-2946) |
| Comoros | 21 (18-25) | 15 (12-18) | 9 (7-11) |
| Congo | 246 (191-306) | 180 (140-224) | 117 (91-142) |
| Costa Rica | 461 (398-528) | 282 (240-328) | 232 (196-272) |
| Cote d'Ivoire | 747 (613-877) | 516 (426-613) | 325 (266-390) |
| Croatia | 1793 (1497-2075) | 1039 (845-1249) | 987 (792-1200) |
| Cuba | 1250 (1035-1459) | 803 (651-953) | 608 (494-728) |
| Cyprus | 116 (88-146) | 56 (42-74) | 78 (60-101) |
| Czech Republic | 3426 (2933-3917) | 1768 (1451-2123) | 2101 (1742-2493) |
| Democratic Republic of the Congo | 891 (466-1439) | 811 (443-1302) | 603 (344-924) |
| Denmark | 493 (376-599) | 96 (60-143) | 436 (334-534) |
| Djibouti | 22 (19-26) | 15 (13-18) | 10 (8-12) |
| Dominica | 7 (6-8) | 5 (4-6) | 3 (3-4) |
| Dominican Republic | 699 (584-822) | 456 (379-542) | 336 (273-410) |
| Ecuador | 759 (633-875) | 501 (409-595) | 347 (282-418) |
| Egypt | 1639 (901-2486) | 1319 (770-1957) | 947 (563-1412) |
| El Salvador | 439 (374-506) | 292 (243-340) | 195 (161-231) |
| Equatorial Guinea | 41 (32-50) | 28 (22-35) | 20 (16-25) |
| Eritrea | 118 (97-143) | 84 (68-103) | 50 (40-63) |
| Estonia | 93 (56-142) | 54 (33-93) | 64 (40-106) |
| Ethiopia | 1268 (1037-1508) | 896 (717-1074) | 543 (429-675) |
| Federated States of Micronesia | 11 (9-13) | 7 (6-9) | 5 (4-6) |
| Fiji | 107 (90-123) | 70 (59-82) | 47 (38-56) |
| Finland | 813 (601-1053) | 277 (194-386) | 680 (494-878) |
| France | 5209 (3760-6675) | 2067 (1490-2787) | 3917 (2881-5069) |
| Gabon | 97 (74-119) | 66 (51-82) | 49 (38-62) |
| Georgia | 1760 (1493-2046) | 1184 (982-1411) | 798 (646-968) |
| Germany | 13658 (10128-17345) | 4717 (3321-6433) | 11014 (8252-13919) |
| Ghana | 846 (731-969) | 586 (497-684) | 366 (305-436) |
| Greece | 2033 (1476-2550) | 1181 (887-1493) | 1184 (895-1512) |
| Greenland | 3 (2-3) | 1 (1-1) | 2 (2-3) |
| Grenada | 7 (6-9) | 5 (4-6) | 4 (3-4) |
| Guatemala | 851 (720-994) | 567 (470-674) | 381 (316-458) |
| Guinea | 410 (334-485) | 293 (238-351) | 177 (143-214) |
| Guinea-Bissau | 61 (50-73) | 44 (35-52) | 26 (21-32) |
| Guyana | 57 (47-66) | 37 (30-44) | 28 (23-34) |
| Haiti | 615 (512-721) | 434 (357-511) | 267 (220-323) |
| Honduras | 1075 (905-1244) | 728 (607-866) | 464 (378-559) |
| Hungary | 4446 (3847-5053) | 2579 (2163-3078) | 2492 (2092-2964) |
| Iceland | 24 (18-30) | 5 (3-7) | 21 (16-26) |
| India | 77615 (67600-86910) | 53258 (45421-60779) | 33023 (27392-38895) |
| Indonesia | 32716 (28413-37484) | 21726 (18632-25473) | 14452 (12093-17088) |
| Iran | 1031 (524-1602) | 851 (447-1320) | 612 (334-968) |
| Iraq | 564 (315-835) | 448 (259-641) | 325 (202-467) |
| Ireland | 215 (154-288) | 21 (13-35) | 207 (147-277) |
| Israel | 416 (327-516) | 135 (101-182) | 333 (261-414) |
| Italy | 10247 (7744-12997) | 4550 (3354-6110) | 7075 (5346-9138) |
| Jamaica | 219 (182-256) | 148 (121-178) | 101 (83-122) |
| Japan | 27220 (22847-32046) | 10921 (8660-13646) | 18813 (15499-22721) |
| Jordan | 63 (35-97) | 50 (29-77) | 37 (21-55) |
| Kazakhstan | 3821 (3207-4495) | 2410 (1982-2846) | 1867 (1536-2271) |
| Kenya | 501 (391-633) | 349 (272-445) | 220 (167-285) |
| Kiribati | 8 (6-9) | 5 (4-6) | 3 (3-4) |
| Kuwait | 72 (56-90) | 36 (28-47) | 49 (39-62) |
| Kyrgyzstan | 908 (767-1060) | 633 (526-749) | 384 (313-469) |
| Laos | 691 (597-793) | 472 (394-547) | 298 (248-349) |
| Latvia | 420 (299-571) | 274 (187-366) | 278 (187-378) |
| Lebanon | 46 (23-81) | 38 (19-67) | 28 (14-49) |
| Lesotho | 78 (54-106) | 63 (44-84) | 41 (29-53) |
| Liberia | 140 (113-166) | 99 (81-120) | 59 (48-72) |
| Libya | 70 (38-108) | 56 (32-87) | 42 (25-66) |
| Lithuania | 951 (736-1170) | 499 (384-640) | 557 (422-708) |
| Luxembourg | 47 (35-58) | 3 (2-4) | 46 (35-56) |
| Macedonia | 1130 (952-1317) | 743 (608-885) | 527 (428-652) |
| Madagascar | 601 (506-700) | 426 (355-501) | 255 (208-307) |
| Malawi | 333 (282-390) | 235 (194-278) | 140 (114-171) |
| Malaysia | 3553 (3098-4037) | 2175 (1868-2530) | 1723 (1461-2038) |
| Maldives | 25 (21-29) | 15 (13-18) | 12 (10-14) |
| Mali | 404 (317-489) | 288 (226-353) | 174 (135-212) |
| Malta | 56 (44-68) | 25 (20-32) | 38 (29-47) |
| Marshall Islands | 4 (4-5) | 3 (2-3) | 2 (2-2) |
| Mauritania | 151 (124-181) | 106 (87-128) | 66 (53-81) |
| Mauritius | 205 (179-236) | 131 (112-153) | 99 (82-116) |
| Mexico | 10036 (8399-11548) | 6342 (5226-7465) | 4920 (4035-5901) |
| Moldova | 636 (453-839) | 482 (335-645) | 346 (246-460) |
| Mongolia | 382 (326-444) | 257 (215-302) | 171 (141-204) |
| Montenegro | 297 (252-344) | 190 (157-227) | 145 (117-175) |
| Morocco | 539 (293-827) | 437 (244-665) | 315 (177-472) |
| Mozambique | 616 (514-737) | 436 (357-527) | 261 (211-323) |
| Myanmar | 6917 (5936-7859) | 4743 (4035-5485) | 2898 (2420-3396) |
| Namibia | 75 (51-102) | 59 (42-78) | 42 (29-56) |
| Nauru | 1 (1-1) | 1 (1-1) | 1 (1-1) |
| Nepal | 3044 (2400-3674) | 2145 (1693-2649) | 1310 (1018-1643) |
| Netherlands | 1337 (1020-1656) | 371 (256-524) | 1126 (868-1405) |
| New Zealand | 312 (228-407) | 119 (81-160) | 255 (186-332) |
| Nicaragua | 440 (378-506) | 301 (253-350) | 186 (156-219) |
| Niger | 394 (316-473) | 281 (227-342) | 168 (134-206) |
| Nigeria | 5984 (4757-7272) | 4371 (3475-5320) | 2736 (2190-3322) |
| Norway | 274 (197-370) | 30 (18-53) | 262 (186-353) |
| Oman | 28 (17-42) | 20 (12-29) | 19 (12-29) |
| Pakistan | 11895 (10375-13619) | 8296 (7086-9685) | 5099 (4271-5995) |
| Palau | 3 (2-3) | 2 (1-2) | 1 (1-2) |
| Panama | 346 (301-399) | 203 (172-241) | 181 (153-213) |
| Papua New Guinea | 379 (296-458) | 263 (201-319) | 168 (128-205) |
| Paraguay | 533 (453-606) | 352 (297-409) | 244 (203-288) |
| Peru | 873 (734-1012) | 573 (464-678) | 402 (329-482) |
| Philippines | 10005 (8543-11432) | 6766 (5735-7868) | 4403 (3692-5233) |
| Poland | 10866 (8979-12830) | 6186 (5028-7619) | 5919 (4772-7203) |
| Portugal | 628 (425-887) | 389 (261-549) | 430 (284-595) |
| Puerto Rico | 305 (254-356) | 144 (115-177) | 201 (163-241) |
| Qatar | 6 (4-9) | 2 (1-3) | 6 (3-8) |
| Romania | 12085 (10402-13854) | 7304 (6054-8709) | 6420 (5344-7692) |
| Russia | 29049 (23367-35060) | 18041 (14366-22025) | 14653 (11782-18266) |
| Rwanda | 239 (198-288) | 168 (138-205) | 102 (80-126) |
| Saint Kitts and Nevis | 4 (3-5) | 2 (2-3) | 2 (2-3) |
| Saint Lucia | 15 (13-18) | 10 (8-12) | 8 (6-10) |
| Saint Vincent and the Grenadines | 12 (10-14) | 8 (6-9) | 6 (5-7) |
| Samoa | 13 (11-16) | 9 (7-11) | 6 (5-7) |
| Sao Tome and Principe | 7 (6-8) | 5 (4-6) | 3 (2-4) |
| Saudi Arabia | 354 (213-521) | 229 (146-319) | 252 (158-357) |
| Senegal | 409 (330-485) | 289 (230-347) | 176 (143-216) |
| Serbia | 3599 (3090-4074) | 2329 (1936-2720) | 1772 (1464-2074) |
| Seychelles | 14 (11-16) | 8 (7-10) | 7 (6-9) |
| Sierra Leone | 268 (215-321) | 191 (154-229) | 113 (91-138) |
| Singapore | 356 (294-415) | 52 (33-78) | 321 (265-376) |
| Slovakia | 1884 (1595-2151) | 1034 (842-1234) | 1083 (895-1302) |
| Slovenia | 586 (479-690) | 296 (231-370) | 373 (299-459) |
| Solomon Islands | 48 (40-57) | 33 (27-40) | 21 (17-25) |
| Somalia | 233 (192-282) | 164 (133-203) | 99 (79-123) |
| South Africa | 1960 (1258-2745) | 1579 (1047-2194) | 1208 (837-1626) |
| South Korea | 5564 (4476-6692) | 2522 (1956-3231) | 3735 (2915-4606) |
| Spain | 788 (307-1736) | 637 (230-1280) | 710 (269-1540) |
| Sri Lanka | 2772 (2355-3234) | 1845 (1552-2173) | 1215 (995-1466) |
| Sudan | 416 (236-623) | 341 (203-497) | 227 (136-332) |
| Suriname | 37 (30-43) | 25 (20-29) | 17 (14-20) |
| Swaziland | 34 (23-45) | 26 (19-34) | 18 (13-24) |
| Sweden | 1188 (901-1490) | 301 (194-436) | 1011 (773-1266) |
| Switzerland | 993 (726-1274) | 93 (59-180) | 946 (694-1213) |
| Syria | 287 (163-444) | 237 (137-354) | 156 (92-235) |
| Taiwan (Province of China) | 2283 (1943-2663) | 1052 (852-1269) | 1484 (1234-1763) |
| Tajikistan | 1002 (849-1169) | 705 (595-835) | 426 (351-508) |
| Tanzania | 1091 (904-1278) | 755 (618-905) | 459 (365-555) |
| Thailand | 8204 (6954-9485) | 5303 (4472-6236) | 3785 (3117-4513) |
| The Bahamas | 24 (20-29) | 13 (10-15) | 16 (13-19) |
| The Gambia | 80 (64-97) | 57 (45-70) | 34 (27-42) |
| Timor-Leste | 131 (111-153) | 88 (74-104) | 56 (46-68) |
| Togo | 239 (195-282) | 170 (138-204) | 102 (82-125) |
| Tonga | 8 (7-10) | 6 (5-7) | 4 (3-4) |
| Trinidad and Tobago | 113 (95-133) | 69 (57-82) | 62 (51-74) |
| Tunisia | 173 (84-282) | 142 (72-230) | 103 (51-167) |
| Turkey | 408 (123-858) | 368 (112-757) | 321 (96-634) |
| Turkmenistan | 1071 (915-1230) | 699 (588-818) | 510 (425-598) |
| Tuvalu | 2 (2-2) | 1 (1-1) | 1 (1-1) |
| Uganda | 570 (462-684) | 399 (318-487) | 241 (191-295) |
| Ukraine | 8720 (6054-12085) | 6786 (4568-9268) | 4832 (3310-6634) |
| United Arab Emirates | 36 (21-51) | 17 (10-25) | 30 (18-43) |
| United Kingdom | 5925 (4519-7392) | 2199 (1634-2849) | 4555 (3500-5725) |
| United States | 42549 (33956-51370) | 8304 (5437-11895) | 38079 (30273-45967) |
| Uruguay | 474 (396-550) | 272 (222-328) | 258 (209-311) |
| Uzbekistan | 5816 (4834-6722) | 4018 (3310-4748) | 2506 (2017-2964) |
| Vanuatu | 27 (23-31) | 18 (15-21) | 12 (10-14) |
| Venezuela | 4145 (3569-4752) | 2829 (2397-3302) | 1753 (1470-2099) |
| Vietnam | 8022 (6864-9357) | 5369 (4497-6340) | 3497 (2888-4206) |
| Yemen | 296 (162-447) | 243 (140-364) | 161 (93-239) |
| Zambia | 351 (287-414) | 247 (199-294) | 150 (121-182) |
| Zimbabwe | 635 (509-762) | 452 (364-545) | 280 (220-340) |

#### b. **Additional deaths in patients with CKD**

| **Country** | **Replacement of discretionary salt and non-discretionary salt Median (95% UI)** | **Replacement of discretionary salt Median (95% UI)** | **Replacement of non-discretionary salt  Median (95% UI)** |
| --- | --- | --- | --- |
| Afghanistan | 24 (11-46) | 18 (9-33) | 11 (6-20) |
| Albania | 148 (105-200) | 76 (52-106) | 47 (32-69) |
| Algeria | 43 (17-85) | 33 (13-64) | 23 (9-46) |
| American Samoa | 1 (0-1) | 0 (0-0) | 0 (0-0) |
| Andorra | 1 (0-1) | 0 (0-0) | 0 (0-1) |
| Angola | 98 (67-138) | 61 (42-87) | 35 (24-51) |
| Antigua and Barbuda | 1 (1-1) | 0 (0-0) | 0 (0-0) |
| Argentina | 324 (222-457) | 161 (109-234) | 119 (81-176) |
| Armenia | 126 (90-178) | 69 (47-97) | 42 (28-60) |
| Australia | 93 (41-174) | 29 (13-55) | 84 (39-153) |
| Austria | 257 (171-359) | 55 (32-88) | 176 (115-256) |
| Azerbaijan | 269 (189-387) | 147 (103-219) | 92 (61-136) |
| Bahrain | 1 (0-2) | 0 (0-1) | 1 (0-1) |
| Bangladesh | 1700 (1207-2316) | 957 (666-1359) | 536 (368-776) |
| Barbados | 2 (2-4) | 1 (1-2) | 1 (1-2) |
| Belarus | 200 (118-336) | 134 (78-222) | 96 (55-157) |
| Belgium | 197 (136-278) | 45 (28-71) | 133 (90-196) |
| Belize | 2 (1-2) | 1 (1-1) | 1 (0-1) |
| Benin | 42 (31-56) | 25 (18-34) | 14 (9-18) |
| Bermuda | 1 (1-1) | 0 (0-0) | 1 (1-1) |
| Bhutan | 7 (5-10) | 4 (3-6) | 2 (2-3) |
| Bolivia | 55 (40-76) | 31 (22-43) | 18 (12-25) |
| Bosnia and Herzegovina | 188 (135-259) | 96 (66-138) | 62 (42-91) |
| Botswana | 6 (4-10) | 5 (3-7) | 3 (2-5) |
| Brazil | 2127 (1576-2767) | 1115 (795-1500) | 754 (521-1012) |
| Brunei | 3 (2-4) | 1 (1-1) | 1 (1-2) |
| Bulgaria | 770 (548-1045) | 360 (249-517) | 277 (187-395) |
| Burkina Faso | 65 (46-90) | 40 (28-56) | 22 (15-31) |
| Burundi | 23 (18-31) | 14 (10-18) | 7 (5-10) |
| Cambodia | 195 (143-251) | 110 (80-147) | 59 (42-81) |
| Cameroon | 129 (90-176) | 78 (54-107) | 43 (30-60) |
| Canada | 369 (242-541) | 90 (54-144) | 262 (173-395) |
| Cape Verde | 4 (3-6) | 3 (2-4) | 2 (1-2) |
| Central African Republic | 19 (13-28) | 13 (9-19) | 7 (5-10) |
| Chad | 45 (31-61) | 27 (19-37) | 15 (10-21) |
| Chile | 128 (89-177) | 58 (40-84) | 52 (35-75) |
| China | 34898 (25826-46618) | 16706 (12073-23151) | 12993 (9314-18291) |
| Colombia | 768 (575-1011) | 407 (292-557) | 254 (179-348) |
| Comoros | 3 (2-3) | 1 (1-2) | 1 (1-1) |
| Congo | 26 (18-38) | 16 (11-24) | 10 (7-14) |
| Costa Rica | 57 (42-78) | 28 (20-39) | 22 (15-31) |
| Cote d'Ivoire | 86 (63-119) | 50 (37-71) | 29 (20-41) |
| Croatia | 261 (189-370) | 113 (78-171) | 105 (72-157) |
| Cuba | 167 (115-225) | 88 (60-122) | 62 (43-87) |
| Cyprus | 15 (10-23) | 6 (4-9) | 9 (6-13) |
| Czech Republic | 515 (379-694) | 191 (133-280) | 242 (171-340) |
| Democratic Republic of the Congo | 82 (38-158) | 73 (34-131) | 51 (27-91) |
| Denmark | 73 (48-105) | 11 (5-18) | 61 (40-88) |
| Djibouti | 3 (2-3) | 1 (1-2) | 1 (1-1) |
| Dominica | 1 (1-1) | 0 (0-1) | 0 (0-0) |
| Dominican Republic | 86 (63-117) | 46 (33-64) | 31 (23-45) |
| Ecuador | 98 (71-131) | 53 (37-73) | 34 (23-46) |
| Egypt | 163 (74-317) | 119 (57-216) | 80 (39-145) |
| El Salvador | 61 (45-78) | 34 (24-44) | 20 (15-28) |
| Equatorial Guinea | 4 (3-6) | 3 (2-4) | 2 (1-3) |
| Eritrea | 14 (10-19) | 8 (6-11) | 4 (3-6) |
| Estonia | 10 (6-17) | 5 (3-9) | 6 (3-11) |
| Ethiopia | 161 (119-214) | 93 (66-125) | 50 (36-70) |
| Federated States of Micronesia | 1 (1-2) | 1 (1-1) | 0 (0-1) |
| Fiji | 14 (10-18) | 7 (5-10) | 4 (3-6) |
| Finland | 105 (65-162) | 27 (16-45) | 79 (50-121) |
| France | 759 (463-1169) | 239 (139-392) | 504 (308-786) |
| Gabon | 10 (7-15) | 6 (4-8) | 4 (3-6) |
| Georgia | 211 (152-290) | 116 (81-162) | 70 (49-103) |
| Germany | 1767 (1082-2651) | 465 (275-754) | 1263 (788-1909) |
| Ghana | 103 (76-137) | 58 (43-79) | 33 (24-46) |
| Greece | 276 (175-411) | 132 (83-199) | 134 (84-201) |
| Greenland | 0 (0-1) | 0 (0-0) | 0 (0-0) |
| Grenada | 1 (1-1) | 0 (0-1) | 0 (0-0) |
| Guatemala | 114 (82-150) | 62 (43-84) | 38 (26-52) |
| Guinea | 50 (35-69) | 31 (22-42) | 17 (12-24) |
| Guinea-Bissau | 7 (5-9) | 4 (3-6) | 2 (2-3) |
| Guyana | 6 (5-9) | 3 (2-5) | 2 (2-3) |
| Haiti | 71 (50-97) | 42 (30-59) | 24 (16-33) |
| Honduras | 127 (91-172) | 70 (49-99) | 40 (28-56) |
| Hungary | 616 (455-831) | 264 (185-378) | 251 (178-358) |
| Iceland | 3 (2-5) | 1 (0-1) | 3 (2-4) |
| India | 10112 (7534-13001) | 5761 (4258-7614) | 3260 (2334-4413) |
| Indonesia | 4356 (3279-5616) | 2380 (1752-3138) | 1419 (1011-1900) |
| Iran | 102 (41-195) | 77 (33-149) | 53 (23-103) |
| Iraq | 56 (25-106) | 40 (19-72) | 28 (13-49) |
| Ireland | 34 (21-53) | 2 (1-5) | 32 (20-50) |
| Israel | 61 (38-87) | 15 (9-24) | 44 (28-63) |
| Italy | 1310 (866-1918) | 443 (283-696) | 765 (488-1157) |
| Jamaica | 27 (19-36) | 15 (10-21) | 9 (7-13) |
| Japan | 3466 (2192-5082) | 986 (570-1602) | 1973 (1208-2937) |
| Jordan | 6 (3-11) | 5 (2-8) | 3 (2-6) |
| Kazakhstan | 490 (344-699) | 249 (169-364) | 181 (124-270) |
| Kenya | 58 (37-82) | 33 (20-49) | 19 (11-28) |
| Kiribati | 1 (1-1) | 1 (0-1) | 0 (0-0) |
| Kuwait | 8 (6-12) | 3 (2-5) | 5 (3-7) |
| Kyrgyzstan | 111 (81-152) | 64 (45-90) | 34 (24-50) |
| Laos | 96 (74-123) | 53 (40-69) | 30 (22-39) |
| Latvia | 53 (32-83) | 28 (17-46) | 29 (17-47) |
| Lebanon | 5 (2-10) | 4 (2-8) | 3 (1-6) |
| Lesotho | 8 (5-13) | 6 (4-9) | 3 (2-5) |
| Liberia | 16 (12-22) | 10 (7-14) | 5 (4-7) |
| Libya | 7 (3-13) | 5 (2-9) | 4 (2-7) |
| Lithuania | 123 (80-185) | 51 (34-80) | 59 (38-93) |
| Luxembourg | 7 (4-10) | 0 (0-1) | 6 (4-9) |
| Macedonia | 156 (106-223) | 79 (52-121) | 51 (32-78) |
| Madagascar | 68 (51-89) | 39 (29-53) | 21 (15-28) |
| Malawi | 40 (29-53) | 23 (17-31) | 12 (9-18) |
| Malaysia | 493 (372-645) | 245 (182-330) | 183 (131-247) |
| Maldives | 4 (3-5) | 2 (1-3) | 1 (1-2) |
| Mali | 48 (33-67) | 29 (20-41) | 16 (11-23) |
| Malta | 8 (5-11) | 3 (2-4) | 4 (3-6) |
| Marshall Islands | 1 (0-1) | 0 (0-0) | 0 (0-0) |
| Mauritania | 19 (13-25) | 11 (8-15) | 6 (4-9) |
| Mauritius | 28 (21-35) | 14 (11-18) | 10 (7-13) |
| Mexico | 1301 (968-1716) | 669 (488-905) | 489 (355-668) |
| Moldova | 69 (43-105) | 46 (28-70) | 32 (20-48) |
| Mongolia | 46 (32-63) | 25 (17-35) | 15 (10-22) |
| Montenegro | 43 (31-61) | 21 (15-32) | 15 (10-22) |
| Morocco | 53 (23-105) | 40 (19-75) | 27 (13-49) |
| Mozambique | 69 (50-92) | 40 (29-55) | 21 (14-30) |
| Myanmar | 935 (713-1242) | 536 (398-722) | 284 (205-392) |
| Namibia | 8 (5-13) | 6 (4-9) | 4 (2-6) |
| Nauru | 0 (0-0) | 0 (0-0) | 0 (0-0) |
| Nepal | 401 (285-545) | 228 (162-318) | 123 (83-174) |
| Netherlands | 207 (131-310) | 44 (25-74) | 160 (102-243) |
| New Zealand | 43 (27-65) | 13 (8-21) | 32 (19-49) |
| Nicaragua | 54 (40-70) | 30 (22-40) | 17 (12-23) |
| Niger | 47 (33-64) | 29 (21-40) | 15 (10-21) |
| Nigeria | 735 (514-1028) | 463 (323-637) | 270 (184-378) |
| Norway | 40 (25-61) | 3 (2-7) | 38 (23-57) |
| Oman | 3 (1-5) | 2 (1-3) | 2 (1-3) |
| Pakistan | 1394 (1048-1886) | 796 (587-1098) | 434 (313-606) |
| Palau | 0 (0-0) | 0 (0-0) | 0 (0-0) |
| Panama | 47 (34-61) | 22 (15-29) | 19 (13-26) |
| Papua New Guinea | 58 (43-77) | 32 (24-44) | 18 (13-25) |
| Paraguay | 64 (46-86) | 34 (24-47) | 22 (15-31) |
| Peru | 118 (84-160) | 63 (44-87) | 40 (28-57) |
| Philippines | 1376 (1051-1746) | 764 (578-1005) | 441 (324-588) |
| Poland | 1572 (1153-2204) | 685 (477-1015) | 648 (444-943) |
| Portugal | 88 (51-147) | 47 (26-77) | 52 (29-87) |
| Puerto Rico | 38 (28-53) | 14 (10-20) | 21 (15-29) |
| Qatar | 1 (0-1) | 0 (0-0) | 1 (0-1) |
| Romania | 1670 (1185-2276) | 753 (502-1086) | 634 (431-926) |
| Russia | 3683 (2442-5541) | 1883 (1223-2854) | 1473 (952-2212) |
| Rwanda | 29 (21-40) | 17 (12-23) | 9 (6-13) |
| Saint Kitts and Nevis | 0 (0-1) | 0 (0-0) | 0 (0-0) |
| Saint Lucia | 2 (1-3) | 1 (1-1) | 1 (1-1) |
| Saint Vincent and the Grenadines | 1 (1-2) | 1 (0-1) | 1 (0-1) |
| Samoa | 2 (1-2) | 1 (1-1) | 1 (0-1) |
| Sao Tome and Principe | 1 (1-1) | 1 (0-1) | 0 (0-0) |
| Saudi Arabia | 36 (18-62) | 20 (11-33) | 23 (12-37) |
| Senegal | 50 (35-69) | 30 (21-42) | 17 (12-24) |
| Serbia | 546 (399-743) | 270 (190-390) | 185 (126-273) |
| Seychelles | 2 (1-2) | 1 (1-1) | 1 (0-1) |
| Sierra Leone | 32 (23-45) | 20 (14-28) | 11 (7-15) |
| Singapore | 46 (32-62) | 4 (2-8) | 38 (27-54) |
| Slovakia | 270 (192-366) | 108 (74-153) | 116 (77-167) |
| Slovenia | 88 (61-122) | 31 (20-46) | 43 (29-63) |
| Solomon Islands | 7 (5-9) | 4 (3-5) | 2 (2-3) |
| Somalia | 26 (19-36) | 15 (11-21) | 8 (5-12) |
| South Africa | 206 (116-341) | 154 (90-241) | 112 (67-171) |
| South Korea | 789 (532-1141) | 242 (150-386) | 411 (260-614) |
| Spain | 89 (30-225) | 67 (18-165) | 78 (23-201) |
| Sri Lanka | 398 (303-527) | 218 (162-297) | 129 (93-175) |
| Sudan | 40 (18-79) | 31 (14-56) | 19 (9-35) |
| Suriname | 4 (3-6) | 3 (2-3) | 2 (1-2) |
| Swaziland | 3 (2-5) | 2 (1-4) | 2 (1-2) |
| Sweden | 168 (103-248) | 31 (18-55) | 133 (81-197) |
| Switzerland | 131 (85-197) | 9 (5-20) | 121 (78-184) |
| Syria | 30 (13-58) | 23 (10-41) | 14 (7-25) |
| Taiwan (Province of China) | 307 (220-417) | 104 (70-148) | 160 (114-224) |
| Tajikistan | 120 (88-163) | 70 (50-96) | 37 (26-53) |
| Tanzania | 143 (107-187) | 82 (61-109) | 44 (31-59) |
| Thailand | 1320 (992-1688) | 694 (512-912) | 454 (324-615) |
| The Bahamas | 3 (2-3) | 1 (1-1) | 1 (1-2) |
| The Gambia | 10 (7-13) | 6 (4-8) | 3 (2-5) |
| Timor-Leste | 18 (13-24) | 10 (7-14) | 5 (4-8) |
| Togo | 28 (20-38) | 17 (12-23) | 9 (7-13) |
| Tonga | 1 (1-2) | 1 (0-1) | 0 (0-1) |
| Trinidad and Tobago | 14 (10-19) | 7 (5-10) | 6 (4-9) |
| Tunisia | 18 (8-37) | 14 (6-27) | 9 (4-19) |
| Turkey | 42 (11-109) | 36 (10-91) | 31 (7-72) |
| Turkmenistan | 130 (95-173) | 69 (49-95) | 47 (33-64) |
| Tuvalu | 0 (0-0) | 0 (0-0) | 0 (0-0) |
| Uganda | 69 (49-93) | 40 (28-56) | 21 (14-29) |
| Ukraine | 1096 (664-1756) | 761 (448-1218) | 507 (311-817) |
| United Arab Emirates | 4 (2-6) | 1 (1-3) | 3 (1-5) |
| United Kingdom | 931 (601-1369) | 269 (170-421) | 634 (413-940) |
| United States | 5621 (3775-7953) | 797 (447-1332) | 4669 (3192-6763) |
| Uruguay | 56 (39-79) | 25 (17-37) | 24 (15-34) |
| Uzbekistan | 714 (527-975) | 412 (293-573) | 227 (162-320) |
| Vanuatu | 3 (3-5) | 2 (1-3) | 1 (1-2) |
| Venezuela | 496 (371-664) | 281 (205-384) | 156 (110-216) |
| Vietnam | 1139 (860-1516) | 627 (463-856) | 368 (264-506) |
| Yemen | 30 (13-60) | 23 (10-43) | 14 (6-26) |
| Zambia | 42 (31-56) | 24 (18-33) | 13 (9-18) |
| Zimbabwe | 70 (50-98) | 43 (30-59) | 24 (17-34) |

#### c. **Net effect in patients with CKD**

| **Country** | **Replacement of discretionary salt and non-discretionary salt Median (95% UI)** | **Replacement of discretionary salt Median (95% UI)** | **Replacement of non-discretionary salt  Median (95% UI)** |
| --- | --- | --- | --- |
| Afghanistan | 204 (111-324) | 171 (95-267) | 114 (64-180) |
| Albania | 862 (713-1010) | 590 (484-708) | 421 (341-520) |
| Algeria | 375 (190-651) | 310 (152-551) | 232 (117-404) |
| American Samoa | 3 (3-4) | 2 (2-3) | 2 (2-2) |
| Andorra | 3 (2-5) | 2 (1-3) | 3 (2-4) |
| Angola | 813 (618-1029) | 607 (464-768) | 388 (294-484) |
| Antigua and Barbuda | 6 (5-7) | 4 (3-5) | 3 (3-4) |
| Argentina | 2529 (2110-3013) | 1636 (1330-1995) | 1292 (1026-1591) |
| Armenia | 835 (675-1013) | 568 (456-692) | 388 (304-486) |
| Australia | 623 (334-985) | 236 (123-391) | 588 (311-926) |
| Austria | 1671 (1313-2018) | 522 (369-713) | 1329 (1045-1669) |
| Azerbaijan | 1948 (1589-2316) | 1325 (1076-1598) | 913 (740-1118) |
| Bahrain | 7 (4-11) | 5 (3-7) | 5 (3-8) |
| Bangladesh | 11745 (9741-13842) | 8337 (6861-9977) | 5284 (4203-6410) |
| Barbados | 17 (12-22) | 11 (8-14) | 11 (8-13) |
| Belarus | 1424 (1002-2000) | 1087 (761-1502) | 821 (579-1133) |
| Belgium | 1104 (887-1313) | 362 (260-484) | 875 (686-1075) |
| Belize | 12 (10-14) | 8 (7-10) | 6 (5-7) |
| Benin | 296 (248-349) | 212 (174-253) | 130 (106-159) |
| Bermuda | 6 (5-7) | 0 (0-1) | 6 (4-7) |
| Bhutan | 52 (43-62) | 37 (30-44) | 24 (19-29) |
| Bolivia | 379 (303-455) | 265 (211-327) | 173 (135-213) |
| Bosnia and Herzegovina | 1072 (889-1278) | 728 (599-890) | 533 (432-660) |
| Botswana | 55 (37-74) | 42 (30-56) | 32 (23-42) |
| Brazil | 14583 (12433-16982) | 9783 (8132-11505) | 7130 (5959-8514) |
| Brunei | 22 (18-26) | 11 (9-13) | 14 (12-17) |
| Bulgaria | 5220 (4339-6064) | 3419 (2750-4117) | 2847 (2304-3451) |
| Burkina Faso | 481 (376-592) | 351 (271-438) | 213 (166-271) |
| Burundi | 175 (147-207) | 126 (105-152) | 76 (62-92) |
| Cambodia | 1162 (930-1413) | 821 (660-1010) | 519 (410-645) |
| Cameroon | 955 (757-1161) | 687 (545-847) | 427 (339-521) |
| Canada | 2306 (1770-2869) | 768 (549-1031) | 1871 (1421-2335) |
| Cape Verde | 29 (23-35) | 20 (16-25) | 13 (10-17) |
| Central African Republic | 169 (125-215) | 129 (98-164) | 80 (60-103) |
| Chad | 330 (264-401) | 240 (191-294) | 145 (112-179) |
| Chile | 1034 (836-1218) | 626 (495-760) | 569 (449-689) |
| China | 213108 (180102-252519) | 136673 (113362-164010) | 113154 (93196-137394) |
| Colombia | 4676 (3961-5465) | 3188 (2691-3817) | 2262 (1861-2720) |
| Comoros | 19 (15-23) | 13 (11-17) | 8 (7-10) |
| Congo | 219 (169-276) | 164 (126-206) | 107 (82-132) |
| Costa Rica | 403 (341-469) | 254 (213-300) | 210 (175-249) |
| Cote d'Ivoire | 659 (535-784) | 466 (374-562) | 296 (240-358) |
| Croatia | 1530 (1234-1805) | 920 (730-1128) | 877 (691-1085) |
| Cuba | 1085 (872-1293) | 712 (567-865) | 546 (435-663) |
| Cyprus | 99 (75-130) | 49 (37-68) | 69 (51-91) |
| Czech Republic | 2907 (2381-3413) | 1571 (1267-1908) | 1854 (1510-2254) |
| Democratic Republic of the Congo | 802 (411-1309) | 739 (398-1193) | 551 (307-865) |
| Denmark | 417 (309-524) | 85 (53-130) | 374 (277-463) |
| Djibouti | 20 (16-23) | 14 (11-17) | 9 (7-11) |
| Dominica | 6 (5-7) | 4 (3-5) | 3 (2-4) |
| Dominican Republic | 612 (498-735) | 409 (333-494) | 304 (243-375) |
| Ecuador | 659 (534-772) | 449 (358-541) | 314 (250-381) |
| Egypt | 1471 (807-2254) | 1193 (699-1805) | 864 (509-1318) |
| El Salvador | 379 (313-445) | 258 (210-304) | 174 (141-209) |
| Equatorial Guinea | 36 (28-45) | 25 (20-32) | 19 (14-24) |
| Eritrea | 104 (83-129) | 76 (59-94) | 46 (36-58) |
| Estonia | 83 (48-129) | 48 (29-86) | 58 (35-98) |
| Ethiopia | 1106 (872-1348) | 800 (629-979) | 495 (383-619) |
| Federated States of Micronesia | 9 (7-11) | 7 (5-8) | 4 (3-5) |
| Fiji | 93 (77-108) | 63 (52-74) | 43 (34-52) |
| Finland | 709 (508-943) | 249 (170-356) | 598 (423-795) |
| France | 4439 (3143-5848) | 1830 (1279-2538) | 3402 (2424-4516) |
| Gabon | 86 (65-108) | 60 (45-75) | 45 (34-58) |
| Georgia | 1544 (1289-1838) | 1066 (863-1290) | 727 (581-893) |
| Germany | 11892 (8609-15414) | 4240 (2934-5861) | 9749 (7083-12615) |
| Ghana | 743 (627-860) | 526 (439-626) | 333 (272-398) |
| Greece | 1739 (1249-2256) | 1040 (766-1367) | 1046 (765-1370) |
| Greenland | 2 (2-3) | 1 (1-1) | 2 (2-3) |
| Grenada | 6 (5-8) | 4 (3-5) | 3 (2-4) |
| Guatemala | 737 (607-874) | 504 (412-613) | 344 (279-417) |
| Guinea | 360 (287-437) | 262 (208-318) | 160 (127-196) |
| Guinea-Bissau | 54 (43-66) | 39 (31-48) | 24 (19-30) |
| Guyana | 50 (41-60) | 33 (27-41) | 26 (21-32) |
| Haiti | 541 (442-642) | 392 (319-469) | 244 (197-298) |
| Honduras | 946 (781-1118) | 656 (541-788) | 423 (342-518) |
| Hungary | 3820 (3198-4441) | 2308 (1901-2788) | 2236 (1833-2710) |
| Iceland | 20 (15-26) | 4 (3-7) | 18 (14-23) |
| India | 67336 (57009-77299) | 47260 (39799-55603) | 29696 (24279-35292) |
| Indonesia | 28433 (24224-33075) | 19373 (16278-22878) | 13031 (10686-15497) |
| Iran | 925 (454-1457) | 768 (395-1212) | 557 (296-893) |
| Iraq | 506 (278-762) | 406 (233-586) | 296 (179-432) |
| Ireland | 180 (122-243) | 18 (11-32) | 174 (119-239) |
| Israel | 353 (267-450) | 120 (88-163) | 288 (217-369) |
| Italy | 8854 (6458-11532) | 4107 (2980-5618) | 6299 (4602-8251) |
| Jamaica | 191 (155-230) | 133 (107-163) | 91 (74-113) |
| Japan | 23743 (19228-28522) | 9919 (7710-12544) | 16747 (13437-20682) |
| Jordan | 57 (32-89) | 46 (26-71) | 34 (19-51) |
| Kazakhstan | 3327 (2696-4039) | 2146 (1720-2586) | 1681 (1348-2090) |
| Kenya | 443 (331-569) | 316 (235-411) | 202 (147-266) |
| Kiribati | 7 (5-8) | 5 (4-6) | 3 (2-4) |
| Kuwait | 63 (48-81) | 33 (25-43) | 44 (34-57) |
| Kyrgyzstan | 794 (660-950) | 565 (459-679) | 349 (281-432) |
| Laos | 596 (500-697) | 417 (342-493) | 268 (220-318) |
| Latvia | 366 (255-501) | 243 (165-331) | 246 (165-344) |
| Lebanon | 41 (19-73) | 34 (16-60) | 25 (12-44) |
| Lesotho | 70 (47-96) | 57 (39-76) | 37 (26-50) |
| Liberia | 123 (99-148) | 89 (71-109) | 54 (43-66) |
| Libya | 63 (34-100) | 51 (29-81) | 39 (22-61) |
| Lithuania | 826 (623-1045) | 447 (329-589) | 497 (365-644) |
| Luxembourg | 40 (29-50) | 3 (2-4) | 39 (28-49) |
| Macedonia | 971 (788-1146) | 661 (526-801) | 477 (378-592) |
| Madagascar | 535 (441-628) | 387 (316-457) | 235 (188-287) |
| Malawi | 293 (241-348) | 211 (171-253) | 128 (103-158) |
| Malaysia | 3054 (2613-3518) | 1932 (1637-2269) | 1540 (1292-1841) |
| Maldives | 21 (18-25) | 14 (11-17) | 11 (9-13) |
| Mali | 355 (272-442) | 258 (198-322) | 158 (120-196) |
| Malta | 48 (37-60) | 23 (17-29) | 33 (25-42) |
| Marshall Islands | 4 (3-5) | 3 (2-3) | 2 (1-2) |
| Mauritania | 132 (107-162) | 95 (76-116) | 59 (47-74) |
| Mauritius | 178 (152-207) | 116 (97-138) | 89 (73-105) |
| Mexico | 8733 (7071-10232) | 5674 (4558-6757) | 4433 (3486-5372) |
| Moldova | 563 (397-759) | 435 (299-588) | 314 (218-423) |
| Mongolia | 335 (279-398) | 231 (189-277) | 155 (127-188) |
| Montenegro | 252 (209-301) | 168 (136-205) | 129 (104-160) |
| Morocco | 482 (253-745) | 395 (215-604) | 287 (160-436) |
| Mozambique | 546 (444-666) | 396 (316-486) | 239 (191-299) |
| Myanmar | 5979 (4986-6897) | 4204 (3497-4936) | 2616 (2141-3068) |
| Namibia | 67 (45-91) | 53 (36-71) | 38 (26-51) |
| Nauru | 1 (1-1) | 1 (1-1) | 1 (0-1) |
| Nepal | 2643 (2026-3260) | 1911 (1462-2416) | 1185 (892-1514) |
| Netherlands | 1127 (843-1439) | 325 (220-474) | 962 (718-1228) |
| New Zealand | 269 (192-351) | 105 (71-143) | 222 (159-295) |
| Nicaragua | 386 (323-451) | 270 (225-318) | 169 (141-202) |
| Niger | 348 (273-424) | 252 (199-311) | 152 (120-190) |
| Nigeria | 5245 (4090-6508) | 3892 (3029-4815) | 2462 (1939-3015) |
| Norway | 233 (161-321) | 27 (16-48) | 224 (155-309) |
| Oman | 25 (14-39) | 18 (11-27) | 18 (10-27) |
| Pakistan | 10497 (9068-12208) | 7509 (6318-8874) | 4653 (3864-5559) |
| Palau | 2 (2-3) | 1 (1-2) | 1 (1-1) |
| Panama | 300 (255-351) | 181 (150-219) | 162 (136-194) |
| Papua New Guinea | 323 (237-397) | 231 (168-285) | 149 (110-185) |
| Paraguay | 468 (390-544) | 316 (263-374) | 222 (183-265) |
| Peru | 752 (613-888) | 510 (402-611) | 360 (290-440) |
| Philippines | 8628 (7192-10019) | 6001 (4974-7125) | 3961 (3256-4753) |
| Poland | 9273 (7289-11317) | 5495 (4249-6891) | 5244 (4110-6542) |
| Portugal | 533 (352-772) | 341 (221-495) | 376 (244-537) |
| Puerto Rico | 266 (214-317) | 130 (101-163) | 179 (143-218) |
| Qatar | 5 (3-8) | 2 (1-2) | 5 (3-7) |
| Romania | 10412 (8606-12136) | 6560 (5315-7917) | 5792 (4680-7047) |
| Russia | 25329 (19661-31479) | 16053 (12476-20049) | 13181 (10298-16674) |
| Rwanda | 210 (169-259) | 151 (121-187) | 93 (72-117) |
| Saint Kitts and Nevis | 4 (3-4) | 2 (2-3) | 2 (2-3) |
| Saint Lucia | 14 (11-16) | 9 (7-11) | 7 (6-9) |
| Saint Vincent and the Grenadines | 10 (8-13) | 7 (5-9) | 5 (4-7) |
| Samoa | 11 (9-14) | 8 (6-10) | 5 (4-7) |
| Sao Tome and Principe | 6 (5-7) | 4 (3-5) | 3 (2-3) |
| Saudi Arabia | 317 (190-474) | 208 (132-297) | 230 (143-327) |
| Senegal | 357 (282-430) | 257 (201-313) | 159 (127-199) |
| Serbia | 3048 (2531-3518) | 2052 (1673-2429) | 1581 (1283-1879) |
| Seychelles | 12 (10-14) | 8 (6-9) | 6 (5-8) |
| Sierra Leone | 235 (186-284) | 171 (134-208) | 103 (82-126) |
| Singapore | 311 (250-370) | 48 (31-72) | 284 (226-336) |
| Slovakia | 1608 (1311-1890) | 922 (733-1123) | 964 (776-1172) |
| Slovenia | 495 (397-598) | 263 (201-335) | 330 (257-411) |
| Solomon Islands | 42 (33-50) | 29 (23-36) | 19 (15-23) |
| Somalia | 206 (165-255) | 149 (118-187) | 91 (71-115) |
| South Africa | 1741 (1114-2462) | 1418 (936-1993) | 1096 (746-1475) |
| South Korea | 4771 (3693-5911) | 2281 (1726-2950) | 3319 (2524-4217) |
| Spain | 696 (270-1579) | 561 (203-1161) | 625 (228-1382) |
| Sri Lanka | 2381 (1963-2813) | 1631 (1325-1945) | 1085 (870-1323) |
| Sudan | 371 (208-563) | 309 (183-460) | 207 (122-305) |
| Suriname | 32 (26-38) | 22 (18-27) | 15 (12-18) |
| Swaziland | 30 (21-41) | 24 (17-32) | 17 (12-22) |
| Sweden | 1015 (748-1312) | 268 (170-391) | 879 (655-1115) |
| Switzerland | 859 (606-1127) | 84 (53-167) | 821 (577-1072) |
| Syria | 256 (140-397) | 212 (122-323) | 142 (84-213) |
| Taiwan (Province of China) | 1980 (1628-2344) | 945 (750-1163) | 1321 (1074-1598) |
| Tajikistan | 881 (731-1042) | 632 (522-763) | 388 (314-467) |
| Tanzania | 948 (761-1136) | 671 (534-815) | 415 (324-510) |
| Thailand | 6877 (5624-8231) | 4603 (3740-5543) | 3322 (2647-4024) |
| The Bahamas | 22 (18-26) | 12 (9-14) | 14 (11-18) |
| The Gambia | 71 (56-86) | 51 (40-64) | 31 (24-39) |
| Timor-Leste | 113 (93-134) | 79 (64-94) | 51 (41-62) |
| Togo | 211 (168-253) | 153 (122-186) | 93 (74-116) |
| Tonga | 7 (6-8) | 5 (4-6) | 3 (3-4) |
| Trinidad and Tobago | 99 (81-119) | 62 (50-75) | 56 (45-68) |
| Tunisia | 155 (74-255) | 127 (63-212) | 93 (46-153) |
| Turkey | 367 (109-773) | 329 (99-693) | 283 (84-578) |
| Turkmenistan | 940 (783-1096) | 628 (518-746) | 463 (376-549) |
| Tuvalu | 2 (1-2) | 1 (1-1) | 1 (1-1) |
| Uganda | 500 (396-615) | 359 (281-447) | 220 (172-275) |
| Ukraine | 7606 (5155-10768) | 5976 (3941-8506) | 4277 (2863-6003) |
| United Arab Emirates | 32 (18-47) | 16 (9-23) | 27 (16-40) |
| United Kingdom | 5002 (3700-6349) | 1922 (1398-2564) | 3909 (2911-4974) |
| United States | 36893 (28666-45238) | 7481 (4902-10719) | 33354 (25789-41103) |
| Uruguay | 417 (339-493) | 247 (195-300) | 234 (184-285) |
| Uzbekistan | 5076 (4133-6021) | 3606 (2908-4306) | 2273 (1805-2729) |
| Vanuatu | 23 (20-27) | 16 (14-19) | 11 (9-13) |
| Venezuela | 3647 (3099-4240) | 2546 (2128-3011) | 1602 (1326-1931) |
| Vietnam | 6888 (5720-8154) | 4750 (3876-5639) | 3131 (2538-3823) |
| Yemen | 261 (142-405) | 218 (123-335) | 145 (82-217) |
| Zambia | 309 (245-371) | 223 (175-268) | 137 (107-168) |
| Zimbabwe | 563 (442-686) | 409 (321-504) | 255 (198-314) |

### **Supplementary table 6. Sensitivity analyses of the net effect on CVD and CKD combined of replacing regular salt with potassium-enriched salt**

| **Analyses** | **Deaths in millions (95% UI)** | **New cases in millions (95% UI)** | **DALYs in millions (95% UI)** |
| --- | --- | --- | --- |
| Sensitivity analysis 1: Replacing 50% of all regular salt among all adults | 1.72 (1.62-1.83) | 5.89 (5.51-6.25) | 40.57 (37.93-42.88) |
| Sensitivity analysis 2: Replacing 20% of all regular salt among all adults | 0.7 (0.7-0.8) | 2.5 (2.3-2.6) | 17.2 (16.1-18.3) |
| Sensitivity analyses 3: Replacing all regular salt among all adults, assuming data on dose-response between sodium reduction and SBP from Filippini et al^41^ | 2.52 (2.38-2.66) | 8.95 (8.43-9.41) | 61.45 (57.91-64.60) |
| Sensitivity analyses 4: Replacing all regular salt among all adults, assuming only 80% of the additive effect of sodium reduction and potassium increase on SBP. | 2.42 (2.30-2.56) | 8.36 (7.88-8.81) | 57.38 (54.18-60.33) |
| Sensitivity analyses 5: Replacing all regular salt among all adults, assuming only 50% effect of potassium increase on blood pressure | 2.35 (2.23-2.48) | 8.06 (7.60-8.48) | 55.09 (52.00-57.83) |
| Sensitivity analysis 6: Assuming potassium supplementation is only effective among people with hypertension | 2.42 (2.29-2.55) | 8.03 (7.59-8.46) | 54.29 (51.35-57.05) |
| Sensitivity analyses 7: Replacing all regular salt among all adults, assuming 85% NaCl+15% KCl salt substitute composition | 2.02 (1.90-2.15) | 6.92 (6.48-7.33) | 47.55 (44.49-50.20) |
| Sensitivity analyses 8: Replacing discretionary salt among all adults, with the proportion of dictionary salt estimated from published data for 24 countries and the rest of the countries estimated from a regression model | 2.10 (1.97-2.24) | 7.07 (6.59-7.50) | 50.20 (46.84-53.18) |
| Sensitivity analyses 9: Replacing non-discretionary salt among all adults, with the proportion of non-dictionary salt estimated from published data for 24 countries and the rest of the countries estimated from a regression model | 1.28 (1.21-1.35) | 4.45 (4.19-4.70) | 29.19 (27.41-30.71) |
| Sensitivity analyses 10: Replacing discretionary salt among adults with diagnosed hypertension, assuming 30% have CKD stage 3-5 in low and middle-income countries and 20% % have CKD stage 3-5 in high-income countries and those with CKD stages 3-5 were excluded from the intervention | 0.53 (0.49-0.56) | 1.55 (1.45-1.66) | 10.40 (9.71-11.14) |
| Sensitivity analyses 11: Replacing discretionary salt among adults with treated hypertension, assuming 30% have CKD stage 3-5 in low and middle-income countries and 20% % have CKD stage 3-5 in high-income countries and those with CKD stages 3-5 were excluded from the intervention | 0.43 (0.40-0.46) | 1.27 (1.18-1.37) | 8.35 (7.75-8.92) |

### **Supplementary figure 1 Types of deaths, new cases and disability-adjusted life years from cardiovascular disease or kidney disease prevented under different implementation scenarios for potassium-enriched salt, worldwide**

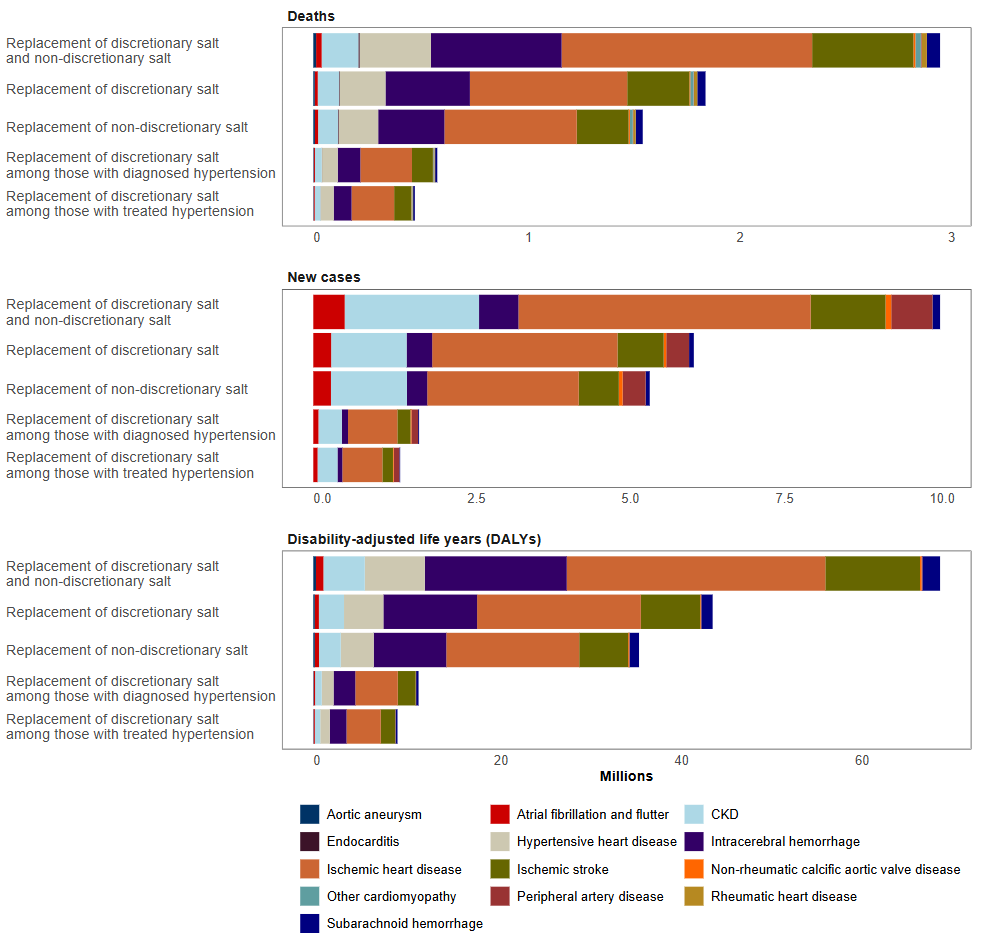

### **Supplementary figure 2. Numbers of new cases caused by cardiovascular disease or kidney disease prevented under different implementation scenarios for potassium-enriched salt for 193 countries and territories**

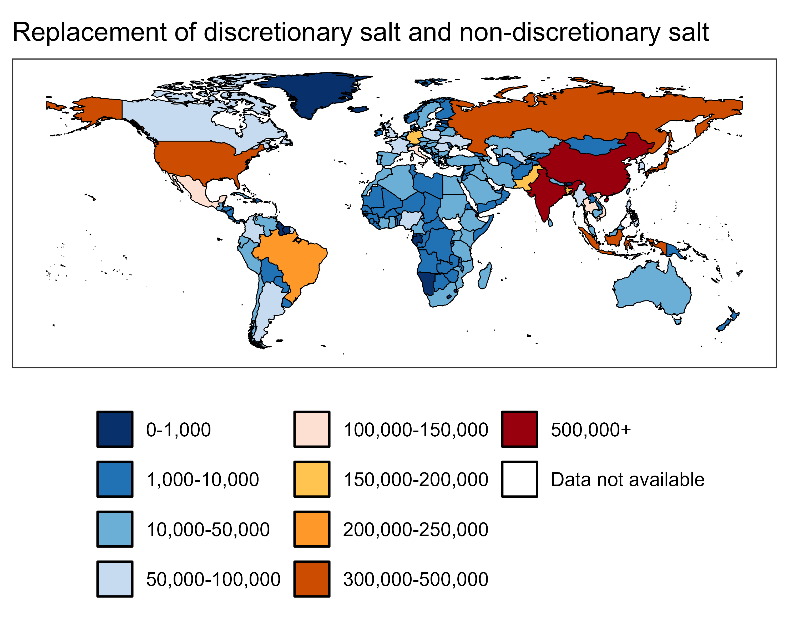

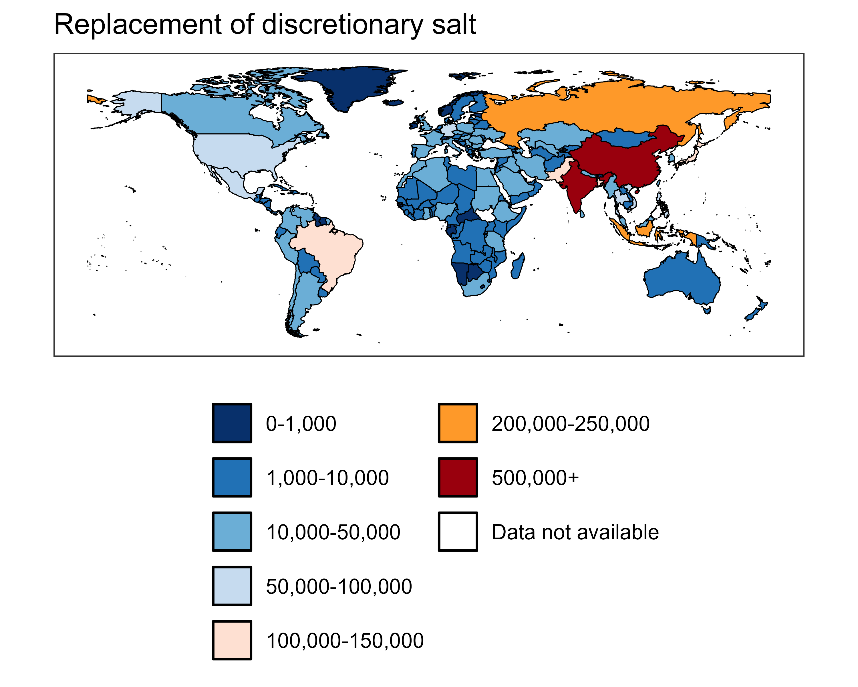

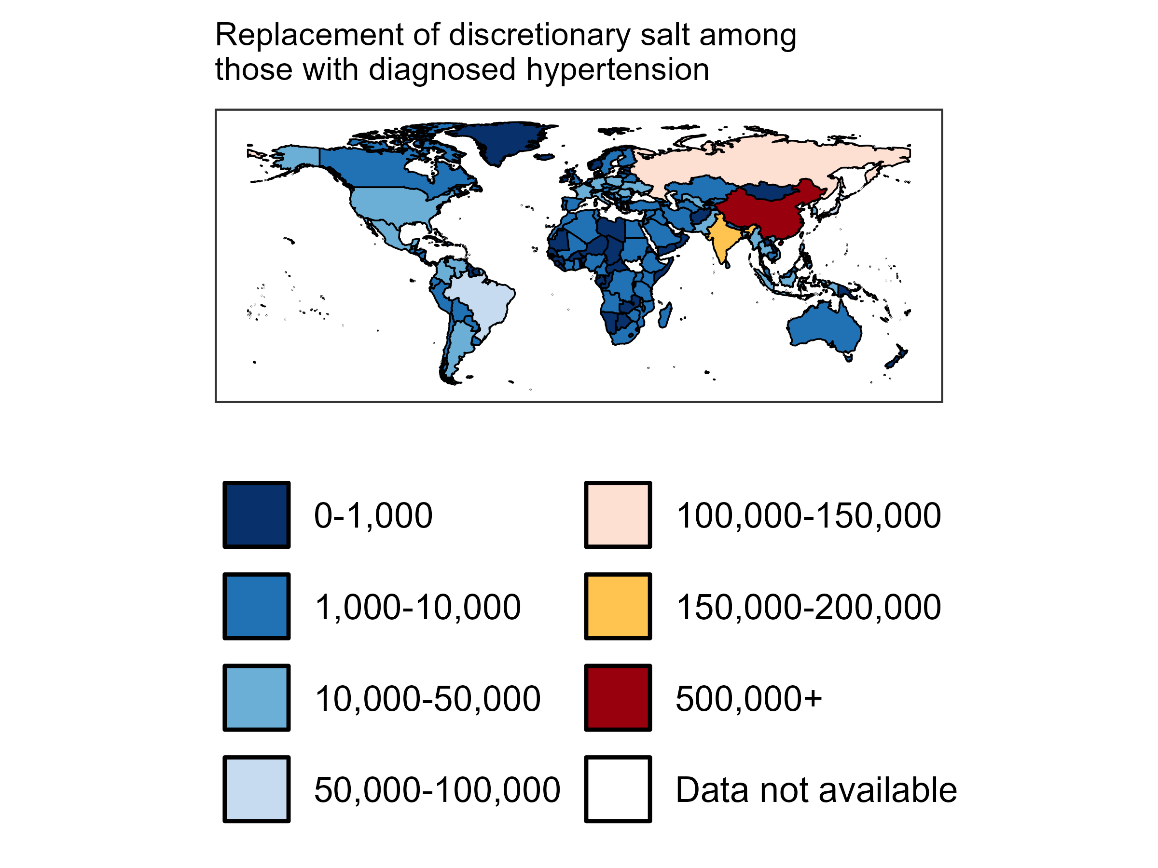

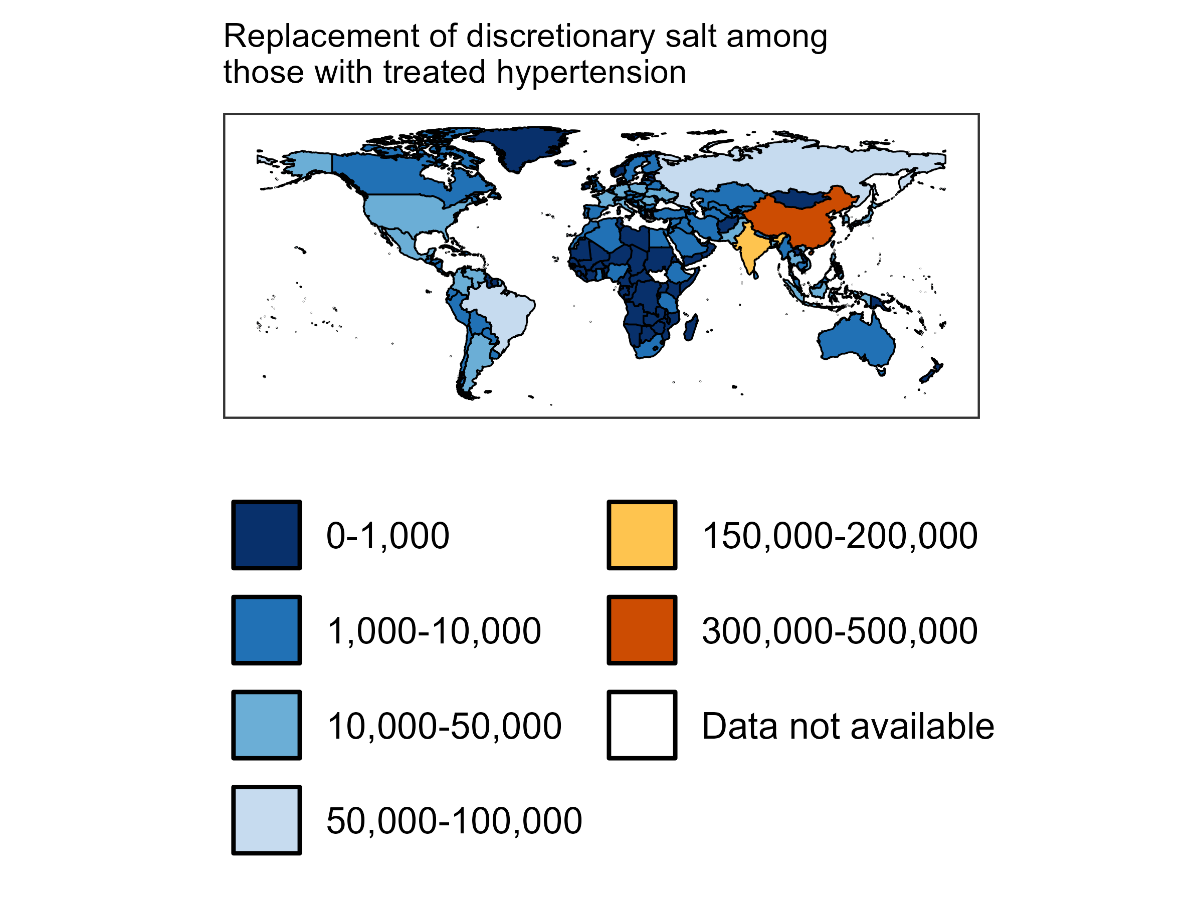

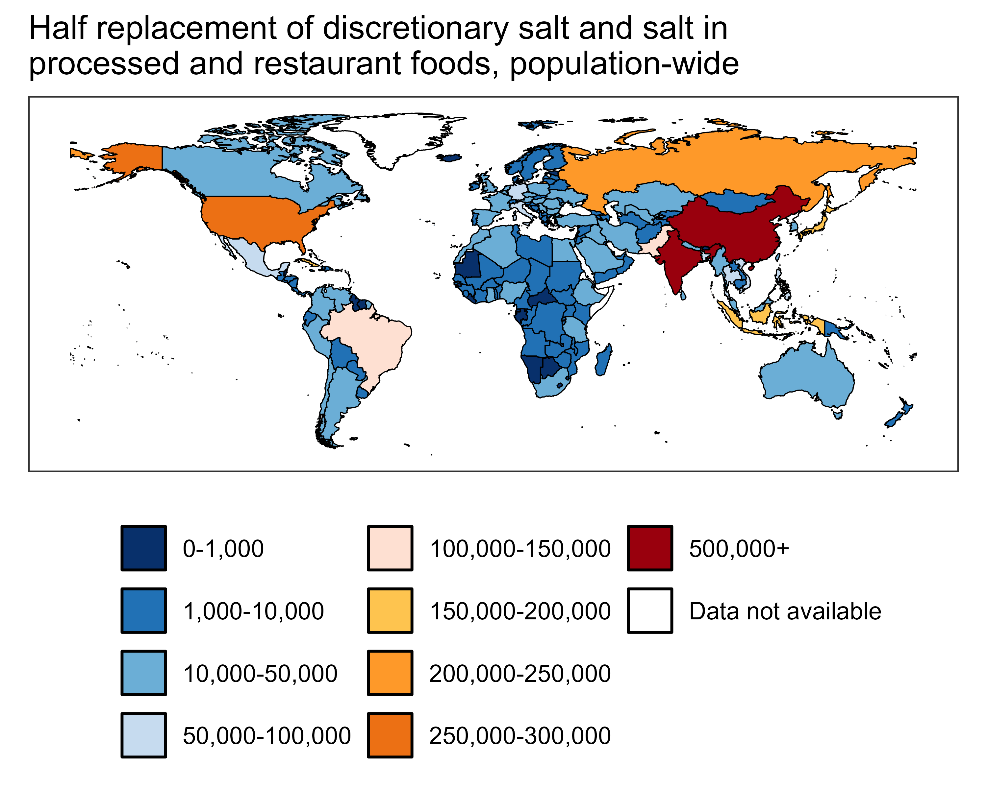

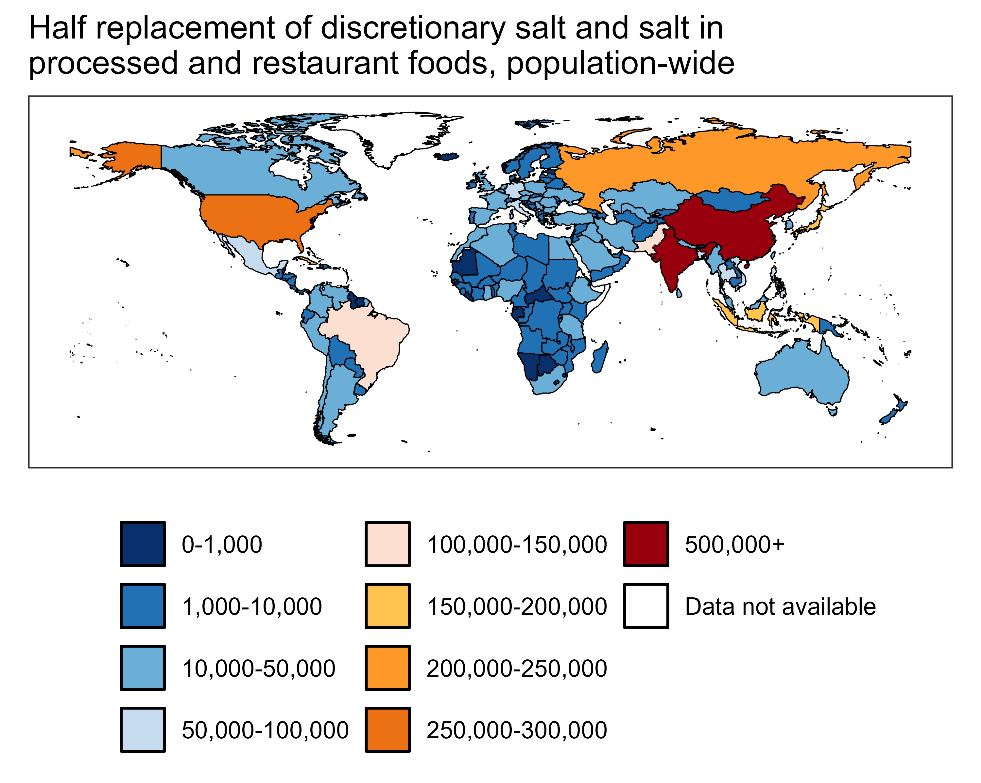

**Legend**

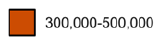

### **Supplementary figure 3. Numbers of disability-adjusted life years caused by cardiovascular disease or kidney disease prevented under different implementation scenarios for potassium-enriched salt for 193 countries and territories**

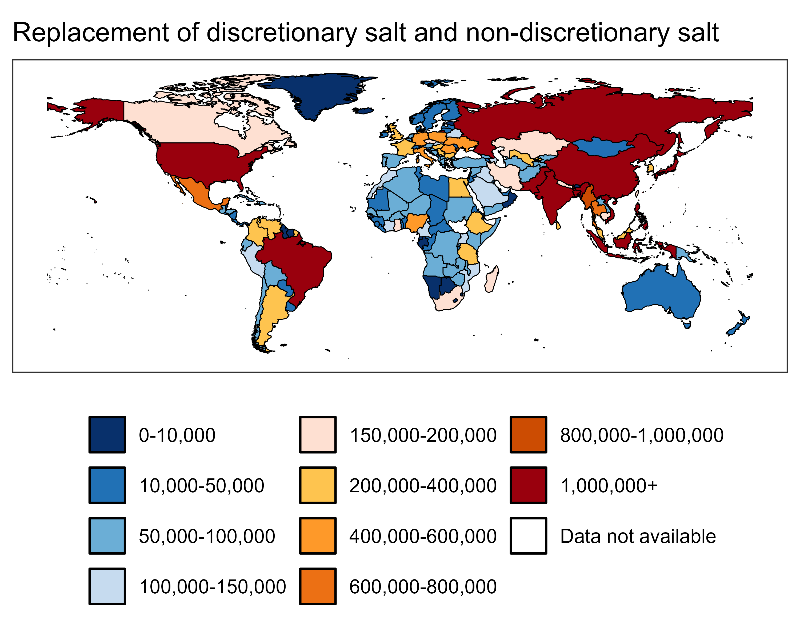

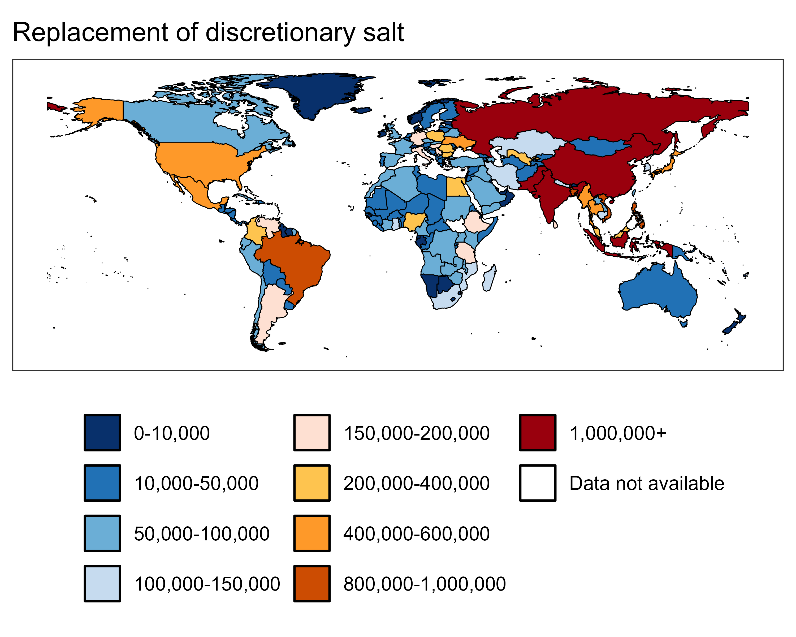

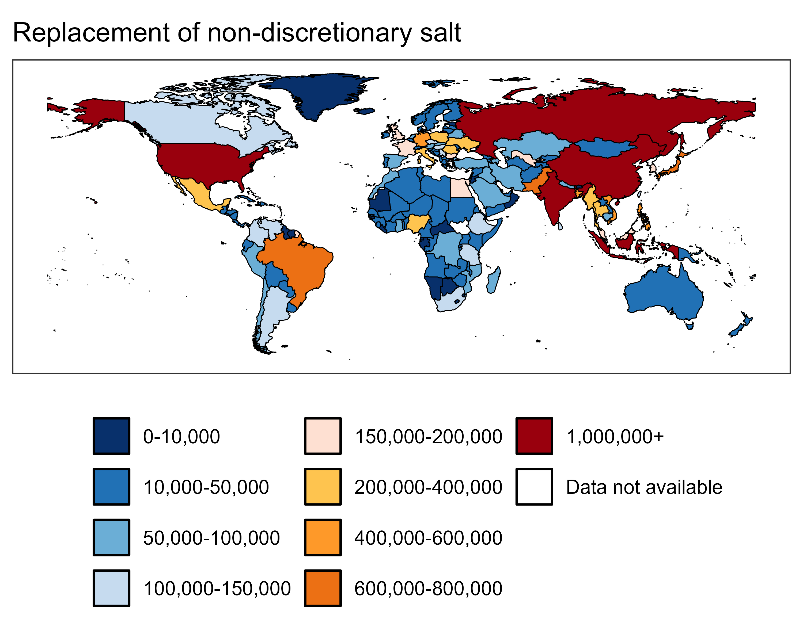

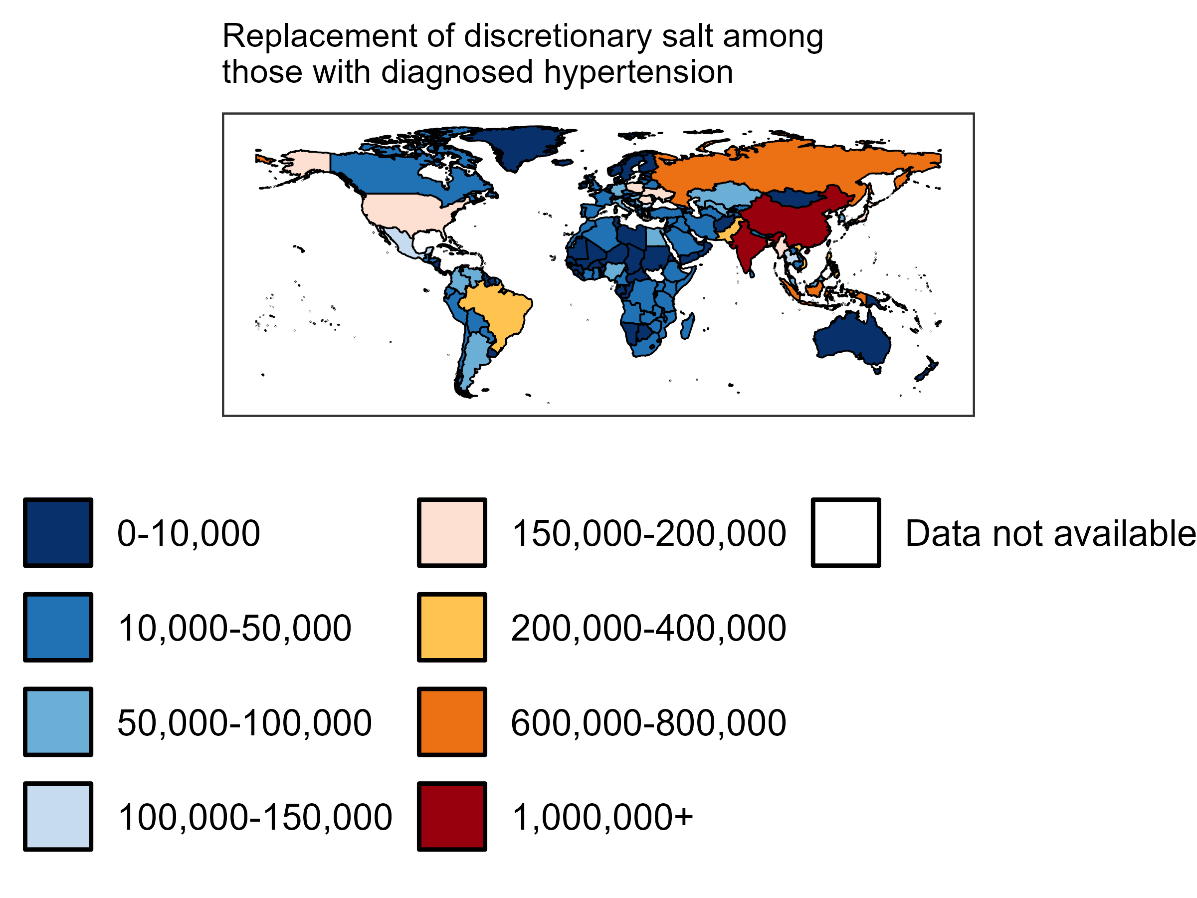

**Legend**

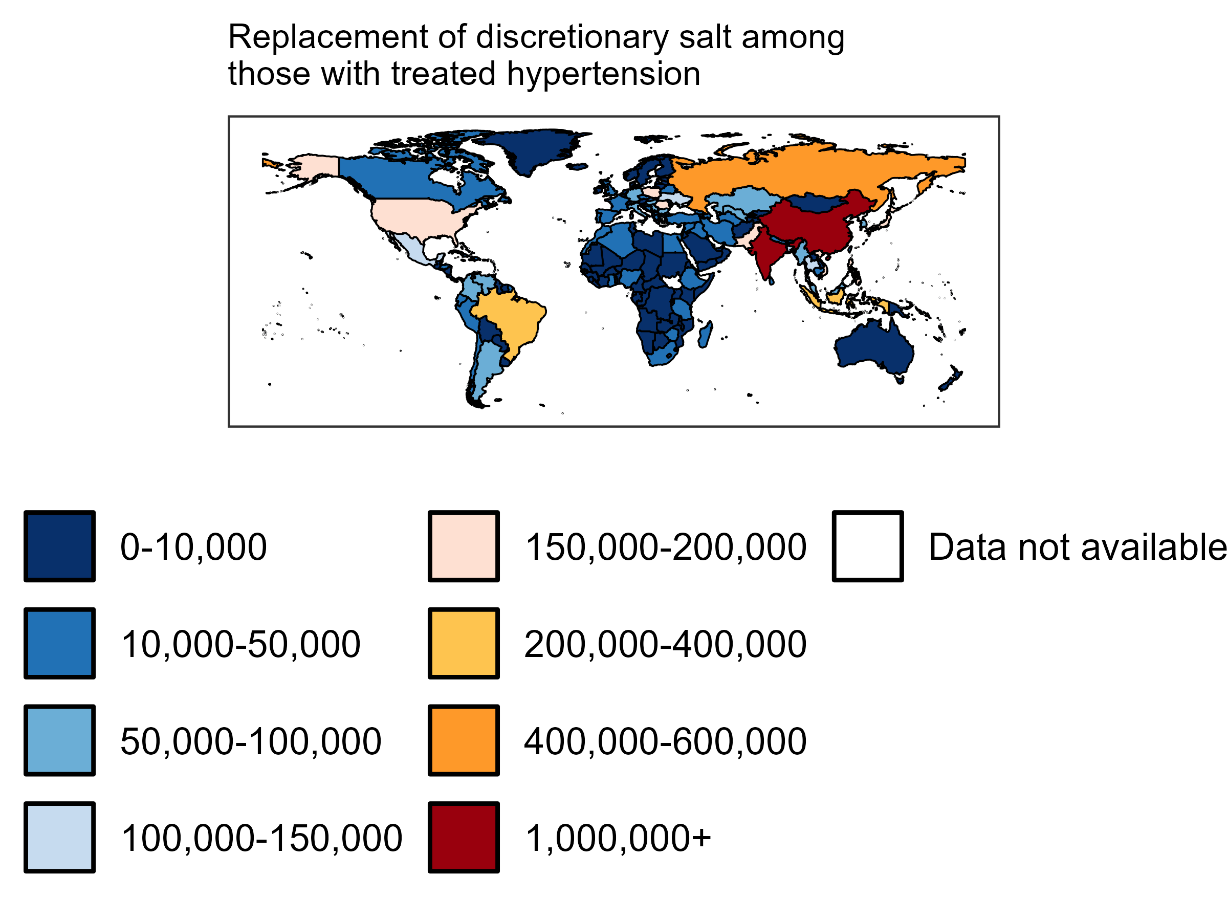

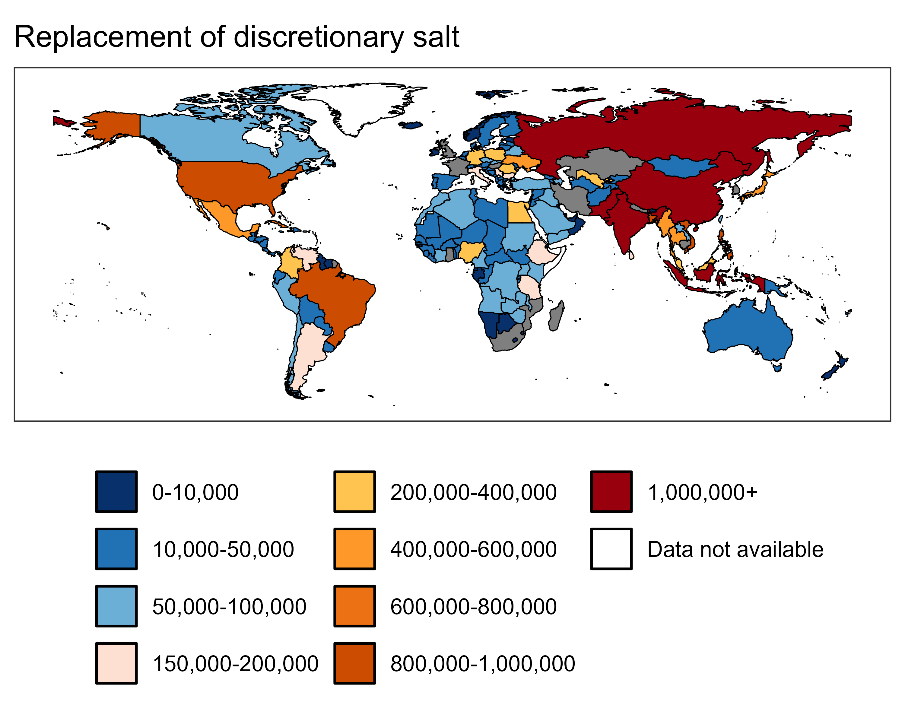

### **Supplementary figure 4. Proportion of national new cases from cardiovascular disease and kidney disease prevented under different implementation scenarios for potassium-enriched salt, for 193 countries and territories**

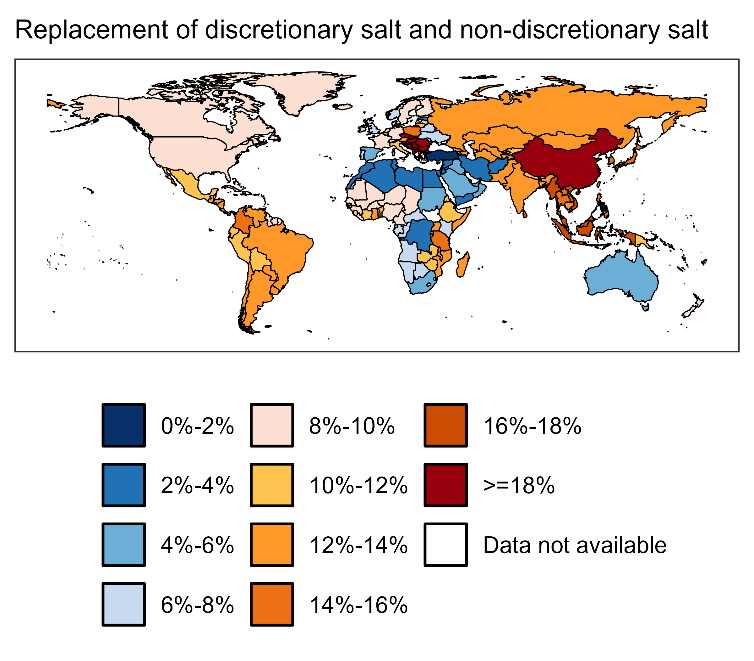

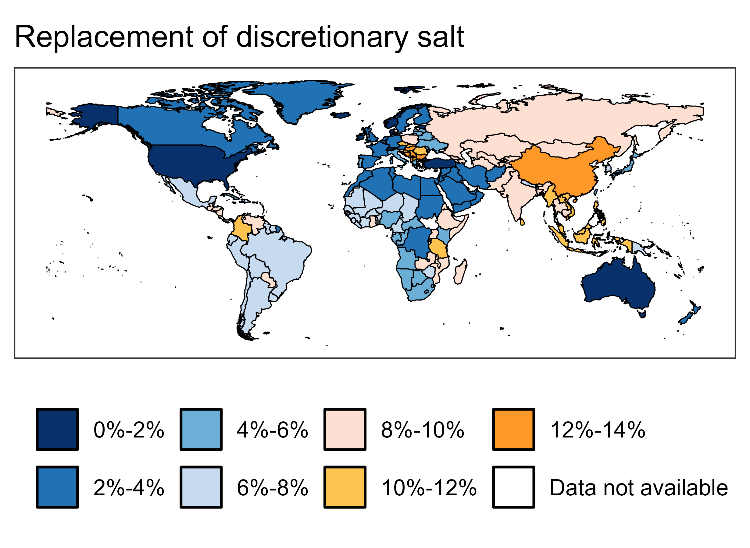

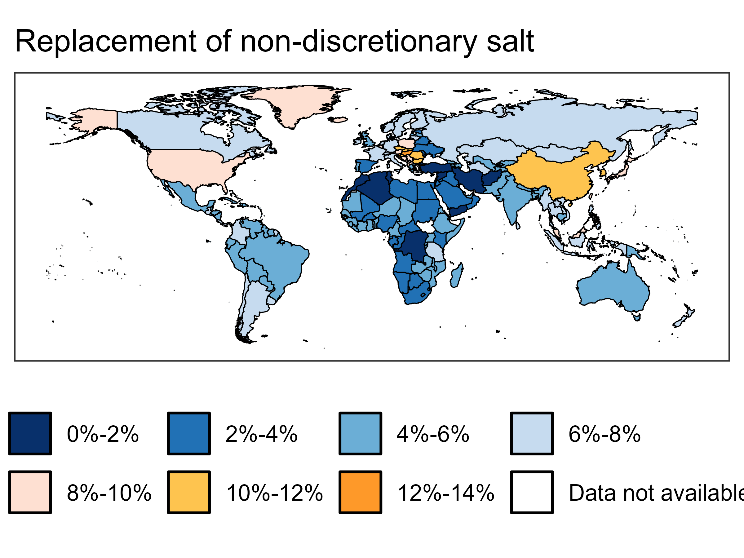

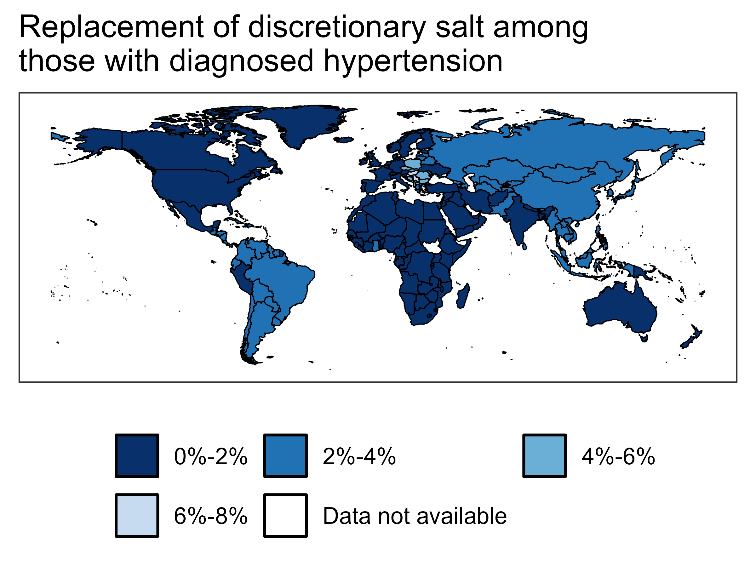

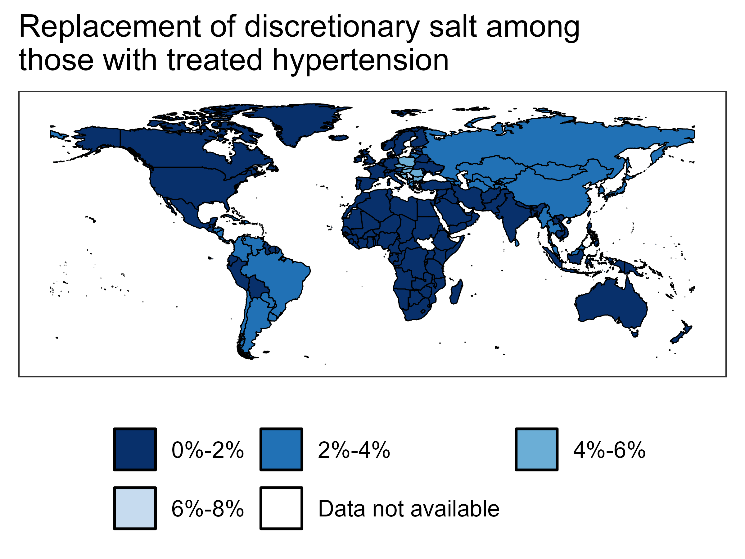

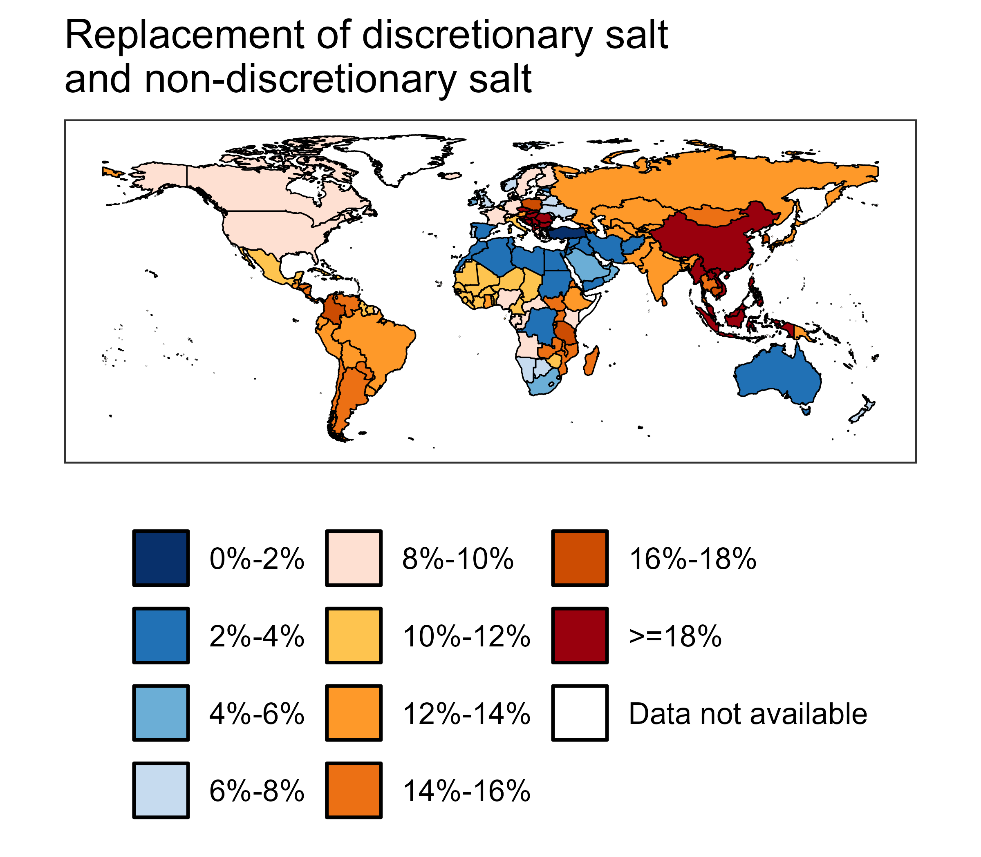

**Legend**

### **Supplementary figure 5. Proportion of national disability adjusted life years from cardiovascular disease and kidney disease prevented under different implementation scenarios for potassium-enriched salt, for 193 countries and territories**

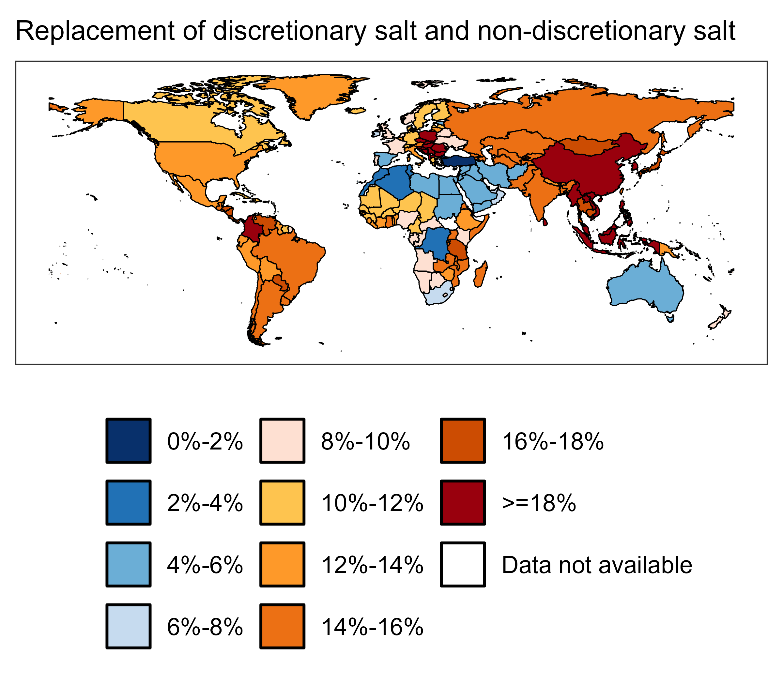

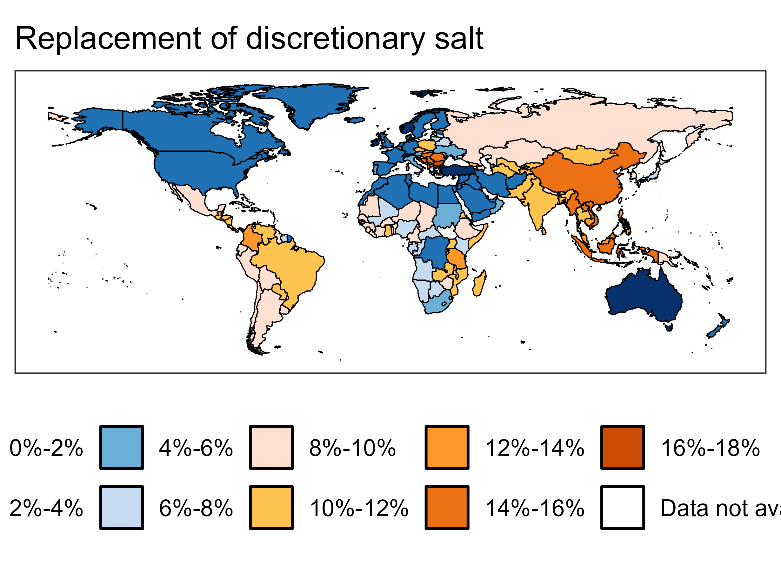

**Legend**

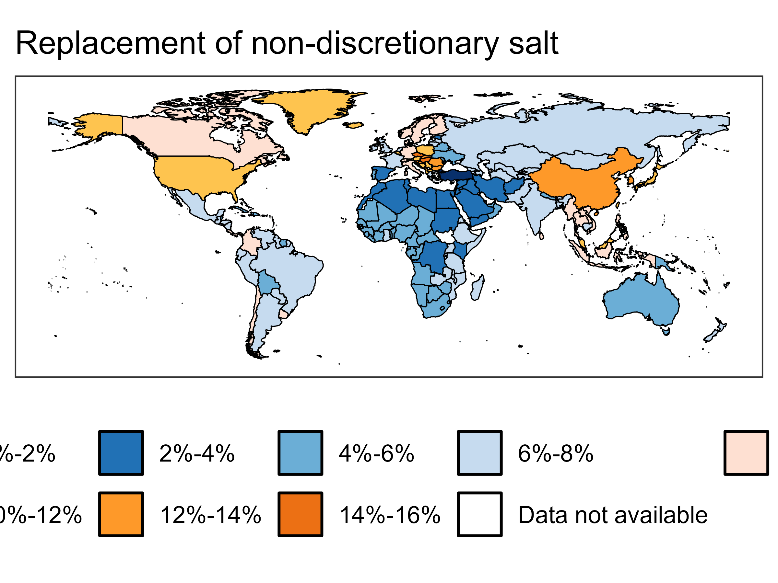

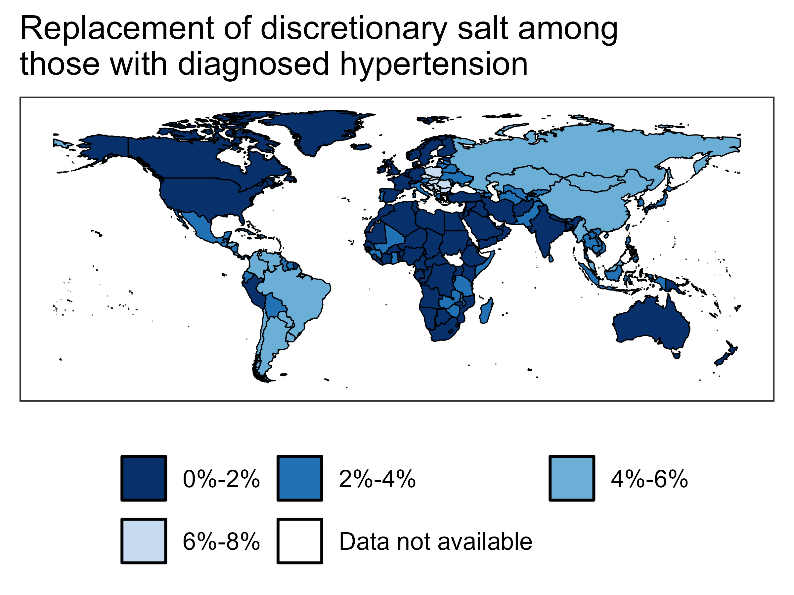

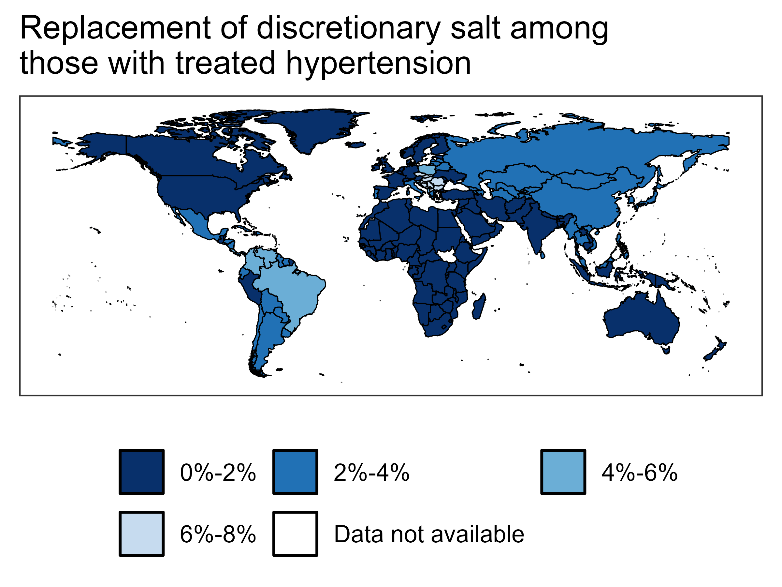

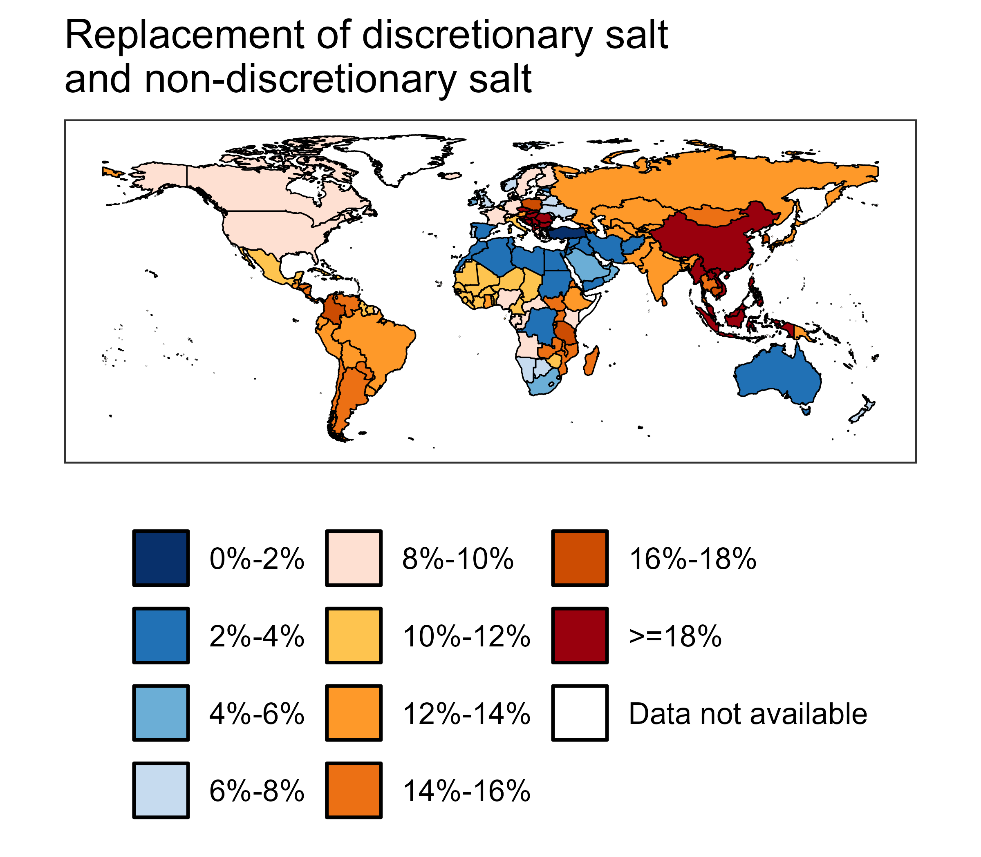

### **Supplementary figure 6. Ten countries with the most averted deaths by replacing regular salt with potassium-enriched salt, by scenario**

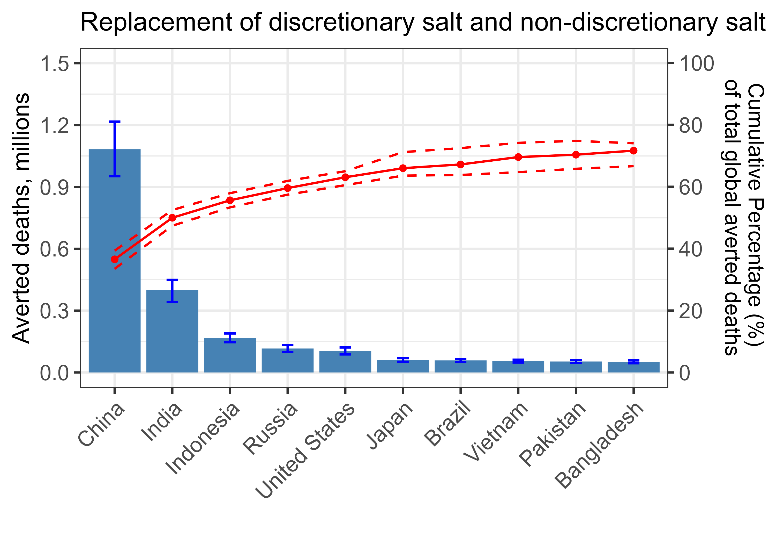

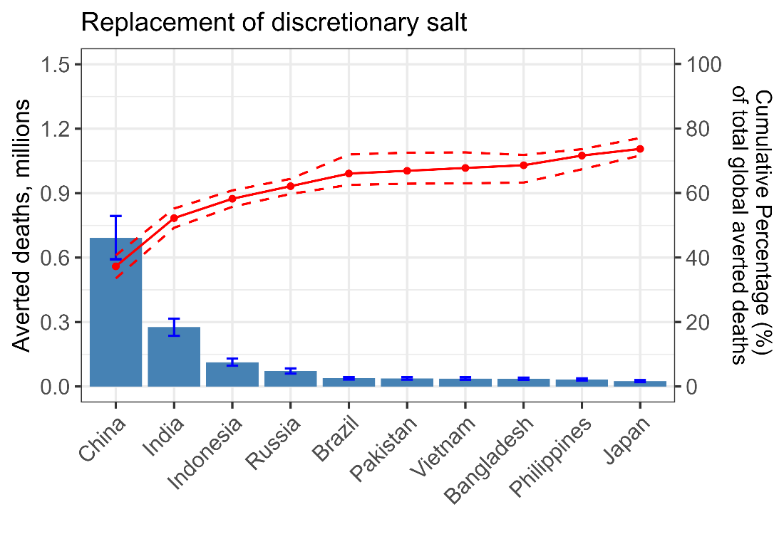

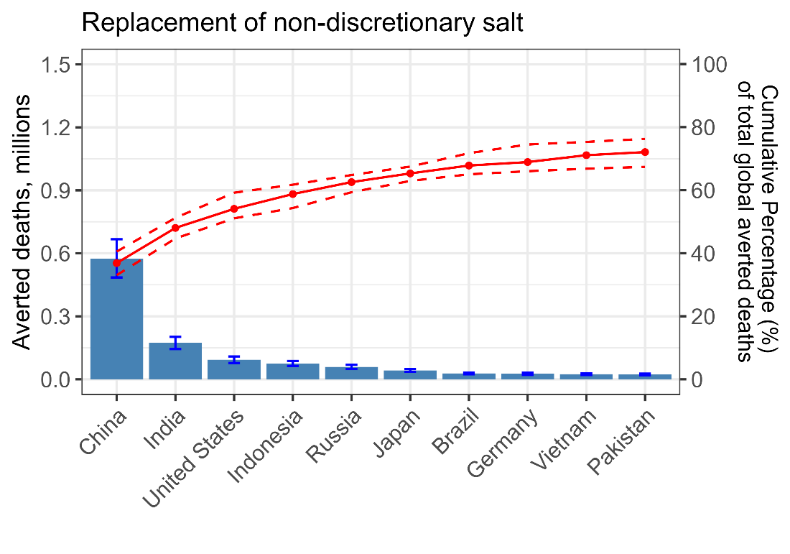

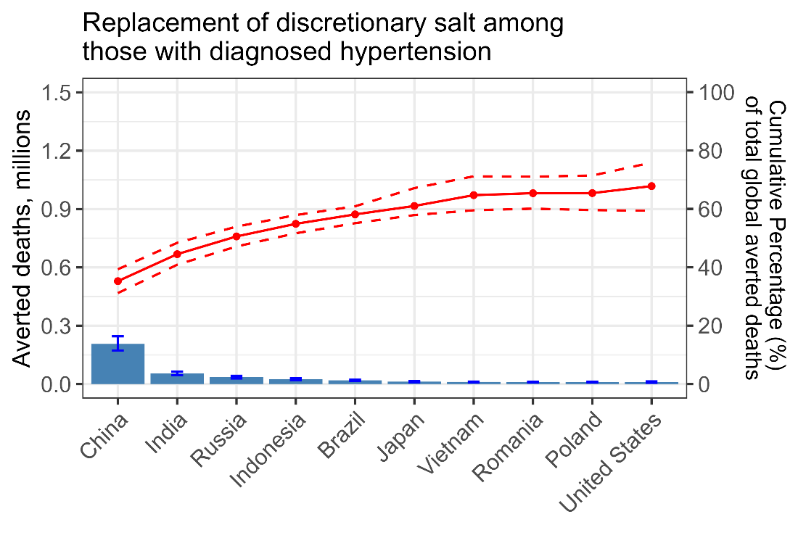

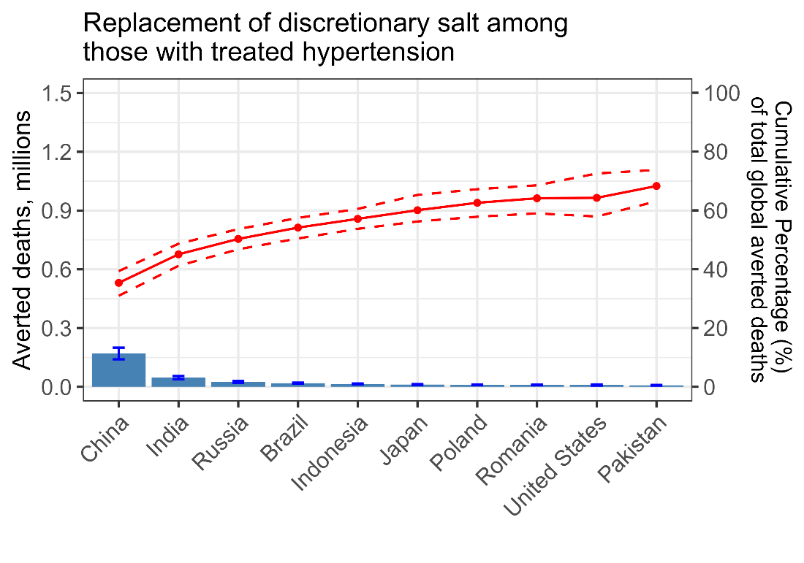

The left-hand axis, the blue columns and the error bars show the number (and 95% uncertainty interval) for CVD and CKD deaths prevented by the intervention strategy in each country. The right-hand axis and the red lines show the cumulative proportion (solid lines) and 95% uncertainty intervals (dotted lines) for all deaths worldwide prevented by the intervention strategy as each country is included moving from left to right.

### **Supplementary figure 7. Ten countries with the most new cases averted by replacing regular salt with potassium-enriched salt**

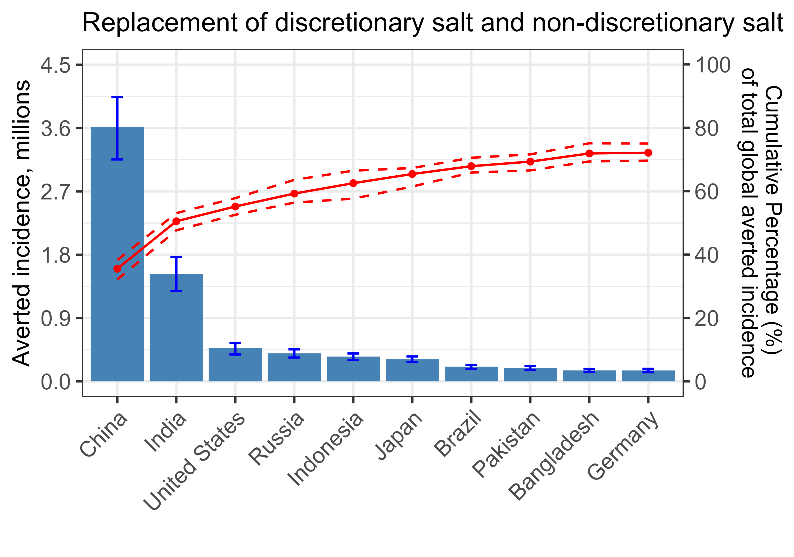

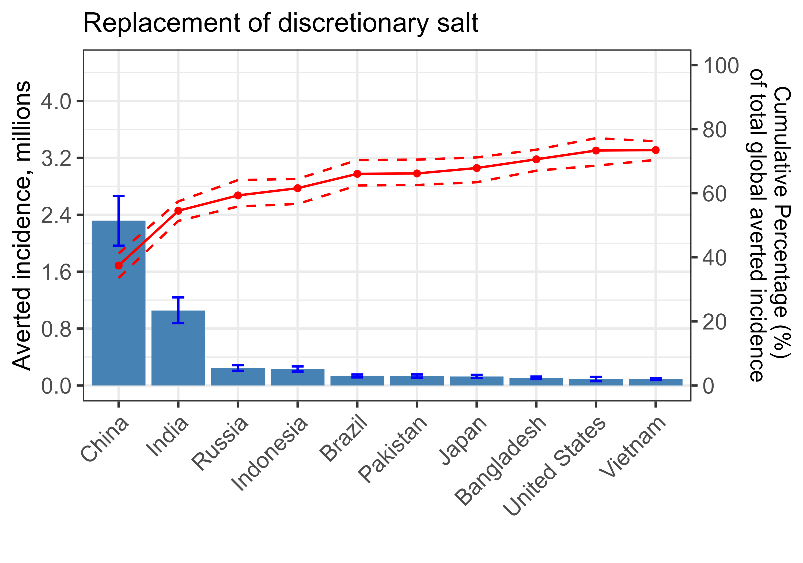

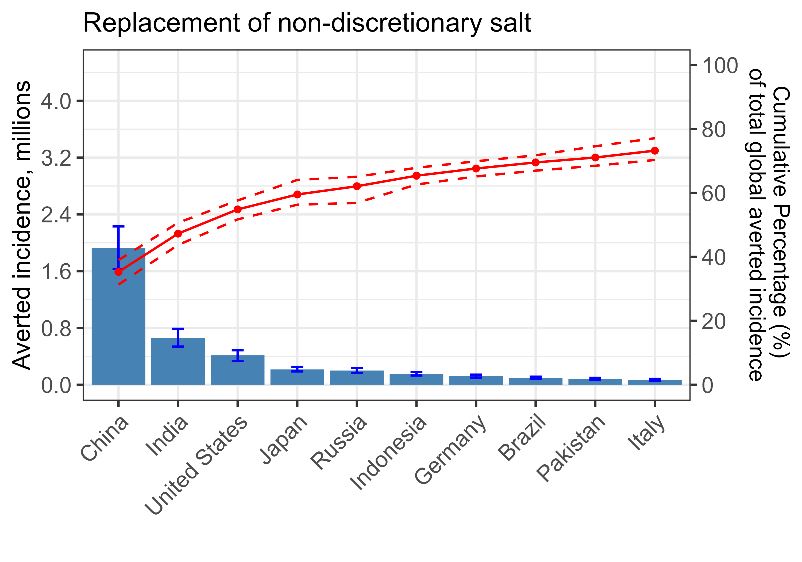

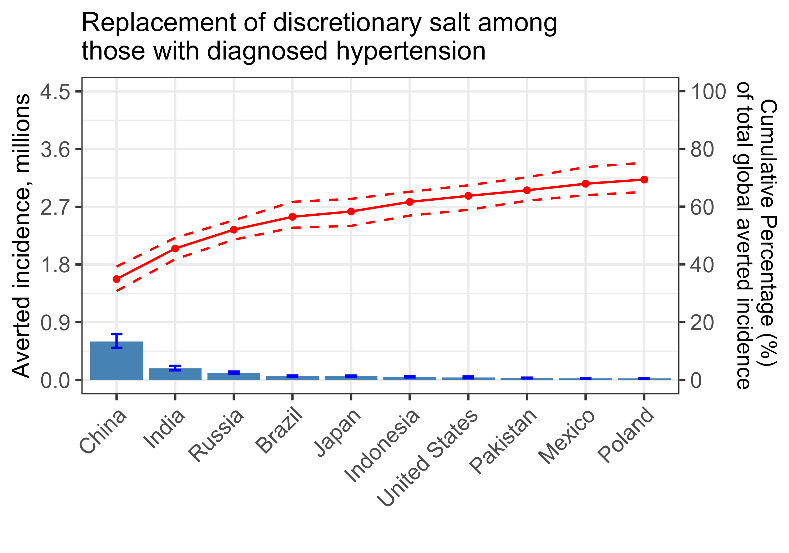

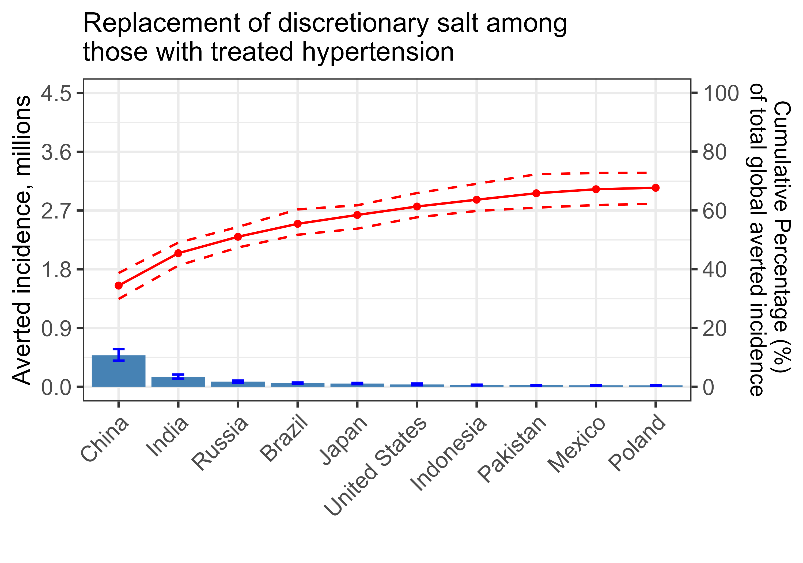

The left-hand axis, the blue columns and the error bars show the number (and 95% uncertainty interval) of new cases (both CVD and CKD) prevented by the intervention strategy in each country. The right-hand axis and the red lines show the cumulative proportion (solid lines) and 95% uncertainty intervals (dotted lines) for all non-fatal events worldwide prevented by the intervention strategy as each country is included moving from left to right.

### **Supplementary figure 8. Ten countries with the most averted disability-adjusted life years by replacing regular salt with potassium-enriched salt**

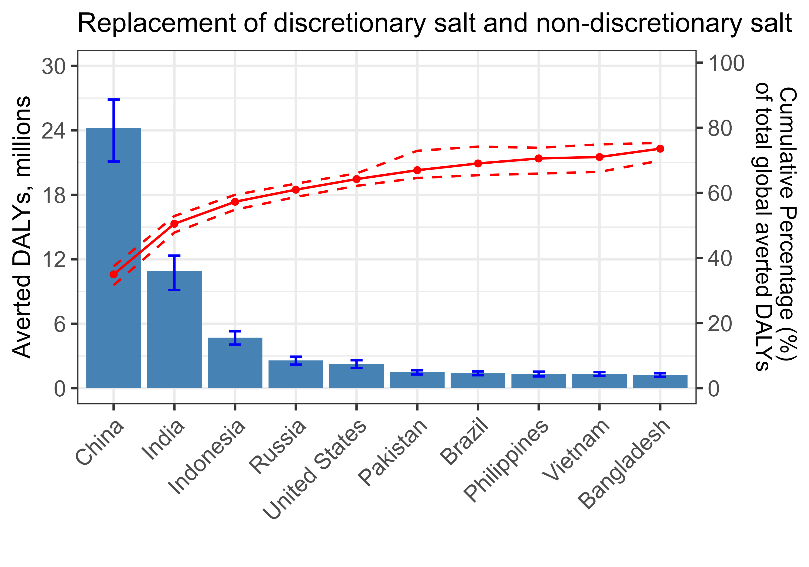

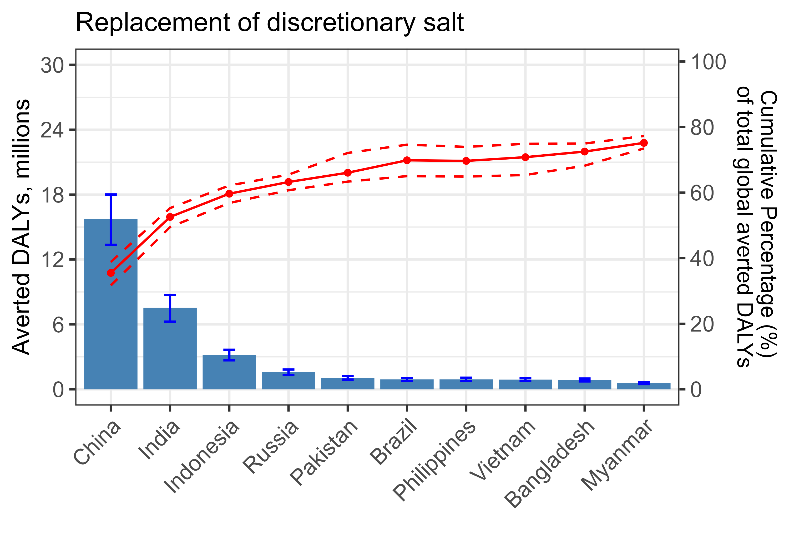

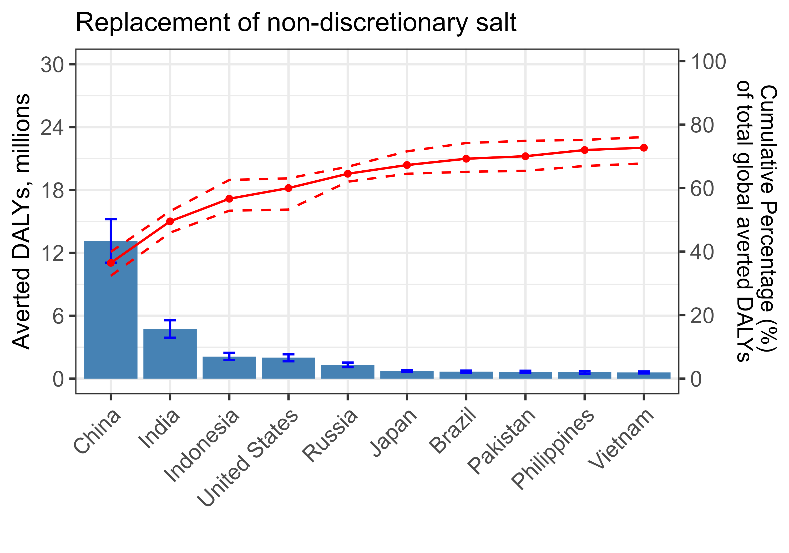

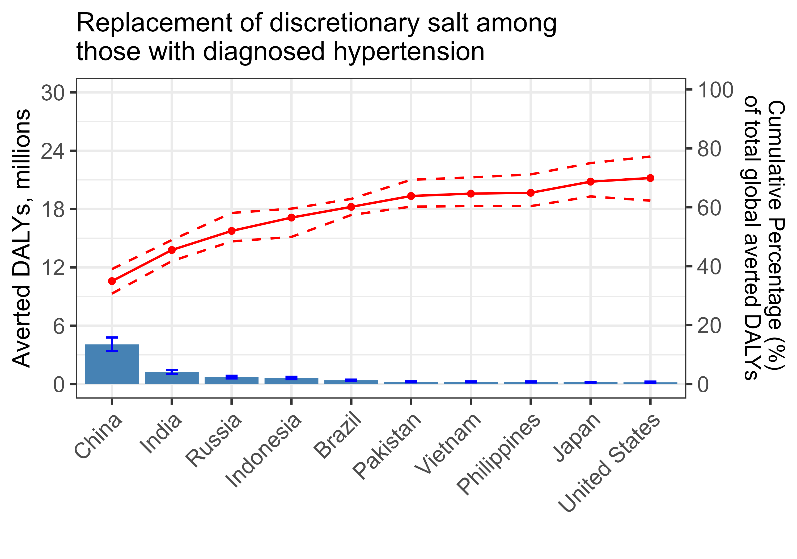

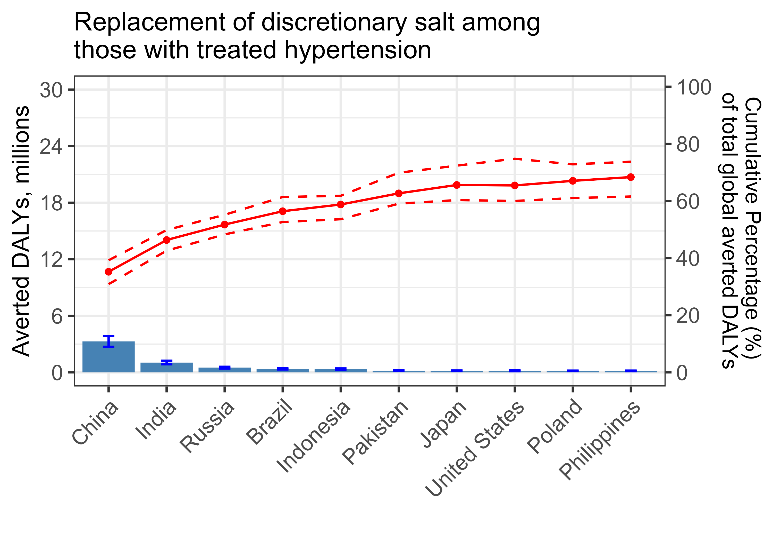

The left-hand axis, the blue columns and the error bars show the number (and 95% uncertainty interval) of disability adjusted life years prevented by the intervention strategy in each country. The right-hand axis and the red lines show the cumulative proportion (solid lines) and 95% uncertainty intervals (dotted lines) for all disability adjusted life years attributable to CVD and CKD worldwide prevented by the intervention strategy as each country is included moving from left to right.

### **Supplementary figure 9. Pre- and post- implementation serum potassium level at different baseline sodium intake under different population wide implementation scenarios**

#### a. **Pre- and post- implementation serum potassium level at different baseline sodium intake under full replacement of both discretionary and non-discretionary salt***

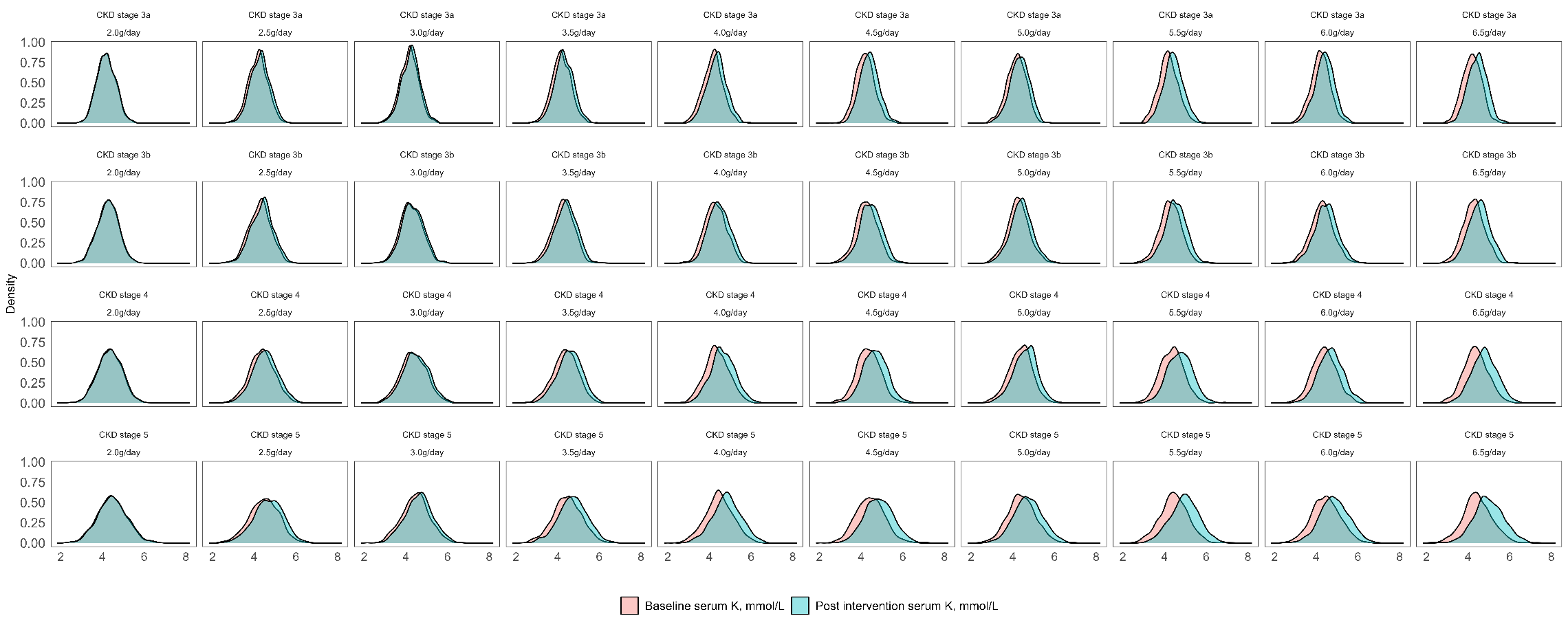

^*Only one age and sex group at each level of sodium intake were chosen to be plotted^

#### b. **Pre- and post- implementation serum potassium level at different baseline sodium intake under full replacement of discretionary salt***

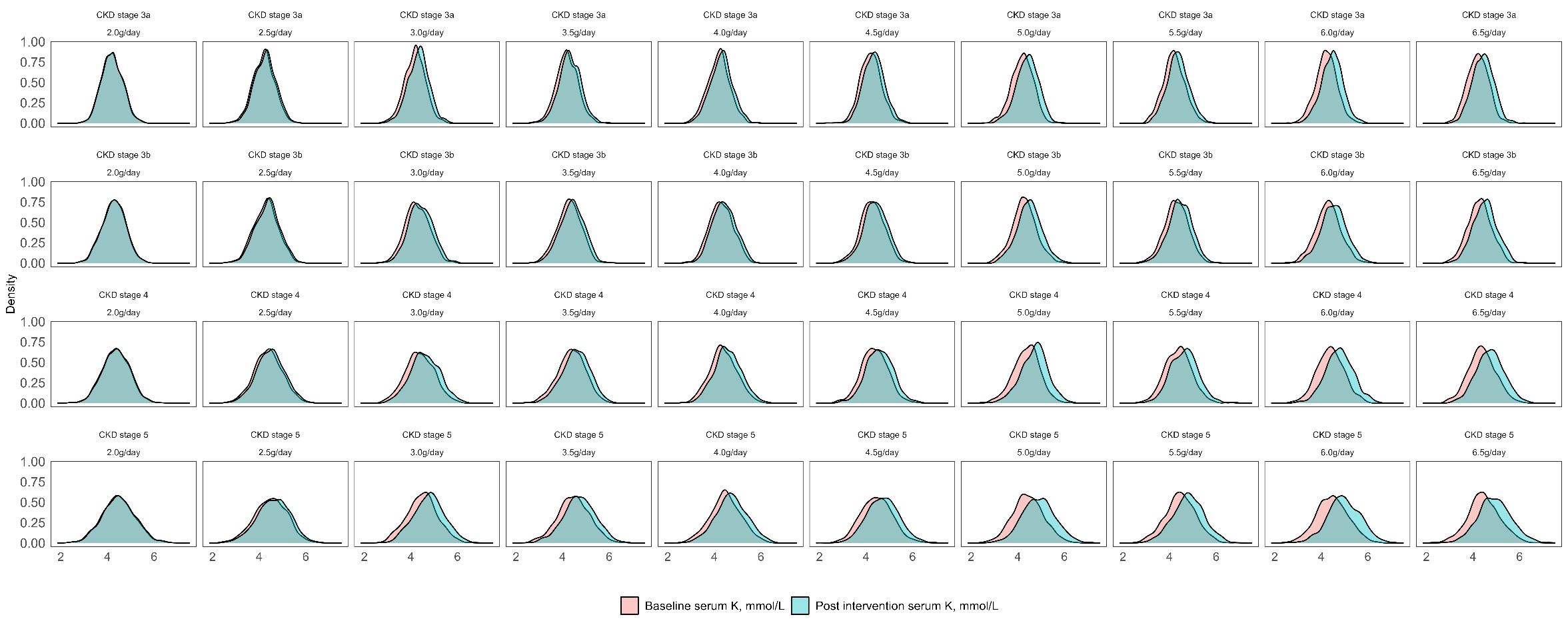

^*Only one age and sex group at each level of sodium intake were chosen to be plotted^

#### c. **Pre- and post- implementation serum potassium level at different baseline sodium intake under full replacement of non-discretionary salt***

^*Only one age and sex group at each level of sodium intake were chosen to be plotted^

### **References**

1.	Guideline: Potassium intake for adults and children. Geneva: World Health Organization; 2012.

2.	Eaton SB, Konner M. Paleolithic nutrition. A consideration of its nature and current implications. *N Engl J Med* 1985; **312**(5): 283–9.

3.	MacKinnon L, Haque O. TeachMePhysiology-Internal Balance of Potassium. 2025. <https://teachmephysiology.com/biochemistry/electrolytes/internal-balance-potassium/> (accessed January 20 2026).

4.	Dogan MF, Yildiz O, Arslan SO, Ulusoy KG. Potassium channels in vascular smooth muscle: a pathophysiological and pharmacological perspective. *Fundam Clin Pharmacol* 2019; **33**(5): 504–23.

5.	Su XT, Yang CL, Ellison DH. Kidney Is Essential for Blood Pressure Modulation by Dietary Potassium. *Curr Cardiol Rep* 2020; **22**(10): 124.

6.	Jackson WF. Potassium Channels in Regulation of Vascular Smooth Muscle Contraction and Growth. *Adv Pharmacol* 2017; **78**: 89–144.

7.	Hoorn EJ, Subramanya AR, Boyd-Shiwarski CR. Potassium and the kidney. *Nat Rev Nephrol* 2026.

8.	Intersalt: an international study of electrolyte excretion and blood pressure. Results for 24 hour urinary sodium and potassium excretion. Intersalt Cooperative Research Group. *BMJ* 1988; **297**(6644): 319–28.

9.	Mente A, O'Donnell MJ, Rangarajan S, et al. Association of urinary sodium and potassium excretion with blood pressure. *N Engl J Med* 2014; **371**(7): 601–11.

10.	Khaw KT, Barrett-Connor E. Dietary potassium and blood pressure in a population. *Am J Clin Nutr* 1984; **39**(6): 963–8.

11.	Li M, Yan S, Li X, et al. Association between blood pressure and dietary intakes of sodium and potassium among US adults using quantile regression analysis NHANES 2007-2014. *J Hum Hypertens* 2020; **34**(5): 346–54.

12.	Ziaei R, Askari G, Foshati S, Zolfaghari H, Clark CCT, Rouhani MH. Association between urinary potassium excretion and blood pressure: A systematic review and meta-analysis of observational studies. *J Res Med Sci* 2020; **25**: 116.

13.	D'Elia L, Barba G, Cappuccio FP, Strazzullo P. Potassium intake, stroke, and cardiovascular disease a meta-analysis of prospective studies. *J Am Coll Cardiol* 2011; **57**(10): 1210–9.

14.	Aburto NJ, Hanson S, Gutierrez H, Hooper L, Elliott P, Cappuccio FP. Effect of increased potassium intake on cardiovascular risk factors and disease: systematic review and meta-analyses. *BMJ* 2013; **346**: f1378.

15.	Larsson SC, Orsini N, Wolk A. Dietary potassium intake and risk of stroke: a dose-response meta-analysis of prospective studies. *Stroke* 2011; **42**(10): 2746–50.

16.	D'Elia L, Iannotta C, Sabino P, Ippolito R. Potassium-rich diet and risk of stroke: updated meta-analysis. *Nutr Metab Cardiovasc Dis* 2014; **24**(6): 585–7.

17.	Filippini T, Violi F, D'Amico R, Vinceti M. The effect of potassium supplementation on blood pressure in hypertensive subjects: A systematic review and meta-analysis. *Int J Cardiol* 2017; **230**: 127–35.

18.	Whelton PK, He J, Cutler JA, et al. Effects of oral potassium on blood pressure. Meta-analysis of randomized controlled clinical trials. *JAMA* 1997; **277**(20): 1624–32.

19.	Granal M, Sourd V, Burnier M, Fauvel JP, Gougeon A. Effect of changes in potassium intake on blood pressure: a dose-response meta-analysis of randomized clinical trials (2000-2024). *Clin Kidney J* 2025; **18**(7): sfaf173.

20.	Filippini T, Naska A, Kasdagli MI, et al. Potassium Intake and Blood Pressure: A Dose-Response Meta-Analysis of Randomized Controlled Trials. *J Am Heart Assoc* 2020; **9**(12): e015719.

21.	Rodrigues S, Baldo M, Machado R, Forechi L, Molina Mdel C, Mill J. High potassium intake blunts the effect of elevated sodium intake on blood pressure levels. *J Am Soc Hypertens* 2014; **8**(4): 232–8.

22.	Gritter M, Rotmans J, Hoorn E. Role of Dietary K(+) in Natriuresis, Blood Pressure Reduction, Cardiovascular Protection, and Renoprotection. *Hypertension* 2019; **73**(1): 15–23.

23.	Sacks F, Svetkey L, Vollmer W, et al. Effects on blood pressure of reduced dietary sodium and the Dietary Approaches to Stop Hypertension (DASH) diet. DASH-Sodium Collaborative Research Group. *N Engl J Med* 2001; **344**(1): 3–10.

24.	Du S, Batis C, Wang H, Zhang B, Zhang J, Popkin B. Understanding the patterns and trends of sodium intake, potassium intake, and sodium to potassium ratio and their effect on hypertension in China. *The American journal of clinical nutrition* 2014; **99**(2): 334–43.

25.	Binia A, Jaeger J, Hu Y, Singh A, Zimmermann D. Daily potassium intake and sodium-to-potassium ratio in the reduction of blood pressure: a meta-analysis of randomized controlled trials. *J Hypertens* 2015; **33**(8): 1509–20.

26.	Rhee M, Shin S, Gu N, et al. Relationship between 24-h urine sodium/potassium ratio and central aortic systolic blood pressure in hypertensive patients. *Hypertens Res* 2016.

27.	Jayedi A, Ghomashi F, Zargar M, Shab-Bidar S. Dietary sodium, sodium-to-potassium ratio, and risk of stroke: A systematic review and nonlinear dose-response meta-analysis. *Clin Nutr* 2018.

28.	Perez V, Chang E. Sodium-to-potassium ratio and blood pressure, hypertension, and related factors. *Adv Nutr* 2014; **5**(6): 712–41.

29.	Rhee M, Lee S. Reduction of dietary sodium/potassium ratio is more effective in lowering of nighttime blood pressure. *European heart journal Conference: european society of cardiology, ESC congress 2017 Spain* 2017; **38**(Supplement 1): 1366.

30.	Pereira T, Mill J, Griep R, Sichieri R, Molina M. Effect of urinary sodium-to-potassium ratio change on blood pressure in participants of the longitudinal health of adults study - ELSA-Brasil. *Medicine (Baltimore)* 2019; **98**(28): e16278.

31.	Mozaffarian D, Fahimi S, Singh GM, et al. Global sodium consumption and death from cardiovascular causes. *N Engl J Med* 2014; **371**(7): 624–34.

32.	Yin X, Rodgers A, Perkovic A, et al. Effects of salt substitutes on clinical outcomes: a systematic review and meta-analysis. *Heart* 2022; **108**(20): 1608–15.

33.	Huang L, Trieu K, Yoshimura S, et al. Effect of dose and duration of reduction in dietary sodium on blood pressure levels: systematic review and meta-analysis of randomised trials. *BMJ* 2020; **368**: m315.

34.	Turban S, Juraschek SP, Miller ER, 3rd, et al. Randomized Trial on the Effects of Dietary Potassium on Blood Pressure and Serum Potassium Levels in Adults with Chronic Kidney Disease. *Nutrients* 2021; **13**(8).

35.	Ogata S, Akashi Y, Kato S, et al. Association Between Dietary Potassium Intake Estimated From Multiple 24-Hour Urine Collections and Serum Potassium in Patients With CKD. *Kidney Int Rep* 2023; **8**(3): 584–95.

36.	Global Burden of Disease Collaborative Network. Global Burden of Disease Study 2021 (GBD 2021) Dietary Risk Exposure Estimates 1990-2021. Seattle, United States of America: Institute for Health Metrics and Evaluation (IHME); 2024.

37.	Bhat S, Marklund M, Henry ME, et al. A Systematic Review of the Sources of Dietary Salt Around the World. *Adv Nutr* 2020; **11**(3): 677–86.

38.	World Bank Group. World Bank national accounts data, and OECD National Accounts data files. 2024. <https://data.worldbank.org/indicator/NY.GDP.PCAP.CN> (accessed August 13 2024).

39.	International Monetary Fund. GDP per capita, current prices. 2021. <https://www.imf.org/external/datamapper/NGDPDPC@WEO/ADVEC/WEOWORLD/TWN/CHN> (accessed April 28 2025).

40.	N. C. D. Risk Factor Collaboration. Worldwide trends in hypertension prevalence and progress in treatment and control from 1990 to 2019: a pooled analysis of 1201 population-representative studies with 104 million participants. *Lancet* 2021; **398**(10304): 957–80.

41.	Filippini T, Malavolti M, Whelton PK, Naska A, Orsini N, Vinceti M. Blood Pressure Effects of Sodium Reduction: Dose-Response Meta-Analysis of Experimental Studies. *Circulation* 2021; **143**(16): 1542–67.

42.	Global Burden of Disease Collaborative Network. Global Burden of Disease Study 2019 (GBD 2019) Relative Risks. Seattle, United States of America: Institute for Health Metrics and Evaluation (IHME) 2020.

43.	Institute for Health Metrics and Evaluation. GBD Results. 2021. <https://vizhub.healthdata.org/gbd-results/> (accessed August 13 2024).

44.	Hallan SI, Matsushita K, Sang Y, et al. Age and association of kidney measures with mortality and end-stage renal disease. *JAMA* 2012; **308**(22): 2349–60.

45.	Global Burden of Disease Collaborative Network. Global Burden of Disease Study 2021 (GBD 2021) Demographics 1950-2021. Seattle, United States of America: Institute for Health Metrics and Evaluation (IHME); 2024.

46.	Matsushita K, Mahmoodi BK, Woodward M, et al. Comparison of risk prediction using the CKD-EPI equation and the MDRD study equation for estimated glomerular filtration rate. *JAMA* 2012; **307**(18): 1941–51.

47.	Kovesdy CP, Matsushita K, Sang Y, et al. Serum potassium and adverse outcomes across the range of kidney function: a CKD Prognosis Consortium meta-analysis. *Eur Heart J* 2018; **39**(17): 1535–42.
